# Phylodynamics reveals the role of human travel and contact tracing in controlling the first wave of COVID-19 in four island nations

**DOI:** 10.1101/2020.08.04.20168518

**Authors:** Jordan Douglas, Fábio K. Mendes, Remco Bouckaert, Dong Xie, Cinthy L. Jiménez-Silva, Christiaan Swanepoel, Joep de Ligt, Xiaoyun Ren, Matt Storey, James Hadfield, Colin R. Simpson, Jemma L. Geoghegan, Alexei J. Drummond, David Welch

## Abstract

**Background:** New Zealand, Australia, Iceland, and Taiwan all saw success at controlling the first wave of the COVID-19 pandemic. As islands, they make excellent case studies for exploring the effects of international travel and human movement on the spread of COVID-19.

**Methods:** We employed a range of robust phylodynamic methods and genome subsampling strategies to infer the epidemiological history of SARS-CoV-2 in these four countries. We compared these results to transmission clusters identified by the New Zealand Ministry of Health by contract tracing strategies.

**Findings:** We estimated the effective reproduction number of COVID-19 as 1–1.4 during early stages of the pandemic, and show that it declined below 1 as human movement was restricted. We also showed that this disease was introduced many times into each country, and that introductions slowed down markedly following the reduction of international travel in mid March 2020. Finally, we confirmed that New Zealand transmission clusters identified via standard health surveillance strategies largely agree with those defined by genomic data.

**Interpretation:** We have demonstrated how the use of genomic data and computational biology methods can assist health officials in characterising the epidemiology of viral epidemics, and for contact tracing.

**Funding:** This research was funded by the Health Research Council of New Zealand, the Ministry of Business, Innovation, and Employment, the Royal Society of New Zealand, and the New Zealand Ministry of Health.

**Research in Context:** *Evidence before this study:* Our study looks at the early months of the COVID-19 pandemic, a period in which the first wave was controlled in four “island” nations – New Zealand, Australia, Taiwan, and Iceland. All prior data used in this study was collected from late 2019 until the end of April 2020. This includes over 3000 SARS-CoV-2 genomic sequences which were collected in this period (and subsequently deposited into GISAID), as well as arrival and departure information (provided by official statistics from each country), human mobility data collected from mobile phones (by Apple), and COVID-19 case data (released by the World Health Organisation). Even early on during the COVID-19 pandemic, the properties of SARS-CoV-2 – including the reproduction number and mutation rate – were well characterised, and a range of these estimates have been covered in our article. Our Bayesian phylodynamic models, including their prior distributions, are informed by all of the above sources of information. Finally, we have incorporated all of the available information on COVID-19 transmission clusters identified by the New Zealand Ministry of Health during this period.

*Added value of this study:* We quantified the decline in the reproduction number of SARS-CoV-2, following the decline in human mobility, in four “island” countries. We also demonstrated how importation events of SARS-CoV-2 into each considered country declined markedly following the reduction of international travel. Our results shed a different light on these patterns because of (i) our locations of choice – the four countries had success in dealing with the first pandemic wave, with their geographic isolation contributing to cleaner signals of human mobility, and (ii) our novel and empirically driven phylodynamic model, which we built from explicitly modelling mobile phone data in the four islands. Furthermore, by crossing epidemiological against ge3nomic data, our paper quantitatively assesses the ability of contact tracing, as implemented by the New Zealand Ministry of Health (NZMH), in identifying COVID-19 transmission clusters. We find evidence for a high efficacy of the specific measures taken – and when they were taken – by the NZMH in identifying transmission clusters, considered worldwide to have been successful in its response to the pandemic. Our analyses also illustrate the power of viral genomic data in assisting contact tracing.

*Implications of all the available evidence:* The conclusions drawn from this research inform effective policy for locations pursuing an elimination strategy. We confirm the accuracy of standard contact tracing methods at identifying clusters and show how these methods are improved using genomic data. We demonstrate how the overseas introduction rates and domestic transmission rates of an infectious viral agent can be surveilled using genomic data, and the important role each plays in overall transmission. Specifically, we have quantified these processes for four countries and have shown that they did decline significantly following declines in human travel and mobility. The phylodynamic methods used in this work is shown to be robust and applicable to a range of scenarios where appropriate subsampling is used.

## Introduction

> Many continued hoping that the epidemic would soon die out and they and their families be spared. Thus they felt under no obligation to make any change in their habits, as yet. Plague was an unwelcome visitant, bound to take its leave one day as unexpectedly as it had come.
>
> — Albert Camus (“The Plague”, 1947)

The respiratory tract illness caused by Severe acute respiratory syndrome coronavirus 2 (SARS-CoV-2) known as Coronavirus Disease 2019 (COVID-19; (Rothan and Byrareddy, 2020)) has caused substantial global health and economic impact, and the global epidemic appears far from over. COVID-19 was first detected in the city of Wuhan, Hubei province (China) in late 2019 (Zhu et al., 2020). By 11 March 2020 there were 100,000 cases across 114 countries leading to the World Health Organization (WHO) declaring a pandemic (World Health Organization, 2020a). By early 2021, over two million deaths had been reported, and over half of the world’s population had received stay-at-home orders (Sandford, 2020).

COVID-19 has now been characterised extensively. The average number of people each case goes on to infect (known as the effective reproduction number, *R*_e_), for example, has been estimated as 1–6 during the early stages of the outbreak (Liu et al., 2020; Binny et al., 2020a; Alimohamadi et al., 2020). Because COVID-19 often causes mild symptoms Day (2020a,b) and surveillance efforts are variable, the difference between the true and reported number of cases could be of an order of magnitude (Li et al., 2020a; Lu et al., 2020). Fortunately, many locations have demonstrated that it is possible to reduce *R*_*e*_ to less than 1 as result of mitigation efforts including contact tracing, case isolation, and travel restrictions (Binny et al., 2020a; Chinazzi et al., 2020).

As we reflect on the early months of the pandemic (until the end of April 2020), different parts of the world tell contrasting stories about their struggle against COVID-195 (**Table 1**). Aotearoa New Zealand, Australia, Iceland, and Taiwan, for example, fared comparatively well against their “first wave”, with smaller infection peaks and lower excess mortalities compared to many mainland countries (World Health Organization, 2020b; FT Visual & Data Journalism team, 2020), with New Zealand and Taiwan achieving elimination or near-elimination of their first waves in that period (Cousins, 2020; Summers et al., 2020). Over the following months, large areas of Iceland, the city of Auckland (New Zealand), and the state of Victoria (Australia) all experienced “second waves” of community transmission (Aljazeera, 2020a,b; Review, 2020) that were eventually brought under control. Taiwan has had no second significant outbreak.

**Table 1:** Summary of the COVID-19 pandemic in the four target islands. Total passenger arrivals into the island in the Jan-Mar period are counted. All dates are in the year 2020. The number of confirmed cases and the percentage of cases with recent overseas travel are reported for the Jan-Apr period. *Visitor arrivals only.

| Island | Population<br>(millions) | Arrivals<br>(millions) | First<br>case | Border<br>close | Confirmed<br>cases | Overseas<br>travel |
| --- | --- | --- | --- | --- | --- | --- |
| New Zealand | 5 | 1.6 (NZ, 2020) | Feb 28 | Mar 19<br>(RNZ, 2020) | 1,129<br>(World Health Organization, 2020b) | 39%<br>(Ministry of Health, Manatū Hauora, 2020) |
| Australia | 25 | 4.3 (of Statistics, 2020) | Jan 25 | Mar 20<br>(Burke, 2020) | 6,746<br>(World Health Organization, 2020b) | 64% (Department of Health, 2020) |
| Iceland | 0.34 | 0.33* (Icelandic Tourist Board, 2020) | Feb 28 | Mar 20<br>(Schengen Visa Info, 2020) | 1,797<br>(World Health Organization, 2020b) | 19% (of Health et al., 2020) |
| Taiwan | 23 | 3.6 (Strong, 2020) | Jan 21 | Mar 19<br>(Aspinwall, 2020) | 429 (Taiwan Centers for Disease Control, 2020a) | 90% (Taiwan Centers for Disease Control, 2020a) |

Irrespective of their current COVID-19 situation, these four countries share in common the effectiveness of their response to the first outbreak, which contributed positively to their swift recovery at the time. In March 2020 amidst the first wave, all four nations closed their borders and required isolation or quarantine of those who were allowed through (Hale et al., 2020), either restricting or discouraging human movement (Apple, 2020). Stay-at-home orders were issued in New Zealand and Australia but not in Iceland or Taiwan (Hale et al., 2020). Stringent contact tracing, coupled with isolation and/or quarantine of cases and their close contacts, proved to be effective strategies in these four locations (Baker et al., 2020; Draper et al., 2020; Gudbjartsson et al., 2020a; Wang et al., 2020). These islands thus provide excellent case studies to characterise the effects of human travel on epidemic spread during the early months of the pandemic, a critical endeavor when facing the threat of global endemicity.

Mathematical methods for studying epidemics are as diverse as the diseases they target, making varied use of surveillance and genetic data, and ranging from models of mathematical epidemiology (Grassly and Fraser, 2008; Binny et al., 2020a; Ferretti et al., 2020; Hethcote, 2000) to phylodynamic models (Volz et al., 2013). The latter take advantage of the comparable timescales of epidemiological and evolutionary processes (Grenfell et al., 2004), which result from short viral generation times and high viral mutation rates (20–40 mutations per year for SARS-CoV-2; (Lai et al., 2020; Li et al., 2020b; Rambaut, 2020)). By modelling the transmission process as phylogenetic trees (Fig. 1), phylodynamic and molecular evolution models can be coupled to reconstruct the mutational history of viral genomes (Li et al., 2020b; Rambaut, 2020; Lai et al., 2020), elucidating the epidemiological processes that shaped them.

**Fig. 1:**
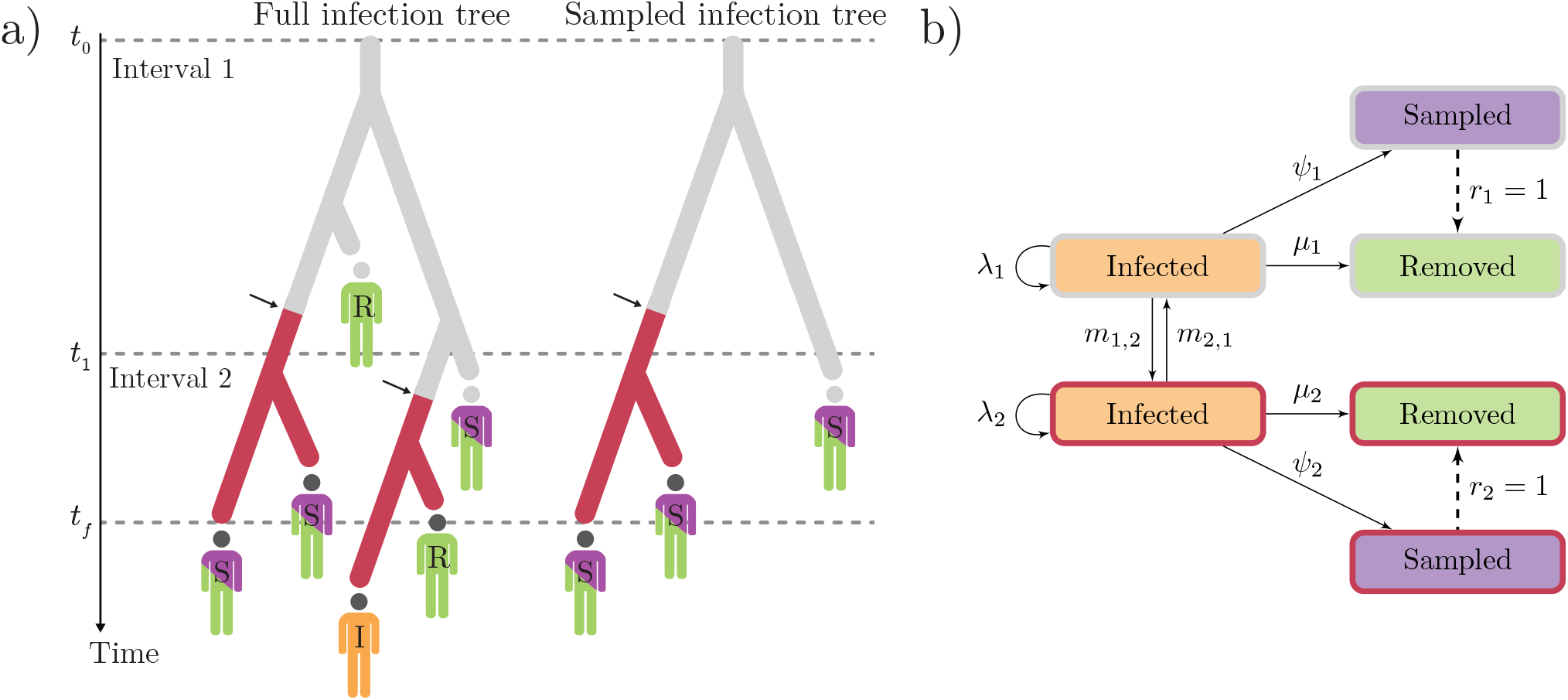
Depiction of the MTBD model. (a) A full infection tree and a sampled infection tree (𝒯), with two time intervals (epochs) and two demes (indicated in gray and red). All lineages in the trees represent infected individuals (I). Branching corresponds to transmission, colour change (arrows) to migration, and infected individuals may be removed (R), or sampled and then removed (S). (b) Transition diagram; solid and dashed arrows labelled with rates and probabilities, respectively.

In this study, we considered disease progression in New Zealand, Australia, Iceland, and Taiwan over the first few months of the pandemic up until the end of April. We hypothesised that the introduction of SARS-CoV-2 would sharply decline following that of international travel, and that viral transmission (*R*_*e*_) would decline following that of human movement. While the notion that restricting human movement constrains the spread of infectious diseases may appear exhaustively addressed, our study is unique in its integration of human mobility data with genomic and epidemiological data, in geographically isolated island settings. There is great benefit in studying the spread of disease in these locations, as their geographic isolation act as a buffer to the statistical noise otherwise introduced by more regular viral imports in mainland countries. We do so by analysing viral genomic data under a range of phylogenetic methods, while at the same time controlling for data subsampling strategies. Lastly, we compare the phylodynamic history of New Zealand’s first outbreak with that inferred by the New Zealand Ministry of Health in order to assess the ability of contact tracing at identifying transmission pathways.

## Methods

In the study of disease phylodynamics, a complete epidemic can be modelled as a rooted, binary phylogenetic tree whose *N* tips are all infected individuals (Fig. 1), and whose characteristics and drivers we wish to infer. In a rapidly growing epidemic the age of this tree is a good proxy for the duration of the infection in the population. Because it is seldom possible to sample the entire infected population, one instead reconstructs a subtree 𝒯 from a sample of c7ases of size *n*, where *n* < *N*. This sample provides *n* viral genome sequences we refer to as *D*, as well as each sequence’s sampling date, collectively represented by vector *y. D* and *y* thus constitute the input data, and are herein referred to as the “samples”. In the case of structured phylodynamic models, samples in *y* are further annotated by “deme” – a geographical unit such as a city, a country, or a continent.

A Bayesian phylodynamic model is characterised by the following unnormalised posterior probability density function:

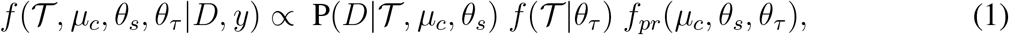

where P(*D*| 𝒯, *µ*_*c*_, *θ*_*s*_) is the phylogenetic likelihood of tree 𝒯 given alignment *D*, whose sequences accumulate substitutions at rate *µ*_*c*_ under a substitution model with parameters *θ*_*s*_. *f* (𝒯 |*θ*_*τ*_) is the probability density function of the tree prior sampling distribution, given tree prior parameters *θ*_*τ*_ (in this study we consider a range of phylodynamic models that differ in their tree prior; see below and Supplementary materials). Finally, *f*_*pr*_(*µ*_*c*_, *θ*_*s*_, *θ*_*τ*_) is the prior distribution on the remaining parameters.

### Multitype birth-death model

The multitype birth-death (MTBD) model (Kühnert et al., 2016; Sciré et al., 2020) is a structured phylodynamic model that assumes infected (“**I**”; Fig. 1) hosts of viral lineages are organised into a set of demes (“types”), and migrate between them at rate *m* (Fig. 1). Infected individuals transmit the infection at birth rate *λ*, and can be sampled (“**S**”; Fig. 1) and sequenced at sampling rate *ψ*, after which they are removed (“**R**”; Fig. 1) with probability *r*. We set *r* = 1 by assuming (sampled) confirmed cases are isolated and are unlikely to cause further infections. Removal without sampling happens at death rate *µ* (Fig. 1). Here, “death” refers to anything causing hosts to no longer be infectious, including recovery, host death, or behavioural changes (e.g., self-isolation).

The probability density function of 𝒯 under the MTBD sampling distribution (Kühnert et al., 2016) and its parameters *θ*_*τ*_ is given by:

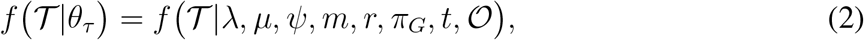

where *π*_*G*_ represents the frequency of lineages in each deme at the root, *and 𝒪* is tree 𝒯 ‘s age.

The MTBD model is reparameterised into a computationally and epidemiologically convenient form, with parameters 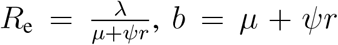, and 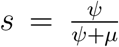. *R*_e_ is the disease’s effective reproductive number (Stadler et al., 2013). *b* is the total rate at which individuals become non-infectious, and *s* is the probability of an individual being a sample (out of all removed individuals). The phylodynamic tree prior under this parameterisation is:

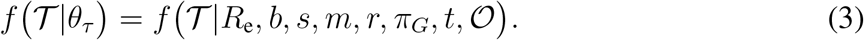

### Alternate phylogenetic models

As a sensitivity analysis, we invoke three additional phylogenetic models on top of MTBD and compare the results between the four models. These models are 1) a discrete phylogeography model (DPG, (Lemey et al., 2009)), 2) a discrete phylogeography model with informed effective population sizes and epochs (DPG+), and 3) the structured coalescent model with informed effective population sizes and epochs (SC, (Müller et al., 2018)). Details of these models are in the Supporting Information.

### SARS-CoV-2 genomic data and subsampling

We sequenced and assembled 217 New Zealand SARS-CoV-2 genomes (representing 19% of confirmed cases during the relevant time period) that are now available on GISAID (Shu and McCauley, 2017), which provided the remaining 3000+ genome sequences analysed here. Due to computational limitations, we subsampled GISAID sequences as follows: (i) two different subsample sizes (“small” vs. “large”), with 200 and 500 ℛ𝒲 (“rest of the world” deme) sequences, and (ii) two sample selection methods (“time” vs. “active”) that chose sequences uniformly across time, or proportionally to the number of active cases over time. Up to 250 sequences from the ℐ𝒮 (island) deme are included in each alignment (where more than 250 were available, we subsampled them using the respective sampling method). We restricted our analyses to the early months of the pandemic (until the end of April 2020) because this time window encompasses largely resolved (i.e., from onset to end) outbreaks in all four countries, while also making dataset sizes more manageable, precluding further subsampling and thinning of data points.

Permutating the two options above yields four subsampling protocols (detailed in SI Appendix), which were applied to the four *ℐ* 𝒮 demes, generating a total of 16 SARS-CoV-2 alignments (30 kb long, 0.3% ambiguous sites on average). Each alignment consists of sequences from one of the four islands (*ℐ𝒮*) and from the rest of the world (*ℛ* 𝒲). Moreover, each alignment was analysed with each phylodynamic models (MTBD, SC, DPG, and DPG+), coming to a total of 48 analyses.

### Demes and infection time intervals

We consider four independent geographical models, each characterised by two demes: a target island deme (New Zealand, Australia, Iceland, or Taiwan; referred to as the “*ℐ*𝒮” deme) and a mutually exclusive “rest-of-the-world” deme (“*ℛ* 𝒲”), which contributes samples from other countries in the world. All *ℛ* 𝒲 specific parameters are considered nuisance parameters.

Under our MTBD model, *R*_*e*_, *b, s*, and *m* are estimated across a series of time intervals (and held constant within intervals) defined by time boundaries *t*, with *t*_0_ < *t*_1_ < … < *t*_*f*_, where *t* is fixed at informed dates. *t*_0_ is the beginning of the infection, and *t*_*f*_ is both the last sampling time and the end of the last interval (Fig. 1). Time intervals are both parameter- and deme-specific.

*R*_*e*_ and *b* are modelled with three epochs *t*_0_ < *t*_1_ < *t*_2_ < *t*_*f*_, where *t*_1_ is the first reported case in each deme and *t*_2_ is the point of human mobility decline, according to a Bayesian hierarchical sigmoidal model trained on Apple mobility data (see Results and Supporting Information). Epoch dates for *s* and *m* can be found in Table S3.

## Results

### Modelling human movement as tracked by mobile phone data

With the goal of characterising the decrease in human movement over the weeks our SARS-CoV-2 samples were collected, we employed a novel Bayesian hierarchical sigmoidal model for the analysis of mobile phone data (Apple, 2020; Fig. 2). This analysis revealed that human movement generally decreased in all four islands, with Taiwan being the first to show a declining trend among the islands, but also the least pronounced one. New Zealand, Australia, and Iceland underwent a marked decline in human movement during March 2020, with a slight recovery starting around mid-April. Weekly travel oscillations can be observed in the original mobility data, but are averaged out by the sigmoidal model.

**Fig. 2:**
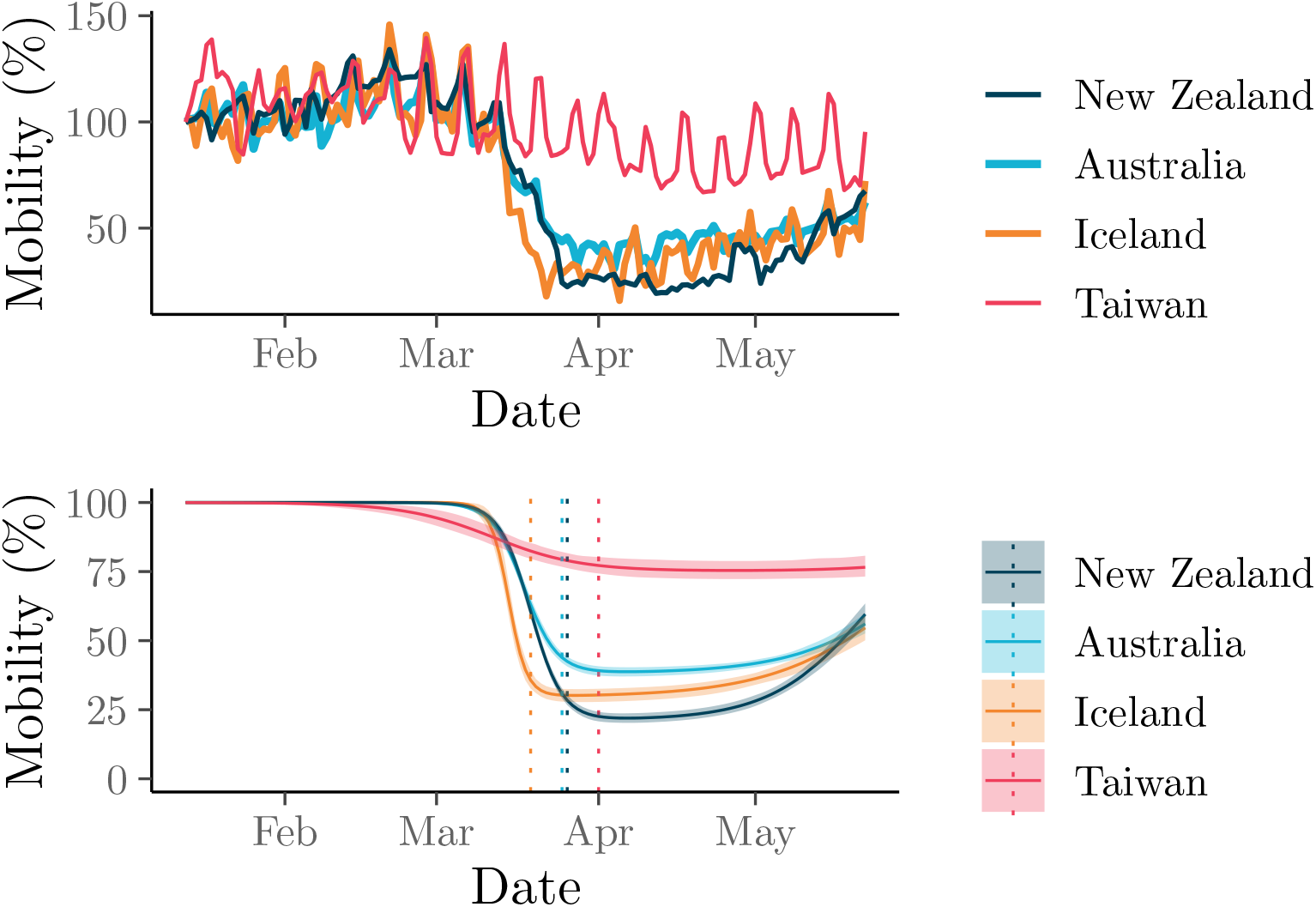
Human walking movement according to cell phone mobility data (Apple, 2020) relative to day 1 (100%). Top: movement data from Apple. Bottom: fitted sigmoid model (solid lines show the median and shaded curves show the 2.5–97.5% quantile interval); vertical dotted lines correspond to the estimated mean date of mobility change, which defines the *t*_2_ epoch date for each deme.

The dates of mobility reduction provided by this mobility model informed the start of the final time interval *t*_2_ in our three-epoch models of *R*_*e*_ and *b*.

### SARS-CoV-2 phylodynamics

We tested four genome subsampling regimes under four phylodynamic models. This sensitivity analysis suggests that inference under the MTBD and SC models is fairly robust to subsampling protocols, as opposed to DPG and DPG2, which yielded unrealistically small estimates of infection duration and of viral introduction counts (see Iceland; Fig. 5), particularly from the “active” method. Such model behavior is unsurprising and has been reported elsewhere (De Maio et al., 2015), and was only slightly alleviated when “large” subsamples were used. Analyses under MTBD and SC were prohibitively time-demanding using the larger datasets. For consistency, we only present results from the “small-time” dataset in the main article, but further results can be found in supplementary information.

We estimated *R*_*e*_ independently for each epoch using the MTBD model (Fig. 3). This analysis estimates *R*_*e*_ as 1.02–1.41 during the early infection of each respective country (following the first case) and later at less than 1 (following human mobility reduction). These estimates were not sensitive to sampling strategies. Each island *IS* saw multiple introductions of SARS-CoV-2 from the rest of the world (*RW;* Fig. 4 and 5). Under MTBD, we estimated the mean number of sample sequence introductions to be 63, 87, 49, and 65 for New Zealand, Australia, Iceland, and Taiwan, respectively. Conversely, we estimated just one virus export event from each country to the rest of the world, across the time period and sample considered. Similar results were observed using the SC model, with neither SC nor MTBD producing drastically different estimates between subsampling strategies.

**Fig. 3:**
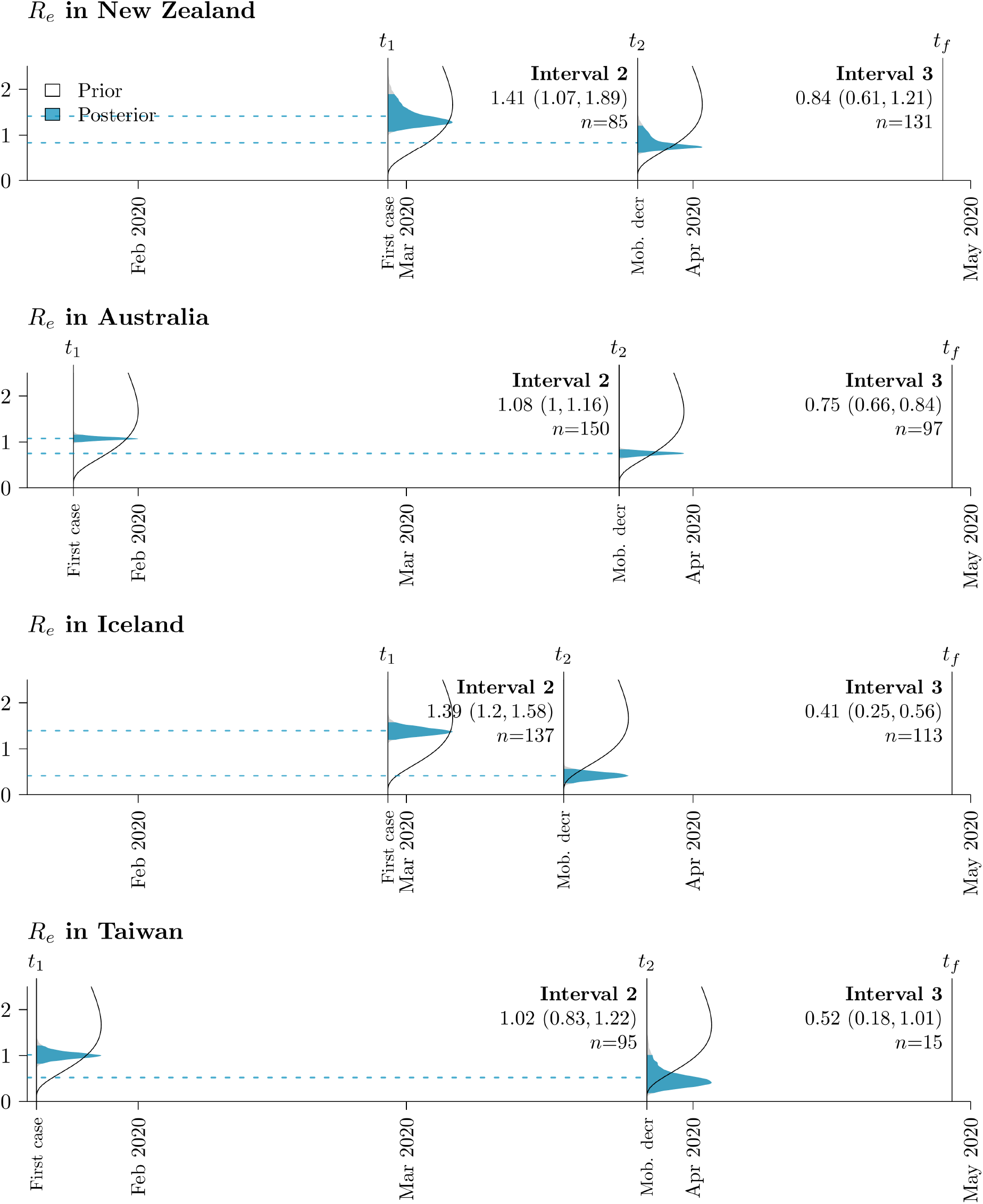
*R*_*e*_ prior (white) and posterior (blue) probability distributions across islands and time intervals (*n* samples in each interval). Posterior means are indicated by dashed lines. Mean estimates (and 95% highest posterior density intervals) are indicated under each time interval: *t*_1_ is the first case in the target deme, *t*_2_ the start of mobility reduction, and *t*_*f*_ the final sample in the dataset.

**Fig. 4:**
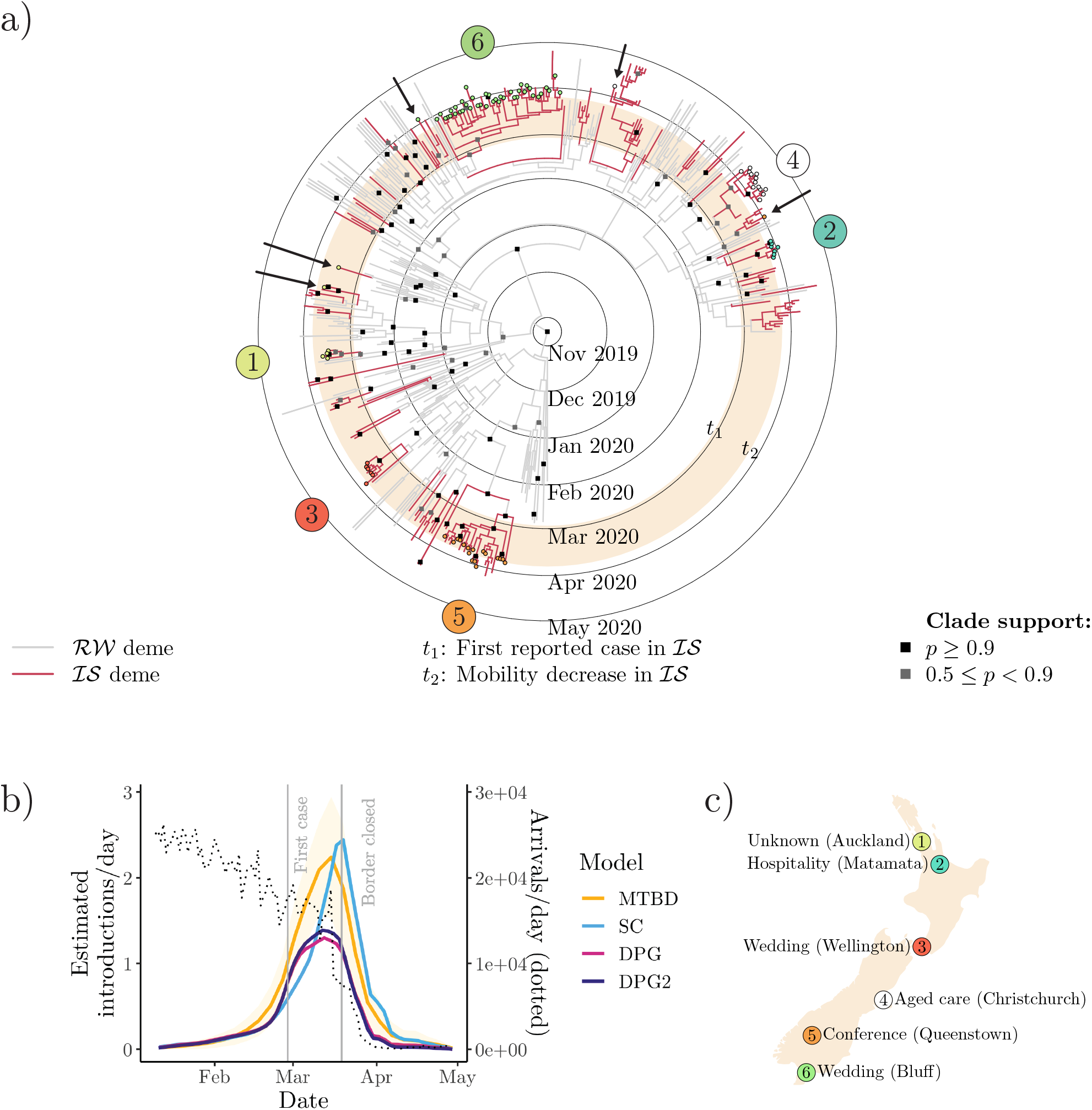
(a) New Zealand COVID-19 maximum-clade-credibility tree (from “small-time” alignment). Samples are labelled according to outbreaks identified by New Zealand Ministry of Health (NZMH). Arrows indicate cases assigned to a cluster by NZMH which are not monophyletic with the rest of the cluster. (b) Number of SARS-CoV-2 introductions in New Zealand over time. (c) New Zealand cluster locations (with > 5 cases).

**Fig. 5:**
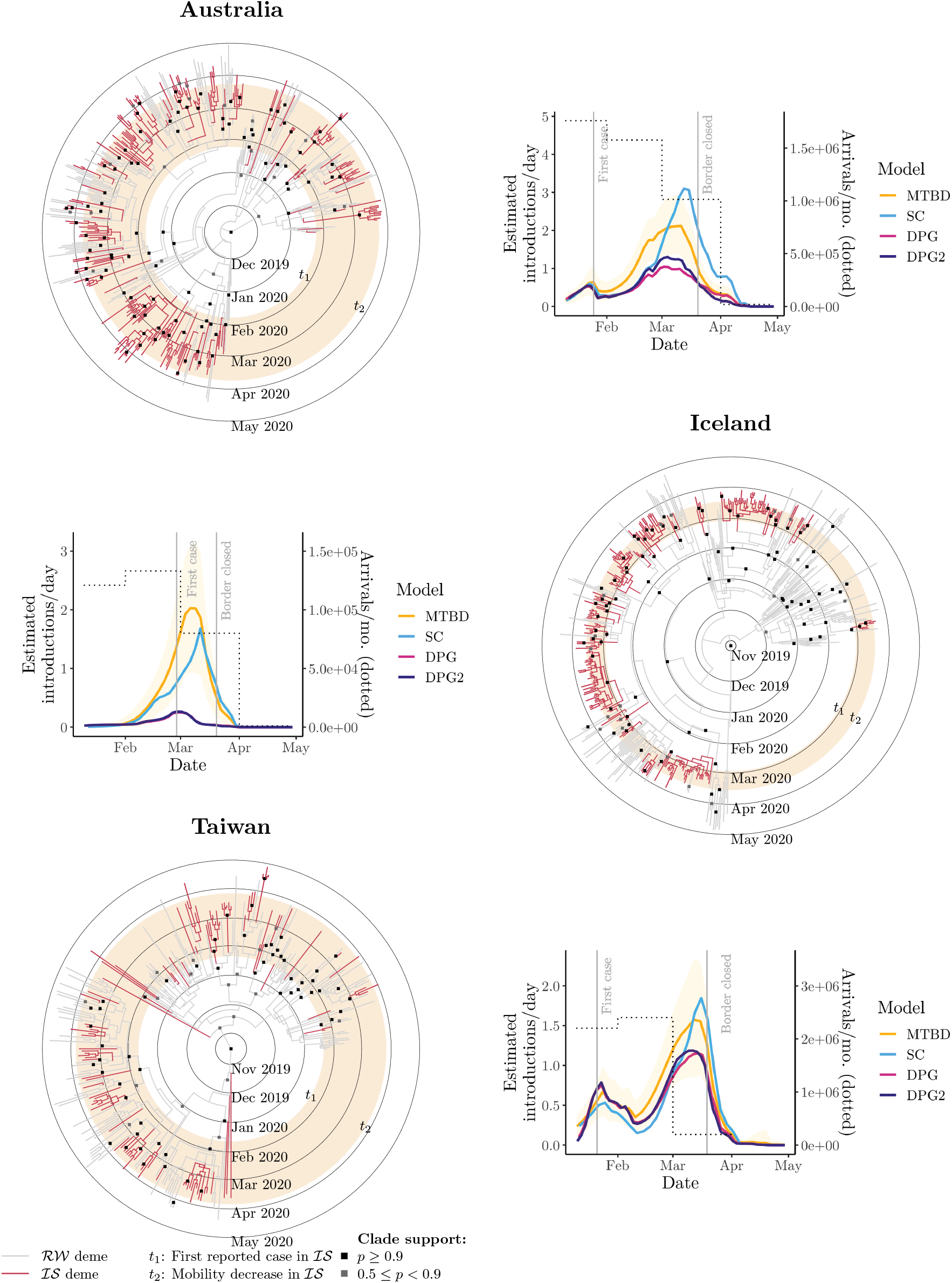
Maximum-clade-credibility COVID-19 tree (from “small-time” alignment) for Australia (top), Iceland (middle), and Taiwan (bottom). Plots show the estimated number of SARS-CoV-2 introductions over time. 18

### Comparison with contact tracing in New Zealand

The New Zealand Ministry of Health (NZMH) defines a significant COVID-19 cluster to comprise ten or more cases connected through transmission, and who are not all part of the same household (New Zealand Ministry of Health, 2020). Five of these clusters are represented by five or more of our samples (Fig. 4a and c), with a sixth non-significant cluster (based in the city of Auckland).

A clade is defined as a set of samples (within a phylogenetic tree) that share an exclusive common ancestor. Here we define a phylogenetic cluster as a clade whose samples share a label and whose origin can be explained by invoking a single introduction into a focal deme. Under this definition (i) case exports are allowed, (ii) isolated labelled tips outside of the clade are considered type-1 contact tracing mismatches, and (iii) unlabelled tips that are both from the focal deme and placed within the clade are considered type-2 mismatches.

Using this definition, all clusters identified by NZMH were recovered (among those represented by our samples). However, there were five type-1 mismatches: two from an Auckland-based cluster, one from Christchurch, one from Queenstown, and one from Bluff (shown by arrows in Fig. 4). We estimated 18 type-2 mismatches.

## Discussion

In this paper, we explored the introduction and transmission history of COVID-19 in New Zealand, Australia, Iceland, and Taiwan – geographically isolated countries that so far have been successful in preventing this disease from wreaking havoc as extensively it did in other parts of the world. We also delved deeper into the outcome of contact tracing within New Zealand specifically. This study is novel in its integration of genomic, epidemiological and mobile phone mobility data through explicit statistical models. The methods employed are robust and transparent, in that they (i) explicitly justify and incorporate prior knowledge about epidemiological parameters (see Supplementary information), (ii) report uncertainty in parameter estimation, and (iii) explore the effect that different data subsampling protocols have on parameter estimation.

### Phylodynamic modelling complements contact tracing

Identifying cases and the sources of their infections is an essential exercise in slowing infectious diseases such as COVID-19 (Hellewell et al., 2020), as recognised by the Director-General of the WHO in his pronouncement: *“You cannot fight a fire blindfolded. And we cannot stop this pandemic if we don’t know who is infected”*. Modern health surveillance strategies include medical databases containing travel history (Wang et al., 2020), the development of mobile phone applications (Ferretti et al., 2020), and the identification of outbreak subgroups known as “clusters”, whose cases share the same origin (New Zealand Ministry of Health, 2020). At the core of health surveillance practice lies contact tracing, the goal of which is to pinpoint the origin of infections by reconstructing the travel history of confirmed cases, and identifying all individuals with whom cases have come in contact.

Nonetheless, contact tracing has its limitations. Establishing the link between cases is restricted, for example, by the ability of cases to recall their recent contacts and travel history, which can lead to type-2 mismatches between epidemiological and genetic clusters. Furthermore, a case may have attended an event causally linked to a cluster but acquired their infection elsewhere and did not infect anyone at the event (type-1 mismatches). The implications of these types of error are manifold. If cluster sizes are underestimated, then so too is the rate of disease spread. If the extent of import-related cases is overestimated, then the impact of international air travel and the extent of community transmission cannot be fully accounted for.

After considering 217 genomes (representing 19% of the cases in New Zealand at the time), our results suggest there are an additional 18 unclassified cases in New Zealand that could have been linked to known clusters but were not (type-2 mismatch), and 5 cases that were linked to a cluster where they did not acquire or transmit the infection (type-1). However, by and large, our phylodynamic analysis is in agreement with conclusions reached by NZMH.

Overall, we have shown that contact tracing has been accurate among the cases considered, and succeeded to a large degree in identifying individuals belonging to the same infection cluster. We have demonstrated how the rapid real-time availability and assessment of viral genomic data can complement and augment health surveillance strategies, and thus improve the accuracy of contact tracing methods. As the pandemic has progressed, real-time whole genome sequence data has been integrated into the health response of the four countries discussed here (Geoghegan et al., 2020; Seemann et al., 2020; Gudbjartsson et al., 2020b; Taiwan Centers for Disease Control, 2020b). Genome sequencing is increasingly affordable and shared global surveillance resources such as NextStrain and GISAID are becoming more complete and useful. We advocate widely incorporating modern methods of genetic epidemiology into standard contact tracing and epidemic surveillance in order to ensure COVID-19 and future epidemics leave as promptly as they come.

### Restricting human movement constrains COVID-19 spread

Our results show that SARS-CoV-2 was introduced into each island on many independent occasions, corroborating the available epidemiological data from health officials (**Table 1**). Viral imports rapidly declined in the days leading to their border closing in mid-late March 2020, as a result of decreased international travel (Fig. 4 and 5). We believe this in turn played a significant role in limiting the spread of COVID-19.

Mobile phone data revealed idiosyncratic human movement patterns (Fig. 2). During the early stages of the pandemic, human mobility rapidly declined in both New Zealand and Australia as a result of stay-at-home orders (Hale et al., 2020), as well as in Iceland despite no such official restraints. Taiwan, on the other hand, did not exhibit as much of a decline in human movement, nor did it enforce stay-at-home orders on the general population. A goal of this study was to characterise the extent to which this decline in human movement could impact the spread of COVID-19.

We estimated effective reproduction numbers, *R*_*e*_, as approximately 1–1.4 at the onset of the pandemic in all four islands (Fig. 3). These estimates are not as high as in other countries where the pandemic was more widespread and where *R*_*e*_ was typically estimated in the range of 2-6 (Liu et al., 2020; Binny et al., 2020a; Alimohamadi et al., 2020). Our low estimates reflect the fact that a large fraction of cases in these countries were travel-related imports, and that community transmission rates were low. *R*_*e*_ declined as human movement decreased in each of the countries studied, reaching values below 1.0 (Fig. 3). Estimates of *R*_*e*_ based only on epidemiological data are also consistently below 1 during these time periods (Binny et al., 2020b,a; Price et al., 2020; Karnakov et al., 2020).

The case of Taiwan illustrates that an extreme restriction of domestic human movement is not required for suppressing a pandemic like COVID-19. In the first three months, Taiwan saw fewer cases-per-capita than New Zealand by an order of magnitude, for instance (**Table 1**), despite the latter enforcing a strict nationwide lockdown while having lower population density. The success of Taiwan has been attributed to the Taiwan Centers for Disease Control (Taiwan CDC) who were well prepared for such an epidemic following their experience with the SARS-CoV-1 outbreak in 2003, the early implementation of mass masking in public spaces, and border control (Summers et al., 2020; Wang et al., 2020).

Nevertheless, a stringent response against COVID-19 greatly restricting mobility such as that adopted by New Zealand is likely to have contributed to the elimination of SARS-CoV-2 by May 9 (Cousins, 2020), thus being the first country with over 1,000 confirmed cases to return to zero active cases, allowing it to re-open its economy, albeit with a restricted border. Most importantly, human morbidity and mortality were limited. Conversely, while Australia also saw a decline in human mobility and initially enjoyed some measure of success against COVID-19, its more lenient approach, compared with either New Zealand’s or Taiwan’s, might underlie Australia’s longer path to elimination (Hale et al., 2020; Aljazeera, 2020b).

In conclusion, we have shown that the introduction of SARS-CoV-2 was significantly inhibited as a result of international travel reduction in March 2020; notably the closing of borders in the four islands examined. We have also shown that spread of SARS-CoV-2 within these communities, while initially lower than other parts of the world, saw a further reduction following the decline of human mobility within the countries. However, positive outcomes against this and similar pandemics will be dependent on a multitude of other factors including contact tracing, case isolation, and hygienic practices (MacIntyre, 2020). As such, none of these should be seen as a silver bullet.

### Ethics and permissions

Nasopharyngeal samples testing positive for SARS-CoV-2 were obtained from public health medical diagnostics laboratories located throughout New Zealand. All samples werede-identified before receipt by the researchers. Under contract for the Ministry of Health, ESR has the approval to conduct genomic sequencing for surveillance of notifiable diseases.

## Data sharing

All genomic sequences used in this study are available on GISAID. BEAST 2 input and output files will be made available with publication.

## Supporting information

GISAID Acknowledgements

## Data Availability

Data will be made available upon publication

## Authors and contributors

J. Douglas - Conceptualisation, Analysis, Investigation, Methodology, Visualisation, Writing.

F.K. Mendes - Conceptualisation, Analysis, Investigation, Methodology, Visualisation, Writing.

R. Bouckaert - Analysis, Investigation, Methodology, Editing.

D. Xie - Analysis, Investigation, Methodology, Visualisation.

C.L. Jiménez-Silva - Analysis, Investigation, Methodology, Writing.

C. Swanepoel - Analysis, Investigation, Methodology, Writing.

J. de Ligt - Analysis, Funding Acquisition, Investigation, Methodology, Editing.

X. Ren - Analysis, Investigation, Methodology, Editing.

M. Storey - Analysis, Investigation, Methodology.

J. Hadfield - Analysis, Investigation, Editing.

C. Simpson - Investigation, Funding Acquisition, Editing.

J. Geoghegan - Analysis, Funding Acquisition, Investigation, Editing.

A.J. Drummond - Conceptualisation, Funding Acquisition, Investigation, Methodology, Editing.

D.Welch - Conceptualisation, Funding Acquisition, Investigation, Methodology, Visualisation, Writing.

The data in this study was verified by J. Douglas, F. Mendes, R. Bouckaert, D. Xie, C.L. Jiménez-Silva, C. Swanepoel, J. de Ligt, X. Ren, M. Storey, J. Hadfield, J. Geoghegan, A.J. Drummond, and D. Welch.

## Acknowledgments

We thank T. Vaughan and J. Sciré for discussions about the MTBD model, and all of the authors who have shared their genomic data on GISAID. This research was supported by a contract from the Health Research Council of New Zealand (20/1018), a COVID-19 Innovation Acceleration Fund (grant CIAF-0470) from the Ministry of Business, Innovation & Employment’s ResearchInfrastructure (MBIE), and Marsden grants 16-UOA-277 and 18-UOA-096 from the Royal Society of New Zealand. Genome sequencing was partially funded by New Zealand Ministry of Health. Finally, we wish to acknowledge the use of New Zealand eScience Infrastructure (NeSI) high performance computing facilities, funded jointly by NeSI’s collaborator institutions and through the MBIE’s Research Infrastructure program.

## SUPPORTING INFORMATION

## 1 Viral genomic sequencing

RNA from 217 confirmed COVID-19 cases in New Zealand were obtained from diagnostic laboratories around the country. Viral RNA is reverse transcribed with SSIV with random hexamers, and then amplified using multiple overlapping PCR reactions spanning the viral genome, by employing Q5 HotStart High-Fidelity DNA Polymerase. Two primer schemes were used in this study: 1) ARTIC network protocol (V1 and V3) and 2) the New South Wales (NSW) primer set described in Eden et al. (2020). Samples processed with the ARTIC protocol were sequenced on R9.4.1 MinION flow cells using the Oxford NanoPore ligation sequencing protocol (Loman et al., 2020). Sample processed with the NSW primer set were sequenced on Illumina NextSeq and MiSeq flowcells in paired-end 300 cycle format using the Nextera-XT library protocol. Table S1 describes the number of genomes processed using each protocol.

**Table S1:**
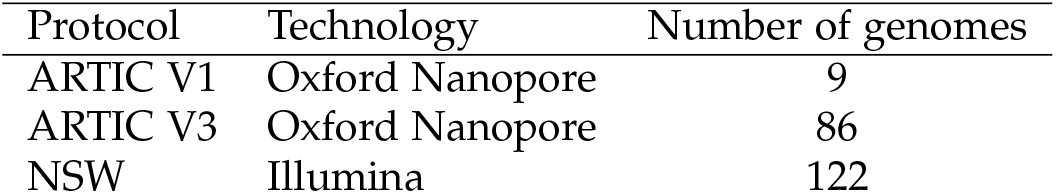
Sequencing protocols used to sequence the 217 New Zealand SARS-CoV-2 genomes.

| Protocol | Technology | Number of genomes |
| --- | --- | --- |
| ARTIC V1 | Oxford Nanopore | 9 |
| ARTIC V3 | Oxford Nanopore | 86 |
| NSW | Illumina | 122 |

For both methods, alignment to the reference genome MN908947.3 was followed by consensus calling for the major alleles on variation sites. Regions are masked with *N*’s in the final genome when amplicon failed to reach sufficient depth. Genomes with fewer than 3000 *N*’s in their consensus genome were used in the analysis presented here. The nanopore reads generated with Nanopore sequencing of ARTIC primer sets (V1 and V3) were mapped and assembled using the ARTIC bioinformatics pipeline (v1.1.0; Loman et al. 2020) with the “–medaka” flag enabled in the minion step. For the NSW primer set, raw reads were quality and adapter trimmed using `trimmomatic` (v0.36; Bolger et al. 2014) with the following settings: removing first 15 and last 20 bases of each read, minimum trailing quality of 30, 4nt-moving average quality of 25, and length >= 100bps.

Trimmed paired reads were mapped to reference using bwa. Primer sequences were masked using iVar (v1.2; Grubaugh et al. 2019). Duplicated reads were marked using Picard (v2.10.10; http://broadinstitute.github.io/picard/) and not used for SNP calling or depth calculation. Single nucleotide polymorphisms (SNPs) were called using bcftools mpileup (v1.9; http://samtools.github.io/bcftools/). SNPs within genomic regions amplified by each primer sets and primer regions were called separately. SNPs were quality trimmed using vcffilter (vcflib v1.0.0; Garrison 2014) requiring 20x depth and overall quality of 30. Positions that are less than 20x were masked as *N* in the final consensus genome. In addition, positions with an alternative allele frequency between 20 and 79% were also masked as *N*.

## 2 Sequence preprocessing

### 2.1 Filtering

Viral genomic sequences were filtered by observing the following constraints:

1. All viral sequences must have a human host;
2. All sequences must have date resolution down to the day of the month;
3. All sequences be more than 25 kb in length;
4. The proportion of ambiguous sites in a sequence must be less than 0.005 (appoximately 150 sites) (due to a shortage in sequences, samples from New Zealand and Taiwan are exempt from this constraint);
5. Sequences on the “NextStrain CoV exclude list” (Hadfield et al., 2018); URL: https://github.com/nextstrain/ncov/blob/master/defaults/exclude.txt) are excluded from the analysis. This list contains sequences which were identified as problematic either through manual inspection, or due to the sample falling outside 4 interquartile ranges from the mean root-to-tip distance (calculated using TreeTime; Sagulenko et al. 2018).

### 2.2 Subsampling

Because analysing all available genomic data using our Bayesian model is prohibitively slow, subsampling the full dataset is necessary. We assess four subsampling schemes producing one alignment each, differing with respect to (i) dataset size *n* (either “small” or “large”), and (ii) the method used to select sequences (either “time” or “active”). In (i), the rest-of-the-world deme (*RW*) contributes 200 or 500 sequences to the “small” and “large” datasets, respectively. Out of these sequences, 40 (for “small”) or 80 (for “large”) always come from China for the phylogenetic signal they carry with respect to the root of the infection tree. The island deme (*IS*) then contributes the remaining sequences to *n*, up to a maximum of 250. Specifically, Australia contributes 250 sequences, Iceland 250, New Zealand 217, and Taiwan 76 (see more details on our deme definitions below). In (ii), the method consists of either (a) randomly choosing one sequence from each of *n* dates sampled with replacement (i.e., picking uniformly through time; the “time” method), or (b) sampling (with replacement) *n* country-date pairs proportionally to the number of active cases reported in that country on that date (according to https://www.worldometers.info/coronavirus/), with one sequence chosen randomly from each pair (the “active” method). 41

### 2.3 Alignment

After filtering and subsampling sequences, alignments are generated using MAFFT under its default settings (Katoh et al., 2009), yielding a total of 16 alignments (four alignments per geographical model).

**Table S2:** Taxa count *N*, alignment length *L*, and A, C, G, T, and ambiguous (?) content are reported for each alignment used in this study.

| Island | Alignment | $N$ | $L$ (kb) | A (%) | C (%) | G (%) | T (%) | ? (%) |
| --- | --- | --- | --- | --- | --- | --- | --- | --- |
| Australia | Small-time | 450 | 30 | 29.7 | 18.3 | 19.5 | 32 | 0.1 |
| Australia | Small-active | 450 | 30 | 29.7 | 18.3 | 19.5 | 32 | 0.1 |
| Australia | Large-time | 750 | 30 | 29.7 | 18.3 | 19.5 | 31.9 | 0.1 |
| Australia | Large-active | 750 | 30 | 29.7 | 18.2 | 19.5 | 31.9 | 0.1 |
| Iceland | Small-time | 450 | 30 | 29.7 | 18.3 | 19.5 | 32 | 0.2 |
| Iceland | Small-active | 450 | 30 | 29.7 | 18.3 | 19.5 | 32 | 0.2 |
| Iceland | Large-time | 750 | 30 | 29.7 | 18.2 | 19.5 | 31.9 | 0.2 |
| Iceland | Large-active | 750 | 30 | 29.7 | 18.3 | 19.5 | 32 | 0.1 |
| New Zealand | Small-time | 417 | 30 | 29.5 | 18.1 | 19.4 | 31.7 | 0.9 |
| New Zealand | Small-active | 417 | 30 | 29.5 | 18.2 | 19.4 | 31.8 | 0.8 |
| New Zealand | Large-time | 717 | 30 | 29.5 | 18.1 | 19.4 | 31.7 | 0.5 |
| New Zealand | Large-active | 717 | 30 | 29.6 | 18.2 | 19.4 | 31.8 | 0.5 |
| Taiwan | Small-time | 310 | 30 | 29.5 | 18.1 | 19.4 | 31.7 | 0.1 |
| Taiwan | Small-active | 310 | 30 | 29.6 | 18.2 | 19.4 | 31.8 | 0.1 |
| Taiwan | Large-time | 610 | 30 | 29.5 | 18.1 | 19.4 | 31.7 | 0.1 |
| Taiwan | Large-active | 610 | 30 | 29.3 | 18 | 19.2 | 31.5 | 0.1 |

## 3 Demes and epochs

We consider four geographical models – each comprised of a target island deme ℐ𝒮 (Australia, Iceland, New Zealand, or Taiwan), and a mutually exclusive “rest-of-the-world” deme ℛ𝒲 The ℛ𝒲 grouping of viral lineages is not biologically meaningful – it is a computational and modelling convenience – as its samples are further structured among themselves and not necessarily more similar to each other than to sequences from island demes. For this reason, the parameters associated with the RW deme are not interpreted and are treated as nuisance parameters.

Our MTBD analyses employ a general model implementation that allows for piecewise rate changes over the course of the infection (Kühnert et al., 2016), i.e., rates are held constant within time intervals, but are allowed to vary between them. An infection process is characterised by the time of the origin at *t*_0_, and the end of its last (*f* -th) interval, *t* _*f*_ (which is also the last sampling time). Each of the *f* intervals is then defined by a boundary time *t*_*i*_ *∈ t* = (*t*_1_, …, *t* _*f* −1_), where *t*_0_ < *t*_1_ < … < *t* _*f* −1_ < *t* _*f*_. Given a parameter *p ∈ {R*_*e*_, *b, s, m}, p*_*i*_ is the rate inside the *i*-th interval [*t*_*i*−1_, *t*_*i*_). Time intervals *t* are both parameter-specific and deme-specific, and are treated as data (i.e., they are not sampled; Table S3).

**Table S3:** Time intervals (epochs) used by the MTBD model, specified as [*t*_*i*_, *t*_*i*+1_). *t*_0_ and *t* _*f*_ correspond to the origin and the date the last sample was collected. All other times are deme-specific, and in the case of *s*, sample specific. The “First reported case” dates are in Table 1 of the main article. For simplicity, the “Border close” date is held at March 20 for all demes, despite a one day difference existing in practice. “Mobility decline” dates are shown in Table S5. The sampling proportion *s* is held constant at 0 from the start of the infection until the first tip within the deme (an alignment-specific date), and then estimated in the second epoch. As there is no migration signal before the first reported case within each island, the migration rate *m*_ℐ𝒮,ℛ𝒲_ is fixed to a negligible quantity (10^−6^) due to its non-identifiability. Nuisance parameters (ℛ𝒲deme) are modelled with a single time interval over which they are sampled.

| Parameter(s) | Deme | $i$ | Time interval | |
| --- | --- | --- | --- | --- |
| | | | Start ($t_i$) | End ($t_{i+1}$) |
| $R_e$ and $b$ | $\mathcal{IS}$ | 0 | $t_0$ | First reported case |
| $R_e$ and $b$ | $\mathcal{IS}$ | 1 | First reported case | Mobility decline |
| $R_e$ and $b$ | $\mathcal{IS}$ | 2 | Mobility decline | $t_f$ |
| $R_e$ and $b$ | $\mathcal{RW}$ | 0 | $t_0$ | $t_f$ |
| $s = 0$ | $\mathcal{IS}$ | 0 | $t_0$ | First sample |
| $s$ | $\mathcal{IS}$ | 1 | First sample | $t_f$ |
| $s = 0$ | $\mathcal{RW}$ | 0 | $t_0$ | First sample |
| $s$ | $\mathcal{RW}$ | 1 | First sample | $t_f$ |
| $m_{\mathcal{IS}, \mathcal{RW}} = 10^{-6}$ | — | 0 | $t_0$ | First reported case |
| $m_{\mathcal{RW}, \mathcal{IS}}$ | — | 0 | $t_0$ | First reported case |
| $m_{\mathcal{IS}, \mathcal{RW}}$ | — | 1 | First reported case | Border close |
| $m_{\mathcal{RW}, \mathcal{IS}}$ | — | 1 | First reported case | Border close |
| $m_{\mathcal{IS}, \mathcal{RW}}$ | — | 2 | Border close | $t_f$ |
| $m_{\mathcal{RW}, \mathcal{IS}}$ | — | 2 | Border close | $t_f$ |

### 3.1 A model of human movement decrease for mobile phone data

Defining time intervals characterised by different levels of human movement is challenging, as the four “island” considered here differ markedly in their response to COVID-19. No single date (such as the day of lockdown, for example) could be used across countries: while the governments of New Zealand and Australia locked human movement down to different extents for several weeks, those of Iceland and Taiwan, on the other hand, found it sufficient to restrict the size of gatherings and encourage social distancing (Hale et al., 2020).

In order to define country-specific time intervals in a statistically principled way, we first parameterise human movement decrease with a sigmoid model describing how mobility (Apple, 2020; Fig. S1) changes as a function of time *t*:

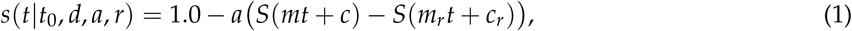

the parameters of which are described in table S4. Here, *S* corresponds to the sigmoid (logistic) function 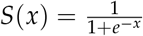, and *m, c, m*_*r*_ and *c*_*r*_ are deterministically defined in terms of other parameters such that:

1. 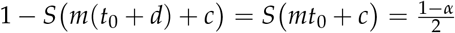, i.e., *α* of the decrease occurs between *t*_0_ and *t*_0_ + *d*,
2. 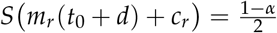, i.e., only 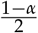 of the recovery has occurred at the end of the decrease, and
3. *S m*_*r*_(*t*_0_ + *d* + *d*_*r*_) + *c*_*r*_ = *r*, i.e., *r* of the recovery has occurred *d*_*r*_ after the end of the decrease.

We apply Equation 1 to our set of four demes ℐ = *{*1, 2, 3, 4*}* by allowing each country *i* ∈ ℐ to have its own parameters controlling the timing of the decrease, *t*_0*i*_ and *d*_*i*_. Additionally, we can consider the three modes of transportation *j* individually (i.e., *j* ∈ 𝒥 = *{*1, 2, 3*}*), and allow each (country *i*–mode of transportation *j*) pair to be characterised by its own decrease amplitude, *a*_*ij*_, recovery amplitude *r*_*ij*_, and mobility standard deviation parameter *v*_*ij*_ (Table S4). Finally, we let each combination of country, mode of transportion, and weekday *k* have its own baseline mobility mean of *b*_*ijk*_. Note that these considerations make all parameters in equation 1 multidimensional (Table S4).

We model the human movement underlying mobility data **M** = (*m*_*ijk*_) (*i*-th country, *j*-th mode of transportation, *k*-th weekday) by assuming *M*_*ijk*_ is sampled from a Normal distribution, letting:

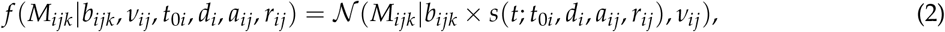

where 𝒩 (·|*µ, σ*) is the probability density function of a normal distribution with mean and standard deviation *σ*. Note that the baseline mean of this distribution, *b*_*ijk*_, is scaled by the sigmoid function *s*(·) describing the mobility decrease effect in equation 1.

**Fig. S1:**
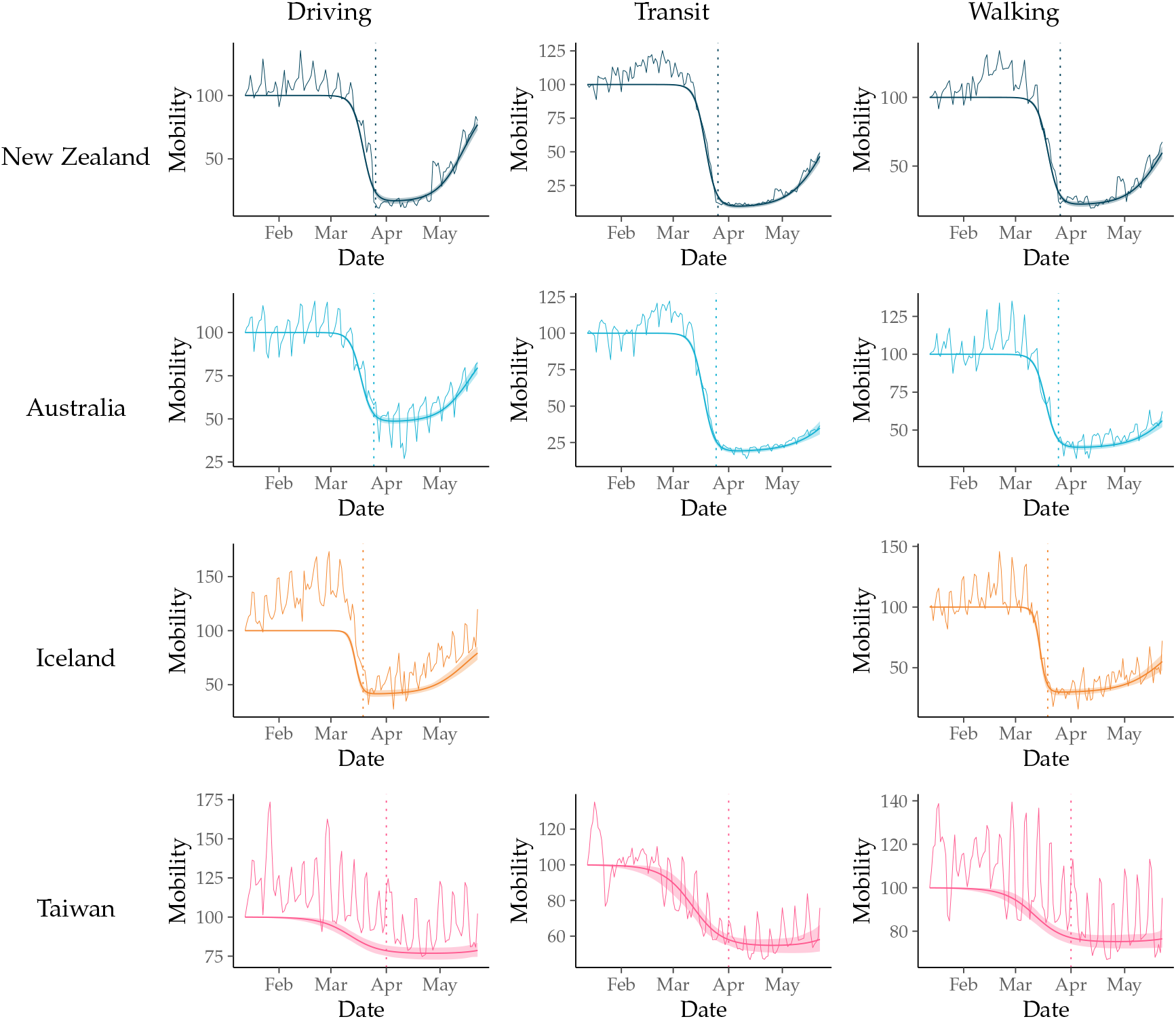
Cell phone mobility data from New Zealand, Australia, Iceland and Taiwan, for three kinds of transportation: driving, transit and walking. The “wiggly” lines correspond to the raw data. Fit sigmoid models are shown in all graphs as a smooth shaded curve (the 2.5–97.5% quantile interval) and a solid line inside of it (the sigmoid function).

Our full model is hierarchical and allows the mobility signal of the different nations to jointly and mutually inform all model parameters. This is done by using hierarchical priors, and estimating shared hyperparameters *θ* = *{µ*_*t*0_, *σ*_*t*0_, *µ*_*d*_, *σ*_*d*_, *α*_*aj*_, *β*_*aj*_, *α*_*rj*_, *β*_*rj*_, *b*_*ijk*_, *v*_*ij*_*}*. The unnormalised posterior density of our full model is given by:

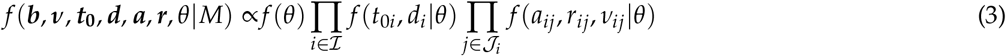

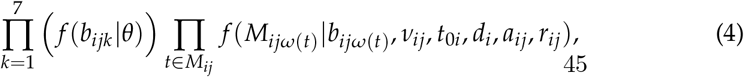

where *ω*(*t*) is a deterministic function that returns the weekday of a time point *t, f* (*θ*) is the hyperprior density on hyperparameters *θ*, and *f* (*t*_0*i*_, *d*_*i*_|*θ*), *f* (*a*_*ij*_, *r*_*ij*_, *v*_*ij*_|*θ*) and *f* (*b*_*ijk*_|*θ*) are the prior densities on country-, transportation mode-, and weekday-level parameters, respectively (Table S4). We chose our priors following recommendation in Gelman et al. (2006). Priors are listed table S4.

We determined the boundary between time intervals characterised by “high” and “low” human movement using New Zealand as a reference. As a post processing step after MCMC, we computed and logged the proportion of mobility that had gone down in New Zealand by 26 March 2020 (according to the fitted sigmoid model), and then deterministically recorded the dates by which the other islands had undergone the same proportional reduction. Let *S* (*t*|*t*_0_, *d*) = *S*(*mt* + *c*), the sigmoid function describing the decrease in mobility, where *m* and *c* are defined as in Equation 1. Let *S*^−1^(*p*|*t*_0_, *d*) be the inverse of this function. Then, for a given set of parameter values, the boundary date for country *i* is:

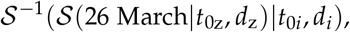

where the subscript *z* refers to the index of New Zealand in the set *I*. We used the posterior mean of this date as the boundary between time intervals.

Our model is implemented in PyMC3 (Salvatier et al., 2016), and we employed the No-U-Turn Sampler (Hoffman and Gelman, 2014) to sample the posterior distribution. We ran three independent chains and sampled each 10000 times (after 1000 tuning samples). The three chains were visually checked for convergence and combined, yielding effective samples sizes larger than 200.

**Table S4:** Parameters of sigmoid model for human movement. These parameters share names with random variables from other models, but represent distinct quantities only used in the sigmoid model. N_*o*_ indicates a bounded Normal distribution conditioned on non-negativity, and HC indicates a Half-Cauchy distribution with location 0 and conditioned on non-negativity.

| Parameter | Description | Prior |
| --- | --- | --- |
| **Mobility model parameters** |  |  |
| $t_0$ | $t_0 = (t_{0i}), i \in \mathcal{I}$, the start of the mobility decrease effect in country $i$ | $N(\mu_{t_0}, \sigma_{t_0})$ |
| $d$ | $d = (d_i), i \in \mathcal{I}$, the duration of the mobility decrease effect in country $i$ | |
| $a$ | $a = (a_{ij}), i \in \mathcal{I}, j \in \mathcal{J}$, the proportional amplitude of the mobility decrease for country $i$ and transportation mode $j$ | $\text{Beta}(\alpha_{aj}, \beta_{aj})$ |
| $r$ | $r = (r_{ij}), i \in \mathcal{I}, j \in \mathcal{J}$, the proportion of mobility decrease that recovers by $(t_0 + d + d_r)$ in country $i$, for transportation mode $j$ | $\text{Beta}(\alpha_{rj}, \beta_{rj})$ |
| $b$ | $b = (b_{ijk}), i \in \mathcal{I}, j \in \mathcal{J}, k \in \mathcal{K}$, the baseline mobility for country $i$, transportation mode $j$ and weekday $k$ | $N_o(\mu_{bjk}, \sigma_{bjk})$ |
| $v$ | $v = (v_{ij}), i \in \mathcal{I}, j \in \mathcal{J}$, the mobility standard deviation for country $i$ and transportation mode $j$ | $\text{HC}(0.1)$ |
| $\alpha$ | Proportion of decrease effect which occurs between $t_0$ and $t_0 + d$ | Fixed to 0.99 |
| $d_r$ | Duration over which recovery is measured | Fixed to 60 |
| **Hyperparameters** |  |  |
| $\mu_{t_0}$ | $t_0$ prior mean | $N(\text{Mar 25, 14 days})$ |
| $\sigma_{t_0}$ | $t_0$ prior standard deviation | $\text{HC}(2.0)$ |
| $\mu_d$ | $d$ prior mean | $N_o(14 \text{ days}, 7 \text{ days})$ |
| $\sigma_d$ | $d$ prior standard deviation | $\text{HC}(2.0)$ |
| $\alpha_a$ | $\alpha_a = (\alpha_{aj}), j \in \mathcal{J}$, $a$ prior shape | $N_o(2.0, 2.0)$ |
| $\beta_a$ | $\beta_a = (\beta_{aj}), j \in \mathcal{J}$, $a$ prior rate | $N_o(2.0, 2.0)$ |
| $\alpha_r$ | $\alpha_r = (\alpha_{rj}), j \in \mathcal{J}$, $r$ prior shape | $N_o(1.5, 2.0)$ |
| $\beta_r$ | $\beta_r = (\beta_{rj}), j \in \mathcal{J}$, $r$ prior rate | $N_o(2.0, 2.0)$ |
| $\mu_b$ | $\mu_b = (\mu_{bjk}), j \in \mathcal{J}, k \in \mathcal{K}$, $b$ prior mean | $N_o(1.0, 0.2)$ |
| $\sigma_b$ | $\sigma_b = (\sigma_{bjk}), j \in \mathcal{J}, k \in \mathcal{K}$, $b$ prior standard deviation | $\text{HC}(1.0)$ |

**Table S5:** Date boundaries of mobility reduction, according to the model described in Section 3.1 which was fit to mobile phone data (Apple, 2020). All dates are within the year of 2020. The start dates were used as epoch boundaries for *R*_*e*_ and *b*.

| $\mathcal{IS}$ | Start | Mean | End |
| --- | --- | --- | --- |
| New Zealand | Mar 26 | Mar 26 | Mar 26 |
| Australia | Mar 24 | Mar 25 | Mar 26 |
| Iceland | Mar 18 | Mar 19 | Mar 20 |
| Taiwan | Mar 27 | Mar 31 | Apr 05 |

## 4 Model definition

### 4.1 Substitution and clock models

In all analyses, the phylogenetic likelihood was evaluated under the HKY substitution model with estimated nucleotide equilibrium frequencies *π*_*D*_. Sites were partitioned into non-coding sites and the three codon positions, with each partition having its own substitution model parameters. The end regions were masked because they are suspected to harbour many sequencing errors (as described by http://virological.org/t/issues-with-sars-cov-2-sequencing-data/473). Because SARS-CoV-2 genomes have undergone a small number of mutations (Lai et al., 2020; Li et al., 2020b; Rambaut, 2020), we assumed a strict molecular clock in all analyses.

#### 4.1.1 Partition schemes and substitution model selection

We compared substitution models and partition schemes based on their posterior distributions over *𝒯*, and picked the simplest model generating the least different posterior distribution. For example, substitution models more complex than HKY did not affect the posterior distribution of *𝒯* relative to HKY, while simpler models such as JC69 (Jukes Cantor 1969) yielded a posterior distribution that was substantially different. Therefore, we selected HKY with estimated frequencies. Model comparison was done with BModelTest (Bouckaert and Drummond, 2017). By the same token, we did not opt for single partition analyses – which led to very different tree posteriors compared to the chosen partition scheme (see main text) – and rejected adding further partitions (e.g., on gene boundaries) because they did not alter the tree posterior significantly.

### 4.2 Phylodynamic models

#### 4.2.1 Discrete phylogeography (DPG)

The DPG model (Lemey et al., 2009) employs a continuous-time Markov chain in similar fashion to substitution models used in molecular evolution studies, but one in which a single “character” is considered: the discrete location (the deme) a lineage occupies in space. Lineages are allowed to change demes over time, with demes being inherited by children lineages according to a phylogenetic tree. As opposed to nucleotide substitution models that emit four discrete states, however, the DPG model can emit *d* states, where *d* is the number of demes represented in the data.

Under the DPG model, state transitions correspond to migrations events (i.e., a lineage inoves from one deme to another) that happen as described by a 2 × 2 symmetric, infinitesimal rate matrix:

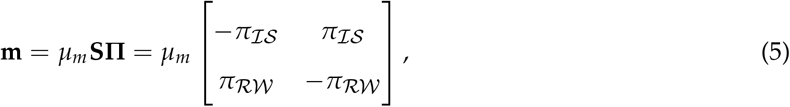

where *µ*_*m*_ is an overall rate scaler, **S** is a matrix of relative rates of changing demes, and **Π** = diag(*π*) (with *π* being the equilibrium deme frequencies; Table S6). Note that (i) **Π** is estimated as opposed to **S**, because the two are non-identifiable (i.e., a symmetric model), (ii) **Π** is normalised such that *µ*_*m*_ reflects the number of migration events per unit time (Lemey et al., 2009), and (iii) the frequencies **Π** at the root are fixed at **Π** = (*π*_*IS*_, *π*_*RW*_) = (0, 1) to incorporate knowledge of the infection originating outside of the four island demes of interest.

Finally, we couple our DPG model implementation to the Bayesian skyline model (Drummond et al., 2005) to allow for effective population size (*N*_*e*_) changes over ten time intervals. Following the notation from the main manuscript, under the DPG we have:

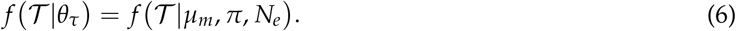

#### 4.2.2 Two epoch discrete phylogeography (DPG2)

The two epoch discrete phylogeography model (DPG2) generalises the DPG model by distinguishing two time intervals: one before and one after *t* =19 March 2020, the date in which both New Zealand and Taiwan closed its borders (Australia and Iceland closed theirs in the following day). These two time intervals are characterised by their own **m** rate matrix. Following the notation from the main manuscript, under the DPG2 model we have:

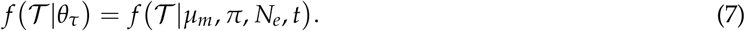

Note that *t* here is fixed and treated as data. In computing this density, only one **m** matrix is used for branches contained in their entirety within a time interval; branches that are intersected by *t*, on the other hand, have transition probabilities computed by combining two different **m** matrices (Bielejec et al., 2014).

#### 4.2.3 Structured coalescent (SC)

The fourth phylodynamic model we used was the structured coalescent model implemented in MASCOT (**M**arginal **A**pproximation of the **S**tructured **Co**alescen**T**; Müller et al. (2018)). Structured coalescent models allow one to estimate demographic parameters and genealogical relationships among sub-populations (demes), but estimation under exact implementations is costly because lineage ancestral states must be sampled with MCMC. The marginal approximation of the structured coalescent (MASCOT) model, on the other hand, circumvents this issue by integrating over all possible migration histories (Müller et al., 2018). Under this model, we estimate population sizes for each deme 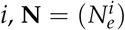, and an among-deme migration rate matrix **m**. Following the notation from the main text:

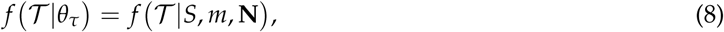

where 𝒮 is treated as data containing the deme information for each sample.

#### 4.2.4 Multi-type birth-death (MTBD)

The full probabilistic graphical model given the MTBD tree prior used in this study can be seen in Fig. S2.

**Fig. S2:**
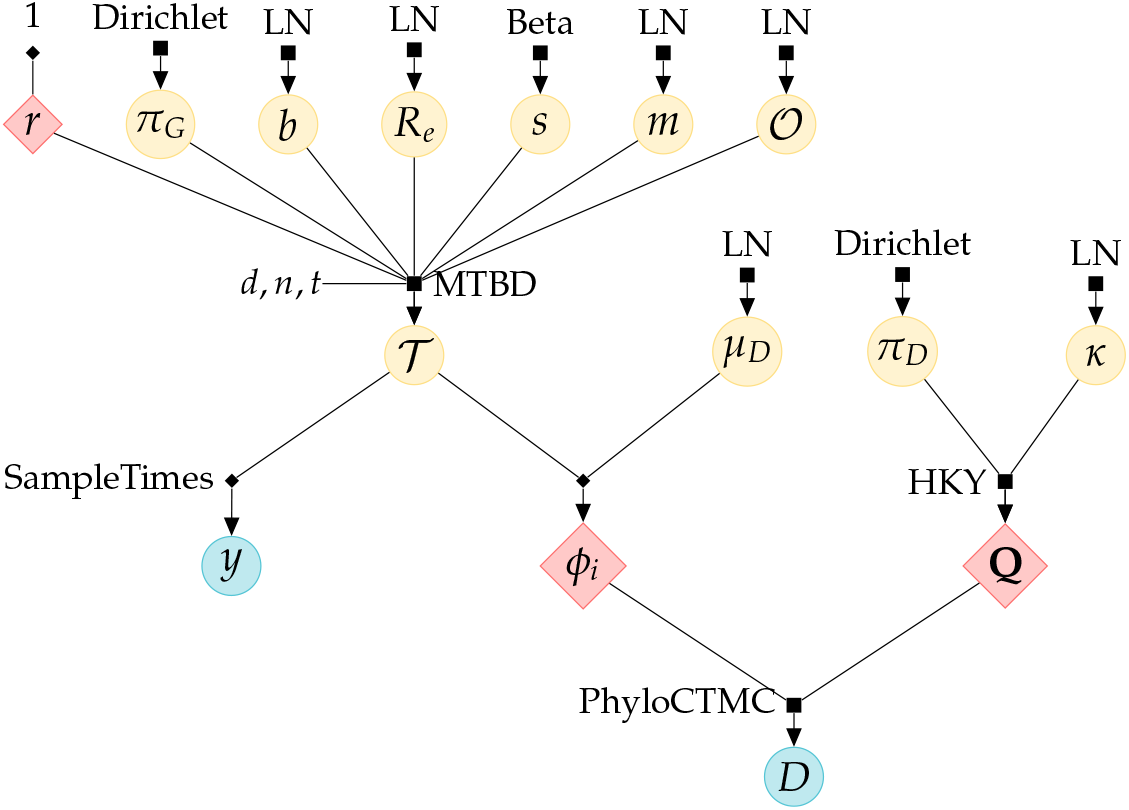
Full MTBD probabilistic graphical model used in our study. Yellow and blue circles correspond to parameters and observed data, respectively. Red diamonds are deterministic functions and their outcomes. Filled squares represent sampling distributions (e.g., “MTBD”, multitype birth-death; “HKY”, Hasegawa-Kishino-Yano substitution model).

### 4.3 Prior distributions

The prior distributions used in this article are summarised in Table S6. Priors for *O, s, m*, and *π*_*G*_ were programatically generated for each alignment and are detailed in the following subsections.

**Table S6:** Summary of prior distributions. Γ(*α, β*) denots a gamma distribution with shape parameter *α* and inverse scale parameter *β* “LN(*µ, σ*)” denotes a log-normal distribution with log-space mean *µ* and log-space standard deviation *σ* “Exp(*µ*)” denotes an exponential distribution with a mean of *µ*. “Dir(*α*_1_, …, *α*_*m*_)” is a Dirichlet distribution with shapes (*α*_1_, …, *α*_*m*_). *Origin time prior distribution has an offset (see Section 4.3.1). \*\**s* is parameterised as *s*^*t*^ (see Section 4.3.2).

| Parameter | Description | Prior distribution |
| --- | --- | --- |
| **DPG/DPG2 parameters** |  |  |
| $\mu_m$ | Migration rate scaler | $\Gamma(0.001, 1000)$ |
| $\pi$ | Equilibrium deme frequencies | Dir(1, 1) |
| $N_e(0)$ | Effective population size baseline | LN(0, 2) |
| $N_e(i), i > 0$ | Effective population size change | Exp($N(i - 1)$) |
| **SC parameters** |  |  |
| $N_e$ | Effective population size | LN(0.007, 1) |
| $m$ | Migration rates | |
| **MTBD parameters** |  |  |
| $R_e$ | Basic reproduction number | LN(1.0, 0.7) |
| $b$ | Rate of becoming non-infectious | LN(4.09, 0.2) |
| $\mathcal{O}$ | Origin time | LN(-1.98, 0.4)* |
| $r$ | Removal probability | 1 |
| $s$ | Sampling proportion | Beta(1.1, 8.0)** |
| $m$ | Migration rates | LN(0, 1) |
| $\pi_G$ | Equilibrium deme frequencies (see Section 4.3.3) | |
| **Substitution model parameters (shared)** |  |  |
| $\kappa$ | HKY transition-transversion ratio | LN(1, 1.25) |
| $\pi_D$ | HKY nucleotide frequencies | Dir(1, 1, 1, 1) |
| $\mu_D^1$ | Molecular clock rate (for DPG and DPG2) | LN(-7, 1.25) |
| $\mu_D^2$ | Molecular clock rate (for MTBD and SC) | LN(-7, 0.25) |

#### 4.3.1 Prior for origin time

As defined in the main text, origin time *O* represents the height of the sampled infection tree (*T*) in years, which goes from the most recent sample to patient zero, the first case of the disease. The time of the most recent sample can vary depending on the sampling scheme (Section 2), but with the exception of one alignment, all alignments had their most recent sample taken on 29 April 2020. Patient zero has been only tentatively placed on 17 November 2020 in Hubei, China, but at least 60 cases had been confirmed with certainty by 20 December in that country (see https://www.scmp.com/news/china/society/article/3074991/coronavirus-chinas-first-confirmed-covid-19-case-traced-back).

We assumed an offset log-normal prior distribution for 𝒪; specifically, (𝒪 − *δ*) *∼* LN(*µ* = −1.98, *σ* = 0.4), where *δ* is an offset defined by the time interval between the alignment-dependent first sample and 20 December 2020, a day we judged to provide positive evidence COVID-19 was spreading in China. By applying variable offsets, one can use the same log-normal parameterisation (i.e., same mean and standard deviation) for all alignments. Therefore, irrespective of the subsampling scheme, this prior translates into the first case having its mean and modal occurrence dates on 26 October and 7 November 2020, respectively. This prior also implies that the first case of COVID-19 has (1) zero probability of having happened after 20 December 2019, and (2) asymptotically zero probability of happening before mid 2019 (Fig. S3).

#### 4.3.2 Priors for sampling proportion *s*

Under the MTBD model, 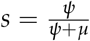 describes the proportion of sampled individuals out of all sampled and removed individuals. In the time interval prior to the first sample no sampling has happened, and hence *s* = 0. After the first sample, *s* has an upper-limit *u* such that *s ≤ u*:

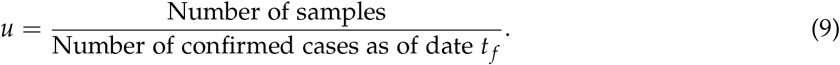

The denominator on the right-hand side comes from the assumption that once an individual has become a confirmed case, they are removed from the infectious pool (i.e., we assume *r* = 1). If a large proportion of infections are asymptomatic or mildly symptomatic, as is the case of COVID-19 (Day, 2020b,a; Li et al., 2020a; Lu et al., 2020), *s* can be an order of magnitude smaller than *u*.

In the equation above, note that *u* can be specific to a deme *d*, in which case we only count individuals from deme *d* when calculating *u*_*d*_. *u*_*d*_ is also alignment specific; for example, in the “small-active” alignment (see Section 2) where New Zealand is the target island, *u*_*IS*_ = 0.147 and *u*_*RW*_ = 8.22 × 10^−5^ (3 sf). The full set of values are available in the GitHub repository accompanying this article. The number of confirmed COVID-19 cases was compiled by https://www.worldometers.info/coronavirus/, and we programmatically accessed these figures from the Worldometers Daily Data GitHub repository by David Bumbeishvili (https://github.com/bumbeishvili/covid19-daily-data).

We reparameterised *s* as 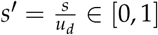 so that a natural prior choice for *s*^*t*^ would be a Beta distribution. More specifically, we assumed *s*^*t*^ *∼* Beta(*α* = 1.1, *β* = 8.0), which sets the mean of *s* at 0.12*u*_*d*_ and bounds it at 0.0 and *u*_*d*_.

**Table S7:** Alignment-specific upper limits for *s* in the MTBD model. Rounded to 3 sf.

| Alignment | Island $\mathcal{IS}$ | $u_{\mathcal{IS}}$ | $u_{\mathcal{RW}}$ |
| --- | --- | --- | --- |
| Small-active | Australia | 0.0373 | $8.62 \times 10^{-5}$ |
| | Iceland | 0.139 | $8.98 \times 10^{-5}$ |
| | New Zealand | 0.147 | $8.22 \times 10^{-5}$ |
| | Taiwan | 0.255 | $8.52 \times 10^{-5}$ |
| Small-time | Australia | 0.0369 | $7.61 \times 10^{-5}$ |
| | Iceland | 0.138 | $8.83 \times 10^{-5}$ |
| | New Zealand | 0.147 | $8.32 \times 10^{-5}$ |
| | Taiwan | 0.255 | $7.44 \times 10^{-5}$ |
| Large-active | Australia | 0.0372 | $1.98 \times 10^{-4}$ |
| | Iceland | 0.14 | $1.96 \times 10^{-4}$ |
| | New Zealand | 0.147 | $2.02 \times 10^{-4}$ |
| | Taiwan | 0.255 | $1.91 \times 10^{-4}$ |
| Large-time | Australia | 0.0369 | $1.89 \times 10^{-4}$ |
| | Iceland | 0.138 | $1.93 \times 10^{-4}$ |
| | New Zealand | 0.147 | $1.8 \times 10^{-4}$ |
| | Taiwan | 0.255 | $1.92 \times 10^{-4}$ |

#### 4.3.3 Priors for MTBD geographical frequencies *π*_*G*_

Under our MTBD model, the equilibrium frequency of each deme is assumed to be proportional to that deme’s human population *H*_*d*_. In the case of *RW, H*_*d*_ is the sum of the population of countries represented in an alignment, so this quantity is thus dependent on the subsampling scheme. *π*_*G*_ is given by:

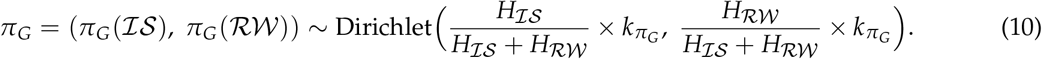

Scalar *k*_*πG*_ controls the variance and was set to 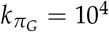. The full set of priors for *π*^*G*^ is presented in Table S8.

**Table S8:** Alignment-specific priors for *π*_*G*_ in the MTBD model. “Dir” stands for Dirichlet.

| Alignment | Island $\mathcal{IS}$ | Prior of $\pi_G$ |
| --- | --- | --- |
| Small-active | Australia | Dir(68.2, 9938.1) |
|  | Iceland | Dir(1, 9999) |
|  | New Zealand | Dir(20.6, 9979.4) |
|  | Taiwan | Dir(66.8, 9933.2) |
| Small-time | Australia | Dir(58.8, 9941.2) |
|  | Iceland | Dir(0.9, 9999.1) |
|  | New Zealand | Dir(11.9, 9988.1) |
|  | Taiwan | Dir(53.3, 9946.7) |
| Large-active | Australia | Dir(63.6, 9936.4) |
|  | Iceland | Dir(0.9, 9999.1) |
|  | New Zealand | Dir(13, 9987) |
|  | Taiwan | Dir(60.1, 9939.9) |
| Large-time | Australia | Dir(55, 9945) |
|  | Iceland | Dir(0.8, 9999.2) |
|  | New Zealand | Dir(10.5, 9989.5) |
|  | Taiwan | Dir(54, 9946) |

**Table S9:**
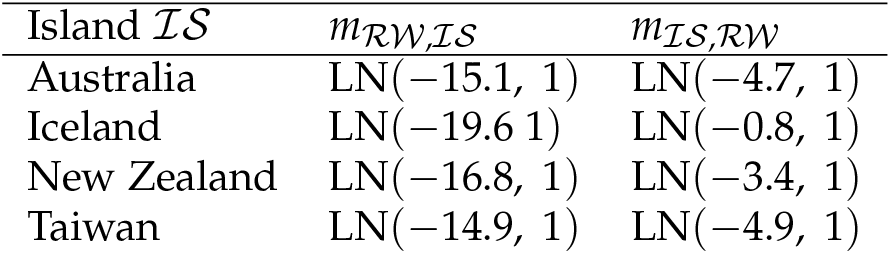
Alignment-specific priors for *m* in the SC model for the “small-active” method. “LN” is a LogNormal distribution.

**Fig. S3:**
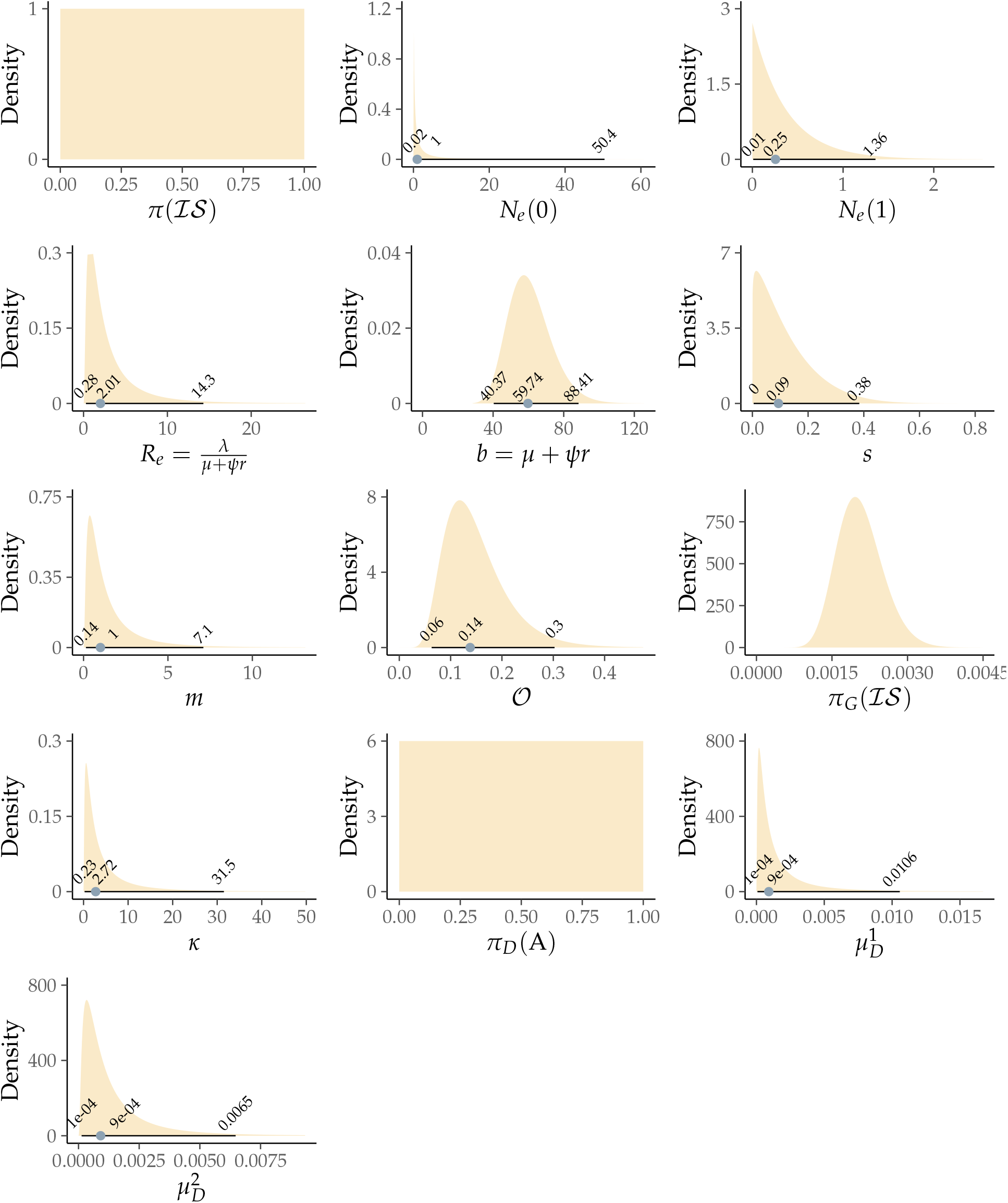
Prior probability distributions used in our models for phylodynamic analyses. Parameters appear in the same order as in Table S6. Black horizontal lines indicate the exact intervals between 2.5% and 97.5% quantiles for the different priors; the blue dot indicates the 50% quantile (Dirichlet priors are multivariate distributions with non-unique quantiles, so those are not shown for these priors). The prior for *N*_*e*_ (1) assumes *N*_*e*_ (1) ∼ Exp(E[Ne (0)]). Only one dimension of each Dirichlet priors is shown. These dimensions are: π(ℛ𝒲) = 1 π() for MTBD geographical frequencies π; πG (ℐ 𝒮) = 1 − *π*_*G*_ (ℛ𝒲) for DPG geographical frequencies *π*_*G*_ ; and *π*_*D*_ (A) = 1 − *π*_*D*_ (C) − *π*_D_ (G) − *π*_*D*_ (T) for nucleotide frequencies *π*_*D*_.

## 5 Model implementation and parameter inference

All models used in this study are implemented in BEAST 2.6 (Bouckaert et al., 2019). Parameter inference is carried out using the Metropolis-Hastings algorithm, which generates a Markov chain that explores the posterior distribution by Monte Carlo simulation (MCMC). We thus employed MCMC to sample (*T, µ*_*c*_, *θ*_*τ*_, *θ*_*s*_) *∼ f* (*𝒯, µ*_*c*_, *θ*_*τ*_, *θ*_*s*_|*D, y*). We used a combination of MCMC and coupled MCMC (MC3; Müller and Bouckaert (2019)), as determined by examining either method’s performance on a chain-by-chain basis. Chain convergence was evaluated by observing a minimum ESS of 200 for the posterior, likelihood, and prior densities, as well as for all reported parameters (Tables S10, S11, S12, S13), with phylogenetic tree convergence inferred from a high correlation between posterior clade probabilities from independent chains (Fig. S4). Due to computational limitations, the “large” alignments did not fully converge for the MTBD or SC methods, and neither did the “small-active” method for MTBD. The results presented in the main text are not derived from the aforementioned chains. This highlights the appeal in the DPG model.

**Table S10:** Effective sample sizes (ESS) under the MTBD model. ESSes are in bold if they are problematic (i.e., less than 100). For vector parameters (such as *κ, R*_*e*_, and *b*), the minimum ESS is reported.

| Alignment | Island | Chain length (10<sup>6</sup>) | posterior | likelihood | prior | $\mu$ | Height | $\kappa$ | $R_e$ | $b$ | $m$ | $s$ | $\pi_G$ |
| --- | --- | --- | --- | --- | --- | --- | --- | --- | --- | --- | --- | --- | --- |
| Large-active | New Zealand | 93 | 158 | 320 | 170 | 187 | 355 | 2911 | **19** | **24** | **19** | 1135 | 4661 |
| Large-active | Australia | 110 | **31** | 214 | **34** | **37** | 129 | 3212 | **18** | **49** | **10** | 1175 | 4942 |
| Large-active | Iceland | 234 | 104 | 104 | 105 | 162 | 1909 | 3271 | 742 | 294 | 546 | 1734 | 11703 |
| Large-active | Taiwan | 162 | 277 | 411 | 238 | 345 | 360 | 4984 | 2075 | 2179 | 2309 | 2188 | 8091 |
| Large-time | New Zealand | 91 | 100 | 101 | 117 | 145 | 404 | 3101 | **23** | **11** | 471 | 971 | 3956 |
| Large-time | Australia | 122 | **65** | 211 | **79** | 195 | 1365 | 3823 | 224 | **42** | 130 | 1830 | 5910 |
| Large-time | Iceland | 218 | **41** | 110 | **44** | **74** | 116 | 6948 | **5** | **5** | **5** | 620 | 10901 |
| Large-time | Taiwan | 137 | 285 | 452 | 279 | 435 | 427 | 4779 | 1250 | **30** | 1597 | 3181 | 6322 |
| Small-active | New Zealand | 158 | **70** | 1497 | **68** | **70** | **98** | 2599 | 1208 | 488 | 125 | 940 | 7496 |
| Small-active | Australia | 122 | **42** | 123 | **43** | **39** | **59** | 3036 | 227 | 243 | 262 | 333 | 5600 |
| Small-active | Iceland | 374 | 173 | 1716 | 162 | 179 | 452 | 7905 | 1558 | 1083 | 434 | 9399 | 18679 |
| Small-active | Taiwan | 268 | 376 | 3165 | 355 | 320 | 444 | 9041 | 147 | 1034 | 143 | 8007 | 13204 |
| Small-time | New Zealand | 174 | 291 | 1930 | 396 | 414 | 976 | 5593 | 812 | 1787 | 840 | 6360 | 8243 |
| Small-time | Australia | 214 | 413 | 2683 | 397 | 573 | 1656 | 8106 | 2820 | 1049 | 1496 | 6411 | 10693 |
| Small-time | Iceland | 409 | 325 | 1030 | 295 | 404 | 1291 | 12747 | 265 | 684 | 159 | 14746 | 20189 |
| Small-time | Taiwan | 280 | 587 | 2128 | 533 | 730 | 2224 | 10535 | 1637 | 628 | 1957 | 7266 | 14023 |

**Table S11:** Effective sample sizes under the SC model. See Table S10 caption for notation.

| Alignment | Island | Chain length (10<sup>6</sup>) | posterior | likelihood | prior | $\mu$ | Height | $\kappa$ | $N_e$ | $m$ |
| --- | --- | --- | --- | --- | --- | --- | --- | --- | --- | --- |
| Large-active | New Zealand | 269 | 121 | **26** | 211 | 310 | 181 | 5289 | 269 | 1703 |
| Large-active | Australia | 204 | 343 | **91** | 294 | 403 | 118 | 15768 | 379 | 1364 |
| Large-active | Iceland | 240 | 321 | **31** | **72** | 171 | 292 | 17602 | 106 | 496 |
| Large-active | Taiwan | 278 | 570 | 163 | 422 | 420 | 157 | 22350 | 536 | 1897 |
| Large-time | New Zealand | 269 | 351 | **43** | 290 | 266 | 381 | 21108 | 240 | 546 |
| Large-time | Australia | 317 | 668 | 340 | 407 | 806 | 299 | 26585 | 787 | 1782 |
| Large-time | Iceland | 253 | 441 | 195 | 374 | 675 | 305 | 20296 | 553 | 1936 |
| Large-time | Taiwan | 279 | 649 | 122 | 437 | 574 | 342 | 18439 | 426 | 1332 |
| Small-active | New Zealand | 270 | 331 | **66** | 1055 | 2036 | 804 | 16208 | 1612 | 3029 |
| Small-active | Australia | 240 | 833 | 334 | 691 | 1054 | 445 | 16094 | 1040 | 2771 |
| Small-active | Iceland | 240 | 1143 | 1106 | 1006 | 1953 | 560 | 12543 | 1650 | 2298 |
| Small-active | Taiwan | 270 | 1240 | 175 | 378 | 556 | 720 | 20527 | 557 | 4134 |
| Small-time | New Zealand | 270 | 770 | 669 | 526 | 217 | 668 | 11967 | 203 | 634 |
| Small-time | Australia | 240 | 768 | 418 | 408 | 986 | 334 | 16274 | 818 | 2333 |
| Small-time | Iceland | 240 | 820 | 158 | 249 | 604 | 1092 | 14314 | 344 | 1236 |
| Small-time | Taiwan | 270 | 1432 | 1735 | 835 | 1136 | 1687 | 15362 | 1111 | 2972 |

**Table S12:**
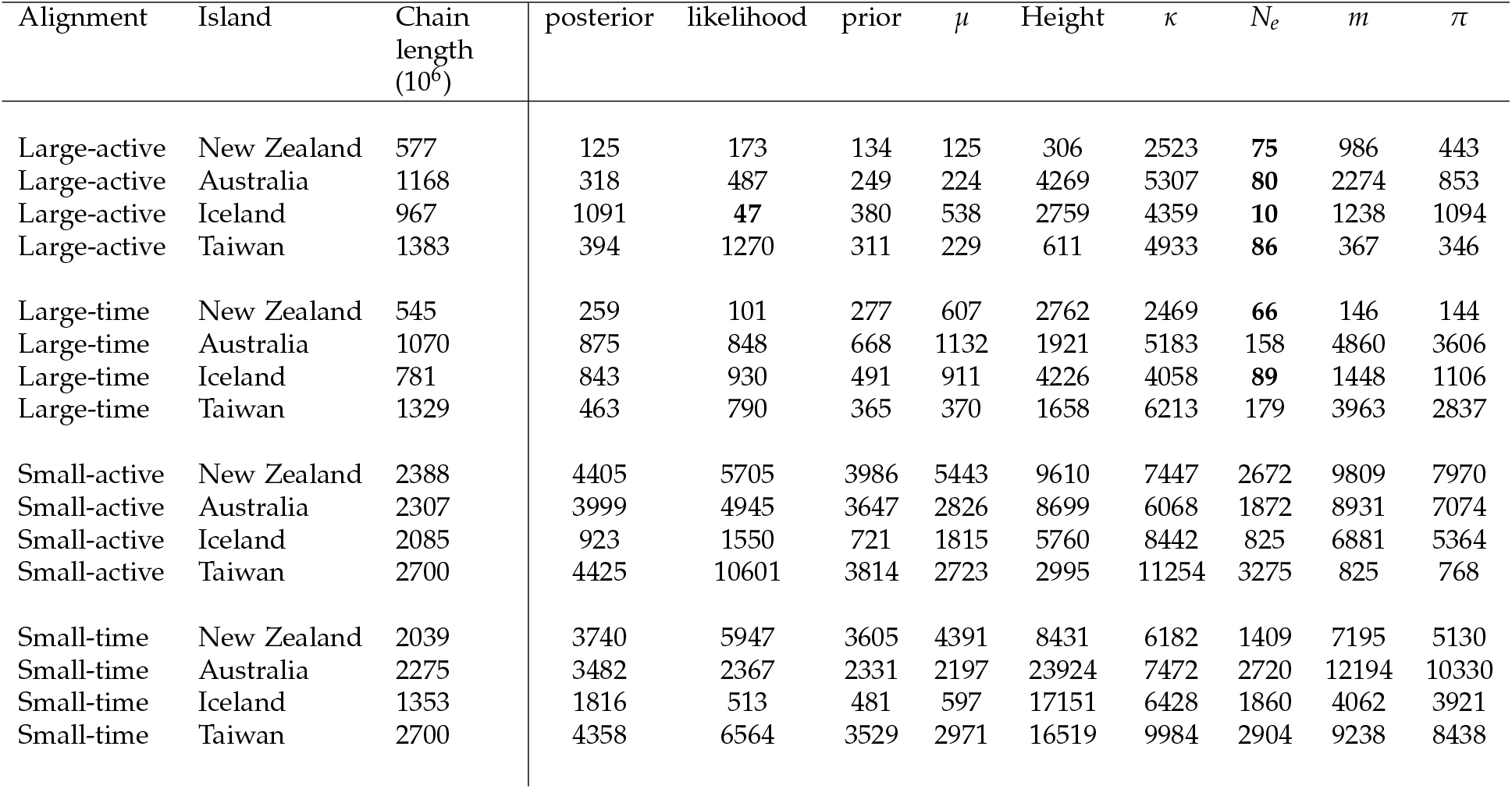
Effective sample sizes under the DPG model. See Table S10 caption for notation.

| Alignment | Island | Chain length (10<sup>6</sup>) | posterior | likelihood | prior | $\mu$ | Height | $\kappa$ | $N_e$ | $m$ | $\pi$ |
| --- | --- | --- | --- | --- | --- | --- | --- | --- | --- | --- | --- |
| Large-active | New Zealand | 577 | 125 | 173 | 134 | 125 | 306 | 2523 | **75** | 986 | 443 |
| Large-active | Australia | 1168 | 318 | 487 | 249 | 224 | 4269 | 5307 | **80** | 2274 | 853 |
| Large-active | Iceland | 967 | 1091 | **47** | 380 | 538 | 2759 | 4359 | **10** | 1238 | 1094 |
| Large-active | Taiwan | 1383 | 394 | 1270 | 311 | 229 | 611 | 4933 | **86** | 367 | 346 |
| Large-time | New Zealand | 545 | 259 | 101 | 277 | 607 | 2762 | 2469 | **66** | 146 | 144 |
| Large-time | Australia | 1070 | 875 | 848 | 668 | 1132 | 1921 | 5183 | 158 | 4860 | 3606 |
| Large-time | Iceland | 781 | 843 | 930 | 491 | 911 | 4226 | 4058 | **89** | 1448 | 1106 |
| Large-time | Taiwan | 1329 | 463 | 790 | 365 | 370 | 1658 | 6213 | 179 | 3963 | 2837 |
| Small-active | New Zealand | 2388 | 4405 | 5705 | 3986 | 5443 | 9610 | 7447 | 2672 | 9809 | 7970 |
| Small-active | Australia | 2307 | 3999 | 4945 | 3647 | 2826 | 8699 | 6068 | 1872 | 8931 | 7074 |
| Small-active | Iceland | 2085 | 923 | 1550 | 721 | 1815 | 5760 | 8442 | 825 | 6881 | 5364 |
| Small-active | Taiwan | 2700 | 4425 | 10601 | 3814 | 2723 | 2995 | 11254 | 3275 | 825 | 768 |
| Small-time | New Zealand | 2039 | 3740 | 5947 | 3605 | 4391 | 8431 | 6182 | 1409 | 7195 | 5130 |
| Small-time | Australia | 2275 | 3482 | 2367 | 2331 | 2197 | 23924 | 7472 | 2720 | 12194 | 10330 |
| Small-time | Iceland | 1353 | 1816 | 513 | 481 | 597 | 17151 | 6428 | 1860 | 4062 | 3921 |
| Small-time | Taiwan | 2700 | 4358 | 6564 | 3529 | 2971 | 16519 | 9984 | 2904 | 9238 | 8438 |

**Table S13:** Effective sample sizes under the DPG2 model. See Table S10 caption for notation.

| Alignment | Island | Chain length<br>(10<sup>6</sup>) | posterior | likelihood | prior | $\mu$ | Height | $\kappa$ | $N_e$ | $m$ | $\pi$ |
| --- | --- | --- | --- | --- | --- | --- | --- | --- | --- | --- | --- |
| Large-active | New Zealand | 328 | 170 | 263 | 294 | 293 | 827 | 3888 | 102 | 2583 | 2229 |
| Large-active | Australia | 357 | 396 | 586 | 412 | 489 | 2905 | 5104 | 189 | 1499 | 1730 |
| Large-active | Iceland | 257 | 100 | 180 | **87** | 162 | 1675 | 3669 | **57** | 1246 | 1001 |
| Large-active | Taiwan | 439 | 359 | 985 | 328 | 286 | 739 | 6557 | 161 | 749 | 538 |
| Large-time | New Zealand | 315 | 386 | **26** | 222 | 351 | 2372 | 5714 | **78** | 2427 | 2408 |
| Large-time | Australia | 320 | 580 | 1139 | 428 | 806 | 2647 | 5170 | 460 | 904 | 848 |
| Large-time | Iceland | 241 | 645 | 791 | 561 | 967 | 3426 | 4569 | 254 | 1326 | 751 |
| Large-time | Taiwan | 418 | 670 | 477 | 644 | 530 | 1038 | 6445 | 188 | 1736 | 1694 |
| Small-active | New Zealand | 180 | 626 | 406 | 474 | 556 | 1019 | 2727 | 651 | 214 | 146 |
| Small-active | Australia | 180 | 662 | 1000 | 659 | 642 | 1408 | 2774 | 431 | 1593 | 1456 |
| Small-active | Iceland | 180 | 921 | 1738 | 800 | 566 | 2135 | 1840 | 256 | 676 | 615 |
| Small-active | Taiwan | 900 | 5977 | 9534 | 4677 | 4292 | 5677 | 16069 | 2076 | 5533 | 5474 |
| Small-time | New Zealand | 180 | 1201 | **81** | 1037 | 551 | 1547 | 3061 | 370 | 1018 | 990 |
| Small-time | Australia | 180 | 797 | 662 | 642 | 768 | 2886 | 2892 | 445 | 687 | 748 |
| Small-time | Iceland | 180 | 807 | 217 | 500 | 521 | 3600 | 2835 | 967 | 2134 | 1948 |
| Small-time | Taiwan | 817 | 3503 | 4816 | 2634 | 2742 | 12256 | 12609 | 3213 | 2686 | 2415 |

**Fig. S4:**
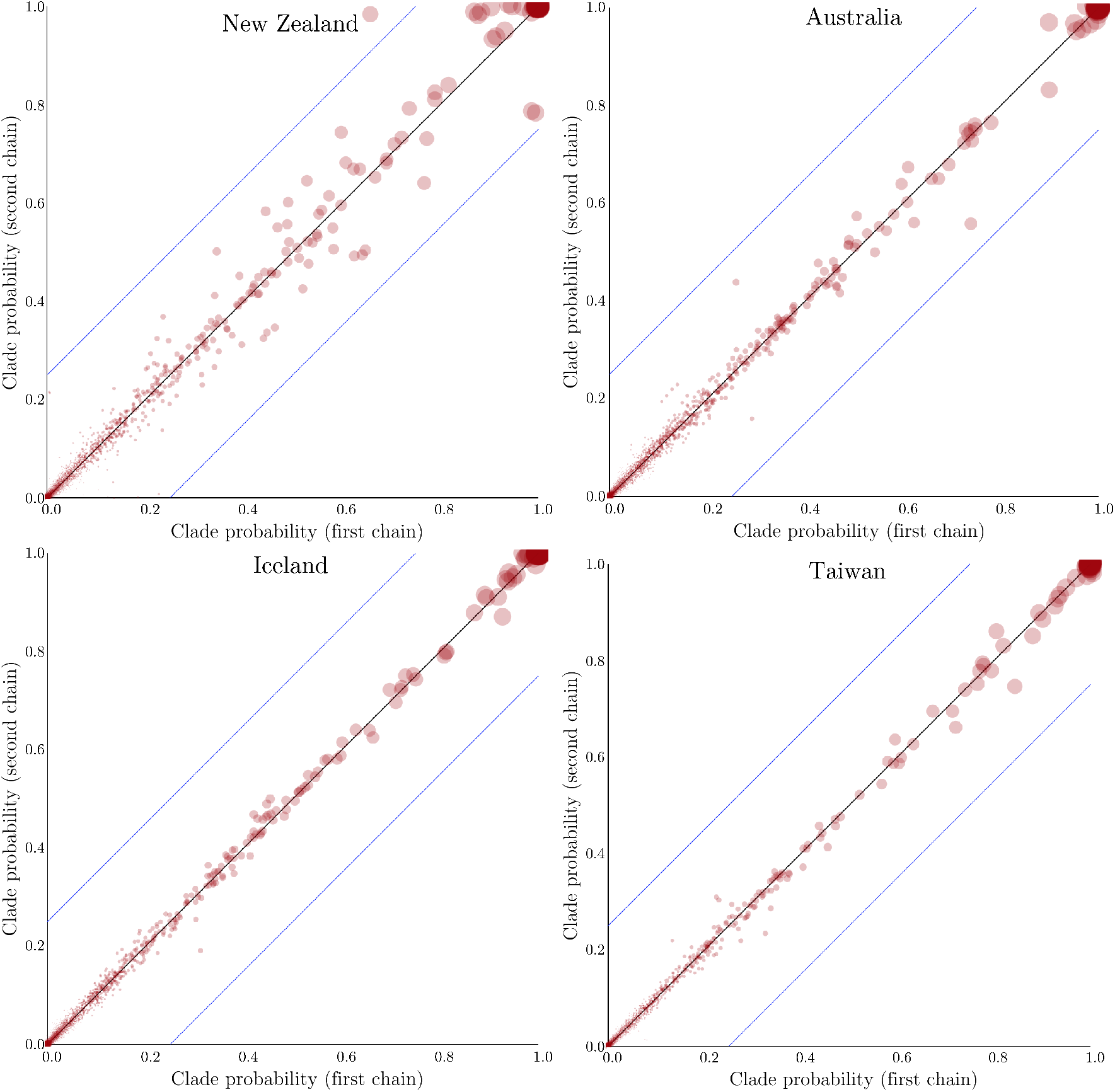
The points on each plot are clade posterior probabilities from two independent MTBD analyses (using the “small-time” alignments). The size of each point is proportional to clade probability. These analysis indicates that tree topologies from independent MCMC chains converged to similar posterior distributions, with most probability differences being significantly less than than 0.25 (blue lines).

## 6 Supplementary results

### 6.1 Parameter estimates

Changes in posterior estimates for MTBD parameters over time intervals (for the “small-active” alignment) are presented in Fig. S5. These results indicate that the rate of becoming non-infectious *b* = *µ* + *ψ* increases over time in all four islands. This results from two mechanisms: 1) in increase in sampling rate *ψ*, likely due to higher rates of SARS-CoV-2 genomic sequencing (Fig. S8), and 2) an increase in death rate *µ*, likely due to improved measures of self-isolation and/or quarantine enforced by the respective governments (Fig. S9). Clock rate and root height estimates vary slightly among differing models and subsampling schemes (Fig. S10). However, the four sampling methods yielded similar results for key MTBD parameters, suggesting that the MTBD analysis was not sensitive to subsampling methods (Fig. S11, S12, S13, and S14).

### 6.2 Introductions through time

The first step in quantifying SARS-CoV-2 introductions into the four *ℐ𝒮* demes is to carry out ancestral state reconstruction (ASR). The main goal of ASR is to sample ancestral states at internal nodes so that states from adjacent nodes can be compared: if the state of a parent node is *ℛ* 𝒲 and that of its child is *ℐ𝒮*, an introduction is inferred. (The sampling procedure is done either simultaneously with the sampling of the tree, or during post-processing by using the parameter values logged during MCMC.) All branches of the tree can then be parsed and introductions annotated according to their chronological distribution, which allows one to plot the number of introductions over time (Fig. S15).

Under all models, ASR is carried out by traversing the tree backward in time, followed by a forward pass during which ancestral states are sampled (note that in a Bayesian framework we sample states at internal nodes, instead of trying to find the marginal or joint collection of states that maximize the likelihood, e.g., Pupko et al., 2000; Yang, 2014). In the case of DPG, DPG2, and MTBD, the tree is peeled backward so that partial likelihoods can be obtained at internal nodes and at the root, and then stochastic mapping is carried out forward in time (Nielsen, 2002; Freyman and Höhna, 2019). Under the SC model, ASR follows an approach similar in spirit to that of Pearl (1982); the procedure has been previously, and thoroughly detailed elsewhere (**?**).

**Fig. S5:**
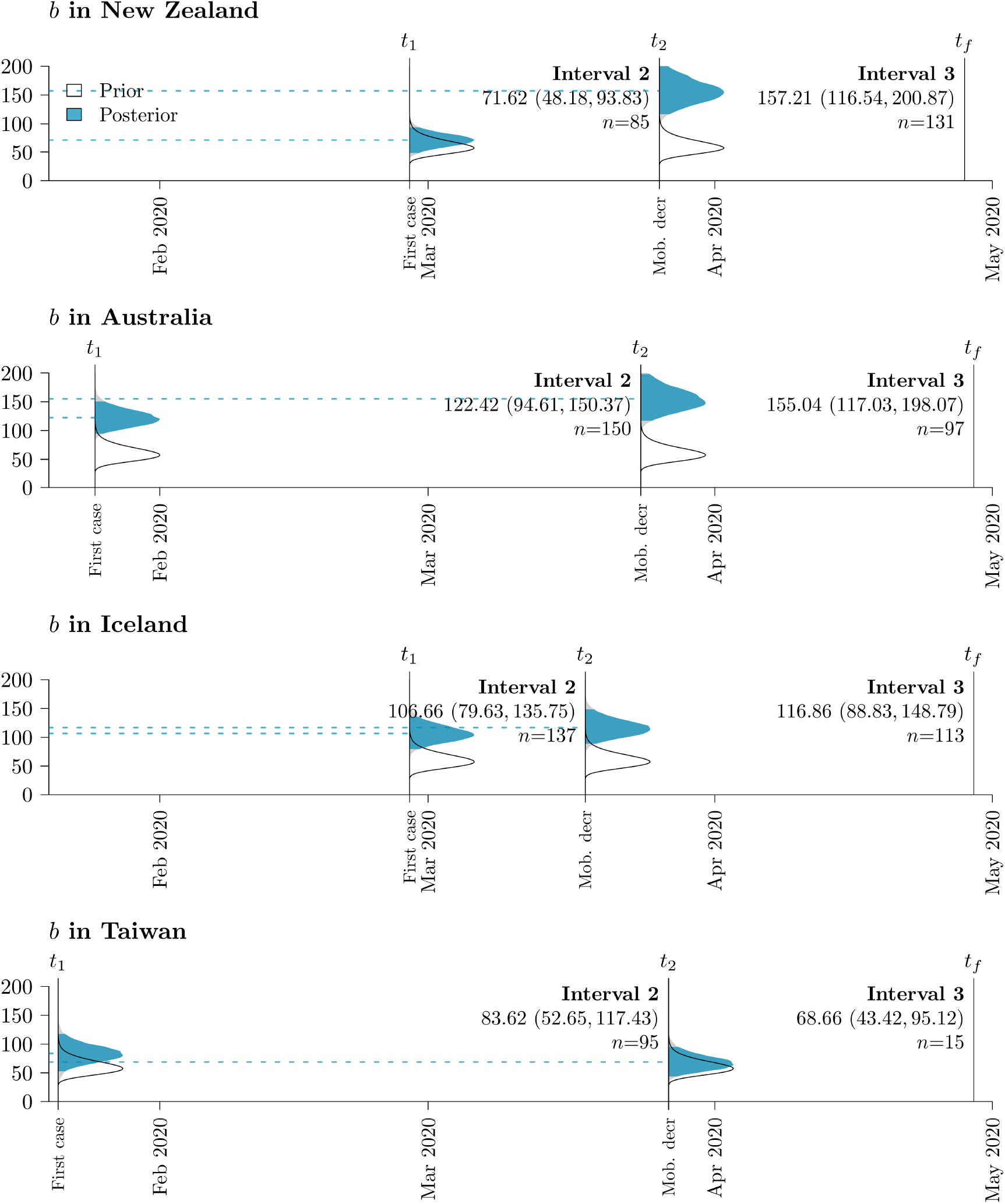
Posterior distribution of the rate of becoming non-infectious *b* across the two epochs following the first reported case in the respective island. Relative prior densities and 95 % HPD intervals (blue) are displayed along the x-axis, with the mean posterior estimate indicated with a dashed line. The number of samples *n* from the specified country within the epoch is reported.

**Fig. S6:**
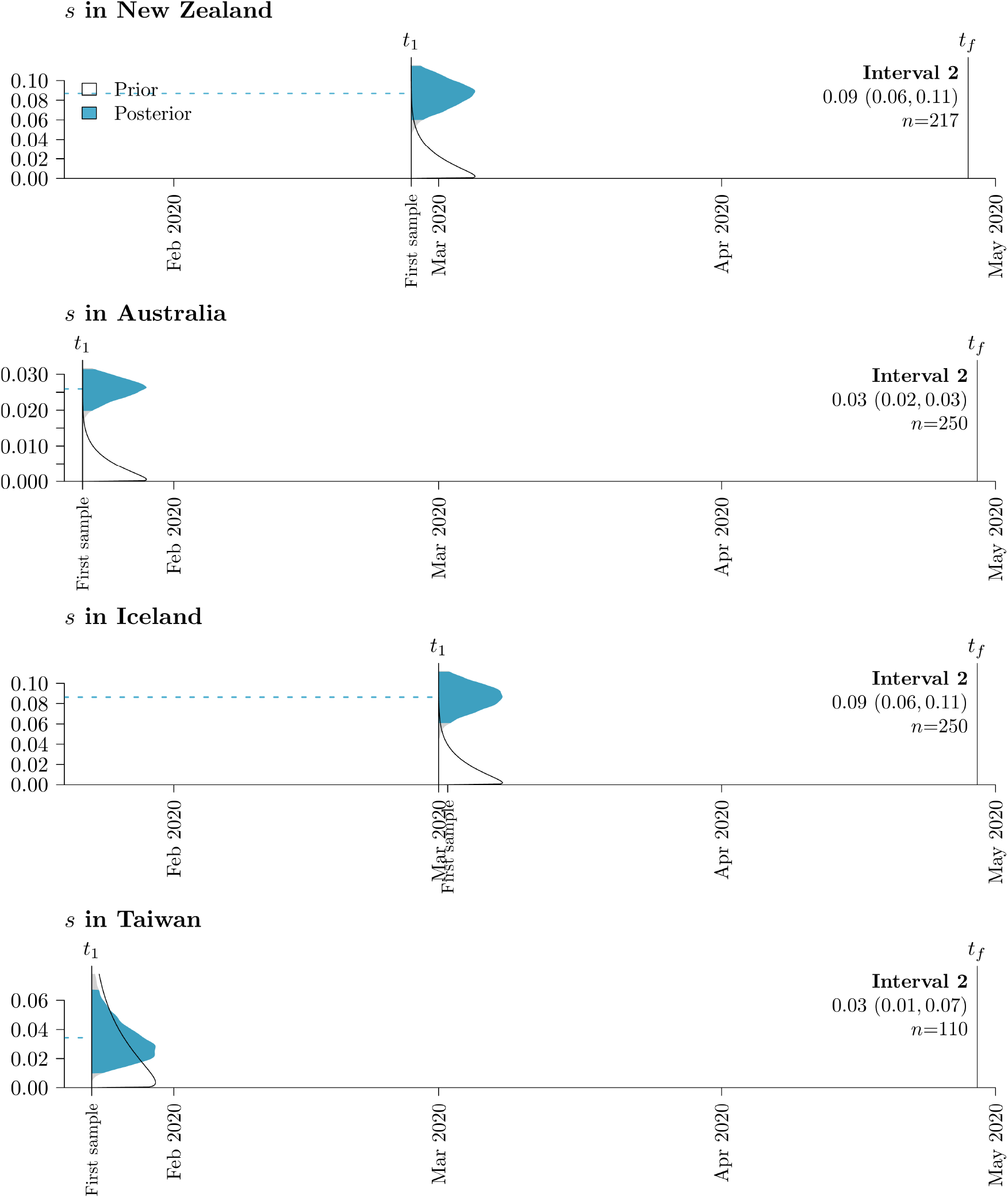
Posterior distribution of sampling proportions *s* after the epoch following the first sample. *s* is held constant at 0 throughout the interval before the first sample. See Fig. S5 for further details on figure notation.

**Fig. S7:**
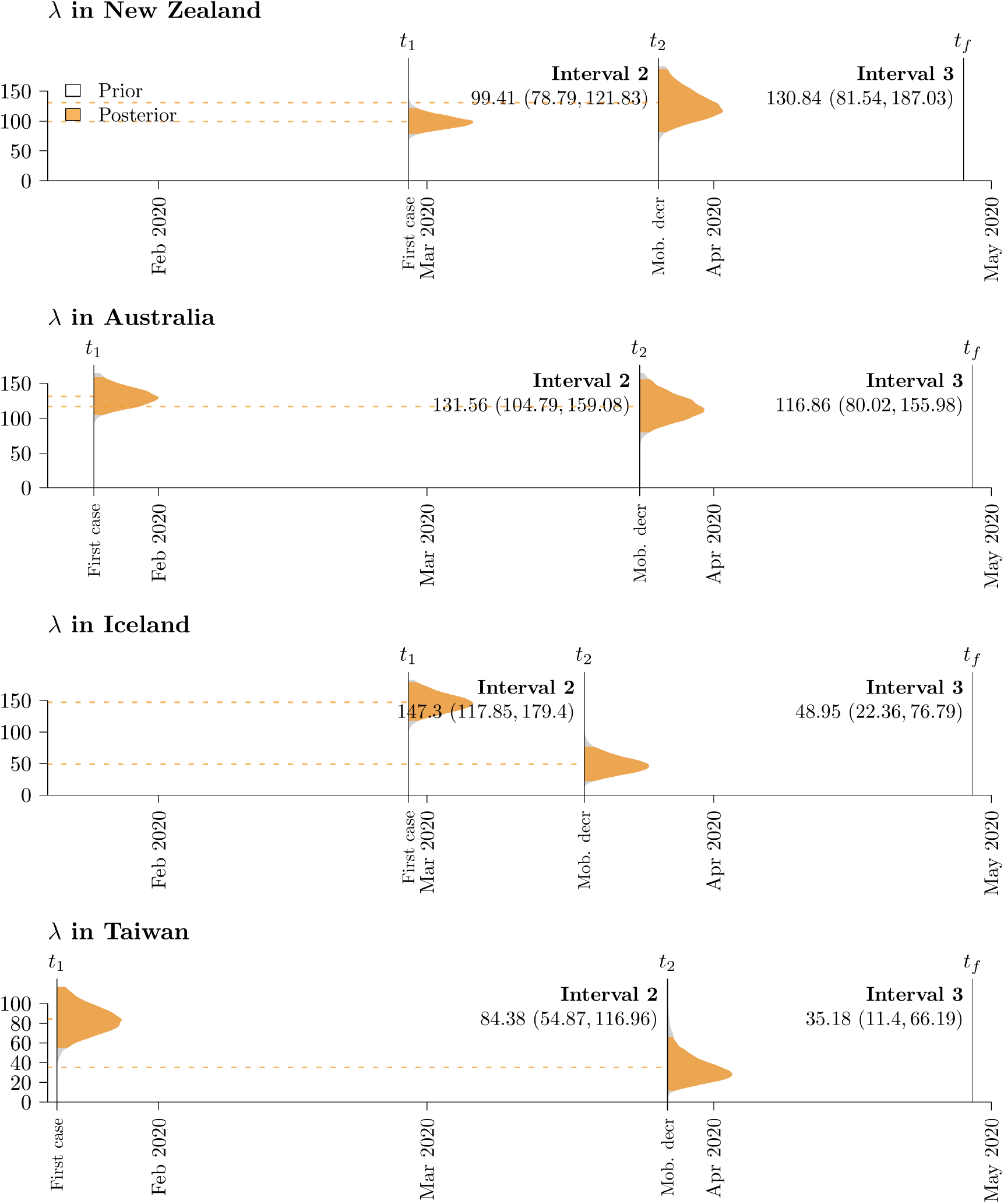
Posterior distribution of the birth rate *λ*, following the first reported case. Although *λ* is not directly estimated as a model parameter, it can be calculated using 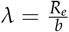. See Fig. S5 for further details on figure notation.

**Fig. S8:**
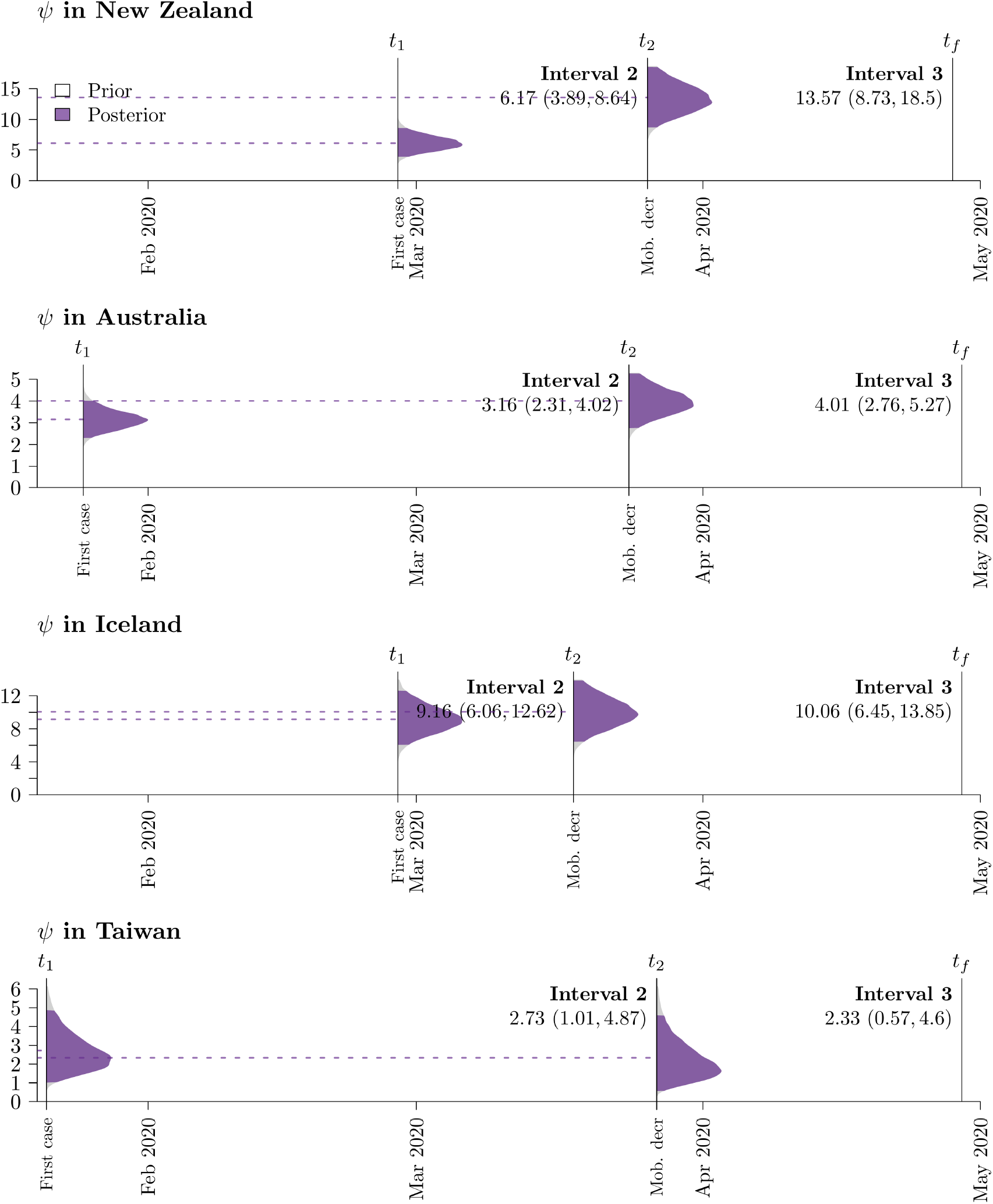
Posterior distribution of the sampling rate *ψ*, following the first reported case. Although *ψ* is not directly estimated as a model parameter, it can be calculated using *ψ* = *sb* (when *r* = 1). See Fig. S5 for further details on figure notation.

**Fig. S9:**
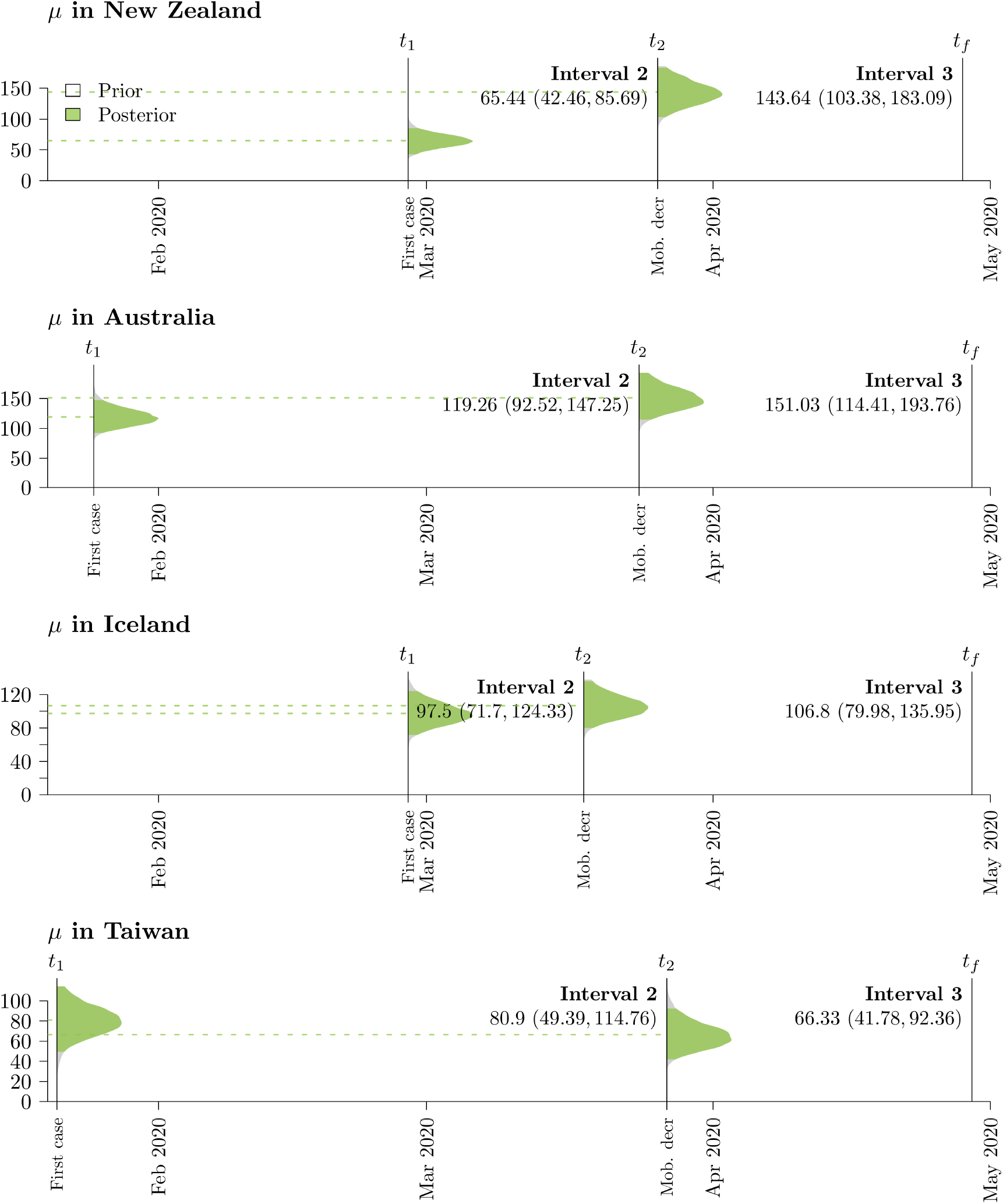
Posterior distribution of the death rate *µ*, following the first reported case. Although *µ* is not directly estimated as a model parameter, it can be calculated using *λ* = *b* −*sb* (when *r* = 1). See Fig. S5 for further details on figure notation.

**Fig. S10:**
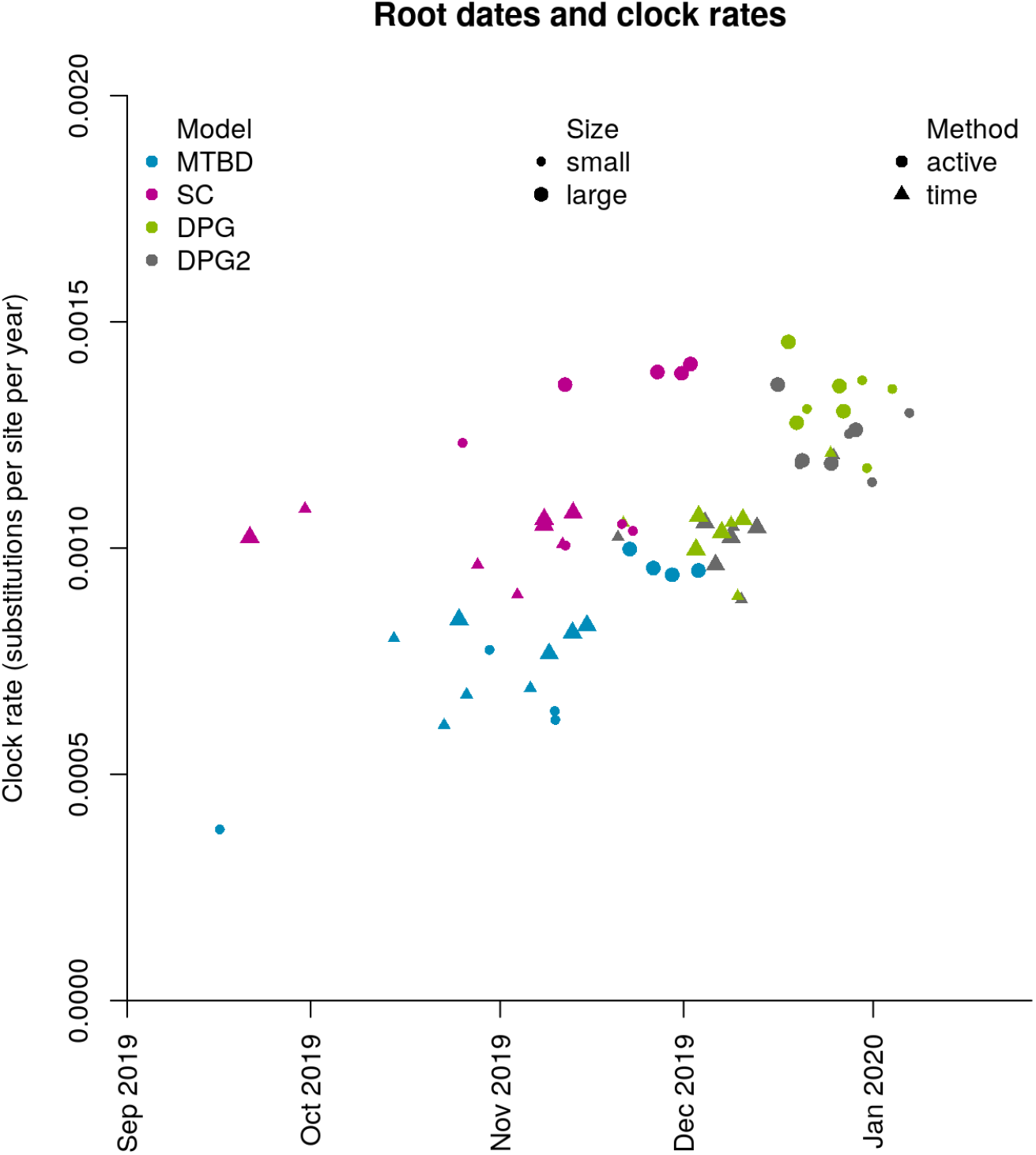
Comparison of mean root height and clock rate estimates across the 64 combinations of sampling methods, models, and islands. These results show that MTBD and SC are less robust to changes in the sample and tend to give, on average, older tree heights. In contrast, DPG and DPG2 both give very late estimates for the root (late Dec - early Jan) when the active sampling method is employed, thus providing further evidence that the active method is not suitable for these models. For MTBD, the “small” datasets yield lower clock rates estimates.

**Fig. S11:**
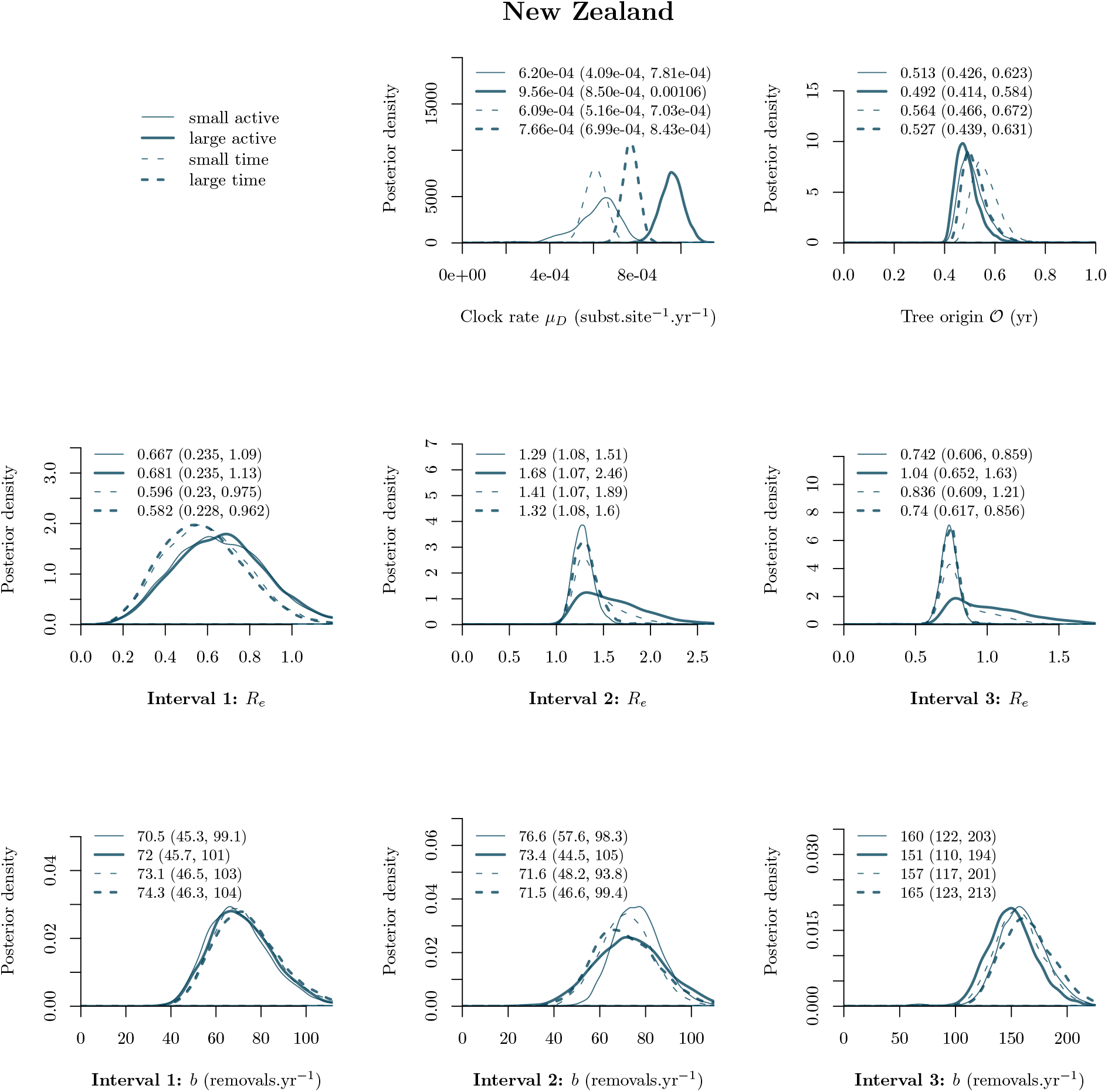
Comparison of the four subsampling methods for New Zealand alignments. Posterior distributions of key parameters from the MTBD analyses are presented above. Alignment-specific mean estimates and 95% highest posterior density intervals are printed above the plots.

**Fig. S12:**
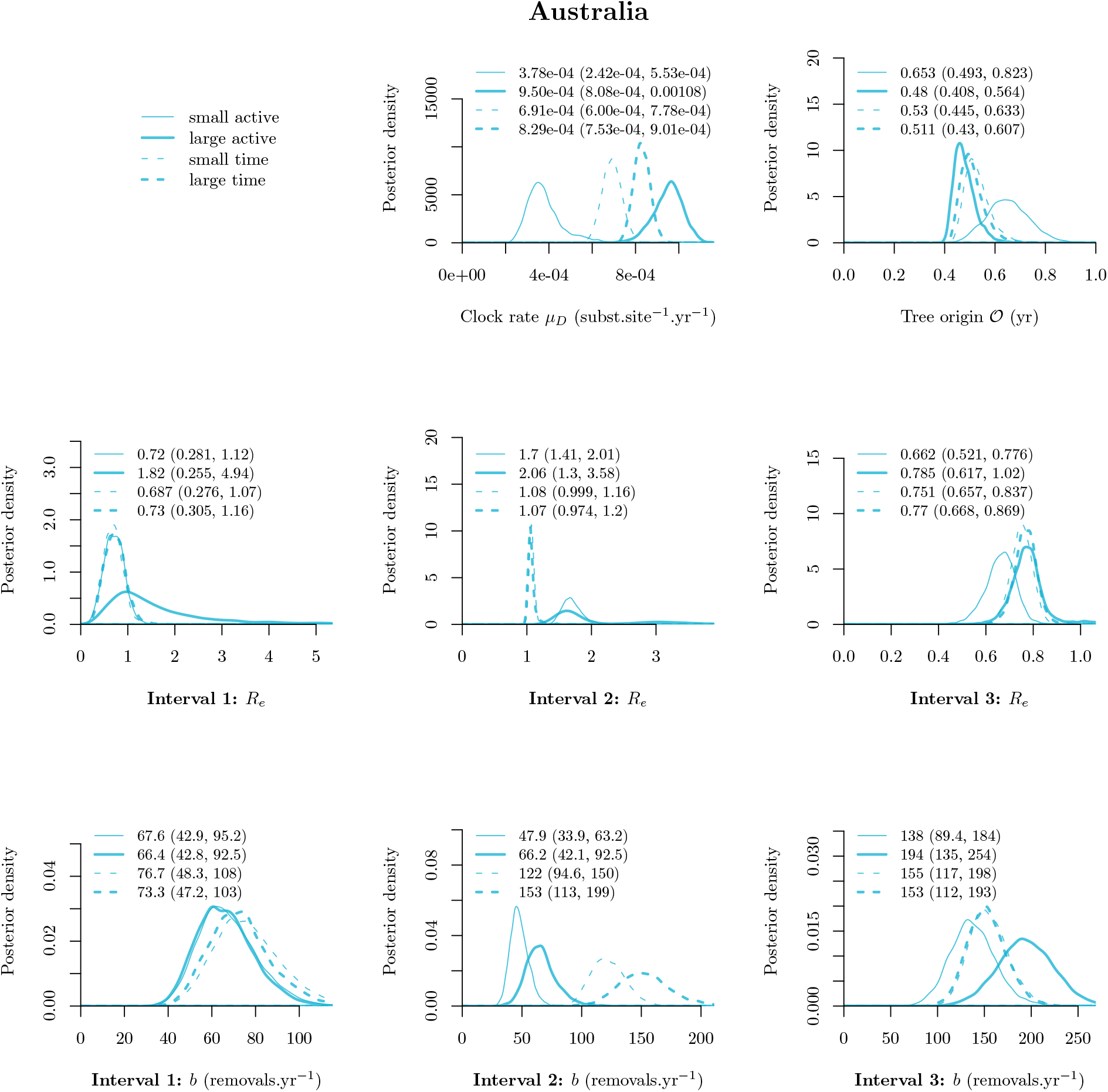
Comparison of subsampling methods for Australia alignments. See Fig. S11 for further details.

**Fig. S13:**
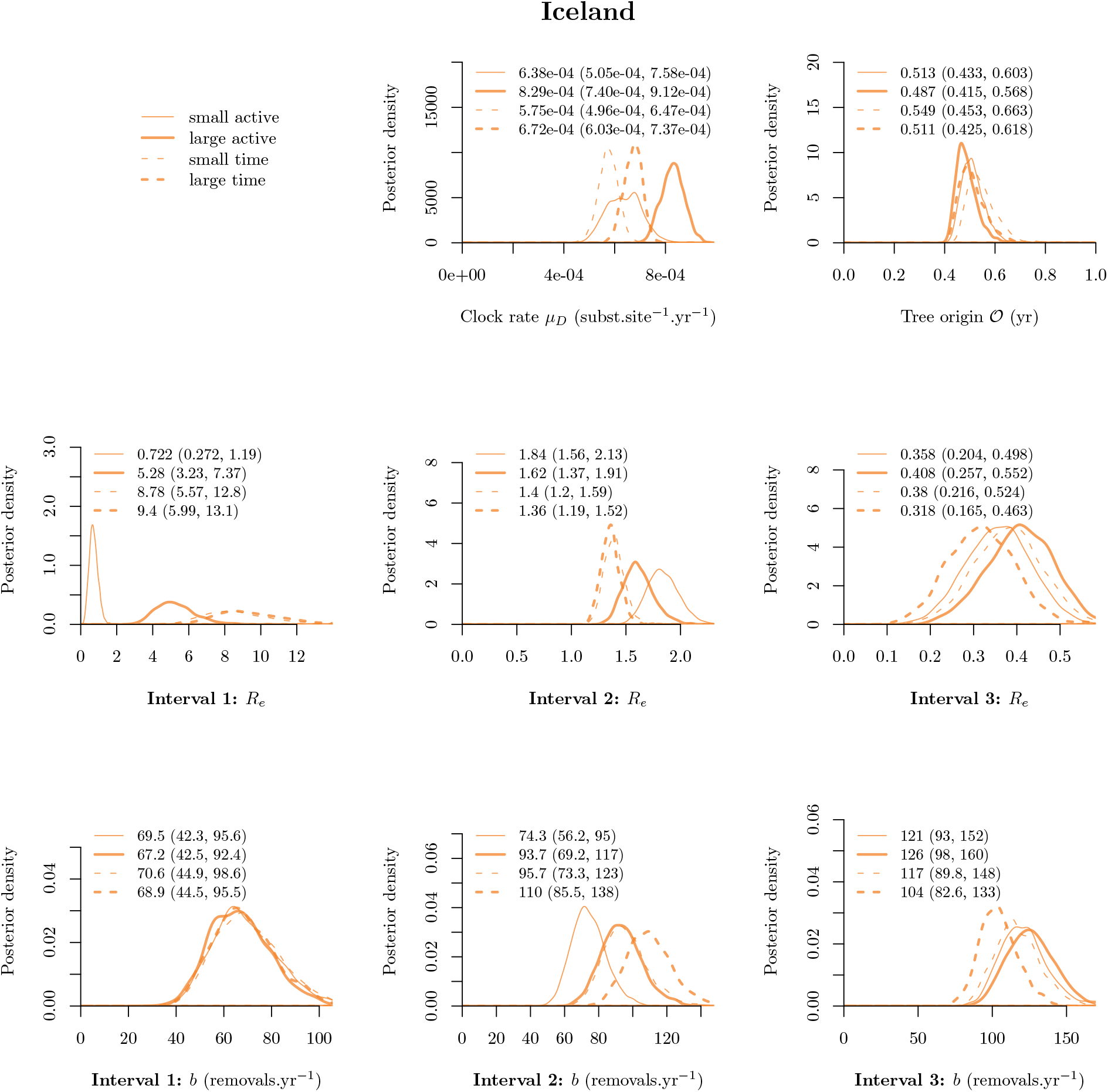
Comparison of subsampling methods for Iceland alignments. See Fig. S11 for further details.

**Fig. S14:**
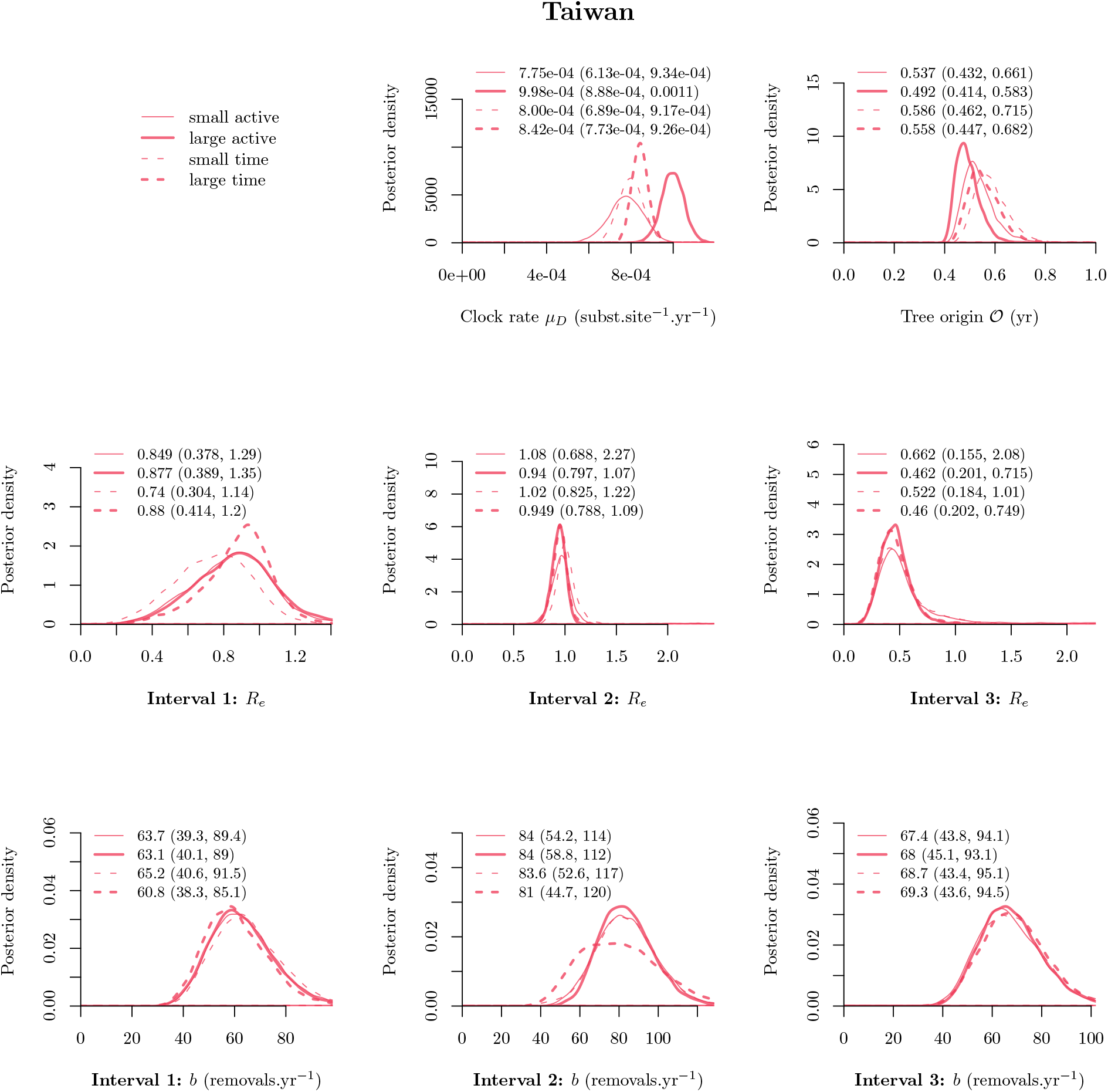
Comparison of subsampling methods for Taiwan alignments. See Fig. S11 for further details.

**Fig. S15:**
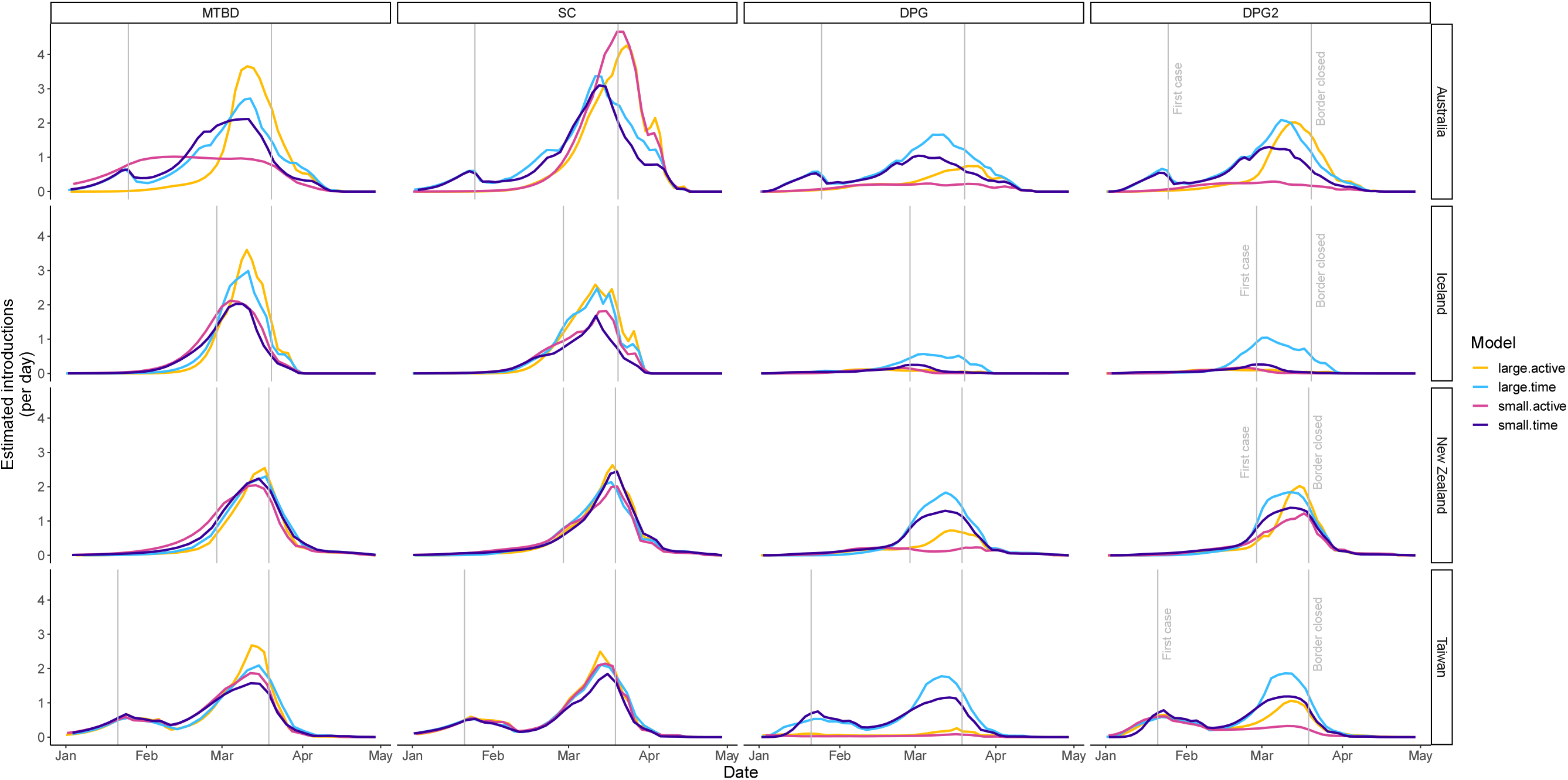
Comparison of subsampling methods on SARS-CoV-2 introductions over time.

**Table S14:** Estimated number of imports and exports (mean and corresponding and 95% HPD intervals) into each target island deme ℐ𝒮 (using the “small-time” subsampling protocol). *Overseas (total)* assumes that the sample is representative of the total proportion of cases linked to overseas travel in Table 1 of the main article. *Overseas (sample)* counts the number of samples that have been marked as having recent overseas travel in the GISAID sequence metadata Shu and McCauley (2017). These counts are likely influenced by missing data. Estimates are highlighted in bold if the expected number is within the 95% HPD interval of either of these two methods.

| Island $\mathcal{IS}$ | Model | Estimated imports | Estimated exports |
| --- | --- | --- | --- |
| New Zealand | *Overseas (total)* | 84 |  |
|  | *Overseas (sample)* | 49 |  |
|  | DPG | **41 [29,51]** | 14[5, 23] |
|  | DPG2 | **41 [30,50]** | 11[4, 20] |
|  | SC | **58 [48,67]** | 0[0, 0] |
|  | MTBD | 63[54, 72] | 1.4[0, 4] |
| Australia | *Overseas (total)* | 159 |  |
|  | *Overseas (sample)* |  |  |
|  | DPG | 49[33, 63] | 34[17, 50] |
|  | DPG2 | 52[38, 65] | 31[16, 44] |
|  | SC | 98[83, 111] | 0[0, 0] |
|  | MTBD | 87[72, 100] | 0.68[0, 3] |
| Iceland | *Overseas (total)* | 47 |  |
|  | *Overseas (sample)* | 68 |  |
|  | DPG | 6.9[6, 9] | 36[26, 46] |
|  | DPG2 | 7[5, 9] | 43[32, 55] |
|  | SC | **37 [25,48]** | 0[0, 0] |
|  | MTBD | **49 [31,64]** | 0.95[0, 3] |
| Taiwan | *Overseas (total)* | 99 |  |
|  | *Overseas (sample)* | 25 |  |
|  | DPG | 48[37, 58] | 21[6, 36] |
|  | DPG2 | 49[38, 58] | 19[6, 32] |
|  | SC | 57[47, 68] | 0[0, 0] |
|  | MTBD | 65[53, 76] | 0.98[0, 4] |

**Table S15:** Estimated imports and exports using the “large-time” method. See Table S14 caption for details.

| Island $\mathcal{IS}$ | Model | Estimated imports | Estimated exports |
| --- | --- | --- | --- |
| New Zealand | *Overseas (total)* | 84 |  |
|  | *Overseas (sample)* | 49 |  |
|  | DPG | **47 [39,55]** | 1.3[0, 4] |
|  | DPG2 | **48 [39,55]** | 2.3[0, 5] |
|  | SC | **55 [47,64]** | 0[0, 0] |
|  | MTBD | 60[50, 67] | 0.52[0, 1] |
| Australia | *Overseas (total)* | 159 |  |
|  | *Overseas (sample)* |  |  |
|  | DPG | 68[55, 80] | 47[27, 67] |
|  | DPG2 | 70[56, 80] | 47[29, 64] |
|  | SC | 110[100, 126] | 0[0, 0] |
|  | MTBD | 92[77, 104] | 2.6[0, 9] |
| Iceland | *Overseas (total)* | 47 |  |
|  | *Overseas (sample)* | 68 |  |
|  | DPG | 19[9, 27] | 65[40, 87] |
|  | DPG2 | 28[18, 37] | 54[31, 76] |
|  | SC | **60 [49,68]** | 0[0, 0] |
|  | MTBD | **62 [53,71]** | 0.7[0, 3] |
| Taiwan | *Overseas (total)* | 99 |  |
|  | *Overseas (sample)* | 25 |  |
|  | DPG | 59[49, 66] | 23[6, 40] |
|  | DPG2 | 60[50, 67] | 29[15, 44] |
|  | SC | 67[59, 74] | 0[0, 0] |
|  | MTBD | 69[60, 77] | 0.93[0, 3] |

**Table S16:** Estimated imports and exports using the “small-active” method. See Table S14 caption for details.

| Island $\mathcal{IS}$ | Model | Estimated imports | Estimated exports |
| --- | --- | --- | --- |
| New Zealand | *Overseas (total)* | 84 |  |
|  | *Overseas (sample)* | 49 |  |
|  | DPG | 13[7, 18] | 41[29, 52] |
|  | DPG2 | **32 [9,49]** | 20[1, 44] |
|  | SC | **54 [43,62]** | 0[0, 0] |
|  | MTBD | 64[55, 71] | 0.13[0, 1] |
| Australia | *Overseas (total)* | 159 |  |
|  | *Overseas (sample)* |  |  |
|  | DPG | 14[8, 20] | 68[52, 80] |
|  | DPG2 | 14[9, 19] | 82[66, 96] |
|  | SC | 120[99, 135] | 0[0, 0] |
|  | MTBD | 72[51, 91] | 5.6[0, 17] |
| Iceland | *Overseas (total)* | 47 |  |
|  | *Overseas (sample)* | 68 |  |
|  | DPG | 4.4[4, 6] | 29[21, 36] |
|  | DPG2 | 4.6[4, 6] | 68[49, 84] |
|  | SC | **47 [36,60]** | 0[0, 0] |
|  | MTBD | **57 [43,69]** | 0.28[0, 1] |
| Taiwan | *Overseas (total)* | 99 |  |
|  | *Overseas (sample)* | 25 |  |
|  | DPG | 4.3[1, 10] | 80[63, 98] |
|  | DPG2 | **26 [16,35]** | 90[68, 110] |
|  | SC | 66[55, 77] | 0[0, 0] |
|  | MTBD | **69 [24,82]** | 11[0, 116] |

**Table S17:**
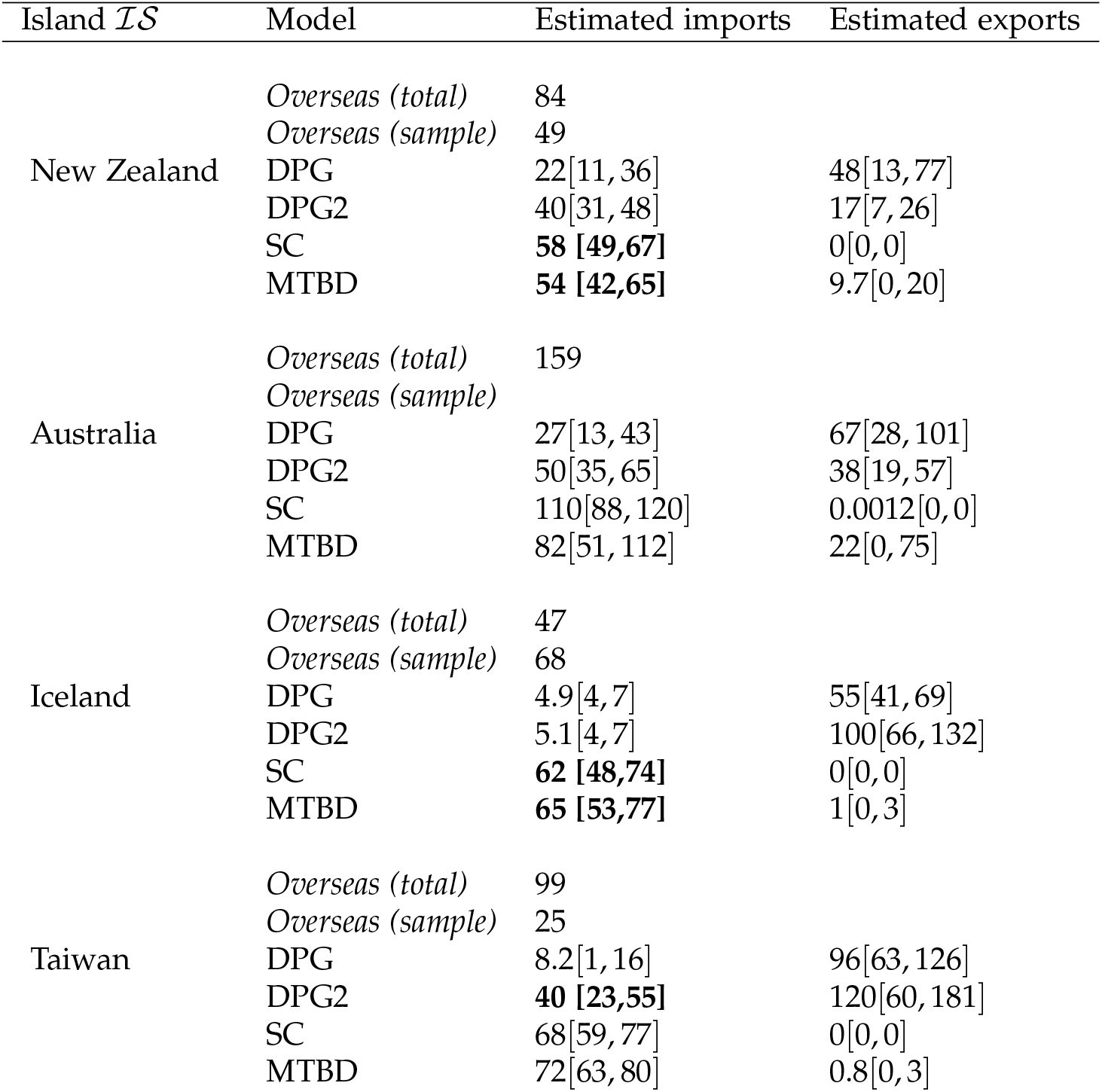
Estimated imports and exports using the “large-active” method. See Table S14 caption for details.

