## Supplementary material for "Phylodynamics reveals the role of human travel and contact tracing in controlling the first wave of COVID-19 in four island nations": GISAID Acknowledgements

We gratefully acknowledge the following Authors from the Originating laboratories responsible for obtaining the specimens and the Submitting laboratories where genetic sequence data were generated and shared via the GISAID Initiative, on which this research is based.

| | Accession | Date | Originating laboratory | Submitting laboratory | Authors |
|---|---|---|---|---|---|
| 1 | Wuhan/IPBCAMS-WH-01/2019 | 2019-12-24 | Institute of Pathogen Biology, Chinese Academy of Medical Sciences & Peking Union Medical College | Institute of Pathogen Biology, Chinese Academy of Medical Sciences & Peking Union Medical College | Lili Ren et al |
| 2 | Wuhan/WH01/2019 | 2019-12-26 | General Hospital of Central Theater Command of People's Liberation Army of China | BGI & Institute of Microbiology, Chinese Academy of Sciences & Shandong First Medical University & Shandong Academy of Medical Sciences & General Hospital of Central Theater Command of People's Liberation Army of China | Weijun Chen et al |
| 3 | Wuhan-Hu-1/2019 | 2019-12-26 | | National Institute for Communicable Disease Control and Prevention (ICDC) Chinese Center for Disease Control and Prevention (China CDC) | Zhang et al |
| 4 | Wuhan/WIV04/2019 | 2019-12-30 | Wuhan Jinyintan Hospital | Wuhan Institute of Virology, Chinese Academy of Sciences | Peng Zhou et al |
| 5 | Wuhan/IPBCAMS-WH-03/2019 | 2019-12-30 | Institute of Pathogen Biology, Chinese Academy of Medical Sciences & Peking Union Medical College | Institute of Pathogen Biology, Chinese Academy of Medical Sciences & Peking Union Medical College | Lili Ren et al |
| 6 | Wuhan/IVDC-HB-05/2019 | 2019-12-30 | National Institute for Viral Disease Control and Prevention, China CDC | National Institute for Viral Disease Control and Prevention, China CDC | Wenjie Tan et al |
| 7 | Wuhan/IPBCAMS-WH-02/2019 | 2019-12-30 | Institute of Pathogen Biology, Chinese Academy of Medical Sciences & Peking Union Medical College | Institute of Pathogen Biology, Chinese Academy of Medical Sciences & Peking Union Medical College | Lili Ren et al |
| 8 | Wuhan/IVDC-HB-GX02/2019 | 2019-12-30 | National Institute for Viral Disease Control and Prevention, China CDC | National Institute for Viral Disease Control and Prevention, China CDC, Yunnan Provincial CDC | Wenjie Tan et al |
| 9 | Wuhan/IVDC-HB-04/2020 | 2020-01-01 | National Institute for Viral Disease Control and Prevention, China CDC | National Institute for Viral Disease Control and Prevention, China CDC | Wenjie Tan et al |
| 10 | Wuhan/WH03/2020 | 2020-01-01 | General Hospital of Central Theater Command of People's Liberation Army of China | BGI & Institute of Microbiology, Chinese Academy of Sciences & Shandong First Medical University & Shandong Academy of Medical Sciences & General Hospital of Central Theater Command of People's Liberation Army of China | Weijun Chen et al |
| 11 | Wuhan/WHU02/2020 | 2020-01-02 | | State Key Laboratory of Virology, Wuhan University | Chen et al |
| 12 | Wuhan/WHU01/2020 | 2020-01-02 | | State Key Laboratory of Virology, Wuhan University | Chen et al |
| 13 | Wuhan/WH04/2020 | 2020-01-05 | General Hospital of Central Theater Command of People's Liberation Army of China | BGI & Institute of Microbiology, Chinese Academy of Sciences & Shandong First Medical University & Shandong Academy of Medical Sciences & General Hospital of Central Theater Command of People's Liberation Army of China | Weijun Chen et al |
| 14 | Thailand/NIH-15/2020 | 2020-01-05 | Bamrasnaradura hospital | National Institute of Health. Department of medical Sciences, Ministry of Public Health, Thailand | Pilailuk et al |
| 15 | Thailand/61/2020 | 2020-01-08 | Bamrasnaradura Hospital | 1. Department of Medical Sciences, Ministry of Public Health, Thailand 2. Thai Red Cross Emerging Infectious Diseases - Health Science Centre 3. Department of Disease | Pilailuk et al |

| | | | | | |
|---|---|---|---|---|---|
| | | | | Control, Ministry of Public Health, Thailand | |
| 16 | China/WH-09/2020 | 2020-01-08 | | Key Laboratory of Human Diseases, Comparative Medicine, Institute of Laboratory Animal Science | Linlin et al |
| 17 | Shenzhen/HKU-SZ-002/2020 | 2020-01-10 | The University of Hong Kong - Shenzhen Hospital | Li Ka Shing Faculty of Medicine, The University of Hong Kong | Chan et al |
| 18 | Jiangxi/IVDC-JX-002/2020 | 2020-01-11 | National Institute for Viral Disease Control and Prevention, China CDC | National Institute for Viral Disease Control & Prevention, CCDC | Wenjie Tan et al |
| 19 | Pingxiang/JX5/2020 | 2020-01-11 | Jiangxi Province Center for Disease Control and Prevention | Jiangxi Province Center for Disease Control and Prevention | JianXiong Li et al |
| 20 | Shenzhen/HKU-SZ-005/2020 | 2020-01-11 | The University of Hong Kong - Shenzhen Hospital | Li Ka Shing Faculty of Medicine, The University of Hong Kong | Chan et al |
| 21 | Shenzhen/SZTH-002/2020 | 2020-01-13 | Shenzhen Key Laboratory of Pathogen and Immunity, National Clinical Research Center for Infectious Disease, Shenzhen Third People's Hospital | Shenzhen Key Laboratory of Pathogen and Immunity, National Clinical Research Center for Infectious Disease, Shenzhen Third People's Hospital | Yang Yang et al |
| 22 | Nepal/61/2020 | 2020-01-13 | National Influenza Centre, National Public Health Laboratory, Kathmandu, Nepal | The University of Hong Kong | Ranjit Sah et al |
| 23 | Thailand/74/2020 | 2020-01-13 | Bamrasnaradura Hospital | 1. Department of Medical Sciences, Ministry of Public Health, Thailand 2. Thai Red Cross Emerging Infectious Diseases - Health Science Centre 3. Department of Disease Control, Ministry of Public Health, Thailand | Pilailuk et al |
| 24 | Guangdong/20SF012/2020 | 2020-01-14 | Guangdong Provincial Center for Diseases Control and Prevention; Guangdong Provincial Public Health | Department of Microbiology, Guangdong Provincial Center for Diseases Control and Prevention | Min Kang et al |
| 25 | Sichuan/IVDC-SC-001/2020 | 2020-01-15 | National Institute for Viral Disease Control and Prevention, China CDC | National Institute for Viral Disease Control & Prevention, CCDC | Wenjie Tan et al |
| 26 | Guangdong/20SF014/2020 | 2020-01-15 | Guangdong Provincial Center for Diseases Control and Prevention; Guangdong Provincial Public Health | Department of Microbiology, Guangdong Provincial Center for Diseases Control and Prevention | Min Kang et al |
| 27 | Guangdong/20SF025/2020 | 2020-01-15 | Guangdong Provincial Center for Diseases Control and Prevention; Guangdong Provincial Public Health | Department of Microbiology, Guangdong Provincial Center for Diseases Control and Prevention | Min Kang et al |
| 28 | Guangdong/20SF013/2020 | 2020-01-15 | Guangdong Provincial Center for Diseases Control and Prevention; Guangdong Provincial Public Health | Department of Microbiology, Guangdong Provincial Center for Diseases Control and Prevention | Min Kang et al |
| 29 | Thailand/NIH-59/2020 | 2020-01-16 | Bamrasnaradura hospital | National Institute of Health. Department of medical Sciences, Ministry of Public Health, Thailand | Pilailuk et al |
| 30 | Shenzhen/SZTH-003/2020 | 2020-01-16 | Shenzhen Key Laboratory of Pathogen and Immunity, National Clinical Research Center for Infectious Disease, Shenzhen Third People's Hospital | Shenzhen Key Laboratory of Pathogen and Immunity, National Clinical Research Center for Infectious Disease, Shenzhen Third People's Hospital | Yang Yang et al |
| 31 | Zhejiang/WZ-01/2020 | 2020-01-16 | Zhejiang Provincial Center for Disease Control and Prevention | Department of Microbiology, Zhejiang Provincial Center for Disease Control and Prevention | Yin Chen et al |
| 32 | Guangdong/20SF028/2020 | 2020-01-17 | Guangdong Provincial Center for Diseases Control and Prevention; Guangdong Provincial Public Health | Department of Microbiology, Guangdong Provincial Center for Diseases Control and Prevention | Min Kang et al |
| 33 | Wuhan/HBCDC-HB-02/2020 | 2020-01-17 | The Central Hospital Of | Hubei Provincial Center for | Bin Fang et al |

| | | | | | |
|---|---|---|---|---|---|
| | | | Wuhan | Disease Control and Prevention | |
| 34 | Zhejiang/WZ-02/2020 | 2020-01-17 | Zhejiang Provincial Center for Disease Control and Prevention | Department of Microbiology, Zhejiang Provincial Center for Disease Control and Prevention | Yanjun Zhang et al |
| 35 | Yunnan/IVDC-YN-003/2020 | 2020-01-17 | National Institute for Viral Disease Control and Prevention, China CDC | National Institute for Viral Disease Control & Prevention, CCDC | Wenjie Tan et al |
| 36 | Wuhan/HBCDC-HB-04/2020 | 2020-01-18 | Union Hospital of Tongji Medical College, Huazhong University of Science and Technology | Hubei Provincial Center for Disease Control and Prevention | Bin Fang et al |
| 37 | Wuhan/HBCDC-HB-03/2020 | 2020-01-18 | Union Hospital of Tongji Medical College, Huazhong University of Science and Technology | Hubei Provincial Center for Disease Control and Prevention | Bin Fang et al |
| 38 | Wuhan/HBCDC-HB-05/2020 | 2020-01-18 | CR&WISCO GENERAL HOSPITAL | Hubei Provincial Center for Disease Control and Prevention | Bin Fang et al |
| 39 | Guangdong/20SF040/2020 | 2020-01-18 | Guangdong Provincial Center for Diseases Control and Prevention; Guangdong Provincial Public Health | Department of Microbiology, Guangdong Provincial Center for Diseases Control and Prevention | Min Kang et al |
| 40 | Chongqing/IVDC-CQ-001/2020 | 2020-01-18 | National Institute for Viral Disease Control and Prevention, China CDC | National Institute for Viral Disease Control & Prevention, CCDC | Wenjie Tan et al |
| 41 | USA/WA1/2020 | 2020-01-19 | Providence Regional Medical Center | Division of Viral Diseases, Centers for Disease Control and Prevention | Queen et al |
| 42 | Jiangsu/IVDC-JS-001/2020 | 2020-01-19 | National Institute for Viral Disease Control and Prevention, China CDC | National Institute for Viral Disease Control & Prevention, CCDC | Wenjie Tan et al |
| 43 | Hangzhou/HZCDC0001/2020 | 2020-01-19 | Hangzhou Center for Disease Control and Prevention | Hangzhou Center for Disease Control and Prevention | Jun Li et al |
| 44 | Shandong/IVDC-SD-001/2020 | 2020-01-19 | National Institute for Viral Disease Control and Prevention, China CDC | National Institute for Viral Disease Control & Prevention, CCDC | Wenjie Tan et al |
| 45 | Hangzhou/HZCDC0013/2020 | 2020-01-20 | Hangzhou Center for Diseases Control and Prevention | Hangzhou Center for Diseases Control and Prevention | Jun Li et al |
| 46 | Hangzhou/HZCDC0012/2020 | 2020-01-20 | Hangzhou Center for Diseases Control and Prevention | Hangzhou Center for Diseases Control and Prevention | Jun Li et al |
| 47 | Hangzhou/HZ-1/2020 | 2020-01-20 | Hangzhou Center for Disease and Control Microbiology Lab | Hangzhou Center for Disease and Control Microbiology Lab | Yu Hua et al |
| 48 | USA/IL1/2020 | 2020-01-21 | IL Department of Public Health Chicago Laboratory | Pathogen Discovery, Respiratory Viruses Branch, Division of Viral Diseases, Centers for Disease Control and Prevention | Ying Tao et al |
| 49 | Shanghai/SH0093/2020 | 2020-01-21 | Shanghai Public Health Clinical Center, Shanghai Medical College, Fudan University | National Research Center for Translational Medicine (Shanghai), Ruijin Hospital affiliated to Shanghai Jiao Tong University School of Medicine & Shanghai Public Health Clinical Center | Shengyue Wang et al |
| 50 | Hangzhou/HZCDC0048/2020 | 2020-01-21 | Hangzhou Center for Diseases Control and Prevention | Hangzhou Center for Diseases Control and Prevention | Jun Li et al |
| 51 | Hangzhou/HZCDC0090L/2020 | 2020-01-21 | Hangzhou Center for Diseases Control and Prevention | Hangzhou Center for Diseases Control and Prevention | Jun Li et al |
| 52 | HongKong/VB20017970/2020 | 2020-01-21 | Prince of Wales Hospital | Hong Kong Department of Health | Alan K.L. Tsang et al |
| 53 | Hangzhou/HZ48/2020 | 2020-01-21 | Hangzhou Center for Disease Control and Prevention | Inspection Center of Hangzhou Center for Disease Control and Prevention | Yu hua et al |
| 54 | Guangdong/20SF117/2020 | 2020-01-21 | Guangdong Provincial Center for Diseases Control and Prevention;Guangdong Provincial Institute of Public Health | School of Public Health, The University of Hong Kong | Bosheng Li et al |
| 55 | Chongqing/YC01/2020 | 2020-01-21 | Yongchuan District Center for Disease Control and Prevention | Chongqing Municipal Center for Disease Control and Prevention | Ye Sheng et al |
| 56 | Hangzhou/HZCDC0119/2020 | 2020-01-22 | Hangzhou Center for Diseases | Hangzhou Center for Diseases | Jun Li et al |

| | | | | | |
|---|---|---|---|---|---|
| | | | Control and Prevention | Control and Prevention | |
| 57 | Thailand/NIH-162/2020 | 2020-01-22 | Bamrasnaradura hospital | National Institute of Health. Department of medical Sciences, Ministry of Public Health, Thailand | Pilailuk et al |
| 58 | Australia/NSW02/2020 | 2020-01-22 | Centre for Infectious Diseases and Microbiology Laboratory Services | NSW Health Pathology - Institute of Clinical Pathology and Medical Research; Westmead Hospital; University of Sydney | Rockett R et al |
| 59 | USA/CA2/2020 | 2020-01-22 | California Department of Public Health | Pathogen Discovery, Respiratory Viruses Branch, Division of Viral Diseases, Centers for Disease Control and Prevention | Anna Uehara et al |
| 60 | USA/AZ1/2020 | 2020-01-22 | Arizona Department of Health Services | Pathogen Discovery, Respiratory Viruses Branch, Division of Viral Diseases, Centers for Disease Control and Prevention | Ying Tao et al |
| 61 | Vietnam/19-01S/2020 | 2020-01-22 | | Microbiology and Immunology department | Cao et al |
| 62 | Vietnam/19-02S/2020 | 2020-01-22 | | Microbiology and Immunology department | Nguyen et al |
| 63 | Fujian/13/2020 | 2020-01-22 | Fujian Center for Disease Control and Prevention | Fujian Center for Disease Control and Prevention | Chen Wei et al |
| 64 | Guangdong/20SF174/2020 | 2020-01-22 | Guangdong Provincial Center for Diseases Control and Prevention; Guangdong Provinical Public Health | Guangdong Provincial Center for Disease Control and Prevention | Min Kang et al |
| 65 | Jiujiang/JX22/2020 | 2020-01-22 | Jiangxi Province Center for Disease Control and Prevention | Jiangxi Province Center for Disease Control and Prevention | JianXiong Li et al |
| 66 | Foshan/20SF211/2020 | 2020-01-22 | Guangdong Provincial Center for Diseases Control and Prevention; Guangdong Provinical Public Health | Guangdong Provincial Center for Diseases Control and Prevention | Min Kang et al |
| 67 | Hangzhou/ZJU-05/2020 | 2020-01-22 | State Key Laboratory for Diagnosis and Treatment of Infectious Diseases, National Clinical Research Center for Infectious Diseases, First Affiliated Hospital, Zhejiang University School of Medicine, Hangzhou, China. 310003 | State Key Laboratory for Diagnosis and Treatment of Infectious Diseases, National Clinical Research Center for Infectious Diseases, First Affiliated Hospital, Zhejiang University School of Medicine, Hangzhou, China. 310003 | Hangping Yao et al |
| 68 | Hangzhou/HZCDC0135/2020 | 2020-01-22 | Hangzhou Center for Diseases Control and Prevention | Hangzhou Center for Diseases Control and Prevention | Jun Li et al |
| 69 | Foshan/20SF210/2020 | 2020-01-22 | Guangdong Provincial Center for Diseases Control and Prevention; Guangdong Provinical Public Health | Guangdong Provincial Center for Diseases Control and Prevention | Min Kang et al |
| 70 | Guangzhou/20SF206/2020 | 2020-01-22 | Guangdong Provincial Center for Diseases Control and Prevention; Guangdong Provinical Public Health | Guangdong Provincial Center for Diseases Control and Prevention | Min Kang et al |
| 71 | Hangzhou/HZ178/2020 | 2020-01-23 | Hangzhou Center for Disease Control and Prevention | Inspection Center of Hangzhou Center for Disease Control and Prevention | Yu hua et al |
| 72 | Taiwan/2/2020 | 2020-01-23 | Centers for Disease Control, R.O.C. (Taiwan) | Centers for Disease Control, R.O.C. (Taiwan) | Ji-Rong Yang et al |
| 73 | Nanchang/JX39/2020 | 2020-01-23 | Jiangxi Province Center for Disease Control and Prevention | Jiangxi Province Center for Disease Control and Prevention | JianXiong Li et al |
| 74 | Japan/OS-20-07-1/2020 | 2020-01-23 | Dept. of Pathology, National Institute of Infectious Diseases | Pathogen Genomics Center, National Institute of Infectious Diseases | Tsuyoshi Sekizuka et al |
| 75 | Canada/ON-VIDO-01/2020 | 2020-01-23 | Public Health Ontario Laboratory | National Microbiology Laboratory | Tyson et al |
| 76 | Singapore/1/2020 | 2020-01-23 | Singapore General Hospital | National Public Health Laboratory | Mak et al |
| 77 | France/IDF0372/2020 | 2020-01-23 | Department of Infectious and Tropical Diseases, Bichat | National Reference Center for Viruses of Respiratory | Mélanie Albert et al |

| | | | | | |
|---|---|---|---|---|---|
| | | | Claude Bernard Hospital, Paris | Infections, Institut Pasteur, Paris | |
| 78 | France/IDF0373/2020 | 2020-01-23 | Department of Infectious and Tropical Diseases, Bichat Claude Bernard Hospital, Paris | National Reference Center for Viruses of Respiratory Infections, Institut Pasteur, Paris | Mélanie Albert et al |
| 79 | Shandong/LY003/2020 | 2020-01-23 | Shandong Provincial Center for Disease Control and Prevention | Beijing Institute of Microbiology and Epidemiology | Jiang et al |
| 80 | Guangdong/20SF273/2020 | 2020-01-23 | Guangdong Provincial Center for Diseases Control and Prevention;Guangdong Provincial Institute of Public Health | School of Public Health, The University of Hong Kong | Bosheng Li et al |
| 81 | USA/CA1/2020 | 2020-01-23 | California Department of Public Health | Pathogen Discovery, Respiratory Viruses Branch, Division of Viral Diseases, Centers for Disease Control and Prevention | Anna Uehara et al |
| 82 | Chongqing/ZX01/2020 | 2020-01-23 | Zhongxian Center for Disease Control and Prevention | Chongqing Municipal Center for Disease Control and Prevention | Ye Sheng et al |
| 83 | Hangzhou/HZ162/2020 | 2020-01-23 | Hangzhou Center for Disease Control and Prevention | Inspection Center of Hangzhou Center for Disease Control and Prevention | Yu hua et al |
| 84 | Taiwan/3/2020 | 2020-01-24 | Taiwan Centers for Disease Control | Taiwan Centers for Disease Control | Ji-Rong Yang et al |
| 85 | Vietnam/VR03-38142/2020 | 2020-01-24 | National Influenza Center - National Institute of Hygiene and Epidemiology (NIHE) | National Influenza Center - National Institute of Hygiene and Epidemiology (NIHE) | Ung Thi Hong Trang et al |
| 86 | Vietnam/38142/2020 | 2020-01-24 | National Influenza Center, National Institute of Hygiene and Epidemiology (NIHE) | National Influenza Center, National Institute of Hygiene and Epidemiology (NIHE) | Le Quynh Mai et al |
| 87 | Australia/NSW01/2020 | 2020-01-24 | Centre for Infectious Diseases and Microbiology Laboratory Services | NSW Health Pathology - Institute of Clinical Pathology and Medical Research; Westmead Hospital; University of Sydney | Eden J-S et al |
| 88 | Australia/NSW04/2020 | 2020-01-24 | Centre for Infectious Diseases and Microbiology Laboratory Services | NSW Health Pathology - Institute of Clinical Pathology and Medical Research; Westmead Hospital; University of Sydney | Eden J-S et al |
| 89 | HongKong/HKU-903a/2020 | 2020-01-24 | | Microbiology | To et al |
| 90 | Beijing/BJ54/2020 | 2020-01-24 | Chinese PLA Institute for Disease Control and Prevention | Chinese PLA Institute for Disease Control and Prevention | Peng Li et al |
| 91 | Anhui/SZ005/2020 | 2020-01-24 | Department of microbiology laboratory,Anhui Provincial Center for Disease Control and Prevention | Department of microbiology laboratory,Anhui Provincial Center for Disease Control and Prevention | Weiwei Li et al |
| 92 | Malaysia/MKAK-CL-2020-5049/2020 | 2020-01-24 | National Public Health Laboratory | Malaysia Genome Institute | Mohd Noor Mat Isa et al |
| 93 | Zhejiang/HZ103/2020 | 2020-01-24 | Zhejiang Provincial Center for Disease Control and Prevention | Zhejiang Provincial Center for Disease Control and Prevention | Yanjun Zhang et al |
| 94 | Jiangsu/JS03/2020 | 2020-01-24 | NHC Key laboratory of Enteric Pathogenic Microbiology, Institute of Pathogenic Microbiology | Jiangsu Provincial Center for Disease Control & Prevention | Kangchen Zhao et al |
| 95 | Jiangsu/JS02/2020 | 2020-01-24 | NHC Key laboratory of Enteric Pathogenic Microbiology, Institute of Pathogenic Microbiology | Jiangsu Provincial Center for Disease Control & Prevention | Kangchen Zhao et al |
| 96 | Hangzhou/HZ477/2020 | 2020-01-24 | Hangzhou Center for Disease Control and Prevention | Inspection Center of Hangzhou Center for Disease Control and Prevention | Yu hua et al |
| 97 | Beijing/BJ53/2020 | 2020-01-24 | Chinese PLA Institute for Disease Control and Prevention | Chinese PLA Institute for Disease Control and Prevention | Peng Li et al |
| 98 | Beijing/Wuhan_IME-BJ01/2020 | 2020-01-24 | | Beijing Institute of Microbiology and Epidemiology | Fan et al |
| 99 | Malaysia/MKAK-CL-2020-5047/2020 | 2020-01-24 | National Public Health Laboratory | Malaysia Genome Institute | Mohd Noor Mat Isa et al |

| | | | | | |
|---|---|---|---|---|---|
| 100 | Hangzhou/ZJU-04/2020 | 2020-01-24 | State Key Laboratory for Diagnosis and Treatment of Infectious Diseases, National Clinical Research Center for Infectious Diseases, First Affiliated Hospital, Zhejiang University School of Medicine, Hangzhou, China 310003 | State Key Laboratory for Diagnosis and Treatment of Infectious Diseases, National Clinical Research Center for Infectious Diseases, First Affiliated Hospital, Zhejiang University School of Medicine, Hangzhou, China 310003 | Hangping Yao et al |
| 101 | HongKong/HKU-902b/2020 | 2020-01-24 | | | To et al |
| 102 | Canada/ON_PHL4181/2020 | 2020-01-25 | Public Health Ontario Laboratories | Public Health Ontario Laboratories | Alireza Eshaghi et al |
| 103 | Australia/NSW03/2020 | 2020-01-25 | Serology, Virology and OTDS Laboratories (SAViD), NSW Health Pathology Randwick | NSW Health Pathology - Institute of Clinical Pathology and Medical Research; Centre for Infectious Diseases and Microbiology Laboratory Services; Westmead Hospital; University of Sydney | Eden J-S et al |
| 104 | Australia/VIC03/2020 | 2020-01-25 | Victorian Infectious Diseases Reference Laboratory (VIDRL) | Victorian Infectious Diseases Reference Laboratory and Microbiological Diagnostic Unit Public Health Laboratory, Doherty Institute | Caly L. et al |
| 105 | Shanghai/SH0003/2020 | 2020-01-25 | Shanghai Public Health Clinical Center, Shanghai Medical College, Fudan University | National Research Center for Translational Medicine (Shanghai), Ruijin Hospital affiliated to Shanghai Jiao Tong University School of Medicine & Shanghai Public Health Clinical Center | Shengyue Wang et al |
| 106 | Hangzhou/HZ551/2020 | 2020-01-25 | Hangzhou Center for Disease Control and Prevention | Inspection Center of Hangzhou Center for Disease Control and Prevention | Yu hua et al |
| 107 | Singapore/2/2020 | 2020-01-25 | Singapore General Hospital | Programme in Emerging Infectious Diseases, Duke-NUS Medical School | Danielle E Anderson et al |
| 108 | SouthKorea/KCDC03/2020 | 2020-01-25 | Korea Centers for Disease Control & Prevention (KCDC) Center for Laboratory Control of Infectious Diseases Division of Viral Diseases | Korea Centers for Disease Control & Prevention (KCDC) Center for Laboratory Control of Infectious Diseases Division of Viral Diseases | Jeong-Min Kim et al |
| 109 | Taiwan/CGMH-CGU-01/2020 | 2020-01-25 | Laboratory Medicine | Department of Laboratory Medicine, Lin-Kou Chang Gung Memorial Hospital, Taoyuan, Taiwan. | Kuo-Chien Tsao et al |
| 110 | Australia/VIC01/2020 | 2020-01-25 | Monash Medical Centre | Collaboration between the University of Melbourne at The Peter Doherty Institute for Infection and Immunity, and the Victorian Infectious Disease Reference Laboratory | Caly et al |
| 111 | HongKong/HKPU6_2101/2020 | 2020-01-25 | Department of Pathology, Princess Margaret Hospital | Department of Health Technology and Informatics, Faculty of Health and Social Science, The Hong Kong Polytechnic University | Kenneth Siu-Sing LEUNG et al |
| 112 | Japan/NA-20-05-1/2020 | 2020-01-25 | Dept. of Pathology, National Institute of Infectious Diseases | Pathogen Genomics Center, National Institute of Infectious Diseases | Tsuyoshi Sekizuka et al |
| 113 | Hangzhou/ZJU-03/2020 | 2020-01-25 | State Key Laboratory for Diagnosis and Treatment of Infectious Diseases, National Clinical Research Center for Infectious Diseases, First Affiliated Hospital, Zhejiang University School of Medicine, Hangzhou, China 310003 | State Key Laboratory for Diagnosis and Treatment of Infectious Diseases, National Clinical Research Center for Infectious Diseases, First Affiliated Hospital, Zhejiang University School of Medicine, Hangzhou, China 310003 | Hangping Yao et al |
| 114 | Shandong/LY004/2020 | 2020-01-26 | Shandong Provincial Center for Disease Control and Prevention | Beijing Institute of Microbiology and Epidemiology | Jiang et al |
| 115 | Hangzhou/ZJU-02/2020 | 2020-01-26 | State Key Laboratory for | State Key Laboratory for | Hangping Yao et al |

| | | | | | |
|---|---|---|---|---|---|
| | | | Diagnosis and Treatment of Infectious Diseases, National Clinical Research Center for Infectious Diseases, First Affiliated Hospital, Zhejiang University School of Medicine, Hangzhou, China. 310003 | Diagnosis and Treatment of Infectious Diseases, National Clinical Research Center for Infectious Diseases, First Affiliated Hospital, Zhejiang University School of Medicine, Hangzhou, China. 310003 | |
| 116 | Xinyu/JX124/2020 | 2020-01-26 | Jiangxi Province Center for Disease Control and Prevention | Jiangxi Province Center for Disease Control and Prevention | JianXiong Li et al |
| 117 | Beijing/105/2020 | 2020-01-26 | | Infectious Disease Control Center | Li et al |
| 118 | Guangdong/20SF630/2020 | 2020-01-26 | Guangdong Provincial Center for Diseases Control and Prevention;Guangdong Provincial Institute of Public Health | School of Public Health, The University of Hong Kong | Bosheng Li et al |
| 119 | Jian/JX129/2020 | 2020-01-26 | Jiangxi Province Center for Disease Control and Prevention | Jiangxi Province Center for Disease Control and Prevention | JianXiong Li et al |
| 120 | Guangdong/20SF629/2020 | 2020-01-26 | Guangdong Provincial Center for Diseases Control and Prevention;Guangdong Provincial Institute of Public Health | School of Public Health, The University of Hong Kong | Bosheng Li et al |
| 121 | Guangdong/20SF684/2020 | 2020-01-27 | Guangdong Provincial Center for Diseases Control and Prevention;Guangdong Provincial Institute of Public Health | School of Public Health, The University of Hong Kong | Bosheng Li et al |
| 122 | Cambodia/0012/2020 | 2020-01-27 | Virology Unit, Institut Pasteur du Cambodge. | Virology Unit, Institut Pasteur du Cambodge (Sequencing done by: Jessica E Manning/Jennifer A Bohl at Malaria and Vector Research Research Laboratory, National Institute of Allergy and Infectious Diseases and Vida Ahyong from Chan-Zuckerberg Biohub) | Erik A Karlsson et al |
| 123 | India/1-27/2020 | 2020-01-27 | Indian Council of Medical Research - National Institute of Virology | National Influenza Center, Indian Council of Medical Research - National Institute of Virology | Potdar V et al |
| 124 | Singapore/15/2020 | 2020-01-27 | National Public Health Laboratory, National Centre for Infectious Diseases | National Public Health Laboratory, National Centre for Infectious Diseases | Mak TM et al |
| 125 | Nanchang/JX149/2020 | 2020-01-27 | Jiangxi Province Center for Disease Control and Prevention | Jiangxi Province Center for Disease Control and Prevention | JianXiong Li et al |
| 126 | HongKong/HKU-908a/2020 | 2020-01-27 | | Microbiology | To et al |
| 127 | Singapore/7/2020 | 2020-01-27 | National Public Health Laboratory, National Centre for Infectious Diseases | National Public Health Laboratory, National Centre for Infectious Diseases | Octavia S et al |
| 128 | USA/CA6/2020 | 2020-01-27 | California Department of Public Health | Pathogen Discovery, Respiratory Viruses Branch, Division of Viral Diseases, Centers for Disease Control and Prevention | Jing Zhang et al |
| 129 | Guangdong/20SF665/2020 | 2020-01-27 | Guangdong Provincial Center for Diseases Control and Prevention;Guangdong Provincial Institute of Public Health | School of Public Health, The University of Hong Kong | Bosheng Li et al |
| 130 | HongKong/HKU-907a/2020 | 2020-01-27 | | Microbiology | To et al |
| 131 | Guangzhou/GZMU0058/2020 | 2020-01-27 | The First Affiliated Hospital of Guangzhou Medical University | BGI-shenzhen & The First Affiliated Hospital of Guangzhou Medical University | et al |
| 132 | HongKong/HKU-903b/2020 | 2020-01-27 | | Microbiology | To et al |
| 133 | Beijing/235/2020 | 2020-01-28 | | Infectious Disease Control Center | Li et al |
| 134 | Germany/BavPat1/2020 | 2020-01-28 | Charité Universitätsmedizin Berlin, Institute of Virology; | Charité Universitätsmedizin Berlin, Institute of Virology | Victor M Corman et al |

| | | | | | |
|---|---|---|---|---|---|
| | | | Institut für Mikrobiologie der Bundeswehr, Munich | | |
| 135 | USA/IL2/2020 | 2020-01-28 | IL Department of Public Health Chicago Laboratory | Pathogen Discovery, Respiratory Viruses Branch, Division of Viral Diseases, Centers for Disease Control and Prevention | Yan Li et al |
| 136 | Australia/QLD01/2020 | 2020-01-28 | Pathology Queensland | Public Health Virology Laboratory | Ben Huang et al |
| 137 | Malaysia/IMR_WC085/2020 | 2020-01-28 | Institute for Medical Research, Infectious Disease Research Centre, National Institutes of Health, Ministry of Health Malaysia | Institute for Medical Research, Infectious Disease Research Centre, National Institutes of Health, Ministry of Health Malaysia | Suppiah.J et al |
| 138 | France/IDF0386-islP1/2020 | 2020-01-28 | Department of Infectious and Tropical Diseases, Bichat Claude Bernard Hospital, Paris | Laboratoire Virpath, CIRI U111, UCBL1, INSERM, CNRS, ENS Lyon | Olivier Terrier et al |
| 139 | Shanghai/SH0007/2020 | 2020-01-28 | Shanghai Public Health Clinical Center, Shanghai Medical College, Fudan University | National Research Center for Translational Medicine (Shanghai), Ruijin Hospital affiliated to Shanghai Jiao Tong University School of Medicine & Shanghai Public Health Clinical Center | Shengyue Wang et al |
| 140 | Taiwan/4/2020 | 2020-01-28 | Taiwan Centers for Disease Control | Taiwan Centers for Disease Control | Ji-Rong Yang et al |
| 141 | Shanghai/SH0005/2020 | 2020-01-28 | Shanghai Public Health Clinical Center, Shanghai Medical College, Fudan University | National Research Center for Translational Medicine (Shanghai), Ruijin Hospital affiliated to Shanghai Jiao Tong University School of Medicine & Shanghai Public Health Clinical Center | Shengyue Wang et al |
| 142 | Shanghai/SH0008/2020 | 2020-01-28 | Shanghai Public Health Clinical Center, Shanghai Medical College, Fudan University | National Research Center for Translational Medicine (Shanghai), Ruijin Hospital affiliated to Shanghai Jiao Tong University School of Medicine & Shanghai Public Health Clinical Center | Shengyue Wang et al |
| 143 | Beijing/Wuhan_IME-BJ03/2020 | 2020-01-28 | | Beijing Institute of Microbiology and Epidemiology | Fan et al |
| 144 | Nanchang/JX155/2020 | 2020-01-29 | Jiangxi Province Center for Disease Control and Prevention | Jiangxi Province Center for Disease Control and Prevention | JianXiong Li et al |
| 145 | Shanghai/SH0009/2020 | 2020-01-29 | Shanghai Public Health Clinical Center, Shanghai Medical College, Fudan University | National Research Center for Translational Medicine (Shanghai), Ruijin Hospital affiliated to Shanghai Jiao Tong University School of Medicine & Shanghai Public Health Clinical Center | Shengyue Wang et al |
| 146 | Beijing/BJ251/2020 | 2020-01-29 | Chinese PLA Institute for Disease Control and Prevention | Chinese PLA Institute for Disease Control and Prevention | Peng Li et al |
| 147 | USA/CA5/2020 | 2020-01-29 | California Department of Health | Pathogen Discovery, Respiratory Viruses Branch, Division of Viral Diseases, Centers for Disease Control and Prevention | Ying Tao et al |
| 148 | Guangzhou/GZMU0051/2020 | 2020-01-29 | The First Affiliated Hospital of Guangzhou Medical University | BGI-shenzhen & The First Affiliated Hospital of Guangzhou Medical University | Yanqun Wang et al |
| 149 | Nanchang/JX176/2020 | 2020-01-29 | Jiangxi Province Center for Disease Control and Prevention | Jiangxi Province Center for Disease Control and Prevention | JianXiong Li et al |
| 150 | Guangzhou/GZMU0052/2020 | 2020-01-29 | The First Affiliated Hospital of Guangzhou Medical University | BGI-shenzhen & The First Affiliated Hospital of Guangzhou Medical University | Yanqun Wang et al |
| 151 | Beijing/BJ288/2020 | 2020-01-29 | Chinese PLA Institute for Disease Control and Prevention | Chinese PLA Institute for Disease Control and Prevention | Peng Li et al |
| 152 | Japan/TY-WK-012/2020 | 2020-01-29 | Dept. of Virology III, National Institute of Infectious Diseases | Pathogen Genomics Center, National Institute of Infectious | Tsuyoshi Sekizuka et al |

| | | | | | |
|---|---|---|---|---|---|
| | | | | Diseases | |
| 153 | Singapore/3Clin/2020 | 2020-01-29 | National Public Health Laboratory, National Centre for Infectious Diseases | National Public Health Laboratory, National Centre for Infectious Diseases | Mak TM et al |
| 154 | France/IDF0515/2020 | 2020-01-29 | Department of Infectious and Tropical Diseases, Bichat Claude Bernard Hospital, Paris | National Reference Center for Viruses of Respiratory Infections, Institut Pasteur, Paris | Mélanie Albert et al |
| 155 | Italy/INMI1-cs/2020 | 2020-01-29 | INMI Lazzaro Spallanzani IRCCS | Laboratory of Virology, INMI Lazzaro Spallanzani IRCCS | Maria R. Capobianchi et al |
| 156 | Shanghai/SH0011/2020 | 2020-01-29 | Shanghai Public Health Clinical Center, Shanghai Medical College, Fudan University | National Research Center for Translational Medicine (Shanghai), Ruijin Hospital affiliated to Shanghai Jiao Tong University School of Medicine & Shanghai Public Health Clinical Center | Shengyue Wang et al |
| 157 | Guangdong/20SF1107/2020 | 2020-01-29 | Guangdong Provincial Center for Diseases Control and Prevention;Guangdong Provincial Institute of Public Health | School of Public Health, The University of Hong Kong | Bosheng Li et al |
| 158 | HongKong/HKU-907b/2020 | 2020-01-29 | | Microbiology | To et al |
| 159 | Guangzhou/GZMU0055/2020 | 2020-01-29 | The First Affiliated Hospital of Guangzhou Medical University | BGI-shenzhen & The First Affiliated Hospital of Guangzhou Medical University | et al |
| 160 | Nanchang/JX174/2020 | 2020-01-29 | Jiangxi Province Center for Disease Control and Prevention | Jiangxi Province Center for Disease Control and Prevention | JianXiong Li et al |
| 161 | Guangdong/20SF840/2020 | 2020-01-29 | Guangdong Provincial Center for Diseases Control and Prevention;Guangdong Provincial Institute of Public Health | School of Public Health, The University of Hong Kong | Bosheng Li et al |
| 162 | Shanghai/SH0041/2020 | 2020-01-29 | Shanghai Public Health Clinical Center, Shanghai Medical College, Fudan University | National Research Center for Translational Medicine (Shanghai), Ruijin Hospital affiliated to Shanghai Jiao Tong University School of Medicine & Shanghai Public Health Clinical Center | Shengyue Wang et al |
| 163 | Italy/SPL1/2020 | 2020-01-29 | Department of Infectious Diseases, Istituto Superiore di Sanità, Rome, Italy | Virology Laboratory, Scientific Department, Army Medical Center | Paola Stefanelli et al |
| 164 | England/02/2020 | 2020-01-29 | Respiratory Virus Unit, Microbiology Services Colindale, Public Health England | Respiratory Virus Unit, Microbiology Services Colindale, Public Health England | Monica Galiano et al |
| 165 | Beijing/Wuhan_IME-BJ07/2020 | 2020-01-29 | | Beijing Institute of Microbiology and Epidemiology | Fan et al |
| 166 | China/IQTC02/2020 | 2020-01-29 | | Technology Centre, Guangzhou Customs | Shi et al |
| 167 | HongKong/HKU-908b/2020 | 2020-01-29 | | Microbiology | To et al |
| 168 | England/01/2020 | 2020-01-29 | Respiratory Virus Unit, Microbiology Services Colindale, Public Health England | Respiratory Virus Unit, Microbiology Services Colindale, Public Health England | Monica Galiano et al |
| 169 | USA/CA4/2020 | 2020-01-29 | California Department of Health | Pathogen Discovery, Respiratory Viruses Branch, Division of Viral Diseases, Centers for Disease Control and Prevention | Krista Queen et al |
| 170 | Singapore/20/2020 | 2020-01-29 | National Public Health Laboratory, National Centre for Infectious Diseases | National Public Health Laboratory, National Centre for Infectious Diseases | Mak TM et al |
| 171 | Beijing/BJ287/2020 | 2020-01-29 | Chinese PLA Institute for Disease Control and Prevention | Chinese PLA Institute for Disease Control and Prevention | Peng Li et al |
| 172 | USA/CA3/2020 | 2020-01-29 | California Department of Health | Pathogen Discovery, Respiratory Viruses Branch, Division of Viral Diseases, Centers for Disease Control and Prevention | Krista Queen et al |
| 173 | Japan/KY-V-029/2020 | 2020-01-29 | Dept. of Virology III, National | Pathogen Genomics Center, | Tsuyoshi Sekizuka et |

| | | | | | |
|---|---|---|---|---|---|
| | | | Institute of Infectious Diseases | National Institute of Infectious Diseases | al |
| 174 | UnitedArabEmirates/L5630/2020 | 2020-01-29 | Mohammed Bin Rashid University of Medicine and Health Sciences | Al Jalila Genomics Center | Ahmad Abou Tayoun et al |
| 175 | Korea/KCDC2002/2020 | 2020-01-30 | Division of Viral Diseases, Center for Laboratory Control of Infectious Diseases, Korea Centers for Diseases Control and Prevention | Division of Viral Diseases, Center for Laboratory Control of Infectious Diseases, Korea Centers for Diseases Control and Prevention | Jeong-Min Kim et al |
| 176 | Australia/QLD02/2020 | 2020-01-30 | Pathology Queensland | Public Health Virology Laboratory | Ben Huang et al |
| 177 | Shanghai/SH0059/2020 | 2020-01-30 | Shanghai Public Health Clinical Center, Shanghai Medical College, Fudan University | National Research Center for Translational Medicine (Shanghai), Ruijin Hospital affiliated to Shanghai Jiao Tong University School of Medicine & Shanghai Public Health Clinical Center | Shengyue Wang et al |
| 178 | Guangdong/2020XN4273-P0036/2020 | 2020-01-30 | Guangdong Provincial Institution of Public Health, Guangdong Provinical Center for Disease Control and Prevention | Guangdong Provincial Institution of Public Health | Jing Lu et al |
| 179 | Shanghai/SH0058/2020 | 2020-01-30 | Shanghai Public Health Clinical Center, Shanghai Medical College, Fudan University | National Research Center for Translational Medicine (Shanghai), Ruijin Hospital affiliated to Shanghai Jiao Tong University School of Medicine & Shanghai Public Health Clinical Center | Shengyue Wang et al |
| 180 | SouthKorea/KCDC05/2020 | 2020-01-30 | Division of Viral Diseases, Center for Laboratory Control of Infectious Diseases, Korea Centers for Diseases Control and Prevention | Division of Viral Diseases, Center for Laboratory Control of Infectious Diseases, Korea Centers for Diseases Control and Prevention | Jeong-Min Kim et al |
| 181 | SouthKorea/KCDC06/2020 | 2020-01-30 | Division of Viral Diseases, Center for Laboratory Control of Infectious Diseases, Korea Centers for Diseases Control and Prevention | Division of Viral Diseases, Center for Laboratory Control of Infectious Diseases, Korea Centers for Diseases Control and Prevention | Jeong-Min Kim et al |
| 182 | Shanghai/SH0013/2020 | 2020-01-30 | Shanghai Public Health Clinical Center, Shanghai Medical College, Fudan University | National Research Center for Translational Medicine (Shanghai), Ruijin Hospital affiliated to Shanghai Jiao Tong University School of Medicine & Shanghai Public Health Clinical Center | Shengyue Wang et al |
| 183 | HongKong/VB20024950/2020 | 2020-01-30 | Hong Kong Department of Health | The University of Hong Kong | Dominic N.C. Tsang et al |
| 184 | Shanghai/SH0022/2020 | 2020-01-30 | Shanghai Public Health Clinical Center, Shanghai Medical College, Fudan University | National Research Center for Translational Medicine (Shanghai), Ruijin Hospital affiliated to Shanghai Jiao Tong University School of Medicine & Shanghai Public Health Clinical Center | Shengyue Wang et al |
| 185 | Guangdong/2020XN4433-P0040/2020 | 2020-01-30 | Guangdong Provincial Institution of Public Health, Guangdong Provinical Center for Disease Control and Prevention | Guangdong Provincial Institution of Public Health | Jing Lu et al |
| 186 | Shandong/LY008/2020 | 2020-01-30 | Shandong Provincial Center for Disease Control and Prevention | Beijing Institute of Microbiology and Epidemiology | Jiang et al |
| 187 | Guangdong/2020XN4475-P0042/2020 | 2020-01-30 | Guangdong Provincial Institution of Public Health, Guangdong Provinical Center for Disease Control and Prevention | Guangdong Provincial Institution of Public Health | Jing Lu et al |
| 188 | Shanghai/SH0023/2020 | 2020-01-30 | Shanghai Public Health Clinical Center, Shanghai | National Research Center for Translational Medicine (Shanghai), Ruijin Hospital | Shengyue Wang et al |

| | | | | | |
|---|---|---|---|---|---|
| | | | Medical College, Fudan University | affiliated to Shanghai Jiao Tong University School of Medicine & Shanghai Public Health Clinical Center | |
| 189 | Guangdong/20SF1152/2020 | 2020-01-30 | Guangdong Provincial Center for Diseases Control and Prevention;Guangdong Provincial Institute of Public Health | School of Public Health, The University of Hong Kong | Bosheng Li et al |
| 190 | Guangdong/20SF1136/2020 | 2020-01-30 | Guangdong Provincial Center for Diseases Control and Prevention;Guangdong Provincial Institute of Public Health | School of Public Health, The University of Hong Kong | Bosheng Li et al |
| 191 | Shanghai/SH0024/2020 | 2020-01-30 | Shanghai Public Health Clinical Center, Shanghai Medical College, Fudan University | National Research Center for Translational Medicine (Shanghai), Ruijin Hospital affiliated to Shanghai Jiao Tong University School of Medicine & Shanghai Public Health Clinical Center | Shengyue Wang et al |
| 192 | Shanghai/SH0043/2020 | 2020-01-30 | Shanghai Public Health Clinical Center, Shanghai Medical College, Fudan University | National Research Center for Translational Medicine (Shanghai), Ruijin Hospital affiliated to Shanghai Jiao Tong University School of Medicine & Shanghai Public Health Clinical Center | Shengyue Wang et al |
| 193 | Shanghai/SH0094/2020 | 2020-01-31 | Shanghai Public Health Clinical Center, Shanghai Medical College, Fudan University | National Research Center for Translational Medicine (Shanghai), Ruijin Hospital affiliated to Shanghai Jiao Tong University School of Medicine & Shanghai Public Health Clinical Center | Shengyue Wang et al |
| 194 | Taiwan/8/2020 | 2020-01-31 | Centers for Disease Control, R.O.C. (Taiwan) | Centers for Disease Control, R.O.C. (Taiwan) | Ji-Rong Yang et al |
| 195 | Australia/VIC13/2020 | 2020-01-31 | Victorian Infectious Diseases Reference Laboratory (VIDRL) | Victorian Infectious Diseases Reference Laboratory and Microbiological Diagnostic Unit Public Health Laboratory, Doherty Institute | Caly L. et al |
| 196 | Taiwan/5/2020 | 2020-01-31 | Centers for Disease Control, R.O.C. (Taiwan) | Centers for Disease Control, R.O.C. (Taiwan) | Ji-Rong Yang et al |
| 197 | Japan/TY-WK-501/2020 | 2020-01-31 | Dept. of Virology III, National Institute of Infectious Diseases | Pathogen Genomics Center, National Institute of Infectious Diseases | Tsuyoshi Sekizuka et al |
| 198 | USA/WI1/2020 | 2020-01-31 | Wisconsin Department of Health Services | Pathogen Discovery, Respiratory Viruses Branch, Division of Viral Diseases, Centers for Disease Control and Prevention | Jing Zhang et al |
| 199 | Guangdong/2020XN4448-P0002/2020 | 2020-01-31 | Guangdong Provincial Institution of Public Health, Guangdong Provinical Center for Disease Control and Prevention | Guangdong Provincial Institution of Public Health | Jing Lu et al |
| 200 | Japan/TY-WK-521/2020 | 2020-01-31 | Dept. of Virology III, National Institute of Infectious Diseases | Pathogen Genomics Center, National Institute of Infectious Diseases | Tsuyoshi Sekizuka et al |
| 201 | SouthKorea/KCDC07/2020 | 2020-01-31 | Division of Viral Diseases, Center for Laboratory Control of Infectious Diseases, Korea Centers for Diseases Control and Prevention | Division of Viral Diseases, Center for Laboratory Control of Infectious Diseases, Korea Centers for Diseases Control and Prevention | Jeong-Min Kim et al |
| 202 | Taiwan/NTU01/2020 | 2020-01-31 | Department of Laboratory Medicine, National Taiwan University Hospital | Microbial Genomics Core Lab, National Taiwan University Centers of Genomic and Precision Medicine | Shiou-Hwei Yeh et al |
| 203 | Guangdong/GD2020087-P0008/2020 | 2020-02-01 | Guangdong Provincial Institution of Public Health, Guangdong Provinical Center for Disease Control and Prevention | Guangdong Provincial Institution of Public Health | Jing Lu et al |

| | | | | | |
|---|---|---|---|---|---|
| 204 | Shanghai/SH0027/2020 | 2020-02-01 | Shanghai Public Health Clinical Center, Shanghai Medical College, Fudan University | National Research Center for Translational Medicine (Shanghai), Ruijin Hospital affiliated to Shanghai Jiao Tong University School of Medicine & Shanghai Public Health Clinical Center | Shengyue Wang et al |
| 205 | Guangzhou/GZMU0035/2020 | 2020-02-01 | The First Affiliated Hospital of Guangzhou Medical University | BGI-shenzhen & The First Affiliated Hospital of Guangzhou Medical University | Yanqun Wang et al |
| 206 | Shanghai/SH0079/2020 | 2020-02-01 | Shanghai Public Health Clinical Center, Shanghai Medical College, Fudan University | National Research Center for Translational Medicine (Shanghai), Ruijin Hospital affiliated to Shanghai Jiao Tong University School of Medicine & Shanghai Public Health Clinical Center | Shengyue Wang et al |
| 207 | Japan/UT-NCGM02/2020 | 2020-02-01 | University of Wisconsin - Madison: Influenza Research Institute | University of Wisconsin Madison, AIDS Vaccine Research Laboratories | Katarina Braun et al |
| 208 | SouthKorea/KCDC12/2020 | 2020-02-01 | Division of Viral Diseases, Center for Laboratory Control of Infectious Diseases, Korea Centers for Diseases Control and Prevention | Division of Viral Diseases, Center for Laboratory Control of Infectious Diseases, Korea Centers for Diseases Control and Prevention | Jeong-Min Kim et al |
| 209 | Singapore/3/2020 | 2020-02-01 | National Centre for Infectious Diseases | Programme in Emerging Infectious Diseases, Duke-NUS Medical School | Danielle E Anderson et al |
| 210 | HongKong/VB20026565/2020 | 2020-02-01 | Hong Kong Department of Health | School of Public Health, The University of Hon g Kong | Dominic N.C. Tsang et al |
| 211 | Shanghai/SH0088/2020 | 2020-02-01 | Shanghai Public Health Clinical Center, Shanghai Medical College, Fudan University | National Research Center for Translational Medicine (Shanghai), Ruijin Hospital affiliated to Shanghai Jiao Tong University School of Medicine & Shanghai Public Health Clinical Center | Shengyue Wang et al |
| 212 | Shanghai/SH0053/2020 | 2020-02-01 | Shanghai Public Health Clinical Center, Shanghai Medical College, Fudan University | National Research Center for Translational Medicine (Shanghai), Ruijin Hospital affiliated to Shanghai Jiao Tong University School of Medicine & Shanghai Public Health Clinical Center | Shengyue Wang et al |
| 213 | Guangdong/GD2020080-P0010/2020 | 2020-02-01 | Guangdong Provincial Institution of Public Health, Guangdong Provinical Center for Disease Control and Prevention | Guangdong Provincial Institution of Public Health | Jing Lu et al |
| 214 | Shanghai/SH0032/2020 | 2020-02-01 | Shanghai Public Health Clinical Center, Shanghai Medical College, Fudan University | National Research Center for Translational Medicine (Shanghai), Ruijin Hospital affiliated to Shanghai Jiao Tong University School of Medicine & Shanghai Public Health Clinical Center | Shengyue Wang et al |
| 215 | Lishui/LS557/2020 | 2020-02-01 | Centers for Disease Control and Prevention of Lishui | Department of Inspection， Centers for Disease Control and Prevention of Lishui | Wang Xiaoguang et al |
| 216 | Guangzhou/GZMU0034/2020 | 2020-02-01 | The First Affiliated Hospital of Guangzhou Medical University | BGI-shenzhen & The First Affiliated Hospital of Guangzhou Medical University | et al |
| 217 | Shanghai/SH0076/2020 | 2020-02-01 | Shanghai Public Health Clinical Center, Shanghai Medical College, Fudan University | National Research Center for Translational Medicine (Shanghai), Ruijin Hospital affiliated to Shanghai Jiao Tong University School of Medicine & Shanghai Public Health Clinical Center | Shengyue Wang et al |
| 218 | Guangdong/GD2020086-P0021/2020 | 2020-02-01 | Guangdong Provincial Institution of Public Health, Guangdong Provinical Center | Guangdong Provincial Institution of Public Health | Jing Lu et al |

| | | | | | |
|---|---|---|---|---|---|
| | | | for Disease Control and Prevention | | |
| 219 | Shanghai/SH0060/2020 | 2020-02-01 | Shanghai Public Health Clinical Center, Shanghai Medical College, Fudan University | National Research Center for Translational Medicine (Shanghai), Ruijin Hospital affiliated to Shanghai Jiao Tong University School of Medicine & Shanghai Public Health Clinical Center | Shengyue Wang et al |
| 220 | Singapore/11/2020 | 2020-02-02 | National Public Health Laboratory | National Public Health Laboratory | Octavia S et al |
| 221 | Korea/KCDC2004/2020 | 2020-02-02 | Division of Viral Diseases, Center for Laboratory Control of Infectious Diseases, Korea Centers for Diseases Control and Prevention | Division of Viral Diseases, Center for Laboratory Control of Infectious Diseases, Korea Centers for Diseases Control and Prevention | Jeong-Min Kim et al |
| 222 | Korea/KCDC2006/2020 | 2020-02-02 | Division of Viral Diseases, Center for Laboratory Control of Infectious Diseases, Korea Centers for Diseases Control and Prevention | Division of Viral Diseases, Center for Laboratory Control of Infectious Diseases, Korea Centers for Diseases Control and Prevention | Jeong-Min Kim et al |
| 223 | Shanghai/SH0107/2020 | 2020-02-02 | Shanghai Public Health Clinical Center, Shanghai Medical College, Fudan University | National Research Center for Translational Medicine (Shanghai), Ruijin Hospital affiliated to Shanghai Jiao Tong University School of Medicine & Shanghai Public Health Clinical Center | Shengyue Wang et al |
| 224 | Shanghai/SH01/2020 | 2020-02-02 | | Key Laboratory of Medical Molecular Virology (MOE/NHC/CAMS) | Zhang et al |
| 225 | Shanghai/SH0121/2020 | 2020-02-02 | Shanghai Public Health Clinical Center, Shanghai Medical College, Fudan University | National Research Center for Translational Medicine (Shanghai), Ruijin Hospital affiliated to Shanghai Jiao Tong University School of Medicine & Shanghai Public Health Clinical Center | Shengyue Wang et al |
| 226 | Shanghai/SH0114/2020 | 2020-02-02 | Shanghai Public Health Clinical Center, Shanghai Medical College, Fudan University | National Research Center for Translational Medicine (Shanghai), Ruijin Hospital affiliated to Shanghai Jiao Tong University School of Medicine & Shanghai Public Health Clinical Center | Shengyue Wang et al |
| 227 | Shanghai/SH0026/2020 | 2020-02-02 | Shanghai Public Health Clinical Center, Shanghai Medical College, Fudan University | National Research Center for Translational Medicine (Shanghai), Ruijin Hospital affiliated to Shanghai Jiao Tong University School of Medicine & Shanghai Public Health Clinical Center | Shengyue Wang et al |
| 228 | Shanghai/SH0128/2020 | 2020-02-02 | Shanghai Public Health Clinical Center, Shanghai Medical College, Fudan University | National Research Center for Translational Medicine (Shanghai), Ruijin Hospital affiliated to Shanghai Jiao Tong University School of Medicine & Shanghai Public Health Clinical Center | Shengyue Wang et al |
| 229 | Shanghai/SH0031/2020 | 2020-02-02 | Shanghai Public Health Clinical Center, Shanghai Medical College, Fudan University | National Research Center for Translational Medicine (Shanghai), Ruijin Hospital affiliated to Shanghai Jiao Tong University School of Medicine & Shanghai Public Health Clinical Center | Shengyue Wang et al |
| 230 | Shanghai/SH0117/2020 | 2020-02-02 | Shanghai Public Health Clinical Center, Shanghai Medical College, Fudan University | National Research Center for Translational Medicine (Shanghai), Ruijin Hospital affiliated to Shanghai Jiao Tong University School of Medicine & Shanghai Public Health Clinical Center | Shengyue Wang et al |
| 231 | Shanghai/SH0012/2020 | 2020-02-02 | Shanghai Public Health | National Research Center for | Shengyue Wang et al |

| | | | | | |
|---|---|---|---|---|---|
| | | | Clinical Center, Shanghai Medical College, Fudan University | Translational Medicine (Shanghai), Ruijin Hospital affiliated to Shanghai Jiao Tong University School of Medicine & Shanghai Public Health Clinical Center | |
| 232 | Shangrao/JX1215/2020 | 2020-02-02 | Jiangxi Province Center for Disease Control and Prevention | Jiangxi Province Center for Disease Control and Prevention | JianXiong Li et al |
| 233 | Shanghai/SH0021/2020 | 2020-02-02 | Shanghai Public Health Clinical Center, Shanghai Medical College, Fudan University | National Research Center for Translational Medicine (Shanghai), Ruijin Hospital affiliated to Shanghai Jiao Tong University School of Medicine & Shanghai Public Health Clinical Center | Shengyue Wang et al |
| 234 | Shanghai/SH0054/2020 | 2020-02-02 | Shanghai Public Health Clinical Center, Shanghai Medical College, Fudan University | National Research Center for Translational Medicine (Shanghai), Ruijin Hospital affiliated to Shanghai Jiao Tong University School of Medicine & Shanghai Public Health Clinical Center | Shengyue Wang et al |
| 235 | Beijing/BJ492/2020 | 2020-02-02 | Chinese PLA Institute for Disease Control and Prevention | Chinese PLA Institute for Disease Control and Prevention | Peng Li et al |
| 236 | Shanghai/SH0109/2020 | 2020-02-02 | Shanghai Public Health Clinical Center, Shanghai Medical College, Fudan University | National Research Center for Translational Medicine (Shanghai), Ruijin Hospital affiliated to Shanghai Jiao Tong University School of Medicine & Shanghai Public Health Clinical Center | Shengyue Wang et al |
| 237 | Hangzhou/ZJU-06/2020 | 2020-02-02 | State Key Laboratory for Diagnosis and Treatment of Infectious Diseases, National Clinical Research Center for Infectious Diseases, First Affiliated Hospital, Zhejiang University School of Medicine, Hangzhou, China 310003 | State Key Laboratory for Diagnosis and Treatment of Infectious Diseases, National Clinical Research Center for Infectious Diseases, First Affiliated Hospital, Zhejiang University School of Medicine, Hangzhou, China 310003 | Hangping Yao et al |
| 238 | Shangrao/JX1178/2020 | 2020-02-03 | Jiangxi Province Center for Disease Control and Prevention | Jiangxi Province Center for Disease Control and Prevention | JianXiong Li et al |
| 239 | Singapore/8/2020 | 2020-02-03 | National Public Health Laboratory, National Centre for Infectious Diseases | National Public Health Laboratory, National Centre for Infectious Diseases | Octavia S et al |
| 240 | Belgium/GHB-03021/2020 | 2020-02-03 | KU Leuven, Clinical and Epidemiological Virology | KU Leuven, Clinical and Epidemiological Virology | Bert Vanmechelen et al |
| 241 | Singapore/4/2020 | 2020-02-03 | National Centre for Infectious Diseases | Programme in Emerging Infectious Diseases, Duke-NUS Medical School | Danielle E Anderson et al |
| 242 | Shangrao/JX1177/2020 | 2020-02-03 | Jiangxi Province Center for Disease Control and Prevention | Jiangxi Province Center for Disease Control and Prevention | JianXiong Li et al |
| 243 | Shangrao/JX1176/2020 | 2020-02-03 | Jiangxi Province Center for Disease Control and Prevention | Jiangxi Province Center for Disease Control and Prevention | JianXiong Li et al |
| 244 | Shanghai/SH0030/2020 | 2020-02-03 | Shanghai Public Health Clinical Center, Shanghai Medical College, Fudan University | National Research Center for Translational Medicine (Shanghai), Ruijin Hospital affiliated to Shanghai Jiao Tong University School of Medicine & Shanghai Public Health Clinical Center | Shengyue Wang et al |
| 245 | Hangzhou/ZJU-010/2020 | 2020-02-03 | State Key Laboratory for Diagnosis and Treatment of Infectious Diseases, National Clinical Research Center for Infectious Diseases, First Affiliated Hospital, Zhejiang University School of | State Key Laboratory for Diagnosis and Treatment of Infectious Diseases, National Clinical Research Center for Infectious Diseases, First Affiliated Hospital, Zhejiang | Hangping Yao et al |

| | | | | | |
|---|---|---|---|---|---|
| | | | Medicine, Hangzhou, China 310003 | University School of Medicine, Hangzhou, China 310003 | |
| 246 | Hangzhou/ZJU-07/2020 | 2020-02-03 | State Key Laboratory for Diagnosis and Treatment of Infectious Diseases, National Clinical Research Center for Infectious Diseases, First Affiliated Hospital, Zhejiang University School of Medicine, Hangzhou, China 310003 | State Key Laboratory for Diagnosis and Treatment of Infectious Diseases, National Clinical Research Center for Infectious Diseases, First Affiliated Hospital, Zhejiang University School of Medicine, Hangzhou, China 310003 | Hangping Yao et al |
| 247 | Malaysia/MKAK-CL-2020-6430/2020 | 2020-02-04 | National Public Health Laboratory | Malaysia Genome Institute | Mohd Noor Mat Isa et al |
| 248 | Shanghai/SH0047/2020 | 2020-02-04 | Shanghai Public Health Clinical Center, Shanghai Medical College, Fudan University | National Research Center for Translational Medicine (Shanghai), Ruijin Hospital affiliated to Shanghai Jiao Tong University School of Medicine & Shanghai Public Health Clinical Center | Shengyue Wang et al |
| 249 | Hangzhou/ZJU-011/2020 | 2020-02-04 | State Key Laboratory for Diagnosis and Treatment of Infectious Diseases, National Clinical Research Center for Infectious Diseases, First Affiliated Hospital, Zhejiang University School of Medicine, Hangzhou, China 310003 | State Key Laboratory for Diagnosis and Treatment of Infectious Diseases, National Clinical Research Center for Infectious Diseases, First Affiliated Hospital, Zhejiang University School of Medicine, Hangzhou, China 310003 | Hangping Yao et al |
| 250 | Shanghai/SH0033/2020 | 2020-02-04 | Shanghai Public Health Clinical Center, Shanghai Medical College, Fudan University | National Research Center for Translational Medicine (Shanghai), Ruijin Hospital affiliated to Shanghai Jiao Tong University School of Medicine & Shanghai Public Health Clinical Center | Shengyue Wang et al |
| 251 | Singapore/10/2020 | 2020-02-04 | National Public Health Laboratory, National Centre for Infectious Diseases | National Centre for Infectious Diseases, National Centre for Infectious Diseases | Octavia S et al |
| 252 | Taiwan/CGMH-CGU-02/2020 | 2020-02-04 | Laboratory Medicine | Department of Laboratory Medicine, Lin-Kou Chang Gung Memorial Hospital, Taoyuan, Taiwan | Kuo-Chien Tsao et al |
| 253 | Korea/KCDC2005/2020 | 2020-02-04 | Division of Viral Diseases, Center for Laboratory Control of Infectious Diseases, Korea Centers for Diseases Control and Prevention | Division of Viral Diseases, Center for Laboratory Control of Infectious Diseases, Korea Centers for Diseases Control and Prevention | Jeong-Min Kim et al |
| 254 | Singapore/9/2020 | 2020-02-04 | National Public Health Laboratory, National Centre for Infectious Diseases | National Public Health Laboratory, National Centre for Infectious Diseases | Octavia S et al |
| 255 | Shanghai/SH0044/2020 | 2020-02-04 | Shanghai Public Health Clinical Center, Shanghai Medical College, Fudan University | National Research Center for Translational Medicine (Shanghai), Ruijin Hospital affiliated to Shanghai Jiao Tong University School of Medicine & Shanghai Public Health Clinical Center | Shengyue Wang et al |
| 256 | Shanghai/SH0028/2020 | 2020-02-04 | Shanghai Public Health Clinical Center, Shanghai Medical College, Fudan University | National Research Center for Translational Medicine (Shanghai), Ruijin Hospital affiliated to Shanghai Jiao Tong University School of Medicine & Shanghai Public Health Clinical Center | Shengyue Wang et al |
| 257 | Guangzhou/GZMU0020/2020 | 2020-02-05 | The First Affiliated Hospital of Guangzhou Medical University | BGI-shenzhen & The First Affiliated Hospital of Guangzhou Medical University | et al |
| 258 | Taiwan/NTU02/2020 | 2020-02-05 | Department of Laboratory Medicine, National Taiwan University Hospital | Microbial Genomics Core Lab, National Taiwan University Centers of Genomic and Precision Medicine | Shiou-Hwei Yeh et al |
| 259 | Australia/QLD04/2020 | 2020-02-05 | Pathology Queensland | Public Health Virology | Ben Huang et al |

| | | | | | |
|---|---|---|---|---|---|
| | | | | Laboratory | |
| 260 | Australia/QLD03/2020 | 2020-02-05 | Pathology Queensland | Public Health Virology Laboratory | Ben Huang et al |
| 261 | Shangrao/JX1948/2020 | 2020-02-05 | Jiangxi Province Center for Disease Control and Prevention | Jiangxi Province Center for Disease Control and Prevention | JianXiong Li et al |
| 262 | China/IQTC01/2020 | 2020-02-05 | | Technology Centre, Guangzhou Customs | Shi et al |
| 263 | England/200641094/2020 | 2020-02-05 | Respiratory Virus Unit, Microbiology Services Colindale, Public Health England | Respiratory Virus Unit, Microbiology Services Colindale, Public Health England | Monica Galiano et al |
| 264 | Korea/KCDC2007/2020 | 2020-02-05 | Division of Viral Diseases, Center for Laboratory Control of Infectious Diseases, Korea Centers for Diseases Control and Prevention | Division of Viral Diseases, Center for Laboratory Control of Infectious Diseases, Korea Centers for Diseases Control and Prevention | Jeong-Min Kim et al |
| 265 | Korea/KCDC2010/2020 | 2020-02-05 | Division of Viral Diseases, Center for Laboratory Control of Infectious Diseases, Korea Centers for Diseases Control and Prevention | Division of Viral Diseases, Center for Laboratory Control of Infectious Diseases, Korea Centers for Diseases Control and Prevention | Jeong-Min Kim et al |
| 266 | Guangzhou/GZMU0019/2020 | 2020-02-05 | The First Affiliated Hospital of Guangzhou Medical University | BGI-shenzhen & The First Affiliated Hospital of Guangzhou Medical University | et al |
| 267 | Shanghai/SH0073/2020 | 2020-02-05 | Shanghai Public Health Clinical Center, Shanghai Medical College, Fudan University | National Research Center for Translational Medicine (Shanghai), Ruijin Hospital affiliated to Shanghai Jiao Tong University School of Medicine & Shanghai Public Health Clinical Center | Shengyue Wang et al |
| 268 | Guangzhou/GZMU0023/2020 | 2020-02-05 | The First Affiliated Hospital of Guangzhou Medical University | BGI-shenzhen & The First Affiliated Hospital of Guangzhou Medical University | Yanqun Wang et al |
| 269 | Guangzhou/GZMU0017/2020 | 2020-02-05 | The First Affiliated Hospital of Guangzhou Medical University | BGI-shenzhen & The First Affiliated Hospital of Guangzhou Medical University | et al |
| 270 | Guangzhou/GZMU0025/2020 | 2020-02-05 | The First Affiliated Hospital of Guangzhou Medical University | BGI-shenzhen & The First Affiliated Hospital of Guangzhou Medical University | et al |
| 271 | Guangzhou/GZMU0024/2020 | 2020-02-05 | The First Affiliated Hospital of Guangzhou Medical University | BGI-shenzhen & The First Affiliated Hospital of Guangzhou Medical University | Yanqun Wang et al |
| 272 | Korea/KCDC2008/2020 | 2020-02-05 | Division of Viral Diseases, Center for Laboratory Control of Infectious Diseases, Korea Centers for Diseases Control and Prevention | Division of Viral Diseases, Center for Laboratory Control of Infectious Diseases, Korea Centers for Diseases Control and Prevention | Jeong-Min Kim et al |
| 273 | Shanghai/SH0025/2020 | 2020-02-06 | Shanghai Public Health Clinical Center, Shanghai Medical College, Fudan University | National Research Center for Translational Medicine (Shanghai), Ruijin Hospital affiliated to Shanghai Jiao Tong University School of Medicine & Shanghai Public Health Clinical Center | Shengyue Wang et al |
| 274 | Guangdong/20SF2546/2020 | 2020-02-06 | Guangdong Provincial Center for Diseases Control and Prevention;Guangdong Provincial Institute of Public Health | School of Public Health, The University of Hong Kong | Bosheng Li et al |
| 275 | Singapore/5/2020 | 2020-02-06 | Singapore General Hospital, Molecular Laboratory, Division of Pathology | Programme in Emerging Infectious Diseases, Duke-NUS Medical School | Danielle E Anderson et al |
| 276 | SouthKorea/KCDC24/2020 | 2020-02-06 | Division of Viral Diseases, Center for Laboratory Control of Infectious Diseases, Korea Centers for Diseases Control and Prevention | Division of Viral Diseases, Center for Laboratory Control of Infectious Diseases, Korea Centers for Diseases Control and Prevention | Jeong-Min Kim et al |
| 277 | USA/CA7/2020 | 2020-02-06 | California Department of Public Health | Pathogen Discovery, Respiratory Viruses Branch, Division of | Krista Queen et al |

| | | | | | |
|---|---|---|---|---|---|
| | | | | Viral Diseases, Centers for Disease Control and Prevention | |
| 278 | Singapore/35/2020 | 2020-02-06 | National Public Health Laboratory, National Centre for Infectious Diseases | National Public Health Laboratory, National Centre for Infectious Diseases | Mak TM et al |
| 279 | Malaysia/MKAK-CL-2020-7554/2020 | 2020-02-06 | National Public Health Laboratory | Malaysia Genome Institute | Mohd Noor Mat Isa et al |
| 280 | SouthKorea/KUMC01/2020 | 2020-02-06 | Department of Microbiology, Institute for Viral Diseases, College of Medicine, Korea University | Department of Microbiology, Institute for Viral Diseases, College of Medicine, Korea University | Changmin Kang et al |
| 281 | Singapore/16/2020 | 2020-02-06 | National Public Health Laboratory, National Centre for Infectious Diseases | National Public Health Laboratory, National Centre for Infectious Diseases | Mak TM et al |
| 282 | SouthKorea/KUMC02/2020 | 2020-02-06 | Department of Microbiology, Institute for Viral Diseases, College of Medicine, Korea University | Department of Microbiology, Institute for Viral Diseases, College of Medicine, Korea University | Changmin Kang et al |
| 283 | Taiwan/TSGH-36/2020 | 2020-02-06 | TSGH-CP molecular lab | TSGH-CP molecular lab | Cherng-Lih Perng et al |
| 284 | Taiwan/TSGH-35/2020 | 2020-02-06 | TSGH-CP molecular lab | TSGH-CP molecular lab | Cherng-Lih Perng et al |
| 285 | England/200690306/2020 | 2020-02-07 | Respiratory Virus Unit, Microbiology Services Colindale, Public Health England | Respiratory Virus Unit, Microbiology Services Colindale, Public Health England | Monica Galiano et al |
| 286 | Guangdong/GD2020233-P0027/2020 | 2020-02-07 | Guangdong Provincial Institution of Public Health, Guangdong Provinical Center for Disease Control and Prevention | Guangdong Provincial Institution of Public Health | Jing Lu et al |
| 287 | Wuhan/HBCDC-HB-06/2020 | 2020-02-07 | Wuhan Lung Hospital | Hubei Provincial Center for Disease Control and Prevention | Bin Fang et al |
| 288 | Shanghai/SH0074/2020 | 2020-02-07 | Shanghai Public Health Clinical Center, Shanghai Medical College, Fudan University | National Research Center for Translational Medicine (Shanghai), Ruijin Hospital affiliated to Shanghai Jiao Tong University School of Medicine & Shanghai Public Health Clinical Center | Shengyue Wang et al |
| 289 | Korea/KCDC2009/2020 | 2020-02-07 | Division of Viral Diseases, Center for Laboratory Control of Infectious Diseases, Korea Centers for Diseases Control and Prevention | Division of Viral Diseases, Center for Laboratory Control of Infectious Diseases, Korea Centers for Diseases Control and Prevention | Jeong-Min Kim et al |
| 290 | Sweden/01/2020 | 2020-02-07 | | Unit for Laboratory Development and Technology Transfer, Public Health Agency of Sweden | Bengner et al |
| 291 | Taiwan/TSGH-01/2020 | 2020-02-07 | TSGH-CP molecular lab | TSGH-CP molecular lab | Cherng-Lih Perng et al |
| 292 | Beijing/BJ710/2020 | 2020-02-07 | Chinese PLA Institute for Disease Control and Prevention | Chinese PLA Institute for Disease Control and Prevention | Peng Li et al |
| 293 | HongKong/CUHK1/2020 | 2020-02-07 | Department of Microbiology, Faculty of Medicine, The Chinese University of Hong Kong, Hong Kong SAR, China | Department of Microbiology, Faculty of Medicine, Chinese University of Hong Kong, Hong Kong SAR, China | Zigui Chen et al |
| 294 | Guangzhou/GZMU0009/2020 | 2020-02-07 | The First Affiliated Hospital of Guangzhou Medical University | BGI-shenzhen & The First Affiliated Hospital of Guangzhou Medical University | et al |
| 295 | Shanghai/SH0070/2020 | 2020-02-07 | Shanghai Public Health Clinical Center, Shanghai Medical College, Fudan University | National Research Center for Translational Medicine (Shanghai), Ruijin Hospital affiliated to Shanghai Jiao Tong University School of Medicine & Shanghai Public Health Clinical Center | Shengyue Wang et al |
| 296 | Guangzhou/GZMU0010/2020 | 2020-02-07 | The First Affiliated Hospital of Guangzhou Medical University | BGI-shenzhen & The First Affiliated Hospital of Guangzhou Medical University | Yanqun Wang et al |
| 297 | Guangzhou/GZMU0008/2020 | 2020-02-07 | The First Affiliated Hospital of Guangzhou Medical University | BGI-shenzhen & The First Affiliated Hospital of Guangzhou Medical University | Yanqun Wang et al |

| | | | | | |
|---|---|---|---|---|---|
| 298 | Guangzhou/GZMU0013/2020 | 2020-02-07 | The First Affiliated Hospital of Guangzhou Medical University | BGI-shenzhen & The First Affiliated Hospital of Guangzhou Medical University | et al |
| 299 | Guangzhou/GZMU0003/2020 | 2020-02-07 | The First Affiliated Hospital of Guangzhou Medical University | BGI-shenzhen & The First Affiliated Hospital of Guangzhou Medical University | Yanqun Wang et al |
| 300 | France/RA739/2020 | 2020-02-08 | CNR Virus des Infections Respiratoires - France SUD | CNR Virus des Infections Respiratoires - France SUD | Bal et al |
| 301 | England/200690300/2020 | 2020-02-08 | Respiratory Virus Unit, Microbiology Services Colindale, Public Health England | Respiratory Virus Unit, Microbiology Services Colindale, Public Health England | Monica Galiano et al |
| 302 | Australia/VIC07/2020 | 2020-02-08 | Victorian Infectious Diseases Reference Laboratory (VIDRL) | Victorian Infectious Diseases Reference Laboratory and Microbiological Diagnostic Unit Public Health Laboratory, Doherty Institute | Caly L. et al |
| 303 | Guangzhou/GZMU0050/2020 | 2020-02-08 | The First Affiliated Hospital of Guangzhou Medical University | BGI-shenzhen & The First Affiliated Hospital of Guangzhou Medical University | et al |
| 304 | Guangzhou/GZMU0046/2020 | 2020-02-08 | The First Affiliated Hospital of Guangzhou Medical University | BGI-shenzhen & The First Affiliated Hospital of Guangzhou Medical University | Yanqun Wang et al |
| 305 | Guangzhou/GZMU0037/2020 | 2020-02-08 | The First Affiliated Hospital of Guangzhou Medical University | BGI-shenzhen & The First Affiliated Hospital of Guangzhou Medical University | Yanqun Wang et al |
| 306 | England/200690245/2020 | 2020-02-08 | Respiratory Virus Unit, Microbiology Services Colindale, Public Health England | Respiratory Virus Unit, Microbiology Services Colindale, Public Health England | Monica Galiano et al |
| 307 | UnitedArabEmirates/L4280/2020 | 2020-02-08 | Mohammed Bin Rashid University of Medicine and Health Sciences | Al Jalila Genomics Center | Ahmad Abou Tayoun et al |
| 308 | England/200690756/2020 | 2020-02-08 | Respiratory Virus Unit, Microbiology Services Colindale, Public Health England | Respiratory Virus Unit, Microbiology Services Colindale, Public Health England | Monica Galiano et al |
| 309 | Guangzhou/GZMU0041/2020 | 2020-02-08 | The First Affiliated Hospital of Guangzhou Medical University | BGI-shenzhen & The First Affiliated Hospital of Guangzhou Medical University | Yanqun Wang et al |
| 310 | HongKong/HKPU29_0102/2020 | 2020-02-08 | Department of Clinical Pathology, Pamela Youde Nethersole Eastern Hospital | Department of Health Technology and Informatics, Faculty of Health and Social Science, The Hong Kong Polytechnic University | Kenneth Siu-Sing LEUNG et al |
| 311 | HongKong/HKPU30_2901/2020 | 2020-02-08 | Department of Clinical Pathology, Pamela Youde Nethersole Eastern Hospital | Department of Health Technology and Informatics, Faculty of Health and Social Science, The Hong Kong Polytechnic University | Kenneth Siu-Sing LEUNG et al |
| 312 | Guangzhou/GZMU0049/2020 | 2020-02-08 | The First Affiliated Hospital of Guangzhou Medical University | BGI-shenzhen & The First Affiliated Hospital of Guangzhou Medical University | et al |
| 313 | Guangzhou/GZMU0039/2020 | 2020-02-08 | The First Affiliated Hospital of Guangzhou Medical University | BGI-shenzhen & The First Affiliated Hospital of Guangzhou Medical University | Yanqun Wang et al |
| 314 | Shanghai/SH0080/2020 | 2020-02-08 | Shanghai Public Health Clinical Center, Shanghai Medical College, Fudan University | National Research Center for Translational Medicine (Shanghai), Ruijin Hospital affiliated to Shanghai Jiao Tong University School of Medicine & Shanghai Public Health Clinical Center | Shengyue Wang et al |
| 315 | Shanghai/SH0055/2020 | 2020-02-08 | Shanghai Public Health Clinical Center, Shanghai Medical College, Fudan University | National Research Center for Translational Medicine (Shanghai), Ruijin Hospital affiliated to Shanghai Jiao Tong University School of Medicine & Shanghai Public Health Clinical Center | Shengyue Wang et al |
| 316 | Shangrao/JX1974/2020 | 2020-02-08 | Jiangxi Province Center for Disease Control and | Jiangxi Province Center for Disease Control and Prevention | JianXiong Li et al |

| | | | | | |
|---|---|---|---|---|---|
| | | | Prevention | | |
| 317 | Guangzhou/GZMU0045/2020 | 2020-02-08 | The First Affiliated Hospital of Guangzhou Medical University | BGI-shenzhen & The First Affiliated Hospital of Guangzhou Medical University | Yanqun Wang et al |
| 318 | Beijing/BJ752/2020 | 2020-02-08 | Chinese PLA Institute for Disease Control and Prevention | Chinese PLA Institute for Disease Control and Prevention | Peng Li et al |
| 319 | Taiwan/TSGH-37/2020 | 2020-02-08 | TSGH-CP molecular lab | TSGH-CP molecular lab | Cherng-Lih Perng et al |
| 320 | Tianmen/HBCDC-HB-07/2020 | 2020-02-08 | Tianmen Center for Disease Control and Prevention | Hubei Provincial Center for Disease Control and Prevention | Bin Fang et al |
| 321 | HongKong/HKPU32_0402/2020 | 2020-02-09 | Department of Clinical Pathology, Pamela Youde Nethersole Eastern Hospital | Department of Health Technology and Informatics, Faculty of Health and Social Science, The Hong Kong Polytechnic University | Kenneth Siu-Sing LEUNG et al |
| 322 | Shanghai/SH0119/2020 | 2020-02-09 | Shanghai Public Health Clinical Center, Shanghai Medical College, Fudan University | National Research Center for Translational Medicine (Shanghai), Ruijin Hospital affiliated to Shanghai Jiao Tong University School of Medicine & Shanghai Public Health Clinical Center | Shengyue Wang et al |
| 323 | Shanghai/SH0101/2020 | 2020-02-09 | Shanghai Public Health Clinical Center, Shanghai Medical College, Fudan University | National Research Center for Translational Medicine (Shanghai), Ruijin Hospital affiliated to Shanghai Jiao Tong University School of Medicine & Shanghai Public Health Clinical Center | Shengyue Wang et al |
| 324 | Korea/KCDC2012/2020 | 2020-02-09 | Division of Viral Diseases, Center for Laboratory Control of Infectious Diseases, Korea Centers for Diseases Control and Prevention | Division of Viral Diseases, Center for Laboratory Control of Infectious Diseases, Korea Centers for Diseases Control and Prevention | Jeong-Min Kim et al |
| 325 | England/09c/2020 | 2020-02-09 | Respiratory Virus Unit, Microbiology Services Colindale, Public Health England | Respiratory Virus Unit, Microbiology Services Colindale, Public Health England | Monica Galiano et al |
| 326 | HongKong/VM20002493/2020 | 2020-02-09 | Hong Kong Department of Health | School of Public Health, The University of Hong Kong | Dominic N.C. Tsang et al |
| 327 | HongKong/HKPU28_3001/2020 | 2020-02-09 | Department of Pathology, United Christian Hospital | Department of Health Technology and Informatics, Faculty of Health and Social Science, The Hong Kong Polytechnic University | Kenneth Siu-Sing LEUNG et al |
| 328 | HongKong/HKPU36-0702/2020 | 2020-02-09 | Department of Clinical Pathology, Pamela Youde Nethersole Eastern Hospital | Department of Health Technology and Informatics, Faculty of Health and Social Science, The Hong Kong Polytechnic University | Kenneth Siu-Sing LEUNG et al |
| 329 | Singapore/6/2020 | 2020-02-09 | Singapore General Hospital, Molecular Laboratory, Division of Pathology | Programme in Emerging Infectious Diseases, Duke-NUS Medical School | Danielle E Anderson et al |
| 330 | HongKong/HKPU34_3001/2020 | 2020-02-09 | Department of Clinical Pathology, Pamela Youde Nethersole Eastern Hospital | Department of Health Technology and Informatics, Faculty of Health and Social Science, The Hong Kong Polytechnic University | Kenneth Siu-Sing LEUNG et al |
| 331 | Guangzhou/GZMU0060/2020 | 2020-02-09 | The First Affiliated Hospital of Guangzhou Medical University | BGI-shenzhen & The First Affiliated Hospital of Guangzhou Medical University | et al |
| 332 | Guangdong/GD2020246-P0028/2020 | 2020-02-09 | Guangdong Provincial Institution of Public Health, Guangdong Provinical Center for Disease Control and Prevention | Guangdong Provincial Institution of Public Health | Jing Lu et al |
| 333 | Shanghai/SH0077/2020 | 2020-02-09 | Shanghai Public Health Clinical Center, Shanghai Medical College, Fudan University | National Research Center for Translational Medicine (Shanghai), Ruijin Hospital affiliated to Shanghai Jiao Tong University School of Medicine | Shengyue Wang et al |

| | | | | | |
|---|---|---|---|---|---|
| | | | | & Shanghai Public Health Clinical Center | |
| 334 | HongKong/HKPU40-2801/2020 | 2020-02-10 | Department of Clinical Pathology, Pamela Youde Nethersole Eastern Hospital | Department of Health Technology and Informatics, Faculty of Health and Social Science, The Hong Kong Polytechnic University | Kenneth Siu-Sing LEUNG et al |
| 335 | HongKong/HKPU39-3001/2020 | 2020-02-10 | Department of Clinical Pathology, Pamela Youde Nethersole Eastern Hospital | Department of Health Technology and Informatics, Faculty of Health and Social Science, The Hong Kong Polytechnic University | Kenneth Siu-Sing LEUNG et al |
| 336 | HongKong/HKPU41-0802/2020 | 2020-02-10 | Department of Clinical Pathology, Pamela Youde Nethersole Eastern Hospital | Department of Health Technology and Informatics, Faculty of Health and Social Science, The Hong Kong Polytechnic University | Kenneth Siu-Sing LEUNG et al |
| 337 | HongKong/VM20002507/2020 | 2020-02-10 | Hong Kong Department of Health | School of Public Health, The University of Hong Kong | Dominic N.C. Tsang et al |
| 338 | Beijing/BJ782/2020 | 2020-02-10 | Chinese PLA Institute for Disease Control and Prevention | Chinese PLA Institute for Disease Control and Prevention | Peng Li et al |
| 339 | Japan/Hu_DP_Kng_19-020/2020 | 2020-02-10 | | Takayuki Hishiki Kanagawa Prefectural Institute of Public Health, Department of Microbiology | Hishiki et al |
| 340 | USA/CA8/2020 | 2020-02-10 | California Department of Public Health | Pathogen Discovery, Respiratory Viruses Branch, Division of Viral Diseases, Centers for Disease Control and Prevention | Krista Queen et al |
| 341 | Singapore/17/2020 | 2020-02-10 | National Public Health Laboratory, National Centre for Infectious Diseases | National Public Health Laboratory, National Centre for Infectious Diseases | Mak TM et al |
| 342 | Japan/Hu_DP_Kng_19-027/2020 | 2020-02-10 | | Takayuki Hishiki Kanagawa Prefectural Institute of Public Health, Department of Microbiology | Hishiki et al |
| 343 | Shanghai/SH0115/2020 | 2020-02-11 | Shanghai Public Health Clinical Center, Shanghai Medical College, Fudan University | National Research Center for Translational Medicine (Shanghai), Ruijin Hospital affiliated to Shanghai Jiao Tong University School of Medicine & Shanghai Public Health Clinical Center | Shengyue Wang et al |
| 344 | Vietnam/CM99/2020 | 2020-02-11 | National Influenza Center, National Institute of Hygiene and Epidemiology (NIHE) | National Influenza Center, National Institute of Hygiene and Epidemiology (NIHE) | Le Quynh Mai et al |
| 345 | USA/TX1/2020 | 2020-02-11 | Texas Department of State Health Services | Pathogen Discovery, Respiratory Viruses Branch, Division of Viral Diseases, Centers for Disease Control and Prevention | Krista Queen et al |
| 346 | HongKong/HKPU48-0202/2020 | 2020-02-11 | Department of Pathology, Princess Margaret Hospital | Department of Health Technology and Informatics, Faculty of Health and Social Science, The Hong Kong Polytechnic University | Kenneth Siu-Sing LEUNG et al |
| 347 | HongKong/VM20002582/2020 | 2020-02-12 | Hong Kong Department of Health | School of Public Health, The University of Hong Kong | Dominic N.C. Tsang et al |
| 348 | Korea/KCDC2014/2020 | 2020-02-12 | Division of Viral Diseases, Center for Laboratory Control of Infectious Diseases, Korea Centers for Diseases Control and Prevention | Division of Viral Diseases, Center for Laboratory Control of Infectious Diseases, Korea Centers for Diseases Control and Prevention | Jeong-Min Kim et al |
| 349 | Malaysia/IMR_WC627/2020 | 2020-02-12 | Institute for Medical Research, Infectious Disease Research Centre, National Institutes of Health, Ministry of Health Malaysia | Institute for Medical Research, Infectious Disease Research Centre, National Institutes of Health, Ministry of Health Malaysia | Suppiah.J et al |
| 350 | HongKong/HKPU52-3101/2020 | 2020-02-13 | Department of Clinical Pathology, Pamela Youde Nethersole Eastern Hospital | Department of Health Technology and Informatics, Faculty of Health and Social | Kenneth Siu-Sing LEUNG et al |

| | | | | | |
|---|---|---|---|---|---|
| | | | | Science, The Hong Kong Polytechnic University | |
| 351 | Singapore/21/2020 | 2020-02-13 | National Public Health Laboratory, National Centre for Infectious Diseases | National Public Health Laboratory, National Centre for Infectious Diseases | Mak TM et al |
| 352 | Singapore/14/2020 | 2020-02-13 | National Centre for Infectious Diseases | Programme in Emerging Infectious Diseases, Duke-NUS Medical School | Danielle E Anderson et al |
| 353 | HongKong/HKPU54-0302/2020 | 2020-02-13 | Department of Clinical Pathology, Pamela Youde Nethersole Eastern Hospital | Department of Health Technology and Informatics, Faculty of Health and Social Science, The Hong Kong Polytechnic University | Kenneth Siu-Sing LEUNG et al |
| 354 | Shanghai/SH0110/2020 | 2020-02-13 | Shanghai Public Health Clinical Center, Shanghai Medical College, Fudan University | National Research Center for Translational Medicine (Shanghai), Ruijin Hospital affiliated to Shanghai Jiao Tong University School of Medicine & Shanghai Public Health Clinical Center | Shengyue Wang et al |
| 355 | Singapore/14Clin/2020 | 2020-02-14 | National Public Health Laboratory, National Centre for Infectious Diseases | National Public Health Laboratory, National Centre for Infectious Diseases | Mak TM et al |
| 356 | Japan/Hu_DP_Kng_19-031/2020 | 2020-02-14 | | Takayuki Hishiki Kanagawa Prefectural Institute of Public Health | Hishiki et al |
| 357 | Shanghai/SH0102/2020 | 2020-02-14 | Shanghai Public Health Clinical Center, Shanghai Medical College, Fudan University | National Research Center for Translational Medicine (Shanghai), Ruijin Hospital affiliated to Shanghai Jiao Tong University School of Medicine & Shanghai Public Health Clinical Center | Shengyue Wang et al |
| 358 | HongKong/HKPU57-0702/2020 | 2020-02-15 | Department of Clinical Pathology, Pamela Youde Nethersole Eastern Hospital | Department of Health Technology and Informatics, Faculty of Health and Social Science, The Hong Kong Polytechnic University | Kenneth Siu-Sing LEUNG et al |
| 359 | Shanghai/SH0126/2020 | 2020-02-15 | Shanghai Public Health Clinical Center, Shanghai Medical College, Fudan University | National Research Center for Translational Medicine (Shanghai), Ruijin Hospital affiliated to Shanghai Jiao Tong University School of Medicine & Shanghai Public Health Clinical Center | Shengyue Wang et al |
| 360 | Shanghai/SH0125/2020 | 2020-02-15 | Shanghai Public Health Clinical Center, Shanghai Medical College, Fudan University | National Research Center for Translational Medicine (Shanghai), Ruijin Hospital affiliated to Shanghai Jiao Tong University School of Medicine & Shanghai Public Health Clinical Center | Shengyue Wang et al |
| 361 | Singapore/43/2020 | 2020-02-16 | National Public Health Laboratory, National Centre for Infectious Diseases | National Public Health Laboratory, National Centre for Infectious Diseases | Mak TM et al |
| 362 | Singapore/36/2020 | 2020-02-16 | National Public Health Laboratory, National Centre for Infectious Diseases | National Public Health Laboratory, National Centre for Infectious Diseases | Mak TM et al |
| 363 | Singapore/12/2020 | 2020-02-17 | National Centre for Infectious Diseases | Programme in Emerging Infectious Diseases, Duke-NUS Medical School | Danielle E Anderson et al |
| 364 | Guangdong/20SF4047/2020 | 2020-02-17 | Guangdong Provincial Center for Diseases Control and Prevention;Guangdong Provincial Institute of Public Health | School of Public Health, The University of Hong Kong | Bosheng Li et al |
| 365 | HongKong/HKPU60-0802/2020 | 2020-02-17 | Department of Clinical Pathology, Pamela Youde Nethersole Eastern Hospital | Department of Health Technology and Informatics, Faculty of Health and Social Science, The Hong Kong Polytechnic University | Kenneth Siu-Sing LEUNG et al |
| 366 | Korea/KCDC2016/2020 | 2020-02-18 | Division of Viral Diseases, Center for Laboratory Control | Division of Viral Diseases, Center for Laboratory Control of | Jeong-Min Kim et al |

| | | | | | |
|---|---|---|---|---|---|
| | | | of Infectious Diseases, Korea Centers for Diseases Control and Prevention | Infectious Diseases, Korea Centers for Diseases Control and Prevention | |
| 367 | Korea/KCDC2019/2020 | 2020-02-18 | Division of Viral Diseases, Center for Laboratory Control of Infectious Diseases, Korea Centers for Diseases Control and Prevention | Division of Viral Diseases, Center for Laboratory Control of Infectious Diseases, Korea Centers for Diseases Control and Prevention | Jeong-Min Kim et al |
| 368 | Korea/KCDC2017/2020 | 2020-02-18 | Division of Viral Diseases, Center for Laboratory Control of Infectious Diseases, Korea Centers for Diseases Control and Prevention | Division of Viral Diseases, Center for Laboratory Control of Infectious Diseases, Korea Centers for Diseases Control and Prevention | Jeong-Min Kim et al |
| 369 | Singapore/13/2020 | 2020-02-18 | National Centre for Infectious Diseases | Programme in Emerging Infectious Diseases, Duke-NUS Medical School | Danielle E Anderson et al |
| 370 | HongKong/HKPU64-1202/2020 | 2020-02-19 | Department of Clinical Pathology, Pamela Youde Nethersole Eastern Hospital | Department of Health Technology and Informatics, Faculty of Health and Social Science, The Hong Kong Polytechnic University | Kenneth Siu-Sing LEUNG et al |
| 371 | Korea/KCDC2018/2020 | 2020-02-19 | Division of Viral Diseases, Center for Laboratory Control of Infectious Diseases, Korea Centers for Diseases Control and Prevention | Division of Viral Diseases, Center for Laboratory Control of Infectious Diseases, Korea Centers for Diseases Control and Prevention | Jeong-Min Kim et al |
| 372 | HongKong/HKPU63-1402/2020 | 2020-02-19 | Department of Clinical Pathology, Tuen Mun Hospital, 23 Tsing Chung Koon Road, Tuen Mun, N.T. | Department of Health Technology and Informatics, Faculty of Health and Social Science, The Hong Kong Polytechnic University | Kenneth Siu-Sing LEUNG et al |
| 373 | USA/WA-S2/2020 | 2020-02-20 | Seattle Flu Study | Seattle Flu Study | Chu et al et al |
| 374 | Canada/ON_PHL5472/2020 | 2020-02-20 | Public Health Ontario Laboratories | Public Health Ontario Laboratories | Alireza Eshaghi et al |
| 375 | Italy/CDG1/2020 | 2020-02-20 | Department of Infectious Diseases, Istituto Superiore di Sanità, Roma , Italy | Virology Laboratory, Scientific Department, Army Medical Center | Paola Stefanelli et al |
| 376 | Japan/TK-20-31-3/2020 | 2020-02-20 | Department of Pathology, Toshima Hospital | Pathogen Genomics Center, National Institute of Infectious Diseases | Tsuyoshi Sekizuka et al |
| 377 | France/HF1463/2020 | 2020-02-21 | Centre Hospitalier Compiègne Laboratoire de Biologie | National Reference Center for Viruses of Respiratory Infections, Institut Pasteur, Paris | Mélanie Albert et al |
| 378 | Australia/NT02/2020 | 2020-02-21 | Royal Darwin Hospital | Victorian Infectious Diseases Reference Laboratory and Microbiological Diagnostic Unit Public Health Laboratory, Doherty Institute | Meumann et al |
| 379 | Australia/NT01/2020 | 2020-02-21 | Royal Darwin Hospital | Victorian Infectious Diseases Reference Laboratory and Microbiological Diagnostic Unit Public Health Laboratory, Doherty Institute | Meumann et al |
| 380 | USA/WA-S84/2020 | 2020-02-21 | Washington State Department of Health | Seattle Flu Study | Chu etl al et al |
| 381 | France/HF1465/2020 | 2020-02-21 | Centre Hospitalier Compiègne Laboratoire de Biologie | National Reference Center for Viruses of Respiratory Infections, Institut Pasteur, Paris | Mélanie Albert et al |
| 382 | USA/WA-S82/2020 | 2020-02-22 | Washington State Department of Health | Seattle Flu Study | Chu etl al et al |
| 383 | Australia/WA11/202 | 2020-02-22 | PathWest Laboratory Medicine WA | PathWest Laboratory Medicine WA | Sikazwe et al |
| 384 | HongKong/VM20002849/2020 | 2020-02-22 | Hong Kong Department of Health | School of Public Health, The University of Hong Kong | Dominic N.C. Tsang et al |
| 385 | USA/WA-S277/2020 | 2020-02-22 | Seattle Flu Study | Seattle Flu Study | Chu et al et al |
| 386 | USA/CA9/2020 | 2020-02-23 | California Department of Public Health | Pathogen Discovery, Respiratory Viruses Branch, Division of Viral Diseases, Centers for Disease Control and Prevention | Krista Queen et al |
| 387 | Israel/ISR_JP0320/2020 | 2020-02-23 | Central Virology Laboratory | Israel Institute for Biological Research | Inbar Cohen-Gihon et al |

| | | | | | |
|---|---|---|---|---|---|
| 388 | Australia/VIC138/2020 | 2020-02-23 | Victorian Infectious Diseases Reference Laboratory (VIDRL) | Victorian Infectious Diseases Reference Laboratory and Microbiological Diagnostic Unit Public Health Laboratory, Doherty Institute | Caly L. et al |
| 389 | Korea/KCDC2013/2020 | 2020-02-23 | Division of Viral Diseases, Center for Laboratory Control of Infectious Diseases, Korea Centers for Diseases Control and Prevention | Division of Viral Diseases, Center for Laboratory Control of Infectious Diseases, Korea Centers for Diseases Control and Prevention | Jeong-Min Kim et al |
| 390 | Korea/KCDC2011/2020 | 2020-02-23 | Division of Viral Diseases, Center for Laboratory Control of Infectious Diseases, Korea Centers for Diseases Control and Prevention | Division of Viral Diseases, Center for Laboratory Control of Infectious Diseases, Korea Centers for Diseases Control and Prevention | Jeong-Min Kim et al |
| 391 | Korea/KCDC2001/2020 | 2020-02-23 | Division of Viral Diseases, Center for Laboratory Control of Infectious Diseases, Korea Centers for Diseases Control and Prevention | Division of Viral Diseases, Center for Laboratory Control of Infectious Diseases, Korea Centers for Diseases Control and Prevention | Jeong-Min Kim et al |
| 392 | Korea/KCDC2003/2020 | 2020-02-23 | Division of Viral Diseases, Center for Laboratory Control of Infectious Diseases, Korea Centers for Diseases Control and Prevention | Division of Viral Diseases, Center for Laboratory Control of Infectious Diseases, Korea Centers for Diseases Control and Prevention | Jeong-Min Kim et al |
| 393 | HongKong/HKPU83-1702/2020 | 2020-02-24 | Department of Clinical Pathology, Pamela Youde Nethersole Eastern Hospital | Department of Health Technology and Informatics, Faculty of Health and Social Science, The Hong Kong Polytechnic University | Kenneth Siu-Sing LEUNG et al |
| 394 | USA/WA2/2020 | 2020-02-24 | Washington State Department of Health | Seattle Flu Study | Helen Chu et al |
| 395 | Italy/UniMI01/2020 | 2020-02-24 | Laboratory of Infectious Diseases, Department of Biomedical and Clinical Sciences L. Sacco, University of Milan | Laboratory of Infectious Diseases, Department of Biomedical and Clinical Sciences L. Sacco, University of Milan | Gianguglielmo Zehender et al |
| 396 | USA/WA-S254/2020 | 2020-02-24 | Washington State Department of Health | Seattle Flu Study | Chu et al et al |
| 397 | Australia/NT05/2020 | 2020-02-24 | Royal Darwin Hospital | Victorian Infectious Diseases Reference Laboratory and Microbiological Diagnostic Unit Public Health Laboratory, Doherty Institute | Meumann et al |
| 398 | HongKong/VM20002868/2020 | 2020-02-24 | Hong Kong Department of Health | School of Public Health, The University of Hong Kong | Dominic N.C. Tsang et al |
| 399 | Italy/UniMI03/2020 | 2020-02-24 | Laboratory of Infectious Diseases, Department of Biomedical and Clinical Sciences L. Sacco, University of Milan | Laboratory of Infectious Diseases, Department of Biomedical and Clinical Sciences L. Sacco, University of Milan | Gianguglielmo Zehender et al |
| 400 | USA/WA-S7/2020 | 2020-02-24 | Seattle Flu Study | Seattle Flu Study | Chu et al et al |
| 401 | Switzerland/TI9486/2020 | 2020-02-24 | Laboratoire de Virologie, HUG | Swiss National Reference Centre for Influenza | LAUBSCHER Florian et al. et al |
| 402 | UnitedArabEmirates/L6599/2020 | 2020-02-25 | Mohammed Bin Rashid University of Medicine and Health Sciences | Al Jalila Genomics Center | Ahmad Abou Tayoun et al |
| 403 | Guangzhou/GZMU0014/2020 | 2020-02-25 | State Key Laboratory of Respiratory Disease, National Clinical Research Center for Respiratory Disease, Guangzhou Institute of Respiratory Health, the First Affiliated Hospital of Guangzhou Medical University | The First Affiliated Hospital of Guangzhou Medical University & BGI-Shenzhen | Zhao et al et al |
| 404 | Guangzhou/GZMU0044/2020 | 2020-02-25 | State Key Laboratory of Respiratory Disease, National Clinical Research Center for Respiratory Disease, Guangzhou Institute of Respiratory Health, the First Affiliated Hospital of | The First Affiliated Hospital of Guangzhou Medical University & BGI-Shenzhen | Zhao et al et al |

| | | | | | |
|---|---|---|---|---|---|
| | | | Guangzhou Medical University | | |
| 405 | Beijing/BJ1112/2020 | 2020-02-25 | Chinese PLA Institute for Disease Control and Prevention | Chinese PLA Institute for Disease Control and Prevention | Peng Li et al |
| 406 | Guangzhou/GZMU0048/2020 | 2020-02-25 | State Key Laboratory of Respiratory Disease, National Clinical Research Center for Respiratory Disease, Guangzhou Institute of Respiratory Health, the First Affiliated Hospital of Guangzhou Medical University | The First Affiliated Hospital of Guangzhou Medical University & BGI-Shenzhen | Zhao et al et al |
| 407 | Guangzhou/GZMU0042/2020 | 2020-02-25 | State Key Laboratory of Respiratory Disease, National Clinical Research Center for Respiratory Disease, Guangzhou Institute of Respiratory Health, the First Affiliated Hospital of Guangzhou Medical University | The First Affiliated Hospital of Guangzhou Medical University & BGI-Shenzhen | Zhao et al et al |
| 408 | HongKong/VM20002907/2020 | 2020-02-25 | Hong Kong Department of Health | School of Public Health, The University of Hong Kong | Dominic N.C. Tsang et al |
| 409 | Spain/Madrid201105/2020 | 2020-02-25 | HOSPITAL UNIVERSITARIO LA PAZ | Instituto de Salud Carlos III | Iglesias-Caballero et al |
| 410 | UnitedArabEmirates/L0184/2020 | 2020-02-25 | Mohammed Bin Rashid University of Medicine and Health Sciences | Al Jalila Genomics Center | Ahmad Abou Tayoun et al |
| 411 | UnitedArabEmirates/L2409/2020 | 2020-02-25 | Mohammed Bin Rashid University of Medicine and Health Sciences | Al Jalila Genomics Center | Ahmad Abou Tayoun et al |
| 412 | UnitedArabEmirates/L4682/2020 | 2020-02-25 | Mohammed Bin Rashid University of Medicine and Health Sciences | Al Jalila Genomics Center | Ahmad Abou Tayoun et al |
| 413 | Germany/NRW-02-1/2020 | 2020-02-25 | Center of Medical Microbiology, Virology, and Hospital Hygiene, University of Duesseldorf | Center of Medical Microbiology, Virology, and Hospital Hygiene, University of Duesseldorf | Ortwin Adams et al |
| 414 | UnitedArabEmirates/L0904/2020 | 2020-02-25 | Mohammed Bin Rashid University of Medicine and Health Sciences | Al Jalila Genomics Center | Ahmad Abou Tayoun et al |
| 415 | Germany/Baden-Wuerttemberg-1/2020 | 2020-02-25 | State Health Office Baden-Wuerttemberg | Charité Universitätsmedizin Berlin, Institute of Virology | Victor M Corman et al |
| 416 | England/200940527/2020 | 2020-02-25 | Respiratory Virus Unit, Microbiology Services Colindale, Public Health England | Respiratory Virus Unit, Microbiology Services Colindale, Public Health England | Monica Galiano et al |
| 417 | Brazil/SPBR-01/2020 | 2020-02-25 | Hospital Israelita Albert Einstein | Instituto Adolfo Lutz Interdisciplinary Procedures Center Strategic Laboratory | Jaqueline Goes de Jesus et al |
| 418 | Taiwan/TSGH-26/2020 | 2020-02-25 | TSGH-CP molecular lab | TSGH-CP molecular lab | Cherng-Lih Perng et al |
| 419 | France/GE1583/2020 | 2020-02-25 | Laboratoire de Virologie Institut de Virologie - INSERM U 1109 Hôpitaux Universitaires de Strasbourg | National Reference Center for Viruses of Respiratory Infections, Institut Pasteur, Paris | Mélnie Albert et al |
| 420 | England/200960515/2020 | 2020-02-25 | Respiratory Virus Unit, Microbiology Services Colindale, Public Health England | Respiratory Virus Unit, Microbiology Services Colindale, Public Health England | Monica Galiano et al |
| 421 | HongKong/HKPU89-2502/2020 | 2020-02-26 | Department of Clinical Pathology, Pamela Youde Nethersole Eastern Hospital | Department of Health Technology and Informatics, Faculty of Health and Social Science, The Hong Kong Polytechnic University | Kenneth Siu-Sing LEUNG et al |
| 422 | Nanchang/JXN3T4/2020 | 2020-02-26 | Jiangxi Province Center for Disease Control and Prevention | Jiangxi Province Center for Disease Control and Prevention | JianXiong Li et al |
| 423 | Korea/KCDC2015/2020 | 2020-02-26 | Division of Viral Diseases, Center for Laboratory Control of Infectious Diseases, Korea | Division of Viral Diseases, Center for Laboratory Control of Infectious Diseases, Korea | Jeong-Min Kim et al |

| | | | | | |
|---|---|---|---|---|---|
| | | | Centers for Diseases Control and Prevention | Centers for Diseases Control and Prevention | |
| 424 | Switzerland/1000477102/2020 | 2020-02-26 | Department of Internal Medicine, Triemli Hospital | Institute of Medical Virology, University of Zurich | Stefan Schmutz et al |
| 425 | England/200990006/2020 | 2020-02-26 | Respiratory Virus Unit, Microbiology Services Colindale, Public Health England | Respiratory Virus Unit, Microbiology Services Colindale, Public Health England | Monica Galiano et al |
| 426 | Norway/1380/2020 | 2020-02-26 | Oslo University Hospital, Department of Medical Microbiology | Norwegian Institute of Public Health, Department of Virology | Kathrine Stene-Johansen et al |
| 427 | England/200981386/2020 | 2020-02-26 | Respiratory Virus Unit, Microbiology Services Colindale, Public Health England | Respiratory Virus Unit, Microbiology Services Colindale, Public Health England | Monica Galiano et al |
| 428 | USA/WA-S272/2020 | 2020-02-26 | Seattle Flu Study | Seattle Flu Study | Chu et al et al |
| 429 | France/N1620/2020 | 2020-02-26 | Centre Hositalier Universitaire de Rouen Laboratoire de Virologie | National Reference Center for Viruses of Respiratory Infections, Institut Pasteur, Paris | Mélnie Albert et al |
| 430 | Taiwan/CGMH-CGU-03/2020 | 2020-02-26 | Laboratory Medicine | Department of Laboratory Medicine, Lin-Kou Chang Gung Memorial Hospital, Taoyuan, Taiwan | Kuo-Chien Tsao et al |
| 431 | USA/CA-SCCPHD-UC101/2020 | 2020-02-26 | Santa Clara County Public Health Department | Chiu Laboratory, University of California, San Francisco | Xianding Deng et al |
| 432 | Austria/CeMM0003/2020 | 2020-02-26 | Center for Virology, Medical University of Vienna | Bergthaler laboratory, CeMM Research Center for Molecular Medicine of the Austrian Academy of Sciences | Alexandra Popa et al |
| 433 | USA/CA_2602/2020 | 2020-02-26 | California Department of Public Health | Pathogen Discovery, Respiratory Viruses Branch, Division of Viral Diseases, Centers for Disease Control and Prevention | Ying Tao et al |
| 434 | France/PL1643/2020 | 2020-02-26 | Centre Hospitalier Régional Universitaire de Nantes Laboratoire de Virologie | National Reference Center for Viruses of Respiratory Infections, Institut Pasteur, Paris | Mélnie Albert et al |
| 435 | France/B2330/2020 | 2020-02-26 | CHRU Pontchaillou - Laboratoire de Virologie | National Reference Center for Viruses of Respiratory Infections, Institut Pasteur, Paris | Mélnie Albert et al |
| 436 | France/B1623/2020 | 2020-02-26 | CHU - Hôpital Cavale Blanche - Labo. de Virologie | National Reference Center for Viruses of Respiratory Infections, Institut Pasteur, Paris | Mélanie Albert et al |
| 437 | USA/CA-CDPH-UC1/2020 | 2020-02-26 | California Department of Public Health | Chiu Laboratory, University of California, San Francisco | Xianding Deng et al |
| 438 | Switzerland/GE3895/2020 | 2020-02-26 | Laboratoire de Virologie, HUG | Swiss National Reference Centre for Influenza | LAUBSCHER Florian et al. et al |
| 439 | USA/WA-S81/2020 | 2020-02-26 | Washington State Department of Health | Seattle Flu Study | Chu etl al et al |
| 440 | NewZealand/20VR0174/2020-02-27 | 2020-02-27 | LabPLUS | Institute of Environmental Science and Research (ESR) Ltd | **Matt Storey et al** |
| 441 | Guangzhou/GZMU0030/2020 | 2020-02-27 | State Key Laboratory of Respiratory Disease, National Clinical Research Center for Respiratory Disease, Guangzhou Institute of Respiratory Health, the First Affiliated Hospital of Guangzhou Medical University | The First Affiliated Hospital of Guangzhou Medical University & BGI-Shenzhen | Zhao et al et al |
| 442 | Switzerland/BL0902/2020 | 2020-02-27 | Laboratoire de Virologie, HUG | Swiss National Reference Centre for Influenza | LAUBSCHER Florian et al. et al |
| 443 | Singapore/51/2020 | 2020-02-27 | National Public Health Laboratory, National Centre for Infectious Diseases | National Public Health Laboratory, National Centre for Infectious Diseases | Mak TM et al |
| 444 | Switzerland/GR3043/2020 | 2020-02-27 | Hôpitaux universitaires de Genève Laboratoire de Virologie | Hôpitaux universitaires de Genève Laboratoire de Virologie | Laubscher F. et al |
| 445 | Taiwan/CGMH-CGU-05/2020 | 2020-02-27 | Laboratory Medicine | Department of Laboratory Medicine, Lin-Kou Chang Gung Memorial Hospital, Taoyuan, Taiwan | Kuo-Chien Tsao et al |

| | | | | | |
|---|---|---|---|---|---|
| 446 | Mexico/CDMX-InDRE_01/2020 | 2020-02-27 | Instituto Nacional de Enfermedades Respiratorias | Instituto de Diagnostico y Referencia Epidemiologicos (INDRE) | Ramirez-Gonzalez Ernesto et al |
| 447 | England/200990660/2020 | 2020-02-27 | Respiratory Virus Unit, Microbiology Services Colindale, Public Health England | Respiratory Virus Unit, Microbiology Services Colindale, Public Health England | Monica Galiano et al |
| 448 | Norway/1379/2020 | 2020-02-27 | Oslo University Hospital, Department of Medical Microbiology | Norwegian Institute of Public Health | Kathrine Stene-Johansen et al |
| 449 | Switzerland/AG0361/2020 | 2020-02-27 | Laboratoire de Virologie, HUG | Swiss National Reference Centre for Influenza | LAUBSCHER Florian et al. et al |
| 450 | SouthKorea/KUMC04/2020 | 2020-02-27 | Department of Microbiology, Institute for Viral Diseases, College of Medicine, Korea University | Department of Microbiology, Institute for Viral Diseases, College of Medicine, Korea University | Changmin Kang et al |
| 451 | Switzerland/GR2988/2020 | 2020-02-27 | Hôpitaux universitaires de Genève Laboratoire de Virologie | Hôpitaux universitaires de Genève Laboratoire de Virologie | Laubscher F. et al |
| 452 | Switzerland/GE5373/2020 | 2020-02-27 | Laboratoire de Virologie, HUG | Swiss National Reference Centre for Influenza | LAUBSCHER Florian et al. et al |
| 453 | England/200990723/2020 | 2020-02-27 | Respiratory Virus Unit, Microbiology Services Colindale, Public Health England | Respiratory Virus Unit, Microbiology Services Colindale, Public Health England | Monica Galiano et al |
| 454 | USA/CA-CDPH-UC3/2020 | 2020-02-27 | California Department of Public Health | Chiu Laboratory, University of California, San Francisco | Xianding Deng et al |
| 455 | Switzerland/GE9586/2020 | 2020-02-27 | Laboratoire de Virologie, HUG | Swiss National Reference Centre for Influenza | LAUBSCHER Florian et al. et al |
| 456 | Korea/BA-ACH_2604/2020 | 2020-02-27 | Brian D. Allgood Army Community Hospital | Pathogen Discovery, Respiratory Viruses Branch, Division of Viral Diseases, Centers for Disease Control and Prevention | Krista Queen et al |
| 457 | Sweden/20-04631/2020 | 2020-02-27 | HS mikrobiologi virus | The Public Health Agency of Sweden | Zhibing Yun et al |
| 458 | USA/WA_5030/2020 | 2020-02-27 | WA State Department of Health | Pathogen Discovery, Respiratory Viruses Branch, Division of Viral Diseases, Centers for Disease Control and Prevention | Ying Tao et al |
| 459 | Taiwan/CGMH-CGU-04/2020 | 2020-02-27 | Laboratory Medicine | Department of Laboratory Medicine, Lin-Kou Chang Gung Memorial Hospital, Taoyuan, Taiwan | Kuo-Chien Tsao et al |
| 460 | England/200960041/2020 | 2020-02-27 | Respiratory Virus Unit, Microbiology Services Colindale, Public Health England | Respiratory Virus Unit, Microbiology Services Colindale, Public Health England | Monica Galiano et al |
| 461 | SouthKorea/KUMC03/2020 | 2020-02-27 | Division of Infectious Diseases, Department of Internal Medicine, Korea University College of Medicine | Department of Microbiology, Institute for Viral Diseases, College of Medicine, Korea University | Changmin Kang et al |
| 462 | England/200990725/2020 | 2020-02-28 | Respiratory Virus Unit, Microbiology Services Colindale, Public Health England | Respiratory Virus Unit, Microbiology Services Colindale, Public Health England | Monica Galiano et al |
| 463 | Australia/NSW09/2020 | 2020-02-28 | Centre for Infectious Diseases and Microbiology Laboratory Services | NSW Health Pathology - Institute of Clinical Pathology and Medical Research; Westmead Hospital; University of Sydney | Rockett R et al |
| 464 | Australia/NSW08/2020 | 2020-02-28 | Centre for Infectious Diseases and Microbiology Laboratory Services | NSW Health Pathology - Institute of Clinical Pathology and Medical Research; Westmead Hospital; University of Sydney | Rockett R et al |
| 465 | Australia/NSW219/202 | 2020-02-28 | Centre for Infectious Diseases and Microbiology Public Health | NSW Health Pathology - Institute of Clinical Pathology and Medical Research; Westmead Hospital; University of Sydney | Timms et al |

| | | | | | |
|---|---|---|---|---|---|
| 466 | Australia/NSW10/2020 | 2020-02-28 | Centre for Infectious Diseases and Microbiology - Public Health | NSW Health Pathology - Institute of Clinical Pathology and Medical Research; Westmead Hospital; University of Sydney | Rockett R et al |
| 467 | Australia/NSW05/2020 | 2020-02-28 | Centre for Infectious Diseases and Microbiology Laboratory Services | NSW Health Pathology - Institute of Clinical Pathology and Medical Research; Westmead Hospital; University of Sydney | Eden J-S et al |
| 468 | Australia/QLDID922/2020 | 2020-02-28 | Mater Pathology | Public Health Virology Laboratory | Bixing Huang et al |
| 469 | USA/WA-S103/2020 | 2020-02-28 | Washington State Department of Health | Seattle Flu Study | Chu etl al et al |
| 470 | Germany/NRW-09/2020 | 2020-02-28 | Center of Medical Microbiology, Virology, and Hospital Hygiene, University of Duesseldorf | Center of Medical Microbiology, Virology, and Hospital Hygiene, University of Duesseldorf | Ortwin Adams et al |
| 471 | USA/WA-S99/2020 | 2020-02-28 | Washington State Department of Health | Seattle Flu Study | Chu etl al et al |
| 472 | USA/WA-S42/2020 | 2020-02-28 | Washington State Department of Health | Seattle Flu Study | Chu etl al et al |
| 473 | Italy/INMI4/2020 | 2020-02-28 | INMI Lazzaro Spallanzani IRCCS | Laboratory of Virology, INMI Lazzaro Spallanzani IRCCS | Cesare E. M. Gruber et al |
| 474 | USA/WA-S96/2020 | 2020-02-28 | Washington State Department of Health | Seattle Flu Study | Chu etl al et al |
| 475 | USA/RI_0520/2020 | 2020-02-28 | RI State Health Laboratories | Pathogen Discovery, Respiratory Viruses Branch, Division of Viral Diseases, Centers for Disease Control and Prevention | Ying Tao et al |
| 476 | USA/WA-S95/2020 | 2020-02-28 | Washington State Department of Health | Seattle Flu Study | Chu etl al et al |
| 477 | Singapore/12Clin/2020 | 2020-02-28 | National Public Health Laboratory, National Centre for Infectious Diseases | National Public Health Laboratory, National Centre for Infectious Diseases | Mak TM et al |
| 478 | USA/WA-S234/2020 | 2020-02-28 | Washington State Department of Health | Seattle Flu Study | Chu et al et al |
| 479 | USA/WA-S97/2020 | 2020-02-28 | Washington State Department of Health | Seattle Flu Study | Chu etl al et al |
| 480 | USA/WA-S105/2020 | 2020-02-28 | Washington State Department of Health | Seattle Flu Study | Chu etl al et al |
| 481 | Switzerland/GE0199/2020 | 2020-02-28 | Hôpitaux universitaires de Genève Laboratoire de Virologie | Hôpitaux universitaires de Genève Laboratoire de Virologie | Laubscher F. et al |
| 482 | USA/FL_5091/2020 | 2020-02-28 | FL Bureau of Public Health Laboratories-Tampa | Pathogen Discovery, Respiratory Viruses Branch, Division of Viral Diseases, Centers for Disease Control and Prevention | Anna Uehara et al |
| 483 | Brazil/SPBR-02/2020 | 2020-02-28 | Hospital Israelita Albert Einstein | Instituto Adolfo Lutz, Interdisciplinary Procedures Center, Strategic Laboratory | Jaqueline Goes de Jesus et al |
| 484 | USA/WA-S40/2020 | 2020-02-28 | Washington State Department of Health | Seattle Flu Study | Chu etl al et al |
| 485 | USA/CA-SCCPHD-UC108/2020 | 2020-02-29 | Santa Clara County Public Health Department | Chiu Laboratory, University of California, San Francisco | Xianding Deng et al |
| 486 | USA/WA-S10/2020 | 2020-02-29 | Seattle Flu Study | Seattle Flu Study | Chu et al et al |
| 487 | Australia/NSW06/2020 | 2020-02-29 | Centre for Infectious Diseases and Microbiology Laboratory Services | NSW Health Pathology - Institute of Clinical Pathology and Medical Research; Westmead Hospital; University of Sydney | Eden J-S et al |
| 488 | Australia/QLD09/2020 | 2020-02-29 | Pathology Queensland | Public Health Virology Laboratory | Bixing Huang et al |
| 489 | Australia/NSW07/2020 | 2020-02-29 | Centre for Infectious Diseases and Microbiology Laboratory Services | NSW Health Pathology - Institute of Clinical Pathology and Medical Research; Westmead Hospital; University of Sydney | Eden J-S et al |
| 490 | Brazil/SPBR-06/2020 | 2020-02-29 | Hospital São Joaquim Beneficencia Portuguesa | Instituto Adolfo Lutz, Interdiciplinary Procedures | Claudio Tavares Sacchi et al |

| | | | | | |
|---|---|---|---|---|---|
| | | | | Center, Strategic Laboratory | |
| 491 | USA/WA6-UW3/2020 | 2020-02-29 | Washington State Public Health Lab | UW Virology Lab | Pavitra Roychoudhury et al |
| 492 | Switzerland/BE6651/2020 | 2020-02-29 | Hôpitaux universitaires de Genève Laboratoire de Virologie | Hôpitaux universitaires de Genève Laboratoire de Virologie | Laubscher F. et al |
| 493 | USA/WA-S123/2020 | 2020-02-29 | Washington State Department of Health | Seattle Flu Study | Chu et al et al |
| 494 | USA/WA-S119/2020 | 2020-02-29 | Washington State Department of Health | Seattle Flu Study | Chu etl al et al |
| 495 | USA/WA-S89/2020 | 2020-02-29 | Washington State Department of Health | Seattle Flu Study | Chu etl al et al |
| 496 | Switzerland/1000477796/2020 | 2020-02-29 | Division of Infectious Diseases, University Hospital Zurich | Institute of Medical Virology, University of Zurich | Stefan Schmutz et al |
| 497 | Malaysia/IMR_WC1098/2020 | 2020-02-29 | Institute for Medical Research, Infectious Disease Research Centre, National Institutes of Health, Ministry of Health Malaysia | Institute for Medical Research, Infectious Disease Research Centre, National Institutes of Health, Ministry of Health Malaysia | Suppiah.J et al |
| 498 | Korea/BA-ACH_2719/2020 | 2020-02-29 | Brian D. Allgood Army Community Hospital | Pathogen Discovery, Respiratory Viruses Branch, Division of Viral Diseases, Centers for Disease Control and Prevention | Krista Queen et al |
| 499 | USA/WA-S47/2020 | 2020-02-29 | Washington State Department of Health | Seattle Flu Study | Chu etl al et al |
| 500 | USA/WA-S55/2020 | 2020-02-29 | Washington State Department of Health | Seattle Flu Study | Chu etl al et al |
| 501 | Luxembourg/LNS0000001/2020 | 2020-02-29 | Laboratoire National de Santé, Microbiology, Virology | Laboratoire National de Santé, Microbiology, Epidemiology and Microbial Genomics | Anke Wienecke-Baldacchino et al |
| 502 | Switzerland/1000477797/2020 | 2020-02-29 | Division of Infectious Diseases, University Hospital Zurich | Institute of Medical Virology, University of Zurich | Stefan Schmutz et al |
| 503 | USA/TX_2039/2020 | 2020-02-29 | Texas DSHS Lab Services | Pathogen Discovery, Respiratory Viruses Branch, Division of Viral Diseases, Centers for Disease Control and Prevention | Krista Queen et al |
| 504 | Greece/127_HPI/2020 | 2020-02-29 | Hellenic Pasteur Institute, Public Health Laboratories | Hellenic Pasteur Institute, Public Health Laboratories, Unit of Bioinformatics and Applied Genomics | Vasiliki Pogka et al |
| 505 | USA/WA-S100/2020 | 2020-02-29 | Washington State Department of Health | Seattle Flu Study | Chu etl al et al |
| 506 | USA/WA-S243/2020 | 2020-02-29 | Washington State Department of Health | Seattle Flu Study | Chu et al et al |
| 507 | USA/WA-S235/2020 | 2020-02-29 | Washington State Department of Health | Seattle Flu Study | Chu et al et al |
| 508 | Taiwan/CGMH-CGU-21/2020 | 2020-02-29 | Laboratory Medicine | Department of Laboratory Medicine, Lin-Kou Chang Gung Memorial Hospital, Taoyuan, Taiwan | Kuo-Chien Tsao et al |
| 509 | USA/WA-S6/2020 | 2020-02-29 | Seattle Flu Study | Seattle Flu Study | Chu et al et al |
| 510 | USA/WA-S106/2020 | 2020-02-29 | Washington State Department of Health | Seattle Flu Study | Chu etl al et al |
| 511 | USA/WA-S93/2020 | 2020-02-29 | Washington State Department of Health | Seattle Flu Study | Chu etl al et al |
| 512 | USA/NY1-PV08001/2020 | 2020-02-29 | MSHS Clinical Microbiology Laboratories | MSHS Pathogen Surveillance Program | Gopi Patel et al |
| 513 | Belgium/VAG-03013/2020 | 2020-03-01 | KU Leuven, Clinical and Epidemiological Virology | KU Leuven, Clinical and Epidemiological Virology | Bert Vanmechelen et al |
| 514 | Spain/CastillaLaMancha201329/2020 | 2020-03-01 | Hospital General y Universitario de Guadalajara | Instituto de Salud Carlos III | Iglesias-Caballero et al |
| 515 | Singapore/18/2020 | 2020-03-01 | National Public Health Laboratory, National Centre for Infectious Diseases | National Public Health Laboratory, National Centre for Infectious Diseases | Mak TM et al |
| 516 | Iceland/14/2020 | 2020-03-01 | The National University Hospital of Iceland | deCODE genetics | Daniel F Gudbjartsson et al |
| 517 | Iceland/12/2020 | 2020-03-01 | The National University | deCODE genetics | Daniel F Gudbjartsson |

| | | | | | |
|---|---|---|---|---|---|
| | | | Hospital of Iceland | | et al |
| 518 | Switzerland/TI2045/2020 | 2020-03-01 | Hôpitaux universitaires de Genève Laboratoire de Virologie | Hôpitaux universitaires de Genève Laboratoire de Virologie | Laubscher F. et al |
| 519 | Norway/1538/2020 | 2020-03-01 | Hospital of Southern Norway - Kristiansand, Department of Medical Microbiology | Norwegian Institute of Public Health, Department of Virology | Kathrine Stene-Johansen et al |
| 520 | Denmark/SSI-03/2020 | 2020-03-01 | Department of Virus and Microbiological Special diagnostics, Statens Serum Institut, Copenhagen, Denmark. | ViFU | Morten Rasmussen et al |
| 521 | Spain/CastillayLeon201323/2020 | 2020-03-01 | COMPLEJO ASISTENCIAL UNIVERSITARIO DE BURGOS | Instituto de Salud Carlos III | Iglesias-Caballero et al |
| 522 | India/GMC-RK100/2020 | 2020-03-01 | Department of Microbiology,Gandhi Medical College and Hospital | Virus Research Laboratory, Department of Zoology, Osmania University,Hyderabad,India | Muttineni Radhakrishna et al |
| 523 | Belgium/CG-030158/2020 | 2020-03-01 | KU Leuven, Clinical and Epidemiological Virology | KU Leuven, Clinical and Epidemiological Virology | Bert Vanmechelen et al |
| 524 | Belgium/BM-03012/2020 | 2020-03-01 | KU Leuven, Clincal and Epidemiological Virology | KU Leuven, Clincal and Epidemiological Virology | Bert Vanmechelen et al |
| 525 | USA/WA-S86/2020 | 2020-03-01 | Washington State Department of Health | Seattle Flu Study | Chu etl al et al |
| 526 | Belgium/SH-03014/2020 | 2020-03-01 | KU Leuven, Clinical and Epidemiological Virology | KU Leuven, Clinical and Epidemiological Virology | Bert Vanmechelen et al |
| 527 | Spain/CastillaLaMancha201328/2020 | 2020-03-01 | Hospital General y Universitario de Guadalajara | Instituto de Salud Carlos III | Iglesias-Caballero et al |
| 528 | France/B2334/2020 | 2020-03-01 | CHRU Pontchaillou - Laboratoire de Virologie | National Reference Center for Viruses of Respiratory Infections, Institut Pasteur, Paris | Mélnie Albert et al |
| 529 | England/200991076/2020 | 2020-03-01 | Respiratory Virus Unit, Microbiology Services Colindale, Public Health England | Respiratory Virus Unit, Microbiology Services Colindale, Public Health England | Monica Galiano et al |
| 530 | USA/WA-S249/2020 | 2020-03-01 | Washington State Department of Health | Seattle Flu Study | Chu et al et al |
| 531 | Argentina/PAIS_A001/2020 | 2020-03-01 | Laboratorio Análisis Clínicos, Unidad de Servicios Diagnósticos, Swiss Medical Group | Área de Secuenciación del Laboratorio de Virología del Hospital de Niños Dr. Ricardo Gutierrez | Nabaes Jodar et al |
| 532 | Spain/Valencia7/2020 | 2020-03-02 | Servicio de Microbiología. Consorcio Hospital General Universitario de Valencia | Sequencing and Bioinformatics Service and Molecular Epidemiology Research Group. FISABIO-Public Health | Neris Garcia-Gonzalez et al |
| 533 | USA/FL_6318/2020 | 2020-03-02 | FL Bureau of Health Laboratories Tampa | Pathogen Discovery, Respiratory Viruses Branch, Division of Viral Diseases, Centers for Disease Control and Prevention | Krista Queen et al |
| 534 | USA/WA-S29/2020 | 2020-03-02 | Washington State Department of Health | Seattle Flu Study | Chu etl al et al |
| 535 | Germany/BavPat3/2020 | 2020-03-02 | Bundeswehr Institute of Microbiology | Bundeswehr Institute of Microbiology | Mathias C Walter et al |
| 536 | Australia/NSW11/2020 | 2020-03-02 | Centre for Infectious Diseases and Microbiology- Public Health | NSW Health Pathology - Institute of Clinical Pathology and Medical Research; Westmead Hospital; University of Sydney | Lam C et al |
| 537 | Australia/VIC04/2020 | 2020-03-02 | Victorian Infectious Diseases Reference Laboratory (VIDRL) | Victorian Infectious Diseases Reference Laboratory and Microbiological Diagnostic Unit Public Health Laboratory, Doherty Institute | Caly L. et al |
| 538 | Australia/NSW217/2020 | 2020-03-02 | Centre for Infectious Diseases and Microbiology Public Health | NSW Health Pathology - Institute of Clinical Pathology and Medical Research; Westmead Hospital; University of Sydney | Rockett R et al |
| 539 | Singapore/23/2020 | 2020-03-02 | National Centre for Infectious Diseases | Programme in Emerging Infectious Diseases, Duke-NUS | Danielle E Anderson et al |

| | | | | | |
|---|---|---|---|---|---|
| | | | | Medical School | |
| 540 | Spain/PaisVasco201382/2020 | 2020-03-02 | HOSPITAL TXAGORRITXU | Instituto de Salud Carlos III | Iglesias-Caballero et al |
| 541 | NewZealand/20VR0189/2020-03-02 | 2020-03-02 | LabPLUS | Institute of Environmental Science and Research (ESR) Ltd | **Matt Storey et al** |
| 542 | Spain/Madrid_R10_33/2020 | 2020-03-02 | Hospital Universitario Ramón y Cajal | Hospital Universitario La Paz | Elias Dahdouh et al |
| 543 | Netherlands/Zeewolde_1365080/2020 | 2020-03-02 | MHC Flevoland | Erasmus Medical Center | David Nieuwenhuijse et al |
| 544 | Singapore/19/2020 | 2020-03-02 | National Public Health Laboratory, National Centre for Infectious Diseases | National Public Health Laboratory, National Centre for Infectious Diseases | Mak TM et al |
| 545 | Belgium/FR-03029/2020 | 2020-03-02 | KU Leuven, Clinical and Epidemiological Virology | KU Leuven, Clinical and Epidemiological Virology | Joan Marti-Carerras et al |
| 546 | USA/WA-S34/2020 | 2020-03-02 | Washington State Department of Health | Seattle Flu Study | Chu etl al et al |
| 547 | Iceland/23/2020 | 2020-03-02 | The National University Hospital of Iceland | deCODE genetics | Daniel F Gudbjartsson et al |
| 548 | Iceland/20/2020 | 2020-03-02 | The National University Hospital of Iceland | deCODE genetics | Daniel F Gudbjartsson et al |
| 549 | Iceland/22/2020 | 2020-03-02 | The National University Hospital of Iceland | deCODE genetics | Daniel F Gudbjartsson et al |
| 550 | Iceland/15/2020 | 2020-03-02 | The National University Hospital of Iceland | deCODE genetics | Daniel F Gudbjartsson et al |
| 551 | Iceland/19/2020 | 2020-03-02 | The National University Hospital of Iceland | deCODE genetics | Daniel F Gudbjartsson et al |
| 552 | Iceland/25/2020 | 2020-03-02 | The National University Hospital of Iceland | deCODE genetics | Daniel F Gudbjartsson et al |
| 553 | Belgium/MTR-03021/2020 | 2020-03-02 | KU Leuven, Clinical and Epidemiological Virology | KU Leuven, Clinical and Epidemiological Virology | Bert Vanmechelen et al |
| 554 | Belgium/MTR-03026/2020 | 2020-03-02 | KU Leuven, Clinical and Epidemiological Virology | KU Leuven, Clinical and Epidemiological Virology | Bert Vanmechelen et al |
| 555 | Taiwan/NTU03/2020 | 2020-03-02 | Department of Laboratory Medicine, National Taiwan University Hospital | Microbial Genomics Core Lab, National Taiwan University Centers of Genomic and Precision Medicine | Shiou-Hwei Yeh et al |
| 556 | Germany/BavPat2/2020 | 2020-03-02 | Bundeswehr Institute of Microbiology | Bundeswehr Institute of Microbiology | Mathias C Walter et al |
| 557 | Netherlands/Utrecht_5/2020 | 2020-03-02 | Dutch COVID-19 response team | Erasmus Medical Center | David Nieuwenhuijse et al |
| 558 | France/IDF2075/2020 | 2020-03-02 | Hôpital Instruction des Armées - BEGIN | National Reference Center for Viruses of Respiratory Infections, Institut Pasteur, Paris | Mélnie Albert et al |
| 559 | USA/WA-S64/2020 | 2020-03-03 | Washington State Department of Health | Seattle Flu Study | Chu etl al et al |
| 560 | Australia/NSW14/2020 | 2020-03-03 | Centre for Infectious Diseases and Microbiology - Public Health | NSW Health Pathology - Institute of Clinical Pathology and Medical Research; Westmead Hospital; University of Sydney | Gall et al |
| 561 | Australia/NSW22/2020 | 2020-03-03 | Centre for Infectious Diseases and Microbiology Public Health | NSW Health Pathology - Institute of Clinical Pathology and Medical Research; Westmead Hospital; University of Sydney | Timms V et al |
| 562 | Australia/NSW15/2020 | 2020-03-03 | Centre for Infectious Diseases and Microbiology Public Health | NSW Health Pathology - Institute of Clinical Pathology and Medical Research; Westmead Hospital; University of Sydney | Rockett R et al |
| 563 | Spain/Madrid_H12_13/2020 | 2020-03-03 | Hospital Universitario 12 de Octubre | Hospital Universitario 12 de Octubre | Sara González et al |
| 564 | USA/WA-S233/2020 | 2020-03-03 | Washington State Department of Health | Seattle Flu Study | Chu et al et al |
| 565 | USA/NY-Wadsworth-10528-01/2020 | 2020-03-03 | Wadsworth Center, New York State Department.of Health | Wadsworth Center, New York State Department.of Health | Kirsten St. George et al |
| 566 | England/20108007302/2020 | 2020-03-03 | Respiratory Virus Unit, Microbiology Services Colindale, Public Health England | Respiratory Virus Unit, Microbiology Services Colindale, Public Health England | Monica Galiano et al |

| | | | | | |
|---|---|---|---|---|---|
| 567 | Netherlands/NoordHolland_2/2020 | 2020-03-03 | Dutch COVID-19 response team | Erasmus Medical Center | David Nieuwenhuijse et al |
| 568 | England/20104002606/2020 | 2020-03-03 | Respiratory Virus Unit, Microbiology Services Colindale, Public Health England | Respiratory Virus Unit, Microbiology Services Colindale, Public Health England | Monica Galiano et al |
| 569 | England/20106004803/2020 | 2020-03-03 | Respiratory Virus Unit, Microbiology Services Colindale, Public Health England | Respiratory Virus Unit, Microbiology Services Colindale, Public Health England | Monica Galiano et al |
| 570 | France/Valence_425/2020 | 2020-03-03 | CNR Virus des Infections Respiratoires - France SUD | CNR Virus des Infections Respiratoires - France SUD | Bal et al |
| 571 | USA/WA-S57/2020 | 2020-03-03 | Washington State Department of Health | Seattle Flu Study | Chu etl al et al |
| 572 | Iceland/33/2020 | 2020-03-03 | The National University Hospital of Iceland | deCODE genetics | Daniel F Gudbjartsson et al |
| 573 | Iceland/31/2020 | 2020-03-03 | The National University Hospital of Iceland | deCODE genetics | Daniel F Gudbjartsson et al |
| 574 | Iceland/27/2020 | 2020-03-03 | The National University Hospital of Iceland | deCODE genetics | Daniel F Gudbjartsson et al |
| 575 | Iceland/30/2020 | 2020-03-03 | The National University Hospital of Iceland | deCODE genetics | Daniel F Gudbjartsson et al |
| 576 | Denmark/SSI-104/2020 | 2020-03-03 | Department of Virus and Microbiological Special diagnostics, Statens Serum Institut, Copenhagen, Denmark. | ViFU | Morten Rasmussen et al |
| 577 | India/c32/2020 | 2020-03-03 | National Influenza Center, Indian Council of Medical Research - National Institute of Virology | Indian Council of Medical Research-National Institute of Virology, Microbial Containment Complex | Pragya D. Yadav. Savita Patil et al |
| 578 | USA/GA_1299/2020 | 2020-03-03 | GA Department of Public Health | Pathogen Discovery, Respiratory Viruses Branch, Division of Viral Diseases, Centers for Disease Control and Prevention | Krista Queen et al |
| 579 | Netherlands/Tilburg_1364286/2020 | 2020-03-03 | Foundation Elisabeth-Tweesteden Ziekenhuis | Erasmus Medical Center | David Nieuwenhuijse et al |
| 580 | Belgium/SN-03031/2020 | 2020-03-03 | KU Leuven, Clinical and Epidemiological Virology | KU Leuven, Clinical and Epidemiological Virology | Bert Vanmechelen et al |
| 581 | Spain/LaRioja201575/2020 | 2020-03-03 | Hospital San Pedro | Instituto de Salud Carlos III | Iglesias-Caballero et al |
| 582 | France/B2337/2020 | 2020-03-03 | CHRU Pontchaillou - Laboratoire de Virologie | National Reference Center for Viruses of Respiratory Infections, Institut Pasteur, Paris | Mélnie Albert et al |
| 583 | USA/GA_1445/2020 | 2020-03-04 | GA Department of Public Health | Pathogen Discovery, Respiratory Viruses Branch, Division of Viral Diseases, Centers for Disease Control and Prevention | Krista Queen et al |
| 584 | England/20108007002/2020 | 2020-03-04 | Respiratory Virus Unit, Microbiology Services Colindale, Public Health England | Respiratory Virus Unit, Microbiology Services Colindale, Public Health England | Monica Galiano et al |
| 585 | Australia/NSW21/2020 | 2020-03-04 | Centre for Infectious Diseases and Microbiology Public Health | NSW Health Pathology - Institute of Clinical Pathology and Medical Research; Westmead Hospital; University of Sydney | Gray K et al |
| 586 | Australia/NSW13/2020 | 2020-03-04 | Centre for Infectious Diseases and Microbiology - Public Health | NSW Health Pathology - Institute of Clinical Pathology and Medical Research; Westmead Hospital; University of Sydney | Timms et al |
| 587 | Australia/NSW66/2020 | 2020-03-04 | Centre for Infectious Diseases and Microbiology Public Health | NSW Health Pathology - Institute of Clinical Pathology and Medical Research; Westmead Hospital; University of Sydney | Timms V et al |
| 588 | Australia/NSW12/2020 | 2020-03-04 | Centre for Infectious Diseases and Microbiology - Public Health | NSW Health Pathology - Institute of Clinical Pathology and Medical Research; | Gray K et al |

| | | | | | |
|---|---|---|---|---|---|
| | | | | Westmead Hospital; University of Sydney | |
| 589 | Australia/NSW23/2020 | 2020-03-04 | Centre for Infectious Diseases and Microbiology Public Health | NSW Health Pathology - Institute of Clinical Pathology and Medical Research; Westmead Hospital; University of Sydney | Gall M et al |
| 590 | Australia/WA06/2020 | 2020-03-04 | Department of Microbiology, PathWest QEII Medical Centre | Department of Microbiology, PathWest QEII Medical Centre | Chisha Sikazwe et al |
| 591 | USA/NY_1922/2020 | 2020-03-04 | NYC Department of Health and Mental Hygiene | Pathogen Discovery, Respiratory Viruses Branch, Division of Viral Diseases, Centers for Disease Control and Prevention | Ying Tao et al |
| 592 | USA/WA-UW28/2020 | 2020-03-04 | UW Virology Lab | UW Virology Lab | Pavitra Roychoudhury et al |
| 593 | USA/WA-S163/2020 | 2020-03-04 | Washington State Department of Health | Seattle Flu Study | Chu et al et al |
| 594 | USA/WA-S68/2020 | 2020-03-04 | Washington State Department of Health | Seattle Flu Study | Chu etl al et al |
| 595 | NewZealand/20VR0195/2020-03-04 | 2020-03-04 | LabPLUS | Institute of Environmental Science and Research (ESR) Ltd | **Matt Storey et al** |
| 596 | Iceland/40/2020 | 2020-03-04 | The National University Hospital of Iceland | deCODE genetics | Daniel F Gudbjartsson et al |
| 597 | Spain/Canarias201495/2020 | 2020-03-04 | Hospital Universitario de Canarias | Instituto de Salud Carlos III | Iglesias-Caballero et al |
| 598 | Belgium/JL-03044/2020 | 2020-03-04 | KU Leuven, Clinical and Epidemiological Virology | KU Leuven, Clinical and Epidemiological Virology | Tony Wawina et al |
| 599 | Iceland/39/2020 | 2020-03-04 | The National University Hospital of Iceland | deCODE genetics | Daniel F Gudbjartsson et al |
| 600 | Iceland/45/2020 | 2020-03-04 | The National University Hospital of Iceland | deCODE genetics | Daniel F Gudbjartsson et al |
| 601 | Iceland/37/2020 | 2020-03-04 | The National University Hospital of Iceland | deCODE genetics | Daniel F Gudbjartsson et al |
| 602 | Iceland/36/2020 | 2020-03-04 | The National University Hospital of Iceland | deCODE genetics | Daniel F Gudbjartsson et al |
| 603 | USA/NY-Wadsworth-10703-01/2020 | 2020-03-04 | Wadsworth Center, New York State Department.of Health | Wadsworth Center, New York State Department.of Health | Kirsten St. George et al |
| 604 | France/Lyon_487/2020 | 2020-03-04 | Institut des Agents Infectieux (IAI) Hospices Civils de Lyon | CNR Virus des Infections Respiratoires - France SUD | Bal et al |
| 605 | Taiwan/NTU04/2020 | 2020-03-04 | Department of Laboratory Medicine, National Taiwan University Hospital | Microbial Genomics Core Lab, National Taiwan University Centers of Genomic and Precision Medicine | Shiou-Hwei Yeh et al |
| 606 | Belgium/SQ-03043/2020 | 2020-03-04 | KU Leuven, Clinical and Epidemiological Virology | KU Leuven, Clinical and Epidemiological Virology | Joan Marti-Carerras et al |
| 607 | Belgium/BGM-030444/2020 | 2020-03-04 | KU Leuven, Clinical and Epidemiological Virology | KU Leuven, Clinical and Epidemiological Virology | Joan Marti-Carreras et al |
| 608 | Spain/CastillayLeon201437/2020 | 2020-03-04 | COMPLEJO ASISTENCIAL UNIVERSITARIO DE BURGOS | Instituto de Salud Carlos III | Iglesias-Caballero et al |
| 609 | USA/WA16-UW12/2020 | 2020-03-05 | UW Virology Lab | UW Virology Lab | Pavitra Roychoudhury et al |
| 610 | Belgium/DD-030593/2020 | 2020-03-05 | KU Leuven, Clinical and Epidemiological Virology | KU Leuven, Clinical and Epidemiological Virology | Joan Marti-Carreras et al |
| 611 | Spain/Andalucia201617/2020 | 2020-03-05 | Hospital Universitario Virgen de las Nieves | Instituto de Salud Carlos III | Iglesias-Caballero et al |
| 612 | USA/WA17-UW13/2020 | 2020-03-05 | UW Virology Lab | UW Virology Lab | Pavitra Roychoudhury et al |
| 613 | Australia/VIC05/2020 | 2020-03-05 | Victorian Infectious Diseases Reference Laboratory (VIDRL) | Victorian Infectious Diseases Reference Laboratory and Microbiological Diagnostic Unit Public Health Laboratory, Doherty Institute | Caly L. et al |
| 614 | Australia/NSW65/2020 | 2020-03-05 | Centre for Infectious Diseases and Microbiology Public Health | NSW Health Pathology - Institute of Clinical Pathology and Medical Research; Westmead Hospital; University of Sydney | Gray K et al |
| 615 | Australia/NSW25/2020 | 2020-03-05 | Centre for Infectious Diseases | NSW Health Pathology - | Gall M et al |

| | | | | | |
|---|---|---|---|---|---|
| | | | and Microbiology Public Health | Institute of Clinical Pathology and Medical Research; Westmead Hospital; University of Sydney | |
| 616 | Australia/NSW18/2020 | 2020-03-05 | Centre for Infectious Diseases and Microbiology Public Health | NSW Health Pathology - Institute of Clinical Pathology and Medical Research; Westmead Hospital; University of Sydney | Eden J-S et al |
| 617 | Iceland/53/2020 | 2020-03-05 | The National University Hospital of Iceland | deCODE genetics | Daniel F Gudbjartsson et al |
| 618 | USA/WA14-UW10/2020 | 2020-03-05 | UW Virology Lab | UW Virology Lab | Pavitra Roychoudhury et al |
| 619 | USA/SC_3572/2020 | 2020-03-05 | SC Dept of Health and Env. Control-Bureau of Laboratories | Pathogen Discovery, Respiratory Viruses Branch, Division of Viral Diseases, Centers for Disease Control and Prevention | Ying Tao et al |
| 620 | NewZealand/20VR0206/2020-03-05 | 2020-03-05 | LabPLUS | Institute of Environmental Science and Research (ESR) Ltd | **Matt Storey et al** |
| 621 | USA/WA-S73/2020 | 2020-03-05 | Washington State Department of Health | Seattle Flu Study | Chu etl al et al |
| 622 | Singapore/25/2020 | 2020-03-05 | National Centre for Infectious Diseases | Programme in Emerging Infectious Diseases, Duke-NUS Medical School | Danielle E Anderson et al |
| 623 | USA/SC_3520/2020 | 2020-03-05 | SC Dept of Health and Env. Control-Bureau of Laboratories | Pathogen Discovery, Respiratory Viruses Branch, Division of Viral Diseases, Centers for Disease Control and Prevention | Ying Tao et al |
| 624 | USA/NE_6605/2020 | 2020-03-05 | NE Public Health Laboratory | Pathogen Discovery, Respiratory Viruses Branch, Division of Viral Diseases, Centers for Disease Control and Prevention | Yan Li et al |
| 625 | Belgium/BG-030551/2020 | 2020-03-05 | KU Leuven, Clinical and Epidemiological Virology | KU Leuven, Clinical and Epidemiological Virology | Bert Vanmechelen et al |
| 626 | Belgium/MCW-030574/2020 | 2020-03-05 | KU Leuven, Clinical and Epidemiological Virology | KU Leuven, Clinical and Epidemiological Virology | Joan Marti-Carreras et al |
| 627 | France/BFC2147/2020 | 2020-03-05 | | National Reference Center for Viruses of Respiratory Infections, Institut Pasteur, Paris | Mélnie Albert et al |
| 628 | Belgium/JK-0305110/2020 | 2020-03-05 | KU Leuven, Clinical and Epidemiological Virology | KU Leuven, Clinical and Epidemiological Virology | Joan Marti-Carreras et al |
| 629 | Brazil/RJ-352/2020 | 2020-03-05 | Instituto Oswaldo Cruz FIOCRUZ - Laboratory of Respiratory Viruses and Measles (LVRS) | Instituto Oswaldo Cruz FIOCRUZ - Laboratory of Respiratory Viruses and Measles (LVRS) | Paola Resende et al |
| 630 | Slovenia/808/2020 | 2020-03-05 | Institute of Microbiology and Immunology, Faculty of Medicine, University of Ljubljana | Institute of Microbiology and Immunology, Faculty of Medicine, University of Ljubljana | Tomaž Mark Zorec et al |
| 631 | Iceland/47/2020 | 2020-03-05 | The National University Hospital of Iceland | deCODE genetics | Daniel F Gudbjartsson et al |
| 632 | USA/WA15-UW11/2020 | 2020-03-05 | UW Virology Lab | UW Virology Lab | Pavitra Roychoudhury et al |
| 633 | Sweden/20-50087/2020 | 2020-03-05 | Klinisk mikrobiologi Orebro | The Public Health Agency of Sweden | Martin Sundqvist et al |
| 634 | Belgium/PAN-030681/2020 | 2020-03-05 | KU Leuven, Clinical and Epidemiological Virology | KU Leuven, Clinical and Epidemiological Virology | Joan Marti-Carreras et al |
| 635 | Belgium/VS-030542/2020 | 2020-03-05 | KU Leuven, Clinical and Epidemiological Virology | KU Leuven, Clinical and Epidemiological Virology | Bert Vanmechelen et al |
| 636 | USA/WA-UW20/2020 | 2020-03-05 | UW Virology Lab | UW Virology Lab | Pavitra Roychoudhury et al |
| 637 | Scotland/CVR76/2020 | 2020-03-05 | West of Scotland Specialist Virology Centre, NHSGGC / MRC-University of Glasgow Centre for Virus Research | COVID-19 Genomics UK (COG-UK) Consortium | Ana da Silva Filipe et al |
| 638 | Taiwan/CGMH-CGU-06/2020 | 2020-03-05 | Laboratory Medicine | Department of Laboratory Medicine, Lin-Kou Chang Gung Memorial Hospital, Taoyuan, Taiwan | Kuo-Chien Tsao et al |
| 639 | Belgium/VDLH-030548/2020 | 2020-03-05 | KU Leuven, Clinical and | KU Leuven, Clinical and | Bert Vanmechelen et al |

| | | | | | |
|---|---|---|---|---|---|
| | | | Epidemiological Virology | Epidemiological Virology | |
| 640 | Belgium/GS-030549/2020 | 2020-03-05 | KU Leuven, Clinical and Epidemiological Virology | KU Leuven, Clinical and Epidemiological Virology | Bert Vanmechelen et al |
| 641 | USA/GA_1847/2020 | 2020-03-05 | GA Department of Public Health Laboratory | Pathogen Discovery, Respiratory Viruses Branch, Division of Viral Diseases, Centers for Disease Control and Prevention | Yan Li et al |
| 642 | Canada/ON_PHL8580/2020 | 2020-03-05 | Public Health Ontario Laboratories | Public Health Ontario Laboratories | Alireza Eshaghi et al |
| 643 | France/HF2039/2020 | 2020-03-05 | | National Reference Center for Viruses of Respiratory Infections, Institut Pasteur, Paris | Mélnie Albert et al |
| 644 | Yunnan/0306-466/2020 | 2020-03-06 | Department of Clinical Laboratory, the First People's Hospital of Yunnan Province | Department of Clinical Laboratory, the First People's Hospital of Yunnan Province | Yi Sun et al |
| 645 | Austria/CeMM0005/2020 | 2020-03-06 | Center for Virology, Medical University of Vienna | Bergthaler laboratory, CeMM Research Center for Molecular Medicine of the Austrian Academy of Sciences | Alexandra Popa et al |
| 646 | USA/WA-S288/2020 | 2020-03-06 | Seattle Flu Study | Seattle Flu Study | Chu et al et al |
| 647 | Australia/NSW24/2020 | 2020-03-06 | Centre for Infectious Diseases and Microbiology Public Health | NSW Health Pathology - Institute of Clinical Pathology and Medical Research; Westmead Hospital; University of Sydney | Timms V et al |
| 648 | Australia/NSW152/2020 | 2020-03-06 | Centre for Infectious Diseases and Microbiology Public Health | NSW Health Pathology - Institute of Clinical Pathology and Medical Research; Westmead Hospital; University of Sydney | Bachmann N et al |
| 649 | Australia/NSW155/2020 | 2020-03-06 | Centre for Infectious Diseases and Microbiology Public Health | NSW Health Pathology - Institute of Clinical Pathology and Medical Research; Westmead Hospital; University of Sydney | Gray K et al |
| 650 | Australia/NSW19/2020 | 2020-03-06 | Centre for Infectious Diseases and Microbiology Public Health | NSW Health Pathology - Institute of Clinical Pathology and Medical Research; Westmead Hospital; University of Sydney | Lam C et al |
| 651 | Netherlands/ZuidHolland_8/2020 | 2020-03-06 | Dutch COVID-19 response team | Erasmus Medical Center | David Nieuwenhuijse et al |
| 652 | France/B2346/2020 | 2020-03-06 | CHRU Pontchaillou - Laboratoire de Virologie | National Reference Center for Viruses of Respiratory Infections, Institut Pasteur, Paris | Mélnie Albert et al |
| 653 | USA/NH_0029/2020 | 2020-03-06 | NH Dept. of Health and Human Services Public Health Labs | Pathogen Discovery, Respiratory Viruses Branch, Division of Viral Diseases, Centers for Disease Control and Prevention | Yan Li et al |
| 654 | Scotland/CVR75/2020 | 2020-03-06 | West of Scotland Specialist Virology Centre, NHSGGC / MRC-University of Glasgow Centre for Virus Research | COVID-19 Genomics UK (COG-UK) Consortium | Ana da Silva Filipe et al |
| 655 | Australia/NSW33/2020 | 2020-03-06 | Centre for Infectious Diseases and Microbiology Public Health | NSW Health Pathology - Institute of Clinical Pathology and Medical Research; Westmead Hospital; University of Sydney | Sadsad R et al |
| 656 | Belgium/VBK-03061/2020 | 2020-03-06 | KU Leuven, Clinical and Epidemiological Virology | KU Leuven, Clinical and Epidemiological Virology | Joan Marti-Carerras et al |
| 657 | Belgium/PA-030680/2020 | 2020-03-06 | KU Leuven, Clinical and Epidemiological Virology | KU Leuven, Clinical and Epidemiological Virology | Joan Marti-Carreras et al |
| 658 | Belgium/VRAR-030643/2020 | 2020-03-06 | KU Leuven, Clinical and Epidemiological Virology | KU Leuven, Clinical and Epidemiological Virology | Bert Vanmechelen et al |
| 659 | Brazil/BA-510/2020 | 2020-03-06 | LACEN-BA - Laboratório Central de Saúde Pública Professor Gonçalo Moniz | Instituto Oswaldo Cruz FIOCRUZ - Laboratory of Respiratory Viruses and Measles (LVRS) | Paola Resende et al |
| 660 | England/20108034006/2020 | 2020-03-06 | Respiratory Virus Unit, Microbiology Services | Respiratory Virus Unit, Microbiology Services | Monica Galiano et al |

| | | | | | |
|---|---|---|---|---|---|
| | | | Colindale, Public Health England | Colindale, Public Health England | |
| 661 | England/20109099006/2020 | 2020-03-06 | Respiratory Virus Unit, Microbiology Services Colindale, Public Health England | Respiratory Virus Unit, Microbiology Services Colindale, Public Health England | Monica Galiano et al |
| 662 | USA/SC_3569/2020 | 2020-03-06 | SC Dept of Health and Env. Control-Bureau of Laboratories | Pathogen Discovery, Respiratory Viruses Branch, Division of Viral Diseases, Centers for Disease Control and Prevention | Yan Li et al |
| 663 | Iceland/61/2020 | 2020-03-06 | The National University Hospital of Iceland | deCODE genetics | Daniel F Gudbjartsson et al |
| 664 | Iceland/57/2020 | 2020-03-06 | The National University Hospital of Iceland | deCODE genetics | Daniel F Gudbjartsson et al |
| 665 | Iceland/54/2020 | 2020-03-06 | The National University Hospital of Iceland | deCODE genetics | Daniel F Gudbjartsson et al |
| 666 | Iceland/59/2020 | 2020-03-06 | The National University Hospital of Iceland | deCODE genetics | Daniel F Gudbjartsson et al |
| 667 | Iceland/56/2020 | 2020-03-06 | The National University Hospital of Iceland | deCODE genetics | Daniel F Gudbjartsson et al |
| 668 | Iceland/62/2020 | 2020-03-06 | The National University Hospital of Iceland | deCODE genetics | Daniel F Gudbjartsson et al |
| 669 | Belgium/LM-030685/2020 | 2020-03-06 | KU Leuven, Clinical and Epidemiological Virology | KU Leuven, Clinical and Epidemiological Virology | Joan Marti-Carreras et al |
| 670 | Ireland/21023/2020 | 2020-03-06 | UCD National Virus Reference Laboratory | UCD National Virus Reference Laboratory | Michael Carr et al |
| 671 | Belgium/RR-030699/2020 | 2020-03-06 | KU Leuven, Clinical and Epidemiological Virology | KU Leuven, Clinical and Epidemiological Virology | Joan Marti-Carreras et al |
| 672 | Belgium/RYR-030649/2020 | 2020-03-06 | KU Leuven, Clinical and Epidemiological Virology | KU Leuven, Clinical and Epidemiological Virology | Joan Marti-Carreras et al |
| 673 | USA/WA-UW39/2020 | 2020-03-06 | UW Virology Lab | UW Virology Lab | Pavitra Roychoudhury et al |
| 674 | USA/NY-Wadsworth-11379-01/2020 | 2020-03-06 | Wadsworth Center, New York State Department.of Health | Wadsworth Center, New York State Department.of Health | Kirsten St. George et al |
| 675 | USA/FL_9656/2020 | 2020-03-06 | FL Bur. of Public Health Laboratories-Jacksonville | Pathogen Discovery, Respiratory Viruses Branch, Division of Viral Diseases, Centers for Disease Control and Prevention | Yan Li et al |
| 676 | USA/MO_4470/2020 | 2020-03-06 | MO State Public Health Laboratory | Pathogen Discovery, Respiratory Viruses Branch, Division of Viral Diseases, Centers for Disease Control and Prevention | Yan Li et al |
| 677 | USA/KS_4126/2020 | 2020-03-06 | KS Health and Environmental Laboratories | Pathogen Discovery, Respiratory Viruses Branch, Division of Viral Diseases, Centers for Disease Control and Prevention | Yan Li et al |
| 678 | Switzerland/GE06207/2020 | 2020-03-06 | Hôpitaux universitaires de Genève Laboratoire de Virologie | Hôpitaux universitaires de Genève Laboratoire de Virologie | Laubscher F. et al |
| 679 | Belgium/WAM-030676/2020 | 2020-03-06 | KU Leuven, Clinical and Epidemiological Virology | KU Leuven, Clinical and Epidemiological Virology | Joan Marti-Carreras et al |
| 680 | Belgium/MJP-0306100/2020 | 2020-03-06 | KU Leuven, Clinical and Epidemiological Virology | KU Leuven, Clinical and Epidemiological Virology | Joan Marti-Carreras et al |
| 681 | Belgium/VMC-030695/2020 | 2020-03-06 | KU Leuven, Clinical and Epidemiological Virology | KU Leuven, Clinical and Epidemiological Virology | Joan Marti-Carreras et al |
| 682 | USA/SC_3575/2020 | 2020-03-07 | SC Dept of Health and Env. Control-Bureau of Laboratories | Pathogen Discovery, Respiratory Viruses Branch, Division of Viral Diseases, Centers for Disease Control and Prevention | Yan Li et al |
| 683 | Australia/VIC-CBA4/2020 | 2020-03-07 | Geelong Centre for Emerging Infectious Diseases | Geelong Centre for Emerging Infectious Diseases | Chamings A. et al |
| 684 | Australia/VIC18/2020 | 2020-03-07 | Victorian Infectious Diseases Reference Laboratory (VIDRL) | Victorian Infectious Diseases Reference Laboratory and Microbiological Diagnostic Unit Public Health Laboratory, Doherty Institute | Caly L. et al |
| 685 | Australia/VIC17/2020 | 2020-03-07 | Victorian Infectious Diseases Reference Laboratory (VIDRL) | Victorian Infectious Diseases Reference Laboratory and Microbiological Diagnostic Unit | Caly L. et al |

| | | | | | |
|---|---|---|---|---|---|
| | | | | Public Health Laboratory, Doherty Institute | |
| 686 | France/B2348/2020 | 2020-03-07 | CHRU Pontchaillou - Laboratoire de Virologie | National Reference Center for Viruses of Respiratory Infections, Institut Pasteur, Paris | Mélnie Albert et al |
| 687 | USA/WA-S253/2020 | 2020-03-07 | Washington State Department of Health | Seattle Flu Study | Chu et al et al |
| 688 | Italy/INMI8/2020 | 2020-03-07 | INMI Lazzaro Spallanzani IRCCS | Laboratory of Virology, INMI Lazzaro Spallanzani IRCCS | Concetta Castilletti et al |
| 689 | France/Macon_06756/2020 | 2020-03-07 | Centre Hospitalier de Macon | CNR Virus des Infections Respiratoires - France SUD | Antonin Bal et al |
| 690 | Spain/Madrid201706/2020 | 2020-03-07 | Fundacion Jimenez Diaz | Instituto de Salud Carlos III | Iglesias-Caballero et al |
| 691 | Scotland/CVR74/2020 | 2020-03-07 | West of Scotland Specialist Virology Centre, NHSGGC / MRC-University of Glasgow Centre for Virus Research | COVID-19 Genomics UK (COG-UK) Consortium | Ana da Silva Filipe et al |
| 692 | USA/PA_2734/2020 | 2020-03-07 | PA Department of Health, Bureau of Laboratories | Pathogen Discovery, Respiratory Viruses Branch, Division of Viral Diseases, Centers for Disease Control and Prevention | Krista Queen et al |
| 693 | USA/WA-UW34/2020 | 2020-03-07 | UW Virology Lab | UW Virology Lab | Pavitra Roychoudhury et al |
| 694 | Iceland/65/2020 | 2020-03-07 | The National University Hospital of Iceland | deCODE genetics | Daniel F Gudbjartsson et al |
| 695 | Iceland/66/2020 | 2020-03-07 | The National University Hospital of Iceland | deCODE genetics | Daniel F Gudbjartsson et al |
| 696 | Iceland/63/2020 | 2020-03-07 | The National University Hospital of Iceland | deCODE genetics | Daniel F Gudbjartsson et al |
| 697 | SaudiArabia/KAIMRC-Alghoribi/2020 | 2020-03-07 | Clinical Microbiology Lab | Infectious Disease Research Department, King Abdullah International Medical Research Center (KAIMRC) | Majed Alghoribi et al |
| 698 | Portugal/PT0007/2020 | 2020-03-07 | CHULC - H D Estefania | Instituto Nacional de Saude (INSA) | Guiomar et al et al |
| 699 | USA/WA-UW38/2020 | 2020-03-07 | UW Virology Lab | UW Virology Lab | Pavitra Roychoudhury et al |
| 700 | Netherlands/NA_43/2020 | 2020-03-07 | Dutch COVID-19 response team | Erasmus Medical Center | Bas Oude Munnink et al |
| 701 | Canada/BC_8718874/2020 | 2020-03-07 | BCCDC Public Health Laboratory | BCCDC Public Health Laboratory | Harrigan et al |
| 702 | USA/PA_2317/2020 | 2020-03-07 | PA Department of Health, Bureau of Laboratories | Pathogen Discovery, Respiratory Viruses Branch, Division of Viral Diseases, Centers for Disease Control and Prevention | Yan Li et al |
| 703 | Netherlands/ZuidHolland_17/2020 | 2020-03-07 | Dutch COVID-19 response team | Erasmus Medical Center | David Nieuwenhuijse et al |
| 704 | USA/VA_6389/2020 | 2020-03-08 | VA-Division of Consolidated Laboratory Services | Pathogen Discovery, Respiratory Viruses Branch, Division of Viral Diseases, Centers for Disease Control and Prevention | Yan Li et al |
| 705 | Australia/NSW35/2020 | 2020-03-08 | Centre for Infectious Diseases and Microbiology Public Health | NSW Health Pathology - Institute of Clinical Pathology and Medical Research; Westmead Hospital; University of Sydney | Holmes EC et al |
| 706 | Australia/NSW27/2020 | 2020-03-08 | Centre for Infectious Diseases and Microbiology Public Health | NSW Health Pathology - Institute of Clinical Pathology and Medical Research; Westmead Hospital; University of Sydney | Sadsad R et al |
| 707 | Australia/NSW29/2020 | 2020-03-08 | Centre for Infectious Diseases and Microbiology Public Health | NSW Health Pathology - Institute of Clinical Pathology and Medical Research; Westmead Hospital; University of Sydney | Arnott A et al |
| 708 | Australia/NSW153/2020 | 2020-03-08 | Centre for Infectious Diseases and Microbiology Public Health | NSW Health Pathology - Institute of Clinical Pathology and Medical Research; Westmead Hospital; University of Sydney | Rockett R et al |

| | | | | | |
|---|---|---|---|---|---|
| 709 | USA/OH_0023/2020 | 2020-03-08 | OH Department of Health Laboratory | Pathogen Discovery, Respiratory Viruses Branch, Division of Viral Diseases, Centers for Disease Control and Prevention | Yan Li et al |
| 710 | USA/WA-UW41/2020 | 2020-03-08 | UW Virology Lab | UW Virology Lab | Pavitra Roychoudhury et al |
| 711 | Sweden/20-50179/2020 | 2020-03-08 | Klinisk mikrobiologi Orebro | The Public Health Agency of Sweden | Martin Sundqvist et al |
| 712 | USA/PA_2887/2020 | 2020-03-08 | PA Department of Health, Bureau of Laboratories | Pathogen Discovery, Respiratory Viruses Branch, Division of Viral Diseases, Centers for Disease Control and Prevention | Krista Queen et al |
| 713 | Australia/VIC20/2020 | 2020-03-08 | Victorian Infectious Diseases Reference Laboratory (VIDRL) | Victorian Infectious Diseases Reference Laboratory and Microbiological Diagnostic Unit Public Health Laboratory, Doherty Institute | Caly L. et al |
| 714 | Australia/NSW28/2020 | 2020-03-08 | Centre for Infectious Diseases and Microbiology Public Health | NSW Health Pathology - Institute of Clinical Pathology and Medical Research; Westmead Hospital; University of Sydney | Carter I et al |
| 715 | USA/IA_6401/2020 | 2020-03-08 | IA State Hygienic Laboratory | Pathogen Discovery, Respiratory Viruses Branch, Division of Viral Diseases, Centers for Disease Control and Prevention | Ying Tao et al |
| 716 | Denmark/ALAB-SSI405/2020 | 2020-03-08 | Department of Virus and Microbiological Special Diagnostics, Statens Serum Institut, Copenhagen, Denmark, Artillerivej 5, 2300 Copenahgen S | Albertsen lab, Department of Chemistry and Bioscience, Aalborg University, Denmark | Rasmus Kirkegaard et al |
| 717 | Sweden/20-50175/2020 | 2020-03-08 | Klinisk mikrobiologi Orebro | The Public Health Agency of Sweden | Martin Sundqvist et al |
| 718 | Denmark/ALAB-SSI121/2020 | 2020-03-08 | Department of Virus and Microbiological Special Diagnostics, Statens Serum Institut, Copenhagen, Denmark, Artillerivej 5, 2300 Copenahgen S | Albertsen lab, Department of Chemistry and Bioscience, Aalborg University, Denmark | Rasmus Kirkegaard et al |
| 719 | England/20109099106/2020 | 2020-03-08 | Respiratory Virus Unit, Microbiology Services Colindale, Public Health England | Respiratory Virus Unit, Microbiology Services Colindale, Public Health England | Monica Galiano et al |
| 720 | USA/CT_9849/2020 | 2020-03-08 | CT-Dr. Katherine A. Kelley State Public Health Lab | Pathogen Discovery, Respiratory Viruses Branch, Division of Viral Diseases, Centers for Disease Control and Prevention | Anna Uehara et al |
| 721 | Ireland/Dublin-22361/2020 | 2020-03-08 | UCD National Virus Reference Laboratory | UCD National Virus Reference Laboratory | Michael Carr et al |
| 722 | Iceland/69/2020 | 2020-03-08 | The National University Hospital of Iceland | deCODE genetics | Daniel F Gudbjartsson et al |
| 723 | Iceland/70/2020 | 2020-03-08 | The National University Hospital of Iceland | deCODE genetics | Daniel F Gudbjartsson et al |
| 724 | Switzerland/GE6679/2020 | 2020-03-08 | Hôpitaux universitaires de Genève Laboratoire de Virologie | Hôpitaux universitaires de Genève Laboratoire de Virologie | Laubscher F. et al |
| 725 | Canada/ON_PHL6883/2020 | 2020-03-08 | Public Health Ontario Laboratories | Public Health Ontario Laboratories | Alireza Eshaghi et al |
| 726 | USA/WA-S112/2020 | 2020-03-08 | Seattle Flu Study | Seattle Flu Study | Chu etl al et al |
| 727 | USA/NY-Wadsworth-11736-02/2020 | 2020-03-08 | Wadsworth Center, New York State Department.of Health | Wadsworth Center, New York State Department.of Health | Kirsten St. George et al |
| 728 | Netherlands/NA_58/2020 | 2020-03-08 | Dutch COVID-19 response team | Erasmus Medical Center | Bas Oude Munnink et al |
| 729 | USA/IN_82003/2020 | 2020-03-08 | IN State Department of Health Laboratory Services | Pathogen Discovery, Respiratory Viruses Branch, Division of Viral Diseases, Centers for Disease Control and Prevention | Krista Queen et al |
| 730 | Denmark/ALAB-SSI123/2020 | 2020-03-08 | Department of Virus and Microbiological Special Diagnostics, Statens Serum | Albertsen lab, Department of Chemistry and Bioscience, Aalborg University, Denmark | Rasmus Kirkegaard et al |

| | | | | | |
|---|---|---|---|---|---|
| | | | Institut, Copenhagen, Denmark, Artillerivej 5, 2300 Copenahgen S | | |
| 731 | Netherlands/NA_21/2020 | 2020-03-08 | Dutch COVID-19 response team | Erasmus Medical Center | David Nieuwenhuijse et al |
| 732 | USA/PA_2937/2020 | 2020-03-08 | PA Department of Health, Bureau of Laboratories | Pathogen Discovery, Respiratory Viruses Branch, Division of Viral Diseases, Centers for Disease Control and Prevention | Yan Li et al |
| 733 | Netherlands/NA_50/2020 | 2020-03-08 | Dutch COVID-19 response team | Erasmus Medical Center | Bas Oude Munnink et al |
| 734 | France/Lyon_06820/2020 | 2020-03-08 | Institut des Agents Infectieux (IAI), Hospices Civils de Lyon | CNR Virus des Infections Respiratoires - France SUD | Antonin Bal et al |
| 735 | Denmark/ALAB-SSI128/2020 | 2020-03-08 | Department of Virus and Microbiological Special Diagnostics, Statens Serum Institut, Copenhagen, Denmark, Artillerivej 5, 2300 Copenahgen S | Albertsen lab, Department of Chemistry and Bioscience, Aalborg University, Denmark | Rasmus Kirkegaard et al |
| 736 | Netherlands/NoordBrabant_75/2020 | 2020-03-09 | Dutch COVID-19 response team | Erasmus Medical Center | Bas Oude Munnink et al |
| 737 | Spain/Madrid_LP16_6193/2020 | 2020-03-09 | Hospital Universitario La Paz | Hospital Universitario 12 de Octubre | Elias Dahdouh et al |
| 738 | Australia/NSW154/2020 | 2020-03-09 | Centre for Infectious Diseases and Microbiology Public Health | NSW Health Pathology - Institute of Clinical Pathology and Medical Research; Westmead Hospital; University of Sydney | Lam C et al |
| 739 | Australia/VIC22/2020 | 2020-03-09 | Victorian Infectious Diseases Reference Laboratory (VIDRL) | Victorian Infectious Diseases Reference Laboratory and Microbiological Diagnostic Unit Public Health Laboratory, Doherty Institute | Caly L. et al |
| 740 | Australia/VIC21/2020 | 2020-03-09 | Victorian Infectious Diseases Reference Laboratory (VIDRL) | Victorian Infectious Diseases Reference Laboratory and Microbiological Diagnostic Unit Public Health Laboratory, Doherty Institute | Caly L. et al |
| 741 | Australia/NSW26/2020 | 2020-03-09 | Centre for Infectious Diseases and Microbiology Public Health | NSW Health Pathology - Institute of Clinical Pathology and Medical Research; Westmead Hospital; University of Sydney | Arnott A et al |
| 742 | Australia/VIC23/2020 | 2020-03-09 | Victorian Infectious Diseases Reference Laboratory (VIDRL) | Victorian Infectious Diseases Reference Laboratory and Microbiological Diagnostic Unit Public Health Laboratory, Doherty Institute | Caly L. et al |
| 743 | Taiwan/CGMH-CGU-07/2020 | 2020-03-09 | Laboratory Medicine | Department of Laboratory Medicine, Lin-Kou Chang Gung Memorial Hospital, Taoyuan, Taiwan | Kuo-Chien Tsao et al |
| 744 | USA/TX-HMH044/2020 | 2020-03-09 | Houston Methodist Hospital | Houston Methodist Hospital | S. Wesley Long et al |
| 745 | Spain/Valencia33/2020 | 2020-03-09 | Servicio de Microbiología. Consorcio Hospital General Universitario de Valencia | Sequencing and Bioinformatics Service and Molecular Epidemiology Research Group. FISABIO-Public Health | Inma Galán Vendrell et al |
| 746 | USA/WA-UW54/2020 | 2020-03-09 | UW Virology Lab | UW Virology Lab | Pavitra Roychoudhury et al |
| 747 | Canada/SK_4/2020 | 2020-03-09 | Roy Romanow Provincial Laboratory | National Microbiology Laboratory | Majer et al |
| 748 | Denmark/ALAB-SSI173/2020 | 2020-03-09 | Department of Virus and Microbiological Special Diagnostics, Statens Serum Institut, Copenhagen, Denmark, Artillerivej 5, 2300 Copenahgen S | Albertsen lab, Department of Chemistry and Bioscience, Aalborg University, Denmark | Rasmus Kirkegaard et al |
| 749 | Denmark/ALAB-SSI143/2020 | 2020-03-09 | Department of Virus and Microbiological Special Diagnostics, Statens Serum Institut, Copenhagen, | Albertsen lab, Department of Chemistry and Bioscience, Aalborg University, Denmark | Rasmus Kirkegaard et al |

| | | | | | |
|---|---|---|---|---|---|
| | | | Denmark, Artillerivej 5, 2300 Copenahgen S | | |
| 750 | Norway/2084/2020 | 2020-03-09 | Oslo University Hospital, Department of Medical Microbiology | Norwegian Institute of Public Health, Department of Virology | Kathrine Stene-Johansen et al |
| 751 | Iceland/83/2020 | 2020-03-09 | The National University Hospital of Iceland | deCODE genetics | Daniel F Gudbjartsson et al |
| 752 | Netherlands/NoordBrabant_52/2020 | 2020-03-09 | Dutch COVID-19 response team | Erasmus Medical Center | David Nieuwenhuijse et al |
| 753 | USA/WA-UW55/2020 | 2020-03-09 | UW Virology Lab | UW Virology Lab | Pavitra Roychoudhury et al |
| 754 | Spain/Madrid_LP12_21/2020 | 2020-03-09 | Hospital Universitario La Paz | Hospital Universitario La Paz | Elias Dahdouh et al |
| 755 | Iceland/80/2020 | 2020-03-09 | The National University Hospital of Iceland | deCODE genetics | Daniel F Gudbjartsson et al |
| 756 | Iceland/73/2020 | 2020-03-09 | The National University Hospital of Iceland | deCODE genetics | Daniel F Gudbjartsson et al |
| 757 | Iceland/82/2020 | 2020-03-09 | The National University Hospital of Iceland | deCODE genetics | Daniel F Gudbjartsson et al |
| 758 | Iceland/78/2020 | 2020-03-09 | The National University Hospital of Iceland | deCODE genetics | Daniel F Gudbjartsson et al |
| 759 | Iceland/81/2020 | 2020-03-09 | The National University Hospital of Iceland | deCODE genetics | Daniel F Gudbjartsson et al |
| 760 | Iceland/77/2020 | 2020-03-09 | The National University Hospital of Iceland | deCODE genetics | Daniel F Gudbjartsson et al |
| 761 | Canada/ON_PHL0539/2020 | 2020-03-09 | Public Health Ontario Laboratories | Public Health Ontario Laboratories | Alireza Eshaghi et al |
| 762 | Netherlands/NA_62/2020 | 2020-03-09 | Dutch COVID-19 response team | Erasmus Medical Center | Bas Oude Munnink et al |
| 763 | Canada/ON_PHL3877/2020 | 2020-03-09 | Public Health Ontario Laboratories | Public Health Ontario Laboratories | Alireza Eshaghi et al |
| 764 | Denmark/ALAB-SSI160/2020 | 2020-03-09 | Department of Virus and Microbiological Special Diagnostics, Statens Serum Institut, Copenhagen, Denmark, Artillerivej 5, 2300 Copenahgen S | Albertsen lab, Department of Chemistry and Bioscience, Aalborg University, Denmark | Rasmus Kirkegaard et al |
| 765 | USA/WA-UW47/2020 | 2020-03-09 | UW Virology Lab | UW Virology Lab | Pavitra Roychoudhury et al |
| 766 | Netherlands/NA_78/2020 | 2020-03-09 | Dutch COVID-19 response team | Erasmus Medical Center | Bas Oude Munnink et al |
| 767 | USA/WA-S280/2020 | 2020-03-09 | Seattle Flu Study | Seattle Flu Study | Chu et al et al |
| 768 | Netherlands/NoordBrabant_104/2020 | 2020-03-09 | Dutch COVID-19 response team | Erasmus Medical Center | Bas Oude Munnink et al |
| 769 | Denmark/ALAB-HH01/2020 | 2020-03-10 | Department of Clinical Microbiology, Copenhagen University Hospital, Hvidovre, Kettegaard Alle 30, 2650 Hvidovre. | Albertsen lab, Department of Chemistry and Bioscience, Aalborg University, Denmark | Rasmus Kirkegaard et al |
| 770 | Australia/NSW37/2020 | 2020-03-10 | Centre for Infectious Diseases and Microbiology Public Health | NSW Health Pathology - Institute of Clinical Pathology and Medical Research; Westmead Hospital; University of Sydney | Sintchenko V et al |
| 771 | Australia/VIC30/2020 | 2020-03-10 | Victorian Infectious Diseases Reference Laboratory (VIDRL) | Victorian Infectious Diseases Reference Laboratory and Microbiological Diagnostic Unit Public Health Laboratory, Doherty Institute | Caly L. et al |
| 772 | Australia/VIC36/2020 | 2020-03-10 | Victorian Infectious Diseases Reference Laboratory (VIDRL) | Victorian Infectious Diseases Reference Laboratory and Microbiological Diagnostic Unit Public Health Laboratory, Doherty Institute | Caly L. et al |
| 773 | Australia/NSW44/2020 | 2020-03-10 | Centre for Infectious Diseases and Microbiology Public Health | NSW Health Pathology - Institute of Clinical Pathology and Medical Research; Westmead Hospital; University of Sydney | Rockett R et al |
| 774 | Australia/NSW31/2020 | 2020-03-10 | Centre for Infectious Diseases and Microbiology Public | NSW Health Pathology - Institute of Clinical Pathology | Rahman H et al |

| | | | | | |
|---|---|---|---|---|---|
| | | | Health | and Medical Research; Westmead Hospital; University of Sydney | |
| 775 | Australia/VIC34/2020 | 2020-03-10 | Victorian Infectious Diseases Reference Laboratory (VIDRL) | Victorian Infectious Diseases Reference Laboratory and Microbiological Diagnostic Unit Public Health Laboratory, Doherty Institute | Caly L. et al |
| 776 | Fuyang/FY002/2020 | 2020-03-10 | | Clinical Laboratory | Ge et al |
| 777 | France/IDF2284/2020 | 2020-03-10 | Hopital franco britannique - Service des Urgences | National Reference Center for Viruses of Respiratory Infections, Institut Pasteur, Paris | Mélnie Albert et al |
| 778 | Australia/VIC29/2020 | 2020-03-10 | Victorian Infectious Diseases Reference Laboratory (VIDRL) | Victorian Infectious Diseases Reference Laboratory and Microbiological Diagnostic Unit Public Health Laboratory, Doherty Institute | Caly L. et al |
| 779 | USA/NY-SURV0863/2020 | 2020-03-10 | NYC Department of Health and Mental Hygiene | Pathogen Discovery, Respiratory Viruses Branch, Division of Viral Diseases, Centers for Disease Control and Prevention | Ying Tao et al |
| 780 | Denmark/ALAB-HH13/2020 | 2020-03-10 | Department of Clinical Microbiology, Copenhagen University Hospital, Hvidovre, Kettegaard Alle 30, 2650 Hvidovre. | Albertsen lab, Department of Chemistry and Bioscience, Aalborg University, Denmark | Rasmus Kirkegaard et al |
| 781 | Denmark/ALAB-SSI298/2020 | 2020-03-10 | Department of Virus and Microbiological Special Diagnostics, Statens Serum Institut, Copenhagen, Denmark, Artillerivej 5, 2300 Copenahgen S | Albertsen lab, Department of Chemistry and Bioscience, Aalborg University, Denmark | Rasmus Kirkegaard et al |
| 782 | Spain/Valencia55/2020 | 2020-03-10 | Servicio de Microbiología. Consorcio Hospital General Universitario de Valencia | Sequencing and Bioinformatics Service and Molecular Epidemiology Research Group. FISABIO-Public Health | David Navarro et al |
| 783 | Netherlands/Gelderland_5/2020 | 2020-03-10 | Dutch COVID-19 response team | Erasmus Medical Center | Bas Oude Munnink et al |
| 784 | Canada/BC_3972884/2020 | 2020-03-10 | BCCDC Public Health Laboratory | BCCDC Public Health Laboratory | Harrigan et al |
| 785 | Japan/P2-1/2020 | 2020-03-10 | Chiba Prefectural Institute of Public Health | Pathogen Genomics Center, National Institute of Infectious Diseases | Tsuyoshi Sekizuka et al |
| 786 | USA/WA-UW95/2020 | 2020-03-10 | UW Virology Lab | UW Virology Lab | Pavitra Roychoudhury et al |
| 787 | Denmark/ALAB-HH03/2020 | 2020-03-10 | Department of Clinical Microbiology, Copenhagen University Hospital, Hvidovre, Kettegaard Alle 30, 2650 Hvidovre. | Albertsen lab, Department of Chemistry and Bioscience, Aalborg University, Denmark | Rasmus Kirkegaard et al |
| 788 | Australia/NSW51/2020 | 2020-03-10 | Centre for Infectious Diseases and Microbiology Public Health | NSW Health Pathology - Institute of Clinical Pathology and Medical Research; Westmead Hospital; University of Sydney | Rockett R et al |
| 789 | Australia/VIC33/2020 | 2020-03-10 | Victorian Infectious Diseases Reference Laboratory (VIDRL) | Victorian Infectious Diseases Reference Laboratory and Microbiological Diagnostic Unit Public Health Laboratory, Doherty Institute | Caly L. et al |
| 790 | Australia/NSW49/2020 | 2020-03-10 | Centre for Infectious Diseases and Microbiology Public Health | NSW Health Pathology - Institute of Clinical Pathology and Medical Research; Westmead Hospital; University of Sydney | Bachmann N et al |
| 791 | Australia/NSW41/2020 | 2020-03-10 | Centre for Infectious Diseases and Microbiology Public Health | NSW Health Pathology - Institute of Clinical Pathology and Medical Research; Westmead Hospital; University of Sydney | Dwyer DE et al |
| 792 | Iceland/103/2020 | 2020-03-10 | The National University Hospital of Iceland | deCODE genetics | Daniel F Gudbjartsson et al |

| | | | | | |
|---|---|---|---|---|---|
| 793 | USA/WA-UW112/2020 | 2020-03-10 | UW Virology Lab | UW Virology Lab | Pavitra Roychoudhury et al |
| 794 | Norway/1953/2020 | 2020-03-10 | Furst Medical Laboratory | Norwegian Institute of Public Health, Department of Virology | Kathrine Stene-Johansen et al |
| 795 | Denmark/ALAB-SSI263/2020 | 2020-03-10 | Department of Virus and Microbiological Special Diagnostics, Statens Serum Institut, Copenhagen, Denmark, Artillerivej 5, 2300 Copenahgen S | Albertsen lab, Department of Chemistry and Bioscience, Aalborg University, Denmark | Rasmus Kirkegaard et al |
| 796 | Spain/Valencia17/2020 | 2020-03-10 | Servicio de Microbiología. Consorcio Hospital General Universitario de Valencia | Sequencing and Bioinformatics Service and Molecular Epidemiology Research Group. FISABIO-Public Health | Maria Dolores Ocete et al |
| 797 | England/20122074602/2020 | 2020-03-10 | Respiratory Virus Unit, Microbiology Services Colindale, Public Health England | Respiratory Virus Unit, Microbiology Services Colindale, Public Health England | Monica Galiano et al |
| 798 | USA/MN5-MDH5/2020 | 2020-03-10 | Minnesota Department of Health, Public Health Laboratory | Minnesota Department of Health, Public Health Laboratory | Matt Plumb et al |
| 799 | Iceland/93/2020 | 2020-03-10 | The National University Hospital of Iceland | deCODE genetics | Daniel F Gudbjartsson et al |
| 800 | Iceland/90/2020 | 2020-03-10 | The National University Hospital of Iceland | deCODE genetics | Daniel F Gudbjartsson et al |
| 801 | Iceland/94/2020 | 2020-03-10 | The National University Hospital of Iceland | deCODE genetics | Daniel F Gudbjartsson et al |
| 802 | Iceland/96/2020 | 2020-03-10 | The National University Hospital of Iceland | deCODE genetics | Daniel F Gudbjartsson et al |
| 803 | Iceland/84/2020 | 2020-03-10 | The National University Hospital of Iceland | deCODE genetics | Daniel F Gudbjartsson et al |
| 804 | Iceland/89/2020 | 2020-03-10 | The National University Hospital of Iceland | deCODE genetics | Daniel F Gudbjartsson et al |
| 805 | Iceland/97/2020 | 2020-03-10 | The National University Hospital of Iceland | deCODE genetics | Daniel F Gudbjartsson et al |
| 806 | Iceland/102/2020 | 2020-03-10 | The National University Hospital of Iceland | deCODE genetics | Daniel F Gudbjartsson et al |
| 807 | Iceland/101/2020 | 2020-03-10 | The National University Hospital of Iceland | deCODE genetics | Daniel F Gudbjartsson et al |
| 808 | Denmark/ALAB-HH11/2020 | 2020-03-10 | Department of Clinical Microbiology, Copenhagen University Hospital, Hvidovre, Kettegaard Alle 30, 2650 Hvidovre. | Albertsen lab, Department of Chemistry and Bioscience, Aalborg University, Denmark | Rasmus Kirkegaard et al |
| 809 | USA/VA-DCLS-0012/2020 | 2020-03-10 | Virginia DCLS | Virginia DCLS | Virginia DCLS et al |
| 810 | Scotland/CVR10/2020 | 2020-03-10 | West of Scotland Specialist Virology Centre, NHSGGC | MRC-University of Glasgow Centre for Virus Research | Kathy Smollett et al |
| 811 | Sweden/20-50234/2020 | 2020-03-10 | Klinisk mikrobiologi Orebro | The Public Health Agency of Sweden | Martin Sundqvist et al |
| 812 | Norway/1917/2020 | 2020-03-10 | Unilabs Laboratory Medicine | Norwegian Institute of Public Health, Department of Virology | Kathrine Stene-Johansen et al |
| 813 | Australia/VIC31/2020 | 2020-03-10 | Victorian Infectious Diseases Reference Laboratory (VIDRL) | Victorian Infectious Diseases Reference Laboratory and Microbiological Diagnostic Unit Public Health Laboratory, Doherty Institute | Caly L. et al |
| 814 | Canada/ON_PHL0977/2020 | 2020-03-10 | Public Health Ontario Laboratories | Public Health Ontario Laboratories | Alireza Eshaghi et al |
| 815 | Portugal/PT0018/2020 | 2020-03-10 | H Braga | Instituto Nacional de Saude (INSA) | Guiomar et al et al |
| 816 | Taiwan/CGMH-CGU-08/2020 | 2020-03-10 | Laboratory Medicine | Department of Laboratory Medicine, Lin-Kou Chang Gung Memorial Hospital, Taoyuan, Taiwan | Kuo-Chien Tsao et al |
| 817 | Netherlands/NA_2/2020 | 2020-03-10 | Dutch COVID-19 response team | Erasmus Medical Center | David Nieuwenhuijse et al |
| 818 | Denmark/ALAB-SSI259/2020 | 2020-03-10 | Department of Virus and Microbiological Special Diagnostics, Statens Serum Institut, Copenhagen, | Albertsen lab, Department of Chemistry and Bioscience, Aalborg University, Denmark | Rasmus Kirkegaard et al |

| | | | | | |
|---|---|---|---|---|---|
| | | | Denmark, Artillerivej 5, 2300 Copenahgen S | | |
| 819 | USA/CA-SCCPHD-UC155/2020 | 2020-03-10 | Santa Clara County Public Health Department | Chiu Laboratory, University of California, San Francisco | Xianding Deng et al |
| 820 | Denmark/ALAB-SSI418/2020 | 2020-03-10 | Department of Virus and Microbiological Special Diagnostics, Statens Serum Institut, Copenhagen, Denmark, Artillerivej 5, 2300 Copenahgen S | Albertsen lab, Department of Chemistry and Bioscience, Aalborg University, Denmark | Rasmus Kirkegaard et al |
| 821 | Netherlands/NA_6/2020 | 2020-03-10 | Dutch COVID-19 response team | Erasmus Medical Center | David Nieuwenhuijse et al |
| 822 | Denmark/ALAB-HH06/2020 | 2020-03-10 | Department of Clinical Microbiology, Copenhagen University Hospital, Hvidovre, Kettegaard Alle 30, 2650 Hvidovre. | Albertsen lab, Department of Chemistry and Bioscience, Aalborg University, Denmark | Rasmus Kirkegaard et al |
| 823 | Australia/VIC27/2020 | 2020-03-10 | Victorian Infectious Diseases Reference Laboratory (VIDRL) | Victorian Infectious Diseases Reference Laboratory and Microbiological Diagnostic Unit Public Health Laboratory, Doherty Institute | Caly L. et al |
| 824 | Australia/NSW103/2020 | 2020-03-10 | Centre for Infectious Diseases and Microbiology Public Health | NSW Health Pathology - Institute of Clinical Pathology and Medical Research; Westmead Hospital; University of Sydney | Rockett R et al |
| 825 | Germany/NRW-12/2020 | 2020-03-10 | Center of Medical Microbiology, Virology, and Hospital Hygiene, University of Duesseldorf | Center of Medical Microbiology, Virology, and Hospital Hygiene, University of Duesseldorf | Ortwin Adams et al |
| 826 | Spain/Madrid_H12_16/2020 | 2020-03-10 | Hospital Universitario 12 de Octubre | Hospital Universitario 12 de Octubre | Sara González et al |
| 827 | Australia/VIC46/2020 | 2020-03-11 | Victorian Infectious Diseases Reference Laboratory (VIDRL) | Victorian Infectious Diseases Reference Laboratory and Microbiological Diagnostic Unit Public Health Laboratory, Doherty Institute | Caly L. et al |
| 828 | Australia/VIC44/2020 | 2020-03-11 | Victorian Infectious Diseases Reference Laboratory (VIDRL) | Victorian Infectious Diseases Reference Laboratory and Microbiological Diagnostic Unit Public Health Laboratory, Doherty Institute | Caly L. et al |
| 829 | Australia/NSW218/2020 | 2020-03-11 | Centre for Infectious Diseases and Microbiology Public Health | NSW Health Pathology - Institute of Clinical Pathology and Medical Research; Westmead Hospital; University of Sydney | Timms V et al |
| 830 | Australia/NSW52/2020 | 2020-03-11 | Centre for Infectious Diseases and Microbiology Public Health | NSW Health Pathology - Institute of Clinical Pathology and Medical Research; Westmead Hospital; University of Sydney | Gall M et al |
| 831 | Australia/VIC38/2020 | 2020-03-11 | Victorian Infectious Diseases Reference Laboratory (VIDRL) | Victorian Infectious Diseases Reference Laboratory and Microbiological Diagnostic Unit Public Health Laboratory, Doherty Institute | Caly L. et al |
| 832 | Australia/NSW34/2020 | 2020-03-11 | Centre for Infectious Diseases and Microbiology Public Health | NSW Health Pathology - Institute of Clinical Pathology and Medical Research; Westmead Hospital; University of Sydney | Draper J et al |
| 833 | Australia/VIC41/2020 | 2020-03-11 | Victorian Infectious Diseases Reference Laboratory (VIDRL) | Victorian Infectious Diseases Reference Laboratory and Microbiological Diagnostic Unit Public Health Laboratory, Doherty Institute | Caly L. et al |
| 834 | Australia/VIC45/2020 | 2020-03-11 | Victorian Infectious Diseases Reference Laboratory (VIDRL) | Victorian Infectious Diseases Reference Laboratory and Microbiological Diagnostic Unit Public Health Laboratory, Doherty Institute | Caly L. et al |

| | | | | | |
|---|---|---|---|---|---|
| 835 | Australia/QLDID920/2020 | 2020-03-11 | Rockhampton Base Hospital | Public Health Virology Laboratory | Bixing Huang et al |
| 836 | Spain/Madrid_LP30_2226/2020 | 2020-03-11 | Hospital Universitario La Paz | Hospital Universitario 12 de Octubre | Elias Dahdouh et al |
| 837 | Iceland/110/2020 | 2020-03-11 | The National University Hospital of Iceland | deCODE genetics | Daniel F Gudbjartsson et al |
| 838 | USA/WA-UW108/2020 | 2020-03-11 | UW Virology Lab | UW Virology Lab | Pavitra Roychoudhury et al |
| 839 | Australia/VIC42/2020 | 2020-03-11 | Victorian Infectious Diseases Reference Laboratory (VIDRL) | Victorian Infectious Diseases Reference Laboratory and Microbiological Diagnostic Unit Public Health Laboratory, Doherty Institute | Caly L. et al |
| 840 | Australia/QLDID919/2020 | 2020-03-11 | Pathology Queensland | Public Health Virology Laboratory | Bixing Huang et al |
| 841 | Australia/VIC39/2020 | 2020-03-11 | Victorian Infectious Diseases Reference Laboratory (VIDRL) | Victorian Infectious Diseases Reference Laboratory and Microbiological Diagnostic Unit Public Health Laboratory, Doherty Institute | Caly L. et al |
| 842 | Germany/NRW-13/2020 | 2020-03-11 | Center of Medical Microbiology, Virology, and Hospital Hygiene, University of Duesseldorf | Center of Medical Microbiology, Virology, and Hospital Hygiene, University of Duesseldorf | Ortwin Adams et al |
| 843 | Germany/NRW-15/2020 | 2020-03-11 | Center of Medical Microbiology, Virology, and Hospital Hygiene, University of Duesseldorf | Center of Medical Microbiology, Virology, and Hospital Hygiene, University of Duesseldorf | Ortwin Adams et al |
| 844 | Russia/Lipetsk-62707/2020 | 2020-03-11 | State Research Center of Virology and Biotechnology VECTOR, Department of Collection of Microorganisms | State Research Center of Virology and Biotechnology VECTOR, Department of Collection of Microorganisms | Oleg V. Pyankov et al |
| 845 | USA/UN-UW-536/2020 | 2020-03-11 | UW Virology Lab | UW Virology Lab | Pavitra Roychoudhury et al |
| 846 | Spain/Madrid_H5_34/2020 | 2020-03-11 | Hospital Universitario 12 de Octubre | Hospital Universitario La Paz | Elias Dahdouh et al |
| 847 | NewZealand/20VR0712/2020-03-11 | 2020-03-11 | PathLab Bay of Plenty | Institute of Environmental Science and Research (ESR) Ltd | **Matt Storey et al** |
| 848 | Denmark/ALAB-SSI419/2020 | 2020-03-11 | Department of Virus and Microbiological Special Diagnostics, Statens Serum Institut, Copenhagen, Denmark, Artillerivej 5, 2300 Copenahgen S | Albertsen lab, Department of Chemistry and Bioscience, Aalborg University, Denmark | Rasmus Kirkegaard et al |
| 849 | Russia/Moscow-62501/2020 | 2020-03-11 | State Research Center of Virology and Biotechnology VECTOR, Department of Collection of Microorganisms | State Research Center of Virology and Biotechnology VECTOR, Department of Collection of Microorganisms | Oleg V. Pyankov et al |
| 850 | Canada/BC_5979789/2020 | 2020-03-11 | BCCDC Public Health Laboratory | BCCDC Public Health Laboratory | Harrigan et al |
| 851 | Netherlands/NoordBrabant_81/2020 | 2020-03-11 | Dutch COVID-19 response team | Erasmus Medical Center | Bas Oude Munnink et al |
| 852 | USA/NY-Wadsworth-13468-02/2020 | 2020-03-11 | Wadsworth Center, New York State Department.of Health | Wadsworth Center, New York State Department.of Health | Kirsten St. George et al |
| 853 | Iceland/9/2020 | 2020-03-11 | The National University Hospital of Iceland | deCODE genetics | Daniel F Gudbjartsson et al |
| 854 | USA/CA-SCCPHD-UC152/2020 | 2020-03-11 | Santa Clara County Public Health Department | Chiu Laboratory, University of California, San Francisco | Xianding Deng et al |
| 855 | USA/VA-DCLS-0019/2020 | 2020-03-11 | Virginia DCLS | Virginia DCLS | Virginia DCLS et al |
| 856 | Iceland/112/2020 | 2020-03-11 | The National University Hospital of Iceland | deCODE genetics | Daniel F Gudbjartsson et al |
| 857 | Iceland/106/2020 | 2020-03-11 | The National University Hospital of Iceland | deCODE genetics | Daniel F Gudbjartsson et al |
| 858 | Iceland/123/2020 | 2020-03-11 | The National University Hospital of Iceland | deCODE genetics | Daniel F Gudbjartsson et al |
| 859 | Iceland/109/2020 | 2020-03-11 | The National University Hospital of Iceland | deCODE genetics | Daniel F Gudbjartsson et al |
| 860 | Iceland/111/2020 | 2020-03-11 | The National University Hospital of Iceland | deCODE genetics | Daniel F Gudbjartsson et al |

| | | | | | |
|---|---|---|---|---|---|
| 861 | Iceland/125/2020 | 2020-03-11 | The National University Hospital of Iceland | deCODE genetics | Daniel F Gudbjartsson et al |
| 862 | Netherlands/NA_132/2020 | 2020-03-11 | Dutch COVID-19 response team | Erasmus Medical Center | Bas Oude Munnink et al |
| 863 | USA/TX-HMH028/2020 | 2020-03-11 | Houston Methodist Hospital | Houston Methodist Hospital | S. Wesley Long et al |
| 864 | Sweden/20-08144/2020 | 2020-03-11 | Klinisk mikrobiologi och vardhygien Halmstad | The Public Health Agency of Sweden | Arne Kotz et al |
| 865 | Taiwan/TSGH-27/2020 | 2020-03-11 | TSGH-CP molecular lab | TSGH-CP molecular lab | Cherng-Lih Perng et al |
| 866 | Spain/Madrid_LP28_2522/2020 | 2020-03-11 | Hospital Universitario La Paz | Hospital Universitario 12 de Octubre | Dahdouh et al |
| 867 | Brunei/3/2020 | 2020-03-11 | National Virology Reference Laboratory | National Public Health Laboratory, National Centre for Infectious Diseases | Mak Tze Minn et al |
| 868 | Sweden/20-08150/2020 | 2020-03-11 | Klinisk mikrobiologi och vardhygien Halmstad | The Public Health Agency of Sweden | Arne Kotz et al |
| 869 | USA/WA-UW105/2020 | 2020-03-11 | UW Virology Lab | UW Virology Lab | Pavitra Roychoudhury et al |
| 870 | Denmark/ALAB-HH26/2020 | 2020-03-11 | Department of Clinical Microbiology, Copenhagen University Hospital, Hvidovre, Kettegaard Alle 30, 2650 Hvidovre. | Albertsen lab, Department of Chemistry and Bioscience, Aalborg University, Denmark | Rasmus Kirkegaard et al |
| 871 | Spain/Madrid_H12_3405/2020 | 2020-03-11 | Hospital Universitario 12 de Octubre | Hospital Universitario 12 de Octubre | Esther Viedma et al |
| 872 | USA/UT-025/2020 | 2020-03-11 | Utah Public Health Laboratory | Utah Public Health Laboratory | Erin Young et al |
| 873 | USA/WA-UW118/2020 | 2020-03-11 | UW Virology Lab | UW Virology Lab | Pavitra Roychoudhury et al |
| 874 | Australia/VIC55/2020 | 2020-03-12 | Victorian Infectious Diseases Reference Laboratory (VIDRL) | Victorian Infectious Diseases Reference Laboratory and Microbiological Diagnostic Unit Public Health Laboratory, Doherty Institute | Caly L. et al |
| 875 | Australia/VIC48/2020 | 2020-03-12 | Victorian Infectious Diseases Reference Laboratory (VIDRL) | Victorian Infectious Diseases Reference Laboratory and Microbiological Diagnostic Unit Public Health Laboratory, Doherty Institute | Caly L. et al |
| 876 | Australia/VIC53/2020 | 2020-03-12 | Victorian Infectious Diseases Reference Laboratory (VIDRL) | Victorian Infectious Diseases Reference Laboratory and Microbiological Diagnostic Unit Public Health Laboratory, Doherty Institute | Caly L. et al |
| 877 | Australia/VIC51/2020 | 2020-03-12 | Victorian Infectious Diseases Reference Laboratory (VIDRL) | Victorian Infectious Diseases Reference Laboratory and Microbiological Diagnostic Unit Public Health Laboratory, Doherty Institute | Caly L. et al |
| 878 | Australia/VIC54/2020 | 2020-03-12 | Victorian Infectious Diseases Reference Laboratory (VIDRL) | Victorian Infectious Diseases Reference Laboratory and Microbiological Diagnostic Unit Public Health Laboratory, Doherty Institute | Caly L. et al |
| 879 | Australia/VIC58/2020 | 2020-03-12 | Victorian Infectious Diseases Reference Laboratory (VIDRL) | Victorian Infectious Diseases Reference Laboratory and Microbiological Diagnostic Unit Public Health Laboratory, Doherty Institute | Caly L. et al |
| 880 | Scotland/CVR18/2020 | 2020-03-12 | West of Scotland Specialist Virology Centre, NHSGGC / MRC-University of Glasgow Centre for Virus Research | COVID-19 Genomics UK (COG-UK) Consortium | Ana da Silva Filipe et al |
| 881 | USA/WA-UW100/2020 | 2020-03-12 | UW Virology Lab | UW Virology Lab | Pavitra Roychoudhury et al |
| 882 | UnitedArabEmirates/L0/2020 | 2020-03-12 | Mohammed Bin Rashid University of Medicine and Health Sciences | Al Jalila Children’s Hospital | Ahmad Abou Tayoun et al |
| 883 | Scotland/CVR19/2020 | 2020-03-12 | West of Scotland Specialist Virology Centre, NHSGGC / MRC-University of Glasgow Centre for Virus Research | COVID-19 Genomics UK (COG-UK) Consortium | Ana da Silva Filipe et al |

| | | | | | |
|---|---|---|---|---|---|
| 884 | Iceland/135/2020 | 2020-03-12 | The National University Hospital of Iceland | deCODE genetics | Daniel F Gudbjartsson et al |
| 885 | Australia/VIC50/2020 | 2020-03-12 | Victorian Infectious Diseases Reference Laboratory (VIDRL) | Victorian Infectious Diseases Reference Laboratory and Microbiological Diagnostic Unit Public Health Laboratory, Doherty Institute | Caly L. et al |
| 886 | Australia/VIC47/2020 | 2020-03-12 | Victorian Infectious Diseases Reference Laboratory (VIDRL) | Victorian Infectious Diseases Reference Laboratory and Microbiological Diagnostic Unit Public Health Laboratory, Doherty Institute | Caly L. et al |
| 887 | Australia/NSW50/2020 | 2020-03-12 | Centre for Infectious Diseases and Microbiology Public Health | NSW Health Pathology - Institute of Clinical Pathology and Medical Research; Westmead Hospital; University of Sydney | Timms V et al |
| 888 | Iceland/127/2020 | 2020-03-12 | The National University Hospital of Iceland | deCODE genetics | Daniel F Gudbjartsson et al |
| 889 | Japan/P4-3/2020 | 2020-03-12 | Saitama Prefectural Institute of Public Health | Pathogen Genomics Center, National Institute of Infectious Diseases | Tsuyoshi Sekizuka et al |
| 890 | USA/WA-UW124/2020 | 2020-03-12 | UW Virology Lab | UW Virology Lab | Pavitra Roychoudhury et al |
| 891 | USA/MN32-MDH32/2020 | 2020-03-12 | Minnesota Department of Health, Public Health Laboratory | Minnesota Department of Health, Public Health Laboratory | Matt Plumb et al |
| 892 | Wales/PHW27/2020 | 2020-03-12 | Wales Specialist Virology Centre | Public Health Wales Microbiology Cardiff | Catherine Moore et al |
| 893 | NewZealand/20VR0334/2020-03-12 | 2020-03-12 | LabPLUS | Institute of Environmental Science and Research (ESR) Ltd | **Matt Storey et al** |
| 894 | USA/WA-UW123/2020 | 2020-03-12 | UW Virology Lab | UW Virology Lab | Pavitra Roychoudhury et al |
| 895 | Scotland/CVR131/2020 | 2020-03-12 | West of Scotland Specialist Virology Centre, NHSGGC / MRC-University of Glasgow Centre for Virus Research | COVID-19 Genomics UK (COG-UK) Consortium | Ana da Silva Filipe et al |
| 896 | Spain/Madrid_H12_32/2020 | 2020-03-12 | Hospital Universitario 12 de Octubre | Hospital Universitario 12 de Octubre | Raúl Recio et al |
| 897 | USA/NY-WCMP2D02P/2020 | 2020-03-12 | NewYork-Presbyterian & Mason Lab | Mason Lab | Daniel J. Butler et al |
| 898 | Iceland/124/2020 | 2020-03-12 | The National University Hospital of Iceland | deCODE genetics | Daniel F Gudbjartsson et al |
| 899 | Iceland/140/2020 | 2020-03-12 | The National University Hospital of Iceland | deCODE genetics | Daniel F Gudbjartsson et al |
| 900 | Iceland/126/2020 | 2020-03-12 | The National University Hospital of Iceland | deCODE genetics | Daniel F Gudbjartsson et al |
| 901 | Iceland/129/2020 | 2020-03-12 | The National University Hospital of Iceland | deCODE genetics | Daniel F Gudbjartsson et al |
| 902 | Iceland/132/2020 | 2020-03-12 | The National University Hospital of Iceland | deCODE genetics | Daniel F Gudbjartsson et al |
| 903 | Iceland/133/2020 | 2020-03-12 | The National University Hospital of Iceland | deCODE genetics | Daniel F Gudbjartsson et al |
| 904 | Iceland/138/2020 | 2020-03-12 | The National University Hospital of Iceland | deCODE genetics | Daniel F Gudbjartsson et al |
| 905 | Iceland/130/2020 | 2020-03-12 | The National University Hospital of Iceland | deCODE genetics | Daniel F Gudbjartsson et al |
| 906 | Canada/ON_PHL5710/2020 | 2020-03-12 | Public Health Ontario Laboratories | Public Health Ontario Laboratories | Alireza Eshaghi et al |
| 907 | Portugal/PT0019/2020 | 2020-03-12 | ARS Algarve - Laboratório Laura Ayres | Instituto Nacional de Saude (INSA) | Guiomar et al et al |
| 908 | Scotland/CVR16/2020 | 2020-03-12 | West of Scotland Specialist Virology Centre, NHSGGC / MRC-University of Glasgow Centre for Virus Research | COVID-19 Genomics UK (COG-UK) Consortium | Ana da Silva Filipe et al |
| 909 | Greece/150/2020 | 2020-03-12 | Laboratory of Microbiology, Medical School, National and Kapodistrian University of Athens | Laboratory of Biology, Department of Medicine, Democritus University of Thrace | Kassela K. et al |

| | | | | | |
|---|---|---|---|---|---|
| 910 | Sweden/20-08169/2020 | 2020-03-12 | Klinisk mikrobiologi och vardhygien Halmstad | The Public Health Agency of Sweden | Arne Kotz et al |
| 911 | France/GE2722/2020 | 2020-03-12 | L'Air du Temps | National Reference Center for Viruses of Respiratory Infections, Institut Pasteur, Paris | Mélanie Albert et al |
| 912 | Iceland/136/2020 | 2020-03-12 | The National University Hospital of Iceland | deCODE genetics | Daniel F Gudbjartsson et al |
| 913 | Iceland/139/2020 | 2020-03-12 | The National University Hospital of Iceland | deCODE genetics | Daniel F Gudbjartsson et al |
| 914 | Iceland/131/2020 | 2020-03-12 | The National University Hospital of Iceland | deCODE genetics | Daniel F Gudbjartsson et al |
| 915 | Iceland/134/2020 | 2020-03-12 | The National University Hospital of Iceland | deCODE genetics | Daniel F Gudbjartsson et al |
| 916 | Iceland/128/2020 | 2020-03-12 | The National University Hospital of Iceland | deCODE genetics | Daniel F Gudbjartsson et al |
| 917 | USA/WA-UW-546/2020 | 2020-03-12 | UW Virology Lab | UW Virology Lab | Pavitra Roychoudhury et al |
| 918 | Spain/Madrid_H12_30/2020 | 2020-03-12 | Hospital Universitario 12 de Octubre | Hospital Universitario 12 de Octubre | Sara González et al |
| 919 | Netherlands/NoordBrabant_82/2020 | 2020-03-12 | Dutch COVID-19 response team | Erasmus Medical Center | Bas Oude Munnink et al |
| 920 | Canada/ON_PHL0142/2020 | 2020-03-12 | Public Health Ontario Laboratories | Public Health Ontario Laboratories | Alireza Eshaghi et al |
| 921 | USA/WA-UW133/2020 | 2020-03-12 | UW Virology Lab | UW Virology Lab | Pavitra Roychoudhury et al |
| 922 | Spain/Madrid_H8_37/2020 | 2020-03-12 | Hospital Universitario 12 de Octubre | Hospital Universitario La Paz | Elias Dahdouh et al |
| 923 | Spain/Vircell001/2020 | 2020-03-12 | Health Sciences Technology Park, Avicena, 8, 18016 Granada. Spain | Sequencing and Bioinformatics Service FISABIO-Public Health | Joaquín Mendoza et al |
| 924 | USA/WA-S273/2020 | 2020-03-12 | Seattle Flu Study | Seattle Flu Study | Chu et al et al |
| 925 | USA/WA-S140/2020 | 2020-03-12 | Washington State Department of Health | Seattle Flu Study | Chu et al et al |
| 926 | USA/WA-UW315/2020 | 2020-03-13 | UW Virology Lab | UW Virology Lab | Pavitra Roychoudhury et al |
| 927 | England/NOTT-10DF32/2020 | 2020-03-13 | Queens Medical Centre, Clinical Microbiology Department / DeepSeq Nottingham | COVID-19 Genomics UK (COG-UK) Consortium | Gemma Clark et al |
| 928 | Australia/VIC69/2020 | 2020-03-13 | Victorian Infectious Diseases Reference Laboratory (VIDRL) | Victorian Infectious Diseases Reference Laboratory and Microbiological Diagnostic Unit Public Health Laboratory, Doherty Institute | Caly L. et al |
| 929 | Australia/VIC66/2020 | 2020-03-13 | Victorian Infectious Diseases Reference Laboratory (VIDRL) | Victorian Infectious Diseases Reference Laboratory and Microbiological Diagnostic Unit Public Health Laboratory, Doherty Institute | Caly L. et al |
| 930 | Australia/VIC59/2020 | 2020-03-13 | Victorian Infectious Diseases Reference Laboratory (VIDRL) | Victorian Infectious Diseases Reference Laboratory and Microbiological Diagnostic Unit Public Health Laboratory, Doherty Institute | Caly L. et al |
| 931 | Australia/VIC62/2020 | 2020-03-13 | Victorian Infectious Diseases Reference Laboratory (VIDRL) | Victorian Infectious Diseases Reference Laboratory and Microbiological Diagnostic Unit Public Health Laboratory, Doherty Institute | Caly L. et al |
| 932 | Australia/NSW45/2020 | 2020-03-13 | Centre for Infectious Diseases and Microbiology Public Health | NSW Health Pathology - Institute of Clinical Pathology and Medical Research; Westmead Hospital; University of Sydney | Eden J-S et al |
| 933 | Australia/WA07/2020 | 2020-03-13 | Department of Microbiology, PathWest QEII Medical Centre | Department of Microbiology, PathWest QEII Medical Centre | Chisha Sikazwe et al |
| 934 | Australia/NSW47/2020 | 2020-03-13 | Centre for Infectious Diseases and Microbiology Public Health | NSW Health Pathology - Institute of Clinical Pathology and Medical Research; | Lam C et al |

| | | | | | |
|---|---|---|---|---|---|
| | | | | Westmead Hospital; University of Sydney | |
| 935 | Australia/VIC68/2020 | 2020-03-13 | Victorian Infectious Diseases Reference Laboratory (VIDRL) | Victorian Infectious Diseases Reference Laboratory and Microbiological Diagnostic Unit Public Health Laboratory, Doherty Institute | Caly L. et al |
| 936 | Australia/VIC70/2020 | 2020-03-13 | Victorian Infectious Diseases Reference Laboratory (VIDRL) | Victorian Infectious Diseases Reference Laboratory and Microbiological Diagnostic Unit Public Health Laboratory, Doherty Institute | Caly L. et al |
| 937 | Scotland/EDB021/2020 | 2020-03-13 | Virology Department, Royal Infirmary of Edinburgh, NHS Lothian / School of Biological Sciences, University of Edinburgh / Institute of Genetics and Molecular Medicine, University of Edinburgh | COVID-19 Genomics UK (COG-UK) Consortium | McHugh M et al |
| 938 | USA/CT-UW271/2020 | 2020-03-13 | UW Virology Lab | UW Virology Lab | Pavitra Roychoudhury et al |
| 939 | Scotland/CVR51/2020 | 2020-03-13 | West of Scotland Specialist Virology Centre, NHSGGC / MRC-University of Glasgow Centre for Virus Research | COVID-19 Genomics UK (COG-UK) Consortium | Ana da Silva Filipe et al |
| 940 | Austria/CeMM0007/2020 | 2020-03-13 | Center for Virology, Medical University of Vienna | Bergthaler laboratory, CeMM Research Center for Molecular Medicine of the Austrian Academy of Sciences | Alexandra Popa et al |
| 941 | USA/VA-DCLS-0024/2020 | 2020-03-13 | Virginia DCLS | Virginia DCLS | Virginia DCLS et al |
| 942 | Sweden/20-05627/2020 | 2020-03-13 | Svardsjo VC | The Public Health Agency of Sweden | Tommy Janers et al |
| 943 | Australia/VIC67/2020 | 2020-03-13 | Victorian Infectious Diseases Reference Laboratory (VIDRL) | Victorian Infectious Diseases Reference Laboratory and Microbiological Diagnostic Unit Public Health Laboratory, Doherty Institute | Caly L. et al |
| 944 | Australia/WA02/2020 | 2020-03-13 | Department of Microbiology, PathWest QEII Medical Centre | Department of Microbiology, PathWest QEII Medical Centre | Chisha Sikazwe et al |
| 945 | Australia/VIC64/2020 | 2020-03-13 | Victorian Infectious Diseases Reference Laboratory (VIDRL) | Victorian Infectious Diseases Reference Laboratory and Microbiological Diagnostic Unit Public Health Laboratory, Doherty Institute | Caly L. et al |
| 946 | USA/CT-UW224/2020 | 2020-03-13 | UW Virology Lab | UW Virology Lab | Pavitra Roychoudhury et al |
| 947 | Australia/VIC60/2020 | 2020-03-13 | Victorian Infectious Diseases Reference Laboratory (VIDRL) | Victorian Infectious Diseases Reference Laboratory and Microbiological Diagnostic Unit Public Health Laboratory, Doherty Institute | Caly L. et al |
| 948 | Fujian/IM3520001T/2020 | 2020-03-13 | Fujian Center for Disease Control and Prevention | Fujian Center for Disease Control and Prevention | Lin Qi et al |
| 949 | NewZealand/20VR0461/2020-03-13 | 2020-03-13 | Southern Community Labs Dunedin | Institute of Environmental Science and Research (ESR) Ltd | **Matt Storey et al** |
| 950 | USA/WA-UW205/2020 | 2020-03-13 | UW Virology Lab | UW Virology Lab | Pavitra Roychoudhury et al |
| 951 | Scotland/CVR107/2020 | 2020-03-13 | West of Scotland Specialist Virology Centre, NHSGGC / MRC-University of Glasgow Centre for Virus Research | COVID-19 Genomics UK (COG-UK) Consortium | Ana da Silva Filipe et al |
| 952 | USA/MN53-MDH53/2020 | 2020-03-13 | Minnesota Department of Health, Public Health Laboratory | Minnesota Department of Health, Public Health Laboratory | Matt Plumb et al |
| 953 | Finland/13M82/2020 | 2020-03-13 | Department of Virology and Immunology, University of Helsinki and Helsinki University Hospital, Huslab Finland | Department of Virology, Faculty of Medicine, University of Helsinki, Helsinki, Finland | Teemu Smura et al |
| 954 | Iceland/151/2020 | 2020-03-13 | The National University | deCODE genetics | Daniel F Gudbjartsson |

| | | | | | |
|---|---|---|---|---|---|
| | | | Hospital of Iceland | | et al |
| 955 | Iceland/168/2020 | 2020-03-13 | The National University Hospital of Iceland | deCODE genetics | Daniel F Gudbjartsson et al |
| 956 | Iceland/146/2020 | 2020-03-13 | The National University Hospital of Iceland | deCODE genetics | Daniel F Gudbjartsson et al |
| 957 | Iceland/152/2020 | 2020-03-13 | The National University Hospital of Iceland | deCODE genetics | Daniel F Gudbjartsson et al |
| 958 | Iceland/145/2020 | 2020-03-13 | The National University Hospital of Iceland | deCODE genetics | Daniel F Gudbjartsson et al |
| 959 | Iceland/157/2020 | 2020-03-13 | The National University Hospital of Iceland | deCODE genetics | Daniel F Gudbjartsson et al |
| 960 | Iceland/150/2020 | 2020-03-13 | The National University Hospital of Iceland | deCODE genetics | Daniel F Gudbjartsson et al |
| 961 | Iceland/147/2020 | 2020-03-13 | The National University Hospital of Iceland | deCODE genetics | Daniel F Gudbjartsson et al |
| 962 | Iceland/142/2020 | 2020-03-13 | The National University Hospital of Iceland | deCODE genetics | Daniel F Gudbjartsson et al |
| 963 | Iceland/3/2020 | 2020-03-13 | The National University Hospital of Iceland | deCODE genetics | Daniel F Gudbjartsson et al |
| 964 | Iceland/144/2020 | 2020-03-13 | The National University Hospital of Iceland | deCODE genetics | Daniel F Gudbjartsson et al |
| 965 | Iceland/137/2020 | 2020-03-13 | The National University Hospital of Iceland | deCODE genetics | Daniel F Gudbjartsson et al |
| 966 | Iceland/6/2020 | 2020-03-13 | The National University Hospital of Iceland | deCODE genetics | Daniel F Gudbjartsson et al |
| 967 | Iceland/158/2020 | 2020-03-13 | The National University Hospital of Iceland | deCODE genetics | Daniel F Gudbjartsson et al |
| 968 | Iceland/148/2020 | 2020-03-13 | The National University Hospital of Iceland | deCODE genetics | Daniel F Gudbjartsson et al |
| 969 | Iceland/153/2020 | 2020-03-13 | The National University Hospital of Iceland | deCODE genetics | Daniel F Gudbjartsson et al |
| 970 | Iceland/156/2020 | 2020-03-13 | The National University Hospital of Iceland | deCODE genetics | Daniel F Gudbjartsson et al |
| 971 | Iceland/166/2020 | 2020-03-13 | The National University Hospital of Iceland | deCODE genetics | Daniel F Gudbjartsson et al |
| 972 | Iceland/335/2020 | 2020-03-13 | deCODE genetics | deCODE genetics | Daniel F Gudbjartsson et al |
| 973 | Iceland/5/2020 | 2020-03-13 | The National University Hospital of Iceland | deCODE genetics | Daniel F Gudbjartsson et al |
| 974 | France/IDF2420/2020 | 2020-03-13 | Clinique AVERAY LA BROUSTE, Med. Polyvalente | National Reference Center for Viruses of Respiratory Infections, Institut Pasteur, Paris | Mélanie Albert et al |
| 975 | Iceland/159/2020 | 2020-03-13 | The National University Hospital of Iceland | deCODE genetics | Daniel F Gudbjartsson et al |
| 976 | Iceland/149/2020 | 2020-03-13 | The National University Hospital of Iceland | deCODE genetics | Daniel F Gudbjartsson et al |
| 977 | USA/IL-UW353/2020 | 2020-03-13 | UW Virology Lab | UW Virology Lab | Pavitra Roychoudhury et al |
| 978 | USA/WA-NH22/2020 | 2020-03-13 | WA State Department of Health | Pathogen Discovery, Respiratory Viruses Branch, Division of Viral Diseases, Centers for Disease Control and Prevention | Ying Tao et al |
| 979 | England/20124097102/2020 | 2020-03-13 | Respiratory Virus Unit, Microbiology Services Colindale, Public Health England | Respiratory Virus Unit, Microbiology Services Colindale, Public Health England | Monica Galiano et al |
| 980 | USA/WA-UW299/2020 | 2020-03-13 | UW Virology Lab | UW Virology Lab | Pavitra Roychoudhury et al |
| 981 | USA/WA-UW163/2020 | 2020-03-13 | UW Virology Lab | UW Virology Lab | Pavitra Roychoudhury et al |
| 982 | Spain/Valencia45/2020 | 2020-03-13 | Servicio de Microbiologia. Hospital Clinico Universitario de Valencia | Sequencing and Bioinformatics Service and Molecular Epidemiology Research Group. FISABIO-Public Health | Inma Galán Vendrell et al |
| 983 | Iceland/169/2020 | 2020-03-13 | The National University Hospital of Iceland | deCODE genetics | Daniel F Gudbjartsson et al |
| 984 | Taiwan/CGMH-CGU-10/2020 | 2020-03-13 | Laboratory Medicine | Department of Laboratory Medicine, Lin-Kou Chang Gung | Kuo-Chien Tsao et al |

| | | | | | |
|---|---|---|---|---|---|
| | | | | Memorial Hospital, Taoyuan, Taiwan | |
| 985 | Taiwan/NTU05/2020 | 2020-03-13 | Department of Laboratory Medicine, National Taiwan University Hospital | Microbial Genomics Core Lab, National Taiwan University Centers of Genomic and Precision Medicine | Shiou-Hwei Yeh et al |
| 986 | Taiwan/CGMH-CGU-09/2020 | 2020-03-13 | Laboratory Medicine | Department of Laboratory Medicine, Lin-Kou Chang Gung Memorial Hospital, Taoyuan, Taiwan | Kuo-Chien Tsao et al |
| 987 | Taiwan/NTU06/2020 | 2020-03-13 | Department of Laboratory Medicine, National Taiwan University Hospital | Microbial Genomics Core Lab, National Taiwan University Centers of Genomic and Precision Medicine | Shiou-Hwei Yeh et al |
| 988 | Taiwan/NTU07/2020 | 2020-03-13 | Department of Laboratory Medicine, National Taiwan University Hospital | Microbial Genomics Core Lab, National Taiwan University Centers of Genomic and Precision Medicine | Shiou-Hwei Yeh et al |
| 989 | Germany/NRW-18/2020 | 2020-03-13 | Center of Medical Microbiology, Virology, and Hospital Hygiene, University of Duesseldorf | Center of Medical Microbiology, Virology, and Hospital Hygiene, University of Duesseldorf | Ortwin Adams et al |
| 990 | Canada/NS_13/2020 | 2020-03-13 | Queen Elizabeth II Health Science Centre | National Microbiology Laboratory | Anna Majer et al |
| 991 | Scotland/CVR108/2020 | 2020-03-13 | West of Scotland Specialist Virology Centre, NHSGGC / MRC-University of Glasgow Centre for Virus Research | COVID-19 Genomics UK (COG-UK) Consortium | Ana da Silva Filipe et al |
| 992 | Russia/Moscow-64601/2020 | 2020-03-13 | State Research Center of Virology and Biotechnology VECTOR, Department of Collection of Microorganisms | State Research Center of Virology and Biotechnology VECTOR, Department of Collection of Microorganisms | Oleg V. Pyankov et al |
| 993 | Senegal/136/2020 | 2020-03-13 | Institut Pasteur Dakar | Institut Pasteur de Dakar | Ndongo Dia et al |
| 994 | USA/UT-021/2020 | 2020-03-14 | Utah Public Health Laboratory | Utah Public Health Laboratory | Erin Young et al |
| 995 | Australia/QLDID923/2020 | 2020-03-14 | Mater Pathology | Public Health Virology Laboratory | Bixing Huang et al |
| 996 | Australia/WA10/2020 | 2020-03-14 | Department of Microbiology, PathWest QEII Medical Centre | Department of Microbiology, PathWest QEII Medical Centre | Chisha Sikazwe et al |
| 997 | Australia/NSW54/2020 | 2020-03-14 | Centre for Infectious Diseases and Microbiology Public Health | NSW Health Pathology - Institute of Clinical Pathology and Medical Research; Westmead Hospital; University of Sydney | Sadsad R et al |
| 998 | Australia/VIC77/2020 | 2020-03-14 | Victorian Infectious Diseases Reference Laboratory (VIDRL) | Victorian Infectious Diseases Reference Laboratory and Microbiological Diagnostic Unit Public Health Laboratory, Doherty Institute | Caly L. et al |
| 999 | Australia/VIC79/2020 | 2020-03-14 | Victorian Infectious Diseases Reference Laboratory (VIDRL) | Victorian Infectious Diseases Reference Laboratory and Microbiological Diagnostic Unit Public Health Laboratory, Doherty Institute | Caly L. et al |
| 1000 | Australia/VIC73/2020 | 2020-03-14 | Victorian Infectious Diseases Reference Laboratory (VIDRL) | Victorian Infectious Diseases Reference Laboratory and Microbiological Diagnostic Unit Public Health Laboratory, Doherty Institute | Caly L. et al |
| 1001 | Hangzhou/HZCDC6706/2020 | 2020-03-14 | Hangzhou Center for Diseases Control and Prevention | Hangzhou Center for Diseases Control and Prevention | Jun Li et al |
| 1002 | Australia/WA03/2020 | 2020-03-14 | Department of Microbiology, PathWest QEII Medical Centre | Department of Microbiology, PathWest QEII Medical Centre | Chisha Sikazwe et al |
| 1003 | Germany/NRW-24/2020 | 2020-03-14 | Center of Medical Microbiology, Virology, and Hospital Hygiene, University of Duesseldorf | Center of Medical Microbiology, Virology, and Hospital Hygiene, University of Duesseldorf | Ortwin Adams et al |
| 1004 | USA/WA-UW316/2020 | 2020-03-14 | UW Virology Lab | UW Virology Lab | Pavitra Roychoudhury et al |
| 1005 | Finland/14M74/2020 | 2020-03-14 | Department of Virology and Immunology, University of Helsinki and Helsinki | Department of Virology, Faculty of Medicine, University of Helsinki, Helsinki, Finland | Teemu Smura et al |

| | | | | | |
|---|---|---|---|---|---|
| | | | University Hospital, Huslab Finland | | |
| 1006 | USA/NY-Wadsworth-15616-01/2020 | 2020-03-14 | Wadsworth Center, New York State Department.of Health | Wadsworth Center, New York State Department.of Health | Kirsten St. George et al |
| 1007 | Australia/WA09/2020 | 2020-03-14 | Department of Microbiology, PathWest QEII Medical Centre | Department of Microbiology, PathWest QEII Medical Centre | Chisha Sikazwe et al |
| 1008 | Australia/VIC81/2020 | 2020-03-14 | Victorian Infectious Diseases Reference Laboratory (VIDRL) | Victorian Infectious Diseases Reference Laboratory and Microbiological Diagnostic Unit Public Health Laboratory, Doherty Institute | Caly L. et al |
| 1009 | Australia/WA01/2020 | 2020-03-14 | Department of Microbiology, PathWest QEII Medical Centre | Department of Microbiology, PathWest QEII Medical Centre | Chisha Sikazwe et al |
| 1010 | Australia/VIC80/2020 | 2020-03-14 | Victorian Infectious Diseases Reference Laboratory (VIDRL) | Victorian Infectious Diseases Reference Laboratory and Microbiological Diagnostic Unit Public Health Laboratory, Doherty Institute | Caly L. et al |
| 1011 | Australia/VIC78/2020 | 2020-03-14 | Victorian Infectious Diseases Reference Laboratory (VIDRL) | Victorian Infectious Diseases Reference Laboratory and Microbiological Diagnostic Unit Public Health Laboratory, Doherty Institute | Caly L. et al |
| 1012 | Australia/VIC82/2020 | 2020-03-14 | Victorian Infectious Diseases Reference Laboratory (VIDRL) | Victorian Infectious Diseases Reference Laboratory and Microbiological Diagnostic Unit Public Health Laboratory, Doherty Institute | Caly L. et al |
| 1013 | Australia/VIC84/2020 | 2020-03-14 | Victorian Infectious Diseases Reference Laboratory (VIDRL) | Victorian Infectious Diseases Reference Laboratory and Microbiological Diagnostic Unit Public Health Laboratory, Doherty Institute | Caly L. et al |
| 1014 | Italy/TE4925/2020 | 2020-03-14 | "Presidio Ospedaliero ""S. Spirito"" - PESCARA" | "Istituto Zooprofilattico Sperimentale dell'Abruzzo e Molise ""G. Caporale""" | Lorusso A et al |
| 1015 | Italy/TE4953/2020 | 2020-03-14 | "Presidio ospedaliero ""Santo Spirito""" | "Istituto Zooprofilattico Sperimentale dell'Abruzzo e Molise ""G. Caporale""" | Lorusso A et al |
| 1016 | Canada/NB_35/2020 | 2020-03-14 | Dr. Georges-L.-Dumont University Hospital Centre | National Microbiology Laboratory | Anna Majer et al |
| 1017 | USA/MN59-MDH59/2020 | 2020-03-14 | Minnesota Department of Health, Public Health Laboratory | Minnesota Department of Health, Public Health Laboratory | Matt Plumb et al |
| 1018 | USA/CT-UW258/2020 | 2020-03-14 | UW Virology Lab | UW Virology Lab | Pavitra Roychoudhury et al |
| 1019 | USA/WA-S147/2020 | 2020-03-14 | Washington State Department of Health | Seattle Flu Study | Chu et al et al |
| 1020 | Germany/NRW-20/2020 | 2020-03-14 | Center of Medical Microbiology, Virology, and Hospital Hygiene, University of Duesseldorf | Center of Medical Microbiology, Virology, and Hospital Hygiene, University of Duesseldorf | Ortwin Adams et al |
| 1021 | Spain/Valencia125/2020 | 2020-03-14 | Servicio de Microbiología. Consorcio Hospital General Universitario de Valencia | Sequencing and Bioinformatics Service and Molecular Epidemiology Research Group. FISABIO-Public Health | Griselda De Marco et al |
| 1022 | France/IDF2534/2020 | 2020-03-14 | LABM GH nord Essonne | National Reference Center for Viruses of Respiratory Infections, Institut Pasteur, Paris | Mélanie Albert et al |
| 1023 | Iceland/331/2020 | 2020-03-14 | deCODE genetics | deCODE genetics | Daniel F Gudbjartsson et al |
| 1024 | Iceland/161/2020 | 2020-03-14 | The National University Hospital of Iceland | deCODE genetics | Daniel F Gudbjartsson et al |
| 1025 | Iceland/164/2020 | 2020-03-14 | The National University Hospital of Iceland | deCODE genetics | Daniel F Gudbjartsson et al |
| 1026 | Iceland/171/2020 | 2020-03-14 | deCODE genetics | deCODE genetics | Daniel F Gudbjartsson et al |
| 1027 | Iceland/162/2020 | 2020-03-14 | The National University Hospital of Iceland | deCODE genetics | Daniel F Gudbjartsson et al |
| 1028 | Iceland/326/2020 | 2020-03-14 | deCODE genetics | deCODE genetics | Daniel F Gudbjartsson |

| | | | | | |
|---|---|---|---|---|---|
| | | | | | et al |
| 1029 | Iceland/336/2020 | 2020-03-14 | deCODE genetics | deCODE genetics | Daniel F Gudbjartsson et al |
| 1030 | Iceland/334/2020 | 2020-03-14 | deCODE genetics | deCODE genetics | Daniel F Gudbjartsson et al |
| 1031 | Iceland/176/2020 | 2020-03-14 | The National University Hospital of Iceland | deCODE genetics | Daniel F Gudbjartsson et al |
| 1032 | Iceland/330/2020 | 2020-03-14 | deCODE genetics | deCODE genetics | Daniel F Gudbjartsson et al |
| 1033 | Iceland/163/2020 | 2020-03-14 | The National University Hospital of Iceland | deCODE genetics | Daniel F Gudbjartsson et al |
| 1034 | Finland/14M77/2020 | 2020-03-14 | Department of Virology and Immunology, University of Helsinki and Helsinki University Hospital, Huslab Finland | Department of Virology, Faculty of Medicine, University of Helsinki, Helsinki, Finland | Teemu Smura et al |
| 1035 | Germany/BAV-V2011395/2020 | 2020-03-14 | Max von Pettenkofer Institute, Virology, National Reference Center for Retroviruses, LMU Munich | Laboratory for Functional Genome Analysis, Dept. Genomics, Gene Center of the LMU Munich | Max Muenchhoff et al |
| 1036 | USA/MN58-MDH58/2020 | 2020-03-14 | Minnesota Department of Health, Public Health Laboratory | Minnesota Department of Health, Public Health Laboratory | Matt Plumb et al |
| 1037 | Netherlands/NA_52/2020 | 2020-03-14 | Dutch COVID-19 response team | Erasmus Medical Center | Bas Oude Munnink et al |
| 1038 | Taiwan/CGMH-CGU-11/2020 | 2020-03-14 | Laboratory Medicine | Department of Laboratory Medicine, Lin-Kou Chang Gung Memorial Hospital, Taoyuan, Taiwan | Kuo-Chien Tsao et al |
| 1039 | Taiwan/CGMH-CGU-12/2020 | 2020-03-14 | Laboratory Medicine | Department of Laboratory Medicine, Lin-Kou Chang Gung Memorial Hospital, Taoyuan, Taiwan | Kuo-Chien Tsao et al |
| 1040 | Taiwan/NTU09/2020 | 2020-03-14 | Department of Laboratory Medicine, National Taiwan University Hospital | Microbial Genomics Core Lab, National Taiwan University Centers of Genomic and Precision Medicine | Shiou-Hwei Yeh et al |
| 1041 | Taiwan/TSGH-05/2020 | 2020-03-14 | TSGH-CP molecular lab | TSGH-CP molecular lab | Cherng-Lih Perng et al |
| 1042 | Taiwan/NTU08/2020 | 2020-03-14 | Department of Laboratory Medicine, National Taiwan University Hospital | Microbial Genomics Core Lab, National Taiwan University Centers of Genomic and Precision Medicine | Shiou-Hwei Yeh et al |
| 1043 | USA/MN63-MDH63/2020 | 2020-03-14 | Minnesota Department of Health, Public Health Laboratory | Minnesota Department of Health, Public Health Laboratory | Matt Plumb et al |
| 1044 | USA/NY-WCMP9B08P/2020 | 2020-03-14 | NewYork-Presbyterian & Mason Lab | Mason Lab | Daniel J. Butler et al |
| 1045 | Wales/PHWC-240E8/2020 | 2020-03-14 | Wales Specialist Virology Centre | Public Health Wales Microbiology Cardiff | Catherine Moore et al |
| 1046 | USA/WA-UW148/2020 | 2020-03-14 | UW Virology Lab | UW Virology Lab | Pavitra Roychoudhury et al |
| 1047 | USA/MN62-MDH62/2020 | 2020-03-14 | Minnesota Department of Health, Public Health Laboratory | Minnesota Department of Health, Public Health Laboratory | Matt Plumb et al |
| 1048 | Wales/PHWC-240F7/2020 | 2020-03-14 | Wales Specialist Virology Centre | Public Health Wales Microbiology Cardiff | Catherine Moore et al |
| 1049 | Fujian/IM3520002T/2020 | 2020-03-15 | Fujian Center for Disease Control and Prevention | Fujian Center for Disease Control and Prevention | Lin Qi et al |
| 1050 | Australia/VIC88/2020 | 2020-03-15 | Victorian Infectious Diseases Reference Laboratory (VIDRL) | Victorian Infectious Diseases Reference Laboratory and Microbiological Diagnostic Unit Public Health Laboratory, Doherty Institute | Caly L. et al |
| 1051 | Australia/VIC85/2020 | 2020-03-15 | Victorian Infectious Diseases Reference Laboratory (VIDRL) | Victorian Infectious Diseases Reference Laboratory and Microbiological Diagnostic Unit Public Health Laboratory, Doherty Institute | Caly L. et al |
| 1052 | Australia/VIC91/2020 | 2020-03-15 | Victorian Infectious Diseases Reference Laboratory | Victorian Infectious Diseases Reference Laboratory and | Caly L. et al |

| | | | | | |
|---|---|---|---|---|---|
| | | | (VIDRL) | Microbiological Diagnostic Unit Public Health Laboratory, Doherty Institute | |
| 1053 | Australia/WA04/2020 | 2020-03-15 | Department of Microbiology, PathWest QEII Medical Centre | Department of Microbiology, PathWest QEII Medical Centre | Chisha Sikazwe et al |
| 1054 | Australia/VIC139/2020 | 2020-03-15 | Victorian Infectious Diseases Reference Laboratory (VIDRL) | Victorian Infectious Diseases Reference Laboratory and Microbiological Diagnostic Unit Public Health Laboratory, Doherty Institute | Caly L. et al |
| 1055 | Australia/VIC86/2020 | 2020-03-15 | Victorian Infectious Diseases Reference Laboratory (VIDRL) | Victorian Infectious Diseases Reference Laboratory and Microbiological Diagnostic Unit Public Health Laboratory, Doherty Institute | Caly L. et al |
| 1056 | Australia/VIC87/2020 | 2020-03-15 | Victorian Infectious Diseases Reference Laboratory (VIDRL) | Victorian Infectious Diseases Reference Laboratory and Microbiological Diagnostic Unit Public Health Laboratory, Doherty Institute | Caly L. et al |
| 1057 | Australia/VIC90/2020 | 2020-03-15 | Victorian Infectious Diseases Reference Laboratory (VIDRL) | Victorian Infectious Diseases Reference Laboratory and Microbiological Diagnostic Unit Public Health Laboratory, Doherty Institute | Caly L. et al |
| 1058 | Australia/NSW112/2020 | 2020-03-15 | Centre for Infectious Diseases and Microbiology Public Health | NSW Health Pathology - Institute of Clinical Pathology and Medical Research; Westmead Hospital; University of Sydney | Bachmann N et al |
| 1059 | Australia/VIC08/2020 | 2020-03-15 | Victorian Infectious Diseases Reference Laboratory (VIDRL) | Victorian Infectious Diseases Reference Laboratory and Microbiological Diagnostic Unit Public Health Laboratory, Doherty Institute | Caly L. et al |
| 1060 | Australia/VIC92/2020 | 2020-03-15 | Victorian Infectious Diseases Reference Laboratory (VIDRL) | Victorian Infectious Diseases Reference Laboratory and Microbiological Diagnostic Unit Public Health Laboratory, Doherty Institute | Caly L. et al |
| 1061 | Australia/VIC89/2020 | 2020-03-15 | Victorian Infectious Diseases Reference Laboratory (VIDRL) | Victorian Infectious Diseases Reference Laboratory and Microbiological Diagnostic Unit Public Health Laboratory, Doherty Institute | Caly L. et al |
| 1062 | Hangzhou/HZCDC6789/2020 | 2020-03-15 | Hangzhou Center for Diseases Control and Prevention | Hangzhou Center for Diseases Control and Prevention | Jun Li et al |
| 1063 | USA/NY-WCMP10H08P/2020 | 2020-03-15 | NewYork-Presbyterian & Mason Lab | Mason Lab | Daniel J. Butler et al |
| 1064 | India/NCDC-01475/2020 | 2020-03-15 | National Centre for Disease control (NCDC), CSIR-Institute of Genomics and Integrative Biology (CSIR-IGIB) | NCDC/CSIR-IGIB | Pramod Kumar et al |
| 1065 | USA/NY-WCMP10B08P/2020 | 2020-03-15 | NewYork-Presbyterian & Mason Lab | Mason Lab | Daniel J. Butler et al |
| 1066 | USA/NY-WCMP10E07P/2020 | 2020-03-15 | NewYork-Presbyterian & Mason Lab | Mason Lab | Daniel J. Butler et al |
| 1067 | USA/WA-UW297/2020 | 2020-03-15 | UW Virology Lab | UW Virology Lab | Pavitra Roychoudhury et al |
| 1068 | Iceland/337/2020 | 2020-03-15 | deCODE genetics | deCODE genetics | Daniel F Gudbjartsson et al |
| 1069 | USA/WA-S286/2020 | 2020-03-15 | Seattle Flu Study | Seattle Flu Study | Chu et al et al |
| 1070 | Germany/NRW-22/2020 | 2020-03-15 | Center of Medical Microbiology, Virology, and Hospital Hygiene, University of Duesseldorf | Center of Medical Microbiology, Virology, and Hospital Hygiene, University of Duesseldorf | Ortwin Adams et al |
| 1071 | USA/WA-UW286/2020 | 2020-03-15 | UW Virology Lab | UW Virology Lab | Pavitra Roychoudhury et al |
| 1072 | USA/AK-PHL002/2020 | 2020-03-15 | Alaska State Virology Laboratory | Alaska State Virology Laboratory | Chen et al |

| | | | | | |
|---|---|---|---|---|---|
| 1073 | NewZealand/20VR0275/2020-03-15 | 2020-03-15 | Wellington SCL (WN) | Institute of Environmental Science and Research (ESR) Ltd | **Matt Storey et al** |
| 1074 | Denmark/ALAB-HH87/2020 | 2020-03-15 | Department of Clinical Microbiology, Copenhagen University Hospital, Hvidovre, Kettegaard Alle 30, 2650 Hvidovre. | Albertsen lab, Department of Chemistry and Bioscience, Aalborg University, Denmark | Rasmus Kirkegaard et al |
| 1075 | USA/AK-PHL003/2020 | 2020-03-15 | Alaska State Virology Laboratory | Alaska State Virology Laboratory | Chen et al |
| 1076 | Portugal/PT0024/2020 | 2020-03-15 | HSE Ilha Terceira - Angra do Heroismo | Instituto Nacional de Saude (INSA) | Guiomar et al et al |
| 1077 | Mexico/CDMX-INER_04/2020 | 2020-03-15 | Instituto Nacional de Enfermedades Respiratorias | Instituto Nacional de Enfermedades Respiratorias | Joel Armando Vázquez Pérez et al |
| 1078 | USA/WA-UW321/2020 | 2020-03-15 | UW Virology Lab | UW Virology Lab | Pavitra Roychoudhury et al |
| 1079 | Luxembourg/LNS4845603/2020 | 2020-03-15 | Laboratoire National de Santé, Microbiology, Virology | Laboratoire National de Santé, Microbiology, Epidemiology and Microbial Genomics | Anke Wienecke-Baldacchino et al |
| 1080 | USA/WA-S144/2020 | 2020-03-15 | Washington State Department of Health | Seattle Flu Study | Chu et al et al |
| 1081 | Spain/Valencia129/2020 | 2020-03-15 | Servicio de Microbiología. Consorcio Hospital General Universitario de Valencia | Sequencing and Bioinformatics Service and Molecular Epidemiology Research Group. FISABIO-Public Health | Neris Garcia-Gonzalez et al |
| 1082 | Denmark/ALAB-HH82/2020 | 2020-03-15 | Department of Clinical Microbiology, Copenhagen University Hospital, Hvidovre, Kettegaard Alle 30, 2650 Hvidovre. | Albertsen lab, Department of Chemistry and Bioscience, Aalborg University, Denmark | Rasmus Kirkegaard et al |
| 1083 | Iceland/167/2020 | 2020-03-15 | The National University Hospital of Iceland | deCODE genetics | Daniel F Gudbjartsson et al |
| 1084 | Iceland/339/2020 | 2020-03-15 | deCODE genetics | deCODE genetics | Daniel F Gudbjartsson et al |
| 1085 | Iceland/8/2020 | 2020-03-15 | The National University Hospital of Iceland | deCODE genetics | Daniel F Gudbjartsson et al |
| 1086 | Iceland/173/2020 | 2020-03-15 | The National University Hospital of Iceland | deCODE genetics | Daniel F Gudbjartsson et al |
| 1087 | Iceland/178/2020 | 2020-03-15 | The National University Hospital of Iceland | deCODE genetics | Daniel F Gudbjartsson et al |
| 1088 | Iceland/338/2020 | 2020-03-15 | deCODE genetics | deCODE genetics | Daniel F Gudbjartsson et al |
| 1089 | Iceland/155/2020 | 2020-03-15 | The National University Hospital of Iceland | deCODE genetics | Daniel F Gudbjartsson et al |
| 1090 | Iceland/341/2020 | 2020-03-15 | deCODE genetics | deCODE genetics | Daniel F Gudbjartsson et al |
| 1091 | Iceland/165/2020 | 2020-03-15 | The National University Hospital of Iceland | deCODE genetics | Daniel F Gudbjartsson et al |
| 1092 | Iceland/7/2020 | 2020-03-15 | The National University Hospital of Iceland | deCODE genetics | Daniel F Gudbjartsson et al |
| 1093 | Iceland/177/2020 | 2020-03-15 | The National University Hospital of Iceland | deCODE genetics | Daniel F Gudbjartsson et al |
| 1094 | Iceland/192/2020 | 2020-03-15 | The National University Hospital of Iceland | deCODE genetics | Daniel F Gudbjartsson et al |
| 1095 | Iceland/172/2020 | 2020-03-15 | The National University Hospital of Iceland | deCODE genetics | Daniel F Gudbjartsson et al |
| 1096 | USA/NY-WCMP10B12P/2020 | 2020-03-15 | NewYork-Presbyterian & Mason Lab | Mason Lab | Daniel J. Butler et al |
| 1097 | Germany/NRW-28/2020 | 2020-03-15 | Center of Medical Microbiology, Virology, and Hospital Hygiene, University of Duesseldorf | Center of Medical Microbiology, Virology, and Hospital Hygiene, University of Duesseldorf | Ortwin Adams et al |
| 1098 | USA/NY-WCMP10A12P/2020 | 2020-03-15 | NewYork-Presbyterian & Mason Lab | Mason Lab | Daniel J. Butler et al |
| 1099 | USA/WY-WYPHL008/2020 | 2020-03-15 | Wyoming Public Health Laboratory | Center for Global Health, University of New Mexico Health Sciences Center | Daryl Domman et al |
| 1100 | Sweden/RV-FOI-1/2020 | 2020-03-15 | Klinisk mikrobiologi, Region Västerbotten | Unit for Biological Agents, Department for CBRN Defence | FOI Bioinformatics team et al |

| | | | | | |
|---|---|---|---|---|---|
| | | | | and Security, Swedish Defence Research Agency | |
| 1101 | USA/WA-UW179/2020 | 2020-03-15 | UW Virology Lab | UW Virology Lab | Pavitra Roychoudhury et al |
| 1102 | Taiwan/TSGH-06/2020 | 2020-03-15 | TSGH-CP molecular lab | TSGH-CP molecular lab | Cherng-Lih Perng et al |
| 1103 | Taiwan/58/2020 | 2020-03-15 | Centers for Disease Control, R.O.C. (Taiwan) | Centers for Disease Control, R.O.C. (Taiwan) | Ji-Rong Yang et al |
| 1104 | Taiwan/NTU10/2020 | 2020-03-15 | Department of Laboratory Medicine, National Taiwan University Hospital | Microbial Genomics Core Lab, National Taiwan University Centers of Genomic and Precision Medicine | Shiou-Hwei Yeh et al |
| 1105 | USA/WA-UW292/2020 | 2020-03-15 | UW Virology Lab | UW Virology Lab | Pavitra Roychoudhury et al |
| 1106 | Vietnam/OUCRU0837/2020 | 2020-03-15 | National Hospital of Tropical Diseases | Oxford University Clinical Research Unit, Hanoi, Vietnam | Nguyen Thi Tam et al |
| 1107 | Fujian/IM3520003T/2020 | 2020-03-16 | Fujian Center for Disease Control and Prevention | Fujian Center for Disease Control and Prevention | Lin Qi et al |
| 1108 | USA/WA-UW388/2020 | 2020-03-16 | UW Virology Lab | UW Virology Lab | Pavitra Roychoudhury et al |
| 1109 | Netherlands/NA_168/2020 | 2020-03-16 | Dutch COVID-19 response team | Erasmus Medical Center | Bas Oude Munnink et al |
| 1110 | Australia/VIC99/2020 | 2020-03-16 | Victorian Infectious Diseases Reference Laboratory (VIDRL) | Victorian Infectious Diseases Reference Laboratory and Microbiological Diagnostic Unit Public Health Laboratory, Doherty Institute | Caly L. et al |
| 1111 | Australia/VIC100/2020 | 2020-03-16 | Victorian Infectious Diseases Reference Laboratory (VIDRL) | Victorian Infectious Diseases Reference Laboratory and Microbiological Diagnostic Unit Public Health Laboratory, Doherty Institute | Caly L. et al |
| 1112 | Australia/NSW190/2020 | 2020-03-16 | Centre for Infectious Diseases and Microbiology Public Health | NSW Health Pathology - Institute of Clinical Pathology and Medical Research; Westmead Hospital; University of Sydney | Draper J et al |
| 1113 | Australia/VIC95/2020 | 2020-03-16 | Victorian Infectious Diseases Reference Laboratory (VIDRL) | Victorian Infectious Diseases Reference Laboratory and Microbiological Diagnostic Unit Public Health Laboratory, Doherty Institute | Caly L. et al |
| 1114 | Australia/VIC102/2020 | 2020-03-16 | Victorian Infectious Diseases Reference Laboratory (VIDRL) | Victorian Infectious Diseases Reference Laboratory and Microbiological Diagnostic Unit Public Health Laboratory, Doherty Institute | Caly L. et al |
| 1115 | Australia/NSW111/2020 | 2020-03-16 | Centre for Infectious Diseases and Microbiology Public Health | NSW Health Pathology - Institute of Clinical Pathology and Medical Research; Westmead Hospital; University of Sydney | Sim E et al |
| 1116 | Australia/VIC98/2020 | 2020-03-16 | Victorian Infectious Diseases Reference Laboratory (VIDRL) | Victorian Infectious Diseases Reference Laboratory and Microbiological Diagnostic Unit Public Health Laboratory, Doherty Institute | Caly L. et al |
| 1117 | Wales/PHWC-2419A/2020 | 2020-03-16 | Wales Specialist Virology Centre | Public Health Wales Microbiology Cardiff | Catherine Moore et al |
| 1118 | USA/NY-WCMP11E05P/2020 | 2020-03-16 | NewYork-Presbyterian & Mason Lab | Mason Lab | Daniel J. Butler et al |
| 1119 | USA/TX-HMH03/2020 | 2020-03-16 | Houston Methodist Hospital | Houston Methodist Hospital | S. Wesley Long et al |
| 1120 | Iceland/100/2020 | 2020-03-16 | deCODE genetics | deCODE genetics | Daniel F Gudbjartsson et al |
| 1121 | Australia/NSW75/2020 | 2020-03-16 | Centre for Infectious Diseases and Microbiology Public Health | NSW Health Pathology - Institute of Clinical Pathology and Medical Research; Westmead Hospital; University of Sydney | Gray K et al |
| 1122 | Australia/NSW104/2020 | 2020-03-16 | Centre for Infectious Diseases and Microbiology Public Health | NSW Health Pathology - Institute of Clinical Pathology and Medical Research; | Lam C et al |

| | | | | | |
|---|---|---|---|---|---|
| | | | | Westmead Hospital; University of Sydney | |
| 1123 | Australia/NSW88/2020 | 2020-03-16 | Centre for Infectious Diseases and Microbiology Public Health | NSW Health Pathology - Institute of Clinical Pathology and Medical Research; Westmead Hospital; University of Sydney | Arnott A et al |
| 1124 | Australia/VIC140/2020 | 2020-03-16 | Victorian Infectious Diseases Reference Laboratory (VIDRL) | Victorian Infectious Diseases Reference Laboratory and Microbiological Diagnostic Unit Public Health Laboratory, Doherty Institute | Caly L. et al |
| 1125 | Australia/VIC141/2020 | 2020-03-16 | Victorian Infectious Diseases Reference Laboratory (VIDRL) | Victorian Infectious Diseases Reference Laboratory and Microbiological Diagnostic Unit Public Health Laboratory, Doherty Institute | Caly L. et al |
| 1126 | Australia/NSW57/2020 | 2020-03-16 | Centre for Infectious Diseases and Microbiology Public Health | NSW Health Pathology - Institute of Clinical Pathology and Medical Research; Westmead Hospital; University of Sydney | Rahman H et al |
| 1127 | England/NOTT-10E001/2020 | 2020-03-16 | Queens Medical Centre, Clinical Microbiology Department / DeepSeq Nottingham | COVID-19 Genomics UK (COG-UK) Consortium | Gemma Clark et al |
| 1128 | USA/WA-UW245/2020 | 2020-03-16 | UW Virology Lab | UW Virology Lab | Pavitra Roychoudhury et al |
| 1129 | Iceland/210/2020 | 2020-03-16 | The National University Hospital of Iceland | deCODE genetics | Daniel F Gudbjartsson et al |
| 1130 | NewZealand/20VR0276/2020-03-16 | 2020-03-16 | Wellington SCL (WN) | Institute of Environmental Science and Research (ESR) Ltd | **Matt Storey et al** |
| 1131 | NewZealand/20VR0462/2020-03-16 | 2020-03-16 | Southern Community Labs Dunedin | Institute of Environmental Science and Research (ESR) Ltd | **Matt Storey et al** |
| 1132 | NewZealand/20VR0993/2020-03-16 | 2020-03-16 | LabPLUS | Institute of Environmental Science and Research (ESR) Ltd | **Matt Storey et al** |
| 1133 | Italy/TE4836/2020 | 2020-03-16 | Ospedale Civile Giuseppe Mazzini | "Istituto Zooprofilattico Sperimentale dell'Abruzzo e Molise ""G. Caporale""" | Lorusso A et al |
| 1134 | Jordan/SR-044/2020 | 2020-03-16 | Biolab Diagnostic Laboratories | Andersen lab at Scripps Research | Issa Abu-Dayyeh et al |
| 1135 | USA/WA-UW248/2020 | 2020-03-16 | UW Virology Lab | UW Virology Lab | Pavitra Roychoudhury et al |
| 1136 | Denmark/ALAB-HH84/2020 | 2020-03-16 | Department of Clinical Microbiology, Copenhagen University Hospital, Hvidovre, Kettegaard Alle 30, 2650 Hvidovre. | Albertsen lab, Department of Chemistry and Bioscience, Aalborg University, Denmark | Rasmus Kirkegaard et al |
| 1137 | Iceland/204/2020 | 2020-03-16 | The National University Hospital of Iceland | deCODE genetics | Daniel F Gudbjartsson et al |
| 1138 | USA/WA-UW371/2020 | 2020-03-16 | UW Virology Lab | UW Virology Lab | Pavitra Roychoudhury et al |
| 1139 | USA/NY-NYUMC7/2020 | 2020-03-16 | NYU Langone Health | Department of Pathology and Medicine, New York University School of Medicine | Margaret Black et al |
| 1140 | Singapore/46/2020 | 2020-03-16 | National Public Health Laboratory, National Centre for Infectious Diseases | National Public Health Laboratory, National Centre for Infectious Diseases | Mak TM et al |
| 1141 | Iceland/203/2020 | 2020-03-16 | The National University Hospital of Iceland | deCODE genetics | Daniel F Gudbjartsson et al |
| 1142 | Iceland/195/2020 | 2020-03-16 | The National University Hospital of Iceland | deCODE genetics | Daniel F Gudbjartsson et al |
| 1143 | Iceland/245/2020 | 2020-03-16 | The National University Hospital of Iceland | deCODE genetics | Daniel F Gudbjartsson et al |
| 1144 | Iceland/2/2020 | 2020-03-16 | deCODE genetics | deCODE genetics | Daniel F Gudbjartsson et al |
| 1145 | Iceland/198/2020 | 2020-03-16 | The National University Hospital of Iceland | deCODE genetics | Daniel F Gudbjartsson et al |
| 1146 | Iceland/10/2020 | 2020-03-16 | deCODE genetics | deCODE genetics | Daniel F Gudbjartsson et al |

| | | | | | |
|---|---|---|---|---|---|
| 1147 | Iceland/175/2020 | 2020-03-16 | The National University Hospital of Iceland | deCODE genetics | Daniel F Gudbjartsson et al |
| 1148 | Iceland/193/2020 | 2020-03-16 | The National University Hospital of Iceland | deCODE genetics | Daniel F Gudbjartsson et al |
| 1149 | Iceland/197/2020 | 2020-03-16 | The National University Hospital of Iceland | deCODE genetics | Daniel F Gudbjartsson et al |
| 1150 | Iceland/98/2020 | 2020-03-16 | deCODE genetics | deCODE genetics | Daniel F Gudbjartsson et al |
| 1151 | Iceland/235/2020 | 2020-03-16 | The National University Hospital of Iceland | deCODE genetics | Daniel F Gudbjartsson et al |
| 1152 | Iceland/226/2020 | 2020-03-16 | The National University Hospital of Iceland | deCODE genetics | Daniel F Gudbjartsson et al |
| 1153 | Iceland/183/2020 | 2020-03-16 | The National University Hospital of Iceland | deCODE genetics | Daniel F Gudbjartsson et al |
| 1154 | Iceland/220/2020 | 2020-03-16 | The National University Hospital of Iceland | deCODE genetics | Daniel F Gudbjartsson et al |
| 1155 | Iceland/218/2020 | 2020-03-16 | The National University Hospital of Iceland | deCODE genetics | Daniel F Gudbjartsson et al |
| 1156 | Iceland/213/2020 | 2020-03-16 | The National University Hospital of Iceland | deCODE genetics | Daniel F Gudbjartsson et al |
| 1157 | Iceland/196/2020 | 2020-03-16 | The National University Hospital of Iceland | deCODE genetics | Daniel F Gudbjartsson et al |
| 1158 | Iceland/224/2020 | 2020-03-16 | The National University Hospital of Iceland | deCODE genetics | Daniel F Gudbjartsson et al |
| 1159 | Iceland/29/2020 | 2020-03-16 | deCODE genetics | deCODE genetics | Daniel F Gudbjartsson et al |
| 1160 | Iceland/217/2020 | 2020-03-16 | The National University Hospital of Iceland | deCODE genetics | Daniel F Gudbjartsson et al |
| 1161 | Germany/NRW-33/2020 | 2020-03-16 | Center of Medical Microbiology, Virology, and Hospital Hygiene, University of Duesseldorf | Center of Medical Microbiology, Virology, and Hospital Hygiene, University of Duesseldorf | Ortwin Adams et al |
| 1162 | Belgium/ULG-7500/2020 | 2020-03-16 | Department of Clinical Microbiology | GIGA Medical Genomics | Keith Durkin et al |
| 1163 | USA/WA-UW336/2020 | 2020-03-16 | UW Virology Lab | UW Virology Lab | Pavitra Roychoudhury et al |
| 1164 | Iceland/205/2020 | 2020-03-16 | The National University Hospital of Iceland | deCODE genetics | Daniel F Gudbjartsson et al |
| 1165 | Iceland/200/2020 | 2020-03-16 | The National University Hospital of Iceland | deCODE genetics | Daniel F Gudbjartsson et al |
| 1166 | Iceland/202/2020 | 2020-03-16 | The National University Hospital of Iceland | deCODE genetics | Daniel F Gudbjartsson et al |
| 1167 | Iceland/186/2020 | 2020-03-16 | The National University Hospital of Iceland | deCODE genetics | Daniel F Gudbjartsson et al |
| 1168 | Iceland/219/2020 | 2020-03-16 | The National University Hospital of Iceland | deCODE genetics | Daniel F Gudbjartsson et al |
| 1169 | Iceland/184/2020 | 2020-03-16 | The National University Hospital of Iceland | deCODE genetics | Daniel F Gudbjartsson et al |
| 1170 | Iceland/182/2020 | 2020-03-16 | The National University Hospital of Iceland | deCODE genetics | Daniel F Gudbjartsson et al |
| 1171 | Iceland/179/2020 | 2020-03-16 | The National University Hospital of Iceland | deCODE genetics | Daniel F Gudbjartsson et al |
| 1172 | Iceland/181/2020 | 2020-03-16 | The National University Hospital of Iceland | deCODE genetics | Daniel F Gudbjartsson et al |
| 1173 | Iceland/180/2020 | 2020-03-16 | The National University Hospital of Iceland | deCODE genetics | Daniel F Gudbjartsson et al |
| 1174 | Iceland/191/2020 | 2020-03-16 | The National University Hospital of Iceland | deCODE genetics | Daniel F Gudbjartsson et al |
| 1175 | France/ARA09686/2020 | 2020-03-16 | Centre Hospitalier de Valence | CNR Virus des Infections Respiratoires - France SUD | Antonin Bal et al |
| 1176 | Iceland/212/2020 | 2020-03-16 | The National University Hospital of Iceland | deCODE genetics | Daniel F Gudbjartsson et al |
| 1177 | Iceland/216/2020 | 2020-03-16 | The National University Hospital of Iceland | deCODE genetics | Daniel F Gudbjartsson et al |
| 1178 | Iceland/222/2020 | 2020-03-16 | The National University Hospital of Iceland | deCODE genetics | Daniel F Gudbjartsson et al |
| 1179 | Iceland/199/2020 | 2020-03-16 | The National University | deCODE genetics | Daniel F Gudbjartsson |

| | | | | | |
|---|---|---|---|---|---|
| | | | Hospital of Iceland | | et al |
| 1180 | Iceland/189/2020 | 2020-03-16 | The National University Hospital of Iceland | deCODE genetics | Daniel F Gudbjartsson et al |
| 1181 | Iceland/187/2020 | 2020-03-16 | The National University Hospital of Iceland | deCODE genetics | Daniel F Gudbjartsson et al |
| 1182 | Iceland/221/2020 | 2020-03-16 | The National University Hospital of Iceland | deCODE genetics | Daniel F Gudbjartsson et al |
| 1183 | Iceland/185/2020 | 2020-03-16 | The National University Hospital of Iceland | deCODE genetics | Daniel F Gudbjartsson et al |
| 1184 | Taiwan/78/2020 | 2020-03-16 | Centers for Disease Control, R.O.C. (Taiwan) | Centers for Disease Control, R.O.C. (Taiwan) | Ji-Rong Yang et al |
| 1185 | Taiwan/CGMH-CGU-16/2020 | 2020-03-16 | Laboratory Medicine | Department of Laboratory Medicine, Lin-Kou Chang Gung Memorial Hospital, Taoyuan, Taiwan | Kuo-Chien Tsao et al |
| 1186 | Taiwan/NTU11/2020 | 2020-03-16 | Department of Laboratory Medicine, National Taiwan University Hospital | Microbial Genomics Core Lab, National Taiwan University Centers of Genomic and Precision Medicine | Shiou-Hwei Yeh et al |
| 1187 | Taiwan/TSGH-28/2020 | 2020-03-16 | TSGH-CP molecular lab | TSGH-CP molecular lab | Cherng-Lih Perng et al |
| 1188 | Taiwan/NTU12/2020 | 2020-03-16 | Department of Laboratory Medicine, National Taiwan University Hospital | Microbial Genomics Core Lab, National Taiwan University Centers of Genomic and Precision Medicine | Shiou-Hwei Yeh et al |
| 1189 | USA/WA-UW322/2020 | 2020-03-16 | UW Virology Lab | UW Virology Lab | Pavitra Roychoudhury et al |
| 1190 | USA/WA-UW337/2020 | 2020-03-16 | UW Virology Lab | UW Virology Lab | Pavitra Roychoudhury et al |
| 1191 | Iceland/174/2020 | 2020-03-16 | The National University Hospital of Iceland | deCODE genetics | Daniel F Gudbjartsson et al |
| 1192 | USA/NY-WCMP12F07P/2020 | 2020-03-16 | NewYork-Presbyterian & Mason Lab | Mason Lab | Daniel J. Butler et al |
| 1193 | France/IDF2684/2020 | 2020-03-16 | LABM GH nord Essonne | National Reference Center for Viruses of Respiratory Infections, Institut Pasteur, Paris | Mélanie Albert et al |
| 1194 | Fujian/IM3520010T/2020 | 2020-03-17 | Fujian Center for Disease Control and Prevention | Fujian Center for Disease Control and Prevention | Lin Qi et al |
| 1195 | USA/WA-S161/2020 | 2020-03-17 | Washington State Department of Health | Seattle Flu Study | Chu et al et al |
| 1196 | England/20124036306/2020 | 2020-03-17 | Respiratory Virus Unit, Microbiology Services Colindale, Public Health England | Respiratory Virus Unit, Microbiology Services Colindale, Public Health England | Monica Galiano et al |
| 1197 | Turkey/HSGM-4706/2020 | 2020-03-17 | Ministry of Health Turkey | Ministry of Health Turkey | Fatma Bayrakdar et al |
| 1198 | Australia/NSW62/2020 | 2020-03-17 | Centre for Infectious Diseases and Microbiology Public Health | NSW Health Pathology - Institute of Clinical Pathology and Medical Research; Westmead Hospital; University of Sydney | Sintchenko V et al |
| 1199 | Australia/VIC327/2020 | 2020-03-17 | Victorian Infectious Diseases Reference Laboratory (VIDRL) | Microbiological Diagnostic Unit Public Health Laboratory and Victorian Infectious Diseases Reference Laboratory, Doherty Institute | Caly L. et al |
| 1200 | Australia/VIC329/2020 | 2020-03-17 | Victorian Infectious Diseases Reference Laboratory (VIDRL) | Microbiological Diagnostic Unit Public Health Laboratory and Victorian Infectious Diseases Reference Laboratory, Doherty Institute | Caly L. et al |
| 1201 | Australia/NSW140/2020 | 2020-03-17 | Centre for Infectious Diseases and Microbiology Public Health | NSW Health Pathology - Institute of Clinical Pathology and Medical Research; Westmead Hospital; University of Sydney | Draper J et al |
| 1202 | Australia/VIC152/2020 | 2020-03-17 | Victorian Infectious Diseases Reference Laboratory (VIDRL) | Victorian Infectious Diseases Reference Laboratory and Microbiological Diagnostic Unit Public Health Laboratory, Doherty Institute | Caly L. et al |
| 1203 | Spain/Valencia288/2020 | 2020-03-17 | Servicio de Microbiología. | Sequencing and Bioinformatics | Maria Dolores Ocete |

| | | | | | |
|---|---|---|---|---|---|
| | | | Consorcio Hospital General Universitario de Valencia | Service and Molecular Epidemiology Research Group. FISABIO-Public Health | et al |
| 1204 | Australia/NSW63/2020 | 2020-03-17 | Centre for Infectious Diseases and Microbiology Public Health | NSW Health Pathology - Institute of Clinical Pathology and Medical Research; Westmead Hospital; University of Sydney | Timms V et al |
| 1205 | Australia/VIC155/2020 | 2020-03-17 | Victorian Infectious Diseases Reference Laboratory (VIDRL) | Victorian Infectious Diseases Reference Laboratory and Microbiological Diagnostic Unit Public Health Laboratory, Doherty Institute | Caly L. et al |
| 1206 | Australia/NSW215/2020 | 2020-03-17 | Centre for Infectious Diseases and Microbiology Public Health | NSW Health Pathology - Institute of Clinical Pathology and Medical Research; Westmead Hospital; University of Sydney | Gray K et al |
| 1207 | Netherlands/NoordBrabant_95/2020 | 2020-03-17 | Dutch COVID-19 response team | Erasmus Medical Center | Bas Oude Munnink et al |
| 1208 | Spain/Valencia94/2020 | 2020-03-17 | Servicio de Microbiología. Consorcio Hospital General Universitario de Valencia | Sequencing and Bioinformatics Service and Molecular Epidemiology Research Group. FISABIO-Public Health | Neris Garcia-Gonzalez et al |
| 1209 | England/20122119302/2020 | 2020-03-17 | Respiratory Virus Unit, Microbiology Services Colindale, Public Health England | Respiratory Virus Unit, Microbiology Services Colindale, Public Health England | Monica Galiano et al |
| 1210 | Iceland/116/2020 | 2020-03-17 | deCODE genetics | deCODE genetics | Daniel F Gudbjartsson et al |
| 1211 | Taiwan/CGMH-CGU-15/2020 | 2020-03-17 | Laboratory Medicine | Department of Laboratory Medicine, Lin-Kou Chang Gung Memorial Hospital, Taoyuan, Taiwan | Kuo-Chien Tsao et al |
| 1212 | Taiwan/TSGH-13/2020 | 2020-03-17 | TSGH-CP molecular lab | TSGH-CP molecular lab | Cherng-Lih Perng et al |
| 1213 | Scotland/CVR164/2020 | 2020-03-17 | West of Scotland Specialist Virology Centre, NHSGGC / MRC-University of Glasgow Centre for Virus Research | COVID-19 Genomics UK (COG-UK) Consortium | Ana da Silva Filipe et al |
| 1214 | Iceland/242/2020 | 2020-03-17 | The National University Hospital of Iceland | deCODE genetics | Daniel F Gudbjartsson et al |
| 1215 | Australia/NSW59/2020 | 2020-03-17 | Centre for Infectious Diseases and Microbiology Public Health | NSW Health Pathology - Institute of Clinical Pathology and Medical Research; Westmead Hospital; University of Sydney | Lam C et al |
| 1216 | Australia/VIC148/2020 | 2020-03-17 | Victorian Infectious Diseases Reference Laboratory (VIDRL) | Victorian Infectious Diseases Reference Laboratory and Microbiological Diagnostic Unit Public Health Laboratory, Doherty Institute | Caly L. et al |
| 1217 | Australia/NSW60/2020 | 2020-03-17 | Centre for Infectious Diseases and Microbiology Public Health | NSW Health Pathology - Institute of Clinical Pathology and Medical Research; Westmead Hospital; University of Sydney | Timms V et al |
| 1218 | Italy/TE5553/2020 | 2020-03-17 | Presidio Ospedaliero Santo Spirito | "Istituto Zooprofilattico Sperimentale dell'Abruzzo e Molise ""G. Caporale""" | Lorusso A et al |
| 1219 | Italy/TE5551/2020 | 2020-03-17 | Presidio Ospedaliero Santo Spirito | "Istituto Zooprofilattico Sperimentale dell'Abruzzo e Molise ""G. Caporale""" | Lorusso A et al |
| 1220 | Norway/2088/2020 | 2020-03-17 | University Hospital of Northern Norway, Department for Microbiology and Infectious Disease Control | Norwegian Institute of Public Health, Department of Virology | Kathrine Stene-Johansen et al |
| 1221 | Senegal/315/2020 | 2020-03-17 | Institut Pasteur Dakar | Institut Pasteur de Dakar | Ndongo Dia et al |
| 1222 | Australia/VIC146/2020 | 2020-03-17 | Victorian Infectious Diseases Reference Laboratory (VIDRL) | Victorian Infectious Diseases Reference Laboratory and Microbiological Diagnostic Unit Public Health Laboratory, Doherty Institute | Caly L. et al |

| | | | | | |
|---|---|---|---|---|---|
| 1223 | Australia/NSW113/2020 | 2020-03-17 | Centre for Infectious Diseases and Microbiology Public Health | NSW Health Pathology - Institute of Clinical Pathology and Medical Research; Westmead Hospital; University of Sydney | Rockett R et al |
| 1224 | Luxembourg/LNS0814536/2020 | 2020-03-17 | Laboratoire National de Sante, Microbiology, Virology | Laboratoire National de Sante, Microbiology, Epidemiology and Microbial Genomics | Anke Wienecke-Baldacchino et al |
| 1225 | USA/WA-UW364/2020 | 2020-03-17 | UW Virology Lab | UW Virology Lab | Pavitra Roychoudhury et al |
| 1226 | USA/NY-NYUMC30/2020 | 2020-03-17 | NYU Langone Health | Department of Pathology and Medicine, New York University School of Medicine | Maria Aguero-Rosenfeld et al |
| 1227 | NewZealand/20VR0987/2020-03-17 | 2020-03-17 | LabPLUS | Institute of Environmental Science and Research (ESR) Ltd | **Matt Storey et al** |
| 1228 | NewZealand/20VR0635/2020-03-17 | 2020-03-17 | Southern Community Labs Dunedin | Institute of Environmental Science and Research (ESR) Ltd | **Matt Storey et al** |
| 1229 | NewZealand/20VR0514/2020-03-17 | 2020-03-17 | Wellington SCL (WN) | Institute of Environmental Science and Research (ESR) Ltd | **Matt Storey et al** |
| 1230 | NewZealand/20VR0465/2020-03-17 | 2020-03-17 | Waikato Hospital | Institute of Environmental Science and Research (ESR) Ltd | **Matt Storey et al** |
| 1231 | NewZealand/20VR0636/2020-03-17 | 2020-03-17 | Southern Community Labs Dunedin | Institute of Environmental Science and Research (ESR) Ltd | **Matt Storey et al** |
| 1232 | NewZealand/20VR0989/2020-03-17 | 2020-03-17 | LabPLUS | Institute of Environmental Science and Research (ESR) Ltd | **Matt Storey et al** |
| 1233 | Scotland/CVR153/2020 | 2020-03-17 | West of Scotland Specialist Virology Centre, NHSGGC / MRC-University of Glasgow Centre for Virus Research | COVID-19 Genomics UK (COG-UK) Consortium | Ana da Silva Filipe et al |
| 1234 | Luxembourg/LNS1644489/2020 | 2020-03-17 | Laboratoire National de Sante, Microbiology, Virology | Laboratoire National de Sante, Microbiology, Epidemiology and Microbial Genomics | Anke Wienecke-Baldacchino et al |
| 1235 | USA/AZ-TG268281/2020 | 2020-03-17 | TGen North | TGen North | Jolene Bowers et al |
| 1236 | Italy/TE5056/2020 | 2020-03-17 | Ospedale Civile Giuseppe Mazzini, Teramo | "Istituto Zooprofilattico Sperimentale dell'Abruzzo e Molise ""G.Caporale""" | Lorusso A et al |
| 1237 | CzechRepublic/Seq6/2020 | 2020-03-17 | Laboratory of Molecular Genetics, 2nd Faculty of Medicine, Charles University in Prague, Prague, Czech Republic | Laboratory of Molecular Genetics, 2nd Faculty of Medicine, Charles University in Prague, Prague, Czech Republic | Lenka Kramna et al |
| 1238 | Luxembourg/LNS7991123/2020 | 2020-03-17 | Laboratoire National de Sante, Microbiology, Virology | Laboratoire National de Sante, Microbiology, Epidemiology and Microbial Genomics | Anke Wienecke-Baldacchino et al |
| 1239 | France/ARA10192/2020 | 2020-03-17 | Institut des Agents Infectieux (IAI), Hospices Civils de Lyon | CNR Virus des Infections Respiratoires - France SUD | Antonin Bal et al |
| 1240 | Iceland/264/2020 | 2020-03-17 | The National University Hospital of Iceland | deCODE genetics | Daniel F Gudbjartsson et al |
| 1241 | Iceland/269/2020 | 2020-03-17 | The National University Hospital of Iceland | deCODE genetics | Daniel F Gudbjartsson et al |
| 1242 | Iceland/230/2020 | 2020-03-17 | The National University Hospital of Iceland | deCODE genetics | Daniel F Gudbjartsson et al |
| 1243 | Iceland/283/2020 | 2020-03-17 | The National University Hospital of Iceland | deCODE genetics | Daniel F Gudbjartsson et al |
| 1244 | Iceland/227/2020 | 2020-03-17 | The National University Hospital of Iceland | deCODE genetics | Daniel F Gudbjartsson et al |
| 1245 | Iceland/237/2020 | 2020-03-17 | The National University Hospital of Iceland | deCODE genetics | Daniel F Gudbjartsson et al |
| 1246 | Iceland/275/2020 | 2020-03-17 | The National University Hospital of Iceland | deCODE genetics | Daniel F Gudbjartsson et al |
| 1247 | Iceland/285/2020 | 2020-03-17 | The National University Hospital of Iceland | deCODE genetics | Daniel F Gudbjartsson et al |
| 1248 | Iceland/120/2020 | 2020-03-17 | deCODE genetics | deCODE genetics | Daniel F Gudbjartsson et al |
| 1249 | Iceland/246/2020 | 2020-03-17 | The National University Hospital of Iceland | deCODE genetics | Daniel F Gudbjartsson et al |
| 1250 | Iceland/118/2020 | 2020-03-17 | deCODE genetics | deCODE genetics | Daniel F Gudbjartsson et al |

| | | | | | |
|---|---|---|---|---|---|
| 1251 | Iceland/119/2020 | 2020-03-17 | deCODE genetics | deCODE genetics | Daniel F Gudbjartsson et al |
| 1252 | Iceland/239/2020 | 2020-03-17 | The National University Hospital of Iceland | deCODE genetics | Daniel F Gudbjartsson et al |
| 1253 | Iceland/243/2020 | 2020-03-17 | The National University Hospital of Iceland | deCODE genetics | Daniel F Gudbjartsson et al |
| 1254 | Iceland/255/2020 | 2020-03-17 | The National University Hospital of Iceland | deCODE genetics | Daniel F Gudbjartsson et al |
| 1255 | Iceland/232/2020 | 2020-03-17 | The National University Hospital of Iceland | deCODE genetics | Daniel F Gudbjartsson et al |
| 1256 | Iceland/247/2020 | 2020-03-17 | The National University Hospital of Iceland | deCODE genetics | Daniel F Gudbjartsson et al |
| 1257 | Iceland/254/2020 | 2020-03-17 | The National University Hospital of Iceland | deCODE genetics | Daniel F Gudbjartsson et al |
| 1258 | Iceland/233/2020 | 2020-03-17 | The National University Hospital of Iceland | deCODE genetics | Daniel F Gudbjartsson et al |
| 1259 | Iceland/170/2020 | 2020-03-17 | deCODE genetics | deCODE genetics | Daniel F Gudbjartsson et al |
| 1260 | Iceland/281/2020 | 2020-03-17 | The National University Hospital of Iceland | deCODE genetics | Daniel F Gudbjartsson et al |
| 1261 | Iceland/284/2020 | 2020-03-17 | The National University Hospital of Iceland | deCODE genetics | Daniel F Gudbjartsson et al |
| 1262 | Iceland/272/2020 | 2020-03-17 | The National University Hospital of Iceland | deCODE genetics | Daniel F Gudbjartsson et al |
| 1263 | Iceland/266/2020 | 2020-03-17 | The National University Hospital of Iceland | deCODE genetics | Daniel F Gudbjartsson et al |
| 1264 | Iceland/270/2020 | 2020-03-17 | The National University Hospital of Iceland | deCODE genetics | Daniel F Gudbjartsson et al |
| 1265 | Iceland/257/2020 | 2020-03-17 | The National University Hospital of Iceland | deCODE genetics | Daniel F Gudbjartsson et al |
| 1266 | Iceland/256/2020 | 2020-03-17 | The National University Hospital of Iceland | deCODE genetics | Daniel F Gudbjartsson et al |
| 1267 | Iceland/258/2020 | 2020-03-17 | The National University Hospital of Iceland | deCODE genetics | Daniel F Gudbjartsson et al |
| 1268 | USA/NY-SURV0996/2020 | 2020-03-17 | NYC Department of Health and Mental Hygiene | Pathogen Discovery, Respiratory Viruses Branch, Division of Viral Diseases, Centers for Disease Control and Prevention | Ying Tao et al |
| 1269 | England/20124003702/2020 | 2020-03-17 | Respiratory Virus Unit, Microbiology Services Colindale, Public Health England | Respiratory Virus Unit, Microbiology Services Colindale, Public Health England | Monica Galiano et al |
| 1270 | Scotland/CVR171/2020 | 2020-03-17 | West of Scotland Specialist Virology Centre, NHSGGC / MRC-University of Glasgow Centre for Virus Research | COVID-19 Genomics UK (COG-UK) Consortium | Ana da Silva Filipe et al |
| 1271 | Thailand/Bangkok-0033/2020 | 2020-03-17 | Ramathibodi Hospital | COVID-19 Network Investigations (CONI) Alliance | Elizabeth Batty et al |
| 1272 | USA/WA-UW-1342/2020 | 2020-03-17 | UW Virology Lab | UW Virology Lab | Pavitra Roychoudhury et al |
| 1273 | Iceland/115/2020 | 2020-03-17 | deCODE genetics | deCODE genetics | Daniel F Gudbjartsson et al |
| 1274 | Iceland/279/2020 | 2020-03-17 | The National University Hospital of Iceland | deCODE genetics | Daniel F Gudbjartsson et al |
| 1275 | Iceland/265/2020 | 2020-03-17 | The National University Hospital of Iceland | deCODE genetics | Daniel F Gudbjartsson et al |
| 1276 | Iceland/236/2020 | 2020-03-17 | The National University Hospital of Iceland | deCODE genetics | Daniel F Gudbjartsson et al |
| 1277 | Iceland/234/2020 | 2020-03-17 | The National University Hospital of Iceland | deCODE genetics | Daniel F Gudbjartsson et al |
| 1278 | Iceland/276/2020 | 2020-03-17 | The National University Hospital of Iceland | deCODE genetics | Daniel F Gudbjartsson et al |
| 1279 | Iceland/228/2020 | 2020-03-17 | The National University Hospital of Iceland | deCODE genetics | Daniel F Gudbjartsson et al |
| 1280 | Iceland/241/2020 | 2020-03-17 | The National University Hospital of Iceland | deCODE genetics | Daniel F Gudbjartsson et al |
| 1281 | Iceland/259/2020 | 2020-03-17 | The National University Hospital of Iceland | deCODE genetics | Daniel F Gudbjartsson et al |

| | | | | | |
|---|---|---|---|---|---|
| 1282 | Switzerland/GE2164/2020 | 2020-03-17 | University Hospitals of Geneva Laboratory of Virology | University Hospitals of Geneva Laboratory of Virology | Laubscher F. et al |
| 1283 | Denmark/ALAB-HH57/2020 | 2020-03-17 | Department of Clinical Microbiology, Copenhagen University Hospital, Hvidovre, Kettegaard Alle 30, 2650 Hvidovre. | Albertsen lab, Department of Chemistry and Bioscience, Aalborg University, Denmark | Rasmus Kirkegaard et al |
| 1284 | Iceland/248/2020 | 2020-03-17 | The National University Hospital of Iceland | deCODE genetics | Daniel F Gudbjartsson et al |
| 1285 | Iceland/229/2020 | 2020-03-17 | The National University Hospital of Iceland | deCODE genetics | Daniel F Gudbjartsson et al |
| 1286 | Iceland/253/2020 | 2020-03-17 | The National University Hospital of Iceland | deCODE genetics | Daniel F Gudbjartsson et al |
| 1287 | Iceland/261/2020 | 2020-03-17 | The National University Hospital of Iceland | deCODE genetics | Daniel F Gudbjartsson et al |
| 1288 | Iceland/249/2020 | 2020-03-17 | The National University Hospital of Iceland | deCODE genetics | Daniel F Gudbjartsson et al |
| 1289 | Iceland/240/2020 | 2020-03-17 | The National University Hospital of Iceland | deCODE genetics | Daniel F Gudbjartsson et al |
| 1290 | Iceland/117/2020 | 2020-03-17 | deCODE genetics | deCODE genetics | Daniel F Gudbjartsson et al |
| 1291 | Iceland/260/2020 | 2020-03-17 | The National University Hospital of Iceland | deCODE genetics | Daniel F Gudbjartsson et al |
| 1292 | Taiwan/CGMH-CGU-17/2020 | 2020-03-17 | Laboratory Medicine | Department of Laboratory Medicine, Lin-Kou Chang Gung Memorial Hospital, Taoyuan, Taiwan | Kuo-Chien Tsao et al |
| 1293 | Taiwan/TSGH-02/2020 | 2020-03-17 | TSGH-CP molecular lab | TSGH-CP molecular lab | Cherng-Lih Perng et al |
| 1294 | Taiwan/TSGH-29/2020 | 2020-03-17 | TSGH-CP molecular lab | TSGH-CP molecular lab | Cherng-Lih Perng et al |
| 1295 | Taiwan/NTU13/2020 | 2020-03-17 | Department of Laboratory Medicine, National Taiwan University Hospital | Microbial Genomics Core Lab, National Taiwan University Centers of Genomic and Precision Medicine | Shiou-Hwei Yeh et al |
| 1296 | Taiwan/CGMH-CGU-14/2020 | 2020-03-17 | Laboratory Medicine | Department of Laboratory Medicine, Lin-Kou Chang Gung Memorial Hospital, Taoyuan, Taiwan | Kuo-Chien Tsao et al |
| 1297 | Taiwan/CGMH-CGU-13/2020 | 2020-03-17 | Laboratory Medicine | Department of Laboratory Medicine, Lin-Kou Chang Gung Memorial Hospital, Taoyuan, Taiwan | Kuo-Chien Tsao et al |
| 1298 | Taiwan/TSGH-03/2020 | 2020-03-17 | TSGH-CP molecular lab | TSGH-CP molecular lab | Cherng-Lih Perng et al |
| 1299 | Taiwan/TSGH-04/2020 | 2020-03-17 | TSGH-CP molecular lab | TSGH-CP molecular lab | Cherng-Lih Perng et al |
| 1300 | USA/AZ-TG269848/2020 | 2020-03-17 | AZ SPHL, Arizona Department of Health Services | TGen North | Jolene Bowers et al |
| 1301 | Netherlands/NA_95/2020 | 2020-03-17 | Dutch COVID-19 response team | Erasmus Medical Center | Bas Oude Munnink et al |
| 1302 | Luxembourg/LNS6689955/2020 | 2020-03-17 | Laboratoire National de Sante, Microbiology, Virology | Laboratoire National de Sante, Microbiology, Epidemiology and Microbial Genomics | Anke Wienecke-Baldacchino et al |
| 1303 | USA/WA-UW-1309/2020 | 2020-03-17 | UW Virology Lab | UW Virology Lab | Pavitra Roychoudhury et al |
| 1304 | Luxembourg/LNS2458673/2020 | 2020-03-17 | Laboratoire National de Sante, Microbiology, Virology | Laboratoire National de Sante, Microbiology, Epidemiology and Microbial Genomics | Anke Wienecke-Baldacchino et al |
| 1305 | USA/NY-PV08453/2020 | 2020-03-17 | MSHS Clinical Microbiology Laboratories | MSHS Pathogen Surveillance Program | Ana S. Gonzalez-Reiche et al |
| 1306 | France/ARA10282/2020 | 2020-03-17 | Centre Hospitalier Lucien Hussel | CNR Virus des Infections Respiratoires - France SUD | Antonin Bal et al |
| 1307 | Iceland/320/2020 | 2020-03-18 | The National University Hospital of Iceland | deCODE genetics | Daniel F Gudbjartsson et al |
| 1308 | Australia/VIC112/2020 | 2020-03-18 | Victorian Infectious Diseases Reference Laboratory (VIDRL) | Victorian Infectious Diseases Reference Laboratory and Microbiological Diagnostic Unit Public Health Laboratory, Doherty Institute | Caly L. et al |
| 1309 | Australia/NSW158/2020 | 2020-03-18 | Centre for Infectious Diseases and Microbiology Public | NSW Health Pathology - Institute of Clinical Pathology | Arnott A et al |

| | | | | | |
|---|---|---|---|---|---|
| | | | Health | and Medical Research; Westmead Hospital; University of Sydney | |
| 1310 | Australia/VIC165/2020 | 2020-03-18 | Victorian Infectious Diseases Reference Laboratory (VIDRL) | Victorian Infectious Diseases Reference Laboratory and Microbiological Diagnostic Unit Public Health Laboratory, Doherty Institute | Caly L. et al |
| 1311 | Australia/VIC166/2020 | 2020-03-18 | Victorian Infectious Diseases Reference Laboratory (VIDRL) | Victorian Infectious Diseases Reference Laboratory and Microbiological Diagnostic Unit Public Health Laboratory, Doherty Institute | Caly L. et al |
| 1312 | Australia/VIC173/2020 | 2020-03-18 | Victorian Infectious Diseases Reference Laboratory (VIDRL) | Victorian Infectious Diseases Reference Laboratory and Microbiological Diagnostic Unit Public Health Laboratory, Doherty Institute | Caly L. et al |
| 1313 | Australia/NSW81/2020 | 2020-03-18 | Centre for Infectious Diseases and Microbiology Public Health | NSW Health Pathology - Institute of Clinical Pathology and Medical Research; Westmead Hospital; University of Sydney | Sim E et al |
| 1314 | Australia/NSW100/2020 | 2020-03-18 | Centre for Infectious Diseases and Microbiology Public Health | NSW Health Pathology - Institute of Clinical Pathology and Medical Research; Westmead Hospital; University of Sydney | Draper J et al |
| 1315 | Australia/VIC175/2020 | 2020-03-18 | Victorian Infectious Diseases Reference Laboratory (VIDRL) | Victorian Infectious Diseases Reference Laboratory and Microbiological Diagnostic Unit Public Health Laboratory, Doherty Institute | Caly L. et al |
| 1316 | Australia/VIC179/2020 | 2020-03-18 | Victorian Infectious Diseases Reference Laboratory (VIDRL) | Victorian Infectious Diseases Reference Laboratory and Microbiological Diagnostic Unit Public Health Laboratory, Doherty Institute | Caly L. et al |
| 1317 | Australia/VIC159/2020 | 2020-03-18 | Victorian Infectious Diseases Reference Laboratory (VIDRL) | Victorian Infectious Diseases Reference Laboratory and Microbiological Diagnostic Unit Public Health Laboratory, Doherty Institute | Caly L. et al |
| 1318 | Australia/QLDID924/2020 | 2020-03-18 | Pathology Queensland | Public Health Virology Laboratory | Bixing Huang et al |
| 1319 | Australia/NSW175/2020 | 2020-03-18 | Centre for Infectious Diseases and Microbiology Public Health | NSW Health Pathology - Institute of Clinical Pathology and Medical Research; Westmead Hospital; University of Sydney | Gray K et al |
| 1320 | Italy/TE5166/2020 | 2020-03-18 | Ospedale Civile Giuseppe Mazzini | "Istituto Zooprofilattico Sperimentale dell'Abruzzo e Molise ""G. Caporale""" | Lorusso A et al |
| 1321 | Spain/Valencia146/2020 | 2020-03-18 | Servicio de Microbiologia. Hospital Universitario Doctor Peset | Sequencing and Bioinformatics Service and Molecular Epidemiology Research Group. FISABIO-Public Health | Juan Alberola Enguídanos et al |
| 1322 | Australia/VIC168/2020 | 2020-03-18 | Victorian Infectious Diseases Reference Laboratory (VIDRL) | Victorian Infectious Diseases Reference Laboratory and Microbiological Diagnostic Unit Public Health Laboratory, Doherty Institute | Caly L. et al |
| 1323 | Australia/VIC108/2020 | 2020-03-18 | Microbiological Diagnostic Unit Public Health Laboratory | Microbiological Diagnostic Unit Public Health Laboratory | Seemann T. et al |
| 1324 | Australia/VIC178/2020 | 2020-03-18 | Victorian Infectious Diseases Reference Laboratory (VIDRL) | Victorian Infectious Diseases Reference Laboratory and Microbiological Diagnostic Unit Public Health Laboratory, Doherty Institute | Caly L. et al |
| 1325 | Australia/VIC162/2020 | 2020-03-18 | Victorian Infectious Diseases Reference Laboratory (VIDRL) | Victorian Infectious Diseases Reference Laboratory and Microbiological Diagnostic Unit | Caly L. et al |

| | | | | | |
|---|---|---|---|---|---|
| | | | | Public Health Laboratory, Doherty Institute | |
| 1326 | Fujian/IM3520017T/2020 | 2020-03-18 | Fujian Center for Disease Control and Prevention | Fujian Center for Disease Control and Prevention | Lin Qi et al |
| 1327 | Argentina/C1374/2020 | 2020-03-18 | Servicio Virosis Respiratorias-Departamento Virología-INEI | Instituto Nacional Enfermedades Infecciosas C.G.Malbran | Baumeister E. et al |
| 1328 | Netherlands/ZuidHolland_41/2020 | 2020-03-18 | Dutch COVID-19 response team | Erasmus Medical Center | Bas Oude Munnink et al |
| 1329 | Belgium/DA-0319106/2020 | 2020-03-18 | KU Leuven, Clinical and Epidemiological Virology | KU Leuven, Clinical and Epidemiological Virology | Joan Marti-Carreras et al |
| 1330 | USA/NJ-NYUMC84/2020 | 2020-03-18 | NYU Langone Health | Departments of Pathology and Medicine, New York University School of Medicine | Maria Aguero-Rosenfeld et al |
| 1331 | USA/NY-NYUMC113/2020 | 2020-03-18 | NYU Langone Health | Departments of Pathology and Medicine, New York University School of Medicine | Maria Aguero-Rosenfeld et al |
| 1332 | USA/NY-NYUMC120/2020 | 2020-03-18 | NYU Langone Health | Departments of Pathology and Medicine, New York University School of Medicine | Maria Aguero-Rosenfeld et al |
| 1333 | USA/NY-PV09097/2020 | 2020-03-18 | MSHS Clinical Microbiology Laboratories | MSHS Pathogen Surveillance Program | Ana S. Gonzalez-Reiche et al |
| 1334 | USA/NY-NYUMC72/2020 | 2020-03-18 | NYU Langone Health | Departments of Pathology and Medicine, New York University School of Medicine | Maria Aguero-Rosenfeld et al |
| 1335 | Australia/NSW141/2020 | 2020-03-18 | Centre for Infectious Diseases and Microbiology Public Health | NSW Health Pathology - Institute of Clinical Pathology and Medical Research; Westmead Hospital; University of Sydney | Sim E et al |
| 1336 | Australia/VIC176/2020 | 2020-03-18 | Victorian Infectious Diseases Reference Laboratory (VIDRL) | Victorian Infectious Diseases Reference Laboratory and Microbiological Diagnostic Unit Public Health Laboratory, Doherty Institute | Caly L. et al |
| 1337 | Australia/VIC109/2020 | 2020-03-18 | Microbiological Diagnostic Unit Public Health Laboratory | Microbiological Diagnostic Unit Public Health Laboratory | Seemann T. et al |
| 1338 | Australia/VIC333/2020 | 2020-03-18 | Victorian Infectious Diseases Reference Laboratory (VIDRL) | Microbiological Diagnostic Unit Public Health Laboratory and Victorian Infectious Diseases Reference Laboratory, Doherty Institute | Caly L. et al |
| 1339 | Iceland/353/2020 | 2020-03-18 | deCODE genetics | deCODE genetics | Daniel F Gudbjartsson et al |
| 1340 | USA/NY-NYUMC46/2020 | 2020-03-18 | NYU Langone Health | Departments of Pathology and Medicine, New York University School of Medicine | Maria Aguero-Rosenfeld et al |
| 1341 | NewZealand/20VR0765/2020-03-18 | 2020-03-18 | Southern Community Labs Dunedin | Institute of Environmental Science and Research (ESR) Ltd | **Matt Storey et al** |
| 1342 | NewZealand/20VR0759/2020-03-18 | 2020-03-18 | Wellington SCL (WN) | Institute of Environmental Science and Research (ESR) Ltd | **Matt Storey et al** |
| 1343 | NewZealand/20VR0761/2020-03-18 | 2020-03-18 | Wellington SCL (WN) | Institute of Environmental Science and Research (ESR) Ltd | **Matt Storey et al** |
| 1344 | NewZealand/20VR1285/2020-03-18 | 2020-03-18 | LabPLUS | Institute of Environmental Science and Research (ESR) Ltd | **Matt Storey et al** |
| 1345 | Greece/16/2020 | 2020-03-18 | Laboratory of Microbiology, Department of Medicine, National and Kapodistrian University of Athens, Greece | Laboratory of Biology, Department of Medicine, Democritus University of Thrace, Greece | Maria Bampali et al |
| 1346 | Scotland/EDB057/2020 | 2020-03-18 | Virology Department, Royal Infirmary of Edinburgh, NHS Lothian / School of Biological Sciences, University of Edinburgh / Institute of Genetics and Molecular Medicine, University of Edinburgh | COVID-19 Genomics UK (COG-UK) Consortium | McHugh M et al |
| 1347 | Portugal/PT0046/2020 | 2020-03-18 | H Dr. Nelio Mendonca - Funchal | Instituto Nacional de Saude (INSA) | Guiomar et al et al |
| 1348 | Spain/Valencia139/2020 | 2020-03-18 | Servicio de Microbiología. | Sequencing and Bioinformatics | Maria Dolores Ocete |

| | | | | | |
|---|---|---|---|---|---|
| | | | Consorcio Hospital General Universitario de Valencia | Service and Molecular Epidemiology Research Group. FISABIO-Public Health | et al |
| 1349 | Turkey/HSGM-5516/2020 | 2020-03-18 | Ministry of Health Turkey | Ministry of Health Turkey | Fatma Bayrakdar et al |
| 1350 | Iceland/379/2020 | 2020-03-18 | The National University Hospital of Iceland | deCODE genetics | Daniel F Gudbjartsson et al |
| 1351 | USA/AZ-TG268360/2020 | 2020-03-18 | TGen North | TGen North | Jolene Bowers et al |
| 1352 | USA/WI-56/2020 | 2020-03-18 | University of Wisconsin-Madison AIDS Vaccine Research Laboratories | University of Wisconsin-Madison AIDS Vaccine Research Laboratories | Moreno et al |
| 1353 | England/NOTT-10E098/2020 | 2020-03-18 | Queens Medical Centre, Clinical Microbiology Department / DeepSeq Nottingham | COVID-19 Genomics UK (COG-UK) Consortium | Gemma Clark et al |
| 1354 | Turkey/HSGM-5624/2020 | 2020-03-18 | Ministry of Health Turkey | Ministry of Health Turkey | Fatma Bayrakdar et al |
| 1355 | Belgium/VD-0318102/2020 | 2020-03-18 | KU Leuven, Clinical and Epidemiological Virology | KU Leuven, Clinical and Epidemiological Virology | Joan Marti-Carreras et al |
| 1356 | USA/WA-UW-1470/2020 | 2020-03-18 | UW Virology Lab | UW Virology Lab | Pavitra Roychoudhury et al |
| 1357 | Iceland/290/2020 | 2020-03-18 | The National University Hospital of Iceland | deCODE genetics | Daniel F Gudbjartsson et al |
| 1358 | Iceland/321/2020 | 2020-03-18 | The National University Hospital of Iceland | deCODE genetics | Daniel F Gudbjartsson et al |
| 1359 | Iceland/301/2020 | 2020-03-18 | The National University Hospital of Iceland | deCODE genetics | Daniel F Gudbjartsson et al |
| 1360 | Iceland/316/2020 | 2020-03-18 | The National University Hospital of Iceland | deCODE genetics | Daniel F Gudbjartsson et al |
| 1361 | Iceland/312/2020 | 2020-03-18 | The National University Hospital of Iceland | deCODE genetics | Daniel F Gudbjartsson et al |
| 1362 | Iceland/292/2020 | 2020-03-18 | The National University Hospital of Iceland | deCODE genetics | Daniel F Gudbjartsson et al |
| 1363 | Iceland/306/2020 | 2020-03-18 | The National University Hospital of Iceland | deCODE genetics | Daniel F Gudbjartsson et al |
| 1364 | Iceland/322/2020 | 2020-03-18 | The National University Hospital of Iceland | deCODE genetics | Daniel F Gudbjartsson et al |
| 1365 | Iceland/313/2020 | 2020-03-18 | The National University Hospital of Iceland | deCODE genetics | Daniel F Gudbjartsson et al |
| 1366 | Iceland/308/2020 | 2020-03-18 | The National University Hospital of Iceland | deCODE genetics | Daniel F Gudbjartsson et al |
| 1367 | Iceland/293/2020 | 2020-03-18 | The National University Hospital of Iceland | deCODE genetics | Daniel F Gudbjartsson et al |
| 1368 | Iceland/287/2020 | 2020-03-18 | The National University Hospital of Iceland | deCODE genetics | Daniel F Gudbjartsson et al |
| 1369 | Iceland/325/2020 | 2020-03-18 | The National University Hospital of Iceland | deCODE genetics | Daniel F Gudbjartsson et al |
| 1370 | Iceland/300/2020 | 2020-03-18 | The National University Hospital of Iceland | deCODE genetics | Daniel F Gudbjartsson et al |
| 1371 | Iceland/324/2020 | 2020-03-18 | The National University Hospital of Iceland | deCODE genetics | Daniel F Gudbjartsson et al |
| 1372 | Iceland/286/2020 | 2020-03-18 | The National University Hospital of Iceland | deCODE genetics | Daniel F Gudbjartsson et al |
| 1373 | Iceland/314/2020 | 2020-03-18 | The National University Hospital of Iceland | deCODE genetics | Daniel F Gudbjartsson et al |
| 1374 | Iceland/296/2020 | 2020-03-18 | The National University Hospital of Iceland | deCODE genetics | Daniel F Gudbjartsson et al |
| 1375 | Iceland/305/2020 | 2020-03-18 | The National University Hospital of Iceland | deCODE genetics | Daniel F Gudbjartsson et al |
| 1376 | Iceland/315/2020 | 2020-03-18 | The National University Hospital of Iceland | deCODE genetics | Daniel F Gudbjartsson et al |
| 1377 | Iceland/318/2020 | 2020-03-18 | The National University Hospital of Iceland | deCODE genetics | Daniel F Gudbjartsson et al |
| 1378 | Iceland/262/2020 | 2020-03-18 | deCODE genetics | deCODE genetics | Daniel F Gudbjartsson et al |
| 1379 | Iceland/323/2020 | 2020-03-18 | The National University Hospital of Iceland | deCODE genetics | Daniel F Gudbjartsson et al |
| 1380 | Iceland/298/2020 | 2020-03-18 | The National University Hospital of Iceland | deCODE genetics | Daniel F Gudbjartsson et al |

| | | | | | |
|---|---|---|---|---|---|
| 1381 | Iceland/310/2020 | 2020-03-18 | The National University Hospital of Iceland | deCODE genetics | Daniel F Gudbjartsson et al |
| 1382 | Iceland/303/2020 | 2020-03-18 | The National University Hospital of Iceland | deCODE genetics | Daniel F Gudbjartsson et al |
| 1383 | Iceland/297/2020 | 2020-03-18 | The National University Hospital of Iceland | deCODE genetics | Daniel F Gudbjartsson et al |
| 1384 | USA/NY-NYUMC78/2020 | 2020-03-18 | NYU Langone Health | Departments of Pathology and Medicine, New York University School of Medicine | Maria Aguero-Rosenfeld et al |
| 1385 | USA/NY-NYUMC41/2020 | 2020-03-18 | NYU Langone Health | Departments of Pathology and Medicine, New York University School of Medicine | Maria Aguero-Rosenfeld et al |
| 1386 | USA/NY-NYUMC73/2020 | 2020-03-18 | NYU Langone Health | Departments of Pathology and Medicine, New York University School of Medicine | Maria Aguero-Rosenfeld et al |
| 1387 | England/20134002104/2020 | 2020-03-18 | Respiratory Virus Unit, Microbiology Services Colindale, Public Health England | Respiratory Virus Unit, Microbiology Services Colindale, Public Health England | Monica Galiano et al |
| 1388 | USA/NY-NYUMC89/2020 | 2020-03-18 | NYU Langone Health | Departments of Pathology and Medicine, New York University School of Medicine | Maria Aguero-Rosenfeld et al |
| 1389 | Iceland/294/2020 | 2020-03-18 | The National University Hospital of Iceland | deCODE genetics | Daniel F Gudbjartsson et al |
| 1390 | USA/NY-NYUMC81/2020 | 2020-03-18 | NYU Langone Health | Departments of Pathology and Medicine, New York University School of Medicine | Maria Aguero-Rosenfeld et al |
| 1391 | Taiwan/TSGH-12/2020 | 2020-03-18 | TSGH-CP molecular lab | TSGH-CP molecular lab | Cherng-Lih Perng et al |
| 1392 | Taiwan/CGMH-CGU-19/2020 | 2020-03-18 | Laboratory Medicine | Department of Laboratory Medicine, Lin-Kou Chang Gung Memorial Hospital, Taoyuan, Taiwan | Kuo-Chien Tsao et al |
| 1393 | Taiwan/CGMH-CGU-20/2020 | 2020-03-18 | Laboratory Medicine | Department of Laboratory Medicine, Lin-Kou Chang Gung Memorial Hospital, Taoyuan, Taiwan | Kuo-Chien Tsao et al |
| 1394 | Taiwan/NTU14/2020 | 2020-03-18 | Department of Laboratory Medicine, National Taiwan University Hospital | Microbial Genomics Core Lab, National Taiwan University Centers of Genomic and Precision Medicine | Shiou-Hwei Yeh et al |
| 1395 | Taiwan/CGMH-CGU-22/2020 | 2020-03-18 | Laboratory Medicine | Department of Laboratory Medicine, Lin-Kou Chang Gung Memorial Hospital, Taoyuan, Taiwan | Kuo-Chien Tsao et al |
| 1396 | Taiwan/NTU15/2020 | 2020-03-18 | Department of Laboratory Medicine, National Taiwan University Hospital | Microbial Genomics Core Lab, National Taiwan University Centers of Genomic and Precision Medicine | Shiou-Hwei Yeh et al |
| 1397 | Taiwan/CGMH-CGU-18/2020 | 2020-03-18 | Laboratory Medicine | Department of Laboratory Medicine, Lin-Kou Chang Gung Memorial Hospital, Taoyuan, Taiwan | Kuo-Chien Tsao et al |
| 1398 | Taiwan/128/2020 | 2020-03-18 | Centers for Disease Control, R.O.C. (Taiwan) | Centers for Disease Control, R.O.C. (Taiwan) | Ji-Rong Yang et al |
| 1399 | USA/NY-NYUMC60/2020 | 2020-03-18 | NYU Langone Health | Departments of Pathology and Medicine, New York University School of Medicine | Maria Aguero-Rosenfeld et al |
| 1400 | Australia/VIC160/2020 | 2020-03-18 | Victorian Infectious Diseases Reference Laboratory (VIDRL) | Victorian Infectious Diseases Reference Laboratory and Microbiological Diagnostic Unit Public Health Laboratory, Doherty Institute | Caly L. et al |
| 1401 | Spain/Valencia285/2020 | 2020-03-18 | Servicio de Microbiologia. Hospital Universitario Doctor Peset | Sequencing and Bioinformatics Service and Molecular Epidemiology Research Group. FISABIO-Public Health | Juan Alberola Enguídanos et al |
| 1402 | Australia/QLDID925/2020 | 2020-03-18 | Pathology Queensland | Public Health Virology Laboratory | Bixing Huang et al |
| 1403 | Switzerland/GE3144/2020 | 2020-03-18 | University Hospitals of Geneva Laboratory of Virology | University Hospitals of Geneva Laboratory of Virology | Laubscher F. et al |

| | | | | | |
|---|---|---|---|---|---|
| 1404 | Germany/NRW-41/2020 | 2020-03-18 | Center of Medical Microbiology, Virology, and Hospital Hygiene, University of Duesseldorf | Center of Medical Microbiology, Virology, and Hospital Hygiene, University of Duesseldorf | Ortwin Adams et al |
| 1405 | Fujian/IM3520013T/2020 | 2020-03-19 | Fujian Center for Disease Control and Prevention | Fujian Center for Disease Control and Prevention | Lin Qi et al |
| 1406 | USA/TX-HMH098/2020 | 2020-03-19 | Houston Methodist Hospital | Houston Methodist Hospital | S. Wesley Long et al |
| 1407 | Iceland/367/2020 | 2020-03-19 | The National University Hospital of Iceland | deCODE genetics | Daniel F Gudbjartsson et al |
| 1408 | Taiwan/144/2020 | 2020-03-19 | Centers for Disease Control, R.O.C. (Taiwan) | Centers for Disease Control, R.O.C. (Taiwan) | Ji-Rong Yang et al |
| 1409 | USA/NY-NYUMC47/2020 | 2020-03-19 | NYU Langone Health | Departments of Pathology and Medicine, New York University School of Medicine | Maria Aguero-Rosenfeld et al |
| 1410 | Australia/VIC198/2020 | 2020-03-19 | Victorian Infectious Diseases Reference Laboratory (VIDRL) | Victorian Infectious Diseases Reference Laboratory and Microbiological Diagnostic Unit Public Health Laboratory, Doherty Institute | Caly L. et al |
| 1411 | Australia/VIC184/2020 | 2020-03-19 | Victorian Infectious Diseases Reference Laboratory (VIDRL) | Victorian Infectious Diseases Reference Laboratory and Microbiological Diagnostic Unit Public Health Laboratory, Doherty Institute | Caly L. et al |
| 1412 | Australia/VIC195/2020 | 2020-03-19 | Victorian Infectious Diseases Reference Laboratory (VIDRL) | Victorian Infectious Diseases Reference Laboratory and Microbiological Diagnostic Unit Public Health Laboratory, Doherty Institute | Caly L. et al |
| 1413 | Australia/VIC207/2020 | 2020-03-19 | Victorian Infectious Diseases Reference Laboratory (VIDRL) | Victorian Infectious Diseases Reference Laboratory and Microbiological Diagnostic Unit Public Health Laboratory, Doherty Institute | Caly L. et al |
| 1414 | Australia/VIC201/2020 | 2020-03-19 | Victorian Infectious Diseases Reference Laboratory (VIDRL) | Victorian Infectious Diseases Reference Laboratory and Microbiological Diagnostic Unit Public Health Laboratory, Doherty Institute | Caly L. et al |
| 1415 | Australia/NSW114/2020 | 2020-03-19 | Centre for Infectious Diseases and Microbiology Public Health | NSW Health Pathology - Institute of Clinical Pathology and Medical Research; Westmead Hospital; University of Sydney | Lam C et al |
| 1416 | Australia/VIC199/2020 | 2020-03-19 | Victorian Infectious Diseases Reference Laboratory (VIDRL) | Victorian Infectious Diseases Reference Laboratory and Microbiological Diagnostic Unit Public Health Laboratory, Doherty Institute | Caly L. et al |
| 1417 | Australia/VIC186/2020 | 2020-03-19 | Victorian Infectious Diseases Reference Laboratory (VIDRL) | Victorian Infectious Diseases Reference Laboratory and Microbiological Diagnostic Unit Public Health Laboratory, Doherty Institute | Caly L. et al |
| 1418 | Switzerland/GE5737/2020 | 2020-03-19 | University Hospitals of Geneva Laboratory of Virology | University Hospitals of Geneva Laboratory of Virology | Laubscher F. et al |
| 1419 | Australia/VIC203/2020 | 2020-03-19 | Victorian Infectious Diseases Reference Laboratory (VIDRL) | Victorian Infectious Diseases Reference Laboratory and Microbiological Diagnostic Unit Public Health Laboratory, Doherty Institute | Caly L. et al |
| 1420 | Australia/VIC181/2020 | 2020-03-19 | Victorian Infectious Diseases Reference Laboratory (VIDRL) | Victorian Infectious Diseases Reference Laboratory and Microbiological Diagnostic Unit Public Health Laboratory, Doherty Institute | Caly L. et al |
| 1421 | Scotland/EDB067/2020 | 2020-03-19 | Virology Department, Royal Infirmary of Edinburgh, NHS Lothian / School of Biological Sciences, University of Edinburgh / Institute of | COVID-19 Genomics UK (COG-UK) Consortium | McHugh M et al |

| | | | | | |
|---|---|---|---|---|---|
| | | | Genetics and Molecular Medicine, University of Edinburgh | | |
| 1422 | England/CAMB-74D5B/2020 | 2020-03-19 | Department of Pathology, University of Cambridge | COVID-19 Genomics UK (COG-UK) Consortium | Luke W Meredith et al |
| 1423 | Iceland/368/2020 | 2020-03-19 | The National University Hospital of Iceland | deCODE genetics | Daniel F Gudbjartsson et al |
| 1424 | Australia/QLDID928/2020 | 2020-03-19 | Pathology Queensland | Public Health Virology Laboratory | Bixing Huang et al |
| 1425 | Australia/VIC185/2020 | 2020-03-19 | Victorian Infectious Diseases Reference Laboratory (VIDRL) | Victorian Infectious Diseases Reference Laboratory and Microbiological Diagnostic Unit Public Health Laboratory, Doherty Institute | Caly L. et al |
| 1426 | Australia/VIC204/2020 | 2020-03-19 | Victorian Infectious Diseases Reference Laboratory (VIDRL) | Victorian Infectious Diseases Reference Laboratory and Microbiological Diagnostic Unit Public Health Laboratory, Doherty Institute | Caly L. et al |
| 1427 | Australia/VIC117/2020 | 2020-03-19 | Microbiological Diagnostic Unit Public Health Laboratory | Microbiological Diagnostic Unit Public Health Laboratory | Seemann T. et al |
| 1428 | Australia/VIC120/2020 | 2020-03-19 | Microbiological Diagnostic Unit Public Health Laboratory | Microbiological Diagnostic Unit Public Health Laboratory | Seemann T. et al |
| 1429 | Australia/VIC190/2020 | 2020-03-19 | Victorian Infectious Diseases Reference Laboratory (VIDRL) | Victorian Infectious Diseases Reference Laboratory and Microbiological Diagnostic Unit Public Health Laboratory, Doherty Institute | Caly L. et al |
| 1430 | Australia/VIC115/2020 | 2020-03-19 | Microbiological Diagnostic Unit Public Health Laboratory | Microbiological Diagnostic Unit Public Health Laboratory | Seemann T. et al |
| 1431 | Australia/VIC197/2020 | 2020-03-19 | Victorian Infectious Diseases Reference Laboratory (VIDRL) | Victorian Infectious Diseases Reference Laboratory and Microbiological Diagnostic Unit Public Health Laboratory, Doherty Institute | Caly L. et al |
| 1432 | Australia/NSW74/2020 | 2020-03-19 | Centre for Infectious Diseases and Microbiology Public Health | NSW Health Pathology - Institute of Clinical Pathology and Medical Research; Westmead Hospital; University of Sydney | Lam C et al |
| 1433 | Netherlands/Limburg_8/2020 | 2020-03-19 | Dutch COVID-19 response team | Erasmus Medical Center | Bas Oude Munnink et al |
| 1434 | Spain/Valencia163/2020 | 2020-03-19 | Servicio de Microbiologia. Hospital Universitario Doctor Peset | Sequencing and Bioinformatics Service and Molecular Epidemiology Research Group. FISABIO-Public Health | Juan Alberola Enguídanos et al |
| 1435 | Italy/TE5476/2020 | 2020-03-19 | Ospedale Civile Giuseppe Mazzini | Istituto Zooprofilattico Sperimentale dell'Abruzzo e Molise "G. Caporale" | Lorusso A et al |
| 1436 | USA/WA-UW-1552/2020 | 2020-03-19 | UW Virology Lab | UW Virology Lab | Pavitra Roychoudhury et al |
| 1437 | Spain/Valencia75/2020 | 2020-03-19 | Servicio de Microbiologia. Hospital Clinico Universitario de Valencia | Sequencing and Bioinformatics Service and Molecular Epidemiology Research Group. FISABIO-Public Health | Inma Galán Vendrell et al |
| 1438 | Spain/Valencia156/2020 | 2020-03-19 | Servicio de Microbiologia. Hospital Universitario Doctor Peset | Sequencing and Bioinformatics Service and Molecular Epidemiology Research Group. FISABIO-Public Health | Juan Alberola Enguídanos et al |
| 1439 | Australia/NSW151/2020 | 2020-03-19 | ACT Pathology, The Canberra Hospital | NSW Health Pathology - Institute of Clinical Pathology and Medical Research; Westmead Hospital; University of Sydney | Sim E et al |
| 1440 | Australia/NSW84/2020 | 2020-03-19 | Centre for Infectious Diseases and Microbiology Public Health | NSW Health Pathology - Institute of Clinical Pathology and Medical Research; Westmead Hospital; University of Sydney | Lam C et al |
| 1441 | USA/TX-HMH096/2020 | 2020-03-19 | Houston Methodist Hospital | Houston Methodist Hospital | S. Wesley Long et al |
| 1442 | USA/AZ-TG268770/2020 | 2020-03-19 | TGen North | TGen North | Jolene Bowers et al |

| | | | | | |
|---|---|---|---|---|---|
| 1443 | NewZealand/20VR0933/2020-03-19 | 2020-03-19 | PathLab Bay of Plenty | Institute of Environmental Science and Research (ESR) Ltd | **Matt Storey et al** |
| 1444 | NewZealand/20VR1110/2020-03-19 | 2020-03-19 | Waikato Hospital | Institute of Environmental Science and Research (ESR) Ltd | **Matt Storey et al** |
| 1445 | NewZealand/20VR1004/2020-03-19 | 2020-03-19 | Southern Community Labs Dunedin | Institute of Environmental Science and Research (ESR) Ltd | **Matt Storey et al** |
| 1446 | NewZealand/20VR1286/2020-03-19 | 2020-03-19 | LabPLUS | Institute of Environmental Science and Research (ESR) Ltd | **Matt Storey et al** |
| 1447 | USA/WA_0429/2020 | 2020-03-19 | WA State Department of Health | Pathogen Discovery, Respiratory Viruses Branch, Division of Viral Diseases, Centers for Disease Control and Prevention | Ying Tao et al |
| 1448 | Portugal/PT0055/2020 | 2020-03-19 | H Beatriz Angelo | Instituto Nacional de Saude (INSA) | Guiomar et al et al |
| 1449 | Iceland/413/2020 | 2020-03-19 | The National University Hospital of Iceland | deCODE genetics | Daniel F Gudbjartsson et al |
| 1450 | Netherlands/NA_135/2020 | 2020-03-19 | Dutch COVID-19 response team | Erasmus Medical Center | Bas Oude Munnink et al |
| 1451 | Fujian/IM3520016T/2020 | 2020-03-19 | Fujian Center for Disease Control and Prevention | Fujian Center for Disease Control and Prevention | Lin Qi et al |
| 1452 | Italy/TE5472/2020 | 2020-03-19 | Ospedale Civile Castel Di Sangro | Istituto Zooprofilattico Sperimentale dell'Abruzzo e Molise "G. Caporale" | Lorusso A et al |
| 1453 | Germany/NRW-42-2/2020 | 2020-03-19 | Center of Medical Microbiology, Virology, and Hospital Hygiene, University of Duesseldorf | Center of Medical Microbiology, Virology, and Hospital Hygiene, University of Duesseldorf | Ortwin Adams et al |
| 1454 | Spain/Valencia155/2020 | 2020-03-19 | Servicio de Microbiologia. Hospital Universitario Doctor Peset | Sequencing and Bioinformatics Service and Molecular Epidemiology Research Group. FISABIO-Public Health | Juan Alberola Enguídanos et al |
| 1455 | Wales/PHWC-248B3/2020 | 2020-03-19 | Wales Specialist Virology Centre | Public Health Wales Microbiology Cardiff | Catherine Moore et al |
| 1456 | Iceland/410/2020 | 2020-03-19 | The National University Hospital of Iceland | deCODE genetics | Daniel F Gudbjartsson et al |
| 1457 | Iceland/400/2020 | 2020-03-19 | The National University Hospital of Iceland | deCODE genetics | Daniel F Gudbjartsson et al |
| 1458 | Iceland/357/2020 | 2020-03-19 | The National University Hospital of Iceland | deCODE genetics | Daniel F Gudbjartsson et al |
| 1459 | Iceland/358/2020 | 2020-03-19 | The National University Hospital of Iceland | deCODE genetics | Daniel F Gudbjartsson et al |
| 1460 | Iceland/414/2020 | 2020-03-19 | The National University Hospital of Iceland | deCODE genetics | Daniel F Gudbjartsson et al |
| 1461 | Iceland/360/2020 | 2020-03-19 | The National University Hospital of Iceland | deCODE genetics | Daniel F Gudbjartsson et al |
| 1462 | Iceland/386/2020 | 2020-03-19 | The National University Hospital of Iceland | deCODE genetics | Daniel F Gudbjartsson et al |
| 1463 | Iceland/422/2020 | 2020-03-19 | The National University Hospital of Iceland | deCODE genetics | Daniel F Gudbjartsson et al |
| 1464 | Iceland/356/2020 | 2020-03-19 | The National University Hospital of Iceland | deCODE genetics | Daniel F Gudbjartsson et al |
| 1465 | Iceland/388/2020 | 2020-03-19 | The National University Hospital of Iceland | deCODE genetics | Daniel F Gudbjartsson et al |
| 1466 | Iceland/366/2020 | 2020-03-19 | The National University Hospital of Iceland | deCODE genetics | Daniel F Gudbjartsson et al |
| 1467 | Iceland/452/2020 | 2020-03-19 | The National University Hospital of Iceland | deCODE genetics | Daniel F Gudbjartsson et al |
| 1468 | Iceland/382/2020 | 2020-03-19 | The National University Hospital of Iceland | deCODE genetics | Daniel F Gudbjartsson et al |
| 1469 | Iceland/398/2020 | 2020-03-19 | The National University Hospital of Iceland | deCODE genetics | Daniel F Gudbjartsson et al |
| 1470 | Iceland/391/2020 | 2020-03-19 | The National University Hospital of Iceland | deCODE genetics | Daniel F Gudbjartsson et al |
| 1471 | Iceland/401/2020 | 2020-03-19 | The National University Hospital of Iceland | deCODE genetics | Daniel F Gudbjartsson et al |
| 1472 | Iceland/374/2020 | 2020-03-19 | The National University Hospital of Iceland | deCODE genetics | Daniel F Gudbjartsson et al |
| 1473 | Iceland/390/2020 | 2020-03-19 | The National University | deCODE genetics | Daniel F Gudbjartsson |

| | | | | | |
|---|---|---|---|---|---|
| | | | Hospital of Iceland | | et al |
| 1474 | Iceland/415/2020 | 2020-03-19 | The National University Hospital of Iceland | deCODE genetics | Daniel F Gudbjartsson et al |
| 1475 | Italy/TE5512/2020 | 2020-03-19 | Ospedale Regionale San Salvatore | Istituto Zooprofilattico Sperimentale dell'Abruzzo e Molise "G. Caporale" | Lorusso A et al |
| 1476 | Spain/Valencia151/2020 | 2020-03-19 | Servicio de Microbiologia. Hospital Universitario Doctor Peset | Sequencing and Bioinformatics Service and Molecular Epidemiology Research Group. FISABIO-Public Health | Juan Alberola Enguídanos et al |
| 1477 | Iceland/377/2020 | 2020-03-19 | The National University Hospital of Iceland | deCODE genetics | Daniel F Gudbjartsson et al |
| 1478 | Iceland/421/2020 | 2020-03-19 | The National University Hospital of Iceland | deCODE genetics | Daniel F Gudbjartsson et al |
| 1479 | Iceland/395/2020 | 2020-03-19 | The National University Hospital of Iceland | deCODE genetics | Daniel F Gudbjartsson et al |
| 1480 | Iceland/399/2020 | 2020-03-19 | The National University Hospital of Iceland | deCODE genetics | Daniel F Gudbjartsson et al |
| 1481 | Iceland/387/2020 | 2020-03-19 | The National University Hospital of Iceland | deCODE genetics | Daniel F Gudbjartsson et al |
| 1482 | Iceland/416/2020 | 2020-03-19 | The National University Hospital of Iceland | deCODE genetics | Daniel F Gudbjartsson et al |
| 1483 | Iceland/361/2020 | 2020-03-19 | The National University Hospital of Iceland | deCODE genetics | Daniel F Gudbjartsson et al |
| 1484 | Iceland/412/2020 | 2020-03-19 | The National University Hospital of Iceland | deCODE genetics | Daniel F Gudbjartsson et al |
| 1485 | Iceland/359/2020 | 2020-03-19 | The National University Hospital of Iceland | deCODE genetics | Daniel F Gudbjartsson et al |
| 1486 | Iceland/405/2020 | 2020-03-19 | The National University Hospital of Iceland | deCODE genetics | Daniel F Gudbjartsson et al |
| 1487 | Iceland/369/2020 | 2020-03-19 | The National University Hospital of Iceland | deCODE genetics | Daniel F Gudbjartsson et al |
| 1488 | Iceland/419/2020 | 2020-03-19 | The National University Hospital of Iceland | deCODE genetics | Daniel F Gudbjartsson et al |
| 1489 | Iceland/380/2020 | 2020-03-19 | The National University Hospital of Iceland | deCODE genetics | Daniel F Gudbjartsson et al |
| 1490 | Iceland/376/2020 | 2020-03-19 | The National University Hospital of Iceland | deCODE genetics | Daniel F Gudbjartsson et al |
| 1491 | USA/WA-UW-1715/2020 | 2020-03-19 | UW Virology Lab | UW Virology Lab | Pavitra Roychoudhury et al |
| 1492 | USA/CA-SCCPHD-UC191/2020 | 2020-03-19 | Santa Clara County Public Health Department | Chiu Laboratory, University of California, San Francisco | Xianding Deng et al |
| 1493 | Australia/VIC189/2020 | 2020-03-19 | Victorian Infectious Diseases Reference Laboratory (VIDRL) | Victorian Infectious Diseases Reference Laboratory and Microbiological Diagnostic Unit Public Health Laboratory, Doherty Institute | Caly L. et al |
| 1494 | Germany/NRW-44/2020 | 2020-03-19 | Center of Medical Microbiology, Virology, and Hospital Hygiene, University of Duesseldorf | Center of Medical Microbiology, Virology, and Hospital Hygiene, University of Duesseldorf | Ortwin Adams et al |
| 1495 | Iceland/418/2020 | 2020-03-19 | The National University Hospital of Iceland | deCODE genetics | Daniel F Gudbjartsson et al |
| 1496 | Iceland/423/2020 | 2020-03-19 | The National University Hospital of Iceland | deCODE genetics | Daniel F Gudbjartsson et al |
| 1497 | Iceland/355/2020 | 2020-03-19 | The National University Hospital of Iceland | deCODE genetics | Daniel F Gudbjartsson et al |
| 1498 | Taiwan/NTU18/2020 | 2020-03-19 | Department of Laboratory Medicine, National Taiwan University Hospital | Microbial Genomics Core Lab, National Taiwan University Centers of Genomic and Precision Medicine | Shiou-Hwei Yeh et al |
| 1499 | Taiwan/NTU20/2020 | 2020-03-19 | Department of Laboratory Medicine, National Taiwan University Hospital | Microbial Genomics Core Lab, National Taiwan University Centers of Genomic and Precision Medicine | Shiou-Hwei Yeh et al |
| 1500 | Taiwan/NTU21/2020 | 2020-03-19 | Department of Laboratory Medicine, National Taiwan University Hospital | Microbial Genomics Core Lab, National Taiwan University Centers of Genomic and Precision Medicine | Shiou-Hwei Yeh et al |

| | | | | | |
|---|---|---|---|---|---|
| 1501 | Taiwan/TSGH-16/2020 | 2020-03-19 | TSGH-CP molecular lab | TSGH-CP molecular lab | Cherng-Lih Perng et al |
| 1502 | Taiwan/NTU17/2020 | 2020-03-19 | Department of Laboratory Medicine, National Taiwan University Hospital | Microbial Genomics Core Lab, National Taiwan University Centers of Genomic and Precision Medicine | Shiou-Hwei Yeh et al |
| 1503 | Taiwan/TSGH-15/2020 | 2020-03-19 | TSGH-CP molecular lab | TSGH-CP molecular lab | """No. 325 et al" |
| 1504 | Taiwan/NTU16/2020 | 2020-03-19 | Department of Laboratory Medicine, National Taiwan University Hospital | Microbial Genomics Core Lab, National Taiwan University Centers of Genomic and Precision Medicine | Shiou-Hwei Yeh et al |
| 1505 | Taiwan/TSGH-17/2020 | 2020-03-19 | TSGH-CP molecular lab | TSGH-CP molecular lab | Cherng-Lih Perng et al |
| 1506 | Taiwan/125/2020 | 2020-03-19 | Centers for Disease Control, R.O.C. (Taiwan) | Centers for Disease Control, R.O.C. (Taiwan) | Ji-Rong Yang et al |
| 1507 | Taiwan/NTU19/2020 | 2020-03-19 | Department of Laboratory Medicine, National Taiwan University Hospital | Microbial Genomics Core Lab, National Taiwan University Centers of Genomic and Precision Medicine | Shiou-Hwei Yeh et al |
| 1508 | USA/NY-NYUMC53/2020 | 2020-03-19 | NYU Langone Health | Departments of Pathology and Medicine, New York University School of Medicine | Maria Aguero-Rosenfeld et al |
| 1509 | Russia/Moscow-71605/2020 | 2020-03-19 | State Research Center of Virology and Biotechnology VECTOR, Department of Collection of Microorganisms | State Research Center of Virology and Biotechnology VECTOR, Department of Collection of Microorganisms | Sergey A. Bodnev et al |
| 1510 | Netherlands/NA_136/2020 | 2020-03-19 | Dutch COVID-19 response team | Erasmus Medical Center | Bas Oude Munnink et al |
| 1511 | USA/NY-PV09176/2020 | 2020-03-19 | MSHS Clinical Microbiology Laboratories | MSHS Pathogen Surveillance Program | Ana S. Gonzalez-Reiche et al |
| 1512 | Luxembourg/LNS9984061/2020 | 2020-03-19 | Laboratoire National de Sante, Microbiology, Virology | Laboratoire National de Sante, Microbiology, Epidemiology and Microbial Genomics | Anke Wienecke-Baldacchino et al |
| 1513 | Spain/Valencia167/2020 | 2020-03-19 | Servicio de Microbiologia. Hospital Universitario Doctor Peset | Sequencing and Bioinformatics Service and Molecular Epidemiology Research Group. FISABIO-Public Health | Juan Alberola Enguídanos et al |
| 1514 | USA/VA-DCLS-0036/2020 | 2020-03-20 | Virginia DCLS | Virginia DCLS | Virginia DCLS et al |
| 1515 | Scotland/CVR309/2020 | 2020-03-20 | West of Scotland Specialist Virology Centre, NHSGGC / MRC-University of Glasgow Centre for Virus Research | COVID-19 Genomics UK (COG-UK) Consortium | Ana da Silva Filipe et al |
| 1516 | Senegal/618/2020 | 2020-03-20 | Institut Pasteur Dakar | Institut Pasteur de Dakar | Ndongo Dia et al |
| 1517 | USA/WA-UW-1640/2020 | 2020-03-20 | UW Virology Lab | UW Virology Lab | Pavitra Roychoudhury et al |
| 1518 | Australia/VIC350/2020 | 2020-03-20 | Victorian Infectious Diseases Reference Laboratory (VIDRL) | Microbiological Diagnostic Unit Public Health Laboratory and Victorian Infectious Diseases Reference Laboratory, Doherty Institute | Caly L. et al |
| 1519 | Australia/VIC339/2020 | 2020-03-20 | Victorian Infectious Diseases Reference Laboratory (VIDRL) | Microbiological Diagnostic Unit Public Health Laboratory and Victorian Infectious Diseases Reference Laboratory, Doherty Institute | Caly L. et al |
| 1520 | Australia/NSW69/2020 | 2020-03-20 | Centre for Infectious Diseases and Microbiology Public Health | NSW Health Pathology - Institute of Clinical Pathology and Medical Research; Westmead Hospital; University of Sydney | Sadsad R et al |
| 1521 | Australia/VIC227/2020 | 2020-03-20 | Victorian Infectious Diseases Reference Laboratory (VIDRL) | Victorian Infectious Diseases Reference Laboratory and Microbiological Diagnostic Unit Public Health Laboratory, Doherty Institute | Caly L. et al |
| 1522 | Australia/VIC226/2020 | 2020-03-20 | Victorian Infectious Diseases Reference Laboratory (VIDRL) | Victorian Infectious Diseases Reference Laboratory and Microbiological Diagnostic Unit Public Health Laboratory, Doherty Institute | Caly L. et al |
| 1523 | Australia/VIC127/2020 | 2020-03-20 | Microbiological Diagnostic Unit Public Health Laboratory | Microbiological Diagnostic Unit Public Health Laboratory | Seemann T. et al |

| | | | | | |
|---|---|---|---|---|---|
| 1524 | Australia/NSW91/2020 | 2020-03-20 | Centre for Infectious Diseases and Microbiology Public Health | NSW Health Pathology - Institute of Clinical Pathology and Medical Research; Westmead Hospital; University of Sydney | Sim E et al |
| 1525 | Australia/VIC219/2020 | 2020-03-20 | Victorian Infectious Diseases Reference Laboratory (VIDRL) | Victorian Infectious Diseases Reference Laboratory and Microbiological Diagnostic Unit Public Health Laboratory, Doherty Institute | Caly L. et al |
| 1526 | Australia/VIC222/2020 | 2020-03-20 | Victorian Infectious Diseases Reference Laboratory (VIDRL) | Victorian Infectious Diseases Reference Laboratory and Microbiological Diagnostic Unit Public Health Laboratory, Doherty Institute | Caly L. et al |
| 1527 | USA/NY-PV09198/2020 | 2020-03-20 | MSHS Clinical Microbiology Laboratories | MSHS Pathogen Surveillance Program | Ana S. Gonzalez-Reiche et al |
| 1528 | Portugal/PT0056/2020 | 2020-03-20 | H Dr. Nelio Mendonca - Funchal | Instituto Nacional de Saude (INSA) | Guiomar et al et al |
| 1529 | USA/WA-UW-1682/2020 | 2020-03-20 | UW Virology Lab | UW Virology Lab | Pavitra Roychoudhury et al |
| 1530 | England/20129018904/2020 | 2020-03-20 | Respiratory Virus Unit, Microbiology Services Colindale, Public Health England | Respiratory Virus Unit, Microbiology Services Colindale, Public Health England | Monica Galiano et al |
| 1531 | Australia/VIC213/2020 | 2020-03-20 | Victorian Infectious Diseases Reference Laboratory (VIDRL) | Victorian Infectious Diseases Reference Laboratory and Microbiological Diagnostic Unit Public Health Laboratory, Doherty Institute | Caly L. et al |
| 1532 | Australia/VIC228/2020 | 2020-03-20 | Victorian Infectious Diseases Reference Laboratory (VIDRL) | Victorian Infectious Diseases Reference Laboratory and Microbiological Diagnostic Unit Public Health Laboratory, Doherty Institute | Caly L. et al |
| 1533 | Australia/VIC209/2020 | 2020-03-20 | Victorian Infectious Diseases Reference Laboratory (VIDRL) | Victorian Infectious Diseases Reference Laboratory and Microbiological Diagnostic Unit Public Health Laboratory, Doherty Institute | Caly L. et al |
| 1534 | Italy/TE5674/2020 | 2020-03-20 | Ospedale Civile Giuseppe Mazzini | "Istituto Zooprofilattico Sperimentale dell'Abruzzo e Molise ""G. Caporale""" | Lorusso A et al |
| 1535 | USA/NY-PV09073/2020 | 2020-03-20 | MSHS Clinical Microbiology Laboratories | MSHS Pathogen Surveillance Program | Ana S. Gonzalez-Reiche et al |
| 1536 | Australia/VIC338/2020 | 2020-03-20 | Victorian Infectious Diseases Reference Laboratory (VIDRL) | Microbiological Diagnostic Unit Public Health Laboratory and Victorian Infectious Diseases Reference Laboratory, Doherty Institute | Caly L. et al |
| 1537 | Australia/VIC347/2020 | 2020-03-20 | Victorian Infectious Diseases Reference Laboratory (VIDRL) | Microbiological Diagnostic Unit Public Health Laboratory and Victorian Infectious Diseases Reference Laboratory, Doherty Institute | Caly L. et al |
| 1538 | Australia/VIC343/2020 | 2020-03-20 | Victorian Infectious Diseases Reference Laboratory (VIDRL) | Microbiological Diagnostic Unit Public Health Laboratory and Victorian Infectious Diseases Reference Laboratory, Doherty Institute | Caly L. et al |
| 1539 | Australia/VIC208/2020 | 2020-03-20 | Victorian Infectious Diseases Reference Laboratory (VIDRL) | Victorian Infectious Diseases Reference Laboratory and Microbiological Diagnostic Unit Public Health Laboratory, Doherty Institute | Caly L. et al |
| 1540 | Iceland/459/2020 | 2020-03-20 | The National University Hospital of Iceland | deCODE genetics | Daniel F Gudbjartsson et al |
| 1541 | Hungary/SRC-00066/2020 | 2020-03-20 | Virological Research Group, Szentágothai Research Centre | Bioinformatics Research Group, Szentágothai Research Centre | Péter Urbán et al |
| 1542 | USA/WA-S186/2020 | 2020-03-20 | Washington State Department of Health | Seattle Flu Study | Chu et al et al |
| 1543 | NewZealand/20VR1018/2020-03-20 | 2020-03-20 | Wellington SCL (WN) | Institute of Environmental | **Matt Storey et al** |

| | | | | | |
|---|---|---|---|---|---|
| | | | | Science and Research (ESR) Ltd | |
| 1544 | NewZealand/20VR1133/2020-03-20 | 2020-03-20 | PathLab Bay of Plenty | Institute of Environmental Science and Research (ESR) Ltd | **Matt Storey et al** |
| 1545 | NewZealand/20VR1020/2020-03-20 | 2020-03-20 | Wellington SCL (WN) | Institute of Environmental Science and Research (ESR) Ltd | **Matt Storey et al** |
| 1546 | NewZealand/20VR1275/2020-03-20 | 2020-03-20 | LabPLUS | Institute of Environmental Science and Research (ESR) Ltd | **Matt Storey et al** |
| 1547 | NewZealand/20VR1283/2020-03-20 | 2020-03-20 | LabPLUS | Institute of Environmental Science and Research (ESR) Ltd | **Matt Storey et al** |
| 1548 | NewZealand/20VR1276/2020-03-20 | 2020-03-20 | LabPLUS | Institute of Environmental Science and Research (ESR) Ltd | **Matt Storey et al** |
| 1549 | NewZealand/20VR1016/2020-03-20 | 2020-03-20 | Wellington SCL (WN) | Institute of Environmental Science and Research (ESR) Ltd | **Matt Storey et al** |
| 1550 | NewZealand/20VR0901/2020-03-20 | 2020-03-20 | Wellington SCL (WR) | Institute of Environmental Science and Research (ESR) Ltd | **Matt Storey et al** |
| 1551 | NewZealand/20VR1022/2020-03-20 | 2020-03-20 | Wellington SCL (WN) | Institute of Environmental Science and Research (ESR) Ltd | **Matt Storey et al** |
| 1552 | NewZealand/20VR1278/2020-03-20 | 2020-03-20 | LabPLUS | Institute of Environmental Science and Research (ESR) Ltd | **Matt Storey et al** |
| 1553 | NewZealand/20VR1114/2020-03-20 | 2020-03-20 | Waikato Hospital | Institute of Environmental Science and Research (ESR) Ltd | **Matt Storey et al** |
| 1554 | NewZealand/20VR1019/2020-03-20 | 2020-03-20 | Wellington SCL (WN) | Institute of Environmental Science and Research (ESR) Ltd | **Matt Storey et al** |
| 1555 | NewZealand/20VR1277/2020-03-20 | 2020-03-20 | LabPLUS | Institute of Environmental Science and Research (ESR) Ltd | **Matt Storey et al** |
| 1556 | NewZealand/20VR1282/2020-03-20 | 2020-03-20 | LabPLUS | Institute of Environmental Science and Research (ESR) Ltd | **Matt Storey et al** |
| 1557 | NewZealand/20VR1280/2020-03-20 | 2020-03-20 | LabPLUS | Institute of Environmental Science and Research (ESR) Ltd | **Matt Storey et al** |
| 1558 | NewZealand/20VR0914/2020-03-20 | 2020-03-20 | Wellington SCL (WR) | Institute of Environmental Science and Research (ESR) Ltd | **Matt Storey et al** |
| 1559 | NewZealand/20VR1281/2020-03-20 | 2020-03-20 | LabPLUS | Institute of Environmental Science and Research (ESR) Ltd | **Matt Storey et al** |
| 1560 | Spain/Valencia81/2020 | 2020-03-20 | Servicio de Microbiologia. Hospital Clinico Universitario de Valencia | Sequencing and Bioinformatics Service and Molecular Epidemiology Research Group. FISABIO-Public Health | Griselda De Marco et al |
| 1561 | Spain/Valencia172/2020 | 2020-03-20 | Servicio de Microbiologia. Hospital Universitario Doctor Peset | Sequencing and Bioinformatics Service and Molecular Epidemiology Research Group. FISABIO-Public Health | Juan Alberola Enguídanos et al |
| 1562 | France/IDF3170/2020 | 2020-03-20 | Service de Biologie Médicale - BP 125 | National Reference Center for Viruses of Respiratory Infections, Institut Pasteur, Paris | Mélanie Albert et al |
| 1563 | USA/WA-UW-1683/2020 | 2020-03-20 | UW Virology Lab | UW Virology Lab | Pavitra Roychoudhury et al |
| 1564 | Iceland/457/2020 | 2020-03-20 | The National University Hospital of Iceland | deCODE genetics | Daniel F Gudbjartsson et al |
| 1565 | USA/MN-MDH-119/2020 | 2020-03-20 | Minnesota Department of Health, Public Health Laboratory | Minnesota Department of Health, Public Health Laboratory | Matt Plumb et al |
| 1566 | USA/NY-PV09079/2020 | 2020-03-20 | MSHS Clinical Microbiology Laboratories | MSHS Pathogen Surveillance Program | Ana S. Gonzalez-Reiche et al |
| 1567 | Japan/P4-7/2020 | 2020-03-20 | Saitama Prefectural Institute of Public Health | Pathogen Genomics Center, National Institute of Infectious Diseases | Tsuyoshi Sekizuka et al |
| 1568 | Iceland/464/2020 | 2020-03-20 | The National University Hospital of Iceland | deCODE genetics | Daniel F Gudbjartsson et al |
| 1569 | Iceland/431/2020 | 2020-03-20 | The National University Hospital of Iceland | deCODE genetics | Daniel F Gudbjartsson et al |
| 1570 | Iceland/480/2020 | 2020-03-20 | The National University Hospital of Iceland | deCODE genetics | Daniel F Gudbjartsson et al |
| 1571 | Iceland/439/2020 | 2020-03-20 | The National University Hospital of Iceland | deCODE genetics | Daniel F Gudbjartsson et al |
| 1572 | Iceland/507/2020 | 2020-03-20 | The National University Hospital of Iceland | deCODE genetics | Daniel F Gudbjartsson et al |
| 1573 | Iceland/438/2020 | 2020-03-20 | The National University Hospital of Iceland | deCODE genetics | Daniel F Gudbjartsson et al |

| | | | | | |
|---|---|---|---|---|---|
| 1574 | Iceland/493/2020 | 2020-03-20 | The National University Hospital of Iceland | deCODE genetics | Daniel F Gudbjartsson et al |
| 1575 | Iceland/435/2020 | 2020-03-20 | The National University Hospital of Iceland | deCODE genetics | Daniel F Gudbjartsson et al |
| 1576 | Iceland/473/2020 | 2020-03-20 | The National University Hospital of Iceland | deCODE genetics | Daniel F Gudbjartsson et al |
| 1577 | Iceland/472/2020 | 2020-03-20 | The National University Hospital of Iceland | deCODE genetics | Daniel F Gudbjartsson et al |
| 1578 | Iceland/426/2020 | 2020-03-20 | The National University Hospital of Iceland | deCODE genetics | Daniel F Gudbjartsson et al |
| 1579 | Iceland/479/2020 | 2020-03-20 | The National University Hospital of Iceland | deCODE genetics | Daniel F Gudbjartsson et al |
| 1580 | Iceland/433/2020 | 2020-03-20 | The National University Hospital of Iceland | deCODE genetics | Daniel F Gudbjartsson et al |
| 1581 | Iceland/446/2020 | 2020-03-20 | The National University Hospital of Iceland | deCODE genetics | Daniel F Gudbjartsson et al |
| 1582 | Iceland/451/2020 | 2020-03-20 | The National University Hospital of Iceland | deCODE genetics | Daniel F Gudbjartsson et al |
| 1583 | Iceland/484/2020 | 2020-03-20 | The National University Hospital of Iceland | deCODE genetics | Daniel F Gudbjartsson et al |
| 1584 | Iceland/477/2020 | 2020-03-20 | The National University Hospital of Iceland | deCODE genetics | Daniel F Gudbjartsson et al |
| 1585 | Iceland/489/2020 | 2020-03-20 | The National University Hospital of Iceland | deCODE genetics | Daniel F Gudbjartsson et al |
| 1586 | Iceland/509/2020 | 2020-03-20 | The National University Hospital of Iceland | deCODE genetics | Daniel F Gudbjartsson et al |
| 1587 | Iceland/443/2020 | 2020-03-20 | The National University Hospital of Iceland | deCODE genetics | Daniel F Gudbjartsson et al |
| 1588 | Iceland/508/2020 | 2020-03-20 | The National University Hospital of Iceland | deCODE genetics | Daniel F Gudbjartsson et al |
| 1589 | Iceland/483/2020 | 2020-03-20 | The National University Hospital of Iceland | deCODE genetics | Daniel F Gudbjartsson et al |
| 1590 | Iceland/462/2020 | 2020-03-20 | The National University Hospital of Iceland | deCODE genetics | Daniel F Gudbjartsson et al |
| 1591 | Iceland/437/2020 | 2020-03-20 | The National University Hospital of Iceland | deCODE genetics | Daniel F Gudbjartsson et al |
| 1592 | Iceland/463/2020 | 2020-03-20 | The National University Hospital of Iceland | deCODE genetics | Daniel F Gudbjartsson et al |
| 1593 | Iceland/490/2020 | 2020-03-20 | The National University Hospital of Iceland | deCODE genetics | Daniel F Gudbjartsson et al |
| 1594 | Iceland/454/2020 | 2020-03-20 | The National University Hospital of Iceland | deCODE genetics | Daniel F Gudbjartsson et al |
| 1595 | Iceland/487/2020 | 2020-03-20 | The National University Hospital of Iceland | deCODE genetics | Daniel F Gudbjartsson et al |
| 1596 | Iceland/455/2020 | 2020-03-20 | The National University Hospital of Iceland | deCODE genetics | Daniel F Gudbjartsson et al |
| 1597 | Iceland/488/2020 | 2020-03-20 | The National University Hospital of Iceland | deCODE genetics | Daniel F Gudbjartsson et al |
| 1598 | Iceland/510/2020 | 2020-03-20 | The National University Hospital of Iceland | deCODE genetics | Daniel F Gudbjartsson et al |
| 1599 | Iceland/442/2020 | 2020-03-20 | The National University Hospital of Iceland | deCODE genetics | Daniel F Gudbjartsson et al |
| 1600 | Iceland/436/2020 | 2020-03-20 | The National University Hospital of Iceland | deCODE genetics | Daniel F Gudbjartsson et al |
| 1601 | Iceland/460/2020 | 2020-03-20 | The National University Hospital of Iceland | deCODE genetics | Daniel F Gudbjartsson et al |
| 1602 | Iceland/450/2020 | 2020-03-20 | The National University Hospital of Iceland | deCODE genetics | Daniel F Gudbjartsson et al |
| 1603 | USA/NY-PV09041/2020 | 2020-03-20 | MSHS Clinical Microbiology Laboratories | MSHS Pathogen Surveillance Program | Ana S. Gonzalez-Reiche et al |
| 1604 | USA/NY-PV09190/2020 | 2020-03-20 | MSHS Clinical Microbiology Laboratories | MSHS Pathogen Surveillance Program | Ana S. Gonzalez-Reiche et al |
| 1605 | USA/WA-S220/2020 | 2020-03-20 | Washington State Department of Health | Seattle Flu Study | Chu et al et al |
| 1606 | USA/WA-S201/2020 | 2020-03-20 | Washington State Department of Health | Seattle Flu Study | Chu et al et al |
| 1607 | Iceland/441/2020 | 2020-03-20 | The National University | deCODE genetics | Daniel F Gudbjartsson |

| | | | | | |
|---|---|---|---|---|---|
| | | | Hospital of Iceland | | et al |
| 1608 | Iceland/445/2020 | 2020-03-20 | The National University Hospital of Iceland | deCODE genetics | Daniel F Gudbjartsson et al |
| 1609 | Iceland/471/2020 | 2020-03-20 | The National University Hospital of Iceland | deCODE genetics | Daniel F Gudbjartsson et al |
| 1610 | Iceland/468/2020 | 2020-03-20 | The National University Hospital of Iceland | deCODE genetics | Daniel F Gudbjartsson et al |
| 1611 | Iceland/449/2020 | 2020-03-20 | The National University Hospital of Iceland | deCODE genetics | Daniel F Gudbjartsson et al |
| 1612 | Iceland/494/2020 | 2020-03-20 | The National University Hospital of Iceland | deCODE genetics | Daniel F Gudbjartsson et al |
| 1613 | Iceland/425/2020 | 2020-03-20 | The National University Hospital of Iceland | deCODE genetics | Daniel F Gudbjartsson et al |
| 1614 | Iceland/448/2020 | 2020-03-20 | The National University Hospital of Iceland | deCODE genetics | Daniel F Gudbjartsson et al |
| 1615 | USA/NY-PV09186/2020 | 2020-03-20 | MSHS Clinical Microbiology Laboratories | MSHS Pathogen Surveillance Program | Ana S. Gonzalez-Reiche et al |
| 1616 | Portugal/PT0057/2020 | 2020-03-20 | H Dr. Nelio Mendonca - Funchal | Instituto Nacional de Saude (INSA) | Guiomar et al et al |
| 1617 | Iceland/461/2020 | 2020-03-20 | The National University Hospital of Iceland | deCODE genetics | Daniel F Gudbjartsson et al |
| 1618 | Iceland/476/2020 | 2020-03-20 | The National University Hospital of Iceland | deCODE genetics | Daniel F Gudbjartsson et al |
| 1619 | Taiwan/107/2020 | 2020-03-20 | Centers for Disease Control, R.O.C. (Taiwan) | Centers for Disease Control, R.O.C. (Taiwan) | Ji-Rong Yang et al |
| 1620 | Taiwan/TSGH-32/2020 | 2020-03-20 | TSGH-CP molecular lab | TSGH-CP molecular lab | Cherng-Lih Perng et al |
| 1621 | Taiwan/CGMH-CGU-25/2020 | 2020-03-20 | Laboratory Medicine | Department of Laboratory Medicine, Lin-Kou Chang Gung Memorial Hospital, Taoyuan, Taiwan | Kuo-Chien Tsao et al |
| 1622 | Taiwan/CGMH-CGU-24/2020 | 2020-03-20 | Laboratory Medicine | Department of Laboratory Medicine, Lin-Kou Chang Gung Memorial Hospital, Taoyuan, Taiwan | Kuo-Chien Tsao et al |
| 1623 | Taiwan/TSGH-30/2020 | 2020-03-20 | TSGH-CP molecular lab | TSGH-CP molecular lab | Cherng-Lih Perng et al |
| 1624 | Spain/Valencia173/2020 | 2020-03-20 | Servicio de Microbiologia. Hospital Universitario Doctor Peset | Sequencing and Bioinformatics Service and Molecular Epidemiology Research Group. FISABIO-Public Health | Juan Alberola Enguídanos et al |
| 1625 | Portugal/PT0064/2020 | 2020-03-20 | H Evora | Instituto Nacional de Saude (INSA) | Guiomar et al et al |
| 1626 | USA/WI-196/2020 | 2020-03-21 | University of Wisconsin-Madison AIDS Vaccine Research Laboratories | University of Wisconsin-Madison AIDS Vaccine Research Laboratories | Gage Moreno and Katarina Braun et al |
| 1627 | Scotland/EDB083/2020 | 2020-03-21 | Virology Department, Royal Infirmary of Edinburgh, NHS Lothian / School of Biological Sciences, University of Edinburgh / Institute of Genetics and Molecular Medicine, University of Edinburgh | COVID-19 Genomics UK (COG-UK) Consortium | McHugh M et al |
| 1628 | Australia/NSW168/2020 | 2020-03-21 | Centre for Infectious Diseases and Microbiology Public Health | NSW Health Pathology - Institute of Clinical Pathology and Medical Research; Westmead Hospital; University of Sydney | Arnott A et al |
| 1629 | Australia/VIC245/2020 | 2020-03-21 | Victorian Infectious Diseases Reference Laboratory (VIDRL) | Victorian Infectious Diseases Reference Laboratory and Microbiological Diagnostic Unit Public Health Laboratory, Doherty Institute | Caly L. et al |
| 1630 | Australia/VIC260/2020 | 2020-03-21 | Victorian Infectious Diseases Reference Laboratory (VIDRL) | Victorian Infectious Diseases Reference Laboratory and Microbiological Diagnostic Unit Public Health Laboratory, Doherty Institute | Caly L. et al |
| 1631 | Australia/VIC239/2020 | 2020-03-21 | Victorian Infectious Diseases Reference Laboratory (VIDRL) | Victorian Infectious Diseases Reference Laboratory and Microbiological Diagnostic Unit | Caly L. et al |

| | | | | | |
|---|---|---|---|---|---|
| | | | | Public Health Laboratory, Doherty Institute | |
| 1632 | Australia/VIC251/2020 | 2020-03-21 | Victorian Infectious Diseases Reference Laboratory (VIDRL) | Victorian Infectious Diseases Reference Laboratory and Microbiological Diagnostic Unit Public Health Laboratory, Doherty Institute | Caly L. et al |
| 1633 | Australia/NSW171/2020 | 2020-03-21 | Centre for Infectious Diseases and Microbiology Public Health | NSW Health Pathology - Institute of Clinical Pathology and Medical Research; Westmead Hospital; University of Sydney | Sim E et al |
| 1634 | Australia/VIC240/2020 | 2020-03-21 | Victorian Infectious Diseases Reference Laboratory (VIDRL) | Victorian Infectious Diseases Reference Laboratory and Microbiological Diagnostic Unit Public Health Laboratory, Doherty Institute | Caly L. et al |
| 1635 | Australia/VIC353/2020 | 2020-03-21 | Victorian Infectious Diseases Reference Laboratory (VIDRL) | Microbiological Diagnostic Unit Public Health Laboratory and Victorian Infectious Diseases Reference Laboratory, Doherty Institute | Caly L. et al |
| 1636 | Australia/VIC243/2020 | 2020-03-21 | Victorian Infectious Diseases Reference Laboratory (VIDRL) | Victorian Infectious Diseases Reference Laboratory and Microbiological Diagnostic Unit Public Health Laboratory, Doherty Institute | Caly L. et al |
| 1637 | Australia/VIC357/2020 | 2020-03-21 | Victorian Infectious Diseases Reference Laboratory (VIDRL) | Microbiological Diagnostic Unit Public Health Laboratory and Victorian Infectious Diseases Reference Laboratory, Doherty Institute | Caly L. et al |
| 1638 | Australia/VIC257/2020 | 2020-03-21 | Victorian Infectious Diseases Reference Laboratory (VIDRL) | Victorian Infectious Diseases Reference Laboratory and Microbiological Diagnostic Unit Public Health Laboratory, Doherty Institute | Caly L. et al |
| 1639 | Australia/VIC252/2020 | 2020-03-21 | Victorian Infectious Diseases Reference Laboratory (VIDRL) | Victorian Infectious Diseases Reference Laboratory and Microbiological Diagnostic Unit Public Health Laboratory, Doherty Institute | Caly L. et al |
| 1640 | Australia/VIC130/2020 | 2020-03-21 | Microbiological Diagnostic Unit Public Health Laboratory | Microbiological Diagnostic Unit Public Health Laboratory | Seemann T. et al |
| 1641 | Australia/NSW116/2020 | 2020-03-21 | Centre for Infectious Diseases and Microbiology Public Health | NSW Health Pathology - Institute of Clinical Pathology and Medical Research; Westmead Hospital; University of Sydney | Timms V et al |
| 1642 | Fujian/IM3520008T/2020 | 2020-03-21 | Fujian Center for Disease Control and Prevention | Fujian Center for Disease Control and Prevention | Lin Qi et al |
| 1643 | England/20134060904/2020 | 2020-03-21 | Respiratory Virus Unit, Microbiology Services Colindale, Public Health England | Respiratory Virus Unit, Microbiology Services Colindale, Public Health England | Monica Galiano et al |
| 1644 | Spain/Madrid_H12_3506/2020 | 2020-03-21 | Hospital Universitario 12 de Octubre | Hospital Universitario 12 de Octubre | Esther Viedma et al |
| 1645 | Netherlands/NoordBrabant_102/2020 | 2020-03-21 | Dutch COVID-19 response team | Erasmus Medical Center | Bas Oude Munnink et al |
| 1646 | Scotland/EDB095/2020 | 2020-03-21 | Virology Department, Royal Infirmary of Edinburgh, NHS Lothian / School of Biological Sciences, University of Edinburgh / Institute of Genetics and Molecular Medicine, University of Edinburgh | COVID-19 Genomics UK (COG-UK) Consortium | McHugh M et al |
| 1647 | Australia/NSW212/2020 | 2020-03-21 | Centre for Infectious Diseases and Microbiology Public Health | NSW Health Pathology - Institute of Clinical Pathology and Medical Research; Westmead Hospital; University of Sydney | Bachmann N et al |

| | | | | | |
|---|---|---|---|---|---|
| 1648 | Australia/NSW83/2020 | 2020-03-21 | ACT Pathology, The Canberra Hospital | NSW Health Pathology - Institute of Clinical Pathology and Medical Research; Westmead Hospital; University of Sydney | Rockett R et al |
| 1649 | Australia/NSW144/2020 | 2020-03-21 | Centre for Infectious Diseases and Microbiology Public Health | NSW Health Pathology - Institute of Clinical Pathology and Medical Research; Westmead Hospital; University of Sydney | Lam C et al |
| 1650 | Australia/VIC356/2020 | 2020-03-21 | Victorian Infectious Diseases Reference Laboratory (VIDRL) | Microbiological Diagnostic Unit Public Health Laboratory and Victorian Infectious Diseases Reference Laboratory, Doherty Institute | Caly L. et al |
| 1651 | Australia/VIC254/2020 | 2020-03-21 | Victorian Infectious Diseases Reference Laboratory (VIDRL) | Victorian Infectious Diseases Reference Laboratory and Microbiological Diagnostic Unit Public Health Laboratory, Doherty Institute | Caly L. et al |
| 1652 | Australia/NSW178/2020 | 2020-03-21 | Centre for Infectious Diseases and Microbiology Public Health | NSW Health Pathology - Institute of Clinical Pathology and Medical Research; Westmead Hospital; University of Sydney | Arnott A et al |
| 1653 | Australia/VIC248/2020 | 2020-03-21 | Victorian Infectious Diseases Reference Laboratory (VIDRL) | Victorian Infectious Diseases Reference Laboratory and Microbiological Diagnostic Unit Public Health Laboratory, Doherty Institute | Caly L. et al |
| 1654 | Australia/NSW182/2020 | 2020-03-21 | Centre for Infectious Diseases and Microbiology Public Health | NSW Health Pathology - Institute of Clinical Pathology and Medical Research; Westmead Hospital; University of Sydney | Bachmann N et al |
| 1655 | Australia/NSW110/2020 | 2020-03-21 | Centre for Infectious Diseases and Microbiology Public Health | NSW Health Pathology - Institute of Clinical Pathology and Medical Research; Westmead Hospital; University of Sydney | Draper J et al |
| 1656 | Australia/VIC256/2020 | 2020-03-21 | Victorian Infectious Diseases Reference Laboratory (VIDRL) | Victorian Infectious Diseases Reference Laboratory and Microbiological Diagnostic Unit Public Health Laboratory, Doherty Institute | Caly L. et al |
| 1657 | Australia/NSW183/2020 | 2020-03-21 | Centre for Infectious Diseases and Microbiology Public Health | NSW Health Pathology - Institute of Clinical Pathology and Medical Research; Westmead Hospital; University of Sydney | Rockett R et al |
| 1658 | Italy/TE5780/2020 | 2020-03-21 | Ospedale Regionale San Salvatore | Istituto Zooprofilattico Sperimentale dell’Abruzzo e Molise “G. Caporale” | Lorusso A et al |
| 1659 | England/SHEF-C07F8/2020 | 2020-03-21 | Virology Department, Sheffield Teaching Hospitals NHS Foundation Trust | Department of Infection, Immunity and Cardiovascular Disease, The Florey Institute, The Medical School, University of Sheffield | Thushan de Silva et al |
| 1660 | Australia/VIC250/2020 | 2020-03-21 | Victorian Infectious Diseases Reference Laboratory (VIDRL) | Victorian Infectious Diseases Reference Laboratory and Microbiological Diagnostic Unit Public Health Laboratory, Doherty Institute | Caly L. et al |
| 1661 | Australia/VIC236/2020 | 2020-03-21 | Victorian Infectious Diseases Reference Laboratory (VIDRL) | Victorian Infectious Diseases Reference Laboratory and Microbiological Diagnostic Unit Public Health Laboratory, Doherty Institute | Caly L. et al |
| 1662 | Australia/NSW201/2020 | 2020-03-21 | Centre for Infectious Diseases and Microbiology Public Health | NSW Health Pathology - Institute of Clinical Pathology and Medical Research; Westmead Hospital; University of Sydney | Sim E et al |

| | | | | | |
|---|---|---|---|---|---|
| 1663 | Australia/NSW174/2020 | 2020-03-21 | Centre for Infectious Diseases and Microbiology Public Health | NSW Health Pathology - Institute of Clinical Pathology and Medical Research; Westmead Hospital; University of Sydney | Lam C et al |
| 1664 | Australia/NSW189/2020 | 2020-03-21 | Centre for Infectious Diseases and Microbiology Public Health | NSW Health Pathology - Institute of Clinical Pathology and Medical Research; Westmead Hospital; University of Sydney | Sadsad R et al |
| 1665 | Australia/VIC231/2020 | 2020-03-21 | Victorian Infectious Diseases Reference Laboratory (VIDRL) | Victorian Infectious Diseases Reference Laboratory and Microbiological Diagnostic Unit Public Health Laboratory, Doherty Institute | Caly L. et al |
| 1666 | Australia/VIC354/2020 | 2020-03-21 | Victorian Infectious Diseases Reference Laboratory (VIDRL) | Microbiological Diagnostic Unit Public Health Laboratory and Victorian Infectious Diseases Reference Laboratory, Doherty Institute | Caly L. et al |
| 1667 | Australia/VIC242/2020 | 2020-03-21 | Victorian Infectious Diseases Reference Laboratory (VIDRL) | Victorian Infectious Diseases Reference Laboratory and Microbiological Diagnostic Unit Public Health Laboratory, Doherty Institute | Caly L. et al |
| 1668 | Australia/NSW205/2020 | 2020-03-21 | Centre for Infectious Diseases and Microbiology Public Health | NSW Health Pathology - Institute of Clinical Pathology and Medical Research; Westmead Hospital; University of Sydney | Gray K et al |
| 1669 | Australia/NSW197/2020 | 2020-03-21 | Centre for Infectious Diseases and Microbiology Public Health | NSW Health Pathology - Institute of Clinical Pathology and Medical Research; Westmead Hospital; University of Sydney | Gall M et al |
| 1670 | USA/WA-UW-1733/2020 | 2020-03-21 | UW Virology Lab | UW Virology Lab | Pavitra Roychoudhury et al |
| 1671 | India/S2/2020 | 2020-03-21 | ICMR-National Institute of Cholera and Enteric Diseases | National Institute of Biomedical Genomics | Arindam Maitra et al |
| 1672 | USA/NY-PV09029/2020 | 2020-03-21 | MSHS Clinical Microbiology Laboratories | MSHS Pathogen Surveillance Program | Ana S. Gonzalez-Reiche et al |
| 1673 | Australia/NSW115/2020 | 2020-03-21 | Centre for Infectious Diseases and Microbiology Public Health | NSW Health Pathology - Institute of Clinical Pathology and Medical Research; Westmead Hospital; University of Sydney | Gray K et al |
| 1674 | NewZealand/20VR1279/2020-03-21 | 2020-03-21 | LabPLUS | Institute of Environmental Science and Research (ESR) Ltd | **Matt Storey et al** |
| 1675 | NewZealand/20VR1290/2020-03-21 | 2020-03-21 | LabPLUS | Institute of Environmental Science and Research (ESR) Ltd | **Matt Storey et al** |
| 1676 | NewZealand/20VR1192/2020-03-21 | 2020-03-21 | PathLab Bay of Plenty | Institute of Environmental Science and Research (ESR) Ltd | **Matt Storey et al** |
| 1677 | NewZealand/20VR1273/2020-03-21 | 2020-03-21 | LabPLUS | Institute of Environmental Science and Research (ESR) Ltd | **Matt Storey et al** |
| 1678 | NewZealand/20VR1452/2020-03-21 | 2020-03-21 | Southern Community Labs Dunedin | Institute of Environmental Science and Research (ESR) Ltd | **Matt Storey et al** |
| 1679 | NewZealand/20VR1272/2020-03-21 | 2020-03-21 | LabPLUS | Institute of Environmental Science and Research (ESR) Ltd | **Matt Storey et al** |
| 1680 | NewZealand/20VR1274/2020-03-21 | 2020-03-21 | LabPLUS | Institute of Environmental Science and Research (ESR) Ltd | **Matt Storey et al** |
| 1681 | France/ARA11939/2020 | 2020-03-21 | Institut des Agents Infectieux (IAI), Hospices Civils de Lyon | CNR Virus des Infections Respiratoires - France SUD | Antonin Bal et al |
| 1682 | Germany/BAV-V2012856/2020 | 2020-03-21 | Max von Pettenkofer Institute, Virology, National Reference Center for Retroviruses, LMU Munich | Laboratory for Functional Genome Analysis, Dept. Genomics, Gene Center of the LMU Munich | Max Muenchhoff et al |
| 1683 | USA/WA-UW-1753/2020 | 2020-03-21 | UW Virology Lab | UW Virology Lab | Pavitra Roychoudhury et al |
| 1684 | USA/WI-180/2020 | 2020-03-21 | University of Wisconsin-Madison AIDS Vaccine Research Laboratories | University of Wisconsin-Madison AIDS Vaccine Research Laboratories | Gage Moreno and Katarina Braun et al |

| | | | | | |
|---|---|---|---|---|---|
| 1685 | Australia/NSW80/2020 | 2020-03-21 | Centre for Infectious Diseases and Microbiology Public Health | NSW Health Pathology - Institute of Clinical Pathology and Medical Research; Westmead Hospital; University of Sydney | Draper J et al |
| 1686 | Spain/Madrid_H12_28/2020 | 2020-03-21 | Hospital Universitario 12 de Octubre | Hospital Universitario 12 de Octubre | Sara González et al |
| 1687 | France/ARA11952/2020 | 2020-03-21 | Institut des Agents Infectieux (IAI), Hospices Civils de Lyon | CNR Virus des Infections Respiratoires - France SUD | Antonin Bal et al |
| 1688 | USA/ID-UW-2127/2020 | 2020-03-21 | UW Virology Lab | UW Virology Lab | Pavitra Roychoudhury et al |
| 1689 | Wales/PHWC-2522D/2020 | 2020-03-21 | Wales Specialist Virology Centre | Public Health Wales Microbiology Cardiff | Catherine Moore et al |
| 1690 | Australia/VIC229/2020 | 2020-03-21 | Victorian Infectious Diseases Reference Laboratory (VIDRL) | Victorian Infectious Diseases Reference Laboratory and Microbiological Diagnostic Unit Public Health Laboratory, Doherty Institute | Caly L. et al |
| 1691 | Iceland/497/2020 | 2020-03-21 | The National University Hospital of Iceland | deCODE genetics | Daniel F Gudbjartsson et al |
| 1692 | Iceland/499/2020 | 2020-03-21 | The National University Hospital of Iceland | deCODE genetics | Daniel F Gudbjartsson et al |
| 1693 | Iceland/502/2020 | 2020-03-21 | The National University Hospital of Iceland | deCODE genetics | Daniel F Gudbjartsson et al |
| 1694 | Iceland/496/2020 | 2020-03-21 | The National University Hospital of Iceland | deCODE genetics | Daniel F Gudbjartsson et al |
| 1695 | Iceland/504/2020 | 2020-03-21 | The National University Hospital of Iceland | deCODE genetics | Daniel F Gudbjartsson et al |
| 1696 | Iceland/500/2020 | 2020-03-21 | The National University Hospital of Iceland | deCODE genetics | Daniel F Gudbjartsson et al |
| 1697 | Iceland/514/2020 | 2020-03-21 | deCODE genetics | deCODE genetics | Daniel F Gudbjartsson et al |
| 1698 | Iceland/470/2020 | 2020-03-21 | The National University Hospital of Iceland | deCODE genetics | Daniel F Gudbjartsson et al |
| 1699 | Iceland/506/2020 | 2020-03-21 | The National University Hospital of Iceland | deCODE genetics | Daniel F Gudbjartsson et al |
| 1700 | Iceland/469/2020 | 2020-03-21 | The National University Hospital of Iceland | deCODE genetics | Daniel F Gudbjartsson et al |
| 1701 | Iceland/501/2020 | 2020-03-21 | The National University Hospital of Iceland | deCODE genetics | Daniel F Gudbjartsson et al |
| 1702 | Iceland/498/2020 | 2020-03-21 | The National University Hospital of Iceland | deCODE genetics | Daniel F Gudbjartsson et al |
| 1703 | Iceland/503/2020 | 2020-03-21 | The National University Hospital of Iceland | deCODE genetics | Daniel F Gudbjartsson et al |
| 1704 | Iceland/513/2020 | 2020-03-21 | deCODE genetics | deCODE genetics | Daniel F Gudbjartsson et al |
| 1705 | Iceland/505/2020 | 2020-03-21 | The National University Hospital of Iceland | deCODE genetics | Daniel F Gudbjartsson et al |
| 1706 | Spain/Madrid_H12_3910/2020 | 2020-03-21 | Hospital Universitario 12 de Octubre | Hospital Universitario 12 de Octubre | Esther Viedma et al |
| 1707 | Spain/Valencia271/2020 | 2020-03-21 | Servicio de Microbiologia. Hospital Clinico Universitario de Valencia | Sequencing and Bioinformatics Service and Molecular Epidemiology Research Group. FISABIO-Public Health | Marta Pla Diaz et al |
| 1708 | Iceland/495/2020 | 2020-03-21 | The National University Hospital of Iceland | deCODE genetics | Daniel F Gudbjartsson et al |
| 1709 | England/NOTT-10E335/2020 | 2020-03-21 | Queens Medical Centre, Clinical Microbiology Department / DeepSeq Nottingham | COVID-19 Genomics UK (COG-UK) Consortium | Gemma Clark et al |
| 1710 | USA/NY-PV09032/2020 | 2020-03-21 | MSHS Clinical Microbiology Laboratories | MSHS Pathogen Surveillance Program | Ana S. Gonzalez-Reiche et al |
| 1711 | USA/TX-HMH0142/2020 | 2020-03-21 | Houston Methodist Hospital | Houston Methodist Hospital | S. Wesley Long et al |
| 1712 | Australia/VIC262/2020 | 2020-03-21 | Victorian Infectious Diseases Reference Laboratory (VIDRL) | Victorian Infectious Diseases Reference Laboratory and Microbiological Diagnostic Unit Public Health Laboratory, Doherty Institute | Caly L. et al |
| 1713 | Taiwan/NTU22/2020 | 2020-03-21 | Department of Laboratory | Microbial Genomics Core Lab, | Shiou-Hwei Yeh et al |

| | | | | | |
|---|---|---|---|---|---|
| | | | Medicine, National Taiwan University Hospital | National Taiwan University Centers of Genomic and Precision Medicine | |
| 1714 | Taiwan/170/2020 | 2020-03-21 | Centers for Disease Control, R.O.C. (Taiwan) | Centers for Disease Control, R.O.C. (Taiwan) | Ji-Rong Yang et al |
| 1715 | Taiwan/CGMH-CGU-23/2020 | 2020-03-21 | Laboratory Medicine | Department of Laboratory Medicine, Lin-Kou Chang Gung Memorial Hospital, Taoyuan, Taiwan | Kuo-Chien Tsao et al |
| 1716 | USA/TX-HMH0129/2020 | 2020-03-21 | Houston Methodist Hospital | Houston Methodist Hospital | S. Wesley Long et al |
| 1717 | Scotland/CVR338/2020 | 2020-03-21 | West of Scotland Specialist Virology Centre, NHSGGC / MRC-University of Glasgow Centre for Virus Research | COVID-19 Genomics UK (COG-UK) Consortium | Ana da Silva Filipe et al |
| 1718 | USA/WA-UW-1879/2020 | 2020-03-21 | UW Virology Lab | UW Virology Lab | Pavitra Roychoudhury et al |
| 1719 | USA/WA-UW-1722/2020 | 2020-03-21 | UW Virology Lab | UW Virology Lab | Pavitra Roychoudhury et al |
| 1720 | Australia/VIC253/2020 | 2020-03-21 | Victorian Infectious Diseases Reference Laboratory (VIDRL) | Victorian Infectious Diseases Reference Laboratory and Microbiological Diagnostic Unit Public Health Laboratory, Doherty Institute | Caly L. et al |
| 1721 | Netherlands/NA_155/2020 | 2020-03-21 | Dutch COVID-19 response team | Erasmus Medical Center | Bas Oude Munnink et al |
| 1722 | USA/NY-PV09037/2020 | 2020-03-21 | MSHS Clinical Microbiology Laboratories | MSHS Pathogen Surveillance Program | Ana S. Gonzalez-Reiche et al |
| 1723 | Australia/VIC230/2020 | 2020-03-21 | Victorian Infectious Diseases Reference Laboratory (VIDRL) | Victorian Infectious Diseases Reference Laboratory and Microbiological Diagnostic Unit Public Health Laboratory, Doherty Institute | Caly L. et al |
| 1724 | Germany/NRW-46/2020 | 2020-03-21 | Center of Medical Microbiology, Virology, and Hospital Hygiene, University of Duesseldorf | Center of Medical Microbiology, Virology, and Hospital Hygiene, University of Duesseldorf | Ortwin Adams et al |
| 1725 | Fujian/IM3520014T/2020 | 2020-03-22 | Fujian Center for Disease Control and Prevention | Fujian Center for Disease Control and Prevention | Lin Qi et al |
| 1726 | France/IDF3230/2020 | 2020-03-22 | Labo BM - Site de Juvisy - Hopital Général | National Reference Center for Viruses of Respiratory Infections, Institut Pasteur, Paris | Mélanie Albert et al |
| 1727 | Australia/VIC135/2020 | 2020-03-22 | Microbiological Diagnostic Unit Public Health Laboratory | Microbiological Diagnostic Unit Public Health Laboratory | Seemann T. et al |
| 1728 | Australia/VIC278/2020 | 2020-03-22 | Victorian Infectious Diseases Reference Laboratory (VIDRL) | Victorian Infectious Diseases Reference Laboratory and Microbiological Diagnostic Unit Public Health Laboratory, Doherty Institute | Caly L. et al |
| 1729 | Australia/NSW118/2020 | 2020-03-22 | Centre for Infectious Diseases and Microbiology Public Health | NSW Health Pathology - Institute of Clinical Pathology and Medical Research; Westmead Hospital; University of Sydney | Arnott A et al |
| 1730 | Australia/NSW187/2020 | 2020-03-22 | Centre for Infectious Diseases and Microbiology Public Health | NSW Health Pathology - Institute of Clinical Pathology and Medical Research; Westmead Hospital; University of Sydney | Gall M et al |
| 1731 | Turkey/HSGM-8010/2020 | 2020-03-22 | Ministry of Health Turkey | Ministry of Health Turkey | Fatma Bayrakdar et al |
| 1732 | Australia/NSW98/2020 | 2020-03-22 | Centre for Infectious Diseases and Microbiology Public Health | NSW Health Pathology - Institute of Clinical Pathology and Medical Research; Westmead Hospital; University of Sydney | Arnott A et al |
| 1733 | Australia/NSW106/2020 | 2020-03-22 | Centre for Infectious Diseases and Microbiology Public Health | NSW Health Pathology - Institute of Clinical Pathology and Medical Research; Westmead Hospital; University of Sydney | Timms V et al |
| 1734 | Australia/VIC275/2020 | 2020-03-22 | Victorian Infectious Diseases Reference Laboratory | Victorian Infectious Diseases Reference Laboratory and | Caly L. et al |

| | | | | | |
|---|---|---|---|---|---|
| | | | (VIDRL) | Microbiological Diagnostic Unit Public Health Laboratory, Doherty Institute | |
| 1735 | Australia/VIC375/2020 | 2020-03-22 | Victorian Infectious Diseases Reference Laboratory (VIDRL) | Microbiological Diagnostic Unit Public Health Laboratory and Victorian Infectious Diseases Reference Laboratory, Doherty Institute | Caly L. et al |
| 1736 | Australia/NSW107/2020 | 2020-03-22 | Centre for Infectious Diseases and Microbiology Public Health | NSW Health Pathology - Institute of Clinical Pathology and Medical Research; Westmead Hospital; University of Sydney | Gall M et al |
| 1737 | Australia/VIC274/2020 | 2020-03-22 | Victorian Infectious Diseases Reference Laboratory (VIDRL) | Victorian Infectious Diseases Reference Laboratory and Microbiological Diagnostic Unit Public Health Laboratory, Doherty Institute | Caly L. et al |
| 1738 | Australia/VIC359/2020 | 2020-03-22 | Victorian Infectious Diseases Reference Laboratory (VIDRL) | Microbiological Diagnostic Unit Public Health Laboratory and Victorian Infectious Diseases Reference Laboratory, Doherty Institute | Caly L. et al |
| 1739 | Australia/VIC365/2020 | 2020-03-22 | Victorian Infectious Diseases Reference Laboratory (VIDRL) | Microbiological Diagnostic Unit Public Health Laboratory and Victorian Infectious Diseases Reference Laboratory, Doherty Institute | Caly L. et al |
| 1740 | Australia/NSW128/2020 | 2020-03-22 | Centre for Infectious Diseases and Microbiology Public Health | NSW Health Pathology - Institute of Clinical Pathology and Medical Research; Westmead Hospital; University of Sydney | Arnott A et al |
| 1741 | USA/TX-HMH0154/2020 | 2020-03-22 | Houston Methodist Hospital | Houston Methodist Hospital | S. Wesley Long et al |
| 1742 | Denmark/ALAB-SSI376/2020 | 2020-03-22 | Department of Virus and Microbiological Special Diagnostics, Statens Serum Institut, Copenhagen, Denmark, Artillerivej 5, 2300 Copenahgen S | Albertsen lab, Department of Chemistry and Bioscience, Aalborg University, Denmark | Rasmus Kirkegaard et al |
| 1743 | USA/ID-UW-1938/2020 | 2020-03-22 | UW Virology Lab | UW Virology Lab | Pavitra Roychoudhury et al |
| 1744 | France/ARA12388/2020 | 2020-03-22 | Institut des Agents Infectieux (IAI), Hospices Civils de Lyon | CNR Virus des Infections Respiratoires - France SUD | Antonin Bal et al |
| 1745 | Australia/VIC134/2020 | 2020-03-22 | Microbiological Diagnostic Unit Public Health Laboratory | Microbiological Diagnostic Unit Public Health Laboratory | Seemann T. et al |
| 1746 | Australia/VIC276/2020 | 2020-03-22 | Victorian Infectious Diseases Reference Laboratory (VIDRL) | Victorian Infectious Diseases Reference Laboratory and Microbiological Diagnostic Unit Public Health Laboratory, Doherty Institute | Caly L. et al |
| 1747 | Australia/VIC378/2020 | 2020-03-22 | Victorian Infectious Diseases Reference Laboratory (VIDRL) | Microbiological Diagnostic Unit Public Health Laboratory and Victorian Infectious Diseases Reference Laboratory, Doherty Institute | Caly L. et al |
| 1748 | Australia/VIC137/2020 | 2020-03-22 | Microbiological Diagnostic Unit Public Health Laboratory | Microbiological Diagnostic Unit Public Health Laboratory | Seemann T. et al |
| 1749 | Australia/NSW120/2020 | 2020-03-22 | Centre for Infectious Diseases and Microbiology Public Health | NSW Health Pathology - Institute of Clinical Pathology and Medical Research; Westmead Hospital; University of Sydney | Draper J et al |
| 1750 | Australia/VIC403/2020 | 2020-03-22 | Victorian Infectious Diseases Reference Laboratory (VIDRL) | Microbiological Diagnostic Unit Public Health Laboratory and Victorian Infectious Diseases Reference Laboratory, Doherty Institute | Caly L. et al |
| 1751 | Australia/VIC286/2020 | 2020-03-22 | Victorian Infectious Diseases Reference Laboratory (VIDRL) | Victorian Infectious Diseases Reference Laboratory and Microbiological Diagnostic Unit | Caly L. et al |

| | | | | | |
|---|---|---|---|---|---|
| | | | | Public Health Laboratory, Doherty Institute | |
| 1752 | Switzerland/GE0636/2020 | 2020-03-22 | University Hospitals of Geneva Laboratory of Virology | University Hospitals of Geneva Laboratory of Virology | Laubscher F. et al |
| 1753 | Greece/48/2020 | 2020-03-22 | Laboratory of Microbiology, Medical School, National and Kapodistrian University of Athens | Laboratory of Biology, Department of Medicine, Democritus University of Thrace | Kassela K. et al |
| 1754 | Australia/VIC132/2020 | 2020-03-22 | Microbiological Diagnostic Unit Public Health Laboratory | Microbiological Diagnostic Unit Public Health Laboratory | Seemann T. et al |
| 1755 | Australia/NSW90/2020 | 2020-03-22 | Centre for Infectious Diseases and Microbiology Public Health | NSW Health Pathology - Institute of Clinical Pathology and Medical Research; Westmead Hospital; University of Sydney | Draper J et al |
| 1756 | Australia/VIC264/2020 | 2020-03-22 | Victorian Infectious Diseases Reference Laboratory (VIDRL) | Victorian Infectious Diseases Reference Laboratory and Microbiological Diagnostic Unit Public Health Laboratory, Doherty Institute | Caly L. et al |
| 1757 | Kazakhstan/NCB-1/2020 | 2020-03-22 | Laboratory of Applied Genetics | "RSE ""National Center for Biotechnology""" | Shevtsov et al |
| 1758 | Wales/PHWC-25111/2020 | 2020-03-22 | Wales Specialist Virology Centre | Public Health Wales Microbiology Cardiff | Catherine Moore et al |
| 1759 | NewZealand/20VR1499/2020-03-22 | 2020-03-22 | PathLab Bay of Plenty | Institute of Environmental Science and Research (ESR) Ltd | **Matt Storey et al** |
| 1760 | NewZealand/20VR1799/2020-03-22 | 2020-03-22 | Middlemore Hospital | Institute of Environmental Science and Research (ESR) Ltd | **Matt Storey et al** |
| 1761 | NewZealand/20VR1078/2020-03-22 | 2020-03-22 | Wellington SCL (WR) | Institute of Environmental Science and Research (ESR) Ltd | **Matt Storey et al** |
| 1762 | NewZealand/20VR1380/2020-03-22 | 2020-03-22 | PathLab Bay of Plenty | Institute of Environmental Science and Research (ESR) Ltd | **Matt Storey et al** |
| 1763 | NewZealand/20VR1454/2020-03-22 | 2020-03-22 | Southern Community Labs Dunedin | Institute of Environmental Science and Research (ESR) Ltd | **Matt Storey et al** |
| 1764 | NewZealand/20VR1076/2020-03-22 | 2020-03-22 | Wellington SCL (WR) | Institute of Environmental Science and Research (ESR) Ltd | **Matt Storey et al** |
| 1765 | NewZealand/20VR1798/2020-03-22 | 2020-03-22 | Middlemore Hospital | Institute of Environmental Science and Research (ESR) Ltd | **Matt Storey et al** |
| 1766 | NewZealand/20VR1449/2020-03-22 | 2020-03-22 | Southern Community Labs Dunedin | Institute of Environmental Science and Research (ESR) Ltd | **Matt Storey et al** |
| 1767 | USA/WI-182/2020 | 2020-03-22 | University of Wisconsin-Madison AIDS Vaccine Research Laboratories | University of Wisconsin-Madison AIDS Vaccine Research Laboratories | Gage Moreno and Katarina Braun et al |
| 1768 | USA/NY-PV09307/2020 | 2020-03-22 | MSHS Clinical Microbiology Laboratories | MSHS Pathogen Surveillance Program | Ana S. Gonzalez-Reiche et al |
| 1769 | Australia/VIC266/2020 | 2020-03-22 | Victorian Infectious Diseases Reference Laboratory (VIDRL) | Victorian Infectious Diseases Reference Laboratory and Microbiological Diagnostic Unit Public Health Laboratory, Doherty Institute | Caly L. et al |
| 1770 | Spain/Madrid_H12_25/2020 | 2020-03-22 | Hospital Universitario 12 de Octubre | Hospital Universitario 12 de Octubre | Sara González et al |
| 1771 | Sweden/20-08687/2020 | 2020-03-22 | Laboratoriemedicin | The Public Health Agency of Sweden | Oskar Karlsson Lindsjo et al |
| 1772 | Sweden/20-08691/2020 | 2020-03-22 | Laboratoriemedicin | The Public Health Agency of Sweden | Oskar Karlsson Lindsjo et al |
| 1773 | USA/WA-UW-1770/2020 | 2020-03-22 | UW Virology Lab | UW Virology Lab | Pavitra Roychoudhury et al |
| 1774 | Australia/VIC265/2020 | 2020-03-22 | Victorian Infectious Diseases Reference Laboratory (VIDRL) | Victorian Infectious Diseases Reference Laboratory and Microbiological Diagnostic Unit Public Health Laboratory, Doherty Institute | Caly L. et al |
| 1775 | England/20132090404/2020 | 2020-03-22 | Respiratory Virus Unit, Microbiology Services Colindale, Public Health England | Respiratory Virus Unit, Microbiology Services Colindale, Public Health England | Monica Galiano et al |
| 1776 | USA/WA-UW-1846/2020 | 2020-03-22 | UW Virology Lab | UW Virology Lab | Pavitra Roychoudhury et al |

| | | | | | |
|---|---|---|---|---|---|
| 1777 | Australia/VIC502/2020 | 2020-03-22 | Victorian Infectious Diseases Reference Laboratory (VIDRL) | Microbiological Diagnostic Unit Public Health Laboratory and Victorian Infectious Diseases Reference Laboratory, Doherty Institute | Caly L. et al |
| 1778 | Taiwan/225/2020 | 2020-03-22 | Centers for Disease Control, R.O.C. (Taiwan) | Centers for Disease Control, R.O.C. (Taiwan) | Ji-Rong Yang et al |
| 1779 | Taiwan/TSGH-07/2020 | 2020-03-22 | TSGH-CP molecular lab | TSGH-CP molecular lab | Cherng-Lih Perng et al |
| 1780 | Taiwan/TSGH-25/2020 | 2020-03-22 | TSGH-CP molecular lab, Division of Clinical Pathology, Department of Pathology | TSGH-CP molecular lab, Division of Clinical Pathology, Department of Pathology | Cherng-Lih Perng et al |
| 1781 | Brazil/DF-891/2020 | 2020-03-22 | Instituto Oswaldo Cruz FIOCRUZ - Laboratory of Respiratory Viruses and Measles (LVRS) | Instituto Oswaldo Cruz FIOCRUZ - Laboratory of Respiratory Viruses and Measles (LVRS) | Paola Resende et al |
| 1782 | Australia/NSW206/2020 | 2020-03-22 | Centre for Infectious Diseases and Microbiology Public Health | NSW Health Pathology - Institute of Clinical Pathology and Medical Research; Westmead Hospital; University of Sydney | Timms V et al |
| 1783 | USA/UN-UW-1817/2020 | 2020-03-22 | UW Virology Lab | UW Virology Lab | Pavitra Roychoudhury et al |
| 1784 | Australia/VIC279/2020 | 2020-03-22 | Victorian Infectious Diseases Reference Laboratory (VIDRL) | Victorian Infectious Diseases Reference Laboratory and Microbiological Diagnostic Unit Public Health Laboratory, Doherty Institute | Caly L. et al |
| 1785 | France/ARA12260/2020 | 2020-03-22 | Institut des Agents Infectieux (IAI), Hospices Civils de Lyon | CNR Virus des Infections Respiratoires - France SUD | Antonin Bal et al |
| 1786 | USA/VA-DCLS-0040/2020 | 2020-03-22 | Virginia DCLS | Virginia DCLS | Virginia DCLS et al |
| 1787 | Australia/VIC285/2020 | 2020-03-22 | Victorian Infectious Diseases Reference Laboratory (VIDRL) | Victorian Infectious Diseases Reference Laboratory and Microbiological Diagnostic Unit Public Health Laboratory, Doherty Institute | Caly L. et al |
| 1788 | Denmark/ALAB-SSI375/2020 | 2020-03-22 | Department of Virus and Microbiological Special Diagnostics, Statens Serum Institut, Copenhagen, Denmark, Artillerivej 5, 2300 Copenahgen S | Albertsen lab, Department of Chemistry and Bioscience, Aalborg University, Denmark | Rasmus Kirkegaard et al |
| 1789 | Australia/VIC364/2020 | 2020-03-22 | Victorian Infectious Diseases Reference Laboratory (VIDRL) | Microbiological Diagnostic Unit Public Health Laboratory and Victorian Infectious Diseases Reference Laboratory, Doherty Institute | Caly L. et al |
| 1790 | USA/WI-46/2020 | 2020-03-22 | University of Wisconsin-Madison AIDS Vaccine Research Laboratories | University of Wisconsin-Madison AIDS Vaccine Research Laboratories | Moreno et al |
| 1791 | USA/WA-UW-1797/2020 | 2020-03-22 | UW Virology Lab | UW Virology Lab | Pavitra Roychoudhury et al |
| 1792 | USA/NY-PV09154/2020 | 2020-03-22 | MSHS Clinical Microbiology Laboratories | MSHS Pathogen Surveillance Program | Ana S. Gonzalez-Reiche et al |
| 1793 | USA/WI-39/2020 | 2020-03-22 | University of Wisconsin-Madison AIDS Vaccine Research Laboratories | University of Wisconsin-Madison AIDS Vaccine Research Laboratories | Moreno et al |
| 1794 | Australia/VIC284/2020 | 2020-03-22 | Victorian Infectious Diseases Reference Laboratory (VIDRL) | Victorian Infectious Diseases Reference Laboratory and Microbiological Diagnostic Unit Public Health Laboratory, Doherty Institute | Caly L. et al |
| 1795 | Netherlands/NA_173/2020 | 2020-03-22 | Dutch COVID-19 response team | Erasmus Medical Center | Bas Oude Munnink et al |
| 1796 | Spain/Valencia302/2020 | 2020-03-22 | Servicio de Microbiologia. Hospital Clinico Universitario de Valencia | Sequencing and Bioinformatics Service and Molecular Epidemiology Research Group. FISABIO-Public Health | Loreto Ferrús Abad et al |
| 1797 | USA/NY-PV09168/2020 | 2020-03-22 | MSHS Clinical Microbiology Laboratories | MSHS Pathogen Surveillance Program | Ana S. Gonzalez-Reiche et al |
| 1798 | France/ARA12996/2020 | 2020-03-23 | Centre Hospitalier de Bourg en Bresse | CNR Virus des Infections Respiratoires - France SUD | Antonin Bal et al |

| | | | | | |
|---|---|---|---|---|---|
| 1799 | Denmark/ALAB-SSI502/2020 | 2020-03-23 | Department of Virus and Microbiological Special Diagnostics, Statens Serum Institut, Copenhagen, Denmark, Artillerivej 5, 2300 Copenahgen S | Albertsen lab, Department of Chemistry and Bioscience, Aalborg University, Denmark | Rasmus Kirkegaard et al |
| 1800 | Belgium/NA-0323141/2020 | 2020-03-23 | KU Leuven, Clinical and Epidemiological Virology | KU Leuven, Clinical and Epidemiological Virology | Joan Marti-Carreras et al |
| 1801 | Australia/NSW137/2020 | 2020-03-23 | Centre for Infectious Diseases and Microbiology Public Health | NSW Health Pathology - Institute of Clinical Pathology and Medical Research; Westmead Hospital; University of Sydney | Gall M et al |
| 1802 | Australia/VIC496/2020 | 2020-03-23 | Victorian Infectious Diseases Reference Laboratory (VIDRL) | Microbiological Diagnostic Unit Public Health Laboratory and Victorian Infectious Diseases Reference Laboratory, Doherty Institute | Caly L. et al |
| 1803 | Australia/VIC295/2020 | 2020-03-23 | Victorian Infectious Diseases Reference Laboratory (VIDRL) | Victorian Infectious Diseases Reference Laboratory and Microbiological Diagnostic Unit Public Health Laboratory, Doherty Institute | Caly L. et al |
| 1804 | Australia/VIC-CBA1/2020 | 2020-03-23 | Geelong Centre for Emerging Infectious Diseases | Geelong Centre for Emerging Infectious Diseases | Chamings et al |
| 1805 | Australia/NSW146/2020 | 2020-03-23 | Centre for Infectious Diseases and Microbiology Public Health | NSW Health Pathology - Institute of Clinical Pathology and Medical Research; Westmead Hospital; University of Sydney | Timms V et al |
| 1806 | Australia/NSW147/2020 | 2020-03-23 | Centre for Infectious Diseases and Microbiology Public Health | NSW Health Pathology - Institute of Clinical Pathology and Medical Research; Westmead Hospital; University of Sydney | Gall M et al |
| 1807 | Australia/VIC304/2020 | 2020-03-23 | Victorian Infectious Diseases Reference Laboratory (VIDRL) | Victorian Infectious Diseases Reference Laboratory and Microbiological Diagnostic Unit Public Health Laboratory, Doherty Institute | Caly L. et al |
| 1808 | Australia/VIC806/2020 | 2020-03-23 | Victorian Infectious Diseases Reference Laboratory (VIDRL) | Microbiological Diagnostic Unit Public Health Laboratory and Victorian Infectious Diseases Reference Laboratory, Doherty Institute | Caly L. et al |
| 1809 | Netherlands/NA_191/2020 | 2020-03-23 | Dutch COVID-19 response team | Erasmus Medical Center | Bas Oude Munnink et al |
| 1810 | Australia/VIC299/2020 | 2020-03-23 | Victorian Infectious Diseases Reference Laboratory (VIDRL) | Victorian Infectious Diseases Reference Laboratory and Microbiological Diagnostic Unit Public Health Laboratory, Doherty Institute | Caly L. et al |
| 1811 | Australia/VIC297/2020 | 2020-03-23 | Victorian Infectious Diseases Reference Laboratory (VIDRL) | Victorian Infectious Diseases Reference Laboratory and Microbiological Diagnostic Unit Public Health Laboratory, Doherty Institute | Caly L. et al |
| 1812 | Australia/VIC307/2020 | 2020-03-23 | Victorian Infectious Diseases Reference Laboratory (VIDRL) | Victorian Infectious Diseases Reference Laboratory and Microbiological Diagnostic Unit Public Health Laboratory, Doherty Institute | Caly L. et al |
| 1813 | Australia/VIC382/2020 | 2020-03-23 | Victorian Infectious Diseases Reference Laboratory (VIDRL) | Microbiological Diagnostic Unit Public Health Laboratory and Victorian Infectious Diseases Reference Laboratory, Doherty Institute | Caly L. et al |
| 1814 | Australia/VIC385/2020 | 2020-03-23 | Victorian Infectious Diseases Reference Laboratory (VIDRL) | Microbiological Diagnostic Unit Public Health Laboratory and Victorian Infectious Diseases Reference Laboratory, Doherty Institute | Caly L. et al |

| | | | | | |
|---|---|---|---|---|---|
| 1815 | Australia/NSW198/2020 | 2020-03-23 | Centre for Infectious Diseases and Microbiology Public Health | NSW Health Pathology - Institute of Clinical Pathology and Medical Research; Westmead Hospital; University of Sydney | Arnott A et al |
| 1816 | Australia/VIC310/2020 | 2020-03-23 | Microbiological Diagnostic Unit Public Health Laboratory | Microbiological Diagnostic Unit Public Health Laboratory | Seemann T. et al |
| 1817 | Australia/NSW194/2020 | 2020-03-23 | Centre for Infectious Diseases and Microbiology Public Health | NSW Health Pathology - Institute of Clinical Pathology and Medical Research; Westmead Hospital; University of Sydney | Lam C et al |
| 1818 | Australia/VIC291/2020 | 2020-03-23 | Victorian Infectious Diseases Reference Laboratory (VIDRL) | Victorian Infectious Diseases Reference Laboratory and Microbiological Diagnostic Unit Public Health Laboratory, Doherty Institute | Caly L. et al |
| 1819 | Australia/NSW209/2020 | 2020-03-23 | Centre for Infectious Diseases and Microbiology Public Health | NSW Health Pathology - Institute of Clinical Pathology and Medical Research; Westmead Hospital; University of Sydney | Sadsad R et al |
| 1820 | Australia/VIC301/2020 | 2020-03-23 | Victorian Infectious Diseases Reference Laboratory (VIDRL) | Victorian Infectious Diseases Reference Laboratory and Microbiological Diagnostic Unit Public Health Laboratory, Doherty Institute | Caly L. et al |
| 1821 | Australia/VIC298/2020 | 2020-03-23 | Victorian Infectious Diseases Reference Laboratory (VIDRL) | Victorian Infectious Diseases Reference Laboratory and Microbiological Diagnostic Unit Public Health Laboratory, Doherty Institute | Caly L. et al |
| 1822 | Australia/VIC516/2020 | 2020-03-23 | Victorian Infectious Diseases Reference Laboratory (VIDRL) | Microbiological Diagnostic Unit Public Health Laboratory and Victorian Infectious Diseases Reference Laboratory, Doherty Institute | Caly L. et al |
| 1823 | Australia/VIC383/2020 | 2020-03-23 | Victorian Infectious Diseases Reference Laboratory (VIDRL) | Microbiological Diagnostic Unit Public Health Laboratory and Victorian Infectious Diseases Reference Laboratory, Doherty Institute | Caly L. et al |
| 1824 | Australia/VIC381/2020 | 2020-03-23 | Victorian Infectious Diseases Reference Laboratory (VIDRL) | Microbiological Diagnostic Unit Public Health Laboratory and Victorian Infectious Diseases Reference Laboratory, Doherty Institute | Caly L. et al |
| 1825 | Australia/NSW148/2020 | 2020-03-23 | Centre for Infectious Diseases and Microbiology Public Health | NSW Health Pathology - Institute of Clinical Pathology and Medical Research; Westmead Hospital; University of Sydney | Arnott A et al |
| 1826 | Australia/VIC544/2020 | 2020-03-23 | Victorian Infectious Diseases Reference Laboratory (VIDRL) | Microbiological Diagnostic Unit Public Health Laboratory and Victorian Infectious Diseases Reference Laboratory, Doherty Institute | Caly L. et al |
| 1827 | NanChang/JX216/2020 | 2020-03-23 | Jiangxi province Center for Disease Control and Prevention | Jiangxi province Center for Disease Control and Prevention | Li jian Xiong et al |
| 1828 | USA/WA-UW-1919/2020 | 2020-03-23 | UW Virology Lab | UW Virology Lab | Pavitra Roychoudhury et al |
| 1829 | USA/WI-186/2020 | 2020-03-23 | University of Wisconsin-Madison AIDS Vaccine Research Laboratories | University of Wisconsin-Madison AIDS Vaccine Research Laboratories | Gage Moreno and Katarina Braun et al |
| 1830 | Scotland/CVR462/2020 | 2020-03-23 | West of Scotland Specialist Virology Centre, NHSGGC / MRC-University of Glasgow Centre for Virus Research | COVID-19 Genomics UK (COG-UK) Consortium | Ana da Silva Filipe et al |
| 1831 | Netherlands/NA_185/2020 | 2020-03-23 | Dutch COVID-19 response team | Erasmus Medical Center | Bas Oude Munnink et al |
| 1832 | France/ARA12485/2020 | 2020-03-23 | Institut des Agents Infectieux | CNR Virus des Infections | Antonin Bal et al |

| | | | | | |
|---|---|---|---|---|---|
| | | | (IAI), Hospices Civils de Lyon | Respiratoires - France SUD | |
| 1833 | Australia/NSW105/2020 | 2020-03-23 | Centre for Infectious Diseases and Microbiology Public Health | NSW Health Pathology - Institute of Clinical Pathology and Medical Research; Westmead Hospital; University of Sydney | Gray K et al |
| 1834 | Australia/VIC384/2020 | 2020-03-23 | Victorian Infectious Diseases Reference Laboratory (VIDRL) | Microbiological Diagnostic Unit Public Health Laboratory and Victorian Infectious Diseases Reference Laboratory, Doherty Institute | Caly L. et al |
| 1835 | USA/TX-HMH0157/2020 | 2020-03-23 | Houston Methodist Hospital | Houston Methodist Hospital | S. Wesley Long et al |
| 1836 | USA/TX-HMH0195/2020 | 2020-03-23 | Houston Methodist Hospital | Houston Methodist Hospital | S. Wesley Long et al |
| 1837 | Spain/Valencia221/2020 | 2020-03-23 | Servicio de Microbiologia. Hospital Clinico Universitario de Valencia | Sequencing and Bioinformatics Service and Molecular Epidemiology Research Group. FISABIO-Public Health | Marta Pla Diaz et al |
| 1838 | Australia/NSW179/2020 | 2020-03-23 | Centre for Infectious Diseases and Microbiology Public Health | NSW Health Pathology - Institute of Clinical Pathology and Medical Research; Westmead Hospital; University of Sydney | Sadsad R et al |
| 1839 | Australia/VIC309/2020 | 2020-03-23 | Microbiological Diagnostic Unit Public Health Laboratory | Microbiological Diagnostic Unit Public Health Laboratory | Seemann T. et al |
| 1840 | Australia/VIC538/2020 | 2020-03-23 | Victorian Infectious Diseases Reference Laboratory (VIDRL) | Microbiological Diagnostic Unit Public Health Laboratory and Victorian Infectious Diseases Reference Laboratory, Doherty Institute | Caly L. et al |
| 1841 | Australia/VIC302/2020 | 2020-03-23 | Victorian Infectious Diseases Reference Laboratory (VIDRL) | Victorian Infectious Diseases Reference Laboratory and Microbiological Diagnostic Unit Public Health Laboratory, Doherty Institute | Caly L. et al |
| 1842 | Australia/VIC296/2020 | 2020-03-23 | Victorian Infectious Diseases Reference Laboratory (VIDRL) | Victorian Infectious Diseases Reference Laboratory and Microbiological Diagnostic Unit Public Health Laboratory, Doherty Institute | Caly L. et al |
| 1843 | Australia/NSW97/2020 | 2020-03-23 | Centre for Infectious Diseases and Microbiology Public Health | NSW Health Pathology - Institute of Clinical Pathology and Medical Research; Westmead Hospital; University of Sydney | Gall M et al |
| 1844 | Australia/VIC520/2020 | 2020-03-23 | Victorian Infectious Diseases Reference Laboratory (VIDRL) | Microbiological Diagnostic Unit Public Health Laboratory and Victorian Infectious Diseases Reference Laboratory, Doherty Institute | Caly L. et al |
| 1845 | Australia/NSW73/2020 | 2020-03-23 | Centre for Infectious Diseases and Microbiology Public Health | NSW Health Pathology - Institute of Clinical Pathology and Medical Research; Westmead Hospital; University of Sydney | Rockett R et al |
| 1846 | Denmark/ALAB-SSI361/2020 | 2020-03-23 | Department of Virus and Microbiological Special Diagnostics, Statens Serum Institut, Copenhagen, Denmark, Artillerivej 5, 2300 Copenahgen S | Albertsen lab, Department of Chemistry and Bioscience, Aalborg University, Denmark | Rasmus Kirkegaard et al |
| 1847 | USA/WA-UW-1801/2020 | 2020-03-23 | UW Virology Lab | UW Virology Lab | Pavitra Roychoudhury et al |
| 1848 | USA/WA-S190/2020 | 2020-03-23 | Washington State Department of Health | Seattle Flu Study | Chu et al et al |
| 1849 | USA/WI-33/2020 | 2020-03-23 | University of Wisconsin-Madison AIDS Vaccine Research Laboratories | University of Wisconsin-Madison AIDS Vaccine Research Laboratories | Moreno et al |
| 1850 | NewZealand/20VR1451/2020-03-23 | 2020-03-23 | Southern Community Labs Dunedin | Institute of Environmental Science and Research (ESR) Ltd | **Matt Storey et al** |
| 1851 | NewZealand/20VR1617/2020-03-23 | 2020-03-23 | Southern Community Labs Dunedin | Institute of Environmental Science and Research (ESR) Ltd | **Matt Storey et al** |

| | | | | | |
|---|---|---|---|---|---|
| 1852 | NewZealand/20VR1791/2020-03-23 | 2020-03-23 | Waikato Hospital | Institute of Environmental Science and Research (ESR) Ltd | **Matt Storey et al** |
| 1853 | NewZealand/20VR1109/2020-03-23 | 2020-03-23 | Waikato Hospital | Institute of Environmental Science and Research (ESR) Ltd | **Matt Storey et al** |
| 1854 | NewZealand/20VR1619/2020-03-23 | 2020-03-23 | Southern Community Labs Dunedin | Institute of Environmental Science and Research (ESR) Ltd | **Matt Storey et al** |
| 1855 | NewZealand/20VR1618/2020-03-23 | 2020-03-23 | Southern Community Labs Dunedin | Institute of Environmental Science and Research (ESR) Ltd | **Matt Storey et al** |
| 1856 | NewZealand/20VR1797/2020-03-23 | 2020-03-23 | Middlemore Hospital | Institute of Environmental Science and Research (ESR) Ltd | **Matt Storey et al** |
| 1857 | NewZealand/20VR2034/2020-03-23 | 2020-03-23 | Wellington SCL (WN) | Institute of Environmental Science and Research (ESR) Ltd | **Matt Storey et al** |
| 1858 | NewZealand/20VR1796/2020-03-23 | 2020-03-23 | Middlemore Hospital | Institute of Environmental Science and Research (ESR) Ltd | **Matt Storey et al** |
| 1859 | NewZealand/20VR2015/2020-03-23 | 2020-03-23 | Wellington SCL (WN) | Institute of Environmental Science and Research (ESR) Ltd | **Matt Storey et al** |
| 1860 | NewZealand/20VR1453/2020-03-23 | 2020-03-23 | Southern Community Labs Dunedin | Institute of Environmental Science and Research (ESR) Ltd | **Matt Storey et al** |
| 1861 | NewZealand/20VR1626/2020-03-23 | 2020-03-23 | Southern Community Labs Dunedin | Institute of Environmental Science and Research (ESR) Ltd | **Matt Storey et al** |
| 1862 | NewZealand/20VR1627/2020-03-23 | 2020-03-23 | Southern Community Labs Dunedin | Institute of Environmental Science and Research (ESR) Ltd | **Matt Storey et al** |
| 1863 | England/20134064004/2020 | 2020-03-23 | Respiratory Virus Unit, Microbiology Services Colindale, Public Health England | Respiratory Virus Unit, Microbiology Services Colindale, Public Health England | Monica Galiano et al |
| 1864 | USA/WA-UW-1860/2020 | 2020-03-23 | UW Virology Lab | UW Virology Lab | Pavitra Roychoudhury et al |
| 1865 | USA/WA-S539/2020 | 2020-03-23 | Washington State Department of Health | Seattle Flu Study | Chu et al et al |
| 1866 | Belgium/MC-0323129/2020 | 2020-03-23 | KU Leuven, Clinical and Epidemiological Virology | KU Leuven, Clinical and Epidemiological Virology | Joan Marti-Carreras et al |
| 1867 | Italy/INMI6/2020 | 2020-03-23 | INMI Lazzaro Spallanzani IRCCS | Laboratory of Virology, INMI Lazzaro Spallanzani IRCCS | Barbara Bartolini et al |
| 1868 | Italy/TE6195/2020 | 2020-03-23 | Ospedale Civile Giuseppe Mazzini | Istituto Zooprofilattico Sperimentale dell'Abruzzo e Molise "G. Caporale" | Lorusso A et al |
| 1869 | Italy/INMI9/2020 | 2020-03-23 | INMI Lazzaro Spallanzani IRCCS | Laboratory of Virology, INMI Lazzaro Spallanzani IRCCS | Fabrizio Carletti et al |
| 1870 | USA/WI-25/2020 | 2020-03-23 | University of Wisconsin-Madison AIDS Vaccine Research Laboratories | University of Wisconsin-Madison AIDS Vaccine Research Laboratories | Moreno et al |
| 1871 | Italy/INMI7/2020 | 2020-03-23 | INMI Lazzaro Spallanzani IRCCS | INMI Lazzaro Spallanzani IRCCS | Antonino Di Caro et al |
| 1872 | USA/WI-55/2020 | 2020-03-23 | University of Wisconsin-Madison AIDS Vaccine Research Laboratories | University of Wisconsin-Madison AIDS Vaccine Research Laboratories | Moreno et al |
| 1873 | USA/OR-UW-1885/2020 | 2020-03-23 | UW Virology Lab | UW Virology Lab | Pavitra Roychoudhury et al |
| 1874 | USA/TX-HMH0160/2020 | 2020-03-23 | Houston Methodist Hospital | Houston Methodist Hospital | S. Wesley Long et al |
| 1875 | USA/WA-UW-1904/2020 | 2020-03-23 | UW Virology Lab | UW Virology Lab | Pavitra Roychoudhury et al |
| 1876 | France/HF3677/2020 | 2020-03-23 | CH Compiègne Laboratoire de Biologie | National Reference Center for Viruses of Respiratory Infections, Institut Pasteur, Paris | Mélanie Albert et al |
| 1877 | England/NOTT-10E5F3/2020 | 2020-03-23 | Queens Medical Centre, Clinical Microbiology Department / DeepSeq Nottingham | COVID-19 Genomics UK (COG-UK) Consortium | Gemma Clark et al |
| 1878 | Russia/Moscow-80402/2020 | 2020-03-23 | State Research Center of Virology and Biotechnology VECTOR, Department of Collection of Microorganisms | State Research Center of Virology and Biotechnology VECTOR, Department of Collection of Microorganisms | Sergey A. Bodnev et al |
| 1879 | England/SHEF-C83ED/2020 | 2020-03-23 | Virology Department, Sheffield Teaching Hospitals NHS Foundation Trust / Virology Department, Sheffield Teaching Hospitals NHS Foundation Trust | COVID-19 Genomics UK (COG-UK) Consortium | Thushan de Silva et al |

| | | | | | |
|---|---|---|---|---|---|
| 1880 | Netherlands/NA_194/2020 | 2020-03-23 | Dutch COVID-19 response team | Erasmus Medical Center | Bas Oude Munnink et al |
| 1881 | Scotland/EDB157/2020 | 2020-03-23 | Virology Department, Royal Infirmary of Edinburgh, NHS Lothian / School of Biological Sciences, University of Edinburgh / Institute of Genetics and Molecular Medicine, University of Edinburgh | COVID-19 Genomics UK (COG-UK) Consortium | McHugh M et al |
| 1882 | Russia/Moscow-80112/2020 | 2020-03-23 | State Research Center of Virology and Biotechnology VECTOR, Department of Collection of Microorganisms | State Research Center of Virology and Biotechnology VECTOR, Department of Collection of Microorganisms | Sergey A. Bodnev et al |
| 1883 | Sweden/20-08683/2020 | 2020-03-23 | Laboratoriemedicin | The Public Health Agency of Sweden | Olov Svartstrom et al |
| 1884 | Italy/6193/2020 | 2020-03-23 | Ospedale Civile Giuseppe Mazzini | Istituto Zooprofilattico Sperimentale dell'Abruzzo e Molise "G. Caporale" | Lorusso A et al |
| 1885 | Taiwan/TSGH-08/2020 | 2020-03-23 | TSGH-CP molecular lab | TSGH-CP molecular lab | Cherng-Lih Perng et al |
| 1886 | Taiwan/108/2020 | 2020-03-23 | Centers for Disease Control, R.O.C. (Taiwan) | Centers for Disease Control, R.O.C. (Taiwan) | Ji-Rong Yang et al |
| 1887 | Taiwan/TSGH-09/2020 | 2020-03-23 | TSGH-CP molecular lab | TSGH-CP molecular lab | Cherng-Lih Perng et al |
| 1888 | USA/ID-UW-2160/2020 | 2020-03-23 | UW Virology Lab | UW Virology Lab | Pavitra Roychoudhury et al |
| 1889 | USA/ID-UW-2174/2020 | 2020-03-23 | UW Virology Lab | UW Virology Lab | Pavitra Roychoudhury et al |
| 1890 | England/20132026904/2020 | 2020-03-23 | Respiratory Virus Unit, Microbiology Services Colindale, Public Health England | Respiratory Virus Unit, Microbiology Services Colindale, Public Health England | Monica Galiano et al |
| 1891 | England/20132017604/2020 | 2020-03-23 | Respiratory Virus Unit, Microbiology Services Colindale, Public Health England | Respiratory Virus Unit, Microbiology Services Colindale, Public Health England | Monica Galiano et al |
| 1892 | France/ARA12632/2020 | 2020-03-23 | Centre Hospitalier de Macon | CNR Virus des Infections Respiratoires - France SUD | Antonin Bal et al |
| 1893 | France/HF3290/2020 | 2020-03-23 | CH Compiègne Laboratoire de Biologie | National Reference Center for Viruses of Respiratory Infections, Institut Pasteur, Paris | Mélanie Albert et al |
| 1894 | NewZealand/20VR1450/2020-03-24 | 2020-03-24 | Southern Community Labs Dunedin | Institute of Environmental Science and Research (ESR) Ltd | **Matt Storey et al** |
| 1895 | PuertoRico/CDC-S11/2020 | 2020-03-24 | Puerto Rico Department of Health | Centers for Disease Control and Prevention, Dengue Branch | Gilberto A. Santiago et al |
| 1896 | Australia/VIC541/2020 | 2020-03-24 | Victorian Infectious Diseases Reference Laboratory (VIDRL) | Microbiological Diagnostic Unit Public Health Laboratory and Victorian Infectious Diseases Reference Laboratory, Doherty Institute | Caly L. et al |
| 1897 | Australia/VIC398/2020 | 2020-03-24 | Victorian Infectious Diseases Reference Laboratory (VIDRL) | Microbiological Diagnostic Unit Public Health Laboratory and Victorian Infectious Diseases Reference Laboratory, Doherty Institute | Caly L. et al |
| 1898 | Australia/VIC319/2020 | 2020-03-24 | Microbiological Diagnostic Unit Public Health Laboratory | Microbiological Diagnostic Unit Public Health Laboratory | Seemann T. et al |
| 1899 | Australia/VIC318/2020 | 2020-03-24 | Microbiological Diagnostic Unit Public Health Laboratory | Microbiological Diagnostic Unit Public Health Laboratory | Seemann T. et al |
| 1900 | Australia/VIC394/2020 | 2020-03-24 | Victorian Infectious Diseases Reference Laboratory (VIDRL) | Microbiological Diagnostic Unit Public Health Laboratory and Victorian Infectious Diseases Reference Laboratory, Doherty Institute | Caly L. et al |
| 1901 | Australia/NSW87/2020 | 2020-03-24 | Centre for Infectious Diseases and Microbiology Public Health | NSW Health Pathology - Institute of Clinical Pathology and Medical Research; Westmead Hospital; University of Sydney | Gall M et al |
| 1902 | Australia/NSW109/2020 | 2020-03-24 | Centre for Infectious Diseases and Microbiology Public Health | NSW Health Pathology - Institute of Clinical Pathology and Medical Research; | Sadsad R et al |

| | | | | | |
|---|---|---|---|---|---|
| | | | | Westmead Hospital; University of Sydney | |
| 1903 | Australia/NSW127/2020 | 2020-03-24 | Centre for Infectious Diseases and Microbiology Public Health | NSW Health Pathology - Institute of Clinical Pathology and Medical Research; Westmead Hospital; University of Sydney | Gall M et al |
| 1904 | Australia/VIC396/2020 | 2020-03-24 | Victorian Infectious Diseases Reference Laboratory (VIDRL) | Microbiological Diagnostic Unit Public Health Laboratory and Victorian Infectious Diseases Reference Laboratory, Doherty Institute | Caly L. et al |
| 1905 | Australia/VIC-CBA2/2020 | 2020-03-24 | Geelong Centre for Emerging Infectious Diseases | Geelong Centre for Emerging Infectious Diseases | Chamings et al |
| 1906 | Spain/Valencia192/2020 | 2020-03-24 | Servicio de Microbiologia. Hospital Clinico Universitario de Valencia | Sequencing and Bioinformatics Service and Molecular Epidemiology Research Group. FISABIO-Public Health | Paula Ruiz-Hueso et al |
| 1907 | Italy/TE6580/2020 | 2020-03-24 | Ospedale Civile Giuseppe Mazzini | "Istituto Zooprofilattico Sperimentale dell'Abruzzo e Molise ""G. Caporale""" | Lorusso A et al |
| 1908 | Italy/TE6644/2020 | 2020-03-24 | Ospedale Civile Giuseppe Mazzini | Istituto Zooprofilattico Sperimentale dell’Abruzzo e Molise “G. Caporale” | Lorusso A et al |
| 1909 | Australia/VIC646/2020 | 2020-03-24 | Victorian Infectious Diseases Reference Laboratory (VIDRL) | Microbiological Diagnostic Unit Public Health Laboratory and Victorian Infectious Diseases Reference Laboratory, Doherty Institute | Caly L. et al |
| 1910 | Australia/VIC323/2020 | 2020-03-24 | Microbiological Diagnostic Unit Public Health Laboratory | Microbiological Diagnostic Unit Public Health Laboratory | Seemann T. et al |
| 1911 | Australia/VIC404/2020 | 2020-03-24 | Victorian Infectious Diseases Reference Laboratory (VIDRL) | Microbiological Diagnostic Unit Public Health Laboratory and Victorian Infectious Diseases Reference Laboratory, Doherty Institute | Caly L. et al |
| 1912 | Australia/QLDID929/2020 | 2020-03-24 | Mater Pathology | Public Health Virology Laboratory | Bixing Huang et al |
| 1913 | Australia/NSW129/2020 | 2020-03-24 | Centre for Infectious Diseases and Microbiology Public Health | NSW Health Pathology - Institute of Clinical Pathology and Medical Research; Westmead Hospital; University of Sydney | Sadsad R et al |
| 1914 | Australia/VIC325/2020 | 2020-03-24 | Microbiological Diagnostic Unit Public Health Laboratory | Microbiological Diagnostic Unit Public Health Laboratory | Seemann T. et al |
| 1915 | Australia/VIC500/2020 | 2020-03-24 | Victorian Infectious Diseases Reference Laboratory (VIDRL) | Microbiological Diagnostic Unit Public Health Laboratory and Victorian Infectious Diseases Reference Laboratory, Doherty Institute | Caly L. et al |
| 1916 | Australia/VIC317/2020 | 2020-03-24 | Microbiological Diagnostic Unit Public Health Laboratory | Microbiological Diagnostic Unit Public Health Laboratory | Seemann T. et al |
| 1917 | USA/WA-UW-2247/2020 | 2020-03-24 | UW Virology Lab | UW Virology Lab | Pavitra Roychoudhury et al |
| 1918 | England/201340127/2020 | 2020-03-24 | Respiratory Virus Unit, Microbiology Services Colindale, Public Health England | Respiratory Virus Unit, Microbiology Services Colindale, Public Health England | Monica Galiano et al |
| 1919 | Australia/NSW138/2020 | 2020-03-24 | Centre for Infectious Diseases and Microbiology Public Health | NSW Health Pathology - Institute of Clinical Pathology and Medical Research; Westmead Hospital; University of Sydney | Arnott A et al |
| 1920 | Australia/VIC395/2020 | 2020-03-24 | Victorian Infectious Diseases Reference Laboratory (VIDRL) | Microbiological Diagnostic Unit Public Health Laboratory and Victorian Infectious Diseases Reference Laboratory, Doherty Institute | Caly L. et al |
| 1921 | Australia/VIC326/2020 | 2020-03-24 | Microbiological Diagnostic Unit Public Health Laboratory | Microbiological Diagnostic Unit Public Health Laboratory | Seemann T. et al |
| 1922 | USA/AK-PHL040/2020 | 2020-03-24 | Alaska State Virology | Alaska State Virology | Jack Chen et al |

| | | | | | |
|---|---|---|---|---|---|
| | | | Laboratory | Laboratory | |
| 1923 | USA/WA-S529/2020 | 2020-03-24 | Washington State Department of Health | Seattle Flu Study | Chu et al et al |
| 1924 | Australia/VIC316/2020 | 2020-03-24 | Microbiological Diagnostic Unit Public Health Laboratory | Microbiological Diagnostic Unit Public Health Laboratory | Seemann T. et al |
| 1925 | Australia/VIC320/2020 | 2020-03-24 | Microbiological Diagnostic Unit Public Health Laboratory | Microbiological Diagnostic Unit Public Health Laboratory | Seemann T. et al |
| 1926 | Australia/VIC399/2020 | 2020-03-24 | Victorian Infectious Diseases Reference Laboratory (VIDRL) | Microbiological Diagnostic Unit Public Health Laboratory and Victorian Infectious Diseases Reference Laboratory, Doherty Institute | Caly L. et al |
| 1927 | Australia/VIC540/2020 | 2020-03-24 | Victorian Infectious Diseases Reference Laboratory (VIDRL) | Microbiological Diagnostic Unit Public Health Laboratory and Victorian Infectious Diseases Reference Laboratory, Doherty Institute | Caly L. et al |
| 1928 | Australia/VIC322/2020 | 2020-03-24 | Microbiological Diagnostic Unit Public Health Laboratory | Microbiological Diagnostic Unit Public Health Laboratory | Seemann T. et al |
| 1929 | Australia/NT17/2020 | 2020-03-24 | Royal Darwin Hospital Pathology | Microbiological Diagnostic Unit Public Health Laboratory and Victorian Infectious Diseases Reference Laboratory, Doherty Institute | Meumann et al |
| 1930 | Australia/NSW139/2020 | 2020-03-24 | Centre for Infectious Diseases and Microbiology Public Health | NSW Health Pathology - Institute of Clinical Pathology and Medical Research; Westmead Hospital; University of Sydney | Sadsad R et al |
| 1931 | Australia/VIC324/2020 | 2020-03-24 | Microbiological Diagnostic Unit Public Health Laboratory | Microbiological Diagnostic Unit Public Health Laboratory | Seemann T. et al |
| 1932 | Taiwan/TSGH-10/2020 | 2020-03-24 | TSGH-CP molecular lab | TSGH-CP molecular lab | Cherng-Lih Perng et al |
| 1933 | Denmark/ALAB-SSI494/2020 | 2020-03-24 | Department of Virus and Microbiological Special Diagnostics, Statens Serum Institut, Copenhagen, Denmark, Artillerivej 5, 2300 Copenahgen S | Albertsen lab, Department of Chemistry and Bioscience, Aalborg University, Denmark | Rasmus Kirkegaard et al |
| 1934 | USA/WA-UW-1880/2020 | 2020-03-24 | UW Virology Lab | UW Virology Lab | Pavitra Roychoudhury et al |
| 1935 | NewZealand/20VR1623/2020-03-24 | 2020-03-24 | Southern Community Labs Dunedin | Institute of Environmental Science and Research (ESR) Ltd | **Matt Storey et al** |
| 1936 | NewZealand/20VR1625/2020-03-24 | 2020-03-24 | Southern Community Labs Dunedin | Institute of Environmental Science and Research (ESR) Ltd | **Matt Storey et al** |
| 1937 | NewZealand/20VR1615/2020-03-24 | 2020-03-24 | Southern Community Labs Dunedin | Institute of Environmental Science and Research (ESR) Ltd | **Matt Storey et al** |
| 1938 | NewZealand/20VR1622/2020-03-24 | 2020-03-24 | Southern Community Labs Dunedin | Institute of Environmental Science and Research (ESR) Ltd | **Matt Storey et al** |
| 1939 | NewZealand/20VR1597/2020-03-24 | 2020-03-24 | PathLab Bay of Plenty | Institute of Environmental Science and Research (ESR) Ltd | **Matt Storey et al** |
| 1940 | NewZealand/20VR1950/2020-03-24 | 2020-03-24 | Wellington SCL (WN) | Institute of Environmental Science and Research (ESR) Ltd | **Matt Storey et al** |
| 1941 | NewZealand/20VR1581/2020-03-24 | 2020-03-24 | PathLab Bay of Plenty | Institute of Environmental Science and Research (ESR) Ltd | **Matt Storey et al** |
| 1942 | NewZealand/20VR1108/2020-03-24 | 2020-03-24 | Waikato Hospital | Institute of Environmental Science and Research (ESR) Ltd | **Matt Storey et al** |
| 1943 | NewZealand/20VR1561/2020-03-24 | 2020-03-24 | PathLab Bay of Plenty | Institute of Environmental Science and Research (ESR) Ltd | **Matt Storey et al** |
| 1944 | NewZealand/20VR1621/2020-03-24 | 2020-03-24 | Southern Community Labs Dunedin | Institute of Environmental Science and Research (ESR) Ltd | **Matt Storey et al** |
| 1945 | NewZealand/20VR1620/2020-03-24 | 2020-03-24 | Southern Community Labs Dunedin | Institute of Environmental Science and Research (ESR) Ltd | **Matt Storey et al** |
| 1946 | France/ARA13160/2020 | 2020-03-24 | Institut des Agents Infectieux (IAI), Hospices Civils de Lyon | CNR Virus des Infections Respiratoires - France SUD | Antonin Bal et al |
| 1947 | USA/TX-HMH0209/2020 | 2020-03-24 | Houston Methodist Hospital | Houston Methodist Hospital | S. Wesley Long et al |
| 1948 | Italy/TE6572/2020 | 2020-03-24 | Ospedale Civile Giuseppe Mazzini | "Istituto Zooprofilattico Sperimentale dell'Abruzzo e Molise ""G. Caporale""" | Lorusso A et al |
| 1949 | USA/TX-HMH0201/2020 | 2020-03-24 | Houston Methodist Hospital | Houston Methodist Hospital | S. Wesley Long et al |

| | | | | | |
|---|---|---|---|---|---|
| 1950 | Belgium/NMA-0324169/2020 | 2020-03-24 | KU Leuven, Clinical and Epidemiological Virology | KU Leuven, Clinical and Epidemiological Virology | Joan Marti-Carreras et al |
| 1951 | USA/WI-200/2020 | 2020-03-24 | University of Wisconsin-Madison AIDS Vaccine Research Laboratories | University of Wisconsin-Madison AIDS Vaccine Research Laboratories | Gage Moreno and Katarina Braun et al |
| 1952 | Spain/VH198152683/2020 | 2020-03-24 | Hospital Universitari Vall d'Hebron (HUVH) - Vall d'Hebron Research Institute (VHIR) | Hospital Universitari Vall d'Hebron (HUVH) - Vall d'Hebron Research Institute (VHIR) | Cristina Andrés et al |
| 1953 | France/HF3534/2020 | 2020-03-24 | CH Compiègne Laboratoire de Biologie | National Reference Center for Viruses of Respiratory Infections, Institut Pasteur, Paris | Mélanie Albert et al |
| 1954 | Scotland/EDB168/2020 | 2020-03-24 | Virology Department, Royal Infirmary of Edinburgh, NHS Lothian / School of Biological Sciences, University of Edinburgh / Institute of Genetics and Molecular Medicine, University of Edinburgh | COVID-19 Genomics UK (COG-UK) Consortium | McHugh M et al |
| 1955 | France/HF3419/2020 | 2020-03-24 | CH Jean de Navarre Laboratoire de Biologie | National Reference Center for Viruses of Respiratory Infections, Institut Pasteur, Paris | Mélanie Albert et al |
| 1956 | Netherlands/NA_192/2020 | 2020-03-24 | Dutch COVID-19 response team | Erasmus Medical Center | Bas Oude Munnink et al |
| 1957 | USA/WA-S541/2020 | 2020-03-24 | Washington State Department of Health | Seattle Flu Study | Chu et al et al |
| 1958 | Denmark/ALAB-SSI488/2020 | 2020-03-24 | Department of Virus and Microbiological Special Diagnostics, Statens Serum Institut, Copenhagen, Denmark, Artillerivej 5, 2300 Copenahgen S | Albertsen lab, Department of Chemistry and Bioscience, Aalborg University, Denmark | Rasmus Kirkegaard et al |
| 1959 | USA/TX-HMH0210/2020 | 2020-03-24 | Houston Methodist Hospital | Houston Methodist Hospital | S. Wesley Long et al |
| 1960 | Netherlands/ZuidHolland_70/2020 | 2020-03-24 | Dutch COVID-19 response team | Erasmus Medical Center | Bas Oude Munnink et al |
| 1961 | Netherlands/NA_201/2020 | 2020-03-24 | Dutch COVID-19 response team | Erasmus Medical Center | Bas Oude Munnink et al |
| 1962 | Belgium/DNS-0324176/2020 | 2020-03-24 | KU Leuven, Clinical and Epidemiological Virology | KU Leuven, Clinical and Epidemiological Virology | Joan Marti-Carreras et al |
| 1963 | England/20132081104/2020 | 2020-03-24 | Respiratory Virus Unit, Microbiology Services Colindale, Public Health England | Respiratory Virus Unit, Microbiology Services Colindale, Public Health England | Monica Galiano et al |
| 1964 | USA/WI-40/2020 | 2020-03-24 | University of Wisconsin-Madison AIDS Vaccine Research Laboratories | University of Wisconsin-Madison AIDS Vaccine Research Laboratories | Moreno et al |
| 1965 | England/SHEF-C0172/2020 | 2020-03-24 | Virology Department, Sheffield Teaching Hospitals NHS Foundation Trust | Department of Infection, Immunity and Cardiovascular Disease, The Florey Institute, The Medical School, University of Sheffield | Thushan de Silva et al |
| 1966 | Taiwan/TSGH-14/2020 | 2020-03-24 | TSGH-CP molecular lab | TSGH-CP molecular lab | Cherng-Lih Perng et al |
| 1967 | Taiwan/TSGH-18/2020 | 2020-03-24 | TSGH-CP molecular lab | TSGH-CP molecular lab | Cherng-Lih Perng et al |
| 1968 | Taiwan/TSGH-38/2020 | 2020-03-24 | TSGH-CP molecular lab | TSGH-CP molecular lab | Cherng-Lih Perng et al |
| 1969 | England/20134001104/2020 | 2020-03-24 | Respiratory Virus Unit, Microbiology Services Colindale, Public Health England | Respiratory Virus Unit, Microbiology Services Colindale, Public Health England | Monica Galiano et al |
| 1970 | Belgium/SEM-0324120/2020 | 2020-03-24 | KU Leuven, Clinical and Epidemiological Virology | KU Leuven, Clinical and Epidemiological Virology | Joan Marti-Carreras et al |
| 1971 | England/201340875/2020 | 2020-03-24 | Respiratory Virus Unit, Microbiology Services Colindale, Public Health England | Respiratory Virus Unit, Microbiology Services Colindale, Public Health England | Monica Galiano et al |
| 1972 | France/ARA1307/2020 | 2020-03-24 | Institut des Agents Infectieux (IAI), Hospices Civils de Lyon | CNR Virus des Infections Respiratoires - France SUD | Antonin Bal et al |
| 1973 | England/20134018902/2020 | 2020-03-24 | Respiratory Virus Unit, Microbiology Services | Respiratory Virus Unit, Microbiology Services | Monica Galiano et al |

| | | | | | |
|---|---|---|---|---|---|
| | | | Colindale, Public Health England | Colindale, Public Health England | |
| 1974 | USA/WA-S502/2020 | 2020-03-24 | Washington State Department of Health | Seattle Flu Study | Chu et al et al |
| 1975 | Denmark/ALAB-SSI450/2020 | 2020-03-24 | Department of Virus and Microbiological Special Diagnostics, Statens Serum Institut, Copenhagen, Denmark, Artillerivej 5, 2300 Copenahgen S | Albertsen lab, Department of Chemistry and Bioscience, Aalborg University, Denmark | Rasmus Kirkegaard et al |
| 1976 | Switzerland/GE5708/2020 | 2020-03-24 | University Hospitals of Geneva Laboratory of Virology | University Hospitals of Geneva Laboratory of Virology | Laubscher F. et al |
| 1977 | Denmark/ALAB-SSI448/2020 | 2020-03-25 | Department of Virus and Microbiological Special Diagnostics, Statens Serum Institut, Copenhagen, Denmark, Artillerivej 5, 2300 Copenahgen S | Albertsen lab, Department of Chemistry and Bioscience, Aalborg University, Denmark | Rasmus Kirkegaard et al |
| 1978 | Australia/NSW204/2020 | 2020-03-25 | Centre for Infectious Diseases and Microbiology Public Health | NSW Health Pathology - Institute of Clinical Pathology and Medical Research; Westmead Hospital; University of Sydney | Lam C et al |
| 1979 | Australia/NSW191/2020 | 2020-03-25 | Centre for Infectious Diseases and Microbiology Public Health | NSW Health Pathology - Institute of Clinical Pathology and Medical Research; Westmead Hospital; University of Sydney | Sim E et al |
| 1980 | Australia/VIC566/2020 | 2020-03-25 | Victorian Infectious Diseases Reference Laboratory (VIDRL) | Microbiological Diagnostic Unit Public Health Laboratory and Victorian Infectious Diseases Reference Laboratory, Doherty Institute | Caly L. et al |
| 1981 | Australia/VIC565/2020 | 2020-03-25 | Victorian Infectious Diseases Reference Laboratory (VIDRL) | Microbiological Diagnostic Unit Public Health Laboratory and Victorian Infectious Diseases Reference Laboratory, Doherty Institute | Caly L. et al |
| 1982 | Australia/VIC407/2020 | 2020-03-25 | Victorian Infectious Diseases Reference Laboratory (VIDRL) | Microbiological Diagnostic Unit Public Health Laboratory and Victorian Infectious Diseases Reference Laboratory, Doherty Institute | Caly L. et al |
| 1983 | Australia/VIC535/2020 | 2020-03-25 | Victorian Infectious Diseases Reference Laboratory (VIDRL) | Microbiological Diagnostic Unit Public Health Laboratory and Victorian Infectious Diseases Reference Laboratory, Doherty Institute | Caly L. et al |
| 1984 | Australia/NSW64/2020 | 2020-03-25 | Centre for Infectious Diseases and Microbiology Public Health | NSW Health Pathology - Institute of Clinical Pathology and Medical Research; Westmead Hospital; University of Sydney | Lam C et al |
| 1985 | Australia/NSW149/2020 | 2020-03-25 | Centre for Infectious Diseases and Microbiology Public Health | NSW Health Pathology - Institute of Clinical Pathology and Medical Research; Westmead Hospital; University of Sydney | Sadsad R et al |
| 1986 | USA/WI-31/2020 | 2020-03-25 | University of Wisconsin-Madison AIDS Vaccine Research Laboratories | University of Wisconsin-Madison AIDS Vaccine Research Laboratories | Moreno et al |
| 1987 | Spain/Valencia273/2020 | 2020-03-25 | Servicio de Microbiologia. Hospital Clinico Universitario de Valencia | Sequencing and Bioinformatics Service and Molecular Epidemiology Research Group. FISABIO-Public Health | Inma Galán Vendrell et al |
| 1988 | Sweden/20-06468/2020 | 2020-03-25 | Uppsala Narakut Aleris | The Public Health Agency of Sweden | Annika Nilsson et al |
| 1989 | Australia/VIC584/2020 | 2020-03-25 | Victorian Infectious Diseases Reference Laboratory (VIDRL) | Microbiological Diagnostic Unit Public Health Laboratory and Victorian Infectious Diseases Reference Laboratory, Doherty Institute | Caly L. et al |

| | | | | | |
|---|---|---|---|---|---|
| 1990 | Australia/NSW122/2020 | 2020-03-25 | Centre for Infectious Diseases and Microbiology Public Health | NSW Health Pathology - Institute of Clinical Pathology and Medical Research; Westmead Hospital; University of Sydney | Bachmann N et al |
| 1991 | Australia/VIC571/2020 | 2020-03-25 | Victorian Infectious Diseases Reference Laboratory (VIDRL) | Microbiological Diagnostic Unit Public Health Laboratory and Victorian Infectious Diseases Reference Laboratory, Doherty Institute | Caly L. et al |
| 1992 | Australia/QLDID935/2020 | 2020-03-25 | Prince Charles Hospital | Public Health Virology Laboratory, Forensics and Scientific Services, Queensland Health | Alyssa Pyke et al |
| 1993 | Australia/VIC558/2020 | 2020-03-25 | Victorian Infectious Diseases Reference Laboratory (VIDRL) | Microbiological Diagnostic Unit Public Health Laboratory and Victorian Infectious Diseases Reference Laboratory, Doherty Institute | Caly L. et al |
| 1994 | Australia/NSW68/2020 | 2020-03-25 | Centre for Infectious Diseases and Microbiology Public Health | NSW Health Pathology - Institute of Clinical Pathology and Medical Research; Westmead Hospital; University of Sydney | Arnott A et al |
| 1995 | Australia/VIC534/2020 | 2020-03-25 | Victorian Infectious Diseases Reference Laboratory (VIDRL) | Microbiological Diagnostic Unit Public Health Laboratory and Victorian Infectious Diseases Reference Laboratory, Doherty Institute | Caly L. et al |
| 1996 | Australia/NSW117/2020 | 2020-03-25 | Centre for Infectious Diseases and Microbiology Public Health | NSW Health Pathology - Institute of Clinical Pathology and Medical Research; Westmead Hospital; University of Sydney | Gall M et al |
| 1997 | Australia/VIC555/2020 | 2020-03-25 | Victorian Infectious Diseases Reference Laboratory (VIDRL) | Microbiological Diagnostic Unit Public Health Laboratory and Victorian Infectious Diseases Reference Laboratory, Doherty Institute | Caly L. et al |
| 1998 | Australia/NSW124/2020 | 2020-03-25 | Centre for Infectious Diseases and Microbiology Public Health | NSW Health Pathology - Institute of Clinical Pathology and Medical Research; Westmead Hospital; University of Sydney | Lam C et al |
| 1999 | Australia/VIC561/2020 | 2020-03-25 | Victorian Infectious Diseases Reference Laboratory (VIDRL) | Microbiological Diagnostic Unit Public Health Laboratory and Victorian Infectious Diseases Reference Laboratory, Doherty Institute | Caly L. et al |
| 2000 | Australia/VIC548/2020 | 2020-03-25 | Victorian Infectious Diseases Reference Laboratory (VIDRL) | Microbiological Diagnostic Unit Public Health Laboratory and Victorian Infectious Diseases Reference Laboratory, Doherty Institute | Caly L. et al |
| 2001 | Australia/QLDID931/2020 | 2020-03-25 | Mater Pathology | Public Health Virology Laboratory | Bixing Huang et al |
| 2002 | Australia/NSW150/2020 | 2020-03-25 | Centre for Infectious Diseases and Microbiology Public Health | NSW Health Pathology - Institute of Clinical Pathology and Medical Research; Westmead Hospital; University of Sydney | Draper J et al |
| 2003 | Australia/NSW131/2020 | 2020-03-25 | Centre for Infectious Diseases and Microbiology Public Health | NSW Health Pathology - Institute of Clinical Pathology and Medical Research; Westmead Hospital; University of Sydney | Sim E et al |
| 2004 | USA/WA-UW-2244/2020 | 2020-03-25 | UW Virology Lab | UW Virology Lab | Pavitra Roychoudhury et al |
| 2005 | England/SHEF-C06DD/2020 | 2020-03-25 | Virology Department, Sheffield Teaching Hospitals NHS Foundation Trust | Department of Infection, Immunity and Cardiovascular Disease, The Florey Institute, The Medical School, University of Sheffield | Thushan de Silva et al |

| | | | | | |
|---|---|---|---|---|---|
| 2006 | Russia/Yakutia-83301/2020 | 2020-03-25 | State Research Center of Virology and Biotechnology VECTOR, Department of Collection of Microorganisms | State Research Center of Virology and Biotechnology VECTOR, Department of Collection of Microorganisms | Oleg V. Pyankov et al |
| 2007 | Wales/PHWC-25FEA/2020 | 2020-03-25 | Wales Specialist Virology Centre | Public Health Wales Microbiology Cardiff | Catherine Moore et al |
| 2008 | Denmark/ALAB-SSI451/2020 | 2020-03-25 | Department of Virus and Microbiological Special Diagnostics, Statens Serum Institut, Copenhagen, Denmark, Artillerivej 5, 2300 Copenahgen S | Albertsen lab, Department of Chemistry and Bioscience, Aalborg University, Denmark | Rasmus Kirkegaard et al |
| 2009 | Wales/PHWC-260F5/2020 | 2020-03-25 | Wales Specialist Virology Centre | Public Health Wales Microbiology Cardiff | Catherine Moore et al |
| 2010 | Australia/VIC551/2020 | 2020-03-25 | Victorian Infectious Diseases Reference Laboratory (VIDRL) | Microbiological Diagnostic Unit Public Health Laboratory and Victorian Infectious Diseases Reference Laboratory, Doherty Institute | Caly L. et al |
| 2011 | Australia/VIC597/2020 | 2020-03-25 | Victorian Infectious Diseases Reference Laboratory (VIDRL) | Microbiological Diagnostic Unit Public Health Laboratory and Victorian Infectious Diseases Reference Laboratory, Doherty Institute | Caly L. et al |
| 2012 | Australia/NT19/2020 | 2020-03-25 | Royal Darwin Hospital Pathology | Microbiological Diagnostic Unit Public Health Laboratory and Victorian Infectious Diseases Reference Laboratory, Doherty Institute | Meumann et al |
| 2013 | Turkey/HSGM-11518/2020 | 2020-03-25 | Ministry of Health Turkey | Ministry of Health Turkey | Fatma Bayrakdar et al |
| 2014 | Italy/VR_20COV21-26/2020 | 2020-03-25 | ULSS9 Distretto di Bussolengo | Istituto Zooprofilattico Sperimentale delle Venezie | Adelaide Milani et al |
| 2015 | USA/TX-HMH0366/2020 | 2020-03-25 | Houston Methodist Hospital | Houston Methodist Hospital | S. Wesley Long et al |
| 2016 | USA/CA-CZB045/2020 | 2020-03-25 | UCSF Clinical Microbiology Laboratory | Chan-Zuckerberg Biohub | CZB Cliahub Consortium et al |
| 2017 | USA/WA-UW-2140/2020 | 2020-03-25 | UW Virology Lab | UW Virology Lab | Pavitra Roychoudhury et al |
| 2018 | USA/WA-UW-2145/2020 | 2020-03-25 | UW Virology Lab | UW Virology Lab | Pavitra Roychoudhury et al |
| 2019 | Spain/Valencia247/2020 | 2020-03-25 | Servicio de Microbiologia. Hospital Clinico Universitario de Valencia | Sequencing and Bioinformatics Service and Molecular Epidemiology Research Group. FISABIO-Public Health | Marta Pla Diaz et al |
| 2020 | Italy/VR_20COV21-37/2020 | 2020-03-25 | ULSS9 Distretto di Bussolengo | Istituto Zooprofilattico Sperimentale delle Venezie | Adelaide Milani et al |
| 2021 | USA/WI-118/2020 | 2020-03-25 | University of Wisconsin-Madison AIDS Vaccine Research Laboratories | University of Wisconsin-Madison AIDS Vaccine Research Laboratories | Moreno et al |
| 2022 | USA/WA-S397/2020 | 2020-03-25 | Washington State Department of Health | Seattle Flu Study | Chu et al et al |
| 2023 | France/IDF3577/2020 | 2020-03-25 | LABM GH nord Essonne de Longjumeau - BP 125 | National Reference Center for Viruses of Respiratory Infections, Institut Pasteur, Paris | Mélanie Albert et al |
| 2024 | Australia/VIC547/2020 | 2020-03-25 | Victorian Infectious Diseases Reference Laboratory (VIDRL) | Microbiological Diagnostic Unit Public Health Laboratory and Victorian Infectious Diseases Reference Laboratory, Doherty Institute | Caly L. et al |
| 2025 | Australia/VIC568/2020 | 2020-03-25 | Victorian Infectious Diseases Reference Laboratory (VIDRL) | Microbiological Diagnostic Unit Public Health Laboratory and Victorian Infectious Diseases Reference Laboratory, Doherty Institute | Caly L. et al |
| 2026 | Australia/VIC599/2020 | 2020-03-25 | Victorian Infectious Diseases Reference Laboratory (VIDRL) | Microbiological Diagnostic Unit Public Health Laboratory and Victorian Infectious Diseases Reference Laboratory, Doherty Institute | Caly L. et al |
| 2027 | Australia/VIC572/2020 | 2020-03-25 | Victorian Infectious Diseases Reference Laboratory (VIDRL) | Microbiological Diagnostic Unit Public Health Laboratory and Victorian Infectious Diseases | Caly L. et al |

| | | | | | |
|---|---|---|---|---|---|
| | | | | Reference Laboratory, Doherty Institute | |
| 2028 | England/NOTT-10E57B/2020 | 2020-03-25 | Queens Medical Centre, Clinical Microbiology Department / DeepSeq Nottingham | COVID-19 Genomics UK (COG-UK) Consortium | Gemma Clark et al |
| 2029 | England/201380049/2020 | 2020-03-25 | Respiratory Virus Unit, Microbiology Services Colindale, Public Health England | Respiratory Virus Unit, Microbiology Services Colindale, Public Health England | Monica Galiano et al |
| 2030 | England/201361022/2020 | 2020-03-25 | Respiratory Virus Unit, Microbiology Services Colindale, Public Health England | Respiratory Virus Unit, Microbiology Services Colindale, Public Health England | Monica Galiano et al |
| 2031 | NewZealand/20VR1778/2020-03-25 | 2020-03-25 | Southern Community Labs Dunedin | Institute of Environmental Science and Research (ESR) Ltd | **Matt Storey et al** |
| 2032 | NewZealand/20VR1765/2020-03-25 | 2020-03-25 | Southern Community Labs Dunedin | Institute of Environmental Science and Research (ESR) Ltd | **Matt Storey et al** |
| 2033 | NewZealand/20VR1763/2020-03-25 | 2020-03-25 | Southern Community Labs Dunedin | Institute of Environmental Science and Research (ESR) Ltd | **Matt Storey et al** |
| 2034 | NewZealand/20VR1984/2020-03-25 | 2020-03-25 | Canterbury Health Laboratories | Institute of Environmental Science and Research (ESR) Ltd | **Matt Storey et al** |
| 2035 | Belgium/LJ-0325148/2020 | 2020-03-25 | KU Leuven, Clinical and Epidemiological Virology | KU Leuven, Clinical and Epidemiological Virology | Joan Marti-Carreras et al |
| 2036 | France/IDF3703/2020 | 2020-03-25 | LABM GH nord Essonne de Longjumeau - BP 125 | National Reference Center for Viruses of Respiratory Infections, Institut Pasteur, Paris | Mélanie Albert et al |
| 2037 | USA/SEARCH-0068-NBG/2020 | 2020-03-25 | Robert Garry lab | Andersen lab at Scripps Research | Allison Smither et al |
| 2038 | Denmark/ALAB-SSI512/2020 | 2020-03-25 | Department of Virus and Microbiological Special Diagnostics, Statens Serum Institut, Copenhagen, Denmark, Artillerivej 5, 2300 Copenahgen S | Albertsen lab, Department of Chemistry and Bioscience, Aalborg University, Denmark | Rasmus Kirkegaard et al |
| 2039 | Netherlands/NA_213/2020 | 2020-03-25 | Dutch COVID-19 response team | Erasmus Medical Center | Bas Oude Munnink et al |
| 2040 | Spain/Valencia86/2020 | 2020-03-25 | Servicio de Microbiologia. Hospital Clinico Universitario de Valencia | Sequencing and Bioinformatics Service and Molecular Epidemiology Research Group. FISABIO-Public Health | Loreto Ferrús Abad et al |
| 2041 | USA/VA-DCLS-0042/2020 | 2020-03-25 | Virginia DCLS | Virginia DCLS | Virginia DCLS et al |
| 2042 | USA/WA-S429/2020 | 2020-03-25 | Washington State Department of Health | Seattle Flu Study | Chu et al et al |
| 2043 | USA/WA-UW-2218/2020 | 2020-03-25 | UW Virology Lab | UW Virology Lab | Pavitra Roychoudhury et al |
| 2044 | USA/WA-UW-2141/2020 | 2020-03-25 | UW Virology Lab | UW Virology Lab | Pavitra Roychoudhury et al |
| 2045 | USA/WA-S405/2020 | 2020-03-25 | Washington State Department of Health | Seattle Flu Study | Chu et al et al |
| 2046 | USA/WA-UW-2245/2020 | 2020-03-25 | UW Virology Lab | UW Virology Lab | Pavitra Roychoudhury et al |
| 2047 | Denmark/ALAB-SSI444/2020 | 2020-03-25 | Department of Virus and Microbiological Special Diagnostics, Statens Serum Institut, Copenhagen, Denmark, Artillerivej 5, 2300 Copenahgen S | Albertsen lab, Department of Chemistry and Bioscience, Aalborg University, Denmark | Rasmus Kirkegaard et al |
| 2048 | Iceland/530/2020 | 2020-03-25 | deCODE genetics | deCODE genetics | Daniel F Gudbjartsson et al |
| 2049 | Iceland/524/2020 | 2020-03-25 | deCODE genetics | deCODE genetics | Daniel F Gudbjartsson et al |
| 2050 | Belgium/VPMV-0325153/2020 | 2020-03-25 | KU Leuven, Clinical and Epidemiological Virology | KU Leuven, Clinical and Epidemiological Virology | Joan Marti-Carreras et al |
| 2051 | USA/WI-67/2020 | 2020-03-25 | University of Wisconsin-Madison AIDS Vaccine Research Laboratories | University of Wisconsin-Madison AIDS Vaccine Research Laboratories | Moreno et al |
| 2052 | USA/SEARCH-0060-NBG/2020 | 2020-03-25 | Robert Garry lab | Andersen lab at Scripps Research | Allison Smither et al |

| | | | | | |
|---|---|---|---|---|---|
| 2053 | USA/WA-S351/2020 | 2020-03-25 | Washington State Department of Health | Seattle Flu Study | Chu et al et al |
| 2054 | Spain/Valencia250/2020 | 2020-03-25 | Servicio de Microbiologia. Hospital Clinico Universitario de Valencia | Sequencing and Bioinformatics Service and Molecular Epidemiology Research Group. FISABIO-Public Health | Sandra Carbo et al |
| 2055 | Taiwan/TSGH-11/2020 | 2020-03-25 | TSGH-CP molecular lab | TSGH-CP molecular lab | Cherng-Lih Perng et al |
| 2056 | Taiwan/NTU24/2020 | 2020-03-25 | Department of Laboratory Medicine, National Taiwan University Hospital | Microbial Genomics Core Lab, National Taiwan University Centers of Genomic and Precision Medicine | Shiou-Hwei Yeh et al |
| 2057 | Taiwan/TSGH-31/2020 | 2020-03-25 | TSGH-CP molecular lab | TSGH-CP molecular lab | Cherng-Lih Perng et al |
| 2058 | Taiwan/NTU23/2020 | 2020-03-25 | Department of Laboratory Medicine, National Taiwan University Hospital | Microbial Genomics Core Lab, National Taiwan University Centers of Genomic and Precision Medicine | Shiou-Hwei Yeh et al |
| 2059 | England/201380221/2020 | 2020-03-25 | Respiratory Virus Unit, Microbiology Services Colindale, Public Health England | Respiratory Virus Unit, Microbiology Services Colindale, Public Health England | Monica Galiano et al |
| 2060 | England/201360221/2020 | 2020-03-25 | Respiratory Virus Unit, Microbiology Services Colindale, Public Health England | Respiratory Virus Unit, Microbiology Services Colindale, Public Health England | Monica Galiano et al |
| 2061 | USA/WA-S500/2020 | 2020-03-25 | Washington State Department of Health | Seattle Flu Study | Chu et al et al |
| 2062 | Luxembourg/LNS3437586/2020 | 2020-03-25 | Laboratoire National de Sante, Microbiology, Virology | Laboratoire National de Sante, Microbiology, Epidemiology and Microbial Genomics | Anke Wienecke-Baldacchino et al |
| 2063 | England/SHEF-C0822/2020 | 2020-03-25 | Virology Department, Sheffield Teaching Hospitals NHS Foundation Trust | Department of Infection, Immunity and Cardiovascular Disease, The Florey Institute, The Medical School, University of Sheffield | Thushan de Silva et al |
| 2064 | England/201380055/2020 | 2020-03-25 | Respiratory Virus Unit, Microbiology Services Colindale, Public Health England | Respiratory Virus Unit, Microbiology Services Colindale, Public Health England | Monica Galiano et al |
| 2065 | USA/WA-S300/2020 | 2020-03-25 | Seattle Flu Study | Seattle Flu Study | Chu et al et al |
| 2066 | England/201380042/2020 | 2020-03-25 | Respiratory Virus Unit, Microbiology Services Colindale, Public Health England | Respiratory Virus Unit, Microbiology Services Colindale, Public Health England | Monica Galiano et al |
| 2067 | USA/CA-CZB09/2020 | 2020-03-25 | UCSF Clinical Microbiology Laboratory | Chan-Zuckerberg Biohub | CZB Cliahub Consortium et al |
| 2068 | USA/WA-UW-2132/2020 | 2020-03-25 | UW Virology Lab | UW Virology Lab | Pavitra Roychoudhury et al |
| 2069 | Wales/PHWC-25E47/2020 | 2020-03-25 | Wales Specialist Virology Centre | Public Health Wales Microbiology Cardiff | Catherine Moore et al |
| 2070 | Australia/VIC617/2020 | 2020-03-26 | Victorian Infectious Diseases Reference Laboratory (VIDRL) | Microbiological Diagnostic Unit Public Health Laboratory and Victorian Infectious Diseases Reference Laboratory, Doherty Institute | Caly L. et al |
| 2071 | Australia/VIC630/2020 | 2020-03-26 | Victorian Infectious Diseases Reference Laboratory (VIDRL) | Microbiological Diagnostic Unit Public Health Laboratory and Victorian Infectious Diseases Reference Laboratory, Doherty Institute | Caly L. et al |
| 2072 | Australia/NSW173/2020 | 2020-03-26 | Centre for Infectious Diseases and Microbiology Public Health | NSW Health Pathology - Institute of Clinical Pathology and Medical Research; Westmead Hospital; University of Sydney | Rockett R et al |
| 2073 | Australia/NSW192/2020 | 2020-03-26 | Centre for Infectious Diseases and Microbiology Public Health | NSW Health Pathology - Institute of Clinical Pathology and Medical Research; Westmead Hospital; University of Sydney | Bachmann N et al |
| 2074 | Australia/NSW196/2020 | 2020-03-26 | Centre for Infectious Diseases and Microbiology Public | NSW Health Pathology - Institute of Clinical Pathology | Timms V et al |

| | | | | | |
|---|---|---|---|---|---|
| | | | Health | and Medical Research; Westmead Hospital; University of Sydney | |
| 2075 | Australia/VIC610/2020 | 2020-03-26 | Victorian Infectious Diseases Reference Laboratory (VIDRL) | Microbiological Diagnostic Unit Public Health Laboratory and Victorian Infectious Diseases Reference Laboratory, Doherty Institute | Caly L. et al |
| 2076 | Australia/VIC458/2020 | 2020-03-26 | Victorian Infectious Diseases Reference Laboratory (VIDRL) | Microbiological Diagnostic Unit Public Health Laboratory and Victorian Infectious Diseases Reference Laboratory, Doherty Institute | Caly L. et al |
| 2077 | Australia/VIC471/2020 | 2020-03-26 | Victorian Infectious Diseases Reference Laboratory (VIDRL) | Microbiological Diagnostic Unit Public Health Laboratory and Victorian Infectious Diseases Reference Laboratory, Doherty Institute | Caly L. et al |
| 2078 | Spain/Valencia260/2020 | 2020-03-26 | Servicio de Microbiologia. Hospital Clinico Universitario de Valencia | Sequencing and Bioinformatics Service and Molecular Epidemiology Research Group. FISABIO-Public Health | Lidia Ruiz Roldan et al |
| 2079 | USA/VA-DCLS-0080/2020 | 2020-03-26 | Virginia DCLS | Virginia DCLS | Virginia DCLS et al |
| 2080 | Australia/VIC603/2020 | 2020-03-26 | Victorian Infectious Diseases Reference Laboratory (VIDRL) | Microbiological Diagnostic Unit Public Health Laboratory and Victorian Infectious Diseases Reference Laboratory, Doherty Institute | Caly L. et al |
| 2081 | Australia/VIC409/2020 | 2020-03-26 | Victorian Infectious Diseases Reference Laboratory (VIDRL) | Microbiological Diagnostic Unit Public Health Laboratory and Victorian Infectious Diseases Reference Laboratory, Doherty Institute | Caly L. et al |
| 2082 | Australia/NSW132/2020 | 2020-03-26 | Centre for Infectious Diseases and Microbiology Public Health | NSW Health Pathology - Institute of Clinical Pathology and Medical Research; Westmead Hospital; University of Sydney | Bachmann N et al |
| 2083 | Australia/NSW185/2020 | 2020-03-26 | Centre for Infectious Diseases and Microbiology Public Health | NSW Health Pathology - Institute of Clinical Pathology and Medical Research; Westmead Hospital; University of Sydney | Gray K et al |
| 2084 | Australia/VIC942/2020 | 2020-03-26 | Microbiological Diagnostic Unit Public Health Laboratory | Microbiological Diagnostic Unit Public Health Laboratory | Seemann T. et al |
| 2085 | Australia/NSWID930/2020 | 2020-03-26 | Pathology North | Public Health Virology Laboratory | Bixing Huang et al |
| 2086 | Australia/VIC951/2020 | 2020-03-26 | Microbiological Diagnostic Unit Public Health Laboratory | Microbiological Diagnostic Unit Public Health Laboratory | Seemann T. et al |
| 2087 | Netherlands/NA_220/2020 | 2020-03-26 | Dutch COVID-19 response team | Erasmus Medical Center | Bas Oude Munnink et al |
| 2088 | USA/AZ-TG271890/2020 | 2020-03-26 | AZ SPHL, Arizona Department of Health Services | TGen North | Jolene Bowers et al |
| 2089 | Spain/Valencia210/2020 | 2020-03-26 | Servicio de Microbiologia. Hospital Clinico Universitario de Valencia | Sequencing and Bioinformatics Service and Molecular Epidemiology Research Group. FISABIO-Public Health | Sandra Carbo et al |
| 2090 | CzechRepublic/IAB_15/2020 | 2020-03-26 | Motol University Hospital | Institute of Applied Biotechnologies a.s. | Petr Brož et al |
| 2091 | England/20138088204/2020 | 2020-03-26 | Respiratory Virus Unit, Microbiology Services Colindale, Public Health England | Respiratory Virus Unit, Microbiology Services Colindale, Public Health England | Monica Galiano et al |
| 2092 | USA/WA-S369/2020 | 2020-03-26 | Washington State Department of Health | Seattle Flu Study | Chu et al et al |
| 2093 | USA/WA-UW-4612/2020 | 2020-03-26 | UW Virology Lab | UW Virology Lab | Pavitra Roychoudhury et al |
| 2094 | USA/WI-230/2020 | 2020-03-26 | University of Wisconsin-Madison AIDS Vaccine Research Laboratories | University of Wisconsin-Madison AIDS Vaccine Research Laboratories | Gage Moreno and Katarina Braun et al |
| 2095 | Russia/Chechenya-83803/2020 | 2020-03-26 | State Research Center of | State Research Center of | Oleg V. Pyankov et al |

| | | | | | |
|---|---|---|---|---|---|
| | | | Virology and Biotechnology VECTOR, Department of Collection of Microorganisms | Virology and Biotechnology VECTOR, Department of Collection of Microorganisms | |
| 2096 | Spain/Valencia233/2020 | 2020-03-26 | Servicio de Microbiologia. Hospital Clinico Universitario de Valencia | Sequencing and Bioinformatics Service and Molecular Epidemiology Research Group. FISABIO-Public Health | Mariana Reyes-Prieto et al |
| 2097 | Spain/Valencia261/2020 | 2020-03-26 | Servicio de Microbiologia. Hospital Clinico Universitario de Valencia | Sequencing and Bioinformatics Service and Molecular Epidemiology Research Group. FISABIO-Public Health | Marta Pla Diaz et al |
| 2098 | Spain/Valencia232/2020 | 2020-03-26 | Servicio de Microbiologia. Hospital Clinico Universitario de Valencia | Sequencing and Bioinformatics Service and Molecular Epidemiology Research Group. FISABIO-Public Health | Paula Ruiz-Hueso et al |
| 2099 | USA/WA-S442/2020 | 2020-03-26 | Washington State Department of Health | Seattle Flu Study | Chu et al et al |
| 2100 | USA/WA-UW-3819/2020 | 2020-03-26 | UW Virology Lab | UW Virology Lab | Pavitra Roychoudhury et al |
| 2101 | NewZealand/20VR1781/2020-03-26 | 2020-03-26 | Southern Community Labs Dunedin | Institute of Environmental Science and Research (ESR) Ltd | **Matt Storey et al** |
| 2102 | NewZealand/20VR1776/2020-03-26 | 2020-03-26 | Southern Community Labs Dunedin | Institute of Environmental Science and Research (ESR) Ltd | **Matt Storey et al** |
| 2103 | NewZealand/20VR1773/2020-03-26 | 2020-03-26 | Southern Community Labs Dunedin | Institute of Environmental Science and Research (ESR) Ltd | **Matt Storey et al** |
| 2104 | NewZealand/20VR1771/2020-03-26 | 2020-03-26 | Southern Community Labs Dunedin | Institute of Environmental Science and Research (ESR) Ltd | **Matt Storey et al** |
| 2105 | NewZealand/20VR1777/2020-03-26 | 2020-03-26 | Southern Community Labs Dunedin | Institute of Environmental Science and Research (ESR) Ltd | **Matt Storey et al** |
| 2106 | NewZealand/20VR1759/2020-03-26 | 2020-03-26 | Southern Community Labs Dunedin | Institute of Environmental Science and Research (ESR) Ltd | **Matt Storey et al** |
| 2107 | NewZealand/20VR1782/2020-03-26 | 2020-03-26 | Southern Community Labs Dunedin | Institute of Environmental Science and Research (ESR) Ltd | **Matt Storey et al** |
| 2108 | NewZealand/20VR1783/2020-03-26 | 2020-03-26 | Southern Community Labs Dunedin | Institute of Environmental Science and Research (ESR) Ltd | **Matt Storey et al** |
| 2109 | NewZealand/20VR1780/2020-03-26 | 2020-03-26 | Southern Community Labs Dunedin | Institute of Environmental Science and Research (ESR) Ltd | **Matt Storey et al** |
| 2110 | NewZealand/20VR1772/2020-03-26 | 2020-03-26 | Southern Community Labs Dunedin | Institute of Environmental Science and Research (ESR) Ltd | **Matt Storey et al** |
| 2111 | NewZealand/20VR1779/2020-03-26 | 2020-03-26 | Southern Community Labs Dunedin | Institute of Environmental Science and Research (ESR) Ltd | **Matt Storey et al** |
| 2112 | NewZealand/20VR1784/2020-03-26 | 2020-03-26 | Southern Community Labs Dunedin | Institute of Environmental Science and Research (ESR) Ltd | **Matt Storey et al** |
| 2113 | NewZealand/20VR1774/2020-03-26 | 2020-03-26 | Southern Community Labs Dunedin | Institute of Environmental Science and Research (ESR) Ltd | **Matt Storey et al** |
| 2114 | NewZealand/20VR2522/2020-03-26 | 2020-03-26 | LabPLUS | Institute of Environmental Science and Research (ESR) Ltd | **Matt Storey et al** |
| 2115 | Scotland/EDB219/2020 | 2020-03-26 | Virology Department, Royal Infirmary of Edinburgh, NHS Lothian / School of Biological Sciences, University of Edinburgh / Institute of Genetics and Molecular Medicine, University of Edinburgh | COVID-19 Genomics UK (COG-UK) Consortium | McHugh M et al |
| 2116 | USA/TX-HMH0266/2020 | 2020-03-26 | Houston Methodist Hospital | Houston Methodist Hospital | S. Wesley Long et al |
| 2117 | USA/ID-UW-3892/2020 | 2020-03-26 | UW Virology Lab | UW Virology Lab | Pavitra Roychoudhury et al |
| 2118 | Iceland/552/2020 | 2020-03-26 | The National University Hospital of Iceland | deCODE genetics | Daniel F Gudbjartsson et al |
| 2119 | USA/WA-UW-3834/2020 | 2020-03-26 | UW Virology Lab | UW Virology Lab | Pavitra Roychoudhury et al |
| 2120 | England/201360281/2020 | 2020-03-26 | Respiratory Virus Unit, Microbiology Services Colindale, Public Health England | Respiratory Virus Unit, Microbiology Services Colindale, Public Health England | Monica Galiano et al |
| 2121 | Denmark/ALAB-SSI497/2020 | 2020-03-26 | Department of Virus and Microbiological Special Diagnostics, Statens Serum | Albertsen lab, Department of Chemistry and Bioscience, Aalborg University, Denmark | Rasmus Kirkegaard et al |

| | | | | | |
|---|---|---|---|---|---|
| | | | Institut, Copenhagen, Denmark, Artillerivej 5, 2300 Copenahgen S | | |
| 2122 | India/S6/2020 | 2020-03-26 | ICMR-National Institute of Cholera and Enteric Diseases | National Institute of Biomedical Genomics | Arindam Maitra et al |
| 2123 | USA/WA-UW-3862/2020 | 2020-03-26 | UW Virology Lab | UW Virology Lab | Pavitra Roychoudhury et al |
| 2124 | USA/TX-HMH0273/2020 | 2020-03-26 | Houston Methodist Hospital | Houston Methodist Hospital | S. Wesley Long et al |
| 2125 | Spain/Valencia231/2020 | 2020-03-26 | Servicio de Microbiologia. Hospital Clinico Universitario de Valencia | Sequencing and Bioinformatics Service and Molecular Epidemiology Research Group. FISABIO-Public Health | Loreto Ferrús Abad et al |
| 2126 | Scotland/EDB214/2020 | 2020-03-26 | Virology Department, Royal Infirmary of Edinburgh, NHS Lothian / School of Biological Sciences, University of Edinburgh / Institute of Genetics and Molecular Medicine, University of Edinburgh | COVID-19 Genomics UK (COG-UK) Consortium | McHugh M et al |
| 2127 | Iceland/542/2020 | 2020-03-26 | deCODE genetics | deCODE genetics | Daniel F Gudbjartsson et al |
| 2128 | Iceland/559/2020 | 2020-03-26 | The National University Hospital of Iceland | deCODE genetics | Daniel F Gudbjartsson et al |
| 2129 | Iceland/590/2020 | 2020-03-26 | The National University Hospital of Iceland | deCODE genetics | Daniel F Gudbjartsson et al |
| 2130 | USA/WA-S391/2020 | 2020-03-26 | Washington State Department of Health | Seattle Flu Study | Chu et al et al |
| 2131 | USA/WI-223/2020 | 2020-03-26 | University of Wisconsin-Madison AIDS Vaccine Research Laboratories | University of Wisconsin-Madison AIDS Vaccine Research Laboratories | Gage Moreno and Katarina Braun et al |
| 2132 | England/20139029404/2020 | 2020-03-26 | Respiratory Virus Unit, Microbiology Services Colindale, Public Health England | Respiratory Virus Unit, Microbiology Services Colindale, Public Health England | Monica Galiano et al |
| 2133 | England/20139027004/2020 | 2020-03-26 | Respiratory Virus Unit, Microbiology Services Colindale, Public Health England | Respiratory Virus Unit, Microbiology Services Colindale, Public Health England | Monica Galiano et al |
| 2134 | France/OCC-11/2020 | 2020-03-26 | CHU Purpan - Laboratoire de Virologie - Institut Fédératif de Biologie | Laboratoire de virologie - École Nationale Vétérinaire de Toulouse | Guillaume Croville et al |
| 2135 | Netherlands/NA_218/2020 | 2020-03-26 | Dutch COVID-19 response team | Erasmus Medical Center | Bas Oude Munnink et al |
| 2136 | Taiwan/TSGH-19/2020 | 2020-03-26 | TSGH-CP molecular lab | TSGH-CP molecular lab | Cherng-Lih Perng et al |
| 2137 | Taiwan/NTU25/2020 | 2020-03-26 | Department of Laboratory Medicine, National Taiwan University Hospital | Microbial Genomics Core Lab, National Taiwan University Centers of Genomic and Precision Medicine | Shiou-Hwei Yeh et al |
| 2138 | Spain/Valencia235/2020 | 2020-03-26 | Servicio de Microbiologia. Hospital Clinico Universitario de Valencia | Sequencing and Bioinformatics Service and Molecular Epidemiology Research Group. FISABIO-Public Health | Ivan Ansari et al |
| 2139 | England/20139027404/2020 | 2020-03-26 | Respiratory Virus Unit, Microbiology Services Colindale, Public Health England | Respiratory Virus Unit, Microbiology Services Colindale, Public Health England | Monica Galiano et al |
| 2140 | USA/SEARCH-0071-NBG/2020 | 2020-03-26 | Robert Garry lab | Andersen lab at Scripps Research | Allison Smither et al |
| 2141 | USA/WA-UW-2240/2020 | 2020-03-26 | UW Virology Lab | UW Virology Lab | Pavitra Roychoudhury et al |
| 2142 | Brazil/RJ-872/2020 | 2020-03-26 | Instituto Oswaldo Cruz FIOCRUZ - Laboratory of Respiratory Viruses and Measles (LVRS) | Instituto Oswaldo Cruz FIOCRUZ - Laboratory of Respiratory Viruses and Measles (LVRS) | Paola Resende et al |
| 2143 | USA/WA-S368/2020 | 2020-03-26 | Washington State Department of Health | Seattle Flu Study | Chu et al et al |
| 2144 | USA/WA-UW-4609/2020 | 2020-03-26 | UW Virology Lab | UW Virology Lab | Pavitra Roychoudhury et al |
| 2145 | Luxembourg/LNS2913521/2020 | 2020-03-26 | Laboratoire National de Sante, | Laboratoire National de Sante, | Anke |

| | | | | | |
|---|---|---|---|---|---|
| | | | Microbiology, Virology | Microbiology, Epidemiology and Microbial Genomics | Wienecke-Baldacchino et al |
| 2146 | Spain/Valencia193/2020 | 2020-03-26 | Servicio de Microbiologia. Hospital Clinico Universitario de Valencia | Sequencing and Bioinformatics Service and Molecular Epidemiology Research Group. FISABIO-Public Health | Mariana Reyes-Prieto et al |
| 2147 | Denmark/ALAB-SSI523/2020 | 2020-03-26 | Department of Virus and Microbiological Special Diagnostics, Statens Serum Institut, Copenhagen, Denmark, Artillerivej 5, 2300 Copenahgen S | Albertsen lab, Department of Chemistry and Bioscience, Aalborg University, Denmark | Rasmus Kirkegaard et al |
| 2148 | NewZealand/20VR2000/2020-03-27 | 2020-03-27 | Wellington SCL (WN) | Institute of Environmental Science and Research (ESR) Ltd | **Matt Storey et al** |
| 2149 | England/CAMB-79A9B/2020 | 2020-03-27 | Department of Pathology, University of Cambridge | COVID-19 Genomics UK (COG-UK) Consortium | Luke W Meredith et al |
| 2150 | Iceland/553/2020 | 2020-03-27 | The National University Hospital of Iceland | deCODE genetics | Daniel F Gudbjartsson et al |
| 2151 | Russia/Buryatia-84508/2020 | 2020-03-27 | State Research Center of Virology and Biotechnology VECTOR, Department of Collection of Microorganisms | State Research Center of Virology and Biotechnology VECTOR, Department of Collection of Microorganisms | Oleg V. Pyankov et al |
| 2152 | England/CAMB-78540/2020 | 2020-03-27 | Department of Pathology, University of Cambridge | COVID-19 Genomics UK (COG-UK) Consortium | Luke W Meredith et al |
| 2153 | USA/WA-UW-3959/2020 | 2020-03-27 | UW Virology Lab | UW Virology Lab | Pavitra Roychoudhury et al |
| 2154 | Australia/VIC668/2020 | 2020-03-27 | Victorian Infectious Diseases Reference Laboratory (VIDRL) | Microbiological Diagnostic Unit Public Health Laboratory and Victorian Infectious Diseases Reference Laboratory, Doherty Institute | Caly L. et al |
| 2155 | Australia/VIC674/2020 | 2020-03-27 | Victorian Infectious Diseases Reference Laboratory (VIDRL) | Microbiological Diagnostic Unit Public Health Laboratory and Victorian Infectious Diseases Reference Laboratory, Doherty Institute | Caly L. et al |
| 2156 | Australia/VIC677/2020 | 2020-03-27 | Victorian Infectious Diseases Reference Laboratory (VIDRL) | Microbiological Diagnostic Unit Public Health Laboratory and Victorian Infectious Diseases Reference Laboratory, Doherty Institute | Caly L. et al |
| 2157 | Australia/VIC661/2020 | 2020-03-27 | Victorian Infectious Diseases Reference Laboratory (VIDRL) | Microbiological Diagnostic Unit Public Health Laboratory and Victorian Infectious Diseases Reference Laboratory, Doherty Institute | Caly L. et al |
| 2158 | Australia/VIC718/2020 | 2020-03-27 | Victorian Infectious Diseases Reference Laboratory (VIDRL) | Microbiological Diagnostic Unit Public Health Laboratory and Victorian Infectious Diseases Reference Laboratory, Doherty Institute | Caly L. et al |
| 2159 | Australia/VIC680/2020 | 2020-03-27 | Victorian Infectious Diseases Reference Laboratory (VIDRL) | Microbiological Diagnostic Unit Public Health Laboratory and Victorian Infectious Diseases Reference Laboratory, Doherty Institute | Caly L. et al |
| 2160 | Australia/VIC606/2020 | 2020-03-27 | Victorian Infectious Diseases Reference Laboratory (VIDRL) | Microbiological Diagnostic Unit Public Health Laboratory and Victorian Infectious Diseases Reference Laboratory, Doherty Institute | Caly L. et al |
| 2161 | Australia/VIC959/2020 | 2020-03-27 | Microbiological Diagnostic Unit Public Health Laboratory | Microbiological Diagnostic Unit Public Health Laboratory | Seemann T. et al |
| 2162 | Australia/VIC676/2020 | 2020-03-27 | Victorian Infectious Diseases Reference Laboratory (VIDRL) | Microbiological Diagnostic Unit Public Health Laboratory and Victorian Infectious Diseases Reference Laboratory, Doherty Institute | Caly L. et al |
| 2163 | Australia/VIC964/2020 | 2020-03-27 | Microbiological Diagnostic Unit Public Health Laboratory | Microbiological Diagnostic Unit Public Health Laboratory | Seemann T. et al |
| 2164 | Australia/VIC672/2020 | 2020-03-27 | Victorian Infectious Diseases Reference Laboratory | Microbiological Diagnostic Unit Public Health Laboratory and | Caly L. et al |

| | | | | | |
|---|---|---|---|---|---|
| | | | (VIDRL) | Victorian Infectious Diseases Reference Laboratory, Doherty Institute | |
| 2165 | Australia/VIC611/2020 | 2020-03-27 | Victorian Infectious Diseases Reference Laboratory (VIDRL) | Microbiological Diagnostic Unit Public Health Laboratory and Victorian Infectious Diseases Reference Laboratory, Doherty Institute | Caly L. et al |
| 2166 | Australia/VIC966/2020 | 2020-03-27 | Microbiological Diagnostic Unit Public Health Laboratory | Microbiological Diagnostic Unit Public Health Laboratory | Seemann T. et al |
| 2167 | Australia/VIC667/2020 | 2020-03-27 | Victorian Infectious Diseases Reference Laboratory (VIDRL) | Microbiological Diagnostic Unit Public Health Laboratory and Victorian Infectious Diseases Reference Laboratory, Doherty Institute | Caly L. et al |
| 2168 | Australia/NT11/2020 | 2020-03-27 | Royal Darwin Hospital Pathology | Microbiological Diagnostic Unit Public Health Laboratory and Victorian Infectious Diseases Reference Laboratory, Doherty Institute | Meumann et al |
| 2169 | Australia/VIC666/2020 | 2020-03-27 | Victorian Infectious Diseases Reference Laboratory (VIDRL) | Microbiological Diagnostic Unit Public Health Laboratory and Victorian Infectious Diseases Reference Laboratory, Doherty Institute | Caly L. et al |
| 2170 | Australia/VIC678/2020 | 2020-03-27 | Victorian Infectious Diseases Reference Laboratory (VIDRL) | Microbiological Diagnostic Unit Public Health Laboratory and Victorian Infectious Diseases Reference Laboratory, Doherty Institute | Caly L. et al |
| 2171 | NewZealand/20VR1816/2020-03-27 | 2020-03-27 | Southern Community Labs Dunedin | Institute of Environmental Science and Research (ESR) Ltd | **Matt Storey et al** |
| 2172 | Iceland/583/2020 | 2020-03-27 | The National University Hospital of Iceland | deCODE genetics | Daniel F Gudbjartsson et al |
| 2173 | England/CAMB-745BD/2020 | 2020-03-27 | Department of Pathology, University of Cambridge | COVID-19 Genomics UK (COG-UK) Consortium | Luke W Meredith et al |
| 2174 | Australia/VIC681/2020 | 2020-03-27 | Victorian Infectious Diseases Reference Laboratory (VIDRL) | Microbiological Diagnostic Unit Public Health Laboratory and Victorian Infectious Diseases Reference Laboratory, Doherty Institute | Caly L. et al |
| 2175 | USA/WA-UW-3895/2020 | 2020-03-27 | UW Virology Lab | UW Virology Lab | Pavitra Roychoudhury et al |
| 2176 | USA/WA-UW-3942/2020 | 2020-03-27 | UW Virology Lab | UW Virology Lab | Pavitra Roychoudhury et al |
| 2177 | USA/WA-UW-3900/2020 | 2020-03-27 | UW Virology Lab | UW Virology Lab | Pavitra Roychoudhury et al |
| 2178 | USA/TX-HMH0283/2020 | 2020-03-27 | Houston Methodist Hospital | Houston Methodist Hospital | S. Wesley Long et al |
| 2179 | Belgium/UGent-40/2020 | 2020-03-27 | Lab voor klinische biologie | Onderzoeksgroep Virologie | Nick Vereecke et al |
| 2180 | Spain/Madrid_H12_2410/2020 | 2020-03-27 | Hospital Universitario 12 de Octubre | Hospital Universitario 12 de Octubre | Esther Viedma et al |
| 2181 | USA/TX-HMH0351/2020 | 2020-03-27 | Houston Methodist Hospital | Houston Methodist Hospital | S. Wesley Long et al |
| 2182 | Australia/VIC664/2020 | 2020-03-27 | Victorian Infectious Diseases Reference Laboratory (VIDRL) | Microbiological Diagnostic Unit Public Health Laboratory and Victorian Infectious Diseases Reference Laboratory, Doherty Institute | Caly L. et al |
| 2183 | Australia/VIC662/2020 | 2020-03-27 | Victorian Infectious Diseases Reference Laboratory (VIDRL) | Microbiological Diagnostic Unit Public Health Laboratory and Victorian Infectious Diseases Reference Laboratory, Doherty Institute | Caly L. et al |
| 2184 | Australia/VIC605/2020 | 2020-03-27 | Victorian Infectious Diseases Reference Laboratory (VIDRL) | Microbiological Diagnostic Unit Public Health Laboratory and Victorian Infectious Diseases Reference Laboratory, Doherty Institute | Caly L. et al |
| 2185 | USA/WA-S379/2020 | 2020-03-27 | Washington State Department of Health | Seattle Flu Study | Chu et al et al |
| 2186 | NewZealand/20VR1818/2020-03-27 | 2020-03-27 | Southern Community Labs Dunedin | Institute of Environmental Science and Research (ESR) Ltd | **Matt Storey et al** |

| | | | | | |
|---|---|---|---|---|---|
| 2187 | NewZealand/20VR1817/2020-03-27 | 2020-03-27 | Southern Community Labs Dunedin | Institute of Environmental Science and Research (ESR) Ltd | **Matt Storey et al** |
| 2188 | NewZealand/20VR1827/2020-03-27 | 2020-03-27 | Southern Community Labs Dunedin | Institute of Environmental Science and Research (ESR) Ltd | **Matt Storey et al** |
| 2189 | NewZealand/20VR1809/2020-03-27 | 2020-03-27 | Southern Community Labs Dunedin | Institute of Environmental Science and Research (ESR) Ltd | **Matt Storey et al** |
| 2190 | NewZealand/20VR1624/2020-03-27 | 2020-03-27 | Southern Community Labs Dunedin | Institute of Environmental Science and Research (ESR) Ltd | **Matt Storey et al** |
| 2191 | NewZealand/20VR1990/2020-03-27 | 2020-03-27 | Wellington SCL (WN) | Institute of Environmental Science and Research (ESR) Ltd | **Matt Storey et al** |
| 2192 | NewZealand/20VR1825/2020-03-27 | 2020-03-27 | Southern Community Labs Dunedin | Institute of Environmental Science and Research (ESR) Ltd | **Matt Storey et al** |
| 2193 | NewZealand/20VR1836/2020-03-27 | 2020-03-27 | Southern Community Labs Dunedin | Institute of Environmental Science and Research (ESR) Ltd | **Matt Storey et al** |
| 2194 | NewZealand/20VR1823/2020-03-27 | 2020-03-27 | Southern Community Labs Dunedin | Institute of Environmental Science and Research (ESR) Ltd | **Matt Storey et al** |
| 2195 | NewZealand/20VR1821/2020-03-27 | 2020-03-27 | Southern Community Labs Dunedin | Institute of Environmental Science and Research (ESR) Ltd | **Matt Storey et al** |
| 2196 | NewZealand/20VR1814/2020-03-27 | 2020-03-27 | Southern Community Labs Dunedin | Institute of Environmental Science and Research (ESR) Ltd | **Matt Storey et al** |
| 2197 | NewZealand/20VR1826/2020-03-27 | 2020-03-27 | Southern Community Labs Dunedin | Institute of Environmental Science and Research (ESR) Ltd | **Matt Storey et al** |
| 2198 | NewZealand/20VR1822/2020-03-27 | 2020-03-27 | Southern Community Labs Dunedin | Institute of Environmental Science and Research (ESR) Ltd | **Matt Storey et al** |
| 2199 | NewZealand/20VR1828/2020-03-27 | 2020-03-27 | Southern Community Labs Dunedin | Institute of Environmental Science and Research (ESR) Ltd | **Matt Storey et al** |
| 2200 | USA/TX-HMH0370/2020 | 2020-03-27 | Houston Methodist Hospital | Houston Methodist Hospital | S. Wesley Long et al |
| 2201 | USA/WA-S382/2020 | 2020-03-27 | Washington State Department of Health | Seattle Flu Study | Chu et al et al |
| 2202 | Luxembourg/LNS7812353/2020 | 2020-03-27 | Laboratoire National de Sante, Microbiology, Virology | Laboratoire National de Sante, Microbiology, Epidemiology and Microbial Genomics | Anke Wienecke-Baldacchino et al |
| 2203 | England/20139009204/2020 | 2020-03-27 | Respiratory Virus Unit, Microbiology Services Colindale, Public Health England | Respiratory Virus Unit, Microbiology Services Colindale, Public Health England | Monica Galiano et al |
| 2204 | England/CAMB-79AF5/2020 | 2020-03-27 | Department of Pathology, University of Cambridge | COVID-19 Genomics UK (COG-UK) Consortium | Luke W Meredith et al |
| 2205 | Scotland/CVR901/2020 | 2020-03-27 | West of Scotland Specialist Virology Centre, NHSGGC / MRC-University of Glasgow Centre for Virus Research | COVID-19 Genomics UK (COG-UK) Consortium | Ana da Silva Filipe et al |
| 2206 | Iceland/544/2020 | 2020-03-27 | The National University Hospital of Iceland | deCODE genetics | Daniel F Gudbjartsson et al |
| 2207 | England/20139008704/2020 | 2020-03-27 | Respiratory Virus Unit, Microbiology Services Colindale, Public Health England | Respiratory Virus Unit, Microbiology Services Colindale, Public Health England | Monica Galiano et al |
| 2208 | England/SHEF-C02BB/2020 | 2020-03-27 | Virology Department, Sheffield Teaching Hospitals NHS Foundation Trust | Department of Infection, Immunity and Cardiovascular Disease, The Florey Institute, The Medical School, University of Sheffield | Thushan de Silva et al |
| 2209 | USA/VA-DCLS-0047/2020 | 2020-03-27 | Virginia DCLS | Virginia DCLS | Virginia DCLS et al |
| 2210 | Spain/Madrid_H12_2902/2020 | 2020-03-27 | Hospital Universitario 12 de Octubre | Hospital Universitario 12 de Octubre | Esther Viedma et al |
| 2211 | USA/WA-UW-3932/2020 | 2020-03-27 | UW Virology Lab | UW Virology Lab | Pavitra Roychoudhury et al |
| 2212 | USA/WA-UW-3843/2020 | 2020-03-27 | UW Virology Lab | UW Virology Lab | Pavitra Roychoudhury et al |
| 2213 | USA/NY-Wadsworth-31617-01/2020 | 2020-03-27 | Wadsworth Center, New York State Department.of Health | Wadsworth Center, New York State Department.of Health | Kirsten St. George et al |
| 2214 | Netherlands/NA_234/2020 | 2020-03-27 | Dutch COVID-19 response team | Erasmus Medical Center | Bas Oude Munnink et al |
| 2215 | Netherlands/NA_229/2020 | 2020-03-27 | Dutch COVID-19 response team | Erasmus Medical Center | Bas Oude Munnink et al |
| 2216 | Iceland/585/2020 | 2020-03-27 | The National University Hospital of Iceland | deCODE genetics | Daniel F Gudbjartsson et al |

| | | | | | |
|---|---|---|---|---|---|
| 2217 | Iceland/582/2020 | 2020-03-27 | The National University Hospital of Iceland | deCODE genetics | Daniel F Gudbjartsson et al |
| 2218 | Iceland/540/2020 | 2020-03-27 | deCODE genetics | deCODE genetics | Daniel F Gudbjartsson et al |
| 2219 | Iceland/562/2020 | 2020-03-27 | The National University Hospital of Iceland | deCODE genetics | Daniel F Gudbjartsson et al |
| 2220 | Iceland/579/2020 | 2020-03-27 | The National University Hospital of Iceland | deCODE genetics | Daniel F Gudbjartsson et al |
| 2221 | Iceland/593/2020 | 2020-03-27 | The National University Hospital of Iceland | deCODE genetics | Daniel F Gudbjartsson et al |
| 2222 | Iceland/537/2020 | 2020-03-27 | The National University Hospital of Iceland | deCODE genetics | Daniel F Gudbjartsson et al |
| 2223 | Iceland/594/2020 | 2020-03-27 | The National University Hospital of Iceland | deCODE genetics | Daniel F Gudbjartsson et al |
| 2224 | Iceland/551/2020 | 2020-03-27 | The National University Hospital of Iceland | deCODE genetics | Daniel F Gudbjartsson et al |
| 2225 | Iceland/556/2020 | 2020-03-27 | The National University Hospital of Iceland | deCODE genetics | Daniel F Gudbjartsson et al |
| 2226 | Iceland/581/2020 | 2020-03-27 | The National University Hospital of Iceland | deCODE genetics | Daniel F Gudbjartsson et al |
| 2227 | Iceland/539/2020 | 2020-03-27 | The National University Hospital of Iceland | deCODE genetics | Daniel F Gudbjartsson et al |
| 2228 | Iceland/577/2020 | 2020-03-27 | The National University Hospital of Iceland | deCODE genetics | Daniel F Gudbjartsson et al |
| 2229 | Iceland/584/2020 | 2020-03-27 | The National University Hospital of Iceland | deCODE genetics | Daniel F Gudbjartsson et al |
| 2230 | Iceland/578/2020 | 2020-03-27 | The National University Hospital of Iceland | deCODE genetics | Daniel F Gudbjartsson et al |
| 2231 | Iceland/554/2020 | 2020-03-27 | The National University Hospital of Iceland | deCODE genetics | Daniel F Gudbjartsson et al |
| 2232 | Iceland/558/2020 | 2020-03-27 | The National University Hospital of Iceland | deCODE genetics | Daniel F Gudbjartsson et al |
| 2233 | Spain/Madrid_H12_2501/2020 | 2020-03-27 | Hospital Universitario 12 de Octubre | Hospital Universitario 12 de Octubre | Esther Viedma et al |
| 2234 | USA/WI-238/2020 | 2020-03-27 | University of Wisconsin-Madison AIDS Vaccine Research Laboratories | University of Wisconsin-Madison AIDS Vaccine Research Laboratories | Gage Moreno and Katarina Braun et al |
| 2235 | Iceland/538/2020 | 2020-03-27 | The National University Hospital of Iceland | deCODE genetics | Daniel F Gudbjartsson et al |
| 2236 | Iceland/543/2020 | 2020-03-27 | The National University Hospital of Iceland | deCODE genetics | Daniel F Gudbjartsson et al |
| 2237 | Iceland/560/2020 | 2020-03-27 | The National University Hospital of Iceland | deCODE genetics | Daniel F Gudbjartsson et al |
| 2238 | Iceland/592/2020 | 2020-03-27 | The National University Hospital of Iceland | deCODE genetics | Daniel F Gudbjartsson et al |
| 2239 | Iceland/557/2020 | 2020-03-27 | The National University Hospital of Iceland | deCODE genetics | Daniel F Gudbjartsson et al |
| 2240 | Iceland/595/2020 | 2020-03-27 | The National University Hospital of Iceland | deCODE genetics | Daniel F Gudbjartsson et al |
| 2241 | Wales/PHWC-2663F/2020 | 2020-03-27 | Wales Specialist Virology Centre | Public Health Wales Microbiology Cardiff | Catherine Moore et al |
| 2242 | Wales/PHWC-26699/2020 | 2020-03-27 | Wales Specialist Virology Centre | Public Health Wales Microbiology Cardiff | Catherine Moore et al |
| 2243 | Spain/Valencia251/2020 | 2020-03-27 | Servicio de Microbiologia. Hospital Clinico Universitario de Valencia | Sequencing and Bioinformatics Service and Molecular Epidemiology Research Group. FISABIO-Public Health | Loreto Ferrús Abad et al |
| 2244 | Belgium/UGent-37/2020 | 2020-03-27 | Lab voor klinische biologie | Onderzoeksgroep Virologie | Laurens Lambrechts et al |
| 2245 | Taiwan/TSGH-20/2020 | 2020-03-27 | TSGH-CP molecular lab | TSGH-CP molecular lab | Cherng-Lih Perng et al |
| 2246 | Scotland/CVR833/2020 | 2020-03-27 | West of Scotland Specialist Virology Centre, NHSGGC / MRC-University of Glasgow Centre for Virus Research | COVID-19 Genomics UK (COG-UK) Consortium | Ana da Silva Filipe et al |
| 2247 | USA/WA-S378/2020 | 2020-03-27 | Washington State Department of Health | Seattle Flu Study | Chu et al et al |
| 2248 | Spain/Madrid_H12_2309/2020 | 2020-03-27 | Hospital Universitario 12 de Octubre | Hospital Universitario 12 de Octubre | Esther Viedma et al |

| | | | | | |
|---|---|---|---|---|---|
| 2249 | USA/AZ-TG271898/2020 | 2020-03-27 | AZ SPHL, Arizona Department of Health Services | TGen North | Jolene Bowers et al |
| 2250 | USA/WA-UW-3908/2020 | 2020-03-27 | UW Virology Lab | UW Virology Lab | Pavitra Roychoudhury et al |
| 2251 | Spain/Madrid_H12_2208/2020 | 2020-03-27 | Hospital Universitario 12 de Octubre | Hospital Universitario 12 de Octubre | Esther Viedma et al |
| 2252 | USA/WI-248/2020 | 2020-03-28 | University of Wisconsin-Madison AIDS Vaccine Research Laboratories | University of Wisconsin-Madison AIDS Vaccine Research Laboratories | Gage Moreno and Katarina Braun et al |
| 2253 | Hungary/SRC-03597w/2020 | 2020-03-28 | Virological Research Group, Szentágothai Research Centre | Bioinformatics Research Group, Szentágothai Research Centre | Péter Urbán et al |
| 2254 | Australia/VIC685/2020 | 2020-03-28 | Victorian Infectious Diseases Reference Laboratory (VIDRL) | Microbiological Diagnostic Unit Public Health Laboratory and Victorian Infectious Diseases Reference Laboratory, Doherty Institute | Caly L. et al |
| 2255 | Australia/VIC657/2020 | 2020-03-28 | Victorian Infectious Diseases Reference Laboratory (VIDRL) | Microbiological Diagnostic Unit Public Health Laboratory and Victorian Infectious Diseases Reference Laboratory, Doherty Institute | Caly L. et al |
| 2256 | Australia/VIC623/2020 | 2020-03-28 | Victorian Infectious Diseases Reference Laboratory (VIDRL) | Microbiological Diagnostic Unit Public Health Laboratory and Victorian Infectious Diseases Reference Laboratory, Doherty Institute | Caly L. et al |
| 2257 | Australia/VIC658/2020 | 2020-03-28 | Victorian Infectious Diseases Reference Laboratory (VIDRL) | Microbiological Diagnostic Unit Public Health Laboratory and Victorian Infectious Diseases Reference Laboratory, Doherty Institute | Caly L. et al |
| 2258 | Australia/VIC697/2020 | 2020-03-28 | Victorian Infectious Diseases Reference Laboratory (VIDRL) | Microbiological Diagnostic Unit Public Health Laboratory and Victorian Infectious Diseases Reference Laboratory, Doherty Institute | Caly L. et al |
| 2259 | Australia/VIC651/2020 | 2020-03-28 | Victorian Infectious Diseases Reference Laboratory (VIDRL) | Microbiological Diagnostic Unit Public Health Laboratory and Victorian Infectious Diseases Reference Laboratory, Doherty Institute | Caly L. et al |
| 2260 | Australia/VIC615/2020 | 2020-03-28 | Victorian Infectious Diseases Reference Laboratory (VIDRL) | Microbiological Diagnostic Unit Public Health Laboratory and Victorian Infectious Diseases Reference Laboratory, Doherty Institute | Caly L. et al |
| 2261 | USA/WI-242/2020 | 2020-03-28 | University of Wisconsin-Madison AIDS Vaccine Research Laboratories | University of Wisconsin-Madison AIDS Vaccine Research Laboratories | Gage Moreno and Katarina Braun et al |
| 2262 | Australia/VIC808/2020 | 2020-03-28 | Victorian Infectious Diseases Reference Laboratory (VIDRL) | Microbiological Diagnostic Unit Public Health Laboratory and Victorian Infectious Diseases Reference Laboratory, Doherty Institute | Caly L. et al |
| 2263 | Australia/VIC656/2020 | 2020-03-28 | Victorian Infectious Diseases Reference Laboratory (VIDRL) | Microbiological Diagnostic Unit Public Health Laboratory and Victorian Infectious Diseases Reference Laboratory, Doherty Institute | Caly L. et al |
| 2264 | Australia/VIC644/2020 | 2020-03-28 | Victorian Infectious Diseases Reference Laboratory (VIDRL) | Microbiological Diagnostic Unit Public Health Laboratory and Victorian Infectious Diseases Reference Laboratory, Doherty Institute | Caly L. et al |
| 2265 | Australia/VIC696/2020 | 2020-03-28 | Victorian Infectious Diseases Reference Laboratory (VIDRL) | Microbiological Diagnostic Unit Public Health Laboratory and Victorian Infectious Diseases Reference Laboratory, Doherty Institute | Caly L. et al |
| 2266 | Australia/VIC660/2020 | 2020-03-28 | Victorian Infectious Diseases Reference Laboratory (VIDRL) | Microbiological Diagnostic Unit Public Health Laboratory and Victorian Infectious Diseases | Caly L. et al |

| | | | | | |
|---|---|---|---|---|---|
| | | | | Reference Laboratory, Doherty Institute | |
| 2267 | Australia/VIC655/2020 | 2020-03-28 | Victorian Infectious Diseases Reference Laboratory (VIDRL) | Microbiological Diagnostic Unit Public Health Laboratory and Victorian Infectious Diseases Reference Laboratory, Doherty Institute | Caly L. et al |
| 2268 | Australia/VIC645/2020 | 2020-03-28 | Victorian Infectious Diseases Reference Laboratory (VIDRL) | Microbiological Diagnostic Unit Public Health Laboratory and Victorian Infectious Diseases Reference Laboratory, Doherty Institute | Caly L. et al |
| 2269 | Australia/VIC-CBA3/2020 | 2020-03-28 | Geelong Centre for Emerging Infectious Diseases | Geelong Centre for Emerging Infectious Diseases | Chamings et al |
| 2270 | Australia/VIC694/2020 | 2020-03-28 | Victorian Infectious Diseases Reference Laboratory (VIDRL) | Microbiological Diagnostic Unit Public Health Laboratory and Victorian Infectious Diseases Reference Laboratory, Doherty Institute | Caly L. et al |
| 2271 | Australia/VIC647/2020 | 2020-03-28 | Victorian Infectious Diseases Reference Laboratory (VIDRL) | Microbiological Diagnostic Unit Public Health Laboratory and Victorian Infectious Diseases Reference Laboratory, Doherty Institute | Caly L. et al |
| 2272 | Australia/VIC652/2020 | 2020-03-28 | Victorian Infectious Diseases Reference Laboratory (VIDRL) | Microbiological Diagnostic Unit Public Health Laboratory and Victorian Infectious Diseases Reference Laboratory, Doherty Institute | Caly L. et al |
| 2273 | Australia/VIC636/2020 | 2020-03-28 | Victorian Infectious Diseases Reference Laboratory (VIDRL) | Microbiological Diagnostic Unit Public Health Laboratory and Victorian Infectious Diseases Reference Laboratory, Doherty Institute | Caly L. et al |
| 2274 | CzechRepublic/IAB_22/2020 | 2020-03-28 | Motol University Hospital | Institute of Applied Biotechnologies a.s. | Petr Brož et al |
| 2275 | USA/CA-CZB084/2020 | 2020-03-28 | UCSF Clinical Microbiology Laboratory | Chan-Zuckerberg Biohub | CZB Cliahub Consortium et al |
| 2276 | USA/TX-HMH0300/2020 | 2020-03-28 | Houston Methodist Hospital | Houston Methodist Hospital | S. Wesley Long et al |
| 2277 | USA/CA-CZB077/2020 | 2020-03-28 | UCSF Clinical Microbiology Laboratory | Chan-Zuckerberg Biohub | CZB Cliahub Consortium et al |
| 2278 | CzechRepublic/IAB_9/2020 | 2020-03-28 | Motol University Hospital | Institute of Applied Biotechnologies a.s. | Petr Brož et al |
| 2279 | USA/WA-S401/2020 | 2020-03-28 | Washington State Department of Health | Seattle Flu Study | Chu et al et al |
| 2280 | Italy/TE7859/2020 | 2020-03-28 | Ospedale Civile S. Liberatore di Atri | "Istituto Zooprofilattico Sperimentale dell'Abruzzo e Molise ""G. Caporale""" | Lorusso A et al |
| 2281 | Spain/Valencia215/2020 | 2020-03-28 | Servicio de Microbiologia. Hospital Clinico Universitario de Valencia | Sequencing and Bioinformatics Service and Molecular Epidemiology Research Group. FISABIO-Public Health | Ivan Ansari et al |
| 2282 | USA/VA-DCLS-0052/2020 | 2020-03-28 | Virginia DCLS | Virginia DCLS | Virginia DCLS et al |
| 2283 | USA/DC-CDC-1738/2020 | 2020-03-28 | DC Public Health Lab Dept of Forensic Science | Pathogen Discovery, Respiratory Viruses Branch, Division of Viral Diseases, Centers for Disease Control and Prevention | Ying Tao et al |
| 2284 | USA/WA-S577/2020 | 2020-03-28 | Washington State Department of Health | Seattle Flu Study | Chu et al et al |
| 2285 | USA/CA-CZB090/2020 | 2020-03-28 | UCSF Clinical Microbiology Laboratory | Chan-Zuckerberg Biohub | CZB Cliahub Consortium et al |
| 2286 | NewZealand/20VR1838/2020-03-28 | 2020-03-28 | Southern Community Labs Dunedin | Institute of Environmental Science and Research (ESR) Ltd | **Matt Storey et al** |
| 2287 | NewZealand/20VR1843/2020-03-28 | 2020-03-28 | Southern Community Labs Dunedin | Institute of Environmental Science and Research (ESR) Ltd | **Matt Storey et al** |
| 2288 | NewZealand/20VR1847/2020-03-28 | 2020-03-28 | Southern Community Labs Dunedin | Institute of Environmental Science and Research (ESR) Ltd | **Matt Storey et al** |
| 2289 | NewZealand/20VR1997/2020-03-28 | 2020-03-28 | Wellington SCL (WN) | Institute of Environmental Science and Research (ESR) Ltd | **Matt Storey et al** |
| 2290 | NewZealand/20VR1839/2020-03-28 | 2020-03-28 | Southern Community Labs | Institute of Environmental | **Matt Storey et al** |

| | | | | | |
|---|---|---|---|---|---|
| | | | Dunedin | Science and Research (ESR) Ltd | |
| 2291 | NewZealand/20VR1769/2020-03-28 | 2020-03-28 | Waikato Hospital | Institute of Environmental Science and Research (ESR) Ltd | **Matt Storey et al** |
| 2292 | USA/WA-S427/2020 | 2020-03-28 | Washington State Department of Health | Seattle Flu Study | Chu et al et al |
| 2293 | USA/TX-HMH0305/2020 | 2020-03-28 | Houston Methodist Hospital | Houston Methodist Hospital | S. Wesley Long et al |
| 2294 | England/20140002804/2020 | 2020-03-28 | Respiratory Virus Unit, Microbiology Services Colindale, Public Health England | Respiratory Virus Unit, Microbiology Services Colindale, Public Health England | Monica Galiano et al |
| 2295 | England/SHEF-C0224/2020 | 2020-03-28 | Virology Department, Sheffield Teaching Hospitals NHS Foundation Trust | Department of Infection, Immunity and Cardiovascular Disease, The Florey Institute, The Medical School, University of Sheffield | Thushan de Silva et al |
| 2296 | England/20139046604/2020 | 2020-03-28 | Respiratory Virus Unit, Microbiology Services Colindale, Public Health England | Respiratory Virus Unit, Microbiology Services Colindale, Public Health England | Monica Galiano et al |
| 2297 | England/CAMB-748E1/2020 | 2020-03-28 | Department of Pathology, University of Cambridge | COVID-19 Genomics UK (COG-UK) Consortium | Luke W Meredith et al |
| 2298 | Iceland/576/2020 | 2020-03-28 | The National University Hospital of Iceland | deCODE genetics | Daniel F Gudbjartsson et al |
| 2299 | India/NCDC-02251/2020 | 2020-03-28 | National Centre for Disease control (NCDC), CSIR-Institute of Genomics and Integrative Biology (CSIR-IGIB) | NCDC/CSIR-IGIB | Pramod Kumar et al |
| 2300 | Spain/Valencia212/2020 | 2020-03-28 | Servicio de Microbiologia. Hospital Clinico Universitario de Valencia | Sequencing and Bioinformatics Service and Molecular Epidemiology Research Group. FISABIO-Public Health | Paula Ruiz-Hueso et al |
| 2301 | USA/CA-CZB0108/2020 | 2020-03-28 | UCSF Clinical Microbiology Laboratory | Chan-Zuckerberg Biohub | CZB Cliahub Consortium et al |
| 2302 | USA/WA-UW-4734/2020 | 2020-03-28 | UW Virology Lab | UW Virology Lab | Pavitra Roychoudhury et al |
| 2303 | USA/WA-S370/2020 | 2020-03-28 | Washington State Department of Health | Seattle Flu Study | Chu et al et al |
| 2304 | Spain/Valencia216/2020 | 2020-03-28 | Servicio de Microbiologia. Hospital Clinico Universitario de Valencia | Sequencing and Bioinformatics Service and Molecular Epidemiology Research Group. FISABIO-Public Health | David Navarro et al |
| 2305 | Netherlands/Friesland_6/2020 | 2020-03-28 | Dutch COVID-19 response team | Erasmus Medical Center | Bas Oude Munnink et al |
| 2306 | Scotland/CVR1053/2020 | 2020-03-28 | West of Scotland Specialist Virology Centre, NHSGGC / MRC-University of Glasgow Centre for Virus Research | COVID-19 Genomics UK (COG-UK) Consortium | Ana da Silva Filipe et al |
| 2307 | USA/CA-CZB069/2020 | 2020-03-28 | UCSF Clinical Microbiology Laboratory | Chan-Zuckerberg Biohub | CZB Cliahub Consortium et al |
| 2308 | Iceland/599/2020 | 2020-03-28 | The National University Hospital of Iceland | deCODE genetics | Daniel F Gudbjartsson et al |
| 2309 | Iceland/586/2020 | 2020-03-28 | The National University Hospital of Iceland | deCODE genetics | Daniel F Gudbjartsson et al |
| 2310 | Iceland/604/2020 | 2020-03-28 | The National University Hospital of Iceland | deCODE genetics | Daniel F Gudbjartsson et al |
| 2311 | Iceland/571/2020 | 2020-03-28 | The National University Hospital of Iceland | deCODE genetics | Daniel F Gudbjartsson et al |
| 2312 | Iceland/597/2020 | 2020-03-28 | The National University Hospital of Iceland | deCODE genetics | Daniel F Gudbjartsson et al |
| 2313 | Iceland/602/2020 | 2020-03-28 | The National University Hospital of Iceland | deCODE genetics | Daniel F Gudbjartsson et al |
| 2314 | Iceland/570/2020 | 2020-03-28 | The National University Hospital of Iceland | deCODE genetics | Daniel F Gudbjartsson et al |
| 2315 | Iceland/561/2020 | 2020-03-28 | The National University Hospital of Iceland | deCODE genetics | Daniel F Gudbjartsson et al |
| 2316 | Iceland/575/2020 | 2020-03-28 | The National University Hospital of Iceland | deCODE genetics | Daniel F Gudbjartsson et al |
| 2317 | Iceland/564/2020 | 2020-03-28 | The National University | deCODE genetics | Daniel F Gudbjartsson |

| | | | | | |
|---|---|---|---|---|---|
| | | | Hospital of Iceland | | et al |
| 2318 | Iceland/596/2020 | 2020-03-28 | The National University Hospital of Iceland | deCODE genetics | Daniel F Gudbjartsson et al |
| 2319 | Iceland/565/2020 | 2020-03-28 | The National University Hospital of Iceland | deCODE genetics | Daniel F Gudbjartsson et al |
| 2320 | Iceland/573/2020 | 2020-03-28 | The National University Hospital of Iceland | deCODE genetics | Daniel F Gudbjartsson et al |
| 2321 | Iceland/568/2020 | 2020-03-28 | The National University Hospital of Iceland | deCODE genetics | Daniel F Gudbjartsson et al |
| 2322 | Iceland/547/2020 | 2020-03-28 | deCODE genetics | deCODE genetics | Daniel F Gudbjartsson et al |
| 2323 | Iceland/601/2020 | 2020-03-28 | The National University Hospital of Iceland | deCODE genetics | Daniel F Gudbjartsson et al |
| 2324 | India/NCDC-02242/2020 | 2020-03-28 | National Centre for Disease control (NCDC), CSIR-Institute of Genomics and Integrative Biology (CSIR-IGIB) | NCDC/CSIR-IGIB | Pramod Kumar et al |
| 2325 | Iceland/550/2020 | 2020-03-28 | deCODE genetics | deCODE genetics | Daniel F Gudbjartsson et al |
| 2326 | Iceland/548/2020 | 2020-03-28 | deCODE genetics | deCODE genetics | Daniel F Gudbjartsson et al |
| 2327 | USA/CA-CZB080/2020 | 2020-03-28 | UCSF Clinical Microbiology Laboratory | Chan-Zuckerberg Biohub | CZB Cliahub Consortium et al |
| 2328 | USA/VA-DCLS-0073/2020 | 2020-03-28 | Virginia DCLS | Virginia DCLS | Virginia DCLS et al |
| 2329 | England/20139046504/2020 | 2020-03-28 | Respiratory Virus Unit, Microbiology Services Colindale, Public Health England | Respiratory Virus Unit, Microbiology Services Colindale, Public Health England | Monica Galiano et al |
| 2330 | USA/VA-DCLS-0046/2020 | 2020-03-28 | Virginia DCLS | Virginia DCLS | Virginia DCLS et al |
| 2331 | USA/CA-CZB085/2020 | 2020-03-28 | UCSF Clinical Microbiology Laboratory | Chan-Zuckerberg Biohub | CZB Cliahub Consortium et al |
| 2332 | USA/WI-245/2020 | 2020-03-28 | University of Wisconsin-Madison AIDS Vaccine Research Laboratories | University of Wisconsin-Madison AIDS Vaccine Research Laboratories | Gage Moreno and Katarina Braun et al |
| 2333 | USA/WA-UW-4775/2020 | 2020-03-28 | UW Virology Lab | UW Virology Lab | Pavitra Roychoudhury et al |
| 2334 | USA/TX-HMH0324/2020 | 2020-03-28 | Houston Methodist Hospital | Houston Methodist Hospital | S. Wesley Long et al |
| 2335 | USA/WA-S432/2020 | 2020-03-28 | Washington State Department of Health | Seattle Flu Study | Chu et al et al |
| 2336 | USA/WA-S400/2020 | 2020-03-28 | Washington State Department of Health | Seattle Flu Study | Chu et al et al |
| 2337 | England/20139059004/2020 | 2020-03-28 | Respiratory Virus Unit, Microbiology Services Colindale, Public Health England | Respiratory Virus Unit, Microbiology Services Colindale, Public Health England | Monica Galiano et al |
| 2338 | Scotland/EDB326/2020 | 2020-03-28 | Virology Department, Royal Infirmary of Edinburgh, NHS Lothian / School of Biological Sciences, University of Edinburgh / Institute of Genetics and Molecular Medicine, University of Edinburgh | COVID-19 Genomics UK (COG-UK) Consortium | McHugh M et al |
| 2339 | Russia/Chelyabinsk-85311/2020 | 2020-03-28 | State Research Center of Virology and Biotechnology VECTOR, Department of Collection of Microorganisms | State Research Center of Virology and Biotechnology VECTOR, Department of Collection of Microorganisms | Sergey A. Bodnev et al |
| 2340 | Denmark/ALAB-SSI592/2020 | 2020-03-28 | Department of Virus and Microbiological Special Diagnostics, Statens Serum Institut, Copenhagen, Denmark, Artillerivej 5, 2300 Copenahgen S | Albertsen lab, Department of Chemistry and Bioscience, Aalborg University, Denmark | Rasmus Kirkegaard et al |
| 2341 | USA/ID-UW-4396/2020 | 2020-03-28 | UW Virology Lab | UW Virology Lab | Pavitra Roychoudhury et al |
| 2342 | USA/WA-S505/2020 | 2020-03-29 | Washington State Department of Health | Seattle Flu Study | Chu et al et al |
| 2343 | Australia/VIC627/2020 | 2020-03-29 | Victorian Infectious Diseases | Microbiological Diagnostic Unit | Caly L. et al |

| | | | | | |
|---|---|---|---|---|---|
| | | | Reference Laboratory (VIDRL) | Public Health Laboratory and Victorian Infectious Diseases Reference Laboratory, Doherty Institute | |
| 2344 | Australia/VIC762/2020 | 2020-03-29 | Victorian Infectious Diseases Reference Laboratory (VIDRL) | Microbiological Diagnostic Unit Public Health Laboratory and Victorian Infectious Diseases Reference Laboratory, Doherty Institute | Caly L. et al |
| 2345 | Australia/VIC639/2020 | 2020-03-29 | Victorian Infectious Diseases Reference Laboratory (VIDRL) | Microbiological Diagnostic Unit Public Health Laboratory and Victorian Infectious Diseases Reference Laboratory, Doherty Institute | Caly L. et al |
| 2346 | USA/TX-HMH0359/2020 | 2020-03-29 | Houston Methodist Hospital | Houston Methodist Hospital | S. Wesley Long et al |
| 2347 | USA/VA-DCLS-0075/2020 | 2020-03-29 | Virginia DCLS | Virginia DCLS | Virginia DCLS et al |
| 2348 | Spain/Valencia246/2020 | 2020-03-29 | Servicio de Microbiologia. Hospital Clinico Universitario de Valencia | Sequencing and Bioinformatics Service and Molecular Epidemiology Research Group. FISABIO-Public Health | Lidia Ruiz Roldan et al |
| 2349 | England/20140066004/2020 | 2020-03-29 | Respiratory Virus Unit, Microbiology Services Colindale, Public Health England | Respiratory Virus Unit, Microbiology Services Colindale, Public Health England | Monica Galiano et al |
| 2350 | USA/WA-S501/2020 | 2020-03-29 | Washington State Department of Health | Seattle Flu Study | Chu et al et al |
| 2351 | Australia/VIC682/2020 | 2020-03-29 | Victorian Infectious Diseases Reference Laboratory (VIDRL) | Microbiological Diagnostic Unit Public Health Laboratory and Victorian Infectious Diseases Reference Laboratory, Doherty Institute | Caly L. et al |
| 2352 | Australia/VIC628/2020 | 2020-03-29 | Victorian Infectious Diseases Reference Laboratory (VIDRL) | Microbiological Diagnostic Unit Public Health Laboratory and Victorian Infectious Diseases Reference Laboratory, Doherty Institute | Caly L. et al |
| 2353 | Australia/VIC638/2020 | 2020-03-29 | Victorian Infectious Diseases Reference Laboratory (VIDRL) | Microbiological Diagnostic Unit Public Health Laboratory and Victorian Infectious Diseases Reference Laboratory, Doherty Institute | Caly L. et al |
| 2354 | Australia/VIC744/2020 | 2020-03-29 | Victorian Infectious Diseases Reference Laboratory (VIDRL) | Microbiological Diagnostic Unit Public Health Laboratory and Victorian Infectious Diseases Reference Laboratory, Doherty Institute | Caly L. et al |
| 2355 | Australia/VIC708/2020 | 2020-03-29 | Victorian Infectious Diseases Reference Laboratory (VIDRL) | Microbiological Diagnostic Unit Public Health Laboratory and Victorian Infectious Diseases Reference Laboratory, Doherty Institute | Caly L. et al |
| 2356 | Australia/VIC710/2020 | 2020-03-29 | Victorian Infectious Diseases Reference Laboratory (VIDRL) | Microbiological Diagnostic Unit Public Health Laboratory and Victorian Infectious Diseases Reference Laboratory, Doherty Institute | Caly L. et al |
| 2357 | Australia/VIC712/2020 | 2020-03-29 | Victorian Infectious Diseases Reference Laboratory (VIDRL) | Microbiological Diagnostic Unit Public Health Laboratory and Victorian Infectious Diseases Reference Laboratory, Doherty Institute | Caly L. et al |
| 2358 | Australia/VIC978/2020 | 2020-03-29 | Victorian Infectious Diseases Reference Laboratory (VIDRL) | Microbiological Diagnostic Unit Public Health Laboratory and Victorian Infectious Diseases Reference Laboratory, The Peter Doherty Institute for Infection and Immunity | Caly L. et al |
| 2359 | India/NCDC-02321/2020 | 2020-03-29 | National Centre for Disease control (NCDC), CSIR-Institute of Genomics and Integrative Biology (CSIR-IGIB) | NCDC/CSIR-IGIB | Pramod Kumar et al |

| | | | | | |
|---|---|---|---|---|---|
| 2360 | USA/ID-UW-4486/2020 | 2020-03-29 | UW Virology Lab | UW Virology Lab | Pavitra Roychoudhury et al |
| 2361 | England/20142026304/2020 | 2020-03-29 | Respiratory Virus Unit, Microbiology Services Colindale, Public Health England | Respiratory Virus Unit, Microbiology Services Colindale, Public Health England | Monica Galiano et al |
| 2362 | England/20139072204/2020 | 2020-03-29 | Respiratory Virus Unit, Microbiology Services Colindale, Public Health England | Respiratory Virus Unit, Microbiology Services Colindale, Public Health England | Monica Galiano et al |
| 2363 | CzechRepublic/IAB_23/2020 | 2020-03-29 | Motol University Hospital | Institute of Applied Biotechnologies a.s. | Petr Brož et al |
| 2364 | Wales/PHWC-2707C/2020 | 2020-03-29 | Wales Specialist Virology Centre | Public Health Wales Microbiology Cardiff | Catherine Moore et al |
| 2365 | England/SHEF-C07BC/2020 | 2020-03-29 | Virology Department, Sheffield Teaching Hospitals NHS Foundation Trust | Department of Infection, Immunity and Cardiovascular Disease, The Florey Institute, The Medical School, University of Sheffield | Thushan de Silva et al |
| 2366 | Belgium/UGent-46/2020 | 2020-03-29 | Lab voor klinische biologie | Onderzoeksgroep Virologie | Nick Vereecke et al |
| 2367 | Belgium/UGent-45/2020 | 2020-03-29 | Lab voor klinische biologie | Onderzoeksgroep Virologie | Nick Vereecke et al |
| 2368 | USA/WA-S566/2020 | 2020-03-29 | Washington State Department of Health | Seattle Flu Study | Chu et al et al |
| 2369 | USA/TX-HMH0353/2020 | 2020-03-29 | Houston Methodist Hospital | Houston Methodist Hospital | S. Wesley Long et al |
| 2370 | Spain/Madrid_H12_1905/2020 | 2020-03-29 | Hospital Universitario 12 de Octubre | Hospital Universitario 12 de Octubre | Esther Viedma et al |
| 2371 | USA/AK-PHL077/2020 | 2020-03-29 | Alaska State Virology Laboratory | Alaska State Virology Laboratory | Chen et al |
| 2372 | USA/WA-UW-3990/2020 | 2020-03-29 | UW Virology Lab | UW Virology Lab | Pavitra Roychoudhury et al |
| 2373 | USA/CA-CZB-1031/2020 | 2020-03-29 | County of Santa Clara Public Health Department | Chan-Zuckerberg Biohub | CZB Cliahub Consortium et al |
| 2374 | NewZealand/20VR2002/2020-03-29 | 2020-03-29 | Wellington SCL (WN) | Institute of Environmental Science and Research (ESR) Ltd | **Matt Storey et al** |
| 2375 | NewZealand/20VR1804/2020-03-29 | 2020-03-29 | Waikato Hospital | Institute of Environmental Science and Research (ESR) Ltd | **Matt Storey et al** |
| 2376 | NewZealand/20VR1837/2020-03-29 | 2020-03-29 | Southern Community Labs Dunedin | Institute of Environmental Science and Research (ESR) Ltd | **Matt Storey et al** |
| 2377 | NewZealand/20VR1845/2020-03-29 | 2020-03-29 | Southern Community Labs Dunedin | Institute of Environmental Science and Research (ESR) Ltd | **Matt Storey et al** |
| 2378 | NewZealand/20VR1805/2020-03-29 | 2020-03-29 | Waikato Hospital | Institute of Environmental Science and Research (ESR) Ltd | **Matt Storey et al** |
| 2379 | USA/WA-UW-3997/2020 | 2020-03-29 | UW Virology Lab | UW Virology Lab | Pavitra Roychoudhury et al |
| 2380 | Spain/Madrid_H12_1804/2021 | 2020-03-29 | Hospital Universitario 12 de Octubre | Hospital Universitario 12 de Octubre | Esther Viedma et al |
| 2381 | England/CAMB-7476C/2020 | 2020-03-29 | Department of Pathology, University of Cambridge | COVID-19 Genomics UK (COG-UK) Consortium | Luke W Meredith et al |
| 2382 | England/NOTT-10E933/2020 | 2020-03-29 | Queens Medical Centre, Clinical Microbiology Department / DeepSeq Nottingham | COVID-19 Genomics UK (COG-UK) Consortium | Gemma Clark et al |
| 2383 | Scotland/CVR1140/2020 | 2020-03-29 | West of Scotland Specialist Virology Centre, NHSGGC / MRC-University of Glasgow Centre for Virus Research | COVID-19 Genomics UK (COG-UK) Consortium | Ana da Silva Filipe et al |
| 2384 | USA/ID-UW-4461/2020 | 2020-03-29 | UW Virology Lab | UW Virology Lab | Pavitra Roychoudhury et al |
| 2385 | Russia/Buryatia-86204/2020 | 2020-03-29 | State Research Center of Virology and Biotechnology VECTOR, Department of Collection of Microorganisms | State Research Center of Virology and Biotechnology VECTOR, Department of Collection of Microorganisms | Sergey A. Bodnev et al |
| 2386 | USA/OR-UW-4388/2020 | 2020-03-29 | UW Virology Lab | UW Virology Lab | Pavitra Roychoudhury et al |
| 2387 | USA/WA-S556/2020 | 2020-03-29 | Washington State Department of Health | Seattle Flu Study | Chu et al et al |
| 2388 | Iceland/535/2020 | 2020-03-29 | The National University Hospital of Iceland | deCODE genetics | Daniel F Gudbjartsson et al |

| | | | | | |
|---|---|---|---|---|---|
| 2389 | Iceland/536/2020 | 2020-03-29 | The National University Hospital of Iceland | deCODE genetics | Daniel F Gudbjartsson et al |
| 2390 | Iceland/589/2020 | 2020-03-29 | deCODE genetics | deCODE genetics | Daniel F Gudbjartsson et al |
| 2391 | USA/ID-UW-4383/2020 | 2020-03-29 | UW Virology Lab | UW Virology Lab | Pavitra Roychoudhury et al |
| 2392 | USA/WA-UW-3988/2020 | 2020-03-29 | UW Virology Lab | UW Virology Lab | Pavitra Roychoudhury et al |
| 2393 | USA/TX-HMH0355/2020 | 2020-03-29 | Houston Methodist Hospital | Houston Methodist Hospital | S. Wesley Long et al |
| 2394 | Scotland/CVR1101/2020 | 2020-03-29 | West of Scotland Specialist Virology Centre, NHSGGC / MRC-University of Glasgow Centre for Virus Research | COVID-19 Genomics UK (COG-UK) Consortium | Ana da Silva Filipe et al |
| 2395 | England/20142024404/2020 | 2020-03-29 | Respiratory Virus Unit, Microbiology Services Colindale, Public Health England | Respiratory Virus Unit, Microbiology Services Colindale, Public Health England | Monica Galiano et al |
| 2396 | Scotland/CVR1093/2020 | 2020-03-29 | West of Scotland Specialist Virology Centre, NHSGGC / MRC-University of Glasgow Centre for Virus Research | COVID-19 Genomics UK (COG-UK) Consortium | Ana da Silva Filipe et al |
| 2397 | England/20139065604/2020 | 2020-03-29 | Respiratory Virus Unit, Microbiology Services Colindale, Public Health England | Respiratory Virus Unit, Microbiology Services Colindale, Public Health England | Monica Galiano et al |
| 2398 | USA/WA-S416/2020 | 2020-03-29 | Washington State Department of Health | Seattle Flu Study | Chu et al et al |
| 2399 | USA/WA-UW-3984/2020 | 2020-03-29 | UW Virology Lab | UW Virology Lab | Pavitra Roychoudhury et al |
| 2400 | USA/WI-GMF-00466/2020 | 2020-03-29 | Gundersen Molecular Diagnostics Laboratory | Kabara Cancer Research Institute | Craig S. Richmond et al |
| 2401 | Taiwan/TSGH-33/2020 | 2020-03-29 | TSGH-CP molecular lab | TSGH-CP molecular lab | Cherng-Lih Perng et al |
| 2402 | Taiwan/299/2020 | 2020-03-29 | Centers for Disease Control, R.O.C. (Taiwan) | Centers for Disease Control, R.O.C. (Taiwan) | Ji-Rong Yang et al |
| 2403 | England/20139065404/2020 | 2020-03-29 | Respiratory Virus Unit, Microbiology Services Colindale, Public Health England | Respiratory Virus Unit, Microbiology Services Colindale, Public Health England | Monica Galiano et al |
| 2404 | England/CAMB-7475D/2020 | 2020-03-29 | Department of Pathology, University of Cambridge | COVID-19 Genomics UK (COG-UK) Consortium | Luke W Meredith et al |
| 2405 | USA/WA-S410/2020 | 2020-03-29 | Washington State Department of Health | Seattle Flu Study | Chu et al et al |
| 2406 | USA/TX-HMH0387/2020 | 2020-03-29 | Houston Methodist Hospital | Houston Methodist Hospital | S. Wesley Long et al |
| 2407 | Switzerland/GE2297/2020 | 2020-03-29 | University Hospitals of Geneva Laboratory of Virology | University Hospitals of Geneva Laboratory of Virology | Laubscher F. et al |
| 2408 | England/20140068904/2020 | 2020-03-29 | Respiratory Virus Unit, Microbiology Services Colindale, Public Health England | Respiratory Virus Unit, Microbiology Services Colindale, Public Health England | Monica Galiano et al |
| 2409 | USA/WA-UW-4703/2020 | 2020-03-29 | UW Virology Lab | UW Virology Lab | Pavitra Roychoudhury et al |
| 2410 | Wales/PHWC-26E37/2020 | 2020-03-29 | Wales Specialist Virology Centre | Public Health Wales Microbiology Cardiff | Catherine Moore et al |
| 2411 | USA/WA-S514/2020 | 2020-03-29 | Washington State Department of Health | Seattle Flu Study | Chu et al et al |
| 2412 | USA/WA-S518/2020 | 2020-03-29 | Washington State Department of Health | Seattle Flu Study | Chu et al et al |
| 2413 | Scotland/EDB407/2020 | 2020-03-30 | Virology Department, Royal Infirmary of Edinburgh, NHS Lothian / School of Biological Sciences, University of Edinburgh / Institute of Genetics and Molecular Medicine, University of Edinburgh | COVID-19 Genomics UK (COG-UK) Consortium | McHugh M et al |
| 2414 | Australia/VIC780/2020 | 2020-03-30 | Microbiological Diagnostic Unit Public Health Laboratory | Microbiological Diagnostic Unit Public Health Laboratory | Seemann T. et al |
| 2415 | France/HF4220/2020 | 2020-03-30 | CH Jeanne de Navarre | National Reference Center for | Mélanie Albert et al |

| | | | | | |
|---|---|---|---|---|---|
| | | | Laboratoire de Biologie | Viruses of Respiratory Infections, Institut Pasteur, Paris | |
| 2416 | USA/NY-NYUMC415/2020 | 2020-03-30 | NYU Langone Health | Departments of Pathology and Medicine, New York University School of Medicine | Maria Aguero-Rosenfeld et al |
| 2417 | Belgium/ULG-10014/2020 | 2020-03-30 | Department of Clinical Microbiology | GIGA Medical Genomics | Keith Durkin et al |
| 2418 | USA/WA-S394/2020 | 2020-03-30 | Washington State Department of Health | Seattle Flu Study | Chu et al et al |
| 2419 | Australia/VIC779/2020 | 2020-03-30 | Microbiological Diagnostic Unit Public Health Laboratory | Microbiological Diagnostic Unit Public Health Laboratory | Seemann T. et al |
| 2420 | Australia/VIC734/2020 | 2020-03-30 | Victorian Infectious Diseases Reference Laboratory (VIDRL) | Microbiological Diagnostic Unit Public Health Laboratory and Victorian Infectious Diseases Reference Laboratory, Doherty Institute | Caly L. et al |
| 2421 | Australia/VIC733/2020 | 2020-03-30 | Victorian Infectious Diseases Reference Laboratory (VIDRL) | Microbiological Diagnostic Unit Public Health Laboratory and Victorian Infectious Diseases Reference Laboratory, Doherty Institute | Caly L. et al |
| 2422 | Australia/VIC753/2020 | 2020-03-30 | Victorian Infectious Diseases Reference Laboratory (VIDRL) | Microbiological Diagnostic Unit Public Health Laboratory and Victorian Infectious Diseases Reference Laboratory, Doherty Institute | Caly L. et al |
| 2423 | Australia/VIC731/2020 | 2020-03-30 | Victorian Infectious Diseases Reference Laboratory (VIDRL) | Microbiological Diagnostic Unit Public Health Laboratory and Victorian Infectious Diseases Reference Laboratory, Doherty Institute | Caly L. et al |
| 2424 | Australia/VIC931/2020 | 2020-03-30 | Victorian Infectious Diseases Reference Laboratory (VIDRL) | Microbiological Diagnostic Unit Public Health Laboratory and Victorian Infectious Diseases Reference Laboratory, Doherty Institute | Caly L. et al |
| 2425 | Australia/VIC741/2020 | 2020-03-30 | Victorian Infectious Diseases Reference Laboratory (VIDRL) | Microbiological Diagnostic Unit Public Health Laboratory and Victorian Infectious Diseases Reference Laboratory, Doherty Institute | Caly L. et al |
| 2426 | Australia/VIC982/2020 | 2020-03-30 | Victorian Infectious Diseases Reference Laboratory (VIDRL) | Microbiological Diagnostic Unit Public Health Laboratory and Victorian Infectious Diseases Reference Laboratory, The Peter Doherty Institute for Infection and Immunity | Caly L. et al |
| 2427 | Australia/VIC783/2020 | 2020-03-30 | Microbiological Diagnostic Unit Public Health Laboratory | Microbiological Diagnostic Unit Public Health Laboratory | Seemann T. et al |
| 2428 | Australia/VIC785/2020 | 2020-03-30 | Microbiological Diagnostic Unit Public Health Laboratory | Microbiological Diagnostic Unit Public Health Laboratory | Seemann T. et al |
| 2429 | Australia/VIC751/2020 | 2020-03-30 | Victorian Infectious Diseases Reference Laboratory (VIDRL) | Microbiological Diagnostic Unit Public Health Laboratory and Victorian Infectious Diseases Reference Laboratory, Doherty Institute | Caly L. et al |
| 2430 | Australia/VIC983/2020 | 2020-03-30 | Victorian Infectious Diseases Reference Laboratory (VIDRL) | Microbiological Diagnostic Unit Public Health Laboratory and Victorian Infectious Diseases Reference Laboratory, The Peter Doherty Institute for Infection and Immunity | Caly L. et al |
| 2431 | Australia/VIC752/2020 | 2020-03-30 | Victorian Infectious Diseases Reference Laboratory (VIDRL) | Microbiological Diagnostic Unit Public Health Laboratory and Victorian Infectious Diseases Reference Laboratory, Doherty Institute | Caly L. et al |
| 2432 | Australia/VIC927/2020 | 2020-03-30 | Victorian Infectious Diseases Reference Laboratory (VIDRL) | Microbiological Diagnostic Unit Public Health Laboratory and Victorian Infectious Diseases Reference Laboratory, Doherty Institute | Caly L. et al |

| | | | | | |
|---|---|---|---|---|---|
| 2433 | Scotland/CVR1022/2020 | 2020-03-30 | West of Scotland Specialist Virology Centre, NHSGGC / MRC-University of Glasgow Centre for Virus Research | COVID-19 Genomics UK (COG-UK) Consortium | Ana da Silva Filipe et al |
| 2434 | England/SHEF-CFE03/2020 | 2020-03-30 | Virology Department, Sheffield Teaching Hospitals NHS Foundation Trust / Virology Department, Sheffield Teaching Hospitals NHS Foundation Trust | COVID-19 Genomics UK (COG-UK) Consortium | Thushan de Silva et al |
| 2435 | Wales/PHWC-26981/2020 | 2020-03-30 | Wales Specialist Virology Centre | Public Health Wales Microbiology Cardiff | Catherine Moore et al |
| 2436 | USA/VA-DCLS-0086/2020 | 2020-03-30 | Virginia DCLS | Virginia DCLS | Virginia DCLS et al |
| 2437 | England/SHEF-D1F56/2020 | 2020-03-30 | Virology Department, Sheffield Teaching Hospitals NHS Foundation Trust / Virology Department, Sheffield Teaching Hospitals NHS Foundation Trust | COVID-19 Genomics UK (COG-UK) Consortium | Thushan de Silva et al |
| 2438 | USA/ID-UW-4598/2020 | 2020-03-30 | UW Virology Lab | UW Virology Lab | Pavitra Roychoudhury et al |
| 2439 | Netherlands/NA_256/2020 | 2020-03-30 | Dutch COVID-19 response team | Erasmus Medical Center | Bas Oude Munnink et al |
| 2440 | USA/WA-S395/2020 | 2020-03-30 | Washington State Department of Health | Seattle Flu Study | Chu et al et al |
| 2441 | Italy/FVG-ICGEB_S14/2020 | 2020-03-30 | Laboratory of Molecular Virology International Center for Genetic Engineering and Biotechnology (ICGEB) | ARGO Open Lab Platform for Genome Sequencing | Licastro D et al |
| 2442 | Italy/FVG-ICGEB_S18/2020 | 2020-03-30 | Laboratory of Molecular Virology International Center for Genetic Engineering and Biotechnology (ICGEB) | ARGO Open Lab Platform for Genome sequencing | Licastro D et al |
| 2443 | USA/NY-NYUMC312/2020 | 2020-03-30 | NYU Langone Health | Departments of Pathology and Medicine, New York University School of Medicine | Maria Aguero-Rosenfeld et al |
| 2444 | Belgium/ULG-10029/2020 | 2020-03-30 | Department of Clinical Microbiology | GIGA Medical Genomics | Keith Durkin et al |
| 2445 | USA/NY-NYUMC309/2020 | 2020-03-30 | NYU Langone Health | Departments of Pathology and Medicine, New York University School of Medicine | Maria Aguero-Rosenfeld et al |
| 2446 | Wales/PHWC-26EBF/2020 | 2020-03-30 | Wales Specialist Virology Centre | Public Health Wales Microbiology Cardiff | Catherine Moore et al |
| 2447 | NewZealand/20VR1857/2020-03-30 | 2020-03-30 | Southern Community Labs Dunedin | Institute of Environmental Science and Research (ESR) Ltd | **Matt Storey et al** |
| 2448 | NewZealand/20VR1803/2020-03-30 | 2020-03-30 | Waikato Hospital | Institute of Environmental Science and Research (ESR) Ltd | **Matt Storey et al** |
| 2449 | NewZealand/20VR1856/2020-03-30 | 2020-03-30 | Southern Community Labs Dunedin | Institute of Environmental Science and Research (ESR) Ltd | **Matt Storey et al** |
| 2450 | NewZealand/20VR1863/2020-03-30 | 2020-03-30 | Southern Community Labs Dunedin | Institute of Environmental Science and Research (ESR) Ltd | **Matt Storey et al** |
| 2451 | NewZealand/20VR1858/2020-03-30 | 2020-03-30 | Southern Community Labs Dunedin | Institute of Environmental Science and Research (ESR) Ltd | **Matt Storey et al** |
| 2452 | NewZealand/20VR1949/2020-03-30 | 2020-03-30 | Wellington SCL (WN) | Institute of Environmental Science and Research (ESR) Ltd | **Matt Storey et al** |
| 2453 | NewZealand/20VR1862/2020-03-30 | 2020-03-30 | Southern Community Labs Dunedin | Institute of Environmental Science and Research (ESR) Ltd | **Matt Storey et al** |
| 2454 | NewZealand/20VR1864/2020-03-30 | 2020-03-30 | Southern Community Labs Dunedin | Institute of Environmental Science and Research (ESR) Ltd | **Matt Storey et al** |
| 2455 | NewZealand/20VR2006/2020-03-30 | 2020-03-30 | Wellington SCL (WN) | Institute of Environmental Science and Research (ESR) Ltd | **Matt Storey et al** |
| 2456 | NewZealand/20VR1833/2020-03-30 | 2020-03-30 | Southern Community Labs Dunedin | Institute of Environmental Science and Research (ESR) Ltd | **Matt Storey et al** |
| 2457 | NewZealand/20VR1870/2020-03-30 | 2020-03-30 | Southern Community Labs Dunedin | Institute of Environmental Science and Research (ESR) Ltd | **Matt Storey et al** |
| 2458 | NewZealand/20VR1829/2020-03-30 | 2020-03-30 | Southern Community Labs Dunedin | Institute of Environmental Science and Research (ESR) Ltd | **Matt Storey et al** |
| 2459 | NewZealand/20VR2004/2020-03-30 | 2020-03-30 | Wellington SCL (WN) | Institute of Environmental Science and Research (ESR) Ltd | **Matt Storey et al** |

| | | | | | |
|---|---|---|---|---|---|
| 2460 | USA/WA-S435/2020 | 2020-03-30 | Washington State Department of Health | Seattle Flu Study | Chu et al et al |
| 2461 | USA/VA-DCLS-0071/2020 | 2020-03-30 | Virginia DCLS | Virginia DCLS | Virginia DCLS et al |
| 2462 | USA/WA-S571/2020 | 2020-03-30 | Washington State Department of Health | Seattle Flu Study | Chu et al et al |
| 2463 | Luxembourg/LNS0756265/2020 | 2020-03-30 | Laboratoire National de Sante, Microbiology, Virology | Laboratoire National de Sante, Microbiology, Epidemiology and Microbial Genomics | Anke Wienecke-Baldacchino et al |
| 2464 | Netherlands/NA_254/2020 | 2020-03-30 | Dutch COVID-19 response team | Erasmus Medical Center | Bas Oude Munnink et al |
| 2465 | Netherlands/NA_261/2020 | 2020-03-30 | Dutch COVID-19 response team | Erasmus Medical Center | Bas Oude Munnink et al |
| 2466 | Russia/Sverdlovsk-86704/2020 | 2020-03-30 | State Research Center of Virology and Biotechnology VECTOR, Department of Collection of Microorganisms | State Research Center of Virology and Biotechnology VECTOR, Department of Collection of Microorganisms | Sergey A. Bodnev et al |
| 2467 | USA/TX-HMH0372/2020 | 2020-03-30 | Houston Methodist Hospital | Houston Methodist Hospital | S. Wesley Long et al |
| 2468 | USA/VA-DCLS-0061/2020 | 2020-03-30 | Virginia DCLS | Virginia DCLS | Virginia DCLS et al |
| 2469 | USA/TX-HMH0402/2020 | 2020-03-30 | Houston Methodist Hospital | Houston Methodist Hospital | S. Wesley Long et al |
| 2470 | USA/NY-NYUMC409/2020 | 2020-03-30 | NYU Langone Health | Departments of Pathology and Medicine, New York University School of Medicine | Maria Aguero-Rosenfeld et al |
| 2471 | USA/WI-106/2020 | 2020-03-30 | University of Wisconsin-Madison AIDS Vaccine Research Laboratories | University of Wisconsin-Madison AIDS Vaccine Research Laboratories | Moreno et al |
| 2472 | England/20142016504/2020 | 2020-03-30 | Respiratory Virus Unit, Microbiology Services Colindale, Public Health England | Respiratory Virus Unit, Microbiology Services Colindale, Public Health England | Monica Galiano et al |
| 2473 | USA/VA-DCLS-0049/2020 | 2020-03-30 | Virginia DCLS | Virginia DCLS | Virginia DCLS et al |
| 2474 | England/20144018204/2020 | 2020-03-30 | Respiratory Virus Unit, Microbiology Services Colindale, Public Health England | Respiratory Virus Unit, Microbiology Services Colindale, Public Health England | Monica Galiano et al |
| 2475 | England/NOTT-10EAC7/2020 | 2020-03-30 | Queens Medical Centre, Clinical Microbiology Department / DeepSeq Nottingham | COVID-19 Genomics UK (COG-UK) Consortium | Gemma Clark et al |
| 2476 | USA/WA-UW-4565/2020 | 2020-03-30 | UW Virology Lab | UW Virology Lab | Pavitra Roychoudhury et al |
| 2477 | USA/WA-S581/2020 | 2020-03-30 | Washington State Department of Health | Seattle Flu Study | Chu et al et al |
| 2478 | USA/TX-HMH0365/2020 | 2020-03-30 | Houston Methodist Hospital | Houston Methodist Hospital | S. Wesley Long et al |
| 2479 | USA/CT-UW-4348/2020 | 2020-03-30 | UW Virology Lab | UW Virology Lab | Pavitra Roychoudhury et al |
| 2480 | USA/CT-UW-4344/2020 | 2020-03-30 | UW Virology Lab | UW Virology Lab | Pavitra Roychoudhury et al |
| 2481 | Sweden/20-06503/2020 | 2020-03-30 | Trollbackens VC | The Public Health Agency of Sweden | Amelie Holmqvist et al |
| 2482 | Taiwan/NTU26/2020 | 2020-03-30 | Department of Laboratory Medicine, National Taiwan University Hospital | Microbial Genomics Core Lab, National Taiwan University Centers of Genomic and Precision Medicine | Shiou-Hwei Yeh et al |
| 2483 | Taiwan/TSGH-21/2020 | 2020-03-30 | TSGH-CP molecular lab | TSGH-CP molecular lab | Cherng-Lih Perng et al |
| 2484 | Taiwan/284/2020 | 2020-03-30 | Centers for Disease Control, R.O.C. (Taiwan) | Centers for Disease Control, R.O.C. (Taiwan) | Ji-Rong Yang et al |
| 2485 | USA/NY-NYUMC307/2020 | 2020-03-30 | NYU Langone Health | Departments of Pathology and Medicine, New York University School of Medicine | Maria Aguero-Rosenfeld et al |
| 2486 | Wales/PHWC-26FCB/2020 | 2020-03-30 | Wales Specialist Virology Centre | Public Health Wales Microbiology Cardiff | Catherine Moore et al |
| 2487 | England/20140070504/2020 | 2020-03-30 | Respiratory Virus Unit, Microbiology Services Colindale, Public Health England | Respiratory Virus Unit, Microbiology Services Colindale, Public Health England | Monica Galiano et al |
| 2488 | USA/NY-NYUMC311/2020 | 2020-03-30 | NYU Langone Health | Departments of Pathology and Medicine, New York University School of Medicine | Maria Aguero-Rosenfeld et al |

| | | | | | |
|---|---|---|---|---|---|
| 2489 | England/20142019704/2020 | 2020-03-30 | Respiratory Virus Unit, Microbiology Services Colindale, Public Health England | Respiratory Virus Unit, Microbiology Services Colindale, Public Health England | Monica Galiano et al |
| 2490 | USA/WI-116/2020 | 2020-03-30 | University of Wisconsin-Madison AIDS Vaccine Research Laboratories | University of Wisconsin-Madison AIDS Vaccine Research Laboratories | Moreno et al |
| 2491 | Australia/VIC989/2020 | 2020-03-31 | Victorian Infectious Diseases Reference Laboratory (VIDRL) | Microbiological Diagnostic Unit Public Health Laboratory and Victorian Infectious Diseases Reference Laboratory, The Peter Doherty Institute for Infection and Immunity | Caly L. et al |
| 2492 | Australia/VIC796/2020 | 2020-03-31 | Microbiological Diagnostic Unit Public Health Laboratory | Microbiological Diagnostic Unit Public Health Laboratory | Seemann T. et al |
| 2493 | Australia/VIC932/2020 | 2020-03-31 | Victorian Infectious Diseases Reference Laboratory (VIDRL) | Microbiological Diagnostic Unit Public Health Laboratory and Victorian Infectious Diseases Reference Laboratory, Doherty Institute | Caly L. et al |
| 2494 | Netherlands/NA_284/2020 | 2020-03-31 | Dutch COVID-19 response team | Erasmus Medical Center | Bas Oude Munnink et al |
| 2495 | Netherlands/ZuidHolland_97/2020 | 2020-03-31 | Dutch COVID-19 response team | Erasmus Medical Center | Bas Oude Munnink et al |
| 2496 | USA/WA-UW-4281/2020 | 2020-03-31 | UW Virology Lab | UW Virology Lab | Pavitra Roychoudhury et al |
| 2497 | USA/WA-S436/2020 | 2020-03-31 | Washington State Department of Health | Seattle Flu Study | Chu et al et al |
| 2498 | Australia/VIC747/2020 | 2020-03-31 | Victorian Infectious Diseases Reference Laboratory (VIDRL) | Microbiological Diagnostic Unit Public Health Laboratory and Victorian Infectious Diseases Reference Laboratory, Doherty Institute | Caly L. et al |
| 2499 | Australia/VIC990/2020 | 2020-03-31 | Victorian Infectious Diseases Reference Laboratory (VIDRL) | Microbiological Diagnostic Unit Public Health Laboratory and Victorian Infectious Diseases Reference Laboratory, The Peter Doherty Institute for Infection and Immunity | Caly L. et al |
| 2500 | Australia/VIC995/2020 | 2020-03-31 | Victorian Infectious Diseases Reference Laboratory (VIDRL) | Microbiological Diagnostic Unit Public Health Laboratory and Victorian Infectious Diseases Reference Laboratory, The Peter Doherty Institute for Infection and Immunity | Caly L. et al |
| 2501 | USA/LA-SR0164/2020 | 2020-03-31 | Robert Garry lab | Andersen lab at Scripps Research | Allison Smither et al |
| 2502 | Belgium/ULG-10030/2020 | 2020-03-31 | Department of Clinical Microbiology | GIGA Medical Genomics | Keith Durkin et al |
| 2503 | Singapore/40/2020 | 2020-03-31 | National Public Health Laboratory, National Centre for Infectious Diseases | National Public Health Laboratory, National Centre for Infectious Diseases | Mak TM et al |
| 2504 | USA/TX-HMH0375/2020 | 2020-03-31 | Houston Methodist Hospital | Houston Methodist Hospital | S. Wesley Long et al |
| 2505 | Wales/PHWC-27759/2020 | 2020-03-31 | Wales Specialist Virology Centre | Public Health Wales Microbiology Cardiff | Catherine Moore et al |
| 2506 | England/NOTT-10F12B/2020 | 2020-03-31 | Queens Medical Centre, Clinical Microbiology Department / DeepSeq Nottingham | COVID-19 Genomics UK (COG-UK) Consortium | Gemma Clark et al |
| 2507 | Netherlands/NA_285/2020 | 2020-03-31 | Dutch COVID-19 response team | Erasmus Medical Center | Bas Oude Munnink et al |
| 2508 | Netherlands/NA_276/2020 | 2020-03-31 | Dutch COVID-19 response team | Erasmus Medical Center | Bas Oude Munnink et al |
| 2509 | Belgium/ULG-10026/2020 | 2020-03-31 | Department of Clinical Microbiology | GIGA Medical Genomics | Keith Durkin et al |
| 2510 | USA/OH-OSUP0016/2020 | 2020-03-31 | The Ohio State University | The Ohio State University-James Molecular Lab at Polaris | Huolin Tu et al |
| 2511 | USA/WA-S526/2020 | 2020-03-31 | Washington State Department of Health | Seattle Flu Study | Chu et al et al |

| | | | | | |
|---|---|---|---|---|---|
| 2512 | USA/WA-UW-4321/2020 | 2020-03-31 | UW Virology Lab | UW Virology Lab | Pavitra Roychoudhury et al |
| 2513 | England/20144014504/2020 | 2020-03-31 | Respiratory Virus Unit, Microbiology Services Colindale, Public Health England | Respiratory Virus Unit, Microbiology Services Colindale, Public Health England | Monica Galiano et al |
| 2514 | NewZealand/20VR1985/2020-03-31 | 2020-03-31 | Canterbury Health Laboratories | Institute of Environmental Science and Research (ESR) Ltd | **Matt Storey et al** |
| 2515 | NewZealand/20VR1899/2020-03-31 | 2020-03-31 | LabPLUS | Institute of Environmental Science and Research (ESR) Ltd | **Matt Storey et al** |
| 2516 | NewZealand/20VR1867/2020-03-31 | 2020-03-31 | Southern Community Labs Dunedin | Institute of Environmental Science and Research (ESR) Ltd | **Matt Storey et al** |
| 2517 | NewZealand/20VR1860/2020-03-31 | 2020-03-31 | Southern Community Labs Dunedin | Institute of Environmental Science and Research (ESR) Ltd | **Matt Storey et al** |
| 2518 | NewZealand/20VR1917/2020-03-31 | 2020-03-31 | Southern Community Labs Dunedin | Institute of Environmental Science and Research (ESR) Ltd | **Matt Storey et al** |
| 2519 | NewZealand/20VR2017/2020-03-31 | 2020-03-31 | Wellington SCL (WN) | Institute of Environmental Science and Research (ESR) Ltd | **Matt Storey et al** |
| 2520 | Russia/Khabarovsk-88201/2020 | 2020-03-31 | State Research Center of Virology and Biotechnology VECTOR, Department of Collection of Microorganisms | State Research Center of Virology and Biotechnology VECTOR, Department of Collection of Microorganisms | Oleg V. Pyankov et al |
| 2521 | England/20144000404/2020 | 2020-03-31 | Respiratory Virus Unit, Microbiology Services Colindale, Public Health England | Respiratory Virus Unit, Microbiology Services Colindale, Public Health England | Monica Galiano et al |
| 2522 | USA/TX-HMH0399/2020 | 2020-03-31 | Houston Methodist Hospital | Houston Methodist Hospital | S. Wesley Long et al |
| 2523 | Singapore/42/2020 | 2020-03-31 | National Public Health Laboratory, National Centre for Infectious Diseases | National Public Health Laboratory, National Centre for Infectious Diseases | Mak TM et al |
| 2524 | USA/WA-UW-4407/2020 | 2020-03-31 | UW Virology Lab | UW Virology Lab | Pavitra Roychoudhury et al |
| 2525 | USA/WA-S437/2020 | 2020-03-31 | Washington State Department of Health | Seattle Flu Study | Chu et al et al |
| 2526 | USA/TX-HMH0412/2020 | 2020-03-31 | Houston Methodist Hospital | Houston Methodist Hospital | S. Wesley Long et al |
| 2527 | Belgium/ULG-10031/2020 | 2020-03-31 | Department of Clinical Microbiology | GIGA Medical Genomics | Keith Durkin et al |
| 2528 | USA/WA-S452/2020 | 2020-03-31 | Washington State Department of Health | Seattle Flu Study | Chu et al et al |
| 2529 | USA/TX-HMH0409/2020 | 2020-03-31 | Houston Methodist Hospital | Houston Methodist Hospital | S. Wesley Long et al |
| 2530 | USA/WI-101/2020 | 2020-03-31 | University of Wisconsin-Madison AIDS Vaccine Research Laboratories | University of Wisconsin-Madison AIDS Vaccine Research Laboratories | Moreno et al |
| 2531 | USA/TX-HMH0403/2020 | 2020-03-31 | Houston Methodist Hospital | Houston Methodist Hospital | S. Wesley Long et al |
| 2532 | Netherlands/NA_288/2020 | 2020-03-31 | Dutch COVID-19 response team | Erasmus Medical Center | Bas Oude Munnink et al |
| 2533 | Wales/PHWC-27786/2020 | 2020-03-31 | Wales Specialist Virology Centre | Public Health Wales Microbiology Cardiff | Catherine Moore et al |
| 2534 | USA/CA-CZB0740/2020 | 2020-03-31 | UCSF Clinical Microbiology Laboratory | Chan-Zuckerberg Biohub | CZB Cliahub Consortium et al |
| 2535 | USA/WA-S299/2020 | 2020-03-31 | Seattle Flu Study | Seattle Flu Study | Chu et al et al |
| 2536 | USA/WA-S520/2020 | 2020-03-31 | Washington State Department of Health | Seattle Flu Study | Chu et al et al |
| 2537 | Netherlands/ZuidHolland_99/2020 | 2020-03-31 | Dutch COVID-19 response team | Erasmus Medical Center | Bas Oude Munnink et al |
| 2538 | Wales/PHWC-27689/2020 | 2020-03-31 | Wales Specialist Virology Centre | Public Health Wales Microbiology Cardiff | Catherine Moore et al |
| 2539 | USA/WA-S528/2020 | 2020-03-31 | Washington State Department of Health | Seattle Flu Study | Chu et al et al |
| 2540 | USA/VA-DCLS-0060/2020 | 2020-03-31 | Virginia DCLS | Virginia DCLS | Virginia DCLS et al |
| 2541 | USA/WI-107/2020 | 2020-03-31 | University of Wisconsin-Madison AIDS Vaccine Research Laboratories | University of Wisconsin-Madison AIDS Vaccine Research Laboratories | Moreno et al |
| 2542 | USA/CA-CZB-1033/2020 | 2020-03-31 | County of Santa Clara Public Health Department | Chan-Zuckerberg Biohub | CZB Cliahub Consortium et al |
| 2543 | USA/WA-UW-4318/2020 | 2020-03-31 | UW Virology Lab | UW Virology Lab | Pavitra Roychoudhury et al |

| | | | | | |
|---|---|---|---|---|---|
| 2544 | USA/CA-CZB-1000/2020 | 2020-03-31 | County of Santa Clara Public Health Department | Chan-Zuckerberg Biohub | CZB Cliahub Consortium et al |
| 2545 | USA/TX-HMH0401/2020 | 2020-03-31 | Houston Methodist Hospital | Houston Methodist Hospital | S. Wesley Long et al |
| 2546 | Taiwan/TSGH-22/2020 | 2020-03-31 | TSGH-CP molecular lab | TSGH-CP molecular lab | Cherng-Lih Perng et al |
| 2547 | USA/WA-S545/2020 | 2020-03-31 | Washington State Department of Health | Seattle Flu Study | Chu et al et al |
| 2548 | USA/WA-UW-4594/2020 | 2020-03-31 | UW Virology Lab | UW Virology Lab | Pavitra Roychoudhury et al |
| 2549 | USA/WA-S454/2020 | 2020-03-31 | Washington State Department of Health | Seattle Flu Study | Chu et al et al |
| 2550 | USA/AZ-TG273300/2020 | 2020-03-31 | AZ SPHL, Arizona Department of Health Services | TGen North | Jolene Bowers et al |
| 2551 | USA/CA-CZB0737/2020 | 2020-03-31 | UCSF Clinical Microbiology Laboratory | Chan-Zuckerberg Biohub | CZB Cliahub Consortium et al |
| 2552 | USA/WI-113/2020 | 2020-03-31 | University of Wisconsin-Madison AIDS Vaccine Research Laboratories | University of Wisconsin-Madison AIDS Vaccine Research Laboratories | Moreno et al |
| 2553 | England/CAMB-739EF/2020 | 2020-03-31 | Department of Pathology, University of Cambridge | COVID-19 Genomics UK (COG-UK) Consortium | Luke W Meredith et al |
| 2554 | USA/VA-DCLS-0057/2020 | 2020-03-31 | Virginia DCLS | Virginia DCLS | Virginia DCLS et al |
| 2555 | Wales/PHWC-273CE/2020 | 2020-03-31 | Wales Specialist Virology Centre | Public Health Wales Microbiology Cardiff | Catherine Moore et al |
| 2556 | USA/WA-UW-4593/2020 | 2020-03-31 | UW Virology Lab | UW Virology Lab | Pavitra Roychoudhury et al |
| 2557 | USA/WA-UW-4370/2020 | 2020-03-31 | UW Virology Lab | UW Virology Lab | Pavitra Roychoudhury et al |
| 2558 | USA/WA-S538/2020 | 2020-03-31 | Washington State Department of Health | Seattle Flu Study | Chu et al et al |
| 2559 | England/CAMB-76EDE/2020 | 2020-03-31 | Department of Pathology, University of Cambridge | COVID-19 Genomics UK (COG-UK) Consortium | Luke W Meredith et al |
| 2560 | Scotland/CVR1358/2020 | 2020-03-31 | West of Scotland Specialist Virology Centre, NHSGGC / MRC-University of Glasgow Centre for Virus Research | COVID-19 Genomics UK (COG-UK) Consortium | Ana da Silva Filipe et al |
| 2561 | USA/WA-S557/2020 | 2020-04-01 | Washington State Department of Health | Seattle Flu Study | Chu et al et al |
| 2562 | Australia/VIC800/2020 | 2020-04-01 | Microbiological Diagnostic Unit Public Health Laboratory | Microbiological Diagnostic Unit Public Health Laboratory | Seemann T. et al |
| 2563 | Australia/VIC1011/2020 | 2020-04-01 | Victorian Infectious Diseases Reference Laboratory (VIDRL) | Microbiological Diagnostic Unit Public Health Laboratory and Victorian Infectious Diseases Reference Laboratory, The Peter Doherty Institute for Infection and Immunity | Caly L. et al |
| 2564 | USA/CA-CZB-1034/2020 | 2020-04-01 | County of Santa Clara Public Health Department | Chan-Zuckerberg Biohub | CZB Cliahub Consortium et al |
| 2565 | Argentina/PAIS_A013/2020 | 2020-04-01 | Laboratorio de Virología del Hospital de Niños Dr. Ricardo Gutierrez | Área de Secuenciación del Laboratorio de Virología del Hospital de Niños Dr. Ricardo Gutierrez | Nabaes Jodar et al |
| 2566 | Netherlands/NA_293/2020 | 2020-04-01 | Dutch COVID-19 response team | Erasmus Medical Center | Bas Oude Munnink et al |
| 2567 | Scotland/EDB625/2020 | 2020-04-01 | Virology Department, Royal Infirmary of Edinburgh, NHS Lothian / School of Biological Sciences, University of Edinburgh / Institute of Genetics and Molecular Medicine, University of Edinburgh | COVID-19 Genomics UK (COG-UK) Consortium | McHugh M et al |
| 2568 | England/CAMB-76FCC/2020 | 2020-04-01 | Department of Pathology, University of Cambridge | COVID-19 Genomics UK (COG-UK) Consortium | Luke W Meredith et al |
| 2569 | USA/CA-CZB-1001/2020 | 2020-04-01 | County of Santa Clara Public Health Department | Chan-Zuckerberg Biohub | CZB Cliahub Consortium et al |
| 2570 | USA/NY-NYUMC145/2020 | 2020-04-01 | NYU Langone Health | Departments of Pathology and Medicine, New York University School of Medicine | Maria Aguero-Rosenfeld et al |
| 2571 | Wales/PHWC-27D39/2020 | 2020-04-01 | Wales Specialist Virology Centre | Public Health Wales Microbiology Cardiff | Catherine Moore et al |

| | | | | | |
|---|---|---|---|---|---|
| 2572 | England/SHEF-C18E5/2020 | 2020-04-01 | Virology Department, Sheffield Teaching Hospitals NHS Foundation Trust / Virology Department, Sheffield Teaching Hospitals NHS Foundation Trust | COVID-19 Genomics UK (COG-UK) Consortium | Thushan de Silva et al |
| 2573 | USA/TX-HMH0425/2020 | 2020-04-01 | Houston Methodist Hospital | Houston Methodist Hospital | S. Wesley Long et al |
| 2574 | Scotland/EDB530/2020 | 2020-04-01 | Virology Department, Royal Infirmary of Edinburgh, NHS Lothian / School of Biological Sciences, University of Edinburgh / Institute of Genetics and Molecular Medicine, University of Edinburgh | COVID-19 Genomics UK (COG-UK) Consortium | McHugh M et al |
| 2575 | USA/NY-NYUMC134/2020 | 2020-04-01 | NYU Langone Health | Departments of Pathology and Medicine, New York University School of Medicine | Maria Aguero-Rosenfeld et al |
| 2576 | USA/WA-S474/2020 | 2020-04-01 | Washington State Department of Health | Seattle Flu Study | Chu et al et al |
| 2577 | USA/CA-CZB-1032/2020 | 2020-04-01 | County of Santa Clara Public Health Department | Chan-Zuckerberg Biohub | CZB Cliahub Consortium et al |
| 2578 | Wales/PHWC-27B5D/2020 | 2020-04-01 | Wales Specialist Virology Centre | Public Health Wales Microbiology Cardiff | Catherine Moore et al |
| 2579 | USA/WA-UW-5177/2020 | 2020-04-01 | UW Virology Lab | UW Virology Lab | Pavitra Roychoudhury et al |
| 2580 | USA/NY-NYUMC188/2020 | 2020-04-01 | NYU Langone Health | Departments of Pathology and Medicine, New York University School of Medicine | Maria Aguero-Rosenfeld et al |
| 2581 | USA/NY-NYUMC123/2020 | 2020-04-01 | NYU Langone Health | Departments of Pathology and Medicine, New York University School of Medicine | Maria Aguero-Rosenfeld et al |
| 2582 | USA/CA-CZB-1035/2020 | 2020-04-01 | County of Santa Clara Public Health Department | Chan-Zuckerberg Biohub | CZB Cliahub Consortium et al |
| 2583 | USA/NY-NYUMC156/2020 | 2020-04-01 | NYU Langone Health | Departments of Pathology and Medicine, New York University School of Medicine | Maria Aguero-Rosenfeld et al |
| 2584 | Wales/PHWC-27CA5/2020 | 2020-04-01 | Wales Specialist Virology Centre | Public Health Wales Microbiology Cardiff | Catherine Moore et al |
| 2585 | NewZealand/20VR1894/2020-04-01 | 2020-04-01 | Southern Community Labs Dunedin | Institute of Environmental Science and Research (ESR) Ltd | **Matt Storey et al** |
| 2586 | NewZealand/20VR1908/2020-04-01 | 2020-04-01 | Southern Community Labs Dunedin | Institute of Environmental Science and Research (ESR) Ltd | **Matt Storey et al** |
| 2587 | NewZealand/20VR1890/2020-04-01 | 2020-04-01 | Southern Community Labs Dunedin | Institute of Environmental Science and Research (ESR) Ltd | **Matt Storey et al** |
| 2588 | NewZealand/20VR1896/2020-04-01 | 2020-04-01 | Southern Community Labs Dunedin | Institute of Environmental Science and Research (ESR) Ltd | **Matt Storey et al** |
| 2589 | NewZealand/20VR1891/2020-04-01 | 2020-04-01 | Southern Community Labs Dunedin | Institute of Environmental Science and Research (ESR) Ltd | **Matt Storey et al** |
| 2590 | NewZealand/20VR2032/2020-04-01 | 2020-04-01 | Wellington SCL | Institute of Environmental Science and Research (ESR) | **Matt Storey et al** |
| 2591 | NewZealand/20VR2001/2020-04-01 | 2020-04-01 | Wellington SCL | Institute of Environmental Science and Research (ESR) | **Matt Storey et al** |
| 2592 | NewZealand/20VR2022/2020-04-01 | 2020-04-01 | Wellington SCL | Institute of Environmental Science and Research (ESR) | **Matt Storey et al** |
| 2593 | NewZealand/20VR1897/2020-04-01 | 2020-04-01 | Southern Community Labs Dunedin | Institute of Environmental Science and Research (ESR) Ltd | **Matt Storey et al** |
| 2594 | NewZealand/20VR2011/2020-04-01 | 2020-04-01 | Wellington SCL | Institute of Environmental Science and Research (ESR) | **Matt Storey et al** |
| 2595 | NewZealand/20VR2019/2020-04-01 | 2020-04-01 | Wellington SCL | Institute of Environmental Science and Research (ESR) | **Matt Storey et al** |
| 2596 | NewZealand/20VR1893/2020-04-01 | 2020-04-01 | Southern Community Labs Dunedin | Institute of Environmental Science and Research (ESR) Ltd | **Matt Storey et al** |
| 2597 | NewZealand/20VR1937/2020-04-01 | 2020-04-01 | Southern Community Labs Dunedin | Institute of Environmental Science and Research (ESR) Ltd | **Matt Storey et al** |
| 2598 | NewZealand/20VR2035/2020-04-01 | 2020-04-01 | Wellington SCL | Institute of Environmental Science and Research (ESR) | **Matt Storey et al** |
| 2599 | NewZealand/20VR2014/2020-04-01 | 2020-04-01 | Wellington SCL | Institute of Environmental Science and Research (ESR) | **Matt Storey et al** |

| | | | | | |
|---|---|---|---|---|---|
| 2600 | NewZealand/20VR1995/2020-04-01 | 2020-04-01 | Wellington SCL | Institute of Environmental Science and Research (ESR) | **Matt Storey et al** |
| 2601 | NewZealand/20VR2037/2020-04-01 | 2020-04-01 | Wellington SCL | Institute of Environmental Science and Research (ESR) | **Matt Storey et al** |
| 2602 | USA/TX-HMH0417/2020 | 2020-04-01 | Houston Methodist Hospital | Houston Methodist Hospital | S. Wesley Long et al |
| 2603 | USA/NY-NYUMC126/2020 | 2020-04-01 | NYU Langone Health | Departments of Pathology and Medicine, New York University School of Medicine | Maria Aguero-Rosenfeld et al |
| 2604 | Scotland/CVR1443/2020 | 2020-04-01 | West of Scotland Specialist Virology Centre, NHSGGC / MRC-University of Glasgow Centre for Virus Research | COVID-19 Genomics UK (COG-UK) Consortium | Ana da Silva Filipe et al |
| 2605 | USA/WA-UW-5203/2020 | 2020-04-01 | UW Virology Lab | UW Virology Lab | Pavitra Roychoudhury et al |
| 2606 | USA/AZ-TG272215/2020 | 2020-04-01 | TGen North | TGen North | Jolene Bowers et al |
| 2607 | Wales/PHWC-27B6C/2020 | 2020-04-01 | Wales Specialist Virology Centre | Public Health Wales Microbiology Cardiff | Catherine Moore et al |
| 2608 | USA/NY-NYUMC180/2020 | 2020-04-01 | NYU Langone Health | Departments of Pathology and Medicine, New York University School of Medicine | Maria Aguero-Rosenfeld et al |
| 2609 | USA/NY-NYUMC325/2020 | 2020-04-01 | NYU Langone Health | Departments of Pathology and Medicine, New York University School of Medicine | Maria Aguero-Rosenfeld et al |
| 2610 | USA/WA-S478/2020 | 2020-04-01 | Washington State Department of Health | Seattle Flu Study | Chu et al et al |
| 2611 | USA/NY-NYUMC164/2020 | 2020-04-01 | NYU Langone Health | Departments of Pathology and Medicine, New York University School of Medicine | Maria Aguero-Rosenfeld et al |
| 2612 | USA/TX-HMH0400/2020 | 2020-04-01 | Houston Methodist Hospital | Houston Methodist Hospital | S. Wesley Long et al |
| 2613 | Netherlands/NA_302/2020 | 2020-04-01 | Dutch COVID-19 response team | Erasmus Medical Center | Bas Oude Munnink et al |
| 2614 | Netherlands/NA_300/2020 | 2020-04-01 | Dutch COVID-19 response team | Erasmus Medical Center | Bas Oude Munnink et al |
| 2615 | USA/WA-S495/2020 | 2020-04-01 | Washington State Department of Health | Seattle Flu Study | Chu et al et al |
| 2616 | England/NOTT-10F2FB/2020 | 2020-04-01 | Queens Medical Centre, Clinical Microbiology Department / DeepSeq Nottingham | COVID-19 Genomics UK (COG-UK) Consortium | Gemma Clark et al |
| 2617 | USA/NY-NYUMC167/2020 | 2020-04-01 | NYU Langone Health | Departments of Pathology and Medicine, New York University School of Medicine | Maria Aguero-Rosenfeld et al |
| 2618 | Netherlands/NA_297/2020 | 2020-04-01 | Dutch COVID-19 response team | Erasmus Medical Center | Bas Oude Munnink et al |
| 2619 | USA/WA-S312/2020 | 2020-04-01 | Washington State Department of Health | Seattle Flu Study | Chu et al et al |
| 2620 | USA/VI-CDC-1993/2020 | 2020-04-01 | US VI Department of Health | Pathogen Discovery, Respiratory Viruses Branch, Division of Viral Diseases, Centers for Disease Control and Prevention | Ying Tao et al |
| 2621 | USA/WA-S326/2020 | 2020-04-01 | Washington State Department of Health | Seattle Flu Study | Chu et al et al |
| 2622 | USA/CA-CZB-1004/2020 | 2020-04-01 | County of Santa Clara Public Health Department | Chan-Zuckerberg Biohub | CZB Cliahub Consortium et al |
| 2623 | USA/WA-S324/2020 | 2020-04-01 | Washington State Department of Health | Seattle Flu Study | Chu et al et al |
| 2624 | USA/CT-UW-6271/2020 | 2020-04-01 | UW Virology Lab | UW Virology Lab | Pavitra Roychoudhury et al |
| 2625 | England/CAMB-76EB0/2020 | 2020-04-01 | Department of Pathology, University of Cambridge | COVID-19 Genomics UK (COG-UK) Consortium | Luke W Meredith et al |
| 2626 | USA/NY-NYUMC199/2020 | 2020-04-01 | NYU Langone Health | Departments of Pathology and Medicine, New York University School of Medicine | Maria Aguero-Rosenfeld et al |
| 2627 | Wales/PHWC-27CD2/2020 | 2020-04-01 | Wales Specialist Virology Centre | Public Health Wales Microbiology Cardiff | Catherine Moore et al |
| 2628 | USA/TX-HMH0382/2020 | 2020-04-01 | Houston Methodist Hospital | Houston Methodist Hospital | S. Wesley Long et al |
| 2629 | USA/NY-NYUMC173/2020 | 2020-04-01 | NYU Langone Health | Departments of Pathology and Medicine, New York University | Maria Aguero-Rosenfeld et |

| | | | | | |
|---|---|---|---|---|---|
| | | | | School of Medicine | al |
| 2630 | USA/NY-NYUMC179/2020 | 2020-04-01 | NYU Langone Health | Departments of Pathology and Medicine, New York University School of Medicine | Maria Aguero-Rosenfeld et al |
| 2631 | USA/VA-DCLS-0056/2020 | 2020-04-01 | Virginia DCLS | Virginia DCLS | Virginia DCLS et al |
| 2632 | USA/CA-CZB-1002/2020 | 2020-04-01 | County of Santa Clara Public Health Department | Chan-Zuckerberg Biohub | CZB Cliahub Consortium et al |
| 2633 | USA/NY-NYUMC153/2020 | 2020-04-01 | NYU Langone Health | Departments of Pathology and Medicine, New York University School of Medicine | Maria Aguero-Rosenfeld et al |
| 2634 | USA/WA-S465/2020 | 2020-04-01 | Washington State Department of Health | Seattle Flu Study | Chu et al et al |
| 2635 | England/SHEF-C1645/2020 | 2020-04-01 | Virology Department, Sheffield Teaching Hospitals NHS Foundation Trust / Virology Department, Sheffield Teaching Hospitals NHS Foundation Trust | COVID-19 Genomics UK (COG-UK) Consortium | Thushan de Silva et al |
| 2636 | Taiwan/TSGH-34/2020 | 2020-04-01 | TSGH-CP molecular lab | TSGH-CP molecular lab | Cherng-Lih Perng et al |
| 2637 | USA/NY-NYUMC193/2020 | 2020-04-01 | NYU Langone Health | Departments of Pathology and Medicine, New York University School of Medicine | Maria Aguero-Rosenfeld et al |
| 2638 | USA/NY-NYUMC143/2020 | 2020-04-01 | NYU Langone Health | Departments of Pathology and Medicine, New York University School of Medicine | Maria Aguero-Rosenfeld et al |
| 2639 | Wales/PHWC-2757D/2020 | 2020-04-01 | Wales Specialist Virology Centre | Public Health Wales Microbiology Cardiff | Catherine Moore et al |
| 2640 | USA/NY-NYUMC326/2020 | 2020-04-01 | NYU Langone Health | Departments of Pathology and Medicine, New York University School of Medicine | Maria Aguero-Rosenfeld et al |
| 2641 | Netherlands/NA_294/2020 | 2020-04-01 | Dutch COVID-19 response team | Erasmus Medical Center | Bas Oude Munnink et al |
| 2642 | USA/WA-S485/2020 | 2020-04-01 | Washington State Department of Health | Seattle Flu Study | Chu et al et al |
| 2643 | USA/NY-NYUMC184/2020 | 2020-04-01 | NYU Langone Health | Departments of Pathology and Medicine, New York University School of Medicine | Maria Aguero-Rosenfeld et al |
| 2644 | Scotland/CVR1414/2020 | 2020-04-02 | West of Scotland Specialist Virology Centre, NHSGGC / MRC-University of Glasgow Centre for Virus Research | COVID-19 Genomics UK (COG-UK) Consortium | Ana da Silva Filipe et al |
| 2645 | USA/VA-DCLS-0064/2020 | 2020-04-02 | Virginia DCLS | Virginia DCLS | Virginia DCLS et al |
| 2646 | Scotland/CVR1429/2020 | 2020-04-02 | West of Scotland Specialist Virology Centre, NHSGGC / MRC-University of Glasgow Centre for Virus Research | COVID-19 Genomics UK (COG-UK) Consortium | Ana da Silva Filipe et al |
| 2647 | Australia/VIC812/2020 | 2020-04-02 | Victorian Infectious Diseases Reference Laboratory (VIDRL) | Microbiological Diagnostic Unit Public Health Laboratory and Victorian Infectious Diseases Reference Laboratory, Doherty Institute | Caly L. et al |
| 2648 | England/CAMB-73C04/2020 | 2020-04-02 | Department of Pathology, University of Cambridge | COVID-19 Genomics UK (COG-UK) Consortium | Luke W Meredith et al |
| 2649 | USA/WA-UW-6335/2020 | 2020-04-02 | UW Virology Lab | UW Virology Lab | Pavitra Roychoudhury et al |
| 2650 | Netherlands/NA_316/2020 | 2020-04-02 | Dutch COVID-19 response team | Erasmus Medical Center | Bas Oude Munnink et al |
| 2651 | USA/WA-S462/2020 | 2020-04-02 | Washington State Department of Health | Seattle Flu Study | Chu et al et al |
| 2652 | USA/TX-HMH0432/2020 | 2020-04-02 | Houston Methodist Hospital | Houston Methodist Hospital | S. Wesley Long et al |
| 2653 | Netherlands/NA_317/2020 | 2020-04-02 | Dutch COVID-19 response team | Erasmus Medical Center | Bas Oude Munnink et al |
| 2654 | USA/UN-UW-5199/2020 | 2020-04-02 | UW Virology Lab | UW Virology Lab | Pavitra Roychoudhury et al |
| 2655 | USA/WA-UW-5200/2020 | 2020-04-02 | UW Virology Lab | UW Virology Lab | Pavitra Roychoudhury et al |
| 2656 | USA/WI-86/2020 | 2020-04-02 | University of Wisconsin-Madison AIDS Vaccine Research Laboratories | University of Wisconsin-Madison AIDS Vaccine Research Laboratories | Moreno et al |

| | | | | | |
|---|---|---|---|---|---|
| 2657 | USA/WA-S332/2020 | 2020-04-02 | Washington State Department of Health | Seattle Flu Study | Chu et al et al |
| 2658 | USA/VA-DCLS-0091/2020 | 2020-04-02 | Virginia DCLS | Virginia DCLS | Virginia DCLS et al |
| 2659 | USA/TX-HMH0427/2020 | 2020-04-02 | Houston Methodist Hospital | Houston Methodist Hospital | S. Wesley Long et al |
| 2660 | USA/WA-UW-5152/2020 | 2020-04-02 | UW Virology Lab | UW Virology Lab | Pavitra Roychoudhury et al |
| 2661 | Scotland/EDB726/2020 | 2020-04-02 | Virology Department, Royal Infirmary of Edinburgh, NHS Lothian / School of Biological Sciences, University of Edinburgh / Institute of Genetics and Molecular Medicine, University of Edinburgh | COVID-19 Genomics UK (COG-UK) Consortium | McHugh M et al |
| 2662 | USA/WA-UW-5205/2020 | 2020-04-02 | UW Virology Lab | UW Virology Lab | Pavitra Roychoudhury et al |
| 2663 | USA/WA-S343/2020 | 2020-04-02 | Washington State Department of Health | Seattle Flu Study | Chu et al et al |
| 2664 | Switzerland/GE0304/2020 | 2020-04-02 | University Hospitals of Geneva Laboratory of Virology | University Hospitals of Geneva Laboratory of Virology | Laubscher F. et al |
| 2665 | USA/WA-S349/2020 | 2020-04-02 | Washington State Department of Health | Seattle Flu Study | Chu et al et al |
| 2666 | USA/WA-S439/2020 | 2020-04-02 | Washington State Department of Health | Seattle Flu Study | Chu et al et al |
| 2667 | USA/VA-DCLS-0089/2020 | 2020-04-02 | Virginia DCLS | Virginia DCLS | Virginia DCLS et al |
| 2668 | NewZealand/20VR1925/2020-04-02 | 2020-04-02 | Southern Community Labs Dunedin | Institute of Environmental Science and Research (ESR) Ltd | **Matt Storey et al** |
| 2669 | NewZealand/20VR1887/2020-04-02 | 2020-04-02 | North Shore Hospital | Institute of Environmental Science and Research (ESR) Ltd | **Matt Storey et al** |
| 2670 | NewZealand/20VR1903/2020-04-02 | 2020-04-02 | Southern Community Labs Dunedin | Institute of Environmental Science and Research (ESR) Ltd | **Matt Storey et al** |
| 2671 | NewZealand/20VR1911/2020-04-02 | 2020-04-02 | Southern Community Labs Dunedin | Institute of Environmental Science and Research (ESR) Ltd | **Matt Storey et al** |
| 2672 | NewZealand/20VR2030/2020-04-02 | 2020-04-02 | Wellington SCL (WN) | Institute of Environmental Science and Research (ESR) Ltd | **Matt Storey et al** |
| 2673 | NewZealand/20VR1910/2020-04-02 | 2020-04-02 | Southern Community Labs Dunedin | Institute of Environmental Science and Research (ESR) Ltd | **Matt Storey et al** |
| 2674 | NewZealand/20VR1952/2020-04-02 | 2020-04-02 | Waikato Hospital | Institute of Environmental Science and Research (ESR) Ltd | **Matt Storey et al** |
| 2675 | NewZealand/20VR1907/2020-04-02 | 2020-04-02 | Southern Community Labs Dunedin | Institute of Environmental Science and Research (ESR) Ltd | **Matt Storey et al** |
| 2676 | USA/WA-UW-5144/2020 | 2020-04-02 | UW Virology Lab | UW Virology Lab | Pavitra Roychoudhury et al |
| 2677 | USA/WA-S342/2020 | 2020-04-02 | Washington State Department of Health | Seattle Flu Study | Chu et al et al |
| 2678 | USA/CA-CZB-1007/2020 | 2020-04-02 | County of Santa Clara Public Health Department | Chan-Zuckerberg Biohub | CZB Cliahub Consortium et al |
| 2679 | England/CAMB-73C5F/2020 | 2020-04-02 | Department of Pathology, University of Cambridge | COVID-19 Genomics UK (COG-UK) Consortium | Luke W Meredith et al |
| 2680 | Russia/StPetersburg-RII4546V/2020 | 2020-04-02 | WHO National Influenza Centre Russian Federation | WHO National Influenza Centre Russian Federation | Andrey Komissarov et al |
| 2681 | USA/TX-HMH0437/2020 | 2020-04-02 | Houston Methodist Hospital | Houston Methodist Hospital | S. Wesley Long et al |
| 2682 | USA/TX-HMH0438/2020 | 2020-04-02 | Houston Methodist Hospital | Houston Methodist Hospital | S. Wesley Long et al |
| 2683 | USA/WA-S467/2020 | 2020-04-02 | Washington State Department of Health | Seattle Flu Study | Chu et al et al |
| 2684 | USA/LA-SR0157/2020 | 2020-04-02 | Robert Garry lab | Andersen lab at Scripps Research | Allison Smither et al |
| 2685 | USA/TX-HMH0439/2020 | 2020-04-02 | Houston Methodist Hospital | Houston Methodist Hospital | S. Wesley Long et al |
| 2686 | USA/NY-NYUMC323/2020 | 2020-04-02 | NYU Langone Health | Departments of Pathology and Medicine, New York University School of Medicine | Maria Aguero-Rosenfeld et al |
| 2687 | Belgium/ITM_C262/2020 | 2020-04-02 | Hospital AZ Rivierenland | Institute of Tropical Medicine | Philippe Selhorst et al |
| 2688 | USA/AK-PHL092/2020 | 2020-04-02 | Alaska State Virology Laboratory | Alaska State Virology Laboratory | Chen et al |
| 2689 | Luxembourg/LNS6326231/2020 | 2020-04-02 | Laboratoire National de Sante, Microbiology, Virology | Laboratoire National de Sante, Microbiology, Epidemiology | Anke Wienecke-Baldacchino |

| | | | | | |
|---|---|---|---|---|---|
| | | | | and Microbial Genomics | et al |
| 2690 | USA/WA-UW-6327/2020 | 2020-04-02 | UW Virology Lab | UW Virology Lab | Pavitra Roychoudhury et al |
| 2691 | USA/CA-CZB-1036/2020 | 2020-04-02 | County of Santa Clara Public Health Department | Chan-Zuckerberg Biohub | CZB Cliahub Consortium et al |
| 2692 | USA/WI-255/2020 | 2020-04-02 | University of Wisconsin-Madison AIDS Vaccine Research Laboratories | University of Wisconsin-Madison AIDS Vaccine Research Laboratories | Moreno et al |
| 2693 | England/CAMB-73BF8/2020 | 2020-04-02 | Department of Pathology, University of Cambridge | COVID-19 Genomics UK (COG-UK) Consortium | Luke W Meredith et al |
| 2694 | USA/CA-CZB-1003/2020 | 2020-04-02 | County of Santa Clara Public Health Department | Chan-Zuckerberg Biohub | CZB Cliahub Consortium et al |
| 2695 | Netherlands/NA_307/2020 | 2020-04-02 | Dutch COVID-19 response team | Erasmus Medical Center | Bas Oude Munnink et al |
| 2696 | USA/WI-UW-277/2020 | 2020-04-02 | University of Wisconsin-Madison AIDS Vaccine Research Laboratories | University of Wisconsin-Madison AIDS Vaccine Research Laboratories | Gage Moreno et al |
| 2697 | USA/WI-87/2020 | 2020-04-02 | University of Wisconsin-Madison AIDS Vaccine Research Laboratories | University of Wisconsin-Madison AIDS Vaccine Research Laboratories | Moreno et al |
| 2698 | USA/AZ-TG273318/2020 | 2020-04-02 | AZ SPHL, Arizona Department of Health Services | TGen North | Jolene Bowers et al |
| 2699 | USA/WA-S484/2020 | 2020-04-02 | Washington State Department of Health | Seattle Flu Study | Chu et al et al |
| 2700 | England/CAMB-73C31/2020 | 2020-04-02 | Department of Pathology, University of Cambridge | COVID-19 Genomics UK (COG-UK) Consortium | Luke W Meredith et al |
| 2701 | USA/WA-S341/2020 | 2020-04-02 | Washington State Department of Health | Seattle Flu Study | Chu et al et al |
| 2702 | Taiwan/TSGH-23/2020 | 2020-04-02 | TSGH-CP molecular lab | TSGH-CP molecular lab | Cherng-Lih Perng et al |
| 2703 | Taiwan/TSGH-24/2020 | 2020-04-02 | TSGH-CP molecular lab | TSGH-CP molecular lab | Cherng-Lih Perng et al |
| 2704 | USA/TX-HMH0348/2020 | 2020-04-02 | Houston Methodist Hospital | Houston Methodist Hospital | S. Wesley Long et al |
| 2705 | USA/WA-S313/2020 | 2020-04-02 | Washington State Department of Health | Seattle Flu Study | Chu et al et al |
| 2706 | Scotland/EDB612/2020 | 2020-04-02 | Virology Department, Royal Infirmary of Edinburgh, NHS Lothian / School of Biological Sciences, University of Edinburgh / Institute of Genetics and Molecular Medicine, University of Edinburgh | COVID-19 Genomics UK (COG-UK) Consortium | McHugh M et al |
| 2707 | USA/WA-S344/2020 | 2020-04-02 | Washington State Department of Health | Seattle Flu Study | Chu et al et al |
| 2708 | USA/WI-GMF-00557/2020 | 2020-04-02 | Gundersen Molecular Diagnostics Laboratory | Kabara Cancer Research Institute | Craig S. Richmond et al |
| 2709 | USA/VA-DCLS-0090/2020 | 2020-04-02 | Virginia DCLS | Virginia DCLS | Virginia DCLS et al |
| 2710 | USA/TX-HMH0435/2020 | 2020-04-02 | Houston Methodist Hospital | Houston Methodist Hospital | S. Wesley Long et al |
| 2711 | USA/WA-S330/2020 | 2020-04-02 | Washington State Department of Health | Seattle Flu Study | Chu et al et al |
| 2712 | USA/NY-NYUMC218/2020 | 2020-04-03 | NYU Langone Health | Departments of Pathology and Medicine, New York University School of Medicine | Maria Aguero-Rosenfeld et al |
| 2713 | England/CAMB-78382/2020 | 2020-04-03 | Department of Pathology, University of Cambridge | COVID-19 Genomics UK (COG-UK) Consortium | Luke W Meredith et al |
| 2714 | Scotland/EDB689/2020 | 2020-04-03 | Virology Department, Royal Infirmary of Edinburgh, NHS Lothian / School of Biological Sciences, University of Edinburgh / Institute of Genetics and Molecular Medicine, University of Edinburgh | COVID-19 Genomics UK (COG-UK) Consortium | McHugh M et al |
| 2715 | Australia/VIC1042/2020 | 2020-04-03 | Victorian Infectious Diseases Reference Laboratory (VIDRL) | Microbiological Diagnostic Unit Public Health Laboratory and Victorian Infectious Diseases Reference Laboratory, The Peter Doherty Institute for Infection and Immunity | Caly L. et al |
| 2716 | USA/NY-NYUMC246/2020 | 2020-04-03 | NYU Langone Health | Departments of Pathology and | Maria |

| | | | | | |
|---|---|---|---|---|---|
| | | | | Medicine, New York University School of Medicine | Aguero-Rosenfeld et al |
| 2717 | USA/WA-S336/2020 | 2020-04-03 | Washington State Department of Health | Seattle Flu Study | Chu et al et al |
| 2718 | Australia/VIC1041/2020 | 2020-04-03 | Victorian Infectious Diseases Reference Laboratory (VIDRL) | Microbiological Diagnostic Unit Public Health Laboratory and Victorian Infectious Diseases Reference Laboratory, The Peter Doherty Institute for Infection and Immunity | Caly L. et al |
| 2719 | Australia/VIC1043/2020 | 2020-04-03 | Victorian Infectious Diseases Reference Laboratory (VIDRL) | Microbiological Diagnostic Unit Public Health Laboratory and Victorian Infectious Diseases Reference Laboratory, The Peter Doherty Institute for Infection and Immunity | Caly L. et al |
| 2720 | Australia/VIC1037/2020 | 2020-04-03 | Victorian Infectious Diseases Reference Laboratory (VIDRL) | Microbiological Diagnostic Unit Public Health Laboratory and Victorian Infectious Diseases Reference Laboratory, The Peter Doherty Institute for Infection and Immunity | Caly L. et al |
| 2721 | Australia/VIC823/2020 | 2020-04-03 | Victorian Infectious Diseases Reference Laboratory (VIDRL) | Microbiological Diagnostic Unit Public Health Laboratory and Victorian Infectious Diseases Reference Laboratory, Doherty Institute | Caly L. et al |
| 2722 | Australia/VIC820/2020 | 2020-04-03 | Victorian Infectious Diseases Reference Laboratory (VIDRL) | Microbiological Diagnostic Unit Public Health Laboratory and Victorian Infectious Diseases Reference Laboratory, Doherty Institute | Caly L. et al |
| 2723 | Australia/VIC1046/2020 | 2020-04-03 | Victorian Infectious Diseases Reference Laboratory (VIDRL) | Microbiological Diagnostic Unit Public Health Laboratory and Victorian Infectious Diseases Reference Laboratory, The Peter Doherty Institute for Infection and Immunity | Caly L. et al |
| 2724 | Australia/VIC824/2020 | 2020-04-03 | Victorian Infectious Diseases Reference Laboratory (VIDRL) | Microbiological Diagnostic Unit Public Health Laboratory and Victorian Infectious Diseases Reference Laboratory, Doherty Institute | Caly L. et al |
| 2725 | Australia/VIC828/2020 | 2020-04-03 | Victorian Infectious Diseases Reference Laboratory (VIDRL) | Microbiological Diagnostic Unit Public Health Laboratory and Victorian Infectious Diseases Reference Laboratory, Doherty Institute | Caly L. et al |
| 2726 | USA/WA-UW-6417/2020 | 2020-04-03 | UW Virology Lab | UW Virology Lab | Pavitra Roychoudhury et al |
| 2727 | USA/WA-UW-6399/2020 | 2020-04-03 | UW Virology Lab | UW Virology Lab | Pavitra Roychoudhury et al |
| 2728 | Belgium/ULG-10078/2020 | 2020-04-03 | Department of Clinical Microbiology | GIGA Medical Genomics | Keith Durkin et al |
| 2729 | Scotland/EDB753/2020 | 2020-04-03 | Virology Department, Royal Infirmary of Edinburgh, NHS Lothian / School of Biological Sciences, University of Edinburgh / Institute of Genetics and Molecular Medicine, University of Edinburgh | COVID-19 Genomics UK (COG-UK) Consortium | McHugh M et al |
| 2730 | USA/WI-GMF-00921/2020 | 2020-04-03 | Gundersen Molecular Diagnostics Laboratory | Kabara Cancer Research Institute | Craig S. Richmond et al |
| 2731 | USA/LA-SR0170/2020 | 2020-04-03 | Robert Garry lab | Andersen lab at Scripps Research | Allison Smither et al |
| 2732 | USA/CA-CZB0741/2020 | 2020-04-03 | UCSF Clinical Microbiology Laboratory | Chan-Zuckerberg Biohub | CZB Cliahub Consortium et al |
| 2733 | Switzerland/GE2453/2020 | 2020-04-03 | University Hospitals of Geneva Laboratory of Virology | University Hospitals of Geneva Laboratory of Virology | Laubscher F. et al |
| 2734 | USA/NY-NYUMC330/2020 | 2020-04-03 | NYU Langone Health | Departments of Pathology and | Maria |

| | | | | | |
|---|---|---|---|---|---|
| | | | | Medicine, New York University School of Medicine | Aguero-Rosenfeld et al |
| 2735 | USA/WI-UW-278/2020 | 2020-04-03 | University of Wisconsin-Madison AIDS Vaccine Research Laboratories | University of Wisconsin-Madison AIDS Vaccine Research Laboratories | Gage Moreno et al |
| 2736 | USA/NY-NYUMC248/2020 | 2020-04-03 | NYU Langone Health | Departments of Pathology and Medicine, New York University School of Medicine | Maria Aguero-Rosenfeld et al |
| 2737 | India/S11/2020 | 2020-04-03 | ICMR-National Institute of Cholera and Enteric Diseases | National Institute of Biomedical Genomics | Arindam Maitra et al |
| 2738 | USA/NY-NYUMC327/2020 | 2020-04-03 | NYU Langone Health | Departments of Pathology and Medicine, New York University School of Medicine | Maria Aguero-Rosenfeld et al |
| 2739 | USA/WI-UW-279/2020 | 2020-04-03 | University of Wisconsin-Madison AIDS Vaccine Research Laboratories | University of Wisconsin-Madison AIDS Vaccine Research Laboratories | Gage Moreno et al |
| 2740 | USA/TX-HMH0440/2020 | 2020-04-03 | Houston Methodist Hospital | Houston Methodist Hospital | S. Wesley Long et al |
| 2741 | USA/TX-HMH0451/2020 | 2020-04-03 | Houston Methodist Hospital | Houston Methodist Hospital | S. Wesley Long et al |
| 2742 | USA/CA-CZB-1010/2020 | 2020-04-03 | County of Santa Clara Public Health Department | Chan-Zuckerberg Biohub | CZB Cliahub Consortium et al |
| 2743 | USA/WA-S328/2020 | 2020-04-03 | Washington State Department of Health | Seattle Flu Study | Chu et al et al |
| 2744 | Switzerland/GE7617/2020 | 2020-04-03 | University Hospitals of Geneva Laboratory of Virology | University Hospitals of Geneva Laboratory of Virology | Laubscher F. et al |
| 2745 | Scotland/EDB773/2020 | 2020-04-03 | Virology Department, Royal Infirmary of Edinburgh, NHS Lothian / School of Biological Sciences, University of Edinburgh / Institute of Genetics and Molecular Medicine, University of Edinburgh | COVID-19 Genomics UK (COG-UK) Consortium | McHugh M et al |
| 2746 | NewZealand/20VR1933/2020-04-03 | 2020-04-03 | Southern Community Labs Dunedin | Institute of Environmental Science and Research (ESR) Ltd | **Matt Storey et al** |
| 2747 | NewZealand/20VR2055/2020-04-03 | 2020-04-03 | Canterbury Health Laboratories | Institute of Environmental Science and Research (ESR) Ltd | **Matt Storey et al** |
| 2748 | NewZealand/20VR2056/2020-04-03 | 2020-04-03 | Canterbury Health Laboratories | Institute of Environmental Science and Research (ESR) Ltd | **Matt Storey et al** |
| 2749 | NewZealand/20VR1947/2020-04-03 | 2020-04-03 | Southern Community Labs Dunedin | Institute of Environmental Science and Research (ESR) Ltd | **Matt Storey et al** |
| 2750 | NewZealand/20VR2054/2020-04-03 | 2020-04-03 | Canterbury Health Laboratories | Institute of Environmental Science and Research (ESR) Ltd | **Matt Storey et al** |
| 2751 | NewZealand/20VR1927/2020-04-03 | 2020-04-03 | Southern Community Labs Dunedin | Institute of Environmental Science and Research (ESR) Ltd | **Matt Storey et al** |
| 2752 | NewZealand/20VR2053/2020-04-03 | 2020-04-03 | Canterbury Health Laboratories | Institute of Environmental Science and Research (ESR) Ltd | **Matt Storey et al** |
| 2753 | NewZealand/20VR1924/2020-04-04 | 2020-04-03 | Southern Community Labs Dunedin | Institute of Environmental Science and Research (ESR) | **Matt Storey et al** |
| 2754 | USA/WA-S314/2020 | 2020-04-03 | Washington State Department of Health | Seattle Flu Study | Chu et al et al |
| 2755 | USA/WA-S473/2020 | 2020-04-03 | Washington State Department of Health | Seattle Flu Study | Chu et al et al |
| 2756 | England/CAMB-73B61/2020 | 2020-04-03 | Department of Pathology, University of Cambridge | COVID-19 Genomics UK (COG-UK) Consortium | Luke W Meredith et al |
| 2757 | USA/WA-UW-6400/2020 | 2020-04-03 | UW Virology Lab | UW Virology Lab | Pavitra Roychoudhury et al |
| 2758 | USA/TX-HMH0441/2020 | 2020-04-03 | Houston Methodist Hospital | Houston Methodist Hospital | S. Wesley Long et al |
| 2759 | USA/VA-DCLS-0081/2020 | 2020-04-03 | Virginia DCLS | Virginia DCLS | Virginia DCLS et al |
| 2760 | USA/VA-DCLS-0067/2020 | 2020-04-03 | Virginia DCLS | Virginia DCLS | Virginia DCLS et al |
| 2761 | Switzerland/GE1184/2020 | 2020-04-03 | University Hospitals of Geneva Laboratory of Virology | University Hospitals of Geneva Laboratory of Virology | Laubscher F. et al |
| 2762 | USA/WI-GMF-00744/2020 | 2020-04-03 | Gundersen Molecular Diagnostics Laboratory | Kabara Cancer Research Institute | Craig S. Richmond et al |
| 2763 | USA/WI-100/2020 | 2020-04-03 | University of Wisconsin-Madison AIDS Vaccine Research Laboratories | University of Wisconsin-Madison AIDS Vaccine Research Laboratories | Moreno et al |

| | | | | | |
|---|---|---|---|---|---|
| 2764 | USA/CA-CZB0750/2020 | 2020-04-03 | UCSF Clinical Microbiology Laboratory | Chan-Zuckerberg Biohub | CZB Cliahub Consortium et al |
| 2765 | Argentina/PAIS_A017/2020 | 2020-04-03 | Departamento de Biología y genética molecular, IACA Laboratorios. | Área de Secuenciación del Laboratorio de Virología del Hospital de Niños Dr. Ricardo Gutierrez | Nabaes Jodar et al |
| 2766 | Scotland/EDB781/2020 | 2020-04-03 | Virology Department, Royal Infirmary of Edinburgh, NHS Lothian / School of Biological Sciences, University of Edinburgh / Institute of Genetics and Molecular Medicine, University of Edinburgh | COVID-19 Genomics UK (COG-UK) Consortium | McHugh M et al |
| 2767 | USA/NY-NYUMC245/2020 | 2020-04-03 | NYU Langone Health | Departments of Pathology and Medicine, New York University School of Medicine | Maria Aguero-Rosenfeld et al |
| 2768 | USA/NY-NYUMC244/2020 | 2020-04-03 | NYU Langone Health | Departments of Pathology and Medicine, New York University School of Medicine | Maria Aguero-Rosenfeld et al |
| 2769 | USA/NY-NYUMC216/2020 | 2020-04-03 | NYU Langone Health | Departments of Pathology and Medicine, New York University School of Medicine | Maria Aguero-Rosenfeld et al |
| 2770 | Scotland/EDB766/2020 | 2020-04-03 | Virology Department, Royal Infirmary of Edinburgh, NHS Lothian / School of Biological Sciences, University of Edinburgh / Institute of Genetics and Molecular Medicine, University of Edinburgh | COVID-19 Genomics UK (COG-UK) Consortium | McHugh M et al |
| 2771 | USA/TX-HMH0450/2020 | 2020-04-03 | Houston Methodist Hospital | Houston Methodist Hospital | S. Wesley Long et al |
| 2772 | England/CAMB-78364/2020 | 2020-04-03 | Department of Pathology, University of Cambridge | COVID-19 Genomics UK (COG-UK) Consortium | Luke W Meredith et al |
| 2773 | USA/VA-DCLS-0066/2020 | 2020-04-03 | Virginia DCLS | Virginia DCLS | Virginia DCLS et al |
| 2774 | USA/WA-S303/2020 | 2020-04-03 | Washington State Department of Health | Seattle Flu Study | Chu et al et al |
| 2775 | USA/WA-UW-6410/2020 | 2020-04-03 | UW Virology Lab | UW Virology Lab | Pavitra Roychoudhury et al |
| 2776 | Russia/StPetersburg-RII4591S/2020 | 2020-04-03 | WHO National Influenza Centre Russian Federation | WHO National Influenza Centre Russian Federation | Andrey Komissarov et al |
| 2777 | USA/LA-SR0166/2020 | 2020-04-03 | Robert Garry lab | Andersen lab at Scripps Research | Allison Smither et al |
| 2778 | USA/TX-HMH0452/2020 | 2020-04-03 | Houston Methodist Hospital | Houston Methodist Hospital | S. Wesley Long et al |
| 2779 | Scotland/CVR1521/2020 | 2020-04-03 | West of Scotland Specialist Virology Centre, NHSGGC / MRC-University of Glasgow Centre for Virus Research | COVID-19 Genomics UK (COG-UK) Consortium | Ana da Silva Filipe et al |
| 2780 | England/SHEF-C715D/2020 | 2020-04-03 | Virology Department, Sheffield Teaching Hospitals NHS Foundation Trust / Virology Department, Sheffield Teaching Hospitals NHS Foundation Trust | COVID-19 Genomics UK (COG-UK) Consortium | Thushan de Silva et al |
| 2781 | Scotland/EDB774/2020 | 2020-04-03 | Virology Department, Royal Infirmary of Edinburgh, NHS Lothian / School of Biological Sciences, University of Edinburgh / Institute of Genetics and Molecular Medicine, University of Edinburgh | COVID-19 Genomics UK (COG-UK) Consortium | McHugh M et al |
| 2782 | Sweden/20-06883/2020 | 2020-04-03 | Lundens VC | The Public Health Agency of Sweden | Marita Dagner et al |
| 2783 | USA/CA-CZB-1040/2020 | 2020-04-03 | County of Santa Clara Public Health Department | Chan-Zuckerberg Biohub | CZB Cliahub Consortium et al |
| 2784 | USA/WI-90/2020 | 2020-04-03 | University of Wisconsin-Madison AIDS Vaccine Research Laboratories | University of Wisconsin-Madison AIDS Vaccine Research Laboratories | Moreno et al |
| 2785 | Belgium/ULG-10081/2020 | 2020-04-03 | Department of Clinical Microbiology | GIGA Medical Genomics | Keith Durkin et al |

| | | | | | |
|---|---|---|---|---|---|
| 2786 | USA/WA-UW-6403/2020 | 2020-04-03 | UW Virology Lab | UW Virology Lab | Pavitra Roychoudhury et al |
| 2787 | USA/NY-NYUMC214/2020 | 2020-04-03 | NYU Langone Health | Departments of Pathology and Medicine, New York University School of Medicine | Maria Aguero-Rosenfeld et al |
| 2788 | Belgium/ULG-10080/2020 | 2020-04-03 | Department of Clinical Microbiology | GIGA Medical Genomics | Keith Durkin et al |
| 2789 | Luxembourg/LNS0078676/2020 | 2020-04-03 | Laboratoire National de Sante, Microbiology, Virology | Laboratoire National de Sante, Microbiology, Epidemiology and Microbial Genomics | Anke Wienecke-Baldacchino et al |
| 2790 | USA/CA-CZB0746/2020 | 2020-04-03 | UCSF Clinical Microbiology Laboratory | Chan-Zuckerberg Biohub | CZB Cliahub Consortium et al |
| 2791 | Greece/53/2020 | 2020-04-04 | Laboratory of Microbiology, Medical School, National and Kapodistrian University of Athens | Laboratory of Biology, Department of Medicine, Democritus University of Thrace | Kassela K. et al |
| 2792 | Australia/VIC856/2020 | 2020-04-04 | Victorian Infectious Diseases Reference Laboratory (VIDRL) | Microbiological Diagnostic Unit Public Health Laboratory and Victorian Infectious Diseases Reference Laboratory, Doherty Institute | Caly L. et al |
| 2793 | Australia/VIC845/2020 | 2020-04-04 | Victorian Infectious Diseases Reference Laboratory (VIDRL) | Microbiological Diagnostic Unit Public Health Laboratory and Victorian Infectious Diseases Reference Laboratory, Doherty Institute | Caly L. et al |
| 2794 | Australia/VIC837/2020 | 2020-04-04 | Victorian Infectious Diseases Reference Laboratory (VIDRL) | Microbiological Diagnostic Unit Public Health Laboratory and Victorian Infectious Diseases Reference Laboratory, Doherty Institute | Caly L. et al |
| 2795 | Australia/VIC1067/2020 | 2020-04-04 | Microbiological Diagnostic Unit Public Health Laboratory | Microbiological Diagnostic Unit Public Health Laboratory | Seemann T. et al |
| 2796 | USA/WA-S464/2020 | 2020-04-04 | Washington State Department of Health | Seattle Flu Study | Chu et al et al |
| 2797 | USA/AK-PHL101/2020 | 2020-04-04 | Alaska State Virology Laboratory | Alaska State Virology Laboratory | Chen et al |
| 2798 | USA/WA-S346/2020 | 2020-04-04 | Washington State Department of Health | Seattle Flu Study | Chu et al et al |
| 2799 | USA/WI-UW-342/2020 | 2020-04-04 | University of Wisconsin-Madison AIDS Vaccine Research Laboratories | University of Wisconsin-Madison AIDS Vaccine Research Laboratories | Gage Moreno et al |
| 2800 | USA/VA-DCLS-0077/2020 | 2020-04-04 | Virginia DCLS | Virginia DCLS | Virginia DCLS et al |
| 2801 | USA/WA-S317/2020 | 2020-04-04 | Washington State Department of Health | Seattle Flu Study | Chu et al et al |
| 2802 | Australia/VIC1062/2020 | 2020-04-04 | Victorian Infectious Diseases Reference Laboratory (VIDRL) | Microbiological Diagnostic Unit Public Health Laboratory and Victorian Infectious Diseases Reference Laboratory, The Peter Doherty Institute for Infection and Immunity | Caly L. et al |
| 2803 | Australia/VIC851/2020 | 2020-04-04 | Victorian Infectious Diseases Reference Laboratory (VIDRL) | Microbiological Diagnostic Unit Public Health Laboratory and Victorian Infectious Diseases Reference Laboratory, Doherty Institute | Caly L. et al |
| 2804 | Australia/VIC843/2020 | 2020-04-04 | Victorian Infectious Diseases Reference Laboratory (VIDRL) | Microbiological Diagnostic Unit Public Health Laboratory and Victorian Infectious Diseases Reference Laboratory, Doherty Institute | Caly L. et al |
| 2805 | Australia/VIC849/2020 | 2020-04-04 | Victorian Infectious Diseases Reference Laboratory (VIDRL) | Microbiological Diagnostic Unit Public Health Laboratory and Victorian Infectious Diseases Reference Laboratory, Doherty Institute | Caly L. et al |
| 2806 | Australia/VIC848/2020 | 2020-04-04 | Victorian Infectious Diseases Reference Laboratory (VIDRL) | Microbiological Diagnostic Unit Public Health Laboratory and Victorian Infectious Diseases Reference Laboratory, Doherty Institute | Caly L. et al |

| | | | | | |
|---|---|---|---|---|---|
| 2807 | Australia/VIC836/2020 | 2020-04-04 | Victorian Infectious Diseases Reference Laboratory (VIDRL) | Microbiological Diagnostic Unit Public Health Laboratory and Victorian Infectious Diseases Reference Laboratory, Doherty Institute | Caly L. et al |
| 2808 | Australia/VIC846/2020 | 2020-04-04 | Victorian Infectious Diseases Reference Laboratory (VIDRL) | Microbiological Diagnostic Unit Public Health Laboratory and Victorian Infectious Diseases Reference Laboratory, Doherty Institute | Caly L. et al |
| 2809 | Australia/VIC850/2020 | 2020-04-04 | Victorian Infectious Diseases Reference Laboratory (VIDRL) | Microbiological Diagnostic Unit Public Health Laboratory and Victorian Infectious Diseases Reference Laboratory, Doherty Institute | Caly L. et al |
| 2810 | Australia/VIC842/2020 | 2020-04-04 | Victorian Infectious Diseases Reference Laboratory (VIDRL) | Microbiological Diagnostic Unit Public Health Laboratory and Victorian Infectious Diseases Reference Laboratory, Doherty Institute | Caly L. et al |
| 2811 | Australia/VIC847/2020 | 2020-04-04 | Victorian Infectious Diseases Reference Laboratory (VIDRL) | Microbiological Diagnostic Unit Public Health Laboratory and Victorian Infectious Diseases Reference Laboratory, Doherty Institute | Caly L. et al |
| 2812 | Australia/VIC853/2020 | 2020-04-04 | Victorian Infectious Diseases Reference Laboratory (VIDRL) | Microbiological Diagnostic Unit Public Health Laboratory and Victorian Infectious Diseases Reference Laboratory, Doherty Institute | Caly L. et al |
| 2813 | Wales/PHWC-29DDD/2020 | 2020-04-04 | Wales Specialist Virology Centre | Public Health Wales Microbiology Cardiff | Catherine Moore et al |
| 2814 | USA/CT-UW-5773/2020 | 2020-04-04 | UW Virology Lab | UW Virology Lab | Pavitra Roychoudhury et al |
| 2815 | Belgium/ULG-10086/2020 | 2020-04-04 | Department of Clinical Microbiology | GIGA Medical Genomics | Keith Durkin et al |
| 2816 | Scotland/EDB868/2020 | 2020-04-04 | Virology Department, Royal Infirmary of Edinburgh, NHS Lothian / School of Biological Sciences, University of Edinburgh / Institute of Genetics and Molecular Medicine, University of Edinburgh | COVID-19 Genomics UK (COG-UK) Consortium | McHugh M et al |
| 2817 | England/CAMB-78461/2020 | 2020-04-04 | Department of Pathology, University of Cambridge | COVID-19 Genomics UK (COG-UK) Consortium | Luke W Meredith et al |
| 2818 | USA/CA-CZB-1006/2020 | 2020-04-04 | County of Santa Clara Public Health Department | Chan-Zuckerberg Biohub | CZB Cliahub Consortium et al |
| 2819 | Scotland/CVR1622/2020 | 2020-04-04 | West of Scotland Specialist Virology Centre, NHSGGC / MRC-University of Glasgow Centre for Virus Research | COVID-19 Genomics UK (COG-UK) Consortium | Ana da Silva Filipe et al |
| 2820 | USA/WA-S479/2020 | 2020-04-04 | Washington State Department of Health | Seattle Flu Study | Chu et al et al |
| 2821 | USA/NY-NYUMC278/2020 | 2020-04-04 | NYU Langone Health | Departments of Pathology and Medicine, New York University School of Medicine | Maria Aguero-Rosenfeld et al |
| 2822 | USA/CT-UW-5772/2020 | 2020-04-04 | UW Virology Lab | UW Virology Lab | Pavitra Roychoudhury et al |
| 2823 | USA/WA-S311/2020 | 2020-04-04 | Washington State Department of Health | Seattle Flu Study | Chu et al et al |
| 2824 | Wales/PHWC-29D91/2020 | 2020-04-04 | Wales Specialist Virology Centre | Public Health Wales Microbiology Cardiff | Catherine Moore et al |
| 2825 | USA/WA-S334/2020 | 2020-04-04 | Washington State Department of Health | Seattle Flu Study | Chu et al et al |
| 2826 | Scotland/EDB880/2020 | 2020-04-04 | Virology Department, Royal Infirmary of Edinburgh, NHS Lothian / School of Biological Sciences, University of Edinburgh / Institute of Genetics and Molecular | COVID-19 Genomics UK (COG-UK) Consortium | McHugh M et al |

| | | | | | |
|---|---|---|---|---|---|
| | | | Medicine, University of Edinburgh | | |
| 2827 | Wales/PHWC-2A097/2020 | 2020-04-04 | Wales Specialist Virology Centre | Public Health Wales Microbiology Cardiff | Catherine Moore et al |
| 2828 | USA/WA-S319/2020 | 2020-04-04 | Washington State Department of Health | Seattle Flu Study | Chu et al et al |
| 2829 | USA/WI-104/2020 | 2020-04-04 | University of Wisconsin-Madison AIDS Vaccine Research Laboratories | University of Wisconsin-Madison AIDS Vaccine Research Laboratories | Moreno et al |
| 2830 | Belgium/ULG-10082/2020 | 2020-04-04 | Department of Clinical Microbiology | GIGA Medical Genomics | Keith Durkin et al |
| 2831 | NewZealand/20VR2063/2020-04-04 | 2020-04-04 | Canterbury Health Laboratories | Institute of Environmental Science and Research (ESR) Ltd | **Matt Storey et al** |
| 2832 | NewZealand/20VR2052/2020-04-04 | 2020-04-04 | Canterbury Health Laboratories | Institute of Environmental Science and Research (ESR) Ltd | **Matt Storey et al** |
| 2833 | NewZealand/20VR1900/2020-04-04 | 2020-04-04 | PathLab Bay of Plenty | Institute of Environmental Science and Research (ESR) Ltd | **Matt Storey et al** |
| 2834 | NewZealand/20VR1941/2020-04-04 | 2020-04-04 | Southern Community Labs Dunedin | Institute of Environmental Science and Research (ESR) Ltd | **Matt Storey et al** |
| 2835 | NewZealand/20VR1936/2020-04-04 | 2020-04-04 | Southern Community Labs Dunedin | Institute of Environmental Science and Research (ESR) Ltd | **Matt Storey et al** |
| 2836 | USA/CA-CZB-1005/2020 | 2020-04-04 | County of Santa Clara Public Health Department | Chan-Zuckerberg Biohub | CZB Cliahub Consortium et al |
| 2837 | Belgium/ULG-10089/2020 | 2020-04-04 | Department of Clinical Microbiology | GIGA Medical Genomics | Keith Durkin et al |
| 2838 | Wales/PHWC-2A088/2020 | 2020-04-04 | Wales Specialist Virology Centre | Public Health Wales Microbiology Cardiff | Catherine Moore et al |
| 2839 | Wales/PHWC-29D82/2020 | 2020-04-04 | Wales Specialist Virology Centre | Public Health Wales Microbiology Cardiff | Catherine Moore et al |
| 2840 | USA/WA-S340/2020 | 2020-04-04 | Washington State Department of Health | Seattle Flu Study | Chu et al et al |
| 2841 | Scotland/EDB889/2020 | 2020-04-04 | Virology Department, Royal Infirmary of Edinburgh, NHS Lothian / School of Biological Sciences, University of Edinburgh / Institute of Genetics and Molecular Medicine, University of Edinburgh | COVID-19 Genomics UK (COG-UK) Consortium | McHugh M et al |
| 2842 | USA/NY-NYUMC277/2020 | 2020-04-04 | NYU Langone Health | Departments of Pathology and Medicine, New York University School of Medicine | Maria Aguero-Rosenfeld et al |
| 2843 | USA/WA-S333/2020 | 2020-04-04 | Washington State Department of Health | Seattle Flu Study | Chu et al et al |
| 2844 | USA/LA-SR0174/2020 | 2020-04-04 | Robert Garry lab | Andersen lab at Scripps Research | Allison Smither et al |
| 2845 | USA/CT-UW-5749/2020 | 2020-04-04 | UW Virology Lab | UW Virology Lab | Pavitra Roychoudhury et al |
| 2846 | USA/CT-Yale-066/2020 | 2020-04-04 | Yale COVID-19 Biorepository | Grubaugh Lab - Yale School of Public Health | Joseph Fauver et al |
| 2847 | USA/CA-CZB-1039/2020 | 2020-04-04 | County of Santa Clara Public Health Department | Chan-Zuckerberg Biohub | CZB Cliahub Consortium et al |
| 2848 | USA/VA-DCLS-0068/2020 | 2020-04-04 | Virginia DCLS | Virginia DCLS | Virginia DCLS et al |
| 2849 | USA/WI-93/2020 | 2020-04-04 | University of Wisconsin-Madison AIDS Vaccine Research Laboratories | University of Wisconsin-Madison AIDS Vaccine Research Laboratories | Moreno et al |
| 2850 | Scotland/CVR1588/2020 | 2020-04-04 | West of Scotland Specialist Virology Centre, NHSGGC / MRC-University of Glasgow Centre for Virus Research | COVID-19 Genomics UK (COG-UK) Consortium | Ana da Silva Filipe et al |
| 2851 | Sweden/20-07042/2020 | 2020-04-04 | Uppsala Narakut Aleris | The Public Health Agency of Sweden | Annika Nilsson et al |
| 2852 | Scotland/EDB788/2020 | 2020-04-04 | Virology Department, Royal Infirmary of Edinburgh, NHS Lothian / School of Biological Sciences, University of Edinburgh / Institute of Genetics and Molecular | COVID-19 Genomics UK (COG-UK) Consortium | McHugh M et al |

| | | | | | |
|---|---|---|---|---|---|
| | | | Medicine, University of Edinburgh | | |
| 2853 | England/CAMB-78294/2020 | 2020-04-04 | Department of Pathology, University of Cambridge | COVID-19 Genomics UK (COG-UK) Consortium | Luke W Meredith et al |
| 2854 | Belgium/ULG-10094/2020 | 2020-04-04 | Department of Clinical Microbiology | GIGA Medical Genomics | Keith Durkin et al |
| 2855 | Scotland/EDB886/2020 | 2020-04-04 | Virology Department, Royal Infirmary of Edinburgh, NHS Lothian / School of Biological Sciences, University of Edinburgh / Institute of Genetics and Molecular Medicine, University of Edinburgh | COVID-19 Genomics UK (COG-UK) Consortium | McHugh M et al |
| 2856 | Scotland/EDB848/2020 | 2020-04-04 | Virology Department, Royal Infirmary of Edinburgh, NHS Lothian / School of Biological Sciences, University of Edinburgh / Institute of Genetics and Molecular Medicine, University of Edinburgh | COVID-19 Genomics UK (COG-UK) Consortium | McHugh M et al |
| 2857 | Scotland/EDB860/2020 | 2020-04-04 | Virology Department, Royal Infirmary of Edinburgh, NHS Lothian / School of Biological Sciences, University of Edinburgh / Institute of Genetics and Molecular Medicine, University of Edinburgh | COVID-19 Genomics UK (COG-UK) Consortium | McHugh M et al |
| 2858 | USA/CA-CZB-1038/2020 | 2020-04-04 | County of Santa Clara Public Health Department | Chan-Zuckerberg Biohub | CZB Cliahub Consortium et al |
| 2859 | Belgium/ULG-10085/2020 | 2020-04-04 | Department of Clinical Microbiology | GIGA Medical Genomics | Keith Durkin et al |
| 2860 | USA/LA-SR0169/2020 | 2020-04-04 | Robert Garry lab | Andersen lab at Scripps Research | Allison Smither et al |
| 2861 | USA/WA-S463/2020 | 2020-04-04 | Washington State Department of Health | Seattle Flu Study | Chu et al et al |
| 2862 | Wales/PHWC-29B79/2020 | 2020-04-04 | Wales Specialist Virology Centre | Public Health Wales Microbiology Cardiff | Catherine Moore et al |
| 2863 | Scotland/CVR1710/2020 | 2020-04-05 | West of Scotland Specialist Virology Centre, NHSGGC / MRC-University of Glasgow Centre for Virus Research | COVID-19 Genomics UK (COG-UK) Consortium | Ana da Silva Filipe et al |
| 2864 | USA/WA-S469/2020 | 2020-04-05 | Washington State Department of Health | Seattle Flu Study | Chu et al et al |
| 2865 | Scotland/CVR1698/2020 | 2020-04-05 | West of Scotland Specialist Virology Centre, NHSGGC / MRC-University of Glasgow Centre for Virus Research | COVID-19 Genomics UK (COG-UK) Consortium | Ana da Silva Filipe et al |
| 2866 | USA/WI-UW-343/2020 | 2020-04-05 | University of Wisconsin-Madison AIDS Vaccine Research Laboratories | University of Wisconsin-Madison AIDS Vaccine Research Laboratories | Gage Moreno et al |
| 2867 | Australia/VIC869/2020 | 2020-04-05 | Victorian Infectious Diseases Reference Laboratory (VIDRL) | Microbiological Diagnostic Unit Public Health Laboratory and Victorian Infectious Diseases Reference Laboratory, Doherty Institute | Caly L. et al |
| 2868 | Australia/VIC874/2020 | 2020-04-05 | Victorian Infectious Diseases Reference Laboratory (VIDRL) | Microbiological Diagnostic Unit Public Health Laboratory and Victorian Infectious Diseases Reference Laboratory, Doherty Institute | Caly L. et al |
| 2869 | Australia/VIC875/2020 | 2020-04-05 | Victorian Infectious Diseases Reference Laboratory (VIDRL) | Microbiological Diagnostic Unit Public Health Laboratory and Victorian Infectious Diseases Reference Laboratory, Doherty Institute | Caly L. et al |
| 2870 | Australia/VIC857/2020 | 2020-04-05 | Victorian Infectious Diseases Reference Laboratory (VIDRL) | Microbiological Diagnostic Unit Public Health Laboratory and Victorian Infectious Diseases | Caly L. et al |

| | | | | | |
|---|---|---|---|---|---|
| | | | | Reference Laboratory, Doherty Institute | |
| 2871 | Australia/VIC858/2020 | 2020-04-05 | Victorian Infectious Diseases Reference Laboratory (VIDRL) | Microbiological Diagnostic Unit Public Health Laboratory and Victorian Infectious Diseases Reference Laboratory, Doherty Institute | Caly L. et al |
| 2872 | Australia/VIC870/2020 | 2020-04-05 | Victorian Infectious Diseases Reference Laboratory (VIDRL) | Microbiological Diagnostic Unit Public Health Laboratory and Victorian Infectious Diseases Reference Laboratory, Doherty Institute | Caly L. et al |
| 2873 | Australia/VIC867/2020 | 2020-04-05 | Victorian Infectious Diseases Reference Laboratory (VIDRL) | Microbiological Diagnostic Unit Public Health Laboratory and Victorian Infectious Diseases Reference Laboratory, Doherty Institute | Caly L. et al |
| 2874 | Australia/VIC876/2020 | 2020-04-05 | Victorian Infectious Diseases Reference Laboratory (VIDRL) | Microbiological Diagnostic Unit Public Health Laboratory and Victorian Infectious Diseases Reference Laboratory, Doherty Institute | Caly L. et al |
| 2875 | Australia/VIC860/2020 | 2020-04-05 | Victorian Infectious Diseases Reference Laboratory (VIDRL) | Microbiological Diagnostic Unit Public Health Laboratory and Victorian Infectious Diseases Reference Laboratory, Doherty Institute | Caly L. et al |
| 2876 | Australia/VIC868/2020 | 2020-04-05 | Victorian Infectious Diseases Reference Laboratory (VIDRL) | Microbiological Diagnostic Unit Public Health Laboratory and Victorian Infectious Diseases Reference Laboratory, Doherty Institute | Caly L. et al |
| 2877 | Scotland/EDB894/2020 | 2020-04-05 | Virology Department, Royal Infirmary of Edinburgh, NHS Lothian / School of Biological Sciences, University of Edinburgh / Institute of Genetics and Molecular Medicine, University of Edinburgh | COVID-19 Genomics UK (COG-UK) Consortium | McHugh M et al |
| 2878 | USA/NY-NYUMC256/2020 | 2020-04-05 | NYU Langone Health | Departments of Pathology and Medicine, New York University School of Medicine | Maria Aguero-Rosenfeld et al |
| 2879 | USA/WA-S483/2020 | 2020-04-05 | Washington State Department of Health | Seattle Flu Study | Chu et al et al |
| 2880 | USA/NY-NYUMC253/2020 | 2020-04-05 | NYU Langone Health | Departments of Pathology and Medicine, New York University School of Medicine | Maria Aguero-Rosenfeld et al |
| 2881 | Australia/VIC871/2020 | 2020-04-05 | Victorian Infectious Diseases Reference Laboratory (VIDRL) | Microbiological Diagnostic Unit Public Health Laboratory and Victorian Infectious Diseases Reference Laboratory, Doherty Institute | Caly L. et al |
| 2882 | Australia/VIC873/2020 | 2020-04-05 | Victorian Infectious Diseases Reference Laboratory (VIDRL) | Microbiological Diagnostic Unit Public Health Laboratory and Victorian Infectious Diseases Reference Laboratory, Doherty Institute | Caly L. et al |
| 2883 | Australia/VIC863/2020 | 2020-04-05 | Victorian Infectious Diseases Reference Laboratory (VIDRL) | Microbiological Diagnostic Unit Public Health Laboratory and Victorian Infectious Diseases Reference Laboratory, Doherty Institute | Caly L. et al |
| 2884 | Australia/VIC862/2020 | 2020-04-05 | Victorian Infectious Diseases Reference Laboratory (VIDRL) | Microbiological Diagnostic Unit Public Health Laboratory and Victorian Infectious Diseases Reference Laboratory, Doherty Institute | Caly L. et al |
| 2885 | England/CAMB-782EF/2020 | 2020-04-05 | Department of Pathology, University of Cambridge | COVID-19 Genomics UK (COG-UK) Consortium | Luke W Meredith et al |
| 2886 | India/NCDC-3230/2020 | 2020-04-05 | National Centre for Disease control (NCDC) | NCDC/CSIR-IGIB | Pramod Kumar et al |

| | | | | | |
|---|---|---|---|---|---|
| 2887 | Wales/PHWC-2A167/2020 | 2020-04-05 | Wales Specialist Virology Centre | Public Health Wales Microbiology Cardiff | Catherine Moore et al |
| 2888 | USA/NY-NYUMC258/2020 | 2020-04-05 | NYU Langone Health | Departments of Pathology and Medicine, New York University School of Medicine | Maria Aguero-Rosenfeld et al |
| 2889 | England/CAMB-782B2/2020 | 2020-04-05 | Department of Pathology, University of Cambridge | COVID-19 Genomics UK (COG-UK) Consortium | Luke W Meredith et al |
| 2890 | USA/CT-UW-5763/2020 | 2020-04-05 | UW Virology Lab | UW Virology Lab | Pavitra Roychoudhury et al |
| 2891 | USA/WA-S345/2020 | 2020-04-05 | Washington State Department of Health | Seattle Flu Study | Chu et al et al |
| 2892 | Russia/StPetersburg-RII4655S/2020 | 2020-04-05 | WHO National Influenza Centre Russian Federation | WHO National Influenza Centre Russian Federation | Andrey Komissarov et al |
| 2893 | USA/WA-S468/2020 | 2020-04-05 | Washington State Department of Health | Seattle Flu Study | Chu et al et al |
| 2894 | Luxembourg/LNS5971805/2020 | 2020-04-05 | Laboratoire National de Sante, Microbiology, Virology | Laboratoire National de Sante, Microbiology, Epidemiology and Microbial Genomics | Anke Wienecke-Baldacchino et al |
| 2895 | USA/NY-NYUMC236/2020 | 2020-04-05 | NYU Langone Health | Departments of Pathology and Medicine, New York University School of Medicine | Maria Aguero-Rosenfeld et al |
| 2896 | USA/WA-UW-4677/2020 | 2020-04-05 | UW Virology Lab | UW Virology Lab | Pavitra Roychoudhury et al |
| 2897 | USA/NY-NYUMC237/2020 | 2020-04-05 | NYU Langone Health | Departments of Pathology and Medicine, New York University School of Medicine | Maria Aguero-Rosenfeld et al |
| 2898 | Russia/StPetersburg-RII4637S/2020 | 2020-04-05 | WHO National Influenza Centre Russian Federation | WHO National Influenza Centre Russian Federation | Andrey Komissarov et al |
| 2899 | Scotland/EDB922/2020 | 2020-04-05 | Virology Department, Royal Infirmary of Edinburgh, NHS Lothian / School of Biological Sciences, University of Edinburgh / Institute of Genetics and Molecular Medicine, University of Edinburgh | COVID-19 Genomics UK (COG-UK) Consortium | McHugh M et al |
| 2900 | NewZealand/20VR2097/2020-04-05 | 2020-04-05 | Middlemore Hospital | Institute of Environmental Science and Research (ESR) Ltd | **Matt Storey et al** |
| 2901 | NewZealand/20VR2058/2020-04-05 | 2020-04-05 | Canterbury Health Laboratories | Institute of Environmental Science and Research (ESR) Ltd | **Matt Storey et al** |
| 2902 | USA/WA-UW-5364/2020 | 2020-04-05 | UW Virology Lab | UW Virology Lab | Pavitra Roychoudhury et al |
| 2903 | USA/WA-UW-5371/2020 | 2020-04-05 | UW Virology Lab | UW Virology Lab | Pavitra Roychoudhury et al |
| 2904 | Wales/PHWC-29C76/2020 | 2020-04-05 | Wales Specialist Virology Centre | Public Health Wales Microbiology Cardiff | Catherine Moore et al |
| 2905 | USA/NY-NYUMC284/2020 | 2020-04-05 | NYU Langone Health | Departments of Pathology and Medicine, New York University School of Medicine | Maria Aguero-Rosenfeld et al |
| 2906 | USA/NY-NYUMC262/2020 | 2020-04-05 | NYU Langone Health | Departments of Pathology and Medicine, New York University School of Medicine | Maria Aguero-Rosenfeld et al |
| 2907 | England/CAMB-785AA/2020 | 2020-04-05 | Department of Pathology, University of Cambridge | COVID-19 Genomics UK (COG-UK) Consortium | Luke W Meredith et al |
| 2908 | Belgium/ULG-10058/2020 | 2020-04-05 | Department of Clinical Microbiology | GIGA Medical Genomics | Keith Durkin et al |
| 2909 | USA/NY-NYUMC285/2020 | 2020-04-05 | NYU Langone Health | Departments of Pathology and Medicine, New York University School of Medicine | Maria Aguero-Rosenfeld et al |
| 2910 | USA/NY-NYUMC255/2020 | 2020-04-05 | NYU Langone Health | Departments of Pathology and Medicine, New York University School of Medicine | Maria Aguero-Rosenfeld et al |
| 2911 | USA/WA-S472/2020 | 2020-04-05 | Washington State Department of Health | Seattle Flu Study | Chu et al et al |
| 2912 | USA/NY-NYUMC279/2020 | 2020-04-05 | NYU Langone Health | Departments of Pathology and Medicine, New York University School of Medicine | Maria Aguero-Rosenfeld et al |
| 2913 | USA/NY-NYUMC241/2020 | 2020-04-05 | NYU Langone Health | Departments of Pathology and Medicine, New York University | Maria Aguero-Rosenfeld et |

| | | | | | |
|---|---|---|---|---|---|
| | | | | School of Medicine | al |
| 2914 | USA/WA-S470/2020 | 2020-04-05 | Washington State Department of Health | Seattle Flu Study | Chu et al et al |
| 2915 | USA/WA-S327/2020 | 2020-04-05 | Washington State Department of Health | Seattle Flu Study | Chu et al et al |
| 2916 | USA/WA-S304/2020 | 2020-04-05 | Washington State Department of Health | Seattle Flu Study | Chu et al et al |
| 2917 | USA/WA-UW-5359/2020 | 2020-04-05 | UW Virology Lab | UW Virology Lab | Pavitra Roychoudhury et al |
| 2918 | Belgium/ULG-10056/2020 | 2020-04-05 | Department of Clinical Microbiology | GIGA Medical Genomics | Keith Durkin et al |
| 2919 | England/CAMB-78407/2020 | 2020-04-05 | Department of Pathology, University of Cambridge | COVID-19 Genomics UK (COG-UK) Consortium | Luke W Meredith et al |
| 2920 | USA/CA-CZB-1011/2020 | 2020-04-05 | County of Santa Clara Public Health Department | Chan-Zuckerberg Biohub | CZB Cliahub Consortium et al |
| 2921 | USA/CT-UW-5754/2020 | 2020-04-05 | UW Virology Lab | UW Virology Lab | Pavitra Roychoudhury et al |
| 2922 | USA/CA-CZB-1009/2020 | 2020-04-05 | County of Santa Clara Public Health Department | Chan-Zuckerberg Biohub | CZB Cliahub Consortium et al |
| 2923 | USA/WA-S460/2020 | 2020-04-05 | Washington State Department of Health | Seattle Flu Study | Chu et al et al |
| 2924 | India/GBRC1s/2020 | 2020-04-05 | Sir M P Shah Government Medical College, Jamnagar | Gujarat Biotechnology Research Centre, Gandhinagar | Ramesh Pandit et al |
| 2925 | England/CAMB-7858C/2020 | 2020-04-05 | Department of Pathology, University of Cambridge | COVID-19 Genomics UK (COG-UK) Consortium | Luke W Meredith et al |
| 2926 | Belgium/ULG-10053/2020 | 2020-04-05 | Department of Clinical Microbiology | GIGA Medical Genomics | Keith Durkin et al |
| 2927 | Scotland/CVR1669/2020 | 2020-04-05 | West of Scotland Specialist Virology Centre, NHSGGC / MRC-University of Glasgow Centre for Virus Research | COVID-19 Genomics UK (COG-UK) Consortium | Ana da Silva Filipe et al |
| 2928 | England/CAMB-7864D/2020 | 2020-04-05 | Department of Pathology, University of Cambridge | COVID-19 Genomics UK (COG-UK) Consortium | Luke W Meredith et al |
| 2929 | USA/WA-UW-5400/2020 | 2020-04-05 | UW Virology Lab | UW Virology Lab | Pavitra Roychoudhury et al |
| 2930 | USA/WA-S477/2020 | 2020-04-05 | Washington State Department of Health | Seattle Flu Study | Chu et al et al |
| 2931 | USA/NY-NYUMC283/2020 | 2020-04-05 | NYU Langone Health | Departments of Pathology and Medicine, New York University School of Medicine | Maria Aguero-Rosenfeld et al |
| 2932 | USA/NY-NYUMC287/2020 | 2020-04-05 | NYU Langone Health | Departments of Pathology and Medicine, New York University School of Medicine | Maria Aguero-Rosenfeld et al |
| 2933 | Scotland/EDB1117/2020 | 2020-04-06 | Virology Department, Royal Infirmary of Edinburgh, NHS Lothian / School of Biological Sciences, University of Edinburgh / Institute of Genetics and Molecular Medicine, University of Edinburgh | COVID-19 Genomics UK (COG-UK) Consortium | McHugh M et al |
| 2934 | USA/NY-NYUMC230/2020 | 2020-04-06 | NYU Langone Health | Departments of Pathology and Medicine, New York University School of Medicine | Maria Aguero-Rosenfeld et al |
| 2935 | USA/NY-NYUMC234/2020 | 2020-04-06 | NYU Langone Health | Departments of Pathology and Medicine, New York University School of Medicine | Maria Aguero-Rosenfeld et al |
| 2936 | Australia/VIC879/2020 | 2020-04-06 | Victorian Infectious Diseases Reference Laboratory (VIDRL) | Microbiological Diagnostic Unit Public Health Laboratory and Victorian Infectious Diseases Reference Laboratory, Doherty Institute | Caly L. et al |
| 2937 | Australia/VIC1080/2020 | 2020-04-06 | Victorian Infectious Diseases Reference Laboratory (VIDRL) | Microbiological Diagnostic Unit Public Health Laboratory and Victorian Infectious Diseases Reference Laboratory, The Peter Doherty Institute for Infection and Immunity | Caly L. et al |
| 2938 | Australia/VIC884/2020 | 2020-04-06 | Victorian Infectious Diseases Reference Laboratory | Microbiological Diagnostic Unit Public Health Laboratory and | Caly L. et al |

| | | | | | |
|---|---|---|---|---|---|
| | | | (VIDRL) | Victorian Infectious Diseases Reference Laboratory, Doherty Institute | |
| 2939 | Australia/VIC878/2020 | 2020-04-06 | Microbiological Diagnostic Unit Public Health Laboratory | Microbiological Diagnostic Unit Public Health Laboratory | Seemann T. et al |
| 2940 | Australia/VIC886/2020 | 2020-04-06 | Victorian Infectious Diseases Reference Laboratory (VIDRL) | Microbiological Diagnostic Unit Public Health Laboratory and Victorian Infectious Diseases Reference Laboratory, Doherty Institute | Caly L. et al |
| 2941 | USA/NY-NYUMC263/2020 | 2020-04-06 | NYU Langone Health | Departments of Pathology and Medicine, New York University School of Medicine | Maria Aguero-Rosenfeld et al |
| 2942 | USA/NY-NYUMC291/2020 | 2020-04-06 | NYU Langone Health | Departments of Pathology and Medicine, New York University School of Medicine | Maria Aguero-Rosenfeld et al |
| 2943 | USA/NY-NYUMC232/2020 | 2020-04-06 | NYU Langone Health | Departments of Pathology and Medicine, New York University School of Medicine | Maria Aguero-Rosenfeld et al |
| 2944 | USA/NY-NYUMC269/2020 | 2020-04-06 | NYU Langone Health | Departments of Pathology and Medicine, New York University School of Medicine | Maria Aguero-Rosenfeld et al |
| 2945 | USA/WI-258/2020 | 2020-04-06 | University of Wisconsin-Madison AIDS Vaccine Research Laboratories | University of Wisconsin-Madison AIDS Vaccine Research Laboratories | Moreno et al |
| 2946 | USA/NY-NYUMC231/2020 | 2020-04-06 | NYU Langone Health | Departments of Pathology and Medicine, New York University School of Medicine | Maria Aguero-Rosenfeld et al |
| 2947 | Australia/VIC883/2020 | 2020-04-06 | Victorian Infectious Diseases Reference Laboratory (VIDRL) | Microbiological Diagnostic Unit Public Health Laboratory and Victorian Infectious Diseases Reference Laboratory, Doherty Institute | Caly L. et al |
| 2948 | Australia/VIC881/2020 | 2020-04-06 | Victorian Infectious Diseases Reference Laboratory (VIDRL) | Microbiological Diagnostic Unit Public Health Laboratory and Victorian Infectious Diseases Reference Laboratory, Doherty Institute | Caly L. et al |
| 2949 | Australia/VIC1086/2020 | 2020-04-06 | Victorian Infectious Diseases Reference Laboratory (VIDRL) | Microbiological Diagnostic Unit Public Health Laboratory and Victorian Infectious Diseases Reference Laboratory, The Peter Doherty Institute for Infection and Immunity | Caly L. et al |
| 2950 | USA/WA-S456/2020 | 2020-04-06 | Washington State Department of Health | Seattle Flu Study | Chu et al et al |
| 2951 | USA/CT-UW-5757/2020 | 2020-04-06 | UW Virology Lab | UW Virology Lab | Pavitra Roychoudhury et al |
| 2952 | USA/WA-S458/2020 | 2020-04-06 | Washington State Department of Health | Seattle Flu Study | Chu et al et al |
| 2953 | USA/WI-152/2020 | 2020-04-06 | University of Wisconsin-Madison AIDS Vaccine Research Laboratories | University of Wisconsin-Madison AIDS Vaccine Research Laboratories | Moreno et al |
| 2954 | USA/WI-UW-285/2020 | 2020-04-06 | University of Wisconsin-Madison AIDS Vaccine Research Laboratories | University of Wisconsin-Madison AIDS Vaccine Research Laboratories | Gage Moreno et al |
| 2955 | England/CAMB-78865/2020 | 2020-04-06 | Department of Pathology, University of Cambridge | COVID-19 Genomics UK (COG-UK) Consortium | Luke W Meredith et al |
| 2956 | USA/NY-NYUMC238/2020 | 2020-04-06 | NYU Langone Health | Departments of Pathology and Medicine, New York University School of Medicine | Maria Aguero-Rosenfeld et al |
| 2957 | USA/WA-S482/2020 | 2020-04-06 | Washington State Department of Health | Seattle Flu Study | Chu et al et al |
| 2958 | England/CAMB-784DA/2020 | 2020-04-06 | Department of Pathology, University of Cambridge | COVID-19 Genomics UK (COG-UK) Consortium | Luke W Meredith et al |
| 2959 | Switzerland/GE6065/2020 | 2020-04-06 | University Hospitals of Geneva Laboratory of Virology | University Hospitals of Geneva Laboratory of Virology | Laubscher F. et al |
| 2960 | England/CAMB-78944/2020 | 2020-04-06 | Department of Pathology, University of Cambridge | COVID-19 Genomics UK (COG-UK) Consortium | Luke W Meredith et al |

| | | | | | |
|---|---|---|---|---|---|
| 2961 | Switzerland/GE6414/2020 | 2020-04-06 | University Hospitals of Geneva Laboratory of Virology | University Hospitals of Geneva Laboratory of Virology | Laubscher F. et al |
| 2962 | USA/WA-S459/2020 | 2020-04-06 | Washington State Department of Health | Seattle Flu Study | Chu et al et al |
| 2963 | USA/NY-NYUMC267/2020 | 2020-04-06 | NYU Langone Health | Departments of Pathology and Medicine, New York University School of Medicine | Maria Aguero-Rosenfeld et al |
| 2964 | NewZealand/20VR2065/2020-04-06 | 2020-04-06 | Canterbury Health Laboratories | Institute of Environmental Science and Research (ESR) Ltd | **Matt Storey et al** |
| 2965 | NewZealand/20VR2067/2020-04-06 | 2020-04-06 | Canterbury Health Laboratories | Institute of Environmental Science and Research (ESR) Ltd | **Matt Storey et al** |
| 2966 | NewZealand/20VR2066/2020-04-06 | 2020-04-06 | Canterbury Health Laboratories | Institute of Environmental Science and Research (ESR) Ltd | **Matt Storey et al** |
| 2967 | USA/NY-NYUMC270/2020 | 2020-04-06 | NYU Langone Health | Departments of Pathology and Medicine, New York University School of Medicine | Maria Aguero-Rosenfeld et al |
| 2968 | USA/NY-NYUMC290/2020 | 2020-04-06 | NYU Langone Health | Departments of Pathology and Medicine, New York University School of Medicine | Maria Aguero-Rosenfeld et al |
| 2969 | Switzerland/GE6099/2020 | 2020-04-06 | University Hospitals of Geneva Laboratory of Virology | University Hospitals of Geneva Laboratory of Virology | Laubscher F. et al |
| 2970 | USA/NY-NYUMC273/2020 | 2020-04-06 | NYU Langone Health | Departments of Pathology and Medicine, New York University School of Medicine | Maria Aguero-Rosenfeld et al |
| 2971 | England/CAMB-78689/2020 | 2020-04-06 | Department of Pathology, University of Cambridge | COVID-19 Genomics UK (COG-UK) Consortium | Luke W Meredith et al |
| 2972 | USA/NY-NYUMC242/2020 | 2020-04-06 | NYU Langone Health | Departments of Pathology and Medicine, New York University School of Medicine | Maria Aguero-Rosenfeld et al |
| 2973 | Scotland/EDB997/2020 | 2020-04-06 | Virology Department, Royal Infirmary of Edinburgh, NHS Lothian / School of Biological Sciences, University of Edinburgh / Institute of Genetics and Molecular Medicine, University of Edinburgh | COVID-19 Genomics UK (COG-UK) Consortium | McHugh M et al |
| 2974 | USA/NY-NYUMC274/2020 | 2020-04-06 | NYU Langone Health | Departments of Pathology and Medicine, New York University School of Medicine | Maria Aguero-Rosenfeld et al |
| 2975 | England/CAMB-7874A/2020 | 2020-04-06 | Department of Pathology, University of Cambridge | COVID-19 Genomics UK (COG-UK) Consortium | Luke W Meredith et al |
| 2976 | USA/VA-DCLS-0084/2020 | 2020-04-06 | Virginia DCLS | Virginia DCLS | Virginia DCLS et al |
| 2977 | Wales/PHWC-2AF03/2020 | 2020-04-06 | Wales Specialist Virology Centre | Public Health Wales Microbiology Cardiff | Catherine Moore et al |
| 2978 | USA/NY-NYUMC233/2020 | 2020-04-06 | NYU Langone Health | Departments of Pathology and Medicine, New York University School of Medicine | Maria Aguero-Rosenfeld et al |
| 2979 | USA/WI-256/2020 | 2020-04-06 | University of Wisconsin-Madison AIDS Vaccine Research Laboratories | University of Wisconsin-Madison AIDS Vaccine Research Laboratories | Moreno et al |
| 2980 | USA/UN-UW-4645/2020 | 2020-04-06 | UW Virology Lab | UW Virology Lab | Pavitra Roychoudhury et al |
| 2981 | USA/WI-UW-268/2020 | 2020-04-06 | University of Wisconsin-Madison AIDS Vaccine Research Laboratories | University of Wisconsin-Madison AIDS Vaccine Research Laboratories | Gage Moreno et al |
| 2982 | USA/WI-UW-287/2020 | 2020-04-06 | University of Wisconsin-Madison AIDS Vaccine Research Laboratories | University of Wisconsin-Madison AIDS Vaccine Research Laboratories | Gage Moreno et al |
| 2983 | USA/WI-UW-344/2020 | 2020-04-06 | University of Wisconsin-Madison AIDS Vaccine Research Laboratories | University of Wisconsin-Madison AIDS Vaccine Research Laboratories | Gage Moreno et al |
| 2984 | USA/NY-NYUMC275/2020 | 2020-04-06 | NYU Langone Health | Departments of Pathology and Medicine, New York University School of Medicine | Maria Aguero-Rosenfeld et al |
| 2985 | Scotland/EDB1072/2020 | 2020-04-06 | Virology Department, Royal Infirmary of Edinburgh, NHS Lothian / School of Biological | COVID-19 Genomics UK (COG-UK) Consortium | McHugh M et al |

| | | | | | |
|---|---|---|---|---|---|
| | | | Sciences, University of Edinburgh / Institute of Genetics and Molecular Medicine, University of Edinburgh | | |
| 2986 | Belgium/ULG-10098/2020 | 2020-04-06 | Department of Clinical Microbiology | GIGA Medical Genomics | Keith Durkin et al |
| 2987 | England/CAMB-788A1/2020 | 2020-04-06 | Department of Pathology, University of Cambridge | COVID-19 Genomics UK (COG-UK) Consortium | Luke W Meredith et al |
| 2988 | Belgium/ULG-10101/2020 | 2020-04-06 | Department of Clinical Microbiology | GIGA Medical Genomics | Keith Durkin et al |
| 2989 | Scotland/EDB986/2020 | 2020-04-06 | Virology Department, Royal Infirmary of Edinburgh, NHS Lothian / School of Biological Sciences, University of Edinburgh / Institute of Genetics and Molecular Medicine, University of Edinburgh | COVID-19 Genomics UK (COG-UK) Consortium | McHugh M et al |
| 2990 | England/SHEF-CE89D/2020 | 2020-04-06 | Virology Department, Sheffield Teaching Hospitals NHS Foundation Trust / Virology Department, Sheffield Teaching Hospitals NHS Foundation Trust | COVID-19 Genomics UK (COG-UK) Consortium | Thushan de Silva et al |
| 2991 | USA/NY-NYUMC276/2020 | 2020-04-06 | NYU Langone Health | Departments of Pathology and Medicine, New York University School of Medicine | Maria Aguero-Rosenfeld et al |
| 2992 | USA/WA-UW-4649/2020 | 2020-04-06 | UW Virology Lab | UW Virology Lab | Pavitra Roychoudhury et al |
| 2993 | USA/NY-NYUMC240/2020 | 2020-04-06 | NYU Langone Health | Departments of Pathology and Medicine, New York University School of Medicine | Maria Aguero-Rosenfeld et al |
| 2994 | USA/NY-NYUMC272/2020 | 2020-04-06 | NYU Langone Health | Departments of Pathology and Medicine, New York University School of Medicine | Maria Aguero-Rosenfeld et al |
| 2995 | USA/WA-UW-5750/2020 | 2020-04-06 | UW Virology Lab | UW Virology Lab | Pavitra Roychoudhury et al |
| 2996 | Belgium/ULG-10108/2020 | 2020-04-06 | Department of Clinical Microbiology | GIGA Medical Genomics | Keith Durkin et al |
| 2997 | USA/WI-144/2020 | 2020-04-06 | University of Wisconsin-Madison AIDS Vaccine Research Laboratories | University of Wisconsin-Madison AIDS Vaccine Research Laboratories | Moreno et al |
| 2998 | Belgium/ULG-10099/2020 | 2020-04-06 | Department of Clinical Microbiology | GIGA Medical Genomics | Keith Durkin et al |
| 2999 | USA/WI-UW-281/2020 | 2020-04-06 | University of Wisconsin-Madison AIDS Vaccine Research Laboratories | University of Wisconsin-Madison AIDS Vaccine Research Laboratories | Gage Moreno et al |
| 3000 | USA/WA-S471/2020 | 2020-04-06 | Washington State Department of Health | Seattle Flu Study | Chu et al et al |
| 3001 | Scotland/EDB999/2020 | 2020-04-06 | Virology Department, Royal Infirmary of Edinburgh, NHS Lothian / School of Biological Sciences, University of Edinburgh / Institute of Genetics and Molecular Medicine, University of Edinburgh | COVID-19 Genomics UK (COG-UK) Consortium | McHugh M et al |
| 3002 | Scotland/EDB1040/2020 | 2020-04-06 | Virology Department, Royal Infirmary of Edinburgh, NHS Lothian / School of Biological Sciences, University of Edinburgh / Institute of Genetics and Molecular Medicine, University of Edinburgh | COVID-19 Genomics UK (COG-UK) Consortium | McHugh M et al |
| 3003 | England/CAMB-7859B/2020 | 2020-04-06 | Department of Pathology, University of Cambridge | COVID-19 Genomics UK (COG-UK) Consortium | Luke W Meredith et al |
| 3004 | USA/WI-GMF-01316/2020 | 2020-04-06 | Gundersen Molecular Diagnostics Laboratory | Kabara Cancer Research Institute | Craig S. Richmond et al |
| 3005 | USA/WI-UW-269/2020 | 2020-04-06 | University of | University of | Gage Moreno et al |

| | | | | | |
|---|---|---|---|---|---|
| | | | Wisconsin-Madison AIDS Vaccine Research Laboratories | Wisconsin-Madison AIDS Vaccine Research Laboratories | |
| 3006 | USA/WA-S461/2020 | 2020-04-06 | Washington State Department of Health | Seattle Flu Study | Chu et al et al |
| 3007 | USA/NY-NYUMC235/2020 | 2020-04-06 | NYU Langone Health | Departments of Pathology and Medicine, New York University School of Medicine | Maria Aguero-Rosenfeld et al |
| 3008 | England/CAMB-78829/2020 | 2020-04-07 | Department of Pathology, University of Cambridge | COVID-19 Genomics UK (COG-UK) Consortium | Luke W Meredith et al |
| 3009 | Scotland/EDB1081/2020 | 2020-04-07 | Virology Department, Royal Infirmary of Edinburgh, NHS Lothian / School of Biological Sciences, University of Edinburgh / Institute of Genetics and Molecular Medicine, University of Edinburgh | COVID-19 Genomics UK (COG-UK) Consortium | McHugh M et al |
| 3010 | USA/CT-Yale-106/2020 | 2020-04-07 | Yale COVID-19 Biorepository | Grubaugh Lab - Yale School of Public Health | Joseph Fauver et al |
| 3011 | Australia/VIC1115/2020 | 2020-04-07 | Victorian Infectious Diseases Reference Laboratory (VIDRL) | Microbiological Diagnostic Unit Public Health Laboratory and Victorian Infectious Diseases Reference Laboratory, The Peter Doherty Institute for Infection and Immunity | Caly L. et al |
| 3012 | Australia/VIC1112/2020 | 2020-04-07 | Victorian Infectious Diseases Reference Laboratory (VIDRL) | Microbiological Diagnostic Unit Public Health Laboratory and Victorian Infectious Diseases Reference Laboratory, The Peter Doherty Institute for Infection and Immunity | Caly L. et al |
| 3013 | Australia/VIC1279/2020 | 2020-04-07 | Microbiological Diagnostic Unit Public Health Laboratory | Microbiological Diagnostic Unit Public Health Laboratory | Seemann T. et al |
| 3014 | USA/AK-PHL151/2020 | 2020-04-07 | Alaska State Virology Laboratory | Alaska State Virology Laboratory | Jack Chen et al |
| 3015 | Italy/TE12337/2020 | 2020-04-07 | Villa Serena del Dr. Leonardo Petruzzi | Istituto Zooprofilattico Sperimentale dell’Abruzzo e Molise “G. Caporale” | Lorusso A et al |
| 3016 | USA/WI-141/2020 | 2020-04-07 | University of Wisconsin-Madison AIDS Vaccine Research Laboratories | University of Wisconsin-Madison AIDS Vaccine Research Laboratories | Moreno et al |
| 3017 | USA/WI-127/2020 | 2020-04-07 | University of Wisconsin-Madison AIDS Vaccine Research Laboratories | University of Wisconsin-Madison AIDS Vaccine Research Laboratories | Moreno et al |
| 3018 | USA/NY-NYUMC333/2020 | 2020-04-07 | NYU Langone Health | Departments of Pathology and Medicine, New York University School of Medicine | Maria Aguero-Rosenfeld et al |
| 3019 | USA/NY-NYUMC334/2020 | 2020-04-07 | NYU Langone Health | Departments of Pathology and Medicine, New York University School of Medicine | Maria Aguero-Rosenfeld et al |
| 3020 | Switzerland/GE3655/2020 | 2020-04-07 | University Hospitals of Geneva Laboratory of Virology | University Hospitals of Geneva Laboratory of Virology | Laubscher F. et al |
| 3021 | USA/AK-PHL153/2020 | 2020-04-07 | Alaska State Virology Laboratory | Alaska State Virology Laboratory | Jack Chen et al |
| 3022 | USA/WI-UW-266/2020 | 2020-04-07 | University of Wisconsin-Madison AIDS Vaccine Research Laboratories | University of Wisconsin-Madison AIDS Vaccine Research Laboratories | Gage Moreno et al |
| 3023 | USA/WI-124/2020 | 2020-04-07 | University of Wisconsin-Madison AIDS Vaccine Research Laboratories | University of Wisconsin-Madison AIDS Vaccine Research Laboratories | Moreno et al |
| 3024 | Sweden/20-07013/2020 | 2020-04-07 | Omtanken Grimmered | The Public Health Agency of Sweden | Bernd Sengpiel et al |
| 3025 | USA/WI-148/2020 | 2020-04-07 | University of Wisconsin-Madison AIDS Vaccine Research Laboratories | University of Wisconsin-Madison AIDS Vaccine Research Laboratories | Moreno et al |
| 3026 | Wales/PHWC-2B00F/2020 | 2020-04-07 | Wales Specialist Virology Centre | Public Health Wales Microbiology Cardiff | Catherine Moore et al |
| 3027 | USA/WA-UW-4921/2020 | 2020-04-07 | UW Virology Lab | UW Virology Lab | Pavitra Roychoudhury et al |

| | | | | | |
|---|---|---|---|---|---|
| 3028 | USA/WI-UW-284/2020 | 2020-04-07 | University of Wisconsin-Madison AIDS Vaccine Research Laboratories | University of Wisconsin-Madison AIDS Vaccine Research Laboratories | Gage Moreno et al |
| 3029 | NewZealand/20VR1986/2020-04-07 | 2020-04-07 | North Shore Hospital | Institute of Environmental Science and Research (ESR) Ltd | **Matt Storey et al** |
| 3030 | USA/NY-NYUMC351/2020 | 2020-04-07 | NYU Langone Health | Departments of Pathology and Medicine, New York University School of Medicine | Maria Aguero-Rosenfeld et al |
| 3031 | USA/WA-UW-5879/2020 | 2020-04-07 | UW Virology Lab | UW Virology Lab | Pavitra Roychoudhury et al |
| 3032 | USA/AK-PHL152/2020 | 2020-04-07 | Alaska State Virology Laboratory | Alaska State Virology Laboratory | Jack Chen et al |
| 3033 | USA/WA-UW-4925/2020 | 2020-04-07 | UW Virology Lab | UW Virology Lab | Pavitra Roychoudhury et al |
| 3034 | USA/CA-CZB-1014/2020 | 2020-04-07 | County of Santa Clara Public Health Department | Chan-Zuckerberg Biohub | CZB Cliahub Consortium et al |
| 3035 | Scotland/EDB1095/2020 | 2020-04-07 | Virology Department, Royal Infirmary of Edinburgh, NHS Lothian / School of Biological Sciences, University of Edinburgh / Institute of Genetics and Molecular Medicine, University of Edinburgh | COVID-19 Genomics UK (COG-UK) Consortium | McHugh M et al |
| 3036 | India/GBRC6/2020 | 2020-04-07 | B.J. Medical College and Civil hospital | Gujarat Biotechnology Research Centre | Janvi Raval et al |
| 3037 | USA/NY-NYUMC354/2020 | 2020-04-07 | NYU Langone Health | Departments of Pathology and Medicine, New York University School of Medicine | Maria Aguero-Rosenfeld et al |
| 3038 | USA/WI-125/2020 | 2020-04-07 | University of Wisconsin-Madison AIDS Vaccine Research Laboratories | University of Wisconsin-Madison AIDS Vaccine Research Laboratories | Moreno et al |
| 3039 | England/CAMB-7AAB8/2020 | 2020-04-07 | Department of Pathology, University of Cambridge | COVID-19 Genomics UK (COG-UK) Consortium | Luke W Meredith et al |
| 3040 | USA/NY-NYUMC345/2020 | 2020-04-07 | NYU Langone Health | Departments of Pathology and Medicine, New York University School of Medicine | Maria Aguero-Rosenfeld et al |
| 3041 | Sweden/20-07010/2020 | 2020-04-07 | Omtanken Grimmered | The Public Health Agency of Sweden | Bernd Sengpiel et al |
| 3042 | USA/CA-CZB-1013/2020 | 2020-04-07 | County of Santa Clara Public Health Department | Chan-Zuckerberg Biohub | CZB Cliahub Consortium et al |
| 3043 | USA/WA-UW-5884/2020 | 2020-04-07 | UW Virology Lab | UW Virology Lab | Pavitra Roychoudhury et al |
| 3044 | USA/WA-UW-4892/2020 | 2020-04-07 | UW Virology Lab | UW Virology Lab | Pavitra Roychoudhury et al |
| 3045 | NewZealand/20VR2046/2020-04-07 | 2020-04-07 | Southern Community Labs Dunedin | Institute of Environmental Science and Research (ESR) Ltd | **Matt Storey et al** |
| 3046 | USA/WA-UW-5911/2020 | 2020-04-07 | UW Virology Lab | UW Virology Lab | Pavitra Roychoudhury et al |
| 3047 | France/OCC-14/2020 | 2020-04-07 | CHU Purpan - Laboratoire de Virologie - Institut Fédératif de Biologie | Laboratoire de virologie - École Nationale Vétérinaire de Toulouse | Guillaume Croville et al |
| 3048 | England/CAMB-78ABA/2020 | 2020-04-07 | Department of Pathology, University of Cambridge | COVID-19 Genomics UK (COG-UK) Consortium | Luke W Meredith et al |
| 3049 | Belgium/ULG-10100/2020 | 2020-04-07 | Department of Clinical Microbiology | GIGA Medical Genomics | Keith Durkin et al |
| 3050 | England/CAMB-78A50/2020 | 2020-04-07 | Department of Pathology, University of Cambridge | COVID-19 Genomics UK (COG-UK) Consortium | Luke W Meredith et al |
| 3051 | USA/WA-UW-5025/2020 | 2020-04-07 | UW Virology Lab | UW Virology Lab | Pavitra Roychoudhury et al |
| 3052 | USA/WI-122/2020 | 2020-04-07 | University of Wisconsin-Madison AIDS Vaccine Research Laboratories | University of Wisconsin-Madison AIDS Vaccine Research Laboratories | Moreno et al |
| 3053 | USA/UN-UW-5742/2020 | 2020-04-07 | UW Virology Lab | UW Virology Lab | Pavitra Roychoudhury et al |
| 3054 | Belgium/ULG-10124/2020 | 2020-04-07 | Department of Clinical Microbiology | GIGA Medical Genomics | Keith Durkin et al |
| 3055 | USA/UN-UW-5769/2020 | 2020-04-07 | UW Virology Lab | UW Virology Lab | Pavitra Roychoudhury et al |

| | | | | | |
|---|---|---|---|---|---|
| 3056 | USA/CT-Yale-105/2020 | 2020-04-07 | Yale COVID-19 Biorepository | Grubaugh Lab - Yale School of Public Health | Joseph Fauver et al |
| 3057 | Russia/StPetersburg-RII4713S/2020 | 2020-04-07 | WHO National Influenza Centre Russian Federation | WHO National Influenza Centre Russian Federation | Andrey Komissarov et al |
| 3058 | USA/NY-NYUMC352/2020 | 2020-04-07 | NYU Langone Health | Departments of Pathology and Medicine, New York University School of Medicine | Maria Aguero-Rosenfeld et al |
| 3059 | England/NOTT-10FB69/2020 | 2020-04-07 | Queens Medical Centre, Clinical Microbiology Department / DeepSeq Nottingham | COVID-19 Genomics UK (COG-UK) Consortium | Gemma Clark et al |
| 3060 | Wales/PHWC-2B1EE/2020 | 2020-04-07 | Wales Specialist Virology Centre | Public Health Wales Microbiology Cardiff | Catherine Moore et al |
| 3061 | Russia/StPetersburg-RII4697S/2020 | 2020-04-07 | WHO National Influenza Centre Russian Federation | WHO National Influenza Centre Russian Federation | Andrey Komissarov et al |
| 3062 | Russia/StPetersburg-RII4694S/2020 | 2020-04-07 | WHO National Influenza Centre Russian Federation | WHO National Influenza Centre Russian Federation | Andrey Komissarov et al |
| 3063 | Russia/StPetersburg-RII4780S/2020 | 2020-04-07 | WHO National Influenza Centre Russian Federation | WHO National Influenza Centre Russian Federation | Andrey Komissarov et al |
| 3064 | England/CAMB-78892/2020 | 2020-04-07 | Department of Pathology, University of Cambridge | COVID-19 Genomics UK (COG-UK) Consortium | Luke W Meredith et al |
| 3065 | USA/NY-NYUMC340/2020 | 2020-04-07 | NYU Langone Health | Departments of Pathology and Medicine, New York University School of Medicine | Maria Aguero-Rosenfeld et al |
| 3066 | USA/WA-UW-5947/2020 | 2020-04-07 | UW Virology Lab | UW Virology Lab | Pavitra Roychoudhury et al |
| 3067 | France/OCC-13/2020 | 2020-04-07 | CHU Purpan - Laboratoire de Virologie - Institut Fédératif de Biologie | Laboratoire de virologie - École Nationale Vétérinaire de Toulouse | Guillaume Croville et al |
| 3068 | USA/CT-UW-5595/2020 | 2020-04-07 | UW Virology Lab | UW Virology Lab | Pavitra Roychoudhury et al |
| 3069 | USA/WI-128/2020 | 2020-04-07 | University of Wisconsin-Madison AIDS Vaccine Research Laboratories | University of Wisconsin-Madison AIDS Vaccine Research Laboratories | Moreno et al |
| 3070 | USA/NY-NYUMC431/2020 | 2020-04-07 | NYU Langone Health | Departments of Pathology and Medicine, New York University School of Medicine | Maria Aguero-Rosenfeld et al |
| 3071 | Scotland/EDB1093/2020 | 2020-04-07 | Virology Department, Royal Infirmary of Edinburgh, NHS Lothian / School of Biological Sciences, University of Edinburgh / Institute of Genetics and Molecular Medicine, University of Edinburgh | COVID-19 Genomics UK (COG-UK) Consortium | McHugh M et al |
| 3072 | Wales/PHWC-2AE7F/2020 | 2020-04-07 | Wales Specialist Virology Centre | Public Health Wales Microbiology Cardiff | Catherine Moore et al |
| 3073 | Russia/StPetersburg-RII4917S/2020 | 2020-04-07 | WHO National Influenza Centre Russian Federation | WHO National Influenza Centre Russian Federation | Andrey Komissarov et al |
| 3074 | Singapore/77/2020 | 2020-04-07 | National Public Health Laboratory, National Centre for Infectious Diseases | National Public Health Laboratory, National Centre for Infectious Diseases | Mak Tze Minn et al |
| 3075 | England/CAMB-7A8FA/2020 | 2020-04-07 | Department of Pathology, University of Cambridge | COVID-19 Genomics UK (COG-UK) Consortium | Luke W Meredith et al |
| 3076 | Scotland/CVR1978/2020 | 2020-04-07 | West of Scotland Specialist Virology Centre, NHSGGC / MRC-University of Glasgow Centre for Virus Research | COVID-19 Genomics UK (COG-UK) Consortium | Ana da Silva Filipe et al |
| 3077 | England/CAMB-789F9/2020 | 2020-04-07 | Department of Pathology, University of Cambridge | COVID-19 Genomics UK (COG-UK) Consortium | Luke W Meredith et al |
| 3078 | England/CAMB-788B0/2020 | 2020-04-07 | Department of Pathology, University of Cambridge | COVID-19 Genomics UK (COG-UK) Consortium | Luke W Meredith et al |
| 3079 | USA/WA-UW-4999/2020 | 2020-04-07 | UW Virology Lab | UW Virology Lab | Pavitra Roychoudhury et al |
| 3080 | Belgium/ULG-10106/2020 | 2020-04-07 | Department of Clinical Microbiology | GIGA Medical Genomics | Keith Durkin et al |
| 3081 | USA/WA-UW-5932/2020 | 2020-04-07 | UW Virology Lab | UW Virology Lab | Pavitra Roychoudhury et al |
| 3082 | England/CAMB-78926/2020 | 2020-04-07 | Department of Pathology, | COVID-19 Genomics UK | Luke W Meredith et al |

| | | | | | |
|---|---|---|---|---|---|
| | | | University of Cambridge | (COG-UK) Consortium | |
| 3083 | USA/WA-UW-5756/2020 | 2020-04-07 | UW Virology Lab | UW Virology Lab | Pavitra Roychoudhury et al |
| 3084 | USA/CA-CZB0156/2020 | 2020-04-08 | UCSF Clinical Microbiology Laboratory | Chan-Zuckerberg Biohub | CZB Cliahub Consortium et al |
| 3085 | Scotland/CVR1971/2020 | 2020-04-08 | West of Scotland Specialist Virology Centre, NHSGGC / MRC-University of Glasgow Centre for Virus Research | COVID-19 Genomics UK (COG-UK) Consortium | Ana da Silva Filipe et al |
| 3086 | Australia/VIC1135/2020 | 2020-04-08 | Victorian Infectious Diseases Reference Laboratory (VIDRL) | Microbiological Diagnostic Unit Public Health Laboratory and Victorian Infectious Diseases Reference Laboratory, The Peter Doherty Institute for Infection and Immunity | Caly L. et al |
| 3087 | Australia/VIC1281/2020 | 2020-04-08 | Microbiological Diagnostic Unit Public Health Laboratory | Microbiological Diagnostic Unit Public Health Laboratory | Seemann T. et al |
| 3088 | Australia/VIC1119/2020 | 2020-04-08 | Victorian Infectious Diseases Reference Laboratory (VIDRL) | Microbiological Diagnostic Unit Public Health Laboratory and Victorian Infectious Diseases Reference Laboratory, The Peter Doherty Institute for Infection and Immunity | Caly L. et al |
| 3089 | Australia/VIC1144/2020 | 2020-04-08 | Victorian Infectious Diseases Reference Laboratory (VIDRL) | Microbiological Diagnostic Unit Public Health Laboratory and Victorian Infectious Diseases Reference Laboratory, The Peter Doherty Institute for Infection and Immunity | Caly L. et al |
| 3090 | Australia/VIC898/2020 | 2020-04-08 | Microbiological Diagnostic Unit Public Health Laboratory | Microbiological Diagnostic Unit Public Health Laboratory | Seemann T. et al |
| 3091 | France/OCC-15/2020 | 2020-04-08 | CHU Purpan - Laboratoire de Virologie - Institut Fédératif de Biologie | Laboratoire de virologie - École Nationale Vétérinaire de Toulouse | Guillaume Croville et al |
| 3092 | Italy/TE12774/2020 | 2020-04-08 | Ospedale SS Annunziata | Istituto Zooprofilattico Sperimentale dell'Abruzzo e Molise "G. Caporale" | Lorusso A et al |
| 3093 | Scotland/EDB1167/2020 | 2020-04-08 | Virology Department, Royal Infirmary of Edinburgh, NHS Lothian / School of Biological Sciences, University of Edinburgh / Institute of Genetics and Molecular Medicine, University of Edinburgh | COVID-19 Genomics UK (COG-UK) Consortium | McHugh M et al |
| 3094 | France/OCC-16/2020 | 2020-04-08 | CHU Purpan - Laboratoire de Virologie - Institut Fédératif de Biologie | Laboratoire de virologie - École Nationale Vétérinaire de Toulouse | Guillaume Croville et al |
| 3095 | USA/NJ-NYUMC344/2020 | 2020-04-08 | NYU Langone Health | Departments of Pathology and Medicine, New York University School of Medicine | Maria Aguero-Rosenfeld et al |
| 3096 | Wales/PHWC-2B078/2020 | 2020-04-08 | Wales Specialist Virology Centre | Public Health Wales Microbiology Cardiff | Catherine Moore et al |
| 3097 | USA/VA-DCLS-0111/2020 | 2020-04-08 | Virginia DCLS | Virginia DCLS | Virginia DCLS et al |
| 3098 | Wales/PHWC-2B281/2020 | 2020-04-08 | Wales Specialist Virology Centre | Public Health Wales Microbiology Cardiff | Catherine Moore et al |
| 3099 | USA/NY-NYUMC337/2020 | 2020-04-08 | NYU Langone Health | Departments of Pathology and Medicine, New York University School of Medicine | Maria Aguero-Rosenfeld et al |
| 3100 | Singapore/59/2020 | 2020-04-08 | National Public Health Laboratory, National Centre for Infectious Diseases | National Public Health Laboratory, National Centre for Infectious Diseases | Mak TM et al |
| 3101 | England/CAMB-7AA8B/2020 | 2020-04-08 | Department of Pathology, University of Cambridge | COVID-19 Genomics UK (COG-UK) Consortium | Luke W Meredith et al |
| 3102 | England/CAMB-7ACD0/2020 | 2020-04-08 | Department of Pathology, University of Cambridge | COVID-19 Genomics UK (COG-UK) Consortium | Luke W Meredith et al |
| 3103 | England/CAMB-7ADBF/2020 | 2020-04-08 | Department of Pathology, University of Cambridge | COVID-19 Genomics UK (COG-UK) Consortium | Luke W Meredith et al |
| 3104 | USA/CA-CZB0142/2020 | 2020-04-08 | UCSF Clinical Microbiology Laboratory | Chan-Zuckerberg Biohub | CZB Cliahub Consortium et al |

| | | | | | |
|---|---|---|---|---|---|
| 3105 | USA/WA-UW-5617/2020 | 2020-04-08 | UW Virology Lab | UW Virology Lab | Pavitra Roychoudhury et al |
| 3106 | USA/WI-UW-290/2020 | 2020-04-08 | University of Wisconsin-Madison AIDS Vaccine Research Laboratories | University of Wisconsin-Madison AIDS Vaccine Research Laboratories | Gage Moreno et al |
| 3107 | USA/CA-CZB0133/2020 | 2020-04-08 | UCSF Clinical Microbiology Laboratory | Chan-Zuckerberg Biohub | CZB Cliahub Consortium et al |
| 3108 | USA/CA-CZB0139/2020 | 2020-04-08 | UCSF Clinical Microbiology Laboratory | Chan-Zuckerberg Biohub | CZB Cliahub Consortium et al |
| 3109 | Italy/TE12759/2020 | 2020-04-08 | Villa Serena del Dr. Leonardo Petruzzi | Istituto Zooprofilattico Sperimentale dell'Abruzzo e Molise "G. Caporale" | Lorusso A et al |
| 3110 | USA/NY-NYUMC347/2020 | 2020-04-08 | NYU Langone Health | Departments of Pathology and Medicine, New York University School of Medicine | Maria Aguero-Rosenfeld et al |
| 3111 | USA/WA-UW-5065/2020 | 2020-04-08 | UW Virology Lab | UW Virology Lab | Pavitra Roychoudhury et al |
| 3112 | Italy/TE13491/2020 | 2020-04-08 | Ospedale SS Annunziata | Istituto Zooprofilattico Sperimentale dell'Abruzzo e Molise "G. Caporale" | Lorusso A et al |
| 3113 | USA/NY-NYUMC346/2020 | 2020-04-08 | NYU Langone Health | Departments of Pathology and Medicine, New York University School of Medicine | Maria Aguero-Rosenfeld et al |
| 3114 | Wales/PHWC-2B254/2020 | 2020-04-08 | Wales Specialist Virology Centre | Public Health Wales Microbiology Cardiff | Catherine Moore et al |
| 3115 | USA/NY-NYUMC348/2020 | 2020-04-08 | NYU Langone Health | Departments of Pathology and Medicine, New York University School of Medicine | Maria Aguero-Rosenfeld et al |
| 3116 | Wales/PHWC-2B236/2020 | 2020-04-08 | Wales Specialist Virology Centre | Public Health Wales Microbiology Cardiff | Catherine Moore et al |
| 3117 | USA/NY-NYUMC335/2020 | 2020-04-08 | NYU Langone Health | Departments of Pathology and Medicine, New York University School of Medicine | Maria Aguero-Rosenfeld et al |
| 3118 | NewZealand/20VR2074/2020-04-08 | 2020-04-08 | Canterbury Health Laboratories | Institute of Environmental Science and Research (ESR) Ltd | **Matt Storey et al** |
| 3119 | Sweden/20-07171/2020 | 2020-04-08 | Hornefors Halsocentral | The Public Health Agency of Sweden | Camilla Eiback et al |
| 3120 | USA/NY-NYUMC359/2020 | 2020-04-08 | NYU Langone Health | Departments of Pathology and Medicine, New York University School of Medicine | Maria Aguero-Rosenfeld et al |
| 3121 | Singapore/56/2020 | 2020-04-08 | National Public Health Laboratory, National Centre for Infectious Diseases | National Public Health Laboratory, National Centre for Infectious Diseases | Mak TM et al |
| 3122 | USA/NY-NYUMC362/2020 | 2020-04-08 | NYU Langone Health | Departments of Pathology and Medicine, New York University School of Medicine | Maria Aguero-Rosenfeld et al |
| 3123 | USA/WA-UW-4897/2020 | 2020-04-08 | UW Virology Lab | UW Virology Lab | Pavitra Roychoudhury et al |
| 3124 | England/CAMB-7AB3D/2020 | 2020-04-08 | Department of Pathology, University of Cambridge | COVID-19 Genomics UK (COG-UK) Consortium | Luke W Meredith et al |
| 3125 | England/CAMB-7AC3A/2020 | 2020-04-08 | Department of Pathology, University of Cambridge | COVID-19 Genomics UK (COG-UK) Consortium | Luke W Meredith et al |
| 3126 | USA/WI-UW-265/2020 | 2020-04-08 | University of Wisconsin-Madison AIDS Vaccine Research Laboratories | University of Wisconsin-Madison AIDS Vaccine Research Laboratories | Gage Moreno et al |
| 3127 | USA/CA-CZB0147/2020 | 2020-04-08 | UCSF Clinical Microbiology Laboratory | Chan-Zuckerberg Biohub | CZB Cliahub Consortium et al |
| 3128 | USA/NY-NYUMC339/2020 | 2020-04-08 | NYU Langone Health | Departments of Pathology and Medicine, New York University School of Medicine | Maria Aguero-Rosenfeld et al |
| 3129 | Belgium/ULG-10134/2020 | 2020-04-08 | Department of Clinical Microbiology | GIGA Medical Genomics | Keith Durkin et al |
| 3130 | USA/NY-NYUMC360/2020 | 2020-04-08 | NYU Langone Health | Departments of Pathology and Medicine, New York University School of Medicine | Maria Aguero-Rosenfeld et al |
| 3131 | USA/UN-UW-5044/2020 | 2020-04-08 | UW Virology Lab | UW Virology Lab | Pavitra Roychoudhury et al |
| 3132 | Belgium/ULG-10142/2020 | 2020-04-08 | Department of Clinical Microbiology | GIGA Medical Genomics | Keith Durkin et al |

| | | | | | |
|---|---|---|---|---|---|
| 3133 | USA/NY-NYUMC349/2020 | 2020-04-08 | NYU Langone Health | Departments of Pathology and Medicine, New York University School of Medicine | Maria Aguero-Rosenfeld et al |
| 3134 | USA/VA-DCLS-0110/2020 | 2020-04-08 | Virginia DCLS | Virginia DCLS | Virginia DCLS et al |
| 3135 | USA/WI-GMF-00822/2020 | 2020-04-08 | Gundersen Molecular Diagnostics Laboratory | Kabara Cancer Research Institute | Craig S. Richmond et al |
| 3136 | Scotland/EDB1106/2020 | 2020-04-08 | Virology Department, Royal Infirmary of Edinburgh, NHS Lothian / School of Biological Sciences, University of Edinburgh / Institute of Genetics and Molecular Medicine, University of Edinburgh | COVID-19 Genomics UK (COG-UK) Consortium | McHugh M et al |
| 3137 | Wales/PHWC-2B0D2/2020 | 2020-04-08 | Wales Specialist Virology Centre | Public Health Wales Microbiology Cardiff | Catherine Moore et al |
| 3138 | USA/WI-UW-267/2020 | 2020-04-08 | University of Wisconsin-Madison AIDS Vaccine Research Laboratories | University of Wisconsin-Madison AIDS Vaccine Research Laboratories | Gage Moreno et al |
| 3139 | England/CAMB-7ACFE/2020 | 2020-04-08 | Department of Pathology, University of Cambridge | COVID-19 Genomics UK (COG-UK) Consortium | Luke W Meredith et al |
| 3140 | Italy/TE13484/2020 | 2020-04-08 | Ospedale SS Annunziata | Istituto Zooprofilattico Sperimentale dell'Abruzzo e Molise "G. Caporale" | Lorusso A et al |
| 3141 | USA/CA-CZB0157/2020 | 2020-04-08 | UCSF Clinical Microbiology Laboratory | Chan-Zuckerberg Biohub | CZB Cliahub Consortium et al |
| 3142 | Scotland/EDB1216/2020 | 2020-04-08 | Virology Department, Royal Infirmary of Edinburgh, NHS Lothian / School of Biological Sciences, University of Edinburgh / Institute of Genetics and Molecular Medicine, University of Edinburgh | COVID-19 Genomics UK (COG-UK) Consortium | McHugh M et al |
| 3143 | Scotland/EDB1161/2020 | 2020-04-08 | Virology Department, Royal Infirmary of Edinburgh, NHS Lothian / School of Biological Sciences, University of Edinburgh / Institute of Genetics and Molecular Medicine, University of Edinburgh | COVID-19 Genomics UK (COG-UK) Consortium | McHugh M et al |
| 3144 | England/CAMB-78917/2020 | 2020-04-08 | Department of Pathology, University of Cambridge | COVID-19 Genomics UK (COG-UK) Consortium | Luke W Meredith et al |
| 3145 | Wales/PHWC-2B2BE/2020 | 2020-04-08 | Wales Specialist Virology Centre | Public Health Wales Microbiology Cardiff | Catherine Moore et al |
| 3146 | USA/CA-CZB0137/2020 | 2020-04-08 | UCSF Clinical Microbiology Laboratory | Chan-Zuckerberg Biohub | CZB Cliahub Consortium et al |
| 3147 | USA/WA-UW-5629/2020 | 2020-04-08 | UW Virology Lab | UW Virology Lab | Pavitra Roychoudhury et al |
| 3148 | USA/CA-CZB0122/2020 | 2020-04-08 | UCSF Clinical Microbiology Laboratory | Chan-Zuckerberg Biohub | CZB Cliahub Consortium et al |
| 3149 | Scotland/EDB1105/2020 | 2020-04-08 | Virology Department, Royal Infirmary of Edinburgh, NHS Lothian / School of Biological Sciences, University of Edinburgh / Institute of Genetics and Molecular Medicine, University of Edinburgh | COVID-19 Genomics UK (COG-UK) Consortium | McHugh M et al |
| 3150 | USA/WI-UW-289/2020 | 2020-04-08 | University of Wisconsin-Madison AIDS Vaccine Research Laboratories | University of Wisconsin-Madison AIDS Vaccine Research Laboratories | Gage Moreno et al |
| 3151 | USA/NY-NYUMC341/2020 | 2020-04-08 | NYU Langone Health | Departments of Pathology and Medicine, New York University School of Medicine | Maria Aguero-Rosenfeld et al |
| 3152 | USA/NY-NYUMC358/2020 | 2020-04-08 | NYU Langone Health | Departments of Pathology and Medicine, New York University School of Medicine | Maria Aguero-Rosenfeld et al |
| 3153 | USA/WI-131/2020 | 2020-04-08 | University of Wisconsin-Madison AIDS | University of Wisconsin-Madison AIDS | Moreno et al |

| | | | | | |
|---|---|---|---|---|---|
| | | | Vaccine Research Laboratories | Vaccine Research Laboratories | |
| 3154 | Scotland/EDB1220/2020 | 2020-04-08 | Virology Department, Royal Infirmary of Edinburgh, NHS Lothian / School of Biological Sciences, University of Edinburgh / Institute of Genetics and Molecular Medicine, University of Edinburgh | COVID-19 Genomics UK (COG-UK) Consortium | McHugh M et al |
| 3155 | USA/NY-NYUMC298/2020 | 2020-04-08 | NYU Langone Health | Departments of Pathology and Medicine, New York University School of Medicine | Maria Aguero-Rosenfeld et al |
| 3156 | Scotland/EDB1107/2020 | 2020-04-08 | Virology Department, Royal Infirmary of Edinburgh, NHS Lothian / School of Biological Sciences, University of Edinburgh / Institute of Genetics and Molecular Medicine, University of Edinburgh | COVID-19 Genomics UK (COG-UK) Consortium | McHugh M et al |
| 3157 | USA/NY-NYUMC338/2020 | 2020-04-08 | NYU Langone Health | Departments of Pathology and Medicine, New York University School of Medicine | Maria Aguero-Rosenfeld et al |
| 3158 | USA/NY-NYUMC332/2020 | 2020-04-08 | NYU Langone Health | Departments of Pathology and Medicine, New York University School of Medicine | Maria Aguero-Rosenfeld et al |
| 3159 | USA/WI-151/2020 | 2020-04-08 | University of Wisconsin-Madison AIDS Vaccine Research Laboratories | University of Wisconsin-Madison AIDS Vaccine Research Laboratories | Moreno et al |
| 3160 | USA/NY-NYUMC356/2020 | 2020-04-08 | NYU Langone Health | Departments of Pathology and Medicine, New York University School of Medicine | Maria Aguero-Rosenfeld et al |
| 3161 | Russia/StPetersburg-RII4936S/2020 | 2020-04-08 | WHO National Influenza Centre Russian Federation | WHO National Influenza Centre Russian Federation | Andrey Komissarov et al |
| 3162 | USA/NY-NYUMC350/2020 | 2020-04-08 | NYU Langone Health | Departments of Pathology and Medicine, New York University School of Medicine | Maria Aguero-Rosenfeld et al |
| 3163 | Russia/StPetersburg-RII4937S/2020 | 2020-04-08 | WHO National Influenza Centre Russian Federation | WHO National Influenza Centre Russian Federation | Andrey Komissarov et al |
| 3164 | USA/WI-UW-263/2020 | 2020-04-08 | University of Wisconsin-Madison AIDS Vaccine Research Laboratories | University of Wisconsin-Madison AIDS Vaccine Research Laboratories | Gage Moreno et al |
| 3165 | Scotland/CVR1935/2020 | 2020-04-08 | West of Scotland Specialist Virology Centre, NHSGGC / MRC-University of Glasgow Centre for Virus Research | COVID-19 Genomics UK (COG-UK) Consortium | Ana da Silva Filipe et al |
| 3166 | USA/NY-NYUMC342/2020 | 2020-04-08 | NYU Langone Health | Departments of Pathology and Medicine, New York University School of Medicine | Maria Aguero-Rosenfeld et al |
| 3167 | USA/WI-UW-291/2020 | 2020-04-08 | University of Wisconsin-Madison AIDS Vaccine Research Laboratories | University of Wisconsin-Madison AIDS Vaccine Research Laboratories | Gage Moreno et al |
| 3168 | NewZealand/20VR2049/2020-04-09 | 2020-04-09 | Southern Community Labs Dunedin | Institute of Environmental Science and Research (ESR) Ltd | **Matt Storey et al** |
| 3169 | Australia/VIC1159/2020 | 2020-04-09 | Victorian Infectious Diseases Reference Laboratory (VIDRL) | Microbiological Diagnostic Unit Public Health Laboratory and Victorian Infectious Diseases Reference Laboratory, The Peter Doherty Institute for Infection and Immunity | Caly L. et al |
| 3170 | Australia/VIC1158/2020 | 2020-04-09 | Victorian Infectious Diseases Reference Laboratory (VIDRL) | Microbiological Diagnostic Unit Public Health Laboratory and Victorian Infectious Diseases Reference Laboratory, The Peter Doherty Institute for Infection and Immunity | Caly L. et al |
| 3171 | Australia/VIC1284/2020 | 2020-04-09 | Victorian Infectious Diseases Reference Laboratory (VIDRL) | Microbiological Diagnostic Unit Public Health Laboratory and Victorian Infectious Diseases Reference Laboratory, The Peter Doherty Institute for Infection and Immunity | Caly L. et al |

| | | | | | |
|---|---|---|---|---|---|
| 3172 | Australia/VIC1157/2020 | 2020-04-09 | Victorian Infectious Diseases Reference Laboratory (VIDRL) | Microbiological Diagnostic Unit Public Health Laboratory and Victorian Infectious Diseases Reference Laboratory, The Peter Doherty Institute for Infection and Immunity | Caly L. et al |
| 3173 | Australia/VIC1161/2020 | 2020-04-09 | Victorian Infectious Diseases Reference Laboratory (VIDRL) | Microbiological Diagnostic Unit Public Health Laboratory and Victorian Infectious Diseases Reference Laboratory, The Peter Doherty Institute for Infection and Immunity | Caly L. et al |
| 3174 | USA/WI-136/2020 | 2020-04-09 | University of Wisconsin-Madison AIDS Vaccine Research Laboratories | University of Wisconsin-Madison AIDS Vaccine Research Laboratories | Moreno et al |
| 3175 | Italy/TE13858/2020 | 2020-04-09 | Servizio di Igiene, Epidemiologia e Sanità Pubblica (SIESP) Avezzano | Istituto Zooprofilattico Sperimentale dell'Abruzzo e Molise "G. Caporale" | Lorusso A et al |
| 3176 | USA/WI-UW-294/2020 | 2020-04-09 | University of Wisconsin-Madison AIDS Vaccine Research Laboratories | University of Wisconsin-Madison AIDS Vaccine Research Laboratories | Gage Moreno et al |
| 3177 | USA/NJ-NYUMC367/2020 | 2020-04-09 | NYU Langone Health | Departments of Pathology and Medicine, New York University School of Medicine | Maria Aguero-Rosenfeld et al |
| 3178 | India/NCDC-3394/2020 | 2020-04-09 | National Centre for Disease control (NCDC) | NCDC/CSIR-IGIB | Pramod Kumar et al |
| 3179 | USA/CA-CZB-1015/2020 | 2020-04-09 | County of Santa Clara Public Health Department | Chan-Zuckerberg Biohub | CZB Cliahub Consortium et al |
| 3180 | Italy/TE14168/2020 | 2020-04-09 | SERVIZIO DI IGIENE E SANITÀ PUBBLICA ASL Teramo | Istituto Zooprofilattico Sperimentale dell'Abruzzo e Molise "G. Caporale" | Lorusso A et al |
| 3181 | USA/WI-137/2020 | 2020-04-09 | University of Wisconsin-Madison AIDS Vaccine Research Laboratories | University of Wisconsin-Madison AIDS Vaccine Research Laboratories | Moreno et al |
| 3182 | Italy/TE14162/2020 | 2020-04-09 | SERVIZIO DI IGIENE E SANITÀ PUBBLICA ASL Teramo | Istituto Zooprofilattico Sperimentale dell'Abruzzo e Molise "G. Caporale" | Lorusso A et al |
| 3183 | India/NCDC-3965/2020 | 2020-04-09 | National Centre for Disease control (NCDC) | NCDC/CSIR-IGIB | Pramod Kumar et al |
| 3184 | USA/NY-NYUMC366/2020 | 2020-04-09 | NYU Langone Health | Departments of Pathology and Medicine, New York University School of Medicine | Maria Aguero-Rosenfeld et al |
| 3185 | USA/CA-CZB-1012/2020 | 2020-04-09 | County of Santa Clara Public Health Department | Chan-Zuckerberg Biohub | CZB Cliahub Consortium et al |
| 3186 | England/CAMB-7AFAA/2020 | 2020-04-09 | Department of Pathology, University of Cambridge | COVID-19 Genomics UK (COG-UK) Consortium | Luke W Meredith et al |
| 3187 | USA/NY-NYUMC357/2020 | 2020-04-09 | NYU Langone Health | Departments of Pathology and Medicine, New York University School of Medicine | Maria Aguero-Rosenfeld et al |
| 3188 | USA/NY-NYUMC364/2020 | 2020-04-09 | NYU Langone Health | Departments of Pathology and Medicine, New York University School of Medicine | Maria Aguero-Rosenfeld et al |
| 3189 | USA/WI-140/2020 | 2020-04-09 | University of Wisconsin-Madison AIDS Vaccine Research Laboratories | University of Wisconsin-Madison AIDS Vaccine Research Laboratories | Moreno et al |
| 3190 | USA/NY-NYUMC381/2020 | 2020-04-09 | NYU Langone Health | Departments of Pathology and Medicine, New York University School of Medicine | Maria Aguero-Rosenfeld et al |
| 3191 | USA/NY-NYUMC384/2020 | 2020-04-09 | NYU Langone Health | Departments of Pathology and Medicine, New York University School of Medicine | Maria Aguero-Rosenfeld et al |
| 3192 | France/OCC-18/2020 | 2020-04-09 | CHU Purpan - Laboratoire de Virologie - Institut Fédératif de Biologie | Laboratoire de virologie - École Nationale Vétérinaire de Toulouse | Guillaume Croville et al |
| 3193 | USA/WI-153/2020 | 2020-04-09 | University of Wisconsin-Madison AIDS Vaccine Research Laboratories | University of Wisconsin-Madison AIDS Vaccine Research Laboratories | Moreno et al |
| 3194 | England/CAMB-7ABF1/2020 | 2020-04-09 | Department of Pathology, University of Cambridge | COVID-19 Genomics UK (COG-UK) Consortium | Luke W Meredith et al |
| 3195 | USA/NY-NYUMC368/2020 | 2020-04-09 | NYU Langone Health | Departments of Pathology and | Maria |

| | | | | | |
|---|---|---|---|---|---|
| | | | | Medicine, New York University School of Medicine | Aguero-Rosenfeld et al |
| 3196 | USA/WI-GMF-00857/2020 | 2020-04-09 | Gundersen Molecular Diagnostics Laboratory | Kabara Cancer Research Institute | Craig S. Richmond et al |
| 3197 | France/OCC-17/2020 | 2020-04-09 | CHU Purpan - Laboratoire de Virologie - Institut Fédératif de Biologie | Laboratoire de virologie - École Nationale Vétérinaire de Toulouse | Guillaume Croville et al |
| 3198 | NewZealand/20VR2077/2020-04-09 | 2020-04-09 | Canterbury Health Laboratories | Institute of Environmental Science and Research (ESR) Ltd | **Matt Storey et al** |
| 3199 | NewZealand/20VR2078/2020-04-09 | 2020-04-09 | Canterbury Health Laboratories | Institute of Environmental Science and Research (ESR) Ltd | **Matt Storey et al** |
| 3200 | NewZealand/20VR2076/2020-04-09 | 2020-04-09 | Canterbury Health Laboratories | Institute of Environmental Science and Research (ESR) Ltd | **Matt Storey et al** |
| 3201 | NewZealand/20VR2075/2020-04-09 | 2020-04-09 | Canterbury Health Laboratories | Institute of Environmental Science and Research (ESR) Ltd | **Matt Storey et al** |
| 3202 | USA/CA-CZB-1016/2020 | 2020-04-09 | County of Santa Clara Public Health Department | Chan-Zuckerberg Biohub | CZB Cliahub Consortium et al |
| 3203 | USA/NY-NYUMC361/2020 | 2020-04-09 | NYU Langone Health | Departments of Pathology and Medicine, New York University School of Medicine | Maria Aguero-Rosenfeld et al |
| 3204 | USA/NY-NYUMC386/2020 | 2020-04-09 | NYU Langone Health | Departments of Pathology and Medicine, New York University School of Medicine | Maria Aguero-Rosenfeld et al |
| 3205 | Russia/StPetersburg-RII5017V/2020 | 2020-04-09 | WHO National Influenza Centre Russian Federation | WHO National Influenza Centre Russian Federation | Andrey Komissarov et al |
| 3206 | USA/WI-135/2020 | 2020-04-09 | University of Wisconsin-Madison AIDS Vaccine Research Laboratories | University of Wisconsin-Madison AIDS Vaccine Research Laboratories | Moreno et al |
| 3207 | Russia/StPetersburg-RII5170S/2020 | 2020-04-09 | WHO National Influenza Centre Russian Federation | WHO National Influenza Centre Russian Federation | Andrey Komissarov et al |
| 3208 | England/CAMB-7AE70/2020 | 2020-04-09 | Department of Pathology, University of Cambridge | COVID-19 Genomics UK (COG-UK) Consortium | Luke W Meredith et al |
| 3209 | USA/NY-NYUMC355/2020 | 2020-04-09 | NYU Langone Health | Departments of Pathology and Medicine, New York University School of Medicine | Maria Aguero-Rosenfeld et al |
| 3210 | Scotland/EDB1317/2020 | 2020-04-09 | Virology Department, Royal Infirmary of Edinburgh, NHS Lothian / School of Biological Sciences, University of Edinburgh / Institute of Genetics and Molecular Medicine, University of Edinburgh | COVID-19 Genomics UK (COG-UK) Consortium | McHugh M et al |
| 3211 | USA/CA-CZB-1020/2020 | 2020-04-09 | County of Santa Clara Public Health Department | Chan-Zuckerberg Biohub | CZB Cliahub Consortium et al |
| 3212 | USA/LA-EVTL01/2020 | 2020-04-09 | LSUHS Emerging Viral Threat Laboratory | Microbial Genome Sequencing Center | Jeremy P. Kamil et al |
| 3213 | USA/AK-PHL193/2020 | 2020-04-09 | Alaska State Virology Laboratory | Alaska State Virology Laboratory | Jack Chen et al |
| 3214 | USA/CA-CZB-1017/2020 | 2020-04-09 | County of Santa Clara Public Health Department | Chan-Zuckerberg Biohub | CZB Cliahub Consortium et al |
| 3215 | India/NCDC-3936/2020 | 2020-04-09 | National Centre for Disease control (NCDC) | NCDC/CSIR-IGIB | Pramod Kumar et al |
| 3216 | USA/VA-DCLS-0095/2020 | 2020-04-09 | Virginia DCLS | Virginia DCLS | Virginia DCLS et al |
| 3217 | England/CAMB-7ACC1/2020 | 2020-04-09 | Department of Pathology, University of Cambridge | COVID-19 Genomics UK (COG-UK) Consortium | Luke W Meredith et al |
| 3218 | USA/WI-UW-296/2020 | 2020-04-09 | University of Wisconsin-Madison AIDS Vaccine Research Laboratories | University of Wisconsin-Madison AIDS Vaccine Research Laboratories | Gage Moreno et al |
| 3219 | Luxembourg/LNS1703112/2020 | 2020-04-09 | Laboratoire National de Sante, Microbiology, Virology | Laboratoire National de Sante, Microbiology, Epidemiology and Microbial Genomics | Anke Wienecke-Baldacchino et al |
| 3220 | England/CAMB-7AE52/2020 | 2020-04-09 | Department of Pathology, University of Cambridge | COVID-19 Genomics UK (COG-UK) Consortium | Luke W Meredith et al |
| 3221 | USA/NY-NYUMC370/2020 | 2020-04-09 | NYU Langone Health | Departments of Pathology and Medicine, New York University School of Medicine | Maria Aguero-Rosenfeld et al |
| 3222 | USA/WI-UW-293/2020 | 2020-04-09 | University of Wisconsin-Madison AIDS | University of Wisconsin-Madison AIDS | Gage Moreno et al |

| | | | | | |
|---|---|---|---|---|---|
| | | | Vaccine Research Laboratories | Vaccine Research Laboratories | |
| 3223 | England/CAMB-7ADEC/2020 | 2020-04-09 | Department of Pathology, University of Cambridge | COVID-19 Genomics UK (COG-UK) Consortium | Luke W Meredith et al |
| 3224 | USA/NJ-NYUMC376/2020 | 2020-04-10 | NYU Langone Health | Departments of Pathology and Medicine, New York University School of Medicine | Maria Aguero-Rosenfeld et al |
| 3225 | USA/NY-NYUMC297/2020 | 2020-04-10 | NYU Langone Health | Departments of Pathology and Medicine, New York University School of Medicine | Maria Aguero-Rosenfeld et al |
| 3226 | Australia/VIC1313/2020 | 2020-04-10 | Victorian Infectious Diseases Reference Laboratory (VIDRL) | Microbiological Diagnostic Unit Public Health Laboratory and Victorian Infectious Diseases Reference Laboratory, The Peter Doherty Institute for Infection and Immunity | Caly L. et al |
| 3227 | Australia/VIC1300/2020 | 2020-04-10 | Microbiological Diagnostic Unit Public Health Laboratory | Microbiological Diagnostic Unit Public Health Laboratory | Seemann T. et al |
| 3228 | Australia/VIC1305/2020 | 2020-04-10 | Victorian Infectious Diseases Reference Laboratory (VIDRL) | Microbiological Diagnostic Unit Public Health Laboratory and Victorian Infectious Diseases Reference Laboratory, The Peter Doherty Institute for Infection and Immunity | Caly L. et al |
| 3229 | Australia/VIC1178/2020 | 2020-04-10 | Microbiological Diagnostic Unit Public Health Laboratory | Microbiological Diagnostic Unit Public Health Laboratory | Seemann T. et al |
| 3230 | USA/NY-NYUMC372/2020 | 2020-04-10 | NYU Langone Health | Departments of Pathology and Medicine, New York University School of Medicine | Maria Aguero-Rosenfeld et al |
| 3231 | USA/NY-NYUMC387/2020 | 2020-04-10 | NYU Langone Health | Departments of Pathology and Medicine, New York University School of Medicine | Maria Aguero-Rosenfeld et al |
| 3232 | Scotland/EDB1340/2020 | 2020-04-10 | Virology Department, Royal Infirmary of Edinburgh, NHS Lothian / School of Biological Sciences, University of Edinburgh / Institute of Genetics and Molecular Medicine, University of Edinburgh | COVID-19 Genomics UK (COG-UK) Consortium | McHugh M et al |
| 3233 | England/CAMB-7AF6E/2020 | 2020-04-10 | Department of Pathology, University of Cambridge | COVID-19 Genomics UK (COG-UK) Consortium | Luke W Meredith et al |
| 3234 | Luxembourg/LNS1658150/2020 | 2020-04-10 | Laboratoire National de Sante, Microbiology, Virology | Laboratoire National de Sante, Microbiology, Epidemiology and Microbial Genomics | Anke Wienecke-Baldacchino et al |
| 3235 | England/CAMB-7B0F1/2020 | 2020-04-10 | Department of Pathology, University of Cambridge | COVID-19 Genomics UK (COG-UK) Consortium | Luke W Meredith et al |
| 3236 | England/CAMB-7B2A0/2020 | 2020-04-10 | Department of Pathology, University of Cambridge | COVID-19 Genomics UK (COG-UK) Consortium | Luke W Meredith et al |
| 3237 | USA/VA-DCLS-0101/2020 | 2020-04-10 | Virginia DCLS | Virginia DCLS | Virginia DCLS et al |
| 3238 | Scotland/EDB1324/2020 | 2020-04-10 | Virology Department, Royal Infirmary of Edinburgh, NHS Lothian / School of Biological Sciences, University of Edinburgh / Institute of Genetics and Molecular Medicine, University of Edinburgh | COVID-19 Genomics UK (COG-UK) Consortium | McHugh M et al |
| 3239 | USA/NY-NYUMC377/2020 | 2020-04-10 | NYU Langone Health | Departments of Pathology and Medicine, New York University School of Medicine | Maria Aguero-Rosenfeld et al |
| 3240 | USA/WI-UW-262/2020 | 2020-04-10 | University of Wisconsin-Madison AIDS Vaccine Research Laboratories | University of Wisconsin-Madison AIDS Vaccine Research Laboratories | Gage Moreno et al |
| 3241 | USA/NY-NYUMC388/2020 | 2020-04-10 | NYU Langone Health | Departments of Pathology and Medicine, New York University School of Medicine | Maria Aguero-Rosenfeld et al |
| 3242 | USA/WI-145/2020 | 2020-04-10 | University of Wisconsin-Madison AIDS Vaccine Research Laboratories | University of Wisconsin-Madison AIDS Vaccine Research Laboratories | Moreno et al |
| 3243 | USA/NY-NYUMC296/2020 | 2020-04-10 | NYU Langone Health | Departments of Pathology and Medicine, New York University | Maria Aguero-Rosenfeld et |

| | | | | | |
|---|---|---|---|---|---|
| | | | | School of Medicine | al |
| 3244 | USA/NY-NYUMC371/2020 | 2020-04-10 | NYU Langone Health | Departments of Pathology and Medicine, New York University School of Medicine | Maria Aguero-Rosenfeld et al |
| 3245 | USA/WI-142/2020 | 2020-04-10 | University of Wisconsin-Madison AIDS Vaccine Research Laboratories | University of Wisconsin-Madison AIDS Vaccine Research Laboratories | Moreno et al |
| 3246 | USA/NY-NYUMC369/2020 | 2020-04-10 | NYU Langone Health | Departments of Pathology and Medicine, New York University School of Medicine | Maria Aguero-Rosenfeld et al |
| 3247 | USA/NY-NYUMC375/2020 | 2020-04-10 | NYU Langone Health | Departments of Pathology and Medicine, New York University School of Medicine | Maria Aguero-Rosenfeld et al |
| 3248 | USA/VA-DCLS-0098/2020 | 2020-04-10 | Virginia DCLS | Virginia DCLS | Virginia DCLS et al |
| 3249 | USA/CT-NYUMC374/2020 | 2020-04-10 | NYU Langone Health | Departments of Pathology and Medicine, New York University School of Medicine | Maria Aguero-Rosenfeld et al |
| 3250 | England/CAMB-7AF7D/2020 | 2020-04-10 | Department of Pathology, University of Cambridge | COVID-19 Genomics UK (COG-UK) Consortium | Luke W Meredith et al |
| 3251 | England/CAMB-7AF13/2020 | 2020-04-10 | Department of Pathology, University of Cambridge | COVID-19 Genomics UK (COG-UK) Consortium | Luke W Meredith et al |
| 3252 | USA/CA-CZB-1024/2020 | 2020-04-10 | County of Santa Clara Public Health Department | Chan-Zuckerberg Biohub | CZB Cliahub Consortium et al |
| 3253 | England/CAMB-7B194/2020 | 2020-04-10 | Department of Pathology, University of Cambridge | COVID-19 Genomics UK (COG-UK) Consortium | Luke W Meredith et al |
| 3254 | NewZealand/20VR2079/2020-04-10 | 2020-04-10 | Canterbury Health Laboratories | Institute of Environmental Science and Research (ESR) Ltd | **Matt Storey et al** |
| 3255 | NewZealand/20VR2045/2020-04-10 | 2020-04-10 | Southern Community Labs Dunedin | Institute of Environmental Science and Research (ESR) Ltd | **Matt Storey et al** |
| 3256 | NewZealand/20VR2093/2020-04-10 | 2020-04-10 | Canterbury Health Laboratories | Institute of Environmental Science and Research (ESR) Ltd | **Matt Storey et al** |
| 3257 | Russia/StPetersburg-RII4980S/2020 | 2020-04-10 | WHO National Influenza Centre Russian Federation | WHO National Influenza Centre Russian Federation | Andrey Komissarov et al |
| 3258 | USA/NY-NYUMC385/2020 | 2020-04-10 | NYU Langone Health | Departments of Pathology and Medicine, New York University School of Medicine | Maria Aguero-Rosenfeld et al |
| 3259 | England/CAMB-7AF31/2020 | 2020-04-10 | Department of Pathology, University of Cambridge | COVID-19 Genomics UK (COG-UK) Consortium | Luke W Meredith et al |
| 3260 | USA/NY-NYUMC380/2020 | 2020-04-10 | NYU Langone Health | Departments of Pathology and Medicine, New York University School of Medicine | Maria Aguero-Rosenfeld et al |
| 3261 | USA/NY-NYUMC379/2020 | 2020-04-10 | NYU Langone Health | Departments of Pathology and Medicine, New York University School of Medicine | Maria Aguero-Rosenfeld et al |
| 3262 | USA/NY-NYUMC382/2020 | 2020-04-10 | NYU Langone Health | Departments of Pathology and Medicine, New York University School of Medicine | Maria Aguero-Rosenfeld et al |
| 3263 | USA/NY-NYUMC383/2020 | 2020-04-10 | NYU Langone Health | Departments of Pathology and Medicine, New York University School of Medicine | Maria Aguero-Rosenfeld et al |
| 3264 | Russia/StPetersburg-RII5029S/2020 | 2020-04-10 | WHO National Influenza Centre Russian Federation | WHO National Influenza Centre Russian Federation | Andrey Komissarov et al |
| 3265 | USA/WI-147/2020 | 2020-04-10 | University of Wisconsin-Madison AIDS Vaccine Research Laboratories | University of Wisconsin-Madison AIDS Vaccine Research Laboratories | Moreno et al |
| 3266 | England/CAMB-7B000/2020 | 2020-04-10 | Department of Pathology, University of Cambridge | COVID-19 Genomics UK (COG-UK) Consortium | Luke W Meredith et al |
| 3267 | England/CAMB-7AF04/2020 | 2020-04-10 | Department of Pathology, University of Cambridge | COVID-19 Genomics UK (COG-UK) Consortium | Luke W Meredith et al |
| 3268 | USA/NY-NYUMC373/2020 | 2020-04-10 | NYU Langone Health | Departments of Pathology and Medicine, New York University School of Medicine | Maria Aguero-Rosenfeld et al |
| 3269 | USA/VA-DCLS-0100/2020 | 2020-04-10 | Virginia DCLS | Virginia DCLS | Virginia DCLS et al |
| 3270 | USA/CA-CZB-1019/2020 | 2020-04-10 | County of Santa Clara Public Health Department | Chan-Zuckerberg Biohub | CZB Cliahub Consortium et al |
| 3271 | Russia/StPetersburg-RII4984S/2020 | 2020-04-10 | WHO National Influenza Centre Russian Federation | WHO National Influenza Centre Russian Federation | Andrey Komissarov et al |
| 3272 | Russia/StPetersburg-RII4985S/2020 | 2020-04-10 | WHO National Influenza | WHO National Influenza Centre | Andrey Komissarov et |

| | | | | | |
|---|---|---|---|---|---|
| | | | Centre Russian Federation | Russian Federation | al |
| 3273 | Luxembourg/LNS0361493/2020 | 2020-04-10 | Laboratoire National de Sante, Microbiology, Virology | Laboratoire National de Sante, Microbiology, Epidemiology and Microbial Genomics | Anke Wienecke-Baldacchino et al |
| 3274 | USA/WI-132/2020 | 2020-04-10 | University of Wisconsin-Madison AIDS Vaccine Research Laboratories | University of Wisconsin-Madison AIDS Vaccine Research Laboratories | Moreno et al |
| 3275 | Russia/StPetersburg-RII4983S/2020 | 2020-04-10 | WHO National Influenza Centre Russian Federation | WHO National Influenza Centre Russian Federation | Andrey Komissarov et al |
| 3276 | Russia/StPetersburg-RII4988S/2020 | 2020-04-10 | WHO National Influenza Centre Russian Federation | WHO National Influenza Centre Russian Federation | Andrey Komissarov et al |
| 3277 | England/CAMB-7AF22/2020 | 2020-04-10 | Department of Pathology, University of Cambridge | COVID-19 Genomics UK (COG-UK) Consortium | Luke W Meredith et al |
| 3278 | USA/NJ-NYUMC378/2020 | 2020-04-10 | NYU Langone Health | Departments of Pathology and Medicine, New York University School of Medicine | Maria Aguero-Rosenfeld et al |
| 3279 | USA/CA-CZB-1018/2020 | 2020-04-10 | County of Santa Clara Public Health Department | Chan-Zuckerberg Biohub | CZB Cliahub Consortium et al |
| 3280 | Singapore/80/2020 | 2020-04-11 | National Public Health Laboratory, National Centre for Infectious Diseases | National Public Health Laboratory, National Centre for Infectious Diseases | Mak Tze Minn et al |
| 3281 | Australia/VIC1316/2020 | 2020-04-11 | Victorian Infectious Diseases Reference Laboratory (VIDRL) | Microbiological Diagnostic Unit Public Health Laboratory and Victorian Infectious Diseases Reference Laboratory, The Peter Doherty Institute for Infection and Immunity | Caly L. et al |
| 3282 | Australia/VIC1315/2020 | 2020-04-11 | Victorian Infectious Diseases Reference Laboratory (VIDRL) | Microbiological Diagnostic Unit Public Health Laboratory and Victorian Infectious Diseases Reference Laboratory, The Peter Doherty Institute for Infection and Immunity | Caly L. et al |
| 3283 | Australia/VIC1318/2020 | 2020-04-11 | Victorian Infectious Diseases Reference Laboratory (VIDRL) | Microbiological Diagnostic Unit Public Health Laboratory and Victorian Infectious Diseases Reference Laboratory, The Peter Doherty Institute for Infection and Immunity | Caly L. et al |
| 3284 | USA/WI-121/2020 | 2020-04-11 | University of Wisconsin-Madison AIDS Vaccine Research Laboratories | University of Wisconsin-Madison AIDS Vaccine Research Laboratories | Moreno et al |
| 3285 | France/OCC-19/2020 | 2020-04-11 | CHU Purpan - Laboratoire de Virologie - Institut Fédératif de Biologie | Laboratoire de virologie - École Nationale Vétérinaire de Toulouse | Guillaume Croville et al |
| 3286 | USA/LA-EVTL03/2020 | 2020-04-11 | LSUHS Emerging Viral Threat Laboratory | Microbial Genome Sequencing Center | Jeremy P. Kamil et al |
| 3287 | USA/WI-UW-295/2020 | 2020-04-11 | University of Wisconsin-Madison AIDS Vaccine Research Laboratories | University of Wisconsin-Madison AIDS Vaccine Research Laboratories | Gage Moreno et al |
| 3288 | USA/VA-DCLS-0099/2020 | 2020-04-11 | Virginia DCLS | Virginia DCLS | Virginia DCLS et al |
| 3289 | USA/WI-126/2020 | 2020-04-11 | University of Wisconsin-Madison AIDS Vaccine Research Laboratories | University of Wisconsin-Madison AIDS Vaccine Research Laboratories | Moreno et al |
| 3290 | USA/WI-123/2020 | 2020-04-11 | University of Wisconsin-Madison AIDS Vaccine Research Laboratories | University of Wisconsin-Madison AIDS Vaccine Research Laboratories | Moreno et al |
| 3291 | England/CAMB-7B237/2020 | 2020-04-11 | Department of Pathology, University of Cambridge | COVID-19 Genomics UK (COG-UK) Consortium | Luke W Meredith et al |
| 3292 | Singapore/79/2020 | 2020-04-11 | National Public Health Laboratory, National Centre for Infectious Diseases | National Public Health Laboratory, National Centre for Infectious Diseases | Mak Tze Minn et al |
| 3293 | USA/NY-NYUMC396/2020 | 2020-04-11 | NYU Langone Health | Departments of Pathology and Medicine, New York University School of Medicine | Maria Aguero-Rosenfeld et al |
| 3294 | Scotland/EDB1457/2020 | 2020-04-11 | Virology Department, Royal Infirmary of Edinburgh, NHS Lothian / School of Biological Sciences, University of Edinburgh / Institute of Genetics and Molecular | COVID-19 Genomics UK (COG-UK) Consortium | McHugh M et al |

| | | | | | |
|---|---|---|---|---|---|
| | | | Medicine, University of Edinburgh | | |
| 3295 | England/CAMB-7B088/2020 | 2020-04-11 | Department of Pathology, University of Cambridge | COVID-19 Genomics UK (COG-UK) Consortium | Luke W Meredith et al |
| 3296 | USA/CA-CZB-1026/2020 | 2020-04-11 | County of Santa Clara Public Health Department | Chan-Zuckerberg Biohub | CZB Cliahub Consortium et al |
| 3297 | USA/LA-EVTL02/2020 | 2020-04-11 | LSUHS Emerging Viral Threat Laboratory | Microbial Genome Sequencing Center | Jeremy P. Kamil et al |
| 3298 | England/CAMB-7B1B2/2020 | 2020-04-11 | Department of Pathology, University of Cambridge | COVID-19 Genomics UK (COG-UK) Consortium | Luke W Meredith et al |
| 3299 | England/CAMB-7B0C4/2020 | 2020-04-11 | Department of Pathology, University of Cambridge | COVID-19 Genomics UK (COG-UK) Consortium | Luke W Meredith et al |
| 3300 | France/OCC-20/2020 | 2020-04-11 | CHU Purpan - Laboratoire de Virologie - Institut Fédératif de Biologie | Laboratoire de virologie - École Nationale Vétérinaire de Toulouse | Guillaume Croville et al |
| 3301 | NewZealand/20VR2081/2020-04-11 | 2020-04-11 | Canterbury Health Laboratories | Institute of Environmental Science and Research (ESR) Ltd | **Matt Storey et al** |
| 3302 | England/CAMB-7B01F/2020 | 2020-04-11 | Department of Pathology, University of Cambridge | COVID-19 Genomics UK (COG-UK) Consortium | Luke W Meredith et al |
| 3303 | Scotland/EDB1470/2020 | 2020-04-11 | Virology Department, Royal Infirmary of Edinburgh, NHS Lothian / School of Biological Sciences, University of Edinburgh / Institute of Genetics and Molecular Medicine, University of Edinburgh | COVID-19 Genomics UK (COG-UK) Consortium | McHugh M et al |
| 3304 | England/CAMB-7B0B5/2020 | 2020-04-11 | Department of Pathology, University of Cambridge | COVID-19 Genomics UK (COG-UK) Consortium | Luke W Meredith et al |
| 3305 | Luxembourg/LNS1368952/2020 | 2020-04-11 | Laboratoire National de Sante, Microbiology, Virology | Laboratoire National de Sante, Microbiology, Epidemiology and Microbial Genomics | Anke Wienecke-Baldacchino et al |
| 3306 | England/CAMB-7B10D/2020 | 2020-04-11 | Department of Pathology, University of Cambridge | COVID-19 Genomics UK (COG-UK) Consortium | Luke W Meredith et al |
| 3307 | USA/CA-CZB-1023/2020 | 2020-04-11 | County of Santa Clara Public Health Department | Chan-Zuckerberg Biohub | CZB Cliahub Consortium et al |
| 3308 | England/CAMB-7B325/2020 | 2020-04-11 | Department of Pathology, University of Cambridge | COVID-19 Genomics UK (COG-UK) Consortium | Luke W Meredith et al |
| 3309 | England/CAMB-7B2EC/2020 | 2020-04-11 | Department of Pathology, University of Cambridge | COVID-19 Genomics UK (COG-UK) Consortium | Luke W Meredith et al |
| 3310 | England/CAMB-7B11C/2020 | 2020-04-11 | Department of Pathology, University of Cambridge | COVID-19 Genomics UK (COG-UK) Consortium | Luke W Meredith et al |
| 3311 | England/CAMB-7B1A3/2020 | 2020-04-11 | Department of Pathology, University of Cambridge | COVID-19 Genomics UK (COG-UK) Consortium | Luke W Meredith et al |
| 3312 | USA/CA-CZB-1025/2020 | 2020-04-11 | County of Santa Clara Public Health Department | Chan-Zuckerberg Biohub | CZB Cliahub Consortium et al |
| 3313 | England/CAMB-7B149/2020 | 2020-04-11 | Department of Pathology, University of Cambridge | COVID-19 Genomics UK (COG-UK) Consortium | Luke W Meredith et al |
| 3314 | USA/CA-CZB-1021/2020 | 2020-04-11 | County of Santa Clara Public Health Department | Chan-Zuckerberg Biohub | CZB Cliahub Consortium et al |
| 3315 | England/CAMB-7B079/2020 | 2020-04-11 | Department of Pathology, University of Cambridge | COVID-19 Genomics UK (COG-UK) Consortium | Luke W Meredith et al |
| 3316 | Scotland/EDB1471/2020 | 2020-04-11 | Virology Department, Royal Infirmary of Edinburgh, NHS Lothian / School of Biological Sciences, University of Edinburgh / Institute of Genetics and Molecular Medicine, University of Edinburgh | COVID-19 Genomics UK (COG-UK) Consortium | McHugh M et al |
| 3317 | Australia/VIC1329/2020 | 2020-04-12 | Victorian Infectious Diseases Reference Laboratory (VIDRL) | Microbiological Diagnostic Unit Public Health Laboratory and Victorian Infectious Diseases Reference Laboratory, The Peter Doherty Institute for Infection and Immunity | Caly L. et al |
| 3318 | Australia/VIC1328/2020 | 2020-04-12 | Victorian Infectious Diseases Reference Laboratory (VIDRL) | Microbiological Diagnostic Unit Public Health Laboratory and Victorian Infectious Diseases Reference Laboratory, The Peter | Caly L. et al |

| | | | | | |
|---|---|---|---|---|---|
| | | | | Doherty Institute for Infection and Immunity | |
| 3319 | USA/WI-154/2020 | 2020-04-12 | University of Wisconsin-Madison AIDS Vaccine Research Laboratories | University of Wisconsin-Madison AIDS Vaccine Research Laboratories | Moreno et al |
| 3320 | England/CAMB-7B1D0/2020 | 2020-04-12 | Department of Pathology, University of Cambridge | COVID-19 Genomics UK (COG-UK) Consortium | Luke W Meredith et al |
| 3321 | USA/NY-NYUMC407/2020 | 2020-04-12 | NYU Langone Health | Departments of Pathology and Medicine, New York University School of Medicine | Maria Aguero-Rosenfeld et al |
| 3322 | USA/OH-OSUP0024/2020 | 2020-04-12 | OSU Wexner Medical Center | James Molecular Lab - OSUWMC | Huolin Tu et al |
| 3323 | Scotland/EDB1510/2020 | 2020-04-12 | Virology Department, Royal Infirmary of Edinburgh, NHS Lothian / School of Biological Sciences, University of Edinburgh / Institute of Genetics and Molecular Medicine, University of Edinburgh | COVID-19 Genomics UK (COG-UK) Consortium | McHugh M et al |
| 3324 | England/CAMB-7B291/2020 | 2020-04-12 | Department of Pathology, University of Cambridge | COVID-19 Genomics UK (COG-UK) Consortium | Luke W Meredith et al |
| 3325 | Scotland/EDB1428/2020 | 2020-04-12 | Virology Department, Royal Infirmary of Edinburgh, NHS Lothian / School of Biological Sciences, University of Edinburgh / Institute of Genetics and Molecular Medicine, University of Edinburgh | COVID-19 Genomics UK (COG-UK) Consortium | McHugh M et al |
| 3326 | England/CAMB-7B307/2020 | 2020-04-12 | Department of Pathology, University of Cambridge | COVID-19 Genomics UK (COG-UK) Consortium | Luke W Meredith et al |
| 3327 | Scotland/EDB1568/2020 | 2020-04-12 | Virology Department, Royal Infirmary of Edinburgh, NHS Lothian / School of Biological Sciences, University of Edinburgh / Institute of Genetics and Molecular Medicine, University of Edinburgh | COVID-19 Genomics UK (COG-UK) Consortium | McHugh M et al |
| 3328 | Scotland/EDB1424/2020 | 2020-04-12 | Virology Department, Royal Infirmary of Edinburgh, NHS Lothian / School of Biological Sciences, University of Edinburgh / Institute of Genetics and Molecular Medicine, University of Edinburgh | COVID-19 Genomics UK (COG-UK) Consortium | McHugh M et al |
| 3329 | USA/AK-PHL200/2020 | 2020-04-12 | Alaska State Virology Laboratory | Alaska State Virology Laboratory | Jack Chen et al |
| 3330 | Scotland/EDB1565/2020 | 2020-04-12 | Virology Department, Royal Infirmary of Edinburgh, NHS Lothian / School of Biological Sciences, University of Edinburgh / Institute of Genetics and Molecular Medicine, University of Edinburgh | COVID-19 Genomics UK (COG-UK) Consortium | McHugh M et al |
| 3331 | USA/WI-138/2020 | 2020-04-12 | University of Wisconsin-Madison AIDS Vaccine Research Laboratories | University of Wisconsin-Madison AIDS Vaccine Research Laboratories | Moreno et al |
| 3332 | USA/WI-UW-264/2020 | 2020-04-12 | University of Wisconsin-Madison AIDS Vaccine Research Laboratories | University of Wisconsin-Madison AIDS Vaccine Research Laboratories | Gage Moreno et al |
| 3333 | England/CAMB-7B404/2020 | 2020-04-12 | Department of Pathology, University of Cambridge | COVID-19 Genomics UK (COG-UK) Consortium | Luke W Meredith et al |
| 3334 | England/CAMB-7B4E6/2020 | 2020-04-12 | Department of Pathology, University of Cambridge | COVID-19 Genomics UK (COG-UK) Consortium | Luke W Meredith et al |
| 3335 | England/CAMB-7B4AA/2020 | 2020-04-12 | Department of Pathology, University of Cambridge | COVID-19 Genomics UK (COG-UK) Consortium | Luke W Meredith et al |
| 3336 | England/CAMB-7B4F5/2020 | 2020-04-12 | Department of Pathology, University of Cambridge | COVID-19 Genomics UK (COG-UK) Consortium | Luke W Meredith et al |

| | | | | | |
|---|---|---|---|---|---|
| 3337 | NewZealand/20VR2088/2020-04-12 | 2020-04-12 | Canterbury Health Laboratories | Institute of Environmental Science and Research (ESR) Ltd | **Matt Storey et al** |
| 3338 | NewZealand/20VR2085/2020-04-12 | 2020-04-12 | Canterbury Health Laboratories | Institute of Environmental Science and Research (ESR) Ltd | **Matt Storey et al** |
| 3339 | England/CAMB-7B3DA/2020 | 2020-04-12 | Department of Pathology, University of Cambridge | COVID-19 Genomics UK (COG-UK) Consortium | Luke W Meredith et al |
| 3340 | Scotland/EDB1536/2020 | 2020-04-12 | Virology Department, Royal Infirmary of Edinburgh, NHS Lothian / School of Biological Sciences, University of Edinburgh / Institute of Genetics and Molecular Medicine, University of Edinburgh | COVID-19 Genomics UK (COG-UK) Consortium | McHugh M et al |
| 3341 | Scotland/EDB1433/2020 | 2020-04-12 | Virology Department, Royal Infirmary of Edinburgh, NHS Lothian / School of Biological Sciences, University of Edinburgh / Institute of Genetics and Molecular Medicine, University of Edinburgh | COVID-19 Genomics UK (COG-UK) Consortium | McHugh M et al |
| 3342 | Scotland/EDB1448/2020 | 2020-04-12 | Virology Department, Royal Infirmary of Edinburgh, NHS Lothian / School of Biological Sciences, University of Edinburgh / Institute of Genetics and Molecular Medicine, University of Edinburgh | COVID-19 Genomics UK (COG-UK) Consortium | McHugh M et al |
| 3343 | Scotland/EDB1559/2020 | 2020-04-12 | Virology Department, Royal Infirmary of Edinburgh, NHS Lothian / School of Biological Sciences, University of Edinburgh / Institute of Genetics and Molecular Medicine, University of Edinburgh | COVID-19 Genomics UK (COG-UK) Consortium | McHugh M et al |
| 3344 | Luxembourg/LNS0188413/2020 | 2020-04-12 | Laboratoire National de Sante, Microbiology, Virology | Laboratoire National de Sante, Microbiology, Epidemiology and Microbial Genomics | Anke Wienecke-Baldacchino et al |
| 3345 | England/CAMB-7B48C/2020 | 2020-04-12 | Department of Pathology, University of Cambridge | COVID-19 Genomics UK (COG-UK) Consortium | Luke W Meredith et al |
| 3346 | India/NCDC-4155/2020 | 2020-04-12 | National Centre for Disease control (NCDC) | NCDC/CSIR-IGIB | Pramod Kumar et al |
| 3347 | Scotland/EDB1526/2020 | 2020-04-12 | Virology Department, Royal Infirmary of Edinburgh, NHS Lothian / School of Biological Sciences, University of Edinburgh / Institute of Genetics and Molecular Medicine, University of Edinburgh | COVID-19 Genomics UK (COG-UK) Consortium | McHugh M et al |
| 3348 | Taiwan/NTU29/2020 | 2020-04-12 | Department of Laboratory Medicine, National Taiwan University Hospital | Microbial Genomics Core Lab, National Taiwan University Centers of Genomic and Precision Medicine | Shiou-Hwei Yeh et al |
| 3349 | England/CAMB-7B431/2020 | 2020-04-12 | Department of Pathology, University of Cambridge | COVID-19 Genomics UK (COG-UK) Consortium | Luke W Meredith et al |
| 3350 | Scotland/EDB1544/2020 | 2020-04-12 | Virology Department, Royal Infirmary of Edinburgh, NHS Lothian / School of Biological Sciences, University of Edinburgh / Institute of Genetics and Molecular Medicine, University of Edinburgh | COVID-19 Genomics UK (COG-UK) Consortium | McHugh M et al |
| 3351 | England/CAMB-7C175/2020 | 2020-04-12 | Department of Pathology, University of Cambridge | COVID-19 Genomics UK (COG-UK) Consortium | Luke W Meredith et al |
| 3352 | England/CAMB-7B2BF/2020 | 2020-04-12 | Department of Pathology, University of Cambridge | COVID-19 Genomics UK (COG-UK) Consortium | Luke W Meredith et al |
| 3353 | Russia/StPetersburg-RII5942S/2020 | 2020-04-13 | WHO National Influenza Centre Russian Federation | WHO National Influenza Centre Russian Federation | Andrey Komissarov et al |

| | | | | | |
|---|---|---|---|---|---|
| 3354 | England/CAMB-7B52F/2020 | 2020-04-13 | Department of Pathology, University of Cambridge | COVID-19 Genomics UK (COG-UK) Consortium | Luke W Meredith et al |
| 3355 | USA/NY-NYUMC392/2020 | 2020-04-13 | NYU Langone Health | Departments of Pathology and Medicine, New York University School of Medicine | Maria Aguero-Rosenfeld et al |
| 3356 | Australia/VIC1334/2020 | 2020-04-13 | Microbiological Diagnostic Unit Public Health Laboratory | Microbiological Diagnostic Unit Public Health Laboratory | Seemann T. et al |
| 3357 | Australia/VIC1336/2020 | 2020-04-13 | Victorian Infectious Diseases Reference Laboratory (VIDRL) | Microbiological Diagnostic Unit Public Health Laboratory and Victorian Infectious Diseases Reference Laboratory, The Peter Doherty Institute for Infection and Immunity | Caly L. et al |
| 3358 | USA/NY-NYUMC404/2020 | 2020-04-13 | NYU Langone Health | Departments of Pathology and Medicine, New York University School of Medicine | Maria Aguero-Rosenfeld et al |
| 3359 | USA/NY-NYUMC395/2020 | 2020-04-13 | NYU Langone Health | Departments of Pathology and Medicine, New York University School of Medicine | Maria Aguero-Rosenfeld et al |
| 3360 | Scotland/EDB1453/2020 | 2020-04-13 | Virology Department, Royal Infirmary of Edinburgh, NHS Lothian / School of Biological Sciences, University of Edinburgh / Institute of Genetics and Molecular Medicine, University of Edinburgh | COVID-19 Genomics UK (COG-UK) Consortium | McHugh M et al |
| 3361 | Scotland/EDB1529/2020 | 2020-04-13 | Virology Department, Royal Infirmary of Edinburgh, NHS Lothian / School of Biological Sciences, University of Edinburgh / Institute of Genetics and Molecular Medicine, University of Edinburgh | COVID-19 Genomics UK (COG-UK) Consortium | McHugh M et al |
| 3362 | USA/WI-UW-299/2020 | 2020-04-13 | University of Wisconsin-Madison AIDS Vaccine Research Laboratories | University of Wisconsin-Madison AIDS Vaccine Research Laboratories | Gage Moreno et al |
| 3363 | USA/WI-143/2020 | 2020-04-13 | University of Wisconsin-Madison AIDS Vaccine Research Laboratories | University of Wisconsin-Madison AIDS Vaccine Research Laboratories | Moreno et al |
| 3364 | USA/NY-NYUMC393/2020 | 2020-04-13 | NYU Langone Health | Departments of Pathology and Medicine, New York University School of Medicine | Maria Aguero-Rosenfeld et al |
| 3365 | USA/WI-130/2020 | 2020-04-13 | University of Wisconsin-Madison AIDS Vaccine Research Laboratories | University of Wisconsin-Madison AIDS Vaccine Research Laboratories | Moreno et al |
| 3366 | USA/WI-UW-301/2020 | 2020-04-13 | University of Wisconsin-Madison AIDS Vaccine Research Laboratories | University of Wisconsin-Madison AIDS Vaccine Research Laboratories | Gage Moreno et al |
| 3367 | USA/WI-UW-298/2020 | 2020-04-13 | University of Wisconsin-Madison AIDS Vaccine Research Laboratories | University of Wisconsin-Madison AIDS Vaccine Research Laboratories | Gage Moreno et al |
| 3368 | USA/NY-NYUMC402/2020 | 2020-04-13 | NYU Langone Health | Departments of Pathology and Medicine, New York University School of Medicine | Maria Aguero-Rosenfeld et al |
| 3369 | England/CAMB-7B440/2020 | 2020-04-13 | Department of Pathology, University of Cambridge | COVID-19 Genomics UK (COG-UK) Consortium | Luke W Meredith et al |
| 3370 | USA/WI-UW-302/2020 | 2020-04-13 | University of Wisconsin-Madison AIDS Vaccine Research Laboratories | University of Wisconsin-Madison AIDS Vaccine Research Laboratories | Gage Moreno et al |
| 3371 | USA/LA-EVTL05/2020 | 2020-04-13 | LSUHS Emerging Viral Threat Laboratory | Microbial Genome Sequencing Center | Jeremy P. Kamil et al |
| 3372 | USA/NY-NYUMC399/2020 | 2020-04-13 | NYU Langone Health | Departments of Pathology and Medicine, New York University School of Medicine | Maria Aguero-Rosenfeld et al |
| 3373 | USA/NY-NYUMC331/2020 | 2020-04-13 | NYU Langone Health | Departments of Pathology and Medicine, New York University School of Medicine | Maria Aguero-Rosenfeld et al |
| 3374 | Turkey/GLAB-CoV008/2020 | 2020-04-13 | Genomic Laboratory (GLAB) (Conjoint lab of Health | Genomic Laboratory (GLAB), Istanbul Technical University | Ilker Karacan et al |

| | | | | | |
|---|---|---|---|---|---|
| | | | Directorate of Istanbul and Istanbul Technical University) | | |
| 3375 | USA/WI-UW-304/2020 | 2020-04-13 | University of Wisconsin-Madison AIDS Vaccine Research Laboratories | University of Wisconsin-Madison AIDS Vaccine Research Laboratories | Gage Moreno et al |
| 3376 | USA/WI-134/2020 | 2020-04-13 | University of Wisconsin-Madison AIDS Vaccine Research Laboratories | University of Wisconsin-Madison AIDS Vaccine Research Laboratories | Moreno et al |
| 3377 | USA/NY-NYUMC437/2020 | 2020-04-13 | NYU Langone Health | Departments of Pathology and Medicine, New York University School of Medicine | Maria Aguero-Rosenfeld et al |
| 3378 | USA/LA-EVTL04/2020 | 2020-04-13 | LSUHS Emerging Viral Threat Laboratory | Microbial Genome Sequencing Center | Jeremy P. Kamil et al |
| 3379 | USA/WI-UW-303/2020 | 2020-04-13 | University of Wisconsin-Madison AIDS Vaccine Research Laboratories | University of Wisconsin-Madison AIDS Vaccine Research Laboratories | Gage Moreno et al |
| 3380 | USA/CA-CZB-1028/2020 | 2020-04-13 | County of Santa Clara Public Health Department | Chan-Zuckerberg Biohub | CZB Cliahub Consortium et al |
| 3381 | USA/NY-NYUMC391/2020 | 2020-04-13 | NYU Langone Health | Departments of Pathology and Medicine, New York University School of Medicine | Maria Aguero-Rosenfeld et al |
| 3382 | USA/NY-NYUMC403/2020 | 2020-04-13 | NYU Langone Health | Departments of Pathology and Medicine, New York University School of Medicine | Maria Aguero-Rosenfeld et al |
| 3383 | Luxembourg/LNS5807524/2020 | 2020-04-13 | Laboratoire National de Sante, Microbiology, Virology | Laboratoire National de Sante, Microbiology, Epidemiology and Microbial Genomics | Anke Wienecke-Baldacchino et al |
| 3384 | NewZealand/20VR2089/2020-04-13 | 2020-04-13 | Canterbury Health Laboratories | Institute of Environmental Science and Research (ESR) Ltd | **Matt Storey et al** |
| 3385 | USA/WI-UW-297/2020 | 2020-04-13 | University of Wisconsin-Madison AIDS Vaccine Research Laboratories | University of Wisconsin-Madison AIDS Vaccine Research Laboratories | Gage Moreno et al |
| 3386 | USA/NY-NYUMC390/2020 | 2020-04-13 | NYU Langone Health | Departments of Pathology and Medicine, New York University School of Medicine | Maria Aguero-Rosenfeld et al |
| 3387 | Scotland/EDB1507/2020 | 2020-04-13 | Virology Department, Royal Infirmary of Edinburgh, NHS Lothian / School of Biological Sciences, University of Edinburgh / Institute of Genetics and Molecular Medicine, University of Edinburgh | COVID-19 Genomics UK (COG-UK) Consortium | McHugh M et al |
| 3388 | USA/NY-NYUMC394/2020 | 2020-04-13 | NYU Langone Health | Departments of Pathology and Medicine, New York University School of Medicine | Maria Aguero-Rosenfeld et al |
| 3389 | USA/NY-NYUMC397/2020 | 2020-04-13 | NYU Langone Health | Departments of Pathology and Medicine, New York University School of Medicine | Maria Aguero-Rosenfeld et al |
| 3390 | Scotland/EDB1580/2020 | 2020-04-13 | Virology Department, Royal Infirmary of Edinburgh, NHS Lothian / School of Biological Sciences, University of Edinburgh / Institute of Genetics and Molecular Medicine, University of Edinburgh | COVID-19 Genomics UK (COG-UK) Consortium | McHugh M et al |
| 3391 | USA/WI-UW-300/2020 | 2020-04-13 | University of Wisconsin-Madison AIDS Vaccine Research Laboratories | University of Wisconsin-Madison AIDS Vaccine Research Laboratories | Gage Moreno et al |
| 3392 | USA/NJ-NYUMC401/2020 | 2020-04-13 | NYU Langone Health | Departments of Pathology and Medicine, New York University School of Medicine | Maria Aguero-Rosenfeld et al |
| 3393 | Scotland/EDB1452/2020 | 2020-04-13 | Virology Department, Royal Infirmary of Edinburgh, NHS Lothian / School of Biological Sciences, University of Edinburgh / Institute of Genetics and Molecular Medicine, University of Edinburgh | COVID-19 Genomics UK (COG-UK) Consortium | McHugh M et al |

| | | | | | |
|---|---|---|---|---|---|
| 3394 | Luxembourg/LNS8742871/2020 | 2020-04-13 | Laboratoire National de Sante, Microbiology, Virology | Laboratoire National de Sante, Microbiology, Epidemiology and Microbial Genomics | Anke Wienecke-Baldacchino et al |
| 3395 | England/CAMB-7C39D/2020 | 2020-04-13 | Department of Pathology, University of Cambridge | COVID-19 Genomics UK (COG-UK) Consortium | Luke W Meredith et al |
| 3396 | USA/NY-NYUMC400/2020 | 2020-04-13 | NYU Langone Health | Departments of Pathology and Medicine, New York University School of Medicine | Maria Aguero-Rosenfeld et al |
| 3397 | India/NCDC-4475/2020 | 2020-04-13 | National Centre for Disease control (NCDC) | NCDC/CSIR-IGIB | Pramod Kumar et al |
| 3398 | England/CAMB-7B510/2020 | 2020-04-13 | Department of Pathology, University of Cambridge | COVID-19 Genomics UK (COG-UK) Consortium | Luke W Meredith et al |
| 3399 | USA/WI-120/2020 | 2020-04-13 | University of Wisconsin-Madison AIDS Vaccine Research Laboratories | University of Wisconsin-Madison AIDS Vaccine Research Laboratories | Moreno et al |
| 3400 | USA/NY-NYUMC398/2020 | 2020-04-13 | NYU Langone Health | Departments of Pathology and Medicine, New York University School of Medicine | Maria Aguero-Rosenfeld et al |
| 3401 | England/CAMB-7B3F8/2020 | 2020-04-13 | Department of Pathology, University of Cambridge | COVID-19 Genomics UK (COG-UK) Consortium | Luke W Meredith et al |
| 3402 | USA/NY-NYUMC406/2020 | 2020-04-13 | NYU Langone Health | Departments of Pathology and Medicine, New York University School of Medicine | Maria Aguero-Rosenfeld et al |
| 3403 | Taiwan/NTU27/2020 | 2020-04-13 | Department of Laboratory Medicine, National Taiwan University Hospital | Microbial Genomics Core Lab, National Taiwan University Centers of Genomic and Precision Medicine | Shiou-Hwei Yeh et al |
| 3404 | India/GBRC3/2020 | 2020-04-13 | B.J. Medical College and Civil hospital | Gujarat Biotechnology Research Centre | Ankit Hinsu et al |
| 3405 | Luxembourg/LNS0972189/2020 | 2020-04-13 | Laboratoire National de Sante, Microbiology, Virology | Laboratoire National de Sante, Microbiology, Epidemiology and Microbial Genomics | Anke Wienecke-Baldacchino et al |
| 3406 | England/CAMB-7B56B/2020 | 2020-04-13 | Department of Pathology, University of Cambridge | COVID-19 Genomics UK (COG-UK) Consortium | Luke W Meredith et al |
| 3407 | India/NCDC-4189/2020 | 2020-04-13 | National Centre for Disease control (NCDC) | NCDC/CSIR-IGIB | Pramod Kumar et al |
| 3408 | USA/NY-NYUMC389/2020 | 2020-04-13 | NYU Langone Health | Departments of Pathology and Medicine, New York University School of Medicine | Maria Aguero-Rosenfeld et al |
| 3409 | England/CAMB-7B422/2020 | 2020-04-13 | Department of Pathology, University of Cambridge | COVID-19 Genomics UK (COG-UK) Consortium | Luke W Meredith et al |
| 3410 | England/CAMB-7C148/2020 | 2020-04-13 | Department of Pathology, University of Cambridge | COVID-19 Genomics UK (COG-UK) Consortium | Luke W Meredith et al |
| 3411 | Scotland/EDB1450/2020 | 2020-04-13 | Virology Department, Royal Infirmary of Edinburgh, NHS Lothian / School of Biological Sciences, University of Edinburgh / Institute of Genetics and Molecular Medicine, University of Edinburgh | COVID-19 Genomics UK (COG-UK) Consortium | McHugh M et al |
| 3412 | Scotland/EDB1519/2020 | 2020-04-13 | Virology Department, Royal Infirmary of Edinburgh, NHS Lothian / School of Biological Sciences, University of Edinburgh / Institute of Genetics and Molecular Medicine, University of Edinburgh | COVID-19 Genomics UK (COG-UK) Consortium | McHugh M et al |
| 3413 | USA/VA-DCLS-0105/2020 | 2020-04-14 | Virginia DCLS | Virginia DCLS | Virginia DCLS et al |
| 3414 | Singapore/82/2020 | 2020-04-14 | National Public Health Laboratory, National Centre for Infectious Diseases | National Public Health Laboratory, National Centre for Infectious Diseases | Mak Tze Minn et al |
| 3415 | Australia/VIC1344/2020 | 2020-04-14 | Victorian Infectious Diseases Reference Laboratory (VIDRL) | Microbiological Diagnostic Unit Public Health Laboratory and Victorian Infectious Diseases Reference Laboratory, The Peter Doherty Institute for Infection and Immunity | Caly L. et al |
| 3416 | Scotland/EDB1629/2020 | 2020-04-14 | Virology Department, Royal Infirmary of Edinburgh, NHS | COVID-19 Genomics UK (COG-UK) Consortium | McHugh M et al |

| | | | | | |
|---|---|---|---|---|---|
| | | | Lothian / School of Biological Sciences, University of Edinburgh / Institute of Genetics and Molecular Medicine, University of Edinburgh | | |
| 3417 | USA/NY-NYUMC443/2020 | 2020-04-14 | NYU Langone Health | Departments of Pathology and Medicine, New York University School of Medicine | Maria Aguero-Rosenfeld et al |
| 3418 | India/GBRC7/2020 | 2020-04-14 | B.J. Medical College and Civil hospital | Gujarat Biotechnology Research Centre | Monika Gandhi et al |
| 3419 | India/NCDC-4208/2020 | 2020-04-14 | National Centre for Disease control (NCDC) | NCDC/CSIR-IGIB | Pramod Kumar et al |
| 3420 | USA/WI-GMF-01047/2020 | 2020-04-14 | Gundersen Molecular Diagnostics Laboratory | Kabara Cancer Research Institute | Craig S. Richmond et al |
| 3421 | USA/WI-UW-305/2020 | 2020-04-14 | University of Wisconsin-Madison AIDS Vaccine Research Laboratories | University of Wisconsin-Madison AIDS Vaccine Research Laboratories | Gage Moreno et al |
| 3422 | USA/WI-UW-306/2020 | 2020-04-14 | University of Wisconsin-Madison AIDS Vaccine Research Laboratories | University of Wisconsin-Madison AIDS Vaccine Research Laboratories | Gage Moreno et al |
| 3423 | Russia/StPetersburg-RII5635S/2020 | 2020-04-14 | WHO National Influenza Centre Russian Federation | WHO National Influenza Centre Russian Federation | Andrey Komissarov et al |
| 3424 | England/CAMB-7C333/2020 | 2020-04-14 | Department of Pathology, University of Cambridge | COVID-19 Genomics UK (COG-UK) Consortium | Luke W Meredith et al |
| 3425 | Luxembourg/LNS2458393/2020 | 2020-04-14 | Laboratoire National de Sante, Microbiology, Virology | Laboratoire National de Sante, Microbiology, Epidemiology and Microbial Genomics | Anke Wienecke-Baldacchino et al |
| 3426 | USA/NY-NYUMC428/2020 | 2020-04-14 | NYU Langone Health | Departments of Pathology and Medicine, New York University School of Medicine | Maria Aguero-Rosenfeld et al |
| 3427 | Scotland/EDB1525/2020 | 2020-04-14 | Virology Department, Royal Infirmary of Edinburgh, NHS Lothian / School of Biological Sciences, University of Edinburgh / Institute of Genetics and Molecular Medicine, University of Edinburgh | COVID-19 Genomics UK (COG-UK) Consortium | McHugh M et al |
| 3428 | CzechRepublic/2308/2020 | 2020-04-14 | The National Institute of Public Health Center for Epidemiology and Microbiology | The National Institute of Public Health Center for Epidemiology and Microbiology | Alexander Nagy et al |
| 3429 | England/CAMB-7C3AC/2020 | 2020-04-14 | Department of Pathology, University of Cambridge | COVID-19 Genomics UK (COG-UK) Consortium | Luke W Meredith et al |
| 3430 | Scotland/EDB1642/2020 | 2020-04-14 | Virology Department, Royal Infirmary of Edinburgh, NHS Lothian / School of Biological Sciences, University of Edinburgh / Institute of Genetics and Molecular Medicine, University of Edinburgh | COVID-19 Genomics UK (COG-UK) Consortium | McHugh M et al |
| 3431 | Scotland/EDB1656/2020 | 2020-04-14 | Virology Department, Royal Infirmary of Edinburgh, NHS Lothian / School of Biological Sciences, University of Edinburgh / Institute of Genetics and Molecular Medicine, University of Edinburgh | COVID-19 Genomics UK (COG-UK) Consortium | McHugh M et al |
| 3432 | Russia/StPetersburg-RII5634S/2020 | 2020-04-14 | WHO National Influenza Centre Russian Federation | WHO National Influenza Centre Russian Federation | Andrey Komissarov et al |
| 3433 | USA/NY-NYUMC426/2020 | 2020-04-14 | NYU Langone Health | Departments of Pathology and Medicine, New York University School of Medicine | Maria Aguero-Rosenfeld et al |
| 3434 | USA/WI-UW-311/2020 | 2020-04-14 | University of Wisconsin-Madison AIDS Vaccine Research Laboratories | University of Wisconsin-Madison AIDS Vaccine Research Laboratories | Gage Moreno et al |
| 3435 | USA/VA-DCLS-0107/2020 | 2020-04-14 | Virginia DCLS | Virginia DCLS | Virginia DCLS et al |
| 3436 | USA/NY-NYUMC446/2020 | 2020-04-14 | NYU Langone Health | Departments of Pathology and | Maria |

| | | | | | |
|---|---|---|---|---|---|
| | | | | Medicine, New York University School of Medicine | Aguero-Rosenfeld et al |
| 3437 | USA/VA-DCLS-0106/2020 | 2020-04-14 | Virginia DCLS | Virginia DCLS | Virginia DCLS et al |
| 3438 | Russia/StPetersburg-RII5633S/2020 | 2020-04-14 | WHO National Influenza Centre Russian Federation | WHO National Influenza Centre Russian Federation | Andrey Komissarov et al |
| 3439 | Sweden/20-07298/2020 | 2020-04-14 | Ulltuna Vardcentral | The Public Health Agency of Sweden | Heidi Lindback et al |
| 3440 | USA/VA-DCLS-0108/2020 | 2020-04-14 | Virginia DCLS | Virginia DCLS | Virginia DCLS et al |
| 3441 | England/CAMB-7C2AF/2020 | 2020-04-14 | Department of Pathology, University of Cambridge | COVID-19 Genomics UK (COG-UK) Consortium | Luke W Meredith et al |
| 3442 | England/CAMB-7C2BE/2020 | 2020-04-14 | Department of Pathology, University of Cambridge | COVID-19 Genomics UK (COG-UK) Consortium | Luke W Meredith et al |
| 3443 | Russia/StPetersburg-RII5654S/2020 | 2020-04-14 | WHO National Influenza Centre Russian Federation | WHO National Influenza Centre Russian Federation | Andrey Komissarov et al |
| 3444 | Russia/StPetersburg-RII6039S/2020 | 2020-04-14 | WHO National Influenza Centre Russian Federation | WHO National Influenza Centre Russian Federation | Andrey Komissarov et al |
| 3445 | Scotland/EDB1523/2020 | 2020-04-14 | Virology Department, Royal Infirmary of Edinburgh, NHS Lothian / School of Biological Sciences, University of Edinburgh / Institute of Genetics and Molecular Medicine, University of Edinburgh | COVID-19 Genomics UK (COG-UK) Consortium | McHugh M et al |
| 3446 | Scotland/EDB1632/2020 | 2020-04-14 | Virology Department, Royal Infirmary of Edinburgh, NHS Lothian / School of Biological Sciences, University of Edinburgh / Institute of Genetics and Molecular Medicine, University of Edinburgh | COVID-19 Genomics UK (COG-UK) Consortium | McHugh M et al |
| 3447 | Russia/StPetersburg-RII6027S/2020 | 2020-04-14 | WHO National Influenza Centre Russian Federation | WHO National Influenza Centre Russian Federation | Andrey Komissarov et al |
| 3448 | Russia/StPetersburg-RII5644S/2020 | 2020-04-14 | WHO National Influenza Centre Russian Federation | WHO National Influenza Centre Russian Federation | Andrey Komissarov et al |
| 3449 | England/CAMB-7C227/2020 | 2020-04-14 | Department of Pathology, University of Cambridge | COVID-19 Genomics UK (COG-UK) Consortium | Luke W Meredith et al |
| 3450 | Scotland/EDB1581/2020 | 2020-04-14 | Virology Department, Royal Infirmary of Edinburgh, NHS Lothian / School of Biological Sciences, University of Edinburgh / Institute of Genetics and Molecular Medicine, University of Edinburgh | COVID-19 Genomics UK (COG-UK) Consortium | McHugh M et al |
| 3451 | Scotland/EDB1647/2020 | 2020-04-14 | Virology Department, Royal Infirmary of Edinburgh, NHS Lothian / School of Biological Sciences, University of Edinburgh / Institute of Genetics and Molecular Medicine, University of Edinburgh | COVID-19 Genomics UK (COG-UK) Consortium | McHugh M et al |
| 3452 | Russia/StPetersburg-RII6031S/2020 | 2020-04-14 | WHO National Influenza Centre Russian Federation | WHO National Influenza Centre Russian Federation | Andrey Komissarov et al |
| 3453 | Russia/StPetersburg-RII5643S/2020 | 2020-04-14 | WHO National Influenza Centre Russian Federation | WHO National Influenza Centre Russian Federation | Andrey Komissarov et al |
| 3454 | England/CAMB-7C360/2020 | 2020-04-14 | Department of Pathology, University of Cambridge | COVID-19 Genomics UK (COG-UK) Consortium | Luke W Meredith et al |
| 3455 | Scotland/EDB1711/2020 | 2020-04-15 | Virology Department, Royal Infirmary of Edinburgh, NHS Lothian / School of Biological Sciences, University of Edinburgh / Institute of Genetics and Molecular Medicine, University of Edinburgh | COVID-19 Genomics UK (COG-UK) Consortium | McHugh M et al |
| 3456 | England/CAMB-7C2DC/2020 | 2020-04-15 | Department of Pathology, University of Cambridge | COVID-19 Genomics UK (COG-UK) Consortium | Luke W Meredith et al |
| 3457 | USA/LA-EVTL08/2020 | 2020-04-15 | LSUHS Emerging Viral Threat | Microbial Genome Sequencing | Jeremy P. Kamil et al |

| | | | | | |
|---|---|---|---|---|---|
| | | | Laboratory | Center | |
| 3458 | USA/WI-UW-310/2020 | 2020-04-15 | University of Wisconsin-Madison AIDS Vaccine Research Laboratories | University of Wisconsin-Madison AIDS Vaccine Research Laboratories | Gage Moreno et al |
| 3459 | USA/NY-NYUMC438/2020 | 2020-04-15 | NYU Langone Health | Departments of Pathology and Medicine, New York University School of Medicine | Maria Aguero-Rosenfeld et al |
| 3460 | USA/NY-NYUMC442/2020 | 2020-04-15 | NYU Langone Health | Departments of Pathology and Medicine, New York University School of Medicine | Maria Aguero-Rosenfeld et al |
| 3461 | USA/LA-EVTL09/2020 | 2020-04-15 | LSUHS Emerging Viral Threat Laboratory | Microbial Genome Sequencing Center | Jeremy P. Kamil et al |
| 3462 | USA/LA-EVTL07/2020 | 2020-04-15 | LSUHS Emerging Viral Threat Laboratory | Microbial Genome Sequencing Center | Jeremy P. Kamil et al |
| 3463 | Russia/StPetersburg-RII6086S/2020 | 2020-04-15 | WHO National Influenza Centre Russian Federation | WHO National Influenza Centre Russian Federation | Andrey Komissarov et al |
| 3464 | USA/NY-NYUMC464/2020 | 2020-04-15 | NYU Langone Health | Departments of Pathology and Medicine, New York University School of Medicine | Maria Aguero-Rosenfeld et al |
| 3465 | USA/WI-UW-307/2020 | 2020-04-15 | University of Wisconsin-Madison AIDS Vaccine Research Laboratories | University of Wisconsin-Madison AIDS Vaccine Research Laboratories | Gage Moreno et al |
| 3466 | Sweden/20-07390/2020 | 2020-04-15 | Lakargruppen | The Public Health Agency of Sweden | Boris Klanger et al |
| 3467 | USA/LA-EVTL011/2020 | 2020-04-15 | LSUHS Emerging Viral Threat Laboratory | Microbial Genome Sequencing Center | Jeremy P. Kamil et al |
| 3468 | Scotland/EDB1694/2020 | 2020-04-15 | Virology Department, Royal Infirmary of Edinburgh, NHS Lothian / School of Biological Sciences, University of Edinburgh / Institute of Genetics and Molecular Medicine, University of Edinburgh | COVID-19 Genomics UK (COG-UK) Consortium | McHugh M et al |
| 3469 | NewZealand/20VR2541/2020-04-25 | 2020-04-15 | LabPLUS | Institute of Environmental Science and Research (ESR) | **Matt Storey et al** |
| 3470 | Luxembourg/LNS5953507/2020 | 2020-04-15 | Laboratoire National de Sante, Microbiology, Virology | Laboratoire National de Sante, Microbiology, Epidemiology and Microbial Genomics | Anke Wienecke-Baldacchino et al |
| 3471 | USA/NY-NYUMC445/2020 | 2020-04-15 | NYU Langone Health | Departments of Pathology and Medicine, New York University School of Medicine | Maria Aguero-Rosenfeld et al |
| 3472 | England/CAMB-7C290/2020 | 2020-04-15 | Department of Pathology, University of Cambridge | COVID-19 Genomics UK (COG-UK) Consortium | Luke W Meredith et al |
| 3473 | USA/LA-EVTL010/2020 | 2020-04-15 | LSUHS Emerging Viral Threat Laboratory | Microbial Genome Sequencing Center | Jeremy P. Kamil et al |
| 3474 | Scotland/EDB1663/2020 | 2020-04-15 | Virology Department, Royal Infirmary of Edinburgh, NHS Lothian / School of Biological Sciences, University of Edinburgh / Institute of Genetics and Molecular Medicine, University of Edinburgh | COVID-19 Genomics UK (COG-UK) Consortium | McHugh M et al |
| 3475 | USA/NY-NYUMC444/2020 | 2020-04-15 | NYU Langone Health | Departments of Pathology and Medicine, New York University School of Medicine | Maria Aguero-Rosenfeld et al |
| 3476 | USA/NY-NYUMC439/2020 | 2020-04-15 | NYU Langone Health | Departments of Pathology and Medicine, New York University School of Medicine | Maria Aguero-Rosenfeld et al |
| 3477 | Sweden/20-07389/2020 | 2020-04-15 | Kungsors VC | The Public Health Agency of Sweden | Jessica Karlsson et al |
| 3478 | England/CAMB-7E164/2020 | 2020-04-15 | Department of Pathology, University of Cambridge | COVID-19 Genomics UK (COG-UK) Consortium | Luke W Meredith et al |
| 3479 | Scotland/EDB1681/2020 | 2020-04-15 | Virology Department, Royal Infirmary of Edinburgh, NHS Lothian / School of Biological Sciences, University of Edinburgh / Institute of Genetics and Molecular | COVID-19 Genomics UK (COG-UK) Consortium | McHugh M et al |

| | | | | | |
|---|---|---|---|---|---|
| | | | Medicine, University of Edinburgh | | |
| 3480 | NewZealand/20VR2109/2020-04-15 | 2020-04-15 | MedLab Central Ltd | Institute of Environmental Science and Research (ESR) Ltd | **Matt Storey et al** |
| 3481 | NewZealand/20VR2108/2020-04-15 | 2020-04-15 | MedLab Central Ltd | Institute of Environmental Science and Research (ESR) Ltd | **Matt Storey et al** |
| 3482 | NewZealand/20VR2562/2020-04-20 | 2020-04-15 | Middlemore Hospital | Institute of Environmental Science and Research (ESR) | **Matt Storey et al** |
| 3483 | USA/LA-EVTL06/2020 | 2020-04-15 | LSUHS Emerging Viral Threat Laboratory | Microbial Genome Sequencing Center | Jeremy P. Kamil et al |
| 3484 | USA/NY-NYUMC429/2020 | 2020-04-15 | NYU Langone Health | Departments of Pathology and Medicine, New York University School of Medicine | Maria Aguero-Rosenfeld et al |
| 3485 | Scotland/EDB1730/2020 | 2020-04-15 | Virology Department, Royal Infirmary of Edinburgh, NHS Lothian / School of Biological Sciences, University of Edinburgh / Institute of Genetics and Molecular Medicine, University of Edinburgh | COVID-19 Genomics UK (COG-UK) Consortium | McHugh M et al |
| 3486 | Russia/StPetersburg-RII6061S/2020 | 2020-04-15 | WHO National Influenza Centre Russian Federation | WHO National Influenza Centre Russian Federation | Andrey Komissarov et al |
| 3487 | Scotland/EDB1697/2020 | 2020-04-15 | Virology Department, Royal Infirmary of Edinburgh, NHS Lothian / School of Biological Sciences, University of Edinburgh / Institute of Genetics and Molecular Medicine, University of Edinburgh | COVID-19 Genomics UK (COG-UK) Consortium | McHugh M et al |
| 3488 | Scotland/EDB1638/2020 | 2020-04-15 | Virology Department, Royal Infirmary of Edinburgh, NHS Lothian / School of Biological Sciences, University of Edinburgh / Institute of Genetics and Molecular Medicine, University of Edinburgh | COVID-19 Genomics UK (COG-UK) Consortium | McHugh M et al |
| 3489 | USA/NY-NYUMC434/2020 | 2020-04-15 | NYU Langone Health | Departments of Pathology and Medicine, New York University School of Medicine | Maria Aguero-Rosenfeld et al |
| 3490 | USA/NY-NYUMC440/2020 | 2020-04-15 | NYU Langone Health | Departments of Pathology and Medicine, New York University School of Medicine | Maria Aguero-Rosenfeld et al |
| 3491 | USA/NY-NYUMC447/2020 | 2020-04-15 | NYU Langone Health | Departments of Pathology and Medicine, New York University School of Medicine | Maria Aguero-Rosenfeld et al |
| 3492 | Scotland/EDB1717/2020 | 2020-04-15 | Virology Department, Royal Infirmary of Edinburgh, NHS Lothian / School of Biological Sciences, University of Edinburgh / Institute of Genetics and Molecular Medicine, University of Edinburgh | COVID-19 Genomics UK (COG-UK) Consortium | McHugh M et al |
| 3493 | Singapore/70/2020 | 2020-04-15 | National Public Health Laboratory, National Centre for Infectious Diseases | National Public Health Laboratory, National Centre for Infectious Diseases | Mak TM et al |
| 3494 | USA/NY-NYUMC441/2020 | 2020-04-15 | NYU Langone Health | Departments of Pathology and Medicine, New York University School of Medicine | Maria Aguero-Rosenfeld et al |
| 3495 | Russia/StPetersburg-RII6053S/2020 | 2020-04-15 | WHO National Influenza Centre Russian Federation | WHO National Influenza Centre Russian Federation | Andrey Komissarov et al |
| 3496 | Scotland/EDB1671/2020 | 2020-04-15 | Virology Department, Royal Infirmary of Edinburgh, NHS Lothian / School of Biological Sciences, University of Edinburgh / Institute of Genetics and Molecular Medicine, University of Edinburgh | COVID-19 Genomics UK (COG-UK) Consortium | McHugh M et al |

| | | | | | |
|---|---|---|---|---|---|
| 3497 | Scotland/EDB1677/2020 | 2020-04-15 | Virology Department, Royal Infirmary of Edinburgh, NHS Lothian / School of Biological Sciences, University of Edinburgh / Institute of Genetics and Molecular Medicine, University of Edinburgh | COVID-19 Genomics UK (COG-UK) Consortium | McHugh M et al |
| 3498 | Singapore/71/2020 | 2020-04-15 | National Public Health Laboratory, National Centre for Infectious Diseases | National Public Health Laboratory, National Centre for Infectious Diseases | Mak TM et al |
| 3499 | Russia/StPetersburg-RII6065S/2020 | 2020-04-15 | WHO National Influenza Centre Russian Federation | WHO National Influenza Centre Russian Federation | Andrey Komissarov et al |
| 3500 | Singapore/69/2020 | 2020-04-15 | National Public Health Laboratory, National Centre for Infectious Diseases | National Public Health Laboratory, National Centre for Infectious Diseases | Mak TM et al |
| 3501 | Scotland/EDB1693/2020 | 2020-04-15 | Virology Department, Royal Infirmary of Edinburgh, NHS Lothian / School of Biological Sciences, University of Edinburgh / Institute of Genetics and Molecular Medicine, University of Edinburgh | COVID-19 Genomics UK (COG-UK) Consortium | McHugh M et al |
| 3502 | USA/NY-NYUMC463/2020 | 2020-04-16 | NYU Langone Health | Departments of Pathology and Medicine, New York University School of Medicine | Maria Aguero-Rosenfeld et al |
| 3503 | England/CAMB-7E119/2020 | 2020-04-16 | Department of Pathology, University of Cambridge | COVID-19 Genomics UK (COG-UK) Consortium | Luke W Meredith et al |
| 3504 | England/CAMB-7FBFC/2020 | 2020-04-16 | Department of Pathology, University of Cambridge | COVID-19 Genomics UK (COG-UK) Consortium | Luke W Meredith et al |
| 3505 | USA/WI-GMF-01158/2020 | 2020-04-16 | Gundersen Molecular Diagnostics Laboratory | Kabara Cancer Research Institute | Craig S. Richmond et al |
| 3506 | USA/WI-UW-313/2020 | 2020-04-16 | University of Wisconsin-Madison AIDS Vaccine Research Laboratories | University of Wisconsin-Madison AIDS Vaccine Research Laboratories | Gage Moreno et al |
| 3507 | USA/NY-NYUMC470/2020 | 2020-04-16 | NYU Langone Health | Departments of Pathology and Medicine, New York University School of Medicine | Maria Aguero-Rosenfeld et al |
| 3508 | USA/WI-UW-309/2020 | 2020-04-16 | University of Wisconsin-Madison AIDS Vaccine Research Laboratories | University of Wisconsin-Madison AIDS Vaccine Research Laboratories | Gage Moreno et al |
| 3509 | USA/NY-NYUMC467/2020 | 2020-04-16 | NYU Langone Health | Departments of Pathology and Medicine, New York University School of Medicine | Maria Aguero-Rosenfeld et al |
| 3510 | USA/NY-NYUMC465/2020 | 2020-04-16 | NYU Langone Health | Departments of Pathology and Medicine, New York University School of Medicine | Maria Aguero-Rosenfeld et al |
| 3511 | England/CAMB-7FC53/2020 | 2020-04-16 | Department of Pathology, University of Cambridge | COVID-19 Genomics UK (COG-UK) Consortium | Luke W Meredith et al |
| 3512 | USA/WI-UW-314/2020 | 2020-04-16 | University of Wisconsin-Madison AIDS Vaccine Research Laboratories | University of Wisconsin-Madison AIDS Vaccine Research Laboratories | Gage Moreno et al |
| 3513 | Scotland/EDB1821/2020 | 2020-04-16 | Virology Department, Royal Infirmary of Edinburgh, NHS Lothian / School of Biological Sciences, University of Edinburgh / Institute of Genetics and Molecular Medicine, University of Edinburgh | COVID-19 Genomics UK (COG-UK) Consortium | McHugh M et al |
| 3514 | England/CAMB-7FC35/2020 | 2020-04-16 | Department of Pathology, University of Cambridge | COVID-19 Genomics UK (COG-UK) Consortium | Luke W Meredith et al |
| 3515 | USA/NY-NYUMC427/2020 | 2020-04-16 | NYU Langone Health | Departments of Pathology and Medicine, New York University School of Medicine | Maria Aguero-Rosenfeld et al |
| 3516 | Sweden/20-07480/2020 | 2020-04-16 | Omtanken Grimmered | The Public Health Agency of Sweden | Bernd Sengpiel et al |
| 3517 | USA/WI-UW-308/2020 | 2020-04-16 | University of Wisconsin-Madison AIDS Vaccine Research Laboratories | University of Wisconsin-Madison AIDS Vaccine Research Laboratories | Gage Moreno et al |

| | | | | | |
|---|---|---|---|---|---|
| 3518 | USA/NY-NYUMC466/2020 | 2020-04-16 | NYU Langone Health | Departments of Pathology and Medicine, New York University School of Medicine | Maria Aguero-Rosenfeld et al |
| 3519 | USA/WI-GMF-01159/2020 | 2020-04-16 | Gundersen Molecular Diagnostics Laboratory | Kabara Cancer Research Institute | Craig S. Richmond et al |
| 3520 | USA/NY-NYUMC468/2020 | 2020-04-16 | NYU Langone Health | Departments of Pathology and Medicine, New York University School of Medicine | Maria Aguero-Rosenfeld et al |
| 3521 | England/CAMB-7E1BF/2020 | 2020-04-16 | Department of Pathology, University of Cambridge | COVID-19 Genomics UK (COG-UK) Consortium | Luke W Meredith et al |
| 3522 | NewZealand/20VR3004/2020-04-16 | 2020-04-16 | North Shore Hospital | Institute of Environmental Science and Research (ESR) Ltd | **Matt Storey et al** |
| 3523 | England/CAMB-7FC08/2020 | 2020-04-16 | Department of Pathology, University of Cambridge | COVID-19 Genomics UK (COG-UK) Consortium | Luke W Meredith et al |
| 3524 | England/CAMB-7FBB0/2020 | 2020-04-16 | Department of Pathology, University of Cambridge | COVID-19 Genomics UK (COG-UK) Consortium | Luke W Meredith et al |
| 3525 | Sweden/20-07440/2020 | 2020-04-16 | Narhalsan Sjobo vardcentral | The Public Health Agency of Sweden | Lovisa Hjerten et al |
| 3526 | India/NCDC-4867/2020 | 2020-04-16 | National Centre for Disease control (NCDC) | NCDC/CSIR-IGIB | Pramod Kumar et al |
| 3527 | USA/CA-CZB-1029/2020 | 2020-04-16 | County of Santa Clara Public Health Department | Chan-Zuckerberg Biohub | CZB Cliahub Consortium et al |
| 3528 | England/CAMB-7E1FB/2020 | 2020-04-16 | Department of Pathology, University of Cambridge | COVID-19 Genomics UK (COG-UK) Consortium | Luke W Meredith et al |
| 3529 | England/CAMB-7FC26/2020 | 2020-04-16 | Department of Pathology, University of Cambridge | COVID-19 Genomics UK (COG-UK) Consortium | Luke W Meredith et al |
| 3530 | Scotland/EDB1829/2020 | 2020-04-16 | Virology Department, Royal Infirmary of Edinburgh, NHS Lothian / School of Biological Sciences, University of Edinburgh / Institute of Genetics and Molecular Medicine, University of Edinburgh | COVID-19 Genomics UK (COG-UK) Consortium | McHugh M et al |
| 3531 | Scotland/EDB1812/2020 | 2020-04-16 | Virology Department, Royal Infirmary of Edinburgh, NHS Lothian / School of Biological Sciences, University of Edinburgh / Institute of Genetics and Molecular Medicine, University of Edinburgh | COVID-19 Genomics UK (COG-UK) Consortium | McHugh M et al |
| 3532 | Scotland/EDB1816/2020 | 2020-04-16 | Virology Department, Royal Infirmary of Edinburgh, NHS Lothian / School of Biological Sciences, University of Edinburgh / Institute of Genetics and Molecular Medicine, University of Edinburgh | COVID-19 Genomics UK (COG-UK) Consortium | McHugh M et al |
| 3533 | England/CAMB-7FB74/2020 | 2020-04-17 | Department of Pathology, University of Cambridge | COVID-19 Genomics UK (COG-UK) Consortium | Luke W Meredith et al |
| 3534 | Turkey/GLAB-CoV012/2020 | 2020-04-17 | Genomic Laboratory (GLAB) (Conjoint lab of Health Directorate of Istanbul and Istanbul Technical University) | Genomic Laboratory (GLAB), Istanbul Technical University | Ilker Karacan et al |
| 3535 | USA/WI-259/2020 | 2020-04-17 | University of Wisconsin-Madison AIDS Vaccine Research Laboratories | University of Wisconsin-Madison AIDS Vaccine Research Laboratories | Moreno et al |
| 3536 | USA/WI-UW-315/2020 | 2020-04-17 | University of Wisconsin-Madison AIDS Vaccine Research Laboratories | University of Wisconsin-Madison AIDS Vaccine Research Laboratories | Gage Moreno et al |
| 3537 | England/CAMB-7FB65/2020 | 2020-04-17 | Department of Pathology, University of Cambridge | COVID-19 Genomics UK (COG-UK) Consortium | Luke W Meredith et al |
| 3538 | England/CAMB-7FB92/2020 | 2020-04-17 | Department of Pathology, University of Cambridge | COVID-19 Genomics UK (COG-UK) Consortium | Luke W Meredith et al |
| 3539 | Scotland/EDB1848/2020 | 2020-04-17 | Virology Department, Royal Infirmary of Edinburgh, NHS Lothian / School of Biological Sciences, University of Edinburgh / Institute of | COVID-19 Genomics UK (COG-UK) Consortium | McHugh M et al |

| | | | | | |
|---|---|---|---|---|---|
| | | | Genetics and Molecular Medicine, University of Edinburgh | | |
| 3540 | England/CAMB-7E322/2020 | 2020-04-17 | Department of Pathology, University of Cambridge | COVID-19 Genomics UK (COG-UK) Consortium | Luke W Meredith et al |
| 3541 | England/CAMB-7E2E9/2020 | 2020-04-17 | Department of Pathology, University of Cambridge | COVID-19 Genomics UK (COG-UK) Consortium | Luke W Meredith et al |
| 3542 | England/CAMB-7E313/2020 | 2020-04-17 | Department of Pathology, University of Cambridge | COVID-19 Genomics UK (COG-UK) Consortium | Luke W Meredith et al |
| 3543 | England/CAMB-7FBA1/2020 | 2020-04-17 | Department of Pathology, University of Cambridge | COVID-19 Genomics UK (COG-UK) Consortium | Luke W Meredith et al |
| 3544 | NewZealand/20VR3045/2020-04-17 | 2020-04-17 | Wellington SCL (WN) | Institute of Environmental Science and Research (ESR) Ltd | **Matt Storey et al** |
| 3545 | Scotland/EDB1897/2020 | 2020-04-17 | Virology Department, Royal Infirmary of Edinburgh, NHS Lothian / School of Biological Sciences, University of Edinburgh / Institute of Genetics and Molecular Medicine, University of Edinburgh | COVID-19 Genomics UK (COG-UK) Consortium | McHugh M et al |
| 3546 | Scotland/EDB1842/2020 | 2020-04-17 | Virology Department, Royal Infirmary of Edinburgh, NHS Lothian / School of Biological Sciences, University of Edinburgh / Institute of Genetics and Molecular Medicine, University of Edinburgh | COVID-19 Genomics UK (COG-UK) Consortium | McHugh M et al |
| 3547 | England/CAMB-7FB83/2020 | 2020-04-17 | Department of Pathology, University of Cambridge | COVID-19 Genomics UK (COG-UK) Consortium | Luke W Meredith et al |
| 3548 | Scotland/EDB1841/2020 | 2020-04-17 | Virology Department, Royal Infirmary of Edinburgh, NHS Lothian / School of Biological Sciences, University of Edinburgh / Institute of Genetics and Molecular Medicine, University of Edinburgh | COVID-19 Genomics UK (COG-UK) Consortium | McHugh M et al |
| 3549 | England/CAMB-7FB47/2020 | 2020-04-17 | Department of Pathology, University of Cambridge | COVID-19 Genomics UK (COG-UK) Consortium | Luke W Meredith et al |
| 3550 | England/CAMB-7FB56/2020 | 2020-04-17 | Department of Pathology, University of Cambridge | COVID-19 Genomics UK (COG-UK) Consortium | Luke W Meredith et al |
| 3551 | Sweden/20-07439/2020 | 2020-04-17 | Narhalsan Molnlycke, Barn och ungdomsmedicin | The Public Health Agency of Sweden | Mats Reimer et al |
| 3552 | Scotland/EDB2050/2020 | 2020-04-18 | Virology Department, Royal Infirmary of Edinburgh, NHS Lothian / School of Biological Sciences, University of Edinburgh / Institute of Genetics and Molecular Medicine, University of Edinburgh | COVID-19 Genomics UK (COG-UK) Consortium | McHugh M et al |
| 3553 | USA/WI-UW-316/2020 | 2020-04-18 | University of Wisconsin-Madison AIDS Vaccine Research Laboratories | University of Wisconsin-Madison AIDS Vaccine Research Laboratories | Gage Moreno et al |
| 3554 | USA/WI-260/2020 | 2020-04-18 | University of Wisconsin-Madison AIDS Vaccine Research Laboratories | University of Wisconsin-Madison AIDS Vaccine Research Laboratories | Moreno et al |
| 3555 | Scotland/EDB2094/2020 | 2020-04-18 | Virology Department, Royal Infirmary of Edinburgh, NHS Lothian / School of Biological Sciences, University of Edinburgh / Institute of Genetics and Molecular Medicine, University of Edinburgh | COVID-19 Genomics UK (COG-UK) Consortium | McHugh M et al |
| 3556 | Scotland/EDB2057/2020 | 2020-04-18 | Virology Department, Royal Infirmary of Edinburgh, NHS Lothian / School of Biological Sciences, University of Edinburgh / Institute of | COVID-19 Genomics UK (COG-UK) Consortium | McHugh M et al |

| | | | | | |
|---|---|---|---|---|---|
| | | | Genetics and Molecular Medicine, University of Edinburgh | | |
| 3557 | Scotland/EDB2049/2020 | 2020-04-18 | Virology Department, Royal Infirmary of Edinburgh, NHS Lothian / School of Biological Sciences, University of Edinburgh / Institute of Genetics and Molecular Medicine, University of Edinburgh | COVID-19 Genomics UK (COG-UK) Consortium | McHugh M et al |
| 3558 | Scotland/EDB2064/2020 | 2020-04-18 | Virology Department, Royal Infirmary of Edinburgh, NHS Lothian / School of Biological Sciences, University of Edinburgh / Institute of Genetics and Molecular Medicine, University of Edinburgh | COVID-19 Genomics UK (COG-UK) Consortium | McHugh M et al |
| 3559 | Argentina/PAIS_A024/2020 | 2020-04-18 | Laboratorio de Virología del Hospital de Niños Dr. Ricardo Gutierrez | Área de Secuenciación del Laboratorio de Virología del Hospital de Niños Dr. Ricardo Gutierrez | Nabaes Jodar et al |
| 3560 | Scotland/EDB2062/2020 | 2020-04-18 | Virology Department, Royal Infirmary of Edinburgh, NHS Lothian / School of Biological Sciences, University of Edinburgh / Institute of Genetics and Molecular Medicine, University of Edinburgh | COVID-19 Genomics UK (COG-UK) Consortium | McHugh M et al |
| 3561 | USA/CA-CZB-1030/2020 | 2020-04-18 | County of Santa Clara Public Health Department | Chan-Zuckerberg Biohub | CZB Cliahub Consortium et al |
| 3562 | Scotland/EDB2051/2020 | 2020-04-18 | Virology Department, Royal Infirmary of Edinburgh, NHS Lothian / School of Biological Sciences, University of Edinburgh / Institute of Genetics and Molecular Medicine, University of Edinburgh | COVID-19 Genomics UK (COG-UK) Consortium | McHugh M et al |
| 3563 | Scotland/EDB2048/2020 | 2020-04-18 | Virology Department, Royal Infirmary of Edinburgh, NHS Lothian / School of Biological Sciences, University of Edinburgh / Institute of Genetics and Molecular Medicine, University of Edinburgh | COVID-19 Genomics UK (COG-UK) Consortium | McHugh M et al |
| 3564 | Scotland/EDB2011/2020 | 2020-04-18 | Virology Department, Royal Infirmary of Edinburgh, NHS Lothian / School of Biological Sciences, University of Edinburgh / Institute of Genetics and Molecular Medicine, University of Edinburgh | COVID-19 Genomics UK (COG-UK) Consortium | McHugh M et al |
| 3565 | Scotland/EDB2095/2020 | 2020-04-18 | Virology Department, Royal Infirmary of Edinburgh, NHS Lothian / School of Biological Sciences, University of Edinburgh / Institute of Genetics and Molecular Medicine, University of Edinburgh | COVID-19 Genomics UK (COG-UK) Consortium | McHugh M et al |
| 3566 | Argentina/PAIS_A026/2020 | 2020-04-18 | Laboratorio de Virología del Hospital de Niños Dr. Ricardo Gutierrez | Área de Secuenciación del Laboratorio de Virología del Hospital de Niños Dr. Ricardo Gutierrez | Nabaes Jodar et al |
| 3567 | Scotland/EDB2065/2020 | 2020-04-18 | Virology Department, Royal Infirmary of Edinburgh, NHS Lothian / School of Biological Sciences, University of Edinburgh / Institute of | COVID-19 Genomics UK (COG-UK) Consortium | McHugh M et al |

| | | | | | |
|---|---|---|---|---|---|
| | | | Genetics and Molecular Medicine, University of Edinburgh | | |
| 3568 | Singapore/84/2020 | 2020-04-18 | National Public Health Laboratory, National Centre for Infectious Diseases | National Public Health Laboratory, National Centre for Infectious Diseases | Mak Tze Minn et al |
| 3569 | Argentina/PAIS_A025/2020 | 2020-04-18 | Laboratorio de Virología del Hospital de Niños Dr. Ricardo Gutierrez | Área de Secuenciación del Laboratorio de Virología del Hospital de Niños Dr. Ricardo Gutierrez | Nabaes Jodar et al |
| 3570 | Scotland/EDB2078/2020 | 2020-04-19 | Virology Department, Royal Infirmary of Edinburgh, NHS Lothian / School of Biological Sciences, University of Edinburgh / Institute of Genetics and Molecular Medicine, University of Edinburgh | COVID-19 Genomics UK (COG-UK) Consortium | McHugh M et al |
| 3571 | Scotland/EDB2097/2020 | 2020-04-19 | Virology Department, Royal Infirmary of Edinburgh, NHS Lothian / School of Biological Sciences, University of Edinburgh / Institute of Genetics and Molecular Medicine, University of Edinburgh | COVID-19 Genomics UK (COG-UK) Consortium | McHugh M et al |
| 3572 | USA/WI-UW-318/2020 | 2020-04-19 | University of Wisconsin-Madison AIDS Vaccine Research Laboratories | University of Wisconsin-Madison AIDS Vaccine Research Laboratories | Gage Moreno et al |
| 3573 | Scotland/EDB2086/2020 | 2020-04-19 | Virology Department, Royal Infirmary of Edinburgh, NHS Lothian / School of Biological Sciences, University of Edinburgh / Institute of Genetics and Molecular Medicine, University of Edinburgh | COVID-19 Genomics UK (COG-UK) Consortium | McHugh M et al |
| 3574 | Scotland/EDB2089/2020 | 2020-04-19 | Virology Department, Royal Infirmary of Edinburgh, NHS Lothian / School of Biological Sciences, University of Edinburgh / Institute of Genetics and Molecular Medicine, University of Edinburgh | COVID-19 Genomics UK (COG-UK) Consortium | McHugh M et al |
| 3575 | Singapore/85/2020 | 2020-04-19 | National Public Health Laboratory, National Centre for Infectious Diseases | National Public Health Laboratory, National Centre for Infectious Diseases | Mak Tze Minn et al |
| 3576 | Scotland/EDB2077/2020 | 2020-04-19 | Virology Department, Royal Infirmary of Edinburgh, NHS Lothian / School of Biological Sciences, University of Edinburgh / Institute of Genetics and Molecular Medicine, University of Edinburgh | COVID-19 Genomics UK (COG-UK) Consortium | McHugh M et al |
| 3577 | Scotland/EDB2092/2020 | 2020-04-19 | Virology Department, Royal Infirmary of Edinburgh, NHS Lothian / School of Biological Sciences, University of Edinburgh / Institute of Genetics and Molecular Medicine, University of Edinburgh | COVID-19 Genomics UK (COG-UK) Consortium | McHugh M et al |
| 3578 | Scotland/EDB2087/2020 | 2020-04-19 | Virology Department, Royal Infirmary of Edinburgh, NHS Lothian / School of Biological Sciences, University of Edinburgh / Institute of Genetics and Molecular Medicine, University of Edinburgh | COVID-19 Genomics UK (COG-UK) Consortium | McHugh M et al |
| 3579 | Scotland/EDB2084/2020 | 2020-04-19 | Virology Department, Royal Infirmary of Edinburgh, NHS | COVID-19 Genomics UK (COG-UK) Consortium | McHugh M et al |

| | | | | | |
|---|---|---|---|---|---|
| | | | Lothian / School of Biological Sciences, University of Edinburgh / Institute of Genetics and Molecular Medicine, University of Edinburgh | | |
| 3580 | Scotland/EDB2085/2020 | 2020-04-19 | Virology Department, Royal Infirmary of Edinburgh, NHS Lothian / School of Biological Sciences, University of Edinburgh / Institute of Genetics and Molecular Medicine, University of Edinburgh | COVID-19 Genomics UK (COG-UK) Consortium | McHugh M et al |
| 3581 | Scotland/EDB2096/2020 | 2020-04-19 | Virology Department, Royal Infirmary of Edinburgh, NHS Lothian / School of Biological Sciences, University of Edinburgh / Institute of Genetics and Molecular Medicine, University of Edinburgh | COVID-19 Genomics UK (COG-UK) Consortium | McHugh M et al |
| 3582 | Scotland/EDB2088/2020 | 2020-04-19 | Virology Department, Royal Infirmary of Edinburgh, NHS Lothian / School of Biological Sciences, University of Edinburgh / Institute of Genetics and Molecular Medicine, University of Edinburgh | COVID-19 Genomics UK (COG-UK) Consortium | McHugh M et al |
| 3583 | Scotland/EDB2098/2020 | 2020-04-19 | Virology Department, Royal Infirmary of Edinburgh, NHS Lothian / School of Biological Sciences, University of Edinburgh / Institute of Genetics and Molecular Medicine, University of Edinburgh | COVID-19 Genomics UK (COG-UK) Consortium | McHugh M et al |
| 3584 | Scotland/EDB2082/2020 | 2020-04-19 | Virology Department, Royal Infirmary of Edinburgh, NHS Lothian / School of Biological Sciences, University of Edinburgh / Institute of Genetics and Molecular Medicine, University of Edinburgh | COVID-19 Genomics UK (COG-UK) Consortium | McHugh M et al |
| 3585 | Taiwan/CGMH-CGU-27/2020 | 2020-04-19 | Laboratory Medicine | Department of Laboratory Medicine, Lin-Kou Chang Gung Memorial Hospital, Taoyuan, Taiwan | Kuo-Chien Tsao et al |
| 3586 | Taiwan/CGMH-CGU-26/2020 | 2020-04-19 | Laboratory Medicine | Department of Laboratory Medicine, Lin-Kou Chang Gung Memorial Hospital, Taoyuan, Taiwan | Kuo-Chien Tsao et al |
| 3587 | Scotland/EDB2079/2020 | 2020-04-19 | Virology Department, Royal Infirmary of Edinburgh, NHS Lothian / School of Biological Sciences, University of Edinburgh / Institute of Genetics and Molecular Medicine, University of Edinburgh | COVID-19 Genomics UK (COG-UK) Consortium | McHugh M et al |
| 3588 | USA/NY-NYUMC469/2020 | 2020-04-20 | NYU Langone Health | Departments of Pathology and Medicine, New York University School of Medicine | Maria Aguero-Rosenfeld et al |
| 3589 | USA/NY-NYUMC433/2020 | 2020-04-20 | NYU Langone Health | Departments of Pathology and Medicine, New York University School of Medicine | Maria Aguero-Rosenfeld et al |
| 3590 | USA/NY-NYUMC449/2020 | 2020-04-20 | NYU Langone Health | Departments of Pathology and Medicine, New York University School of Medicine | Maria Aguero-Rosenfeld et al |
| 3591 | USA/NY-NYUMC459/2020 | 2020-04-20 | NYU Langone Health | Departments of Pathology and Medicine, New York University School of Medicine | Maria Aguero-Rosenfeld et al |

| | | | | | |
|---|---|---|---|---|---|
| 3592 | USA/NY-NYUMC455/2020 | 2020-04-20 | NYU Langone Health | Departments of Pathology and Medicine, New York University School of Medicine | Maria Aguero-Rosenfeld et al |
| 3593 | USA/NJ-NYUMC452/2020 | 2020-04-20 | NYU Langone Health | Departments of Pathology and Medicine, New York University School of Medicine | Maria Aguero-Rosenfeld et al |
| 3594 | USA/NY-NYUMC432/2020 | 2020-04-20 | NYU Langone Health | Departments of Pathology and Medicine, New York University School of Medicine | Maria Aguero-Rosenfeld et al |
| 3595 | USA/NY-NYUMC462/2020 | 2020-04-20 | NYU Langone Health | Departments of Pathology and Medicine, New York University School of Medicine | Maria Aguero-Rosenfeld et al |
| 3596 | India/NCDC-4859/2020 | 2020-04-20 | National Centre for Disease control (NCDC) | NCDC/CSIR-IGIB | Pramod Kumar et al |
| 3597 | USA/CA-CZB-1042/2020 | 2020-04-20 | County of Santa Clara Public Health Department | Chan-Zuckerberg Biohub | CZB Cliahub Consortium et al |
| 3598 | India/NCDC-4876/2020 | 2020-04-20 | National Centre for Disease control (NCDC) | NCDC/CSIR-IGIB | Pramod Kumar et al |
| 3599 | USA/NY-NYUMC473/2020 | 2020-04-20 | NYU Langone Health | Departments of Pathology and Medicine, New York University School of Medicine | Maria Aguero-Rosenfeld et al |
| 3600 | USA/NY-NYUMC458/2020 | 2020-04-20 | NYU Langone Health | Departments of Pathology and Medicine, New York University School of Medicine | Maria Aguero-Rosenfeld et al |
| 3601 | USA/NY-NYUMC453/2020 | 2020-04-20 | NYU Langone Health | Departments of Pathology and Medicine, New York University School of Medicine | Maria Aguero-Rosenfeld et al |
| 3602 | USA/NY-NYUMC451/2020 | 2020-04-20 | NYU Langone Health | Departments of Pathology and Medicine, New York University School of Medicine | Maria Aguero-Rosenfeld et al |
| 3603 | USA/LA-EVTL039/2020 | 2020-04-20 | LSUHS Emerging Viral Threat Laboratory | Microbial Genome Sequencing Center | Jeremy P. Kamil et al |
| 3604 | USA/NY-NYUMC457/2020 | 2020-04-20 | NYU Langone Health | Departments of Pathology and Medicine, New York University School of Medicine | Maria Aguero-Rosenfeld et al |
| 3605 | USA/NY-NYUMC460/2020 | 2020-04-20 | NYU Langone Health | Departments of Pathology and Medicine, New York University School of Medicine | Maria Aguero-Rosenfeld et al |
| 3606 | USA/NY-NYUMC448/2020 | 2020-04-20 | NYU Langone Health | Departments of Pathology and Medicine, New York University School of Medicine | Maria Aguero-Rosenfeld et al |
| 3607 | USA/NY-NYUMC436/2020 | 2020-04-20 | NYU Langone Health | Departments of Pathology and Medicine, New York University School of Medicine | Maria Aguero-Rosenfeld et al |
| 3608 | USA/NY-NYUMC461/2020 | 2020-04-20 | NYU Langone Health | Departments of Pathology and Medicine, New York University School of Medicine | Maria Aguero-Rosenfeld et al |
| 3609 | USA/NY-NYUMC456/2020 | 2020-04-20 | NYU Langone Health | Departments of Pathology and Medicine, New York University School of Medicine | Maria Aguero-Rosenfeld et al |
| 3610 | India/NCDC-4874/2020 | 2020-04-20 | National Centre for Disease control (NCDC) | NCDC/CSIR-IGIB | Pramod Kumar et al |
| 3611 | USA/NY-NYUMC471/2020 | 2020-04-20 | NYU Langone Health | Departments of Pathology and Medicine, New York University School of Medicine | Maria Aguero-Rosenfeld et al |
| 3612 | Sweden/20-07616/2020 | 2020-04-20 | Surbrunns VC | The Public Health Agency of Sweden | Erik Embring et al |
| 3613 | USA/NY-NYUMC425/2020 | 2020-04-20 | NYU Langone Health | Departments of Pathology and Medicine, New York University School of Medicine | Maria Aguero-Rosenfeld et al |
| 3614 | USA/NY-NYUMC472/2020 | 2020-04-20 | NYU Langone Health | Departments of Pathology and Medicine, New York University School of Medicine | Maria Aguero-Rosenfeld et al |
| 3615 | USA/NY-NYUMC450/2020 | 2020-04-20 | NYU Langone Health | Departments of Pathology and Medicine, New York University School of Medicine | Maria Aguero-Rosenfeld et al |
| 3616 | Singapore/86/2020 | 2020-04-20 | National Public Health Laboratory, National Centre for Infectious Diseases | National Public Health Laboratory, National Centre for Infectious Diseases | Mak Tze Minn et al |
| 3617 | USA/NY-NYUMC454/2020 | 2020-04-20 | NYU Langone Health | Departments of Pathology and | Maria |

| | | | | | |
|---|---|---|---|---|---|
| | | | | Medicine, New York University School of Medicine | Aguero-Rosenfeld et al |
| 3618 | India/NCDC-4877/2020 | 2020-04-20 | National Centre for Disease control (NCDC) | NCDC/CSIR-IGIB | Pramod Kumar et al |
| 3619 | Sweden/20-07607/2020 | 2020-04-20 | Surbrunns VC | The Public Health Agency of Sweden | Erik Embring et al |
| 3620 | Singapore/91/2020 | 2020-04-20 | National Public Health Laboratory, National Centre for Infectious Diseases | National Public Health Laboratory, National Centre for Infectious Diseases | Mak Tze Minn et al |
| 3621 | Luxembourg/LNS1673015/2020 | 2020-04-21 | Laboratoire National de Sante, Microbiology, Virology | Laboratoire National de Sante, Microbiology, Epidemiology and Microbial Genomics | Anke Wienecke-Baldacchino et al |
| 3622 | USA/NY-NYUMC475/2020 | 2020-04-21 | NYU Langone Health | Departments of Pathology and Medicine, New York University School of Medicine | Maria Aguero-Rosenfeld et al |
| 3623 | USA/WI-UW-346/2020 | 2020-04-21 | University of Wisconsin-Madison AIDS Vaccine Research Laboratories | University of Wisconsin-Madison AIDS Vaccine Research Laboratories | Gage Moreno et al |
| 3624 | USA/WI-GMF-01455/2020 | 2020-04-21 | Gundersen Molecular Diagnostics Laboratory | Kabara Cancer Research Institute | Craig S. Richmond et al |
| 3625 | USA/WI-UW-320/2020 | 2020-04-21 | University of Wisconsin-Madison AIDS Vaccine Research Laboratories | University of Wisconsin-Madison AIDS Vaccine Research Laboratories | Gage Moreno et al |
| 3626 | USA/NY-NYUMC430/2020 | 2020-04-21 | NYU Langone Health | Departments of Pathology and Medicine, New York University School of Medicine | Maria Aguero-Rosenfeld et al |
| 3627 | USA/NY-NYUMC474/2020 | 2020-04-21 | NYU Langone Health | Departments of Pathology and Medicine, New York University School of Medicine | Maria Aguero-Rosenfeld et al |
| 3628 | USA/WI-GMF-01535/2020 | 2020-04-21 | Gundersen Molecular Diagnostics Laboratory | Kabara Cancer Research Institute | Craig S. Richmond et al |
| 3629 | USA/WI-GMF-01481/2020 | 2020-04-21 | Gundersen Molecular Diagnostics Laboratory | Kabara Cancer Research Institute | Craig S. Richmond et al |
| 3630 | USA/WI-UW-319/2020 | 2020-04-21 | University of Wisconsin-Madison AIDS Vaccine Research Laboratories | University of Wisconsin-Madison AIDS Vaccine Research Laboratories | Gage Moreno et al |
| 3631 | USA/NY-NYUMC476/2020 | 2020-04-21 | NYU Langone Health | Departments of Pathology and Medicine, New York University School of Medicine | Maria Aguero-Rosenfeld et al |
| 3632 | India/GBRC9/2020 | 2020-04-21 | Gujarat Biotechnology Research Centre | Gujarat Biotechnology Research Centre | Maharshi Pandya et al |
| 3633 | Sweden/20-07572/2020 | 2020-04-21 | Narhalsan Backa vardcentral | The Public Health Agency of Sweden | Mats Olsson et al |
| 3634 | Singapore/88/2020 | 2020-04-21 | National Public Health Laboratory, National Centre for Infectious Diseases | National Public Health Laboratory, National Centre for Infectious Diseases | Mak Tze Minn et al |
| 3635 | Luxembourg/LNS2435702/2020 | 2020-04-21 | Laboratoire National de Sante, Microbiology, Virology | Laboratoire National de Sante, Microbiology, Epidemiology and Microbial Genomics | Anke Wienecke-Baldacchino et al |
| 3636 | Singapore/87/2020 | 2020-04-21 | National Public Health Laboratory, National Centre for Infectious Diseases | National Public Health Laboratory, National Centre for Infectious Diseases | Mak Tze Minn et al |
| 3637 | USA/NY-NYUMC424/2020 | 2020-04-22 | NYU Langone Health | Departments of Pathology and Medicine, New York University School of Medicine | Maria Aguero-Rosenfeld et al |
| 3638 | USA/NY-NYUMC420/2020 | 2020-04-22 | NYU Langone Health | Departments of Pathology and Medicine, New York University School of Medicine | Maria Aguero-Rosenfeld et al |
| 3639 | USA/NY-NYUMC478/2020 | 2020-04-22 | NYU Langone Health | Departments of Pathology and Medicine, New York University School of Medicine | Maria Aguero-Rosenfeld et al |
| 3640 | USA/NY-NYUMC477/2020 | 2020-04-22 | NYU Langone Health | Departments of Pathology and Medicine, New York University School of Medicine | Maria Aguero-Rosenfeld et al |
| 3641 | USA/NY-NYUMC479/2020 | 2020-04-22 | NYU Langone Health | Departments of Pathology and Medicine, New York University School of Medicine | Maria Aguero-Rosenfeld et al |
| 3642 | USA/NJ-NYUMC421/2020 | 2020-04-22 | NYU Langone Health | Departments of Pathology and Medicine, New York University School of Medicine | Maria Aguero-Rosenfeld et al |

| | | | | | |
|---|---|---|---|---|---|
| 3643 | USA/NY-NYUMC423/2020 | 2020-04-22 | NYU Langone Health | Departments of Pathology and Medicine, New York University School of Medicine | Maria Aguero-Rosenfeld et al |
| 3644 | Singapore/89/2020 | 2020-04-22 | National Public Health Laboratory, National Centre for Infectious Diseases | National Public Health Laboratory, National Centre for Infectious Diseases | Mak Tze Minn et al |
| 3645 | USA/WI-GMF-01551/2020 | 2020-04-22 | Gundersen Molecular Diagnostics Laboratory | Kabara Cancer Research Institute | Craig S. Richmond et al |
| 3646 | USA/WI-GMF-01591/2020 | 2020-04-23 | Gundersen Molecular Diagnostics Laboratory | Kabara Cancer Research Institute | Craig S. Richmond et al |
| 3647 | USA/NY-NYUMC480/2020 | 2020-04-23 | NYU Langone Health | Departments of Pathology and Medicine, New York University School of Medicine | Maria Aguero-Rosenfeld et al |
| 3648 | USA/WI-UW-321/2020 | 2020-04-23 | University of Wisconsin-Madison AIDS Vaccine Research Laboratories | University of Wisconsin-Madison AIDS Vaccine Research Laboratories | Gage Moreno et al |
| 3649 | USA/WI-GMF-01602/2020 | 2020-04-23 | Gundersen Molecular Diagnostics Laboratory | Kabara Cancer Research Institute | Craig S. Richmond et al |
| 3650 | USA/WI-UW-322/2020 | 2020-04-23 | University of Wisconsin-Madison AIDS Vaccine Research Laboratories | University of Wisconsin-Madison AIDS Vaccine Research Laboratories | Gage Moreno et al |
| 3651 | USA/WI-UW-323/2020 | 2020-04-23 | University of Wisconsin-Madison AIDS Vaccine Research Laboratories | University of Wisconsin-Madison AIDS Vaccine Research Laboratories | Gage Moreno et al |
| 3652 | Singapore/90/2020 | 2020-04-23 | National Public Health Laboratory, National Centre for Infectious Diseases | National Public Health Laboratory, National Centre for Infectious Diseases | Mak Tze Minn et al |
| 3653 | USA/LA-EVTL032/2020 | 2020-04-24 | LSUHS Emerging Viral Threat Laboratory | Microbial Genome Sequencing Center | Jeremy P. Kamil et al |
| 3654 | USA/LA-EVTL021/2020 | 2020-04-24 | LSUHS Emerging Viral Threat Laboratory | Microbial Genome Sequencing Center | John A. Vanchiere et al |
| 3655 | USA/LA-EVTL029/2020 | 2020-04-24 | LSUHS Emerging Viral Threat Laboratory | Microbial Genome Sequencing Center | John A. Vanchiere et al |
| 3656 | USA/LA-EVTL031/2020 | 2020-04-24 | LSUHS Emerging Viral Threat Laboratory | Microbial Genome Sequencing Center | Jeremy P. Kamil et al |
| 3657 | USA/LA-EVTL041/2020 | 2020-04-24 | LSUHS Emerging Viral Threat Laboratory | Microbial Genome Sequencing Center | Jeremy P. Kamil et al |
| 3658 | USA/WI-UW-324/2020 | 2020-04-24 | University of Wisconsin-Madison AIDS Vaccine Research Laboratories | University of Wisconsin-Madison AIDS Vaccine Research Laboratories | Gage Moreno et al |
| 3659 | USA/LA-EVTL023/2020 | 2020-04-24 | LSUHS Emerging Viral Threat Laboratory | Microbial Genome Sequencing Center | John A. Vanchiere et al |
| 3660 | USA/LA-EVTL028/2020 | 2020-04-24 | LSUHS Emerging Viral Threat Laboratory | Microbial Genome Sequencing Center | John A. Vanchiere et al |
| 3661 | USA/LA-EVTL035/2020 | 2020-04-24 | LSUHS Emerging Viral Threat Laboratory | Microbial Genome Sequencing Center | Jeremy P. Kamil et al |
| 3662 | USA/LA-EVTL027/2020 | 2020-04-24 | LSUHS Emerging Viral Threat Laboratory | Microbial Genome Sequencing Center | John A. Vanchiere et al |
| 3663 | USA/LA-EVTL026/2020 | 2020-04-24 | LSUHS Emerging Viral Threat Laboratory | Microbial Genome Sequencing Center | John A. Vanchiere et al |
| 3664 | USA/NY-NYUMC483/2020 | 2020-04-24 | NYU Langone Health | Departments of Pathology and Medicine, New York University School of Medicine | Maria Aguero-Rosenfeld et al |
| 3665 | USA/LA-EVTL030/2020 | 2020-04-24 | LSUHS Emerging Viral Threat Laboratory | Microbial Genome Sequencing Center | John A. Vanchiere et al |
| 3666 | USA/LA-EVTL022/2020 | 2020-04-24 | LSUHS Emerging Viral Threat Laboratory | Microbial Genome Sequencing Center | John A. Vanchiere et al |
| 3667 | USA/LA-EVTL040/2020 | 2020-04-24 | LSUHS Emerging Viral Threat Laboratory | Microbial Genome Sequencing Center | Jeremy P. Kamil et al |
| 3668 | USA/NY-NYUMC435/2020 | 2020-04-24 | NYU Langone Health | Departments of Pathology and Medicine, New York University School of Medicine | Maria Aguero-Rosenfeld et al |
| 3669 | USA/LA-EVTL019/2020 | 2020-04-24 | LSUHS Emerging Viral Threat Laboratory | Microbial Genome Sequencing Center | John A. Vanchiere et al |
| 3670 | USA/WI-UW-325/2020 | 2020-04-24 | University of Wisconsin-Madison AIDS Vaccine Research Laboratories | University of Wisconsin-Madison AIDS Vaccine Research Laboratories | Gage Moreno et al |
| 3671 | USA/LA-EVTL017/2020 | 2020-04-24 | LSUHS Emerging Viral Threat | Microbial Genome Sequencing | John A. Vanchiere et al |

| | | | | | |
|---|---|---|---|---|---|
| | | | Laboratory | Center | |
| 3672 | USA/LA-EVTL020/2020 | 2020-04-24 | LSUHS Emerging Viral Threat Laboratory | Microbial Genome Sequencing Center | John A. Vanchiere et al |
| 3673 | USA/LA-EVTL034/2020 | 2020-04-24 | LSUHS Emerging Viral Threat Laboratory | Microbial Genome Sequencing Center | Jeremy P. Kamil et al |
| 3674 | USA/LA-EVTL037/2020 | 2020-04-24 | LSUHS Emerging Viral Threat Laboratory | Microbial Genome Sequencing Center | Jeremy P. Kamil et al |
| 3675 | USA/LA-EVTL036/2020 | 2020-04-24 | LSUHS Emerging Viral Threat Laboratory | Microbial Genome Sequencing Center | Jeremy P. Kamil et al |
| 3676 | USA/LA-EVTL025/2020 | 2020-04-24 | LSUHS Emerging Viral Threat Laboratory | Microbial Genome Sequencing Center | John A. Vanchiere et al |
| 3677 | USA/NY-NYUMC481/2020 | 2020-04-24 | NYU Langone Health | Departments of Pathology and Medicine, New York University School of Medicine | Maria Aguero-Rosenfeld et al |
| 3678 | USA/LA-EVTL033/2020 | 2020-04-24 | LSUHS Emerging Viral Threat Laboratory | Microbial Genome Sequencing Center | Jeremy P. Kamil et al |
| 3679 | USA/LA-EVTL018/2020 | 2020-04-24 | LSUHS Emerging Viral Threat Laboratory | Microbial Genome Sequencing Center | John A. Vanchiere et al |
| 3680 | USA/LA-EVTL038/2020 | 2020-04-24 | LSUHS Emerging Viral Threat Laboratory | Microbial Genome Sequencing Center | Jeremy P. Kamil et al |
| 3681 | USA/LA-EVTL024/2020 | 2020-04-24 | LSUHS Emerging Viral Threat Laboratory | Microbial Genome Sequencing Center | John A. Vanchiere et al |
| 3682 | USA/NY-NYUMC484/2020 | 2020-04-24 | NYU Langone Health | Departments of Pathology and Medicine, New York University School of Medicine | Maria Aguero-Rosenfeld et al |
| 3683 | NewZealand/20VR3038/2020-04-25 | 2020-04-25 | Wellington SCL | Institute of Environmental Science and Research (ESR) | **Matt Storey et al** |
| 3684 | USA/LA-EVTL014/2020 | 2020-04-25 | LSUHS Emerging Viral Threat Laboratory | Microbial Genome Sequencing Center | Rona S. Scott et al |
| 3685 | USA/WI-UW-329/2020 | 2020-04-25 | University of Wisconsin-Madison AIDS Vaccine Research Laboratories | University of Wisconsin-Madison AIDS Vaccine Research Laboratories | Gage Moreno et al |
| 3686 | NewZealand/20VR2931/2020-04-25 | 2020-04-25 | LabPLUS | Institute of Environmental Science and Research (ESR) | **Matt Storey et al** |
| 3687 | USA/LA-EVTL012/2020 | 2020-04-25 | LSUHS Emerging Viral Threat Laboratory | Microbial Genome Sequencing Center | Rona S. Scott et al |
| 3688 | USA/LA-EVTL016/2020 | 2020-04-25 | LSUHS Emerging Viral Threat Laboratory | Microbial Genome Sequencing Center | Rona S. Scott et al |
| 3689 | USA/LA-EVTL013/2020 | 2020-04-25 | LSUHS Emerging Viral Threat Laboratory | Microbial Genome Sequencing Center | Rona S. Scott et al |
| 3690 | USA/WI-UW-326/2020 | 2020-04-25 | University of Wisconsin-Madison AIDS Vaccine Research Laboratories | University of Wisconsin-Madison AIDS Vaccine Research Laboratories | Gage Moreno et al |
| 3691 | USA/LA-EVTL015/2020 | 2020-04-25 | LSUHS Emerging Viral Threat Laboratory | Microbial Genome Sequencing Center | Rona S. Scott et al |
| 3692 | USA/WI-UW-327/2020 | 2020-04-25 | University of Wisconsin-Madison AIDS Vaccine Research Laboratories | University of Wisconsin-Madison AIDS Vaccine Research Laboratories | Gage Moreno et al |
| 3693 | NewZealand/20VR3034/2020-04-25 | 2020-04-25 | Wellington SCL | Institute of Environmental Science and Research (ESR) | **Matt Storey et al** |
| 3694 | NewZealand/20VR2922/2020-04-25 | 2020-04-25 | LabPLUS | Institute of Environmental Science and Research (ESR) | **Matt Storey et al** |
| 3695 | NewZealand/20VR3027/2020-04-25 | 2020-04-25 | Wellington SCL | Institute of Environmental Science and Research (ESR) | **Matt Storey et al** |
| 3696 | NewZealand/20VR2717/2020-04-25 | 2020-04-25 | LabPLUS | Institute of Environmental Science and Research (ESR) | **Matt Storey et al** |
| 3697 | NewZealand/20VR2659/2020-04-25 | 2020-04-25 | LabPLUS | Institute of Environmental Science and Research (ESR) | **Matt Storey et al** |
| 3698 | USA/WI-UW-271/2020 | 2020-04-26 | University of Wisconsin-Madison AIDS Vaccine Research Laboratories | University of Wisconsin-Madison AIDS Vaccine Research Laboratories | Gage Moreno et al |
| 3699 | USA/WI-UW-332/2020 | 2020-04-26 | University of Wisconsin-Madison AIDS Vaccine Research Laboratories | University of Wisconsin-Madison AIDS Vaccine Research Laboratories | Gage Moreno et al |
| 3700 | USA/WI-GMF-01828/2020 | 2020-04-26 | Gundersen Molecular Diagnostics Laboratory | Kabara Cancer Research Institute | Craig S. Richmond et al |
| 3701 | USA/WI-UW-328/2020 | 2020-04-26 | University of Wisconsin-Madison AIDS | University of Wisconsin-Madison AIDS | Gage Moreno et al |

| | | | | | |
|---|---|---|---|---|---|
| | | | Vaccine Research Laboratories | Vaccine Research Laboratories | |
| 3702 | USA/WI-UW-331/2020 | 2020-04-26 | University of Wisconsin-Madison AIDS Vaccine Research Laboratories | University of Wisconsin-Madison AIDS Vaccine Research Laboratories | Gage Moreno et al |
| 3703 | USA/WI-UW-312/2020 | 2020-04-27 | University of Wisconsin-Madison AIDS Vaccine Research Laboratories | University of Wisconsin-Madison AIDS Vaccine Research Laboratories | Gage Moreno et al |
| 3704 | Taiwan/NTU28/2020 | 2020-04-27 | Department of Laboratory Medicine, National Taiwan University Hospital | Microbial Genomics Core Lab, National Taiwan University Centers of Genomic and Precision Medicine | Shiou-Hwei Yeh et al |
| 3705 | USA/CA-SR0175/2020 | 2020-04-28 | Rady's Childrens Hospital | Andersen lab at Scripps Research | SEARCH Alliance San Diego et al |
| 3706 | USA/CA-SR0176/2020 | 2020-04-29 | Rady's Childrens Hospital | Andersen lab at Scripps Research | SEARCH Alliance San Diego et al |
